# Assessing the utility of COVID-19 case reports as a leading indicator for hospitalization forecasting in the United States

**DOI:** 10.1101/2023.03.08.23286582

**Authors:** Nicholas G Reich, Yijin Wang, Meagan Burns, Rosa Ergas, Estee Y Cramer, Evan L Ray

## Abstract

Identifying data streams that can consistently improve the accuracy of epidemiological forecasting models is challenging. Using models designed to predict daily state-level hospital admissions due to COVID-19 in California and Massachusetts, we investigated whether incorporating COVID-19 case data systematically improved forecast accuracy. Additionally, we considered whether using case data aggregated by date of test or by date of report from a surveillance system made a difference to the forecast accuracy. Evaluating forecast accuracy in a test period, after first having selected the best-performing methods in a validation period, we found that overall the difference in accuracy between approaches was small, especially at forecast horizons of less than two weeks. However, forecasts from models using cases aggregated by test date showed lower accuracy at longer horizons and at key moments in the pandemic, such as the peak of the Omicron wave in January 2022. Overall, these results highlight the challenge of finding a modeling approach that can generate accurate forecasts of outbreak trends both during periods of relative stability and during periods that show rapid growth or decay of transmission rates. While COVID-19 case counts seem to be a natural choice to help predict COVID-19 hospitalizations, in practice any benefits we observed were small and inconsistent.

## Introduction

During the COVID-19 pandemic, real-time data signals from a variety of sources have provided important insights into trends of SARS-CoV-2 transmission for aggregated geospatial units, such as counties and states in the US. However, identifying which signals can provide reliable, consistent, and accurate indicators for monitoring epidemics generally poses a challenge, as many types of signals exist. One class of signals include data such as reported case numbers, hospitalizations, and deaths, that typically are reported through public health organizations or epidemiological data collection systems led by academic groups[1, 2]. Other signals include less commonly used sources of information on outbreaks, such as high-volume surveys on social media platforms, search query data, or data from electronic health records[3, 4, 5].

In the US, one of the most important and reliable indicators of SARS-CoV-2 transmission during the first three years of the pandemic has been the data on hospital admissions that are recorded as part of the HHS Protect system. These data exist at a daily level for individual facilities, and typically have a coverage of over 95% of all hospitals in the country each day. These data have been reported under a mandate from HHS since July 2020, meaning that compliance with reporting for this system has been high. However, these data are reported directly by facilities, with variance in the data collection and reporting methodology. These methods include manual data entry and place a burden on reporting facilities. At the time of writing, this mandate is in place until spring 2024. Additionally, hospitalizations are a relevant indicator for public health decision-making, as high levels are indicative of strain on the healthcare system.

Both prior to and during the COVID-19 pandemic, forecasts of public health indicators around outbreaks have been used as inputs to decision-making [6]. Hospitalization forecasts can provide useful information for healthcare providers to better allocate medical resources and communicate treatment plans [7]. There are clear examples of forecasts, in conjunction with other modeling and surveillance efforts, being used to guide local public health officials to make real-time operational decisions [8]. Accurate forecasts can improve decision making while inaccurate forecasts could negatively impact public health response. In this paper, we investigate the value added to forecasts of COVID-19 hospitalizations from the use of data measuring the number of COVID-19 cases reported through state public health surveillance systems.

Models have used a variety of data inputs to predict hospitalizations due to COVID-19. Some of these data inputs include previous hospitalization data [9, 10], population mobility data [11, 9], realtime hospital occupancy data [12], trends in genomic variants [13], and internet search queries and chats from a public-facing Health Bot [14]. Though forecasts of hospitalizations are important, these forecasts were sometimes inaccurate at different stages of the pandemic due to characteristics of the data inputs. For example, the utility of mobility data as a predictor of transmission and subsequent hospitalizations declined as the pandemic evolved and human behavior and response changed over time[15]. The utility of these data sources in informing disease transmission varies with public health precautionary policies [11], the capacity of healthcare systems, and missing data [12].

In addition to these data sources, one important data input often used to forecast hospitalizations is the number of reported cases in a geographic location [16, 11, 12]. Epidemiological intuition suggests that there should be a lagged relationship between diagnosis of a COVID-19 case and subsequent hospitalization. However, some cases are confirmed and reported upon a positive test when admitted to a hospital, suggesting that in some instances hospital admissions may not lag case reports.

Several papers have published models in which cases were used as inputs to forecast hospitalization metrics [17, 11, 9]. However these efforts do not provide clear evidence that the inclusion of case data as a “covariate” of the model materially improves forecast accuracy above and beyond including just data on prior hospitalizations. Also, none of this work includes modeling beyond July 2021 and therefore does not speak to how the relationship between reported cases and hospitalizations might have changed during the larger Delta and Omicron waves when at-home rapid antigen testing became more widespread.

During the first three years of the pandemic in the US, state-level public health agencies reported new cases due to COVID-19 in ways that changed over time. Some states reported case data recorded by the date that a public health agency released data including an individual’s positive test result, which we refer to throughout this work as the “report date”. This was also the standard method used by data aggregation teams at Johns Hopkins University Center for Systems Science and Engineering (JHU CSSE), New York Times, and USAFacts. A small number of states used alternate methods to record dates associated more closely with the actual onset of symptoms. For example, California makes case counts available by date (defined as the “episode date”) which is the “earliest of the … date received, date of diagnosis, date of symptom onset, specimen collection date, or date of death”[18]. Whereas Massachusetts makes case counts available by the date of a laboratory test confirming an individual has COVID-19. Because these methods are similar in that they try to define a date earlier than the “report date” and closer to symptom onset, we group these two data sources together and refer to them both as referring to a “test date” in the work that follows. While aggregating counts by report date has been standard practice during the first three years of the COVID-19 pandemic, counts aggregated by test date may be seen as more beneficial because they have more epidemiological relevance.

In this paper, we investigate the utility of reported COVID-19 cases as an input to forecasting daily new hospitalization admissions, using data from October 1, 2020 through July 26, 2022. We used data from Massachusetts and California, as these states had publicly available case data aggregated by test date on their Department of Public Health websites. We created multiple distinct time series models that use different versions of case data (aggregated by report date, aggregated by test date, and no case data). A subset of models that performed well in a validation period were used to create forecasts in an out-of-sample test period. The comparative performance of these models can help us assess the extent to which case data are or are not helpful inputs for predicting hospitalizations and whether case data by test date are more helpful than case data by report date.

## Methods

### Data

We used data from Massachusetts and California as the basis for our analysis because the Departments of Public Health from these states recorded and publicly reported state-level new cases aggregated by both report date and test date (Table 1 and Figure 1).

**Table 1:** A list of the data signals used for each state in the analyses presented in this work. Cases aggregated by date of report, or first appearance in a public reporting system, are called “report-date cases.” Cases aggregated by date of test are called “test-date cases.”

| Data signal | State | Source |
| --- | --- | --- |
| hospital admissions | CA | HHS protect [19] |
|  | MA | HHS protect][19] |
| report-date cases | CA | JHU CSSE[1] |
|  | CA | CA DPH[20] |
|  | MA | JHU CSSE[1] |
| test-date cases | CA | CA DPH[20] |
|  | MA | MA DPH[21] |

**Figure 1:**
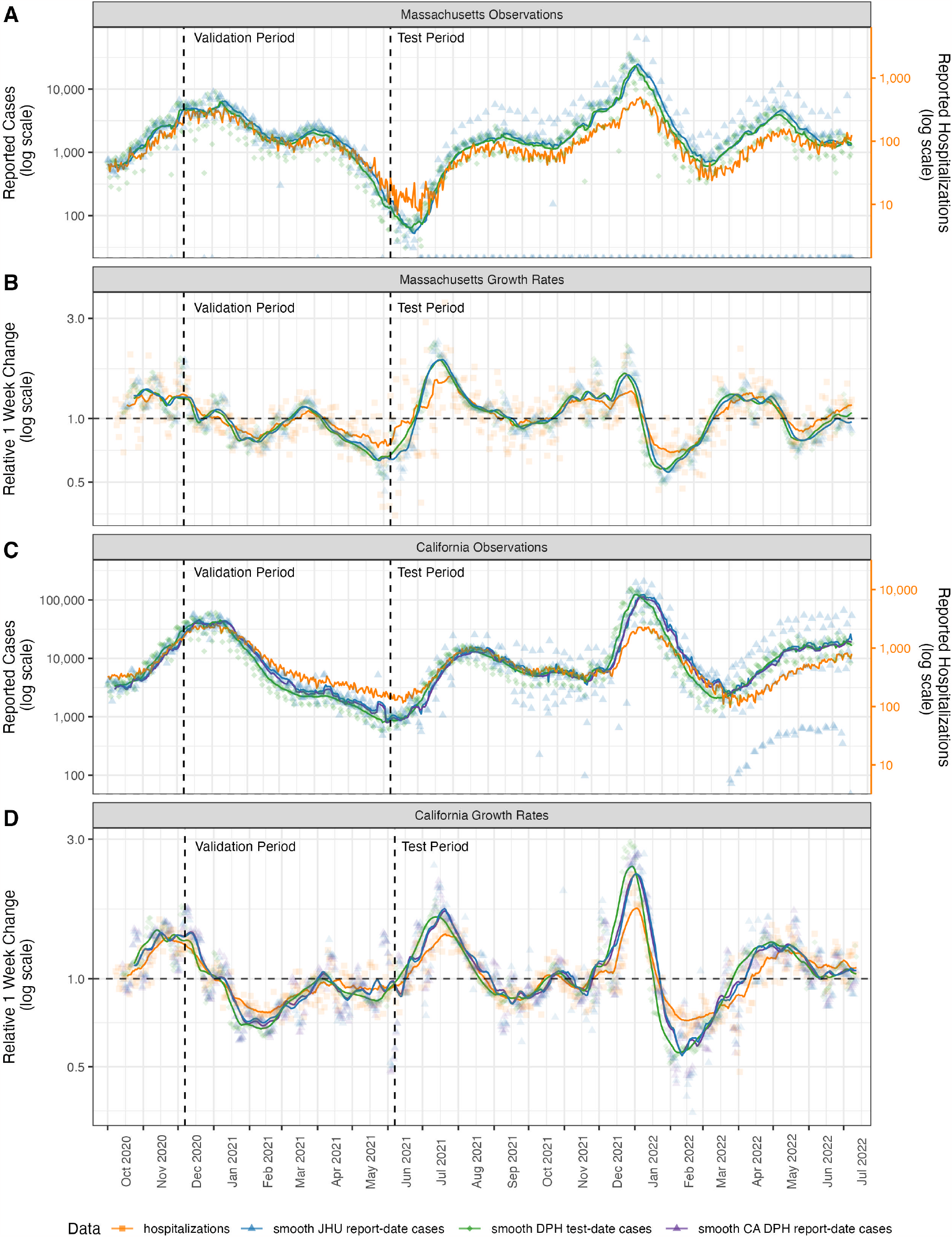
Observations and rates of change of case count data and hospitalization admission data. Panels A and B show observations and growth rates, respectively, for Massachusetts data sources. Panels C and D show observations and growth rates, respectively, for California data sources. For California only, the report-date case counts from CA DPH (purple triangles and lines) were different than those retrieved from JHU CSSE (blue triangles and lines), so both signals are shown. Test-date case counts reported by MA and CA DPH are shown in green diamonds and lines. Hospitalization data (orange squares and lines) were observed at roughly an order of magnitude lower than case data. Hospitalization data in panels A and C were scaled so they could be displayed on the same figure as case data and separate axes are provided on the right of those panels. The “Relative 1 week change” shown in panels B and D is computed for every day as 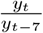. In all plots the lines represent moving averages of the raw data that compute a trailing 1-week average of the current day and the prior six days of observations.

We used data on both COVID-19 hospitalization admissions and new cases in our modeling experiments. COVID-19 hospitalization data for both Massachusetts and California were collected in the Department of Health & Human Services’ Protect Public Data Hub (HHS Protect)[19]. This centralized source contains time series data for daily hospitalization utilization at the facility and state level. For our analyses, we focused on the metric of new daily hospital admissions with a confirmed COVID-19 diagnosis. For some data summaries presented (e.g., Figure 1B and D), we computed a measure of relative one-week change for a given signal measured on day *t* as 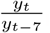.

For both states, test-date case data (daily counts of new confirmed COVID-19 cases, aggregated by date of test) were collected by the respective Departments of Public Health (CA DPH and MA DPH)[20, 21]. We downloaded data from these two web sources. Report-date case data (daily counts of new confirmed COVID-19 cases, aggregated by date on which the case was reported into a surveillance system) for both locations were collected and reported publicly by JHU CSSE. For California, due to differences in data aggregation methodologies, report-date case data reported publicly by CA DPH did not always agree with the same signal from JHU CSSE. We included both signals for completeness, although the 7-day smoothed averages of the two signals are virtually identical (Figure 1).

### Data vintage used in modeling

Data for both hospitalizations and cases are sometimes revised by the surveillance system after initially being reported. The revisions can sometimes be substantial and occur as much as weeks or months after the data were initially reported (Supplemental File 3). Data revisions have been shown to have a substantial impact on forecast accuracy[22, 23]. To focus our model comparisons on how models could perform in an idealized setting where the final data were available in real-time, we fit and evaluated all models based on a “finalized” version of the data retrieved on July 26, 2022. Since the latest date for which forecasts were evaluated was June 26, 2022 (based on final forecast date of May 30, 2022, see “Forecast validation” section below), this allowed for a roughly one-month period to allow the latest data used in evaluation to be revised. Nonetheless, some of the more recent data in 2022 may have experienced revisions after our final date of retrieval.

In the experiments described below, forecasts were generated for each week, where the model was given finalized data from October 1, 2020 through the Monday of that week. While forecasts were created in retrospect, this weekly “timing” of forecast generation mimics the scheduled generation of real-time forecasts from the COVID-19 Forecast Hub[24]. However, the data that would have been available in real-time on a given Monday would in practice include from one to three fewer days of data. For instance, Monday counts are typically not released on that same day, and yet we allowed our models to see those data. Additionally, some values available in real time may have been subject to revision. Snapshots of data revisions are documented in Supplemental Files 1 and 3. These reporting effects impact the various signals differentially, and in the discussion we return to this point in the context of interpreting the results of our analysis.

### Models

#### Model variations based on data inputs

We specified four model variations to investigate the value of different data inputs for forecasting COVID-19 hospital admissions. All models used past hospitalization admissions data as an input to forecasting, and the others additionally incorporated data on COVID-19 cases.

The model variations we considered are:

1. **HospOnly**: A reference model that forecasts hospitalizations using only the past reported values of hospitalizations as an input.
2. **ReportCase-CSSE**: In addition to past hospitalizations, this model uses report-date cases as reported by JHU CSSE as an input to forecasting hospitalizations.
3. **ReportCase-DPH**: In addition to past hospitalizations, this model uses report-date cases as reported by the state Department of Public Health as an input to forecasting hospitalizations. This model was only used in California, as the daily DPH and CSSE numbers did not always agree.
4. **TestCase**: In addition to past hospitalizations, this model uses test-date cases as an input to forecasting hospitalizations.

We formulated our research questions about the utility of COVID-19 cases as an input to forecasting hospitalizations in terms of comparisons of the forecast skill of these models:

1. Are cases helpful for forecasting hospitalizations? This question is addressed with a comparison of the **HospOnly** and **ReportCase** models, and a comparison of the **HospOnly** and **TestCase** models.
2. In an idealized setting without reporting delays, is the more-epidemiologically-relevant signal of test date cases more helpful for forecasting hospitalizations than the signal of cases by report date? This question relates to a comparison of the **ReportCase** and **TestCase** models.

#### Data transformation

We use the notation *c*_*t*_ and *h*_*t*_ to refer to the observed counts of daily cases and hospitalizations at times *t* = 1, …, *T* respectively, after initial data preprocessing steps that were performed in the following order:

1. In instances where there was a run of one or more consecutive reported zeros, we replaced the zeros and the value on the following day by the mean of the replaced observations. This occurred only for report-date cases, and addressed situations where no values were reported on some dates, followed by catch-up reporting.
2. In some model variations, we smoothed the case data using a 7-day trailing average.
3. We used a fourth-root transformation of both the case data and the hospitalization data to stabilize the variability of the signal around the local trend.
4. In some model variations, we used initial non-seasonal or seasonal (with a one-week “seasonal period”) differencing to obtain a stationary series.

We considered four variations for differencing, where the order of non-seasonal and seasonal differencing (denoted by *d* and *D* respectively) could each take values of 0 and 1.

In model variations where differencing was applied, we used recursive differencing[25]. Here we let *y*_*t*_ refer to the time series to be differenced and 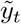 be the resulting time series:

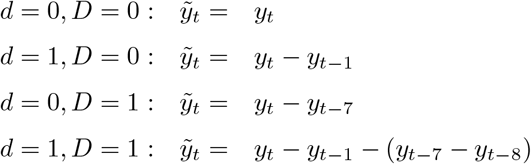

#### Model Specifications

We used seasonal vector autoregressive models to model the case and hospitalizations data jointly, with weekly (7-day) seasonality to capture day-of-week reporting effects.

Our model specifies that these transformed and possibly differenced counts at time *t* follow a normal distribution with a mean that is a linear function of past values of transformed hospitalizations (*h*_*t*_) and/or transformed cases (*c*_*t*_). To encode the epidemiological understanding that an individual will generally be infected before they are hospitalized (other than hospital-acquired infections), we allow hospitalizations to depend on past observed hospitalizations and cases, but cases only depend on past cases. As with standard autoregressive model notation, the *p* and *P* parameters determine the lags included in the model (with *P* determining the ‘seasonal’ lag), while the *d* and *D* parameters determine how the data were differenced. The parameters *p, d, P* and *D* were fixed to be the same values for both cases and hospitalizations. All combinations of (*p, d, P, D*) where *p* ∈ {0, 1, 2, 3, 4}, *d* ∈ {0, 1}, *P* ∈ {0, 1, 2}, *D* ∈ {0, 1} were considered, except for models where both *p* and *P* were zero, leaving 56 valid model specifications per model input specification. The models can be expressed as follows:

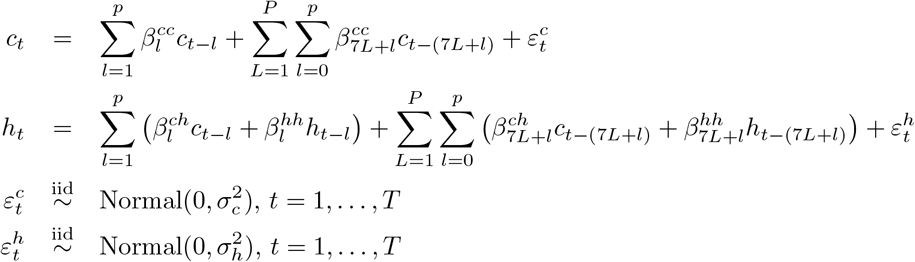

Priors for the model parameters were specified as

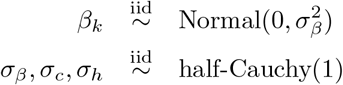

where *β*_*k*_ represents any of the *β* coefficients in the model definition above. This model specification assumes that on average, after transformation, new values of cases are linear combinations of prior values of cases and new values of hospitalizations are linear combinations of a specific set of prior values of cases and hospitalizations.

We fit the models in a Bayesian framework and obtained forecasts from the posterior predictive distribution by iteratively simulating one-step-ahead forecasts of both cases and hospitalizations until 28 days had been forecasted.

### Forecast structure and evaluation

Following the forecast format used by several COVID-19 modeling hubs, we generated probabilistic forecasts in a quantile-based format[24]. A prediction target was defined as a combination of location *l*, week in which the forecast was made *w*, and forecast horizon in days *d* which indicates how many days into the future the forecast was made for, relative to the starting point of the forecast. In this structure, a specific prediction is represented as a set of *K* = 23 symmetric quantiles (including the median) of a predictive distribution for a particular target. We denote the observed data as *y*_*l,w,d*_ and the quantiles as *q*_*l,w,d,k*_, for *k* = 1, …, *K*. Below, we use *k*^*med*^ to denote the index of the quantiles which corresponds to the median.

The forecast evaluations presented below primarily focus on three metrics that together assess the aggregated bias, sharpness and calibration of the predictive distributions generated by the models. Let *N* be the total number of unique combinations of *l, w*, and *d* to be evaluated. The mean absolute error (MAE) is a measure of accuracy of the point estimate (median, or *q*_*l,w,d,k*_*med*) of the forecast and is computed as:

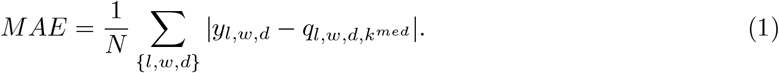

The weighted interval score (WIS)[26] is a proper scoring rule[27] that measures the sharpness and calibration of the entire predictive distribution. For a single observation *y* and a predictive distribution represented by quantiles *q*_1:*K*_, the WIS is computed as

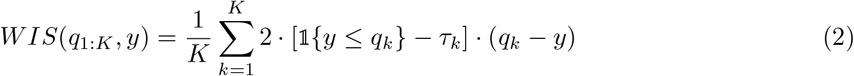

where *τ*_*k*_ is the quantile level (e.g. if *q*_*k*_ is the 95th quantile, then *τ*_*k*_ = 0.95) and 1{*y* ≤ *q*_*k*_} is the indicator function such that it equals 1 when *y* ≤ *q*_*k*_. The mean weighted interval score is defined as

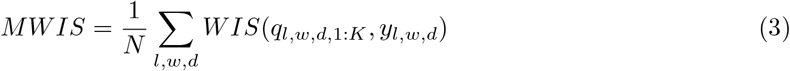

The 95% prediction interval (PI) coverage is the empirical coverage of the nominal 95% prediction intervals as given by the set of quantiles. Letting 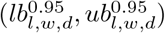 be the quantiles that correspond to the central 95% prediction interval, we compute the 95% PI coverage as:

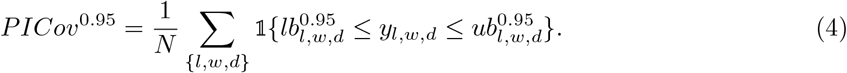

### Forecast validation and testing periods

We used a two-phase validation and test period approach to guard against overfitting in the models and provide an out-of-sample evaluation of the modeling approaches. Prior to fitting models, we established a validation period as running from forecast dates of 2020-12-07 to 2021-06-07 (Figure 1). We performed extensive exploratory analyses on data from the validation period, including examining plots of data, model fitting, model diagnostics and analyses of forecast errors. The test period was defined to include forecasts made on 2021-06-14 through 2022-05-30.

In California, 224 models were evaluated in the validation period, with 56 different autoregressive specifications for each of four different input data types (“HospOnly”, “ReportCase-CSSE”, “ReportCase-DPH”, and “TestCase”). In Massachusetts, 168 models were evaluated in the validation period, with 56 different auto-regressive specifications for each of three different input data types (‘HospOnly”, “ReportCase-CSSE”, and “TestCase”). When case data were used, they were pre-smoothed using a trailing 7-day average before being passed to the model fitting algorithms. Results from models using unsmoothed data are reported in Supplemental File 3. The highest performing models for each of the input data types in the validation period (3 for MA, 4 for CA) were passed to the test period (Table 2)

**Table 2:** Test and validation period accuracy metrics for forecasts of California and Massachusetts hospital admissions. The models shown include the best individual autoregressive models from the training phase that used test-date data (TestCase), report-date data (ReportCase) and no case data (HospOnly) as inputs. The mean weighted interval score (MWIS), mean absolute error (MAE) and 95% prediction interval coverage (PIcov^0.95^) scores are shown for each model with the best scores in the test and validation periods highlighted. Within each state, the models are sorted by highest accuracy (lowest MWIS) scores at the top. The model parameters for the auto-regressive model are also provided in the (p,d,P,D) column. The TestCase model was the most accurate (lowest MWIS) in the validation period for both states, but also was the least accurate in the test period.

| case type | (p,d,P,D) | test period |  |  | validation period |  |  |  |
| --- | --- | --- | --- | --- | --- | --- | --- | --- |
|  |  | MWIS | MAE | PIcov<sup>0.95</sup> | MWIS | MAE | PIcov<sup>0.95</sup> | rank |
| **California** |  |  |  |  |  |  |  |  |
| HospOnly | (1,1,1,0) | **130.0** | 196.1 | 0.94 | 124.8 | 203.7 | 0.99 | 18/224 |
| ReportCase-CSSE | (4,1,0,0) | 130.2 | **192.7** | 0.92 | 114.1 | 172.8 | 1.00 | 6/224 |
| ReportCase-DPH | (4,1,0,0) | 139.3 | 204.9 | 0.92 | 115.4 | 171.9 | 0.99 | 8/224 |
| TestCase | (2,0,0,1) | 141.3 | 206.2 | 0.89 | **104.8** | **159.4** | 0.98 | 1/224 |
| **Massachusetts** |  |  |  |  |  |  |  |  |
| ReportCase-CSSE | (4,1,0,0) | **24.1** | **37.6** | 0.94 | 18.6 | 27.4 | 0.98 | 8/168 |
| HospOnly | (1,0,1,1) | 26.4 | 40.7 | 0.91 | 19.6 | 28.7 | 0.97 | 16/168 |
| TestCase | (1,0,1,1) | 29.2 | 41.8 | 0.93 | **17.2** | **26.5** | 0.99 | 1/168 |

We note that one post hoc adjustment was made to the modeling procedure after an initial view of test period forecasts. Because we had been careful not to look in detail at the data in the test period while fitting models to data from the validation period, we did not initially realize that systematic patterns of reporting of zero cases changed meaningfully between the validation and test periods. Starting in July 2021 for Massachusetts and April 2022 for California, report-date case counts for weekends and holidays were consistently reported to be zero due to changes in data publication schedules (Figure 1). In the entire validation period, zero counts happened very rarely. This caused models that had performed well in the validation period to perform substantially worse in the test period, as zero counts can cause problems in certain model specifications without additional intervention on the raw data to smooth out these systematic changes in reporting. Therefore, after an initial run of models in both the validation and test periods, we went back and updated the methods to smooth input data and re-ran models for the validation period and re-selected models to pass to the test period. While this procedure goes against standard principles of re-fitting models in the validation phase after seeing test phase results, we felt that the resulting modifications were (a) simple and (b) something like what might have been implemented by a modeler in real-time who noticed the systematic changes in reporting of case data. Nonetheless, this revisting of the underlying data processing pipeline after “breaking of the seal” on the test phase data introduces the possibility of additional over-fitting to the data and is a limitation of the current work.

### Data and code availability

All data and code for this project are available publicly at https://github.com/reichlab/covid-hosp-forecasts-with-cases. Models were fit and forecasts were generated using Python version 3.8.2 and numpyro version 0.7.2; the model is implemented in the sarix package, available at https://github.com/elray1/sarix. Post-processing and analyses of forecasts were performed in R version 4.2.0[28].

We followed the EPIFORGE 2020 reporting guidelines for studies with epidemiological forecasting (Supplemental File 3)[29].

## Results

The Results are organized into four discrete sections. First, we discuss results about the degree to which case counts showed empirical correlations as a leading and lagging indicator of hospitalizations in California and Massachusetts. Then we present two moment-in-time case-studies of forecasts generated for case counts and hospital admissions at key moments during the COVID-19 pandemic. Finally, we present a retrospective comparison of forecast accuracy for predictions of hospital admissions from models that both do and do not use case data as inputs.

### Cases and hospitalizations are correlated, with inconsistent leading patterns

Over the course of the COVID-19 pandemic from October 2020 through July 2022 in both Massachusetts and California, finalized case data aggregated by test date or report date did not show strong evidence of being a leading or lagging indicator for hospitalizations. The raw daily case counts for all data streams showed substantial variability (Figure 1A and C). Trailing 7-day averages of final data showed that trends in cases aggregated by test-date led trends of cases aggregated by report-date, as would be expected due to the lag of typically at least one day between a test being performed and being reported by a surveillance system.

However, the degree to which either of these signals led hospitalizations appeared to vary across the pandemic. For example, using a smoothed measure of the relative one week change to indicate whether a signal was increasing or decreasing, hospitalizations in Massachusetts appeared to increase after both case signals in the Alpha wave in spring 2021. Nonetheless, in summer 2022, the hospitalizations started to increase before cases (Figure 1B). In California, hospitalizations appeared to increase before cases in October 2021, but only increased several weeks after rises in cases were observed in spring 2022 (Figure 1D).

Aggregated measures of correlation across the entire time period included in our analysis (October 2020 through July 2022) support the conclusion that there is not strong evidence of any case data stream consistently leading or lagging hospitalizations over the course of the first two and a half years of the pandemic. Cross-correlations were computed for each daily lag from −25 to +25 days between hospitalizations and report-date and test-date case signals. We observed high correlations (greater than approximately 0.75) between hospitalizations and any case signal at lags of +/- 7 days (Figure 2).

**Figure 2:**
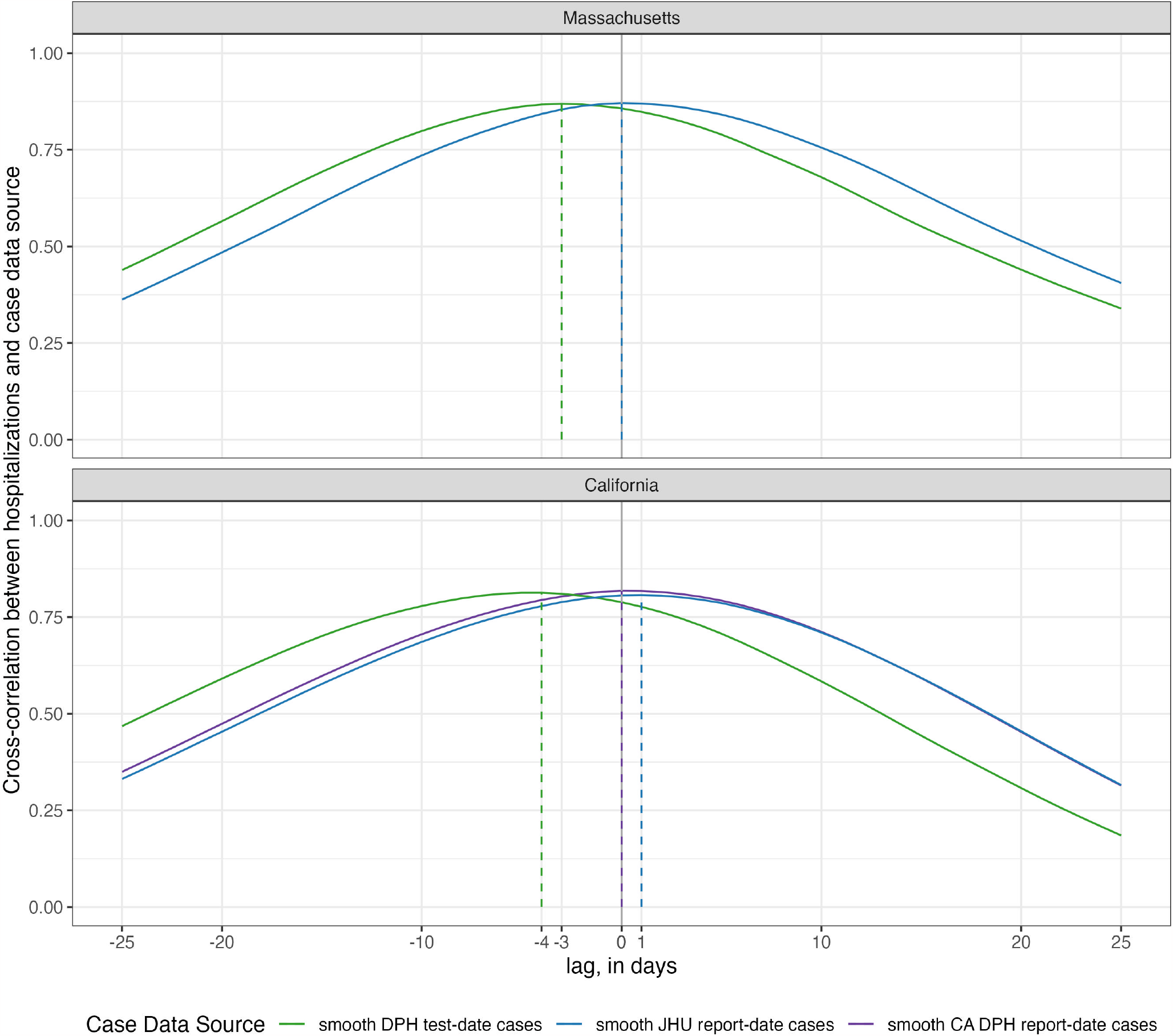
Cross-correlations between hospitalization and case data, by case data source and state. The height of the lines represents the observed of correlation between the time-series of two data sources that are offset by the value on the x-axis. For example, when the lag is zero days (x=0) the cross-correlation value is the correlation between the two time-series. When the lag is −3 days (x=-3), the value of the line reflects the correlation of the hospitalization counts at time *t* and the case counts at time *t* − 3. Test-date cases were observed to have the highest correlation with hospitalizations at lags of −3 days in Massachusetts and −4 days in California. Report-date case counts from JHU CSSE were observed to have the highest correlation with hospitalizations at lags of 0 days in Massachusetts and 1 day in California. Report-date case counts from DPH were observed to have the highest correlation with hospitalizations at a lag of 0 days in California (DPH and JHU CSSE data were identical in Massachusetts). Correlations were above 0.75 for both states and nearly all sources for +/- 7 days of lag.

Test-date cases in both states achieved a maximum correlation with hospitalizations by being aligned as a leading indicator of hospitalizations, although correlations with some positive lag values were also high. New hospitalizations at the current day showed the highest correlation with test-date cases from three days earlier in Massachusetts and four days earlier in California. These correlations reach a level of 0.87 and 0.81 for Massachusetts and California, respectively. With report-date cases, the maximum correlation is achieved at 0 days in both Massachusetts (r=0.87) and 0 or a 1 day lag in California (for DPH or JHU report-date cases, respectively, r=0.82 and 0.81).

### Case Study 1: California, July 2021 – the start of the Delta wave

As one case-study, we examine in detail the data and forecasts from California on Monday, July 12, 2021 (Figure 3). At that time, hospitalization admissions and case counts had shown a steady decline since early January 2021. However, as of early July 2021, both of these signals show clear upward trends in the finalized case counts. We note that real-time data also showed upward trends, although the test-date case count signal would eventually be revised to show a more pronounced increasing trend (Supplemental File 1).

**Figure 3:**
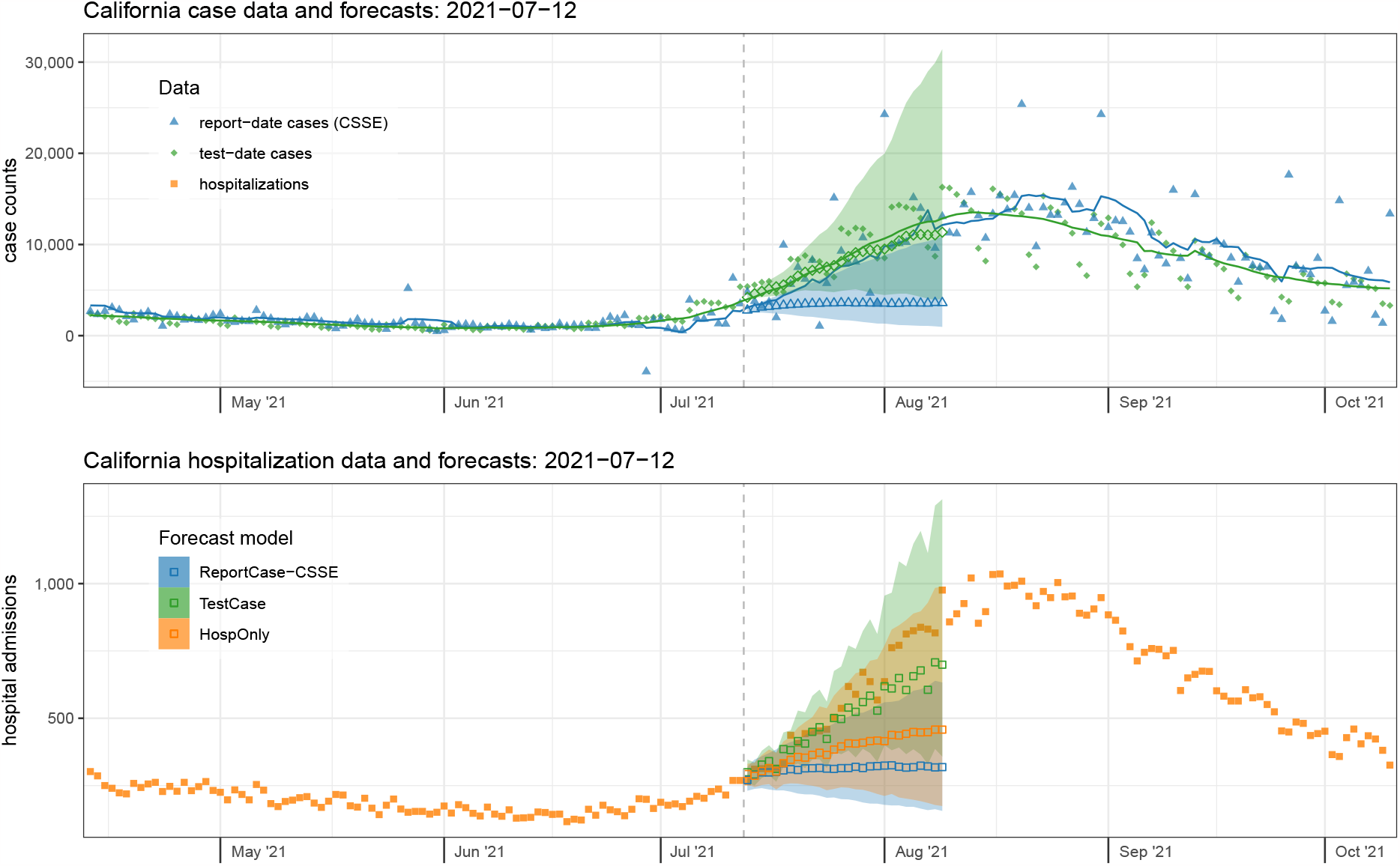
Case data and forecasts (top panel) and hospitalization data and forecasts (bottom panel) from California on July 12, 2021. Finalized case data aggregated by test-date (top panel, solid green diamonds) were slightly higher at this time than the report-date cases (top panel, solid blue triangles). Forecasts of test-date cases (open green diamonds) better reflected the rise in trends over the subsequent four weeks than forecasts of report-date cases (open blue triangles). Similarly, the accompanying forecasts of hospitalizations from the TestCase model that used test-date cases (open green squares) were higher and more accurate than those using cases aggregated by report-date from the ReportCase-CSSE model (open blue squares). The forecasts of hospitalizations (bottom panel) were more accurate on this date for the TestCase model than from any of the other models. Observed hospitalization admission counts (bottom panel) are shown in solid orange squares, and forecasts from the model that only used these data as inputs are shown as open orange squares. Forecasts were similar for the ReportCase-CSSE model (open blue triangles and squares) and ReportCase-DPH model (not shown). Shaded regions around forecasts indicate 80% prediction intervals.

Test-date case forecasts more clearly detected the increasing trend, with a median prediction of over 11,343 test-date cases per day for Monday, August 9th, 2021 (80% PI: 4,029 - 31,412). On this day, the 7-day trailing average case count was 12,840. However, report-date case forecasts did not foresee this steep increase as clearly, predicting 3,602 report-date cases per day for August 9th (80% PI: 966 - 10,853), with the eventual observation being 12,137, landing between the 90th and 95th quantile of the predictive distribution of cases.

The more accurate test-date case forecasts subsequently translated into more accurate forecasts of hospitalizations for this time-period as well, although at horizons beyond two weeks all models underpredicted the eventual observations. The TestCase hospitalization forecast model predicted 699 new admissions on Monday August 9th (80% PI: 358 - 1313) and the ReportCase-CSSE model predicted 319 (80% PI: 156 - 633). The HospOnly forecasts were slightly higher than those from ReportCase-CSSE, with 458 predicted hospitalizations on August 9th (80% PI: 174 - 989). The eventual observed hospital admissions for this day was 976.

### Case Study 2: Massachusetts, January 2022 – nearing the peak of the Omicron wave

As a second case-study, we look in detail at the data and forecasts from Massachusetts on Monday, January 3, 2022 (Figure 4). Hospitalization admissions and case counts had been rising steeply since early November 2021, and they would reach their peak values by the end of the second week of January 2022. Forecasts near the peak of COVID-19 waves have been shown to be unreliable[30, 31], and the forecasts here follow that pattern. The real-time data available at this time showed steady increases, with a slight flattening in the recent trailing averages of counts. The report-date cases were fully reported on this date, and the test-date cases from MA DPH were slightly lagged in reporting (Supplemental File 1).

**Figure 4:**
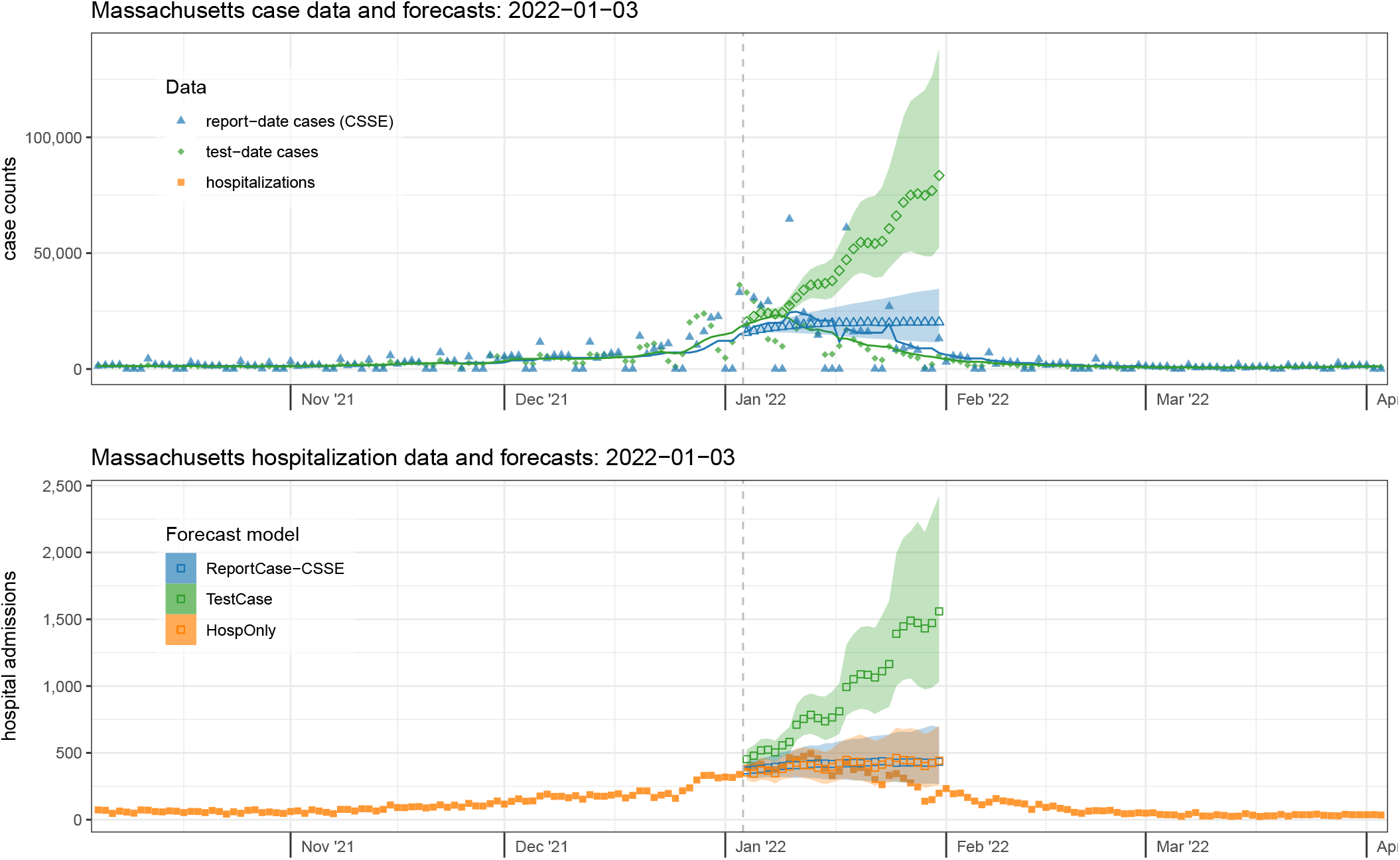
Case data and forecasts (top panel) and hospitalization data and forecasts (bottom panel) from Massachusetts on January 3, 2022. Finalized case data aggregated by test-date (top panel, solid green diamonds) were slightly higher at this time than report-date case counts (solid blue triangles). However, the Omicron wave was near the peak. The forecasts of test-date cases (open green diamonds), which predicted a continued rise, were less accurate than those for cases aggregated by report-date (open blue triangles). The accompanying forecasts of hospitalization admission data (solid orange squares, bottom panel) were less accurate on this date when coming from models that used test-date cases (TestCase model, open green squares) than from models that used only hospitalization data (HospOnly, open orange squares) or report-date case data (ReportCase-CSSE model, open blue squares), as they predicted a continued increase and did not anticipate the peak of the Omicron wave that occurred in mid-January. Forecasts were similar for the ReportCase-CSSE model (open blue triangles and squares) and ReportCase-DPH model (not shown). Shaded regions around forecasts indicate 80% prediction intervals.

The forecasts made on January 3rd, 2022 extended 28 days until January 31st. The number of test-date cases in Massachusetts on January 3rd (based on a 7-day trailing average) was 18,031, up from 4,461 just three weeks earlier. This count peaked on January 8th at 23,204 and on January 31st had dropped to 4,726. Test-date case forecasts made using the data through January 3rd were more aggressively pessimistic, with a median prediction of over 83,520 test-date cases per day for Monday, January 31st, 2022 (80% PI: 52,472 - 138,315). However, report-date case forecasts essentially forecasted a flat line (with a very slight increase) from the most recent observation, predicting 20,243 report-date cases per day for January 31st (80% PI: 11,381 - 34,626).

The number of new hospital admissions in Massachusetts on January 3rd was 339. This count peaked on January 13th at 495 and on January 31st had dropped to 198. The patterns observed in the case forecasts were passed through to the hospitalization forecasts, with all forecasts over-predicting at a 28-day horizon and only the TestCase model overpredicting at all horizons. The TestCase hospitalization forecast model predicted 1,559 new admissions for Monday, January 31st (80% PI: 1,032 - 2,423) and the ReportCase-CSSE model predicted 433 (80% PI: 268 - 691). The HospOnly predictions were similar to ReportCase-CSSE with 441 predicted hospitalizations on January 31st (80% PI: 258 - 703).

### Comparative forecast model results using different case data

When aggregated across all horizons and dates in the test period, forecasts from the HospOnly and ReportCase-CSSE models were similarly accurate according to weighted interval score (WIS), and TestCase-DPH models were the least accurate (Table 2). The chosen specifications for the ReportCase models in both California and Massachusetts was (*p*=4,*d*=1,*P* =0,*D*=0), suggesting that trends in only the last 5 days of report-case data were needed in this modeling setup to generate optimal predictions. The most accurate HospOnly model in the validation period for both states included a *P* = 1 parameter, indicating that observations from prior weeks were also used. TestCase-DPH models showed the highest accuracy (lowest WIS score) of the selected models in the validation period but then had slightly lower accuracy in the test period than the other selected models.

However, the selected TestCase-DPH models consistently performed similarly or better than HospOnly and ReportCase models in specific settings, with dramatically worse performance around the peak hospitalizations in January 2022 that dragged down the overall performance metrics. In Massachusetts for example, forecasts from the selected TestCase-DPH model were more accurate than forecasts from the other models in all but two weeks during a 15 week span from November 2021 through February 2022 (Figure 5). But in the two weeks where the TestCase-DPH forecasts had larger error, the errors were dramatically higher, at a critically important turning point of the epidemic. These large errors were due to TestCase-DPH forecasts predicting a continued increase into early February 2022 when in fact the case counts had just peaked (Figure 4, Supplemental File 2). Looking at aggregate errors by forecast horizon in both states, the errors for the TestCase-DPH forecasts are similar at horizons less than 14 days, and notably larger at horizons beyond 21 days (Figure 6).

**Figure 5:**
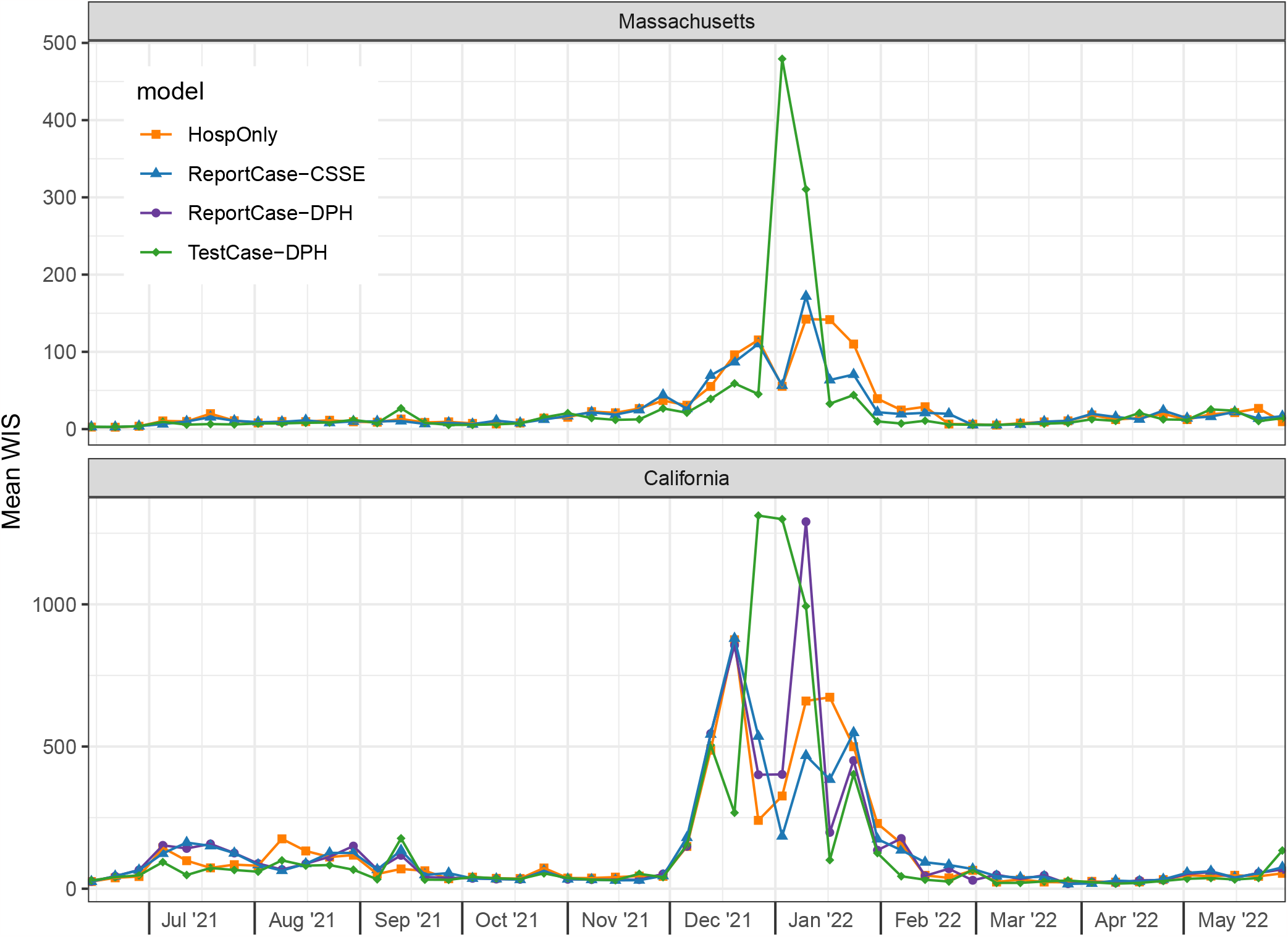
Mean WIS (*MWIS*) by forecast date in test period. Each point represents, for a given model and forecast date, the mean WIS of all the forecasts made on the given forecast date, averaging across all of the 1 through 28 day-ahead forecast horizons. Lower WIS scores indicate higher probabilistic accuracy. In Massachusetts (top panel), forecasts from the TestCase-DPH model (green diamonds) tended to have lower error during the Omicron wave except for two weeks where forecasts from this model had much higher error than other models.In California (bottom panel), WIS scores for all models were similar, although during the Omicron wave of December 2021 through February 2022, the scores were higher and showed greater variability.

**Figure 6:**
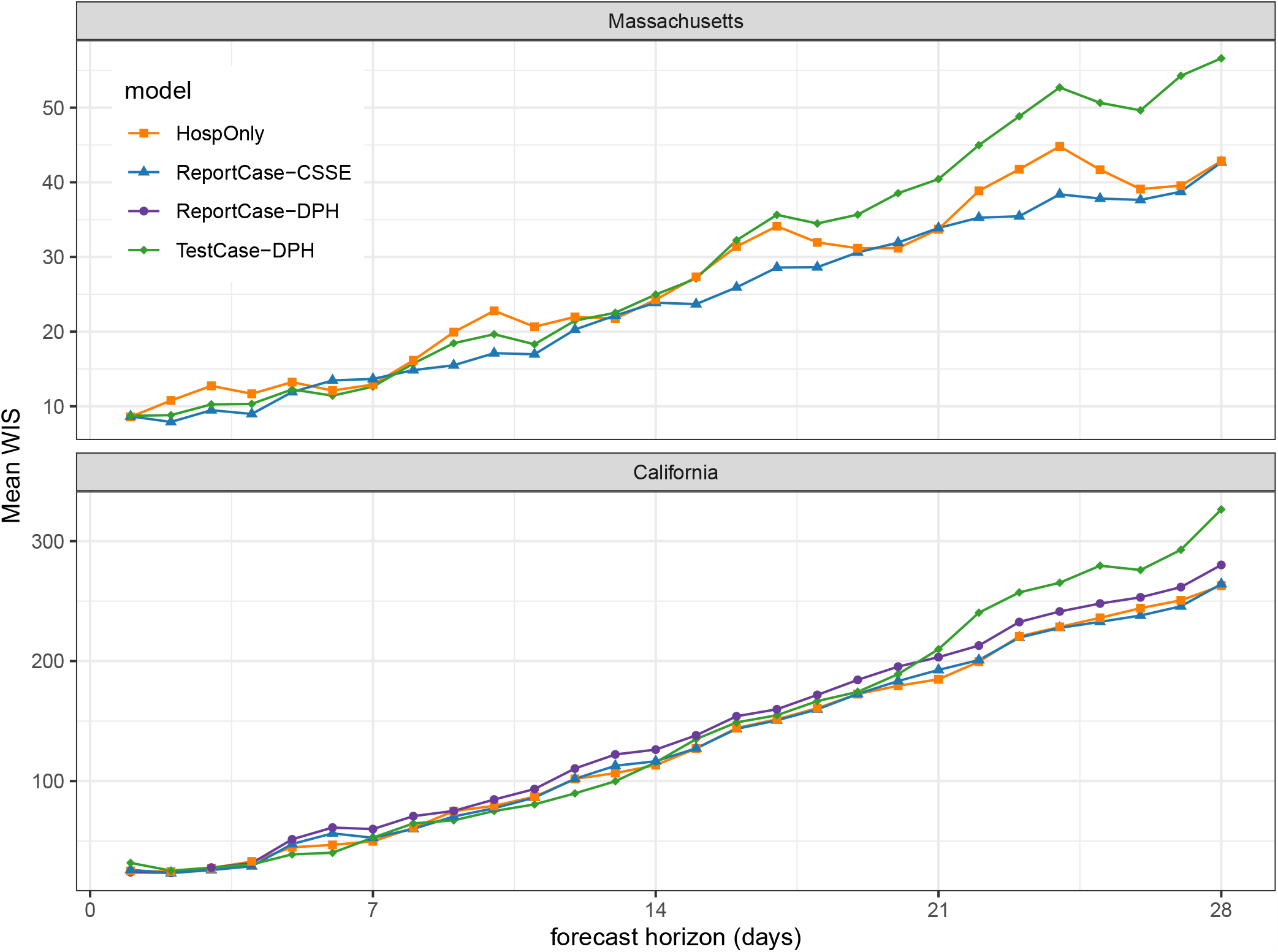
Mean WIS (*MWIS*) by forecast horizon in test period. Each point represents, for a given model and forecast horizon, the mean WIS of all the forecasts made during the test period for the given horizon, averaging across all forecast dates. Lower WIS scores indicate better probabilistic accuracy. In both California (bottom panel) and Massachusetts (top panel), the mean WIS for all models was similar up until between 14 and 21 days. For horizons longer than 21 days, forecasts from the TestCase-DPH model (solid green diamonds) showed higher error than the other models.

Prediction intervals were conservatively wide during the validation period and showed a moderate lack of calibration in the test period (Table 2). The 95% prediction intervals for each of the selected models in the validation phase had empirical coverage ranging from 97% to just under 100%, averaged across all horizons and all dates. In the test period, all empirical coverage rates were under 95%, but never lower than 89%, indicating a fairly good calibration. There were not clear patterns across both locations, with one model type consistently showing better or worse calibration than others. For example, the TestCase models covered the truth 89% of the time in California (the lowest of any model in that state) but showed 93% calibration overall in Massachusetts.

In California, where we compared models with two different report-case data inputs, little difference was observed in the validation period between the two models and a modest difference was observed in the test period (Table 2). In the validation period, the ReportCase-CSSE and ReportCase-DPH models showed virtually identical metrics, with a less than 1% difference in WIS, MAE and PI coverage. In the test period, the ReportCase-CSSE forecasts showed slightly higher error, due largely to one week in early January 2022 with a forecast that did not anticipate the rapid decline (Figure 5).

## Discussion

In this work we investigate whether COVID-19 case counts, as reported by state departments of public health, are useful as indicators and predictors of future trends in hospitalizations. Overall, trends in case counts (whether aggregated by date of report or date of test) were only marginally “ahead of” trends in hospitalizations, and we were unable to find a model that was able to show consistent improvement in forecast accuracy by including recent case data. These results were consistent across two states (California and Massachusetts) with different public health surveillance systems and reporting processes.

Earlier in the paper, we posed two questions, and we summarize our answers below.

1. **Are cases helpful for forecasting hospitalizations?** The answer is mixed. In general, forecasts that only used hospitalization data (the HospOnly model) and forecasts that used report-date cases (the ReportCase model) showed little difference. This suggests that using report-date cases does not significantly impact forecasts of hospitalizations one way or another. However, using test-date cases does impact forecasts of hospitalizations, but in ways that are sometimes helpful and sometimes not. Forecasts that used test-date cases (from TestCase model) showed lower accuracy overall than forecasts that used only hospitalization data (the HospOnly model). Despite having consistently higher accuracy in many weeks, the forecasts that used test-date cases had several weeks with substantially lower accuracy and this drove the overall accuracy down.
2. **Are test-date cases more helpful for forecasting hospitalizations than report-date cases?** Again the answer is mixed. In general, the differences between forecasts that used test-date cases (from TestCase models) and those that used report-date cases (from ReportCase models) followed similar patterns to those discussed above. Forecasts using test-date cases showed higher overall error in the test period, mostly reflecting substantially worse performance in several weeks where the forecasts were too pessimistic at the peak of the Omicron wave, despite showing higher accuracy for many other weeks.

Finding leading indicators that can improve predictive outbreak modeling is a challenging endeavor. Indicators that can be used to improve predictive models must be both strongly correlated with the main signal of interest and have similar dynamics that occur reliably earlier than the main signal. Any temporal relationship between the main signal and a potential indicator must remain consistent over time so that a model has sufficient data to first see and fit the relationship and then have that relationship persist into the future.

In the specific setting of using reported COVID-19 case counts to predict hospitalizations, there are several obstacles that may make the relationship between these two signals difficult to model. First, case reports occur at various times during an individual’s course of infection. Some cases may be diagnosed near to onset of symptoms and days before admission to a hospital because of severe COVID-19 symptoms. But other cases may be diagnosed in asymptomatic individuals who are hospitalized for other reasons and happen to also be infected with COVID-19. Additionally, case diagnoses may take several days to be reported into the public health surveillance system, with lags that may vary depending on how burdened the healthcare system is at a given time. These different ways of having reportable cases arise lead to variation in the difference between the time of case diagnosis and the time of hospitalization, which thus leads to variation in how reliably the case signal precedes hospitalizations. Second, the ways in which cases have been diagnosed has varied over the course of the pandemic and the period studied. For example, with the advent of widespread at-home rapid diagnostic tests for COVID-19 in 2021 and 2022, care-seeking behaviors changed along with the patterns by which cases are reported into the surveillance system.

An additional important factor in considering real-time outbreak forecasting experiments that are conducted in retrospect is the vintage of the data that was used in the experiments. In our experiments, the models were provided with “finalized” data (that is, data as they were reported on July 26, 2022), not with the data as they were available in real-time. This creates an idealized forecasting experiment, as all of the data signals in question (hospitalizations, along with test-date and report-date case counts) are subject to varying levels of revision after an initial report. The degree and nature of the revisions depend on the location and the signal (see Supplemental File 3). Our results therefore illustrate how models would perform in an idealized setting, where the “finalized” data were available immediately, which is rarely the case with public health data. The fact that models were unable, even with the advantage of seeing the final data, to improve on hospitalization forecasts by using final case data, indicates that trying to use such a signal in real-time would be even more difficult. We also note that this may impact the sources of case data differently, for example test-date cases are more impacted and thus might experience a larger decay in forecast performance if real-time data were used to construct forecasts. Furthermore, reporting systems in different states or countries may show fundamentally different delays, which could lead to differences in performance than what was observed here.

There are several other limitations with the current study. The presented results may be dependent on specific modeling structures or choices. For example, in the large-scale comparisons of forecast accuracy that have been enabled by the COVID-19 Forecast Hub efforts, many models have failed to see rapid changes in trend at the onset and peak of pandemic waves[30, 31]. It is possible that additional model structure could ameliorate the problem of dramatic over-prediction near the peak, and that traditional autoregressive time-series approaches require alterations to be able to make probabilistically accurate forecasts in systems, like outbreaks, that experience exponential growth or decay. We also have made specific assumptions about model structure, such as the choice of autoregressive linear models plus the requirement that the choice of (*p, d, P, D*) parameterization be the same for both models for cases and hospitalizations. That said, we believe that the general conclusions from this work, namely that using COVID-19 case counts as a predictor of hospitalizations is not an “easy win” to improve forecast accuracy, would hold up over a wide range of different modeling choices. The generalizability of these results to other pathogens and outbreak settings would depend on the time-scales of the generation time of the pathogen in question, as well as time-to-onset of severe disease.

Finally, we note that “test-date” cases were defined slightly differently in Massachusetts and California. The main difference is that in California, some “episode dates” would have preceded the “test date” as defined in the Massachusetts data, making it closer to the date of symptom onset and perhaps more epidemiologically relevant. We think, however, that this would have only made the California test-date data stream more of a leading indicator, and would have biased the results in favor of seeing improvement in forecast accuracy. Since that is not the case, we think this distinction has limited impact on the results of the paper as stated.

Increasingly, models are used as one input to public health decisions. During the COVID-19 pandemic, forecasts and scenario projections were used to assist with decision-making at local, state and federal levels[6]. This work highlights how careful consideration of key data signals can provide important real-time insight, both through close monitoring of different representations of a signal (e.g. considering different aggregation approaches for case counts, and looking at both raw and smoothed counts) and through studying outputs from forecast models. In short, additional research is needed to continue to advance our understanding of which data streams can be used by models to improve prediction of public health indicators.

## Data Availability

All data produced are available online at https://github.com/reichlab/covid-hosp-forecasts-with-cases.

## Acknowledgments

We wish to acknowledge the efforts of staff at both California and Massachusetts Departments of Public Health who have worked tirelessly on collecting data and making it available in a timely and convenient fashion. We also acknowledge Tomas Leon and Ryan McCorvie from California Department of Public Health and Catherine Brown, Kevin Cranston and Dylan Tierney from Massachusetts Department of Public Health for providing insights and comments on an early version of the manuscript.

## Supplementary material

**Supplementary File 1**: A booklet of figures showing report-date case signals from JHU CSSE (blue triangles) and test-date case signals from CA and MA DPH (green diamonds). Each page of the booklet shows data for one state and one date. The plots show open shapes for the “finalized” observations as of July 26, 2022 and solid shapes as vintages of data specific to every Monday date from January 4, 2021 through July 26, 2022. The solid and dashed lines shows a trailing 7-day average for the real-time vintages of data and the finalized data, respectively. The shaded region highlights how the last 3 (for MA) or 7 (for CA) days of data, especially for the test-date cases, tend to be under-reported.

**Supplementary File 2**: A booklet of figures showing data and forecasts for both cases and hospitalizations in Massachusetts and California. Each page shows one week’s data and forecasts for a given state. The top panel on each page shows finalized case data (solid triangles for report-date cases, solid diamonds for test-date cases), a 7-day trailing average of the observations (thin lines), and the forecasts for each data source (open shapes, with 80% prediction interval shown as a shaded region). Bottom panels show hospital admission observations (orange squares) and forecasts from the three selected models (open squares), including an 80% prediction interval.

**Supplemental File 3**: A file containing additional figures and analyses not included in the main manuscript.

