## Supplemental File 1 for "Assessing the utility of COVID-19 case reports as a leading indicator for hospitalization forecasting in the United States"

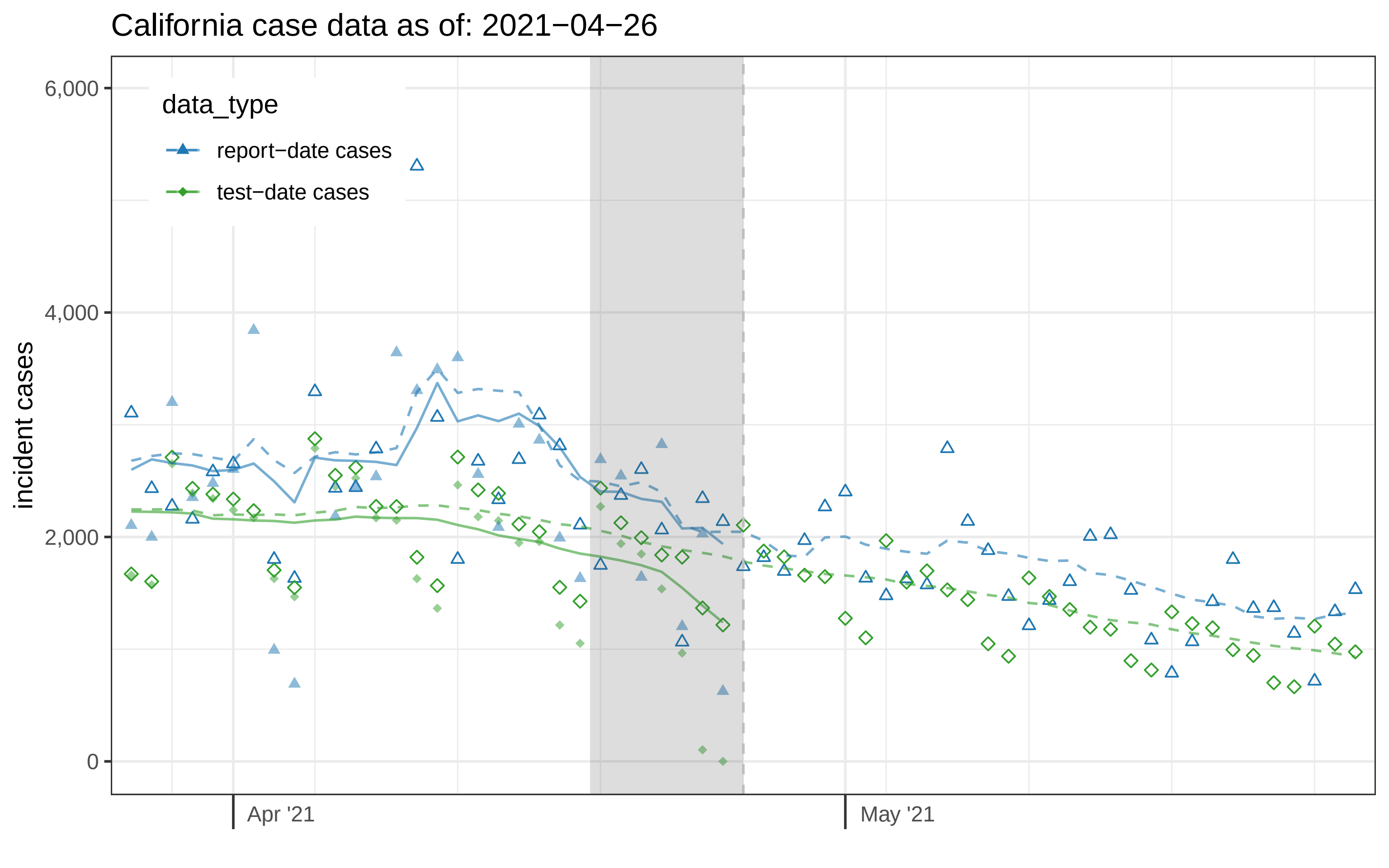
California case data as of: 2021−04−26
data_type
report−date cases
test−date cases
incident cases
6,000
4,000
2,000
0
Apr '21
May '21

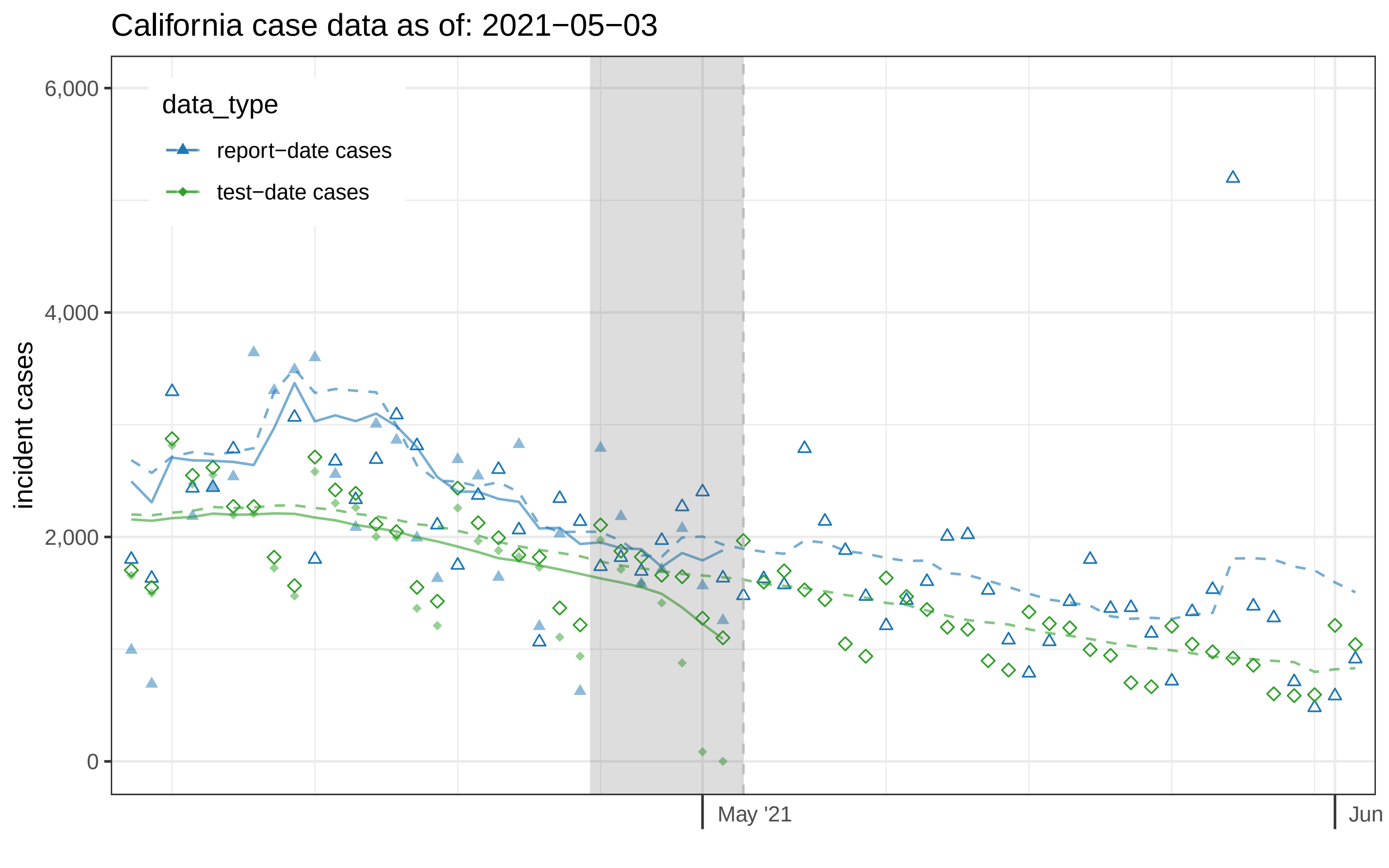

California case data as of: 2021−05−03
data_type
report−date cases
test−date cases
incident cases
6,000
4,000
2,000
0
May '21
Jun

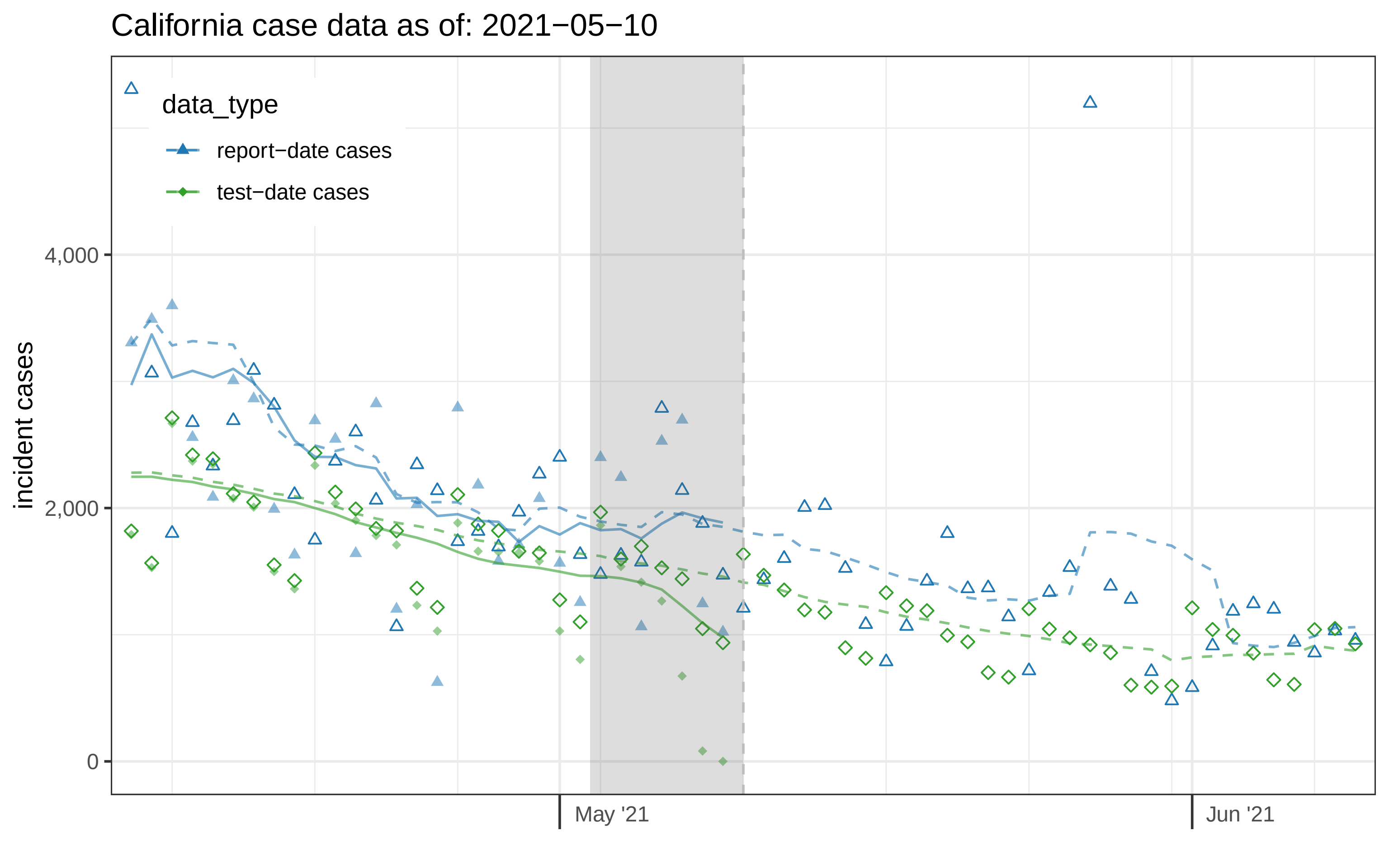
California case data as of: 2021−05−10
data_type
report−date cases
test−date cases
incident cases
4,000
2,000
0
May '21
Jun '21

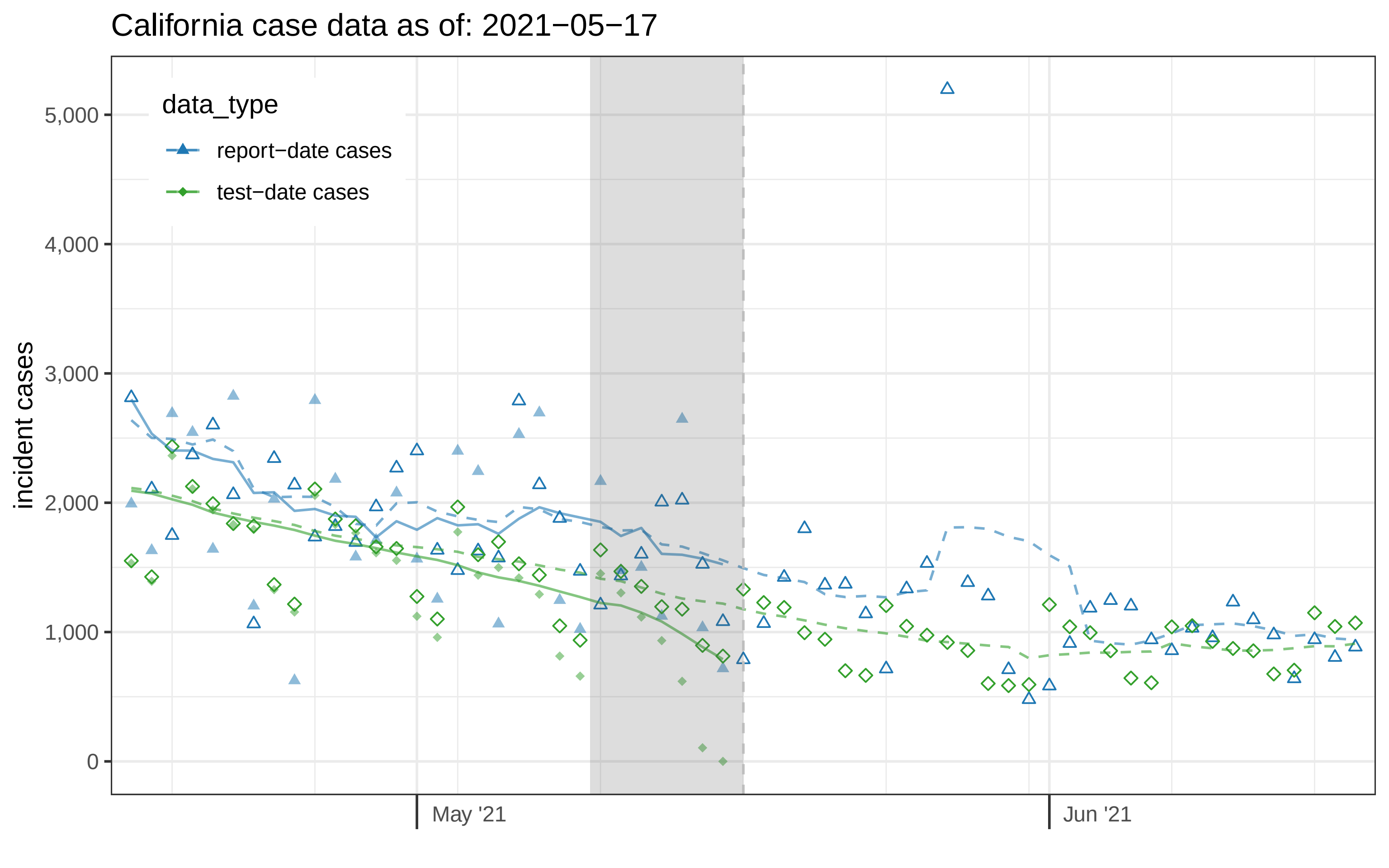

California case data as of: 2021−05−17
data_type
report−date cases
test−date cases
incident cases
5,000
4,000
3,000
2,000
1,000
0
May '21
Jun '21

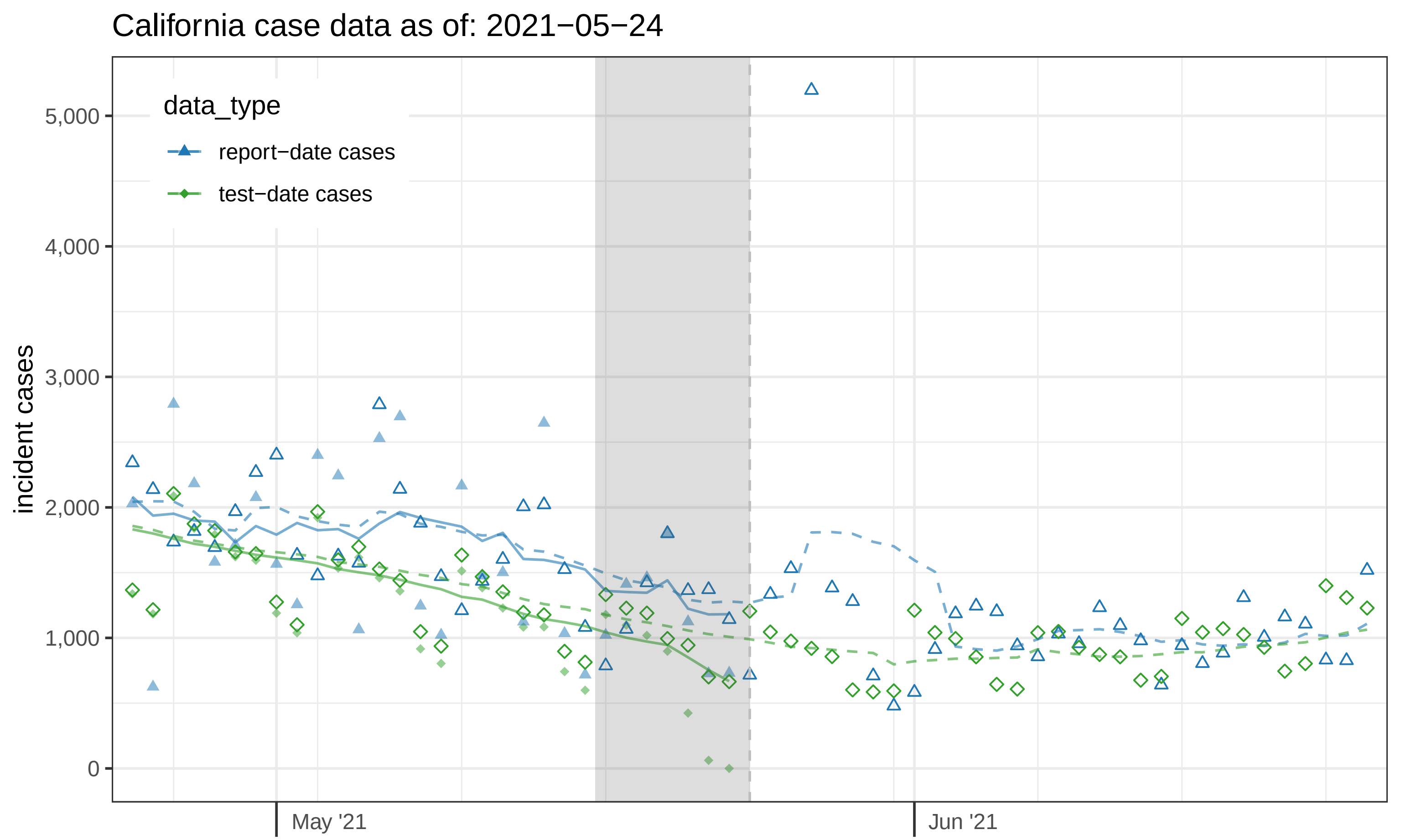

California case data as of: 2021−05−24
data_type
report−date cases
test−date cases
incident cases
5,000
4,000
3,000
2,000
1,000
0
May '21
Jun '21

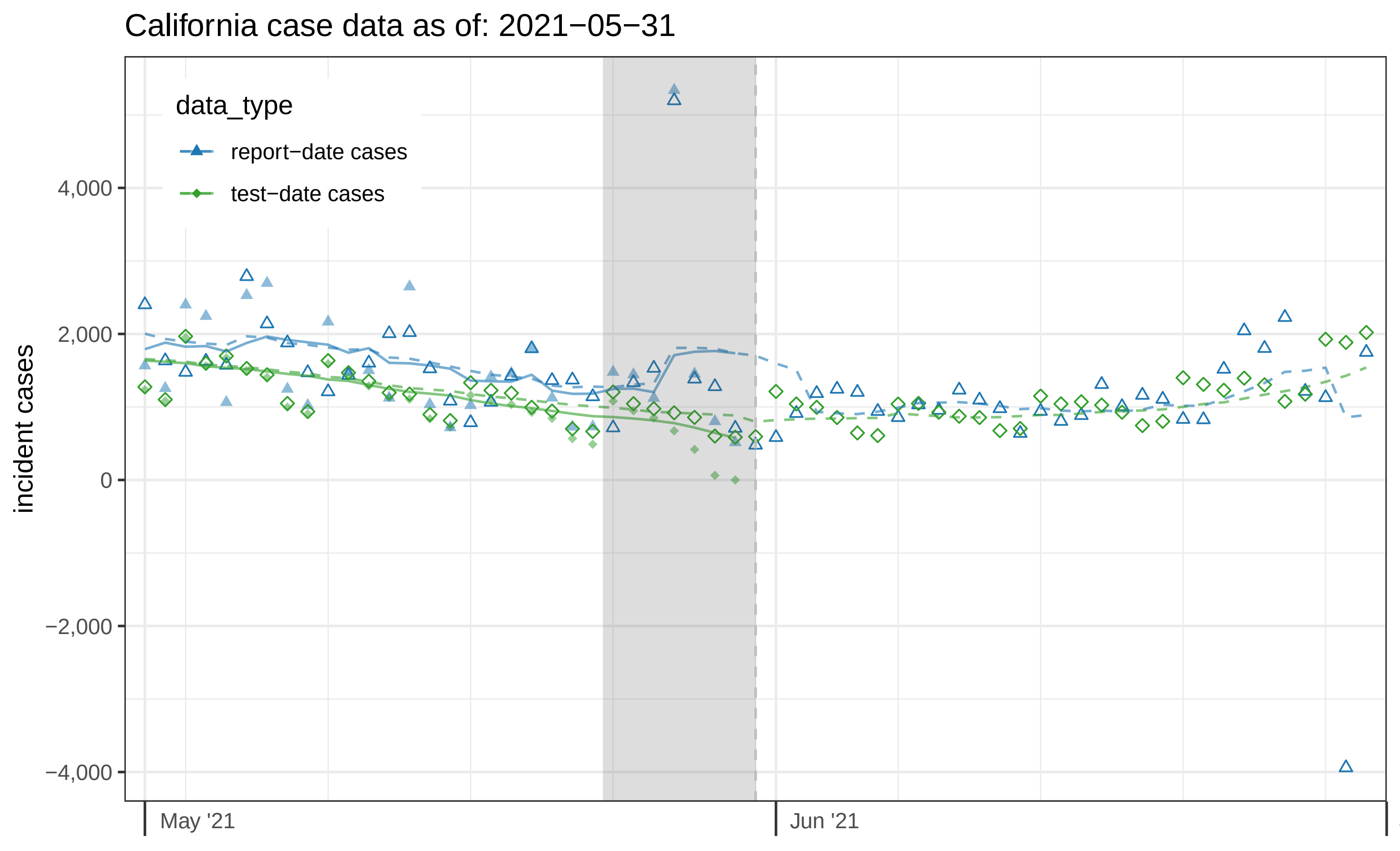
California case data as of: 2021−05−31
data_type
report−date cases
test−date cases
incident cases
4,000
2,000
0
−2,000
−4,000
May '21
Jun '21

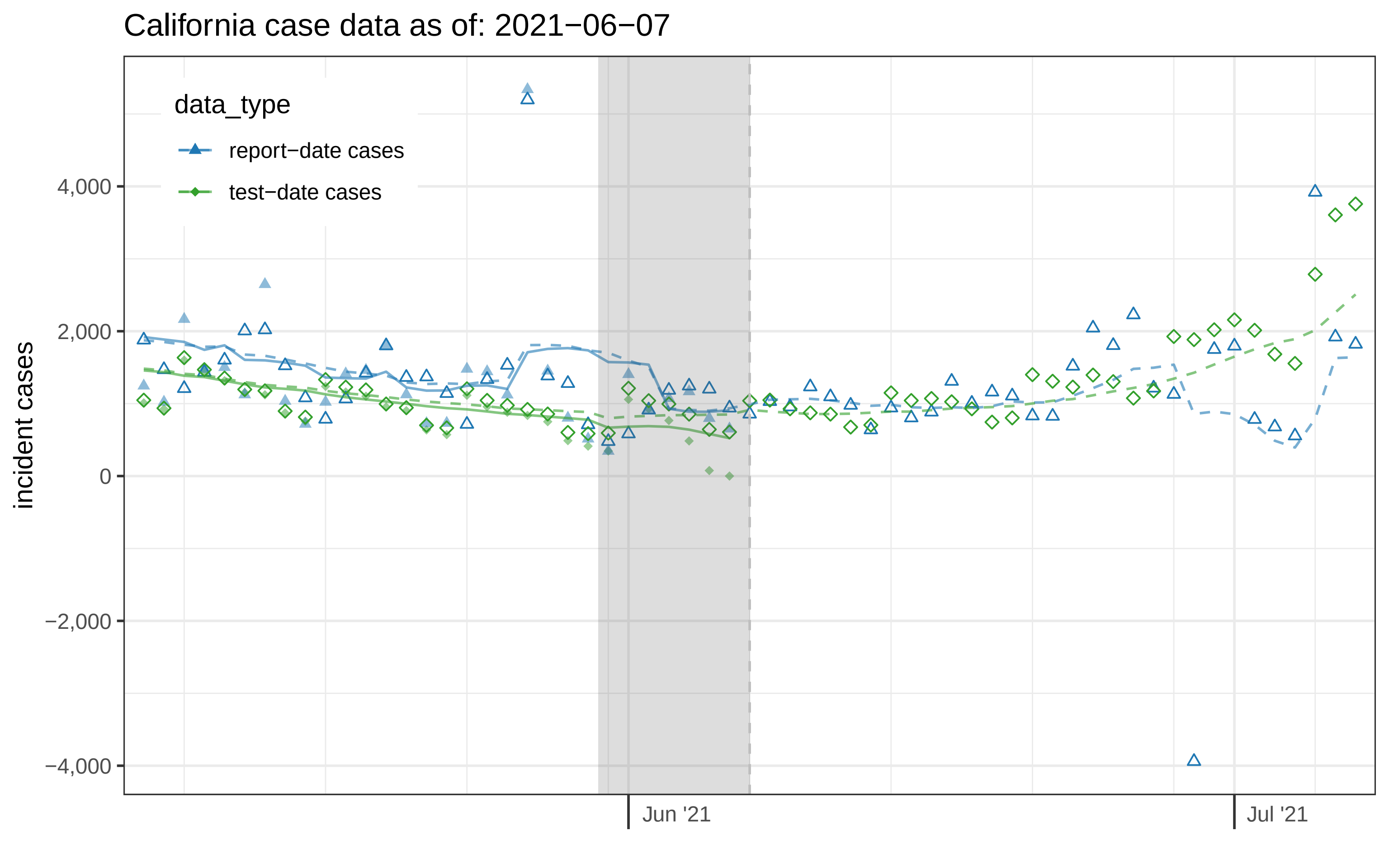
California case data as of: 2021−06−07
data_type
report−date cases
test−date cases
incident cases
4,000
2,000
0
−2,000
−4,000
Jun '21
Jul '21

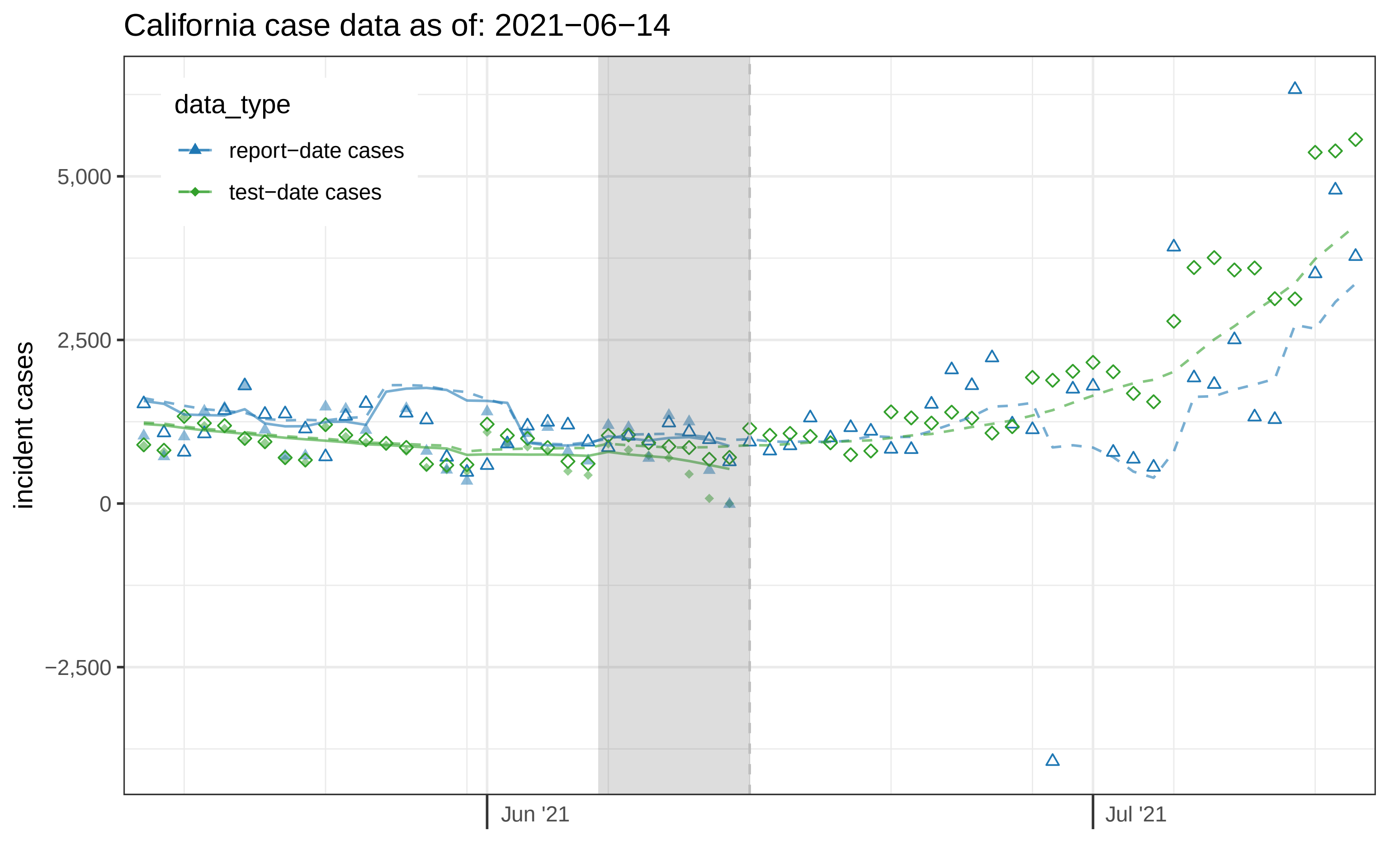

California case data as of: 2021−06−14
data_type
report−date cases
test−date cases
incident cases
5,000
2,500
0
−2,500
Jun '21
Jul '21

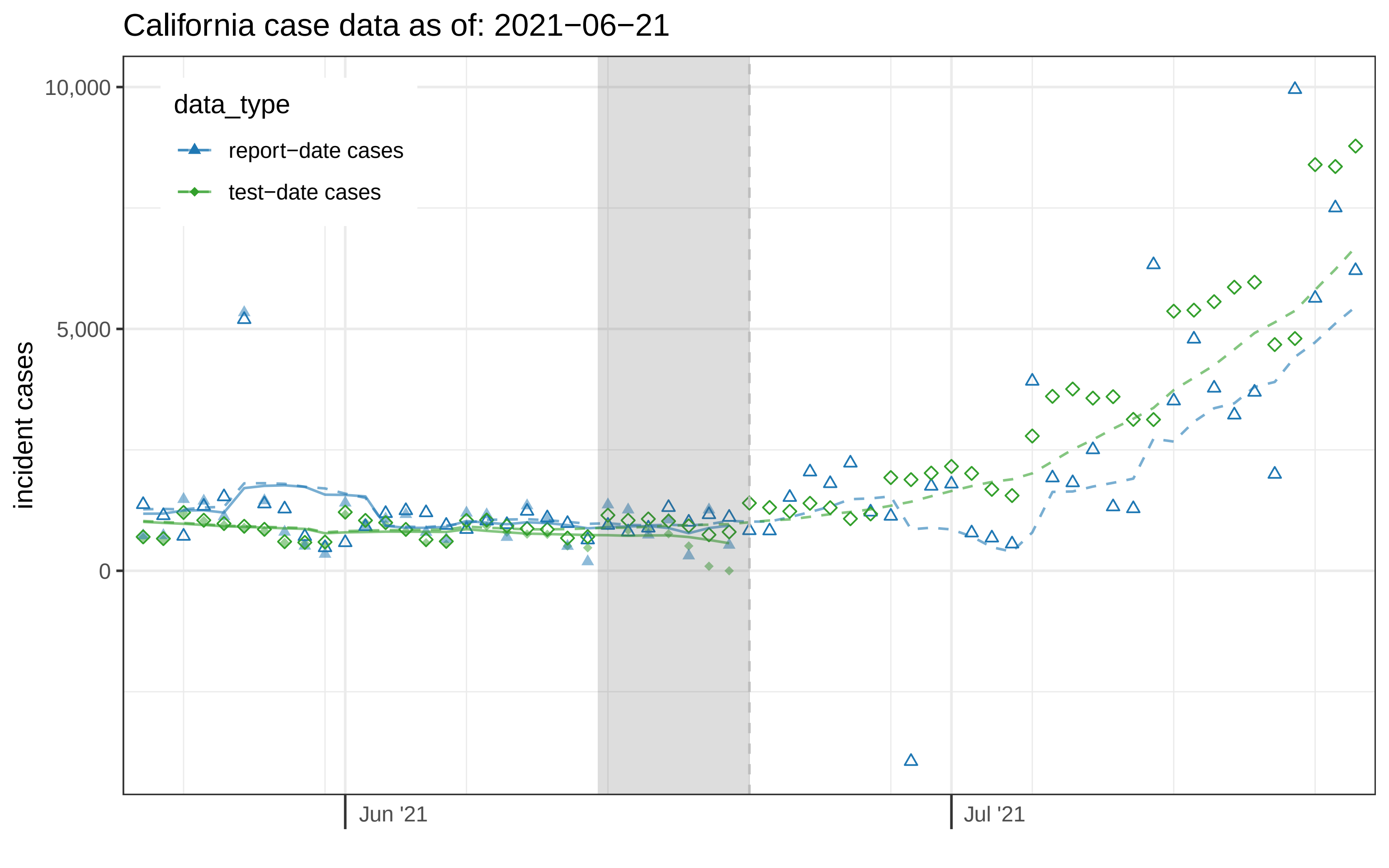
California case data as of: 2021−06−21
data_type
report−date cases
test−date cases
incident cases
10,000
5,000
0
Jun '21
Jul '21

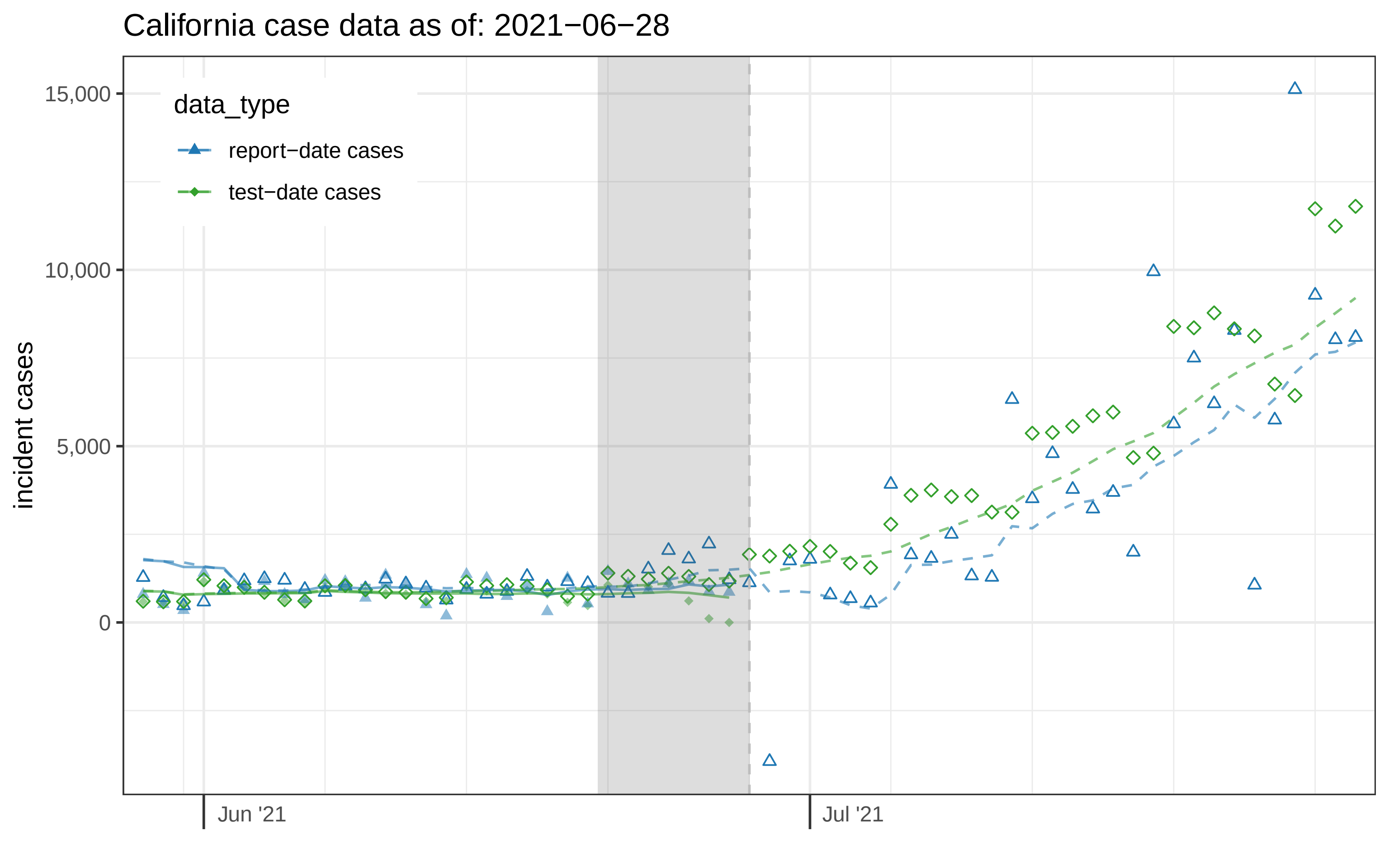
California case data as of: 2021−06−28
data_type
report−date cases
test−date cases
incident cases
15,000
10,000
5,000
0
Jun '21
Jul '21

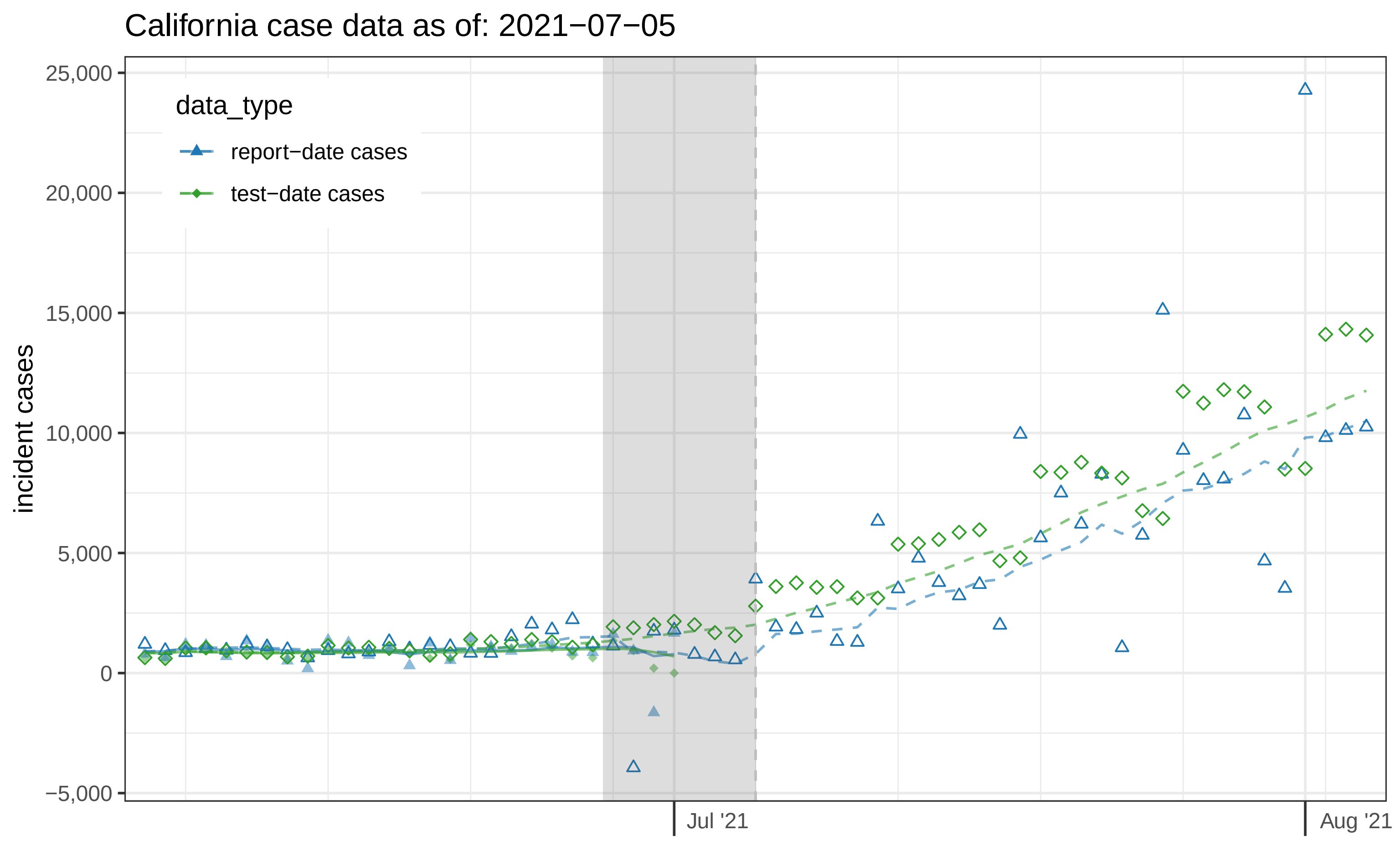
California case data as of: 2021−07−05
data_type
report−date cases
test−date cases
incident cases
25,000
20,000
15,000
10,000
5,000
0
−5,000
Jul '21
Aug '21

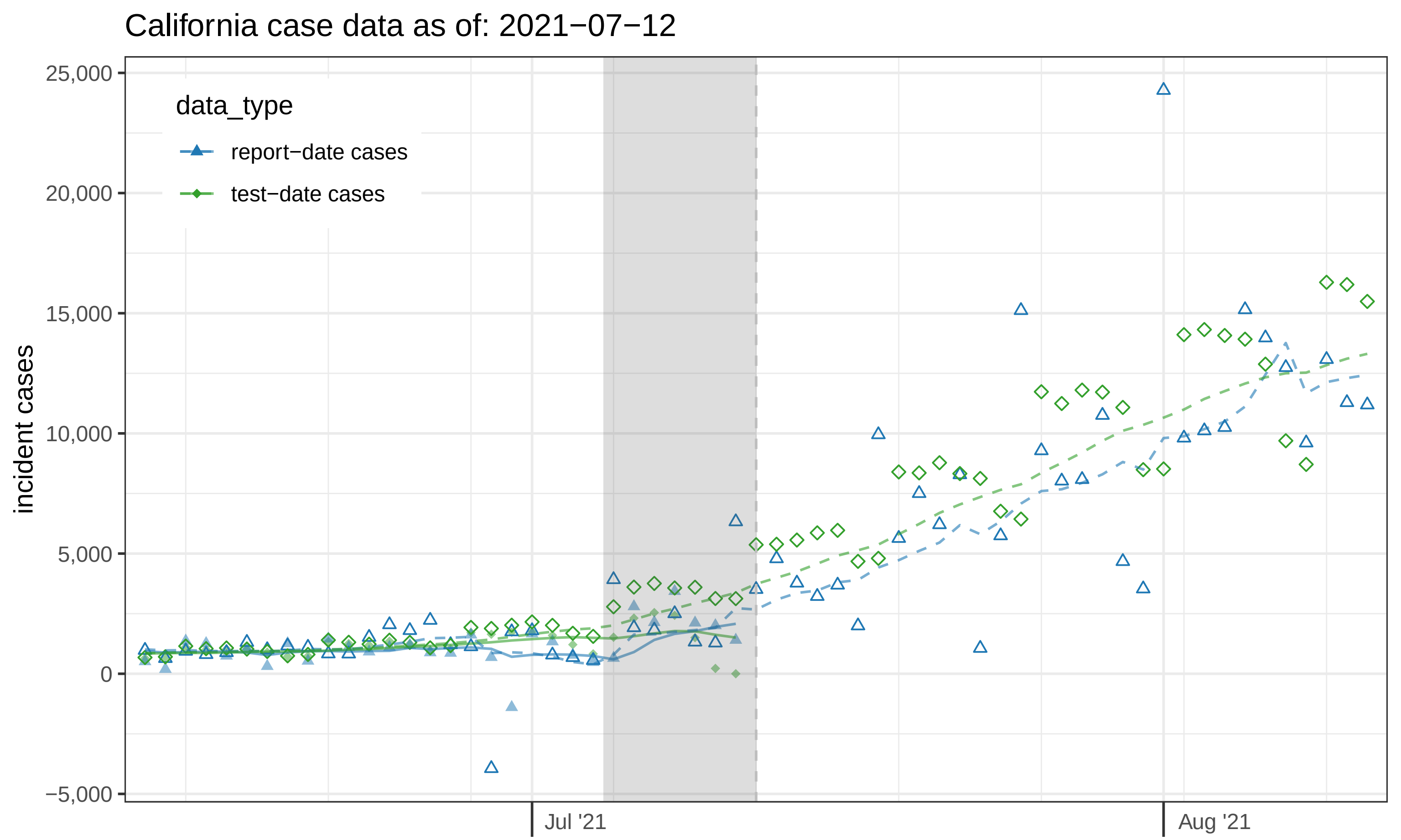

California case data as of: 2021−07−12
incident cases
25,000
20,000
15,000
10,000
5,000
0
−5,000
Jul '21
Aug '21
data_type
report−date cases
test−date cases

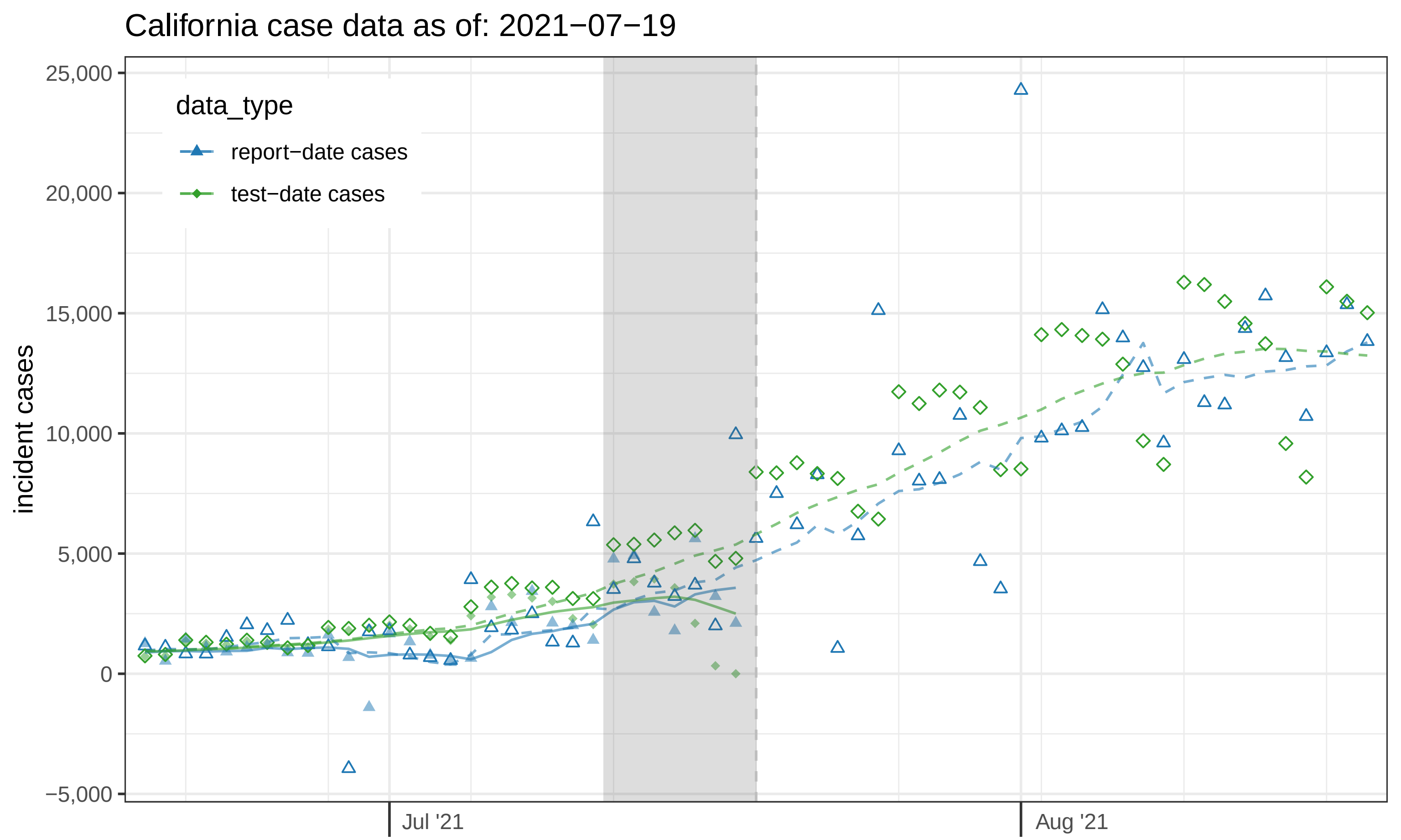

California case data as of: 2021−07−19
data_type
report−date cases
test−date cases
incident cases
25,000
20,000
15,000
10,000
5,000
0
−5,000
Jul '21
Aug '21

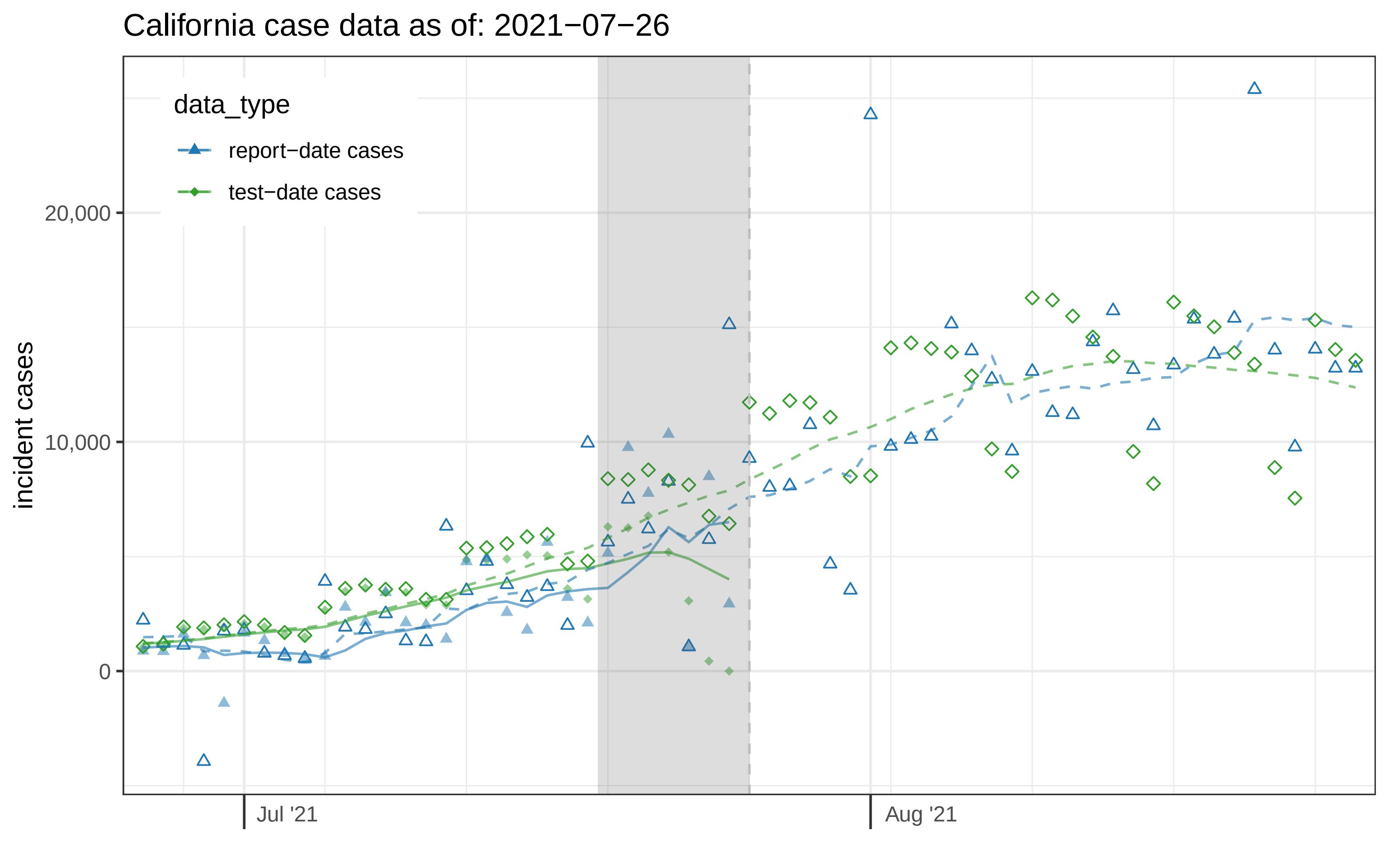

California case data as of: 2021−07−26
data_type
report−date cases
test−date cases
incident cases
20,000
10,000
0
Jul '21
Aug '21

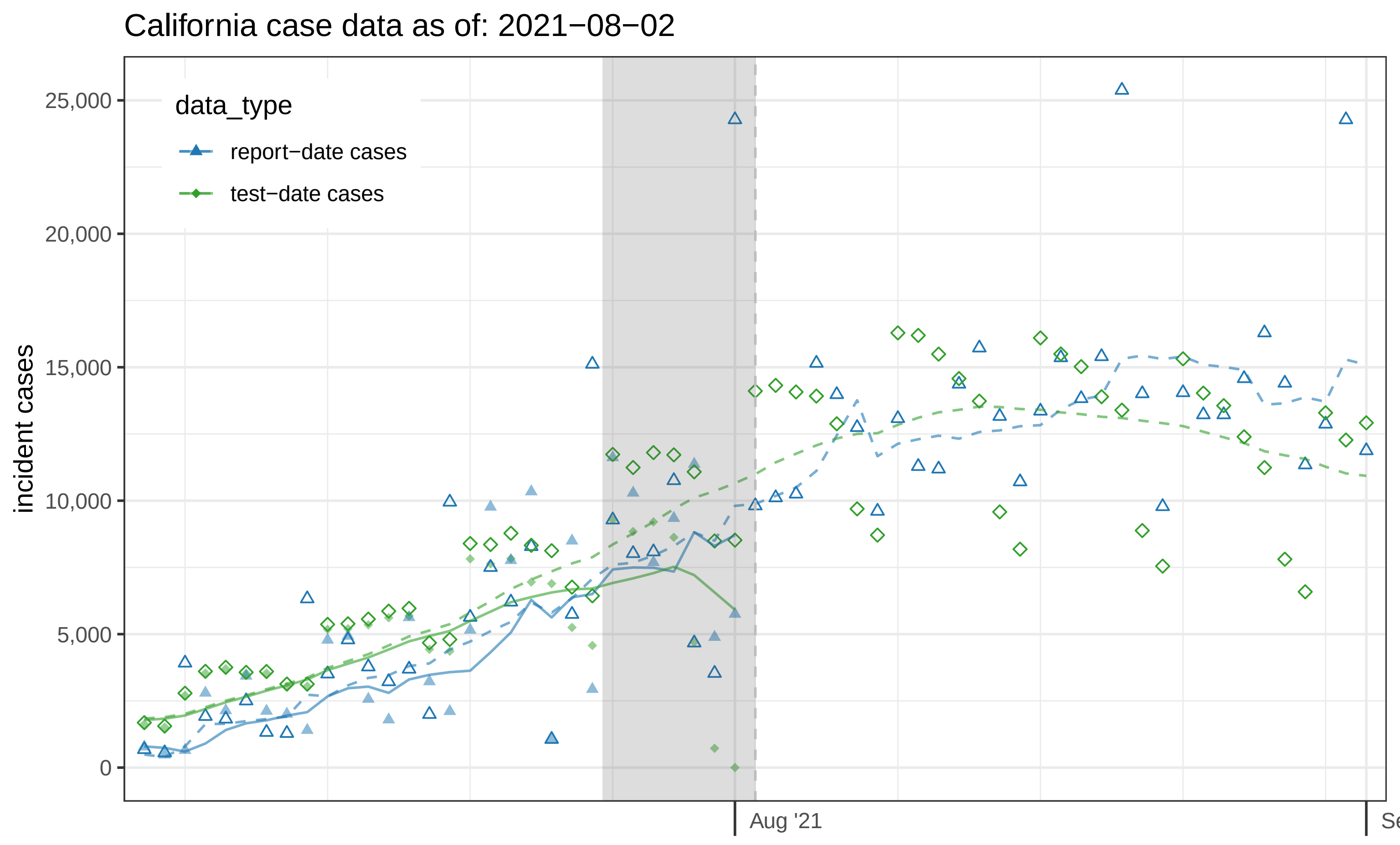

California case data as of: 2021−08−02
data_type
report−date cases
test−date cases
incident cases
25,000
20,000
15,000
10,000
5,000
0
Aug '21

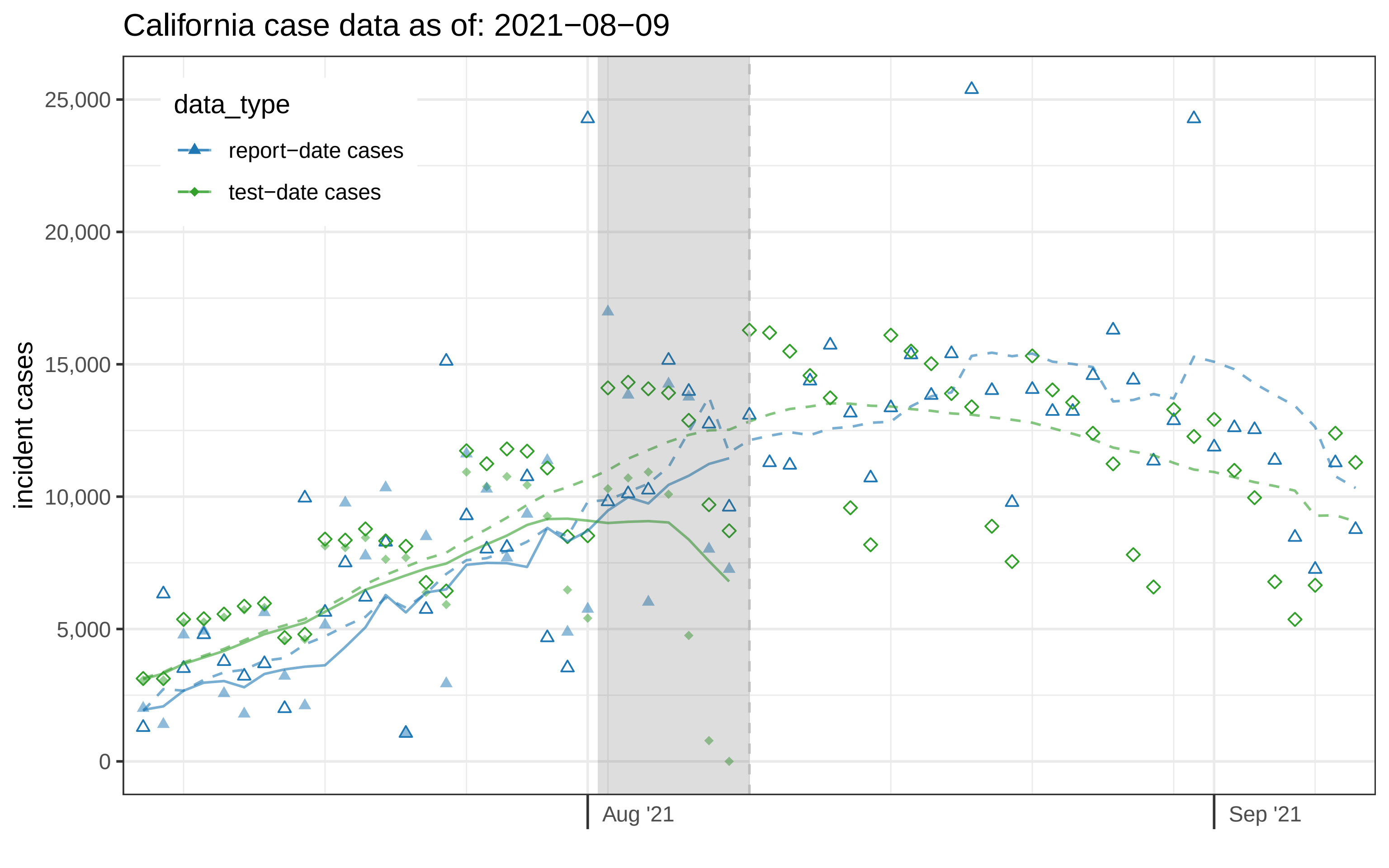
California case data as of: 2021−08−09
data_type
report−date cases
test−date cases
incident cases
25,000
20,000
15,000
10,000
5,000
0
Aug '21
Sep '21

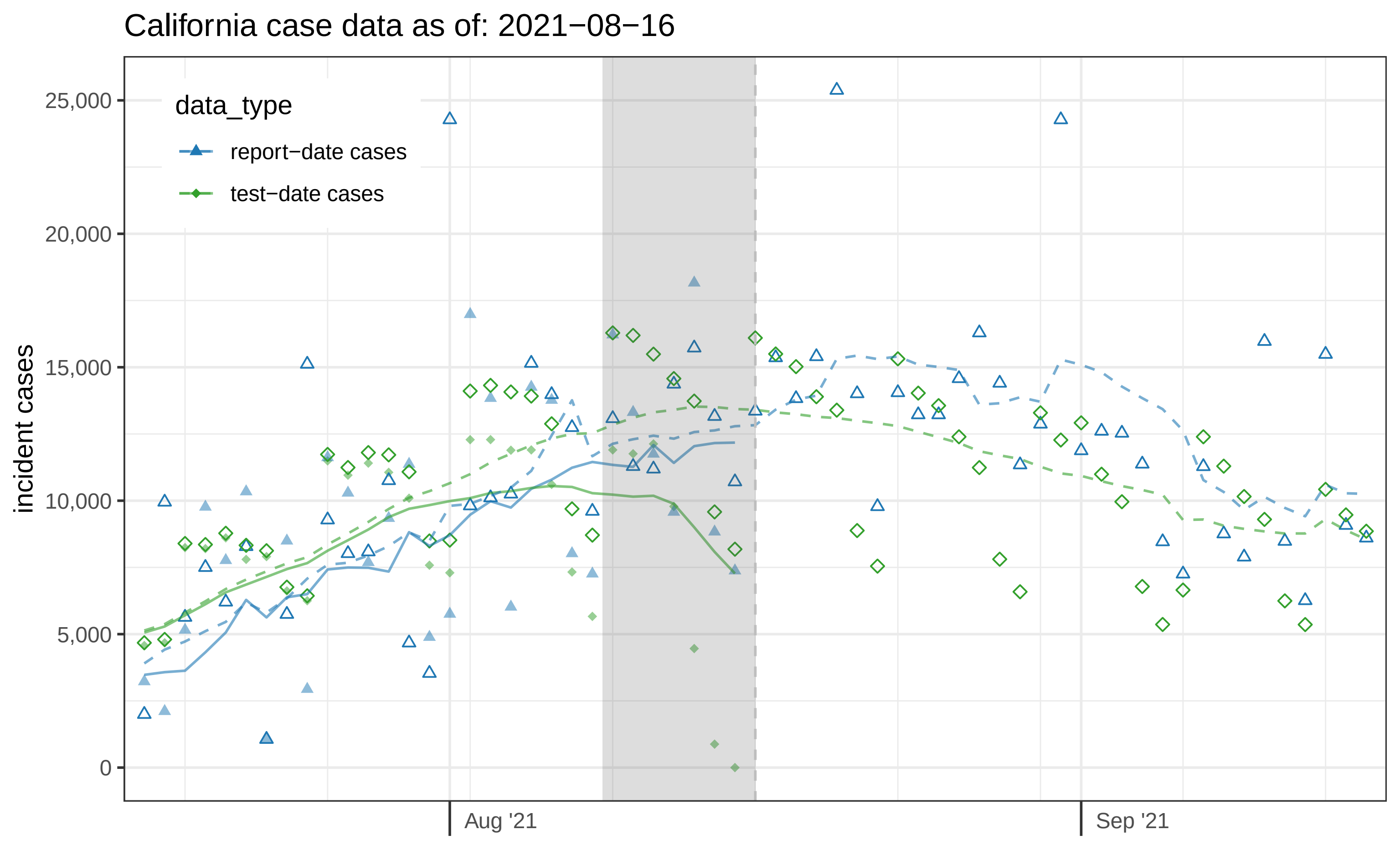
California case data as of: 2021−08−16
data_type
report−date cases
test−date cases
incident cases
25,000
20,000
15,000
10,000
5,000
0
Aug '21
Sep '21

### California case data as of: 2021−08−23

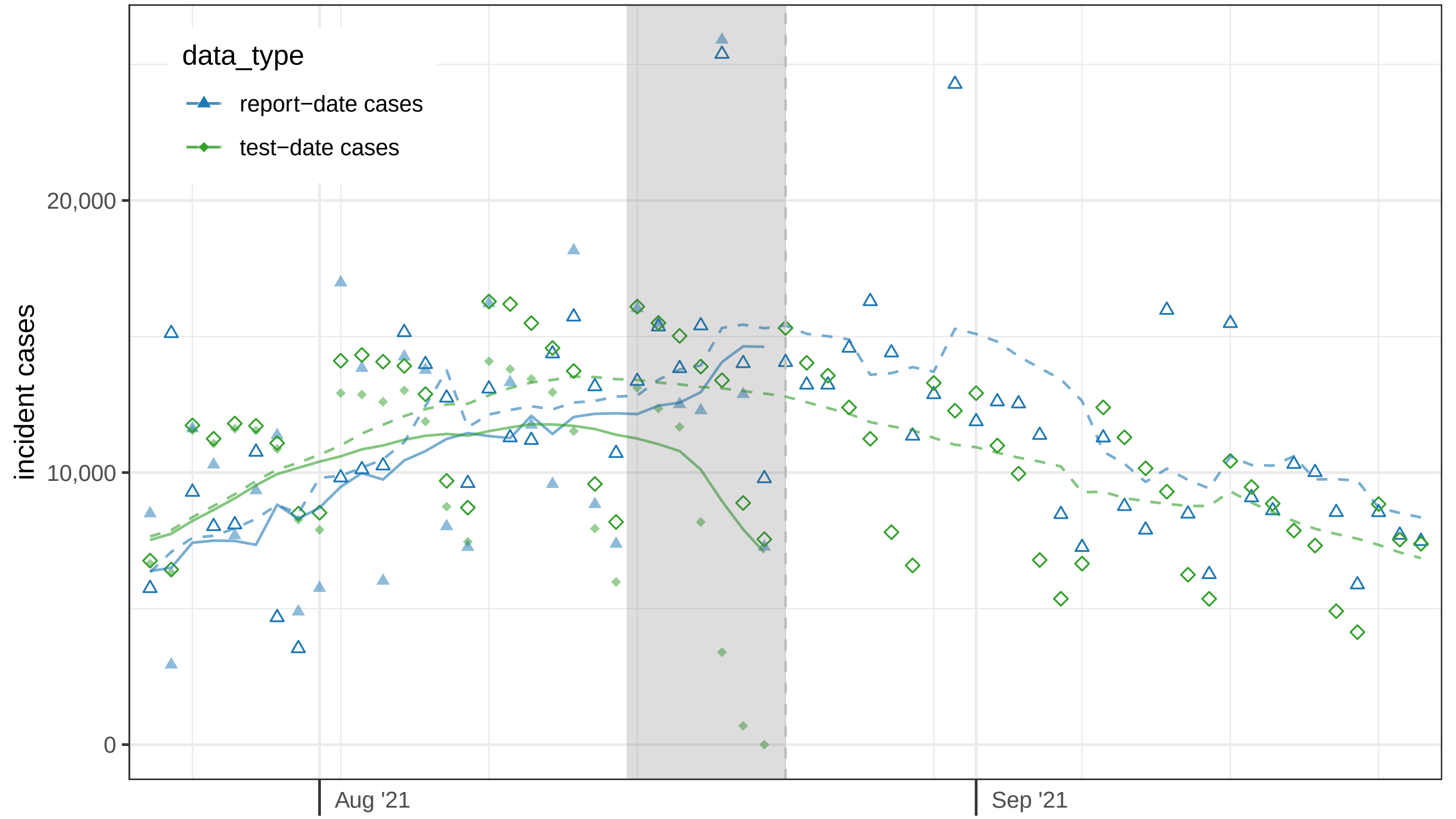

#### California case data as of: 2021−08−30

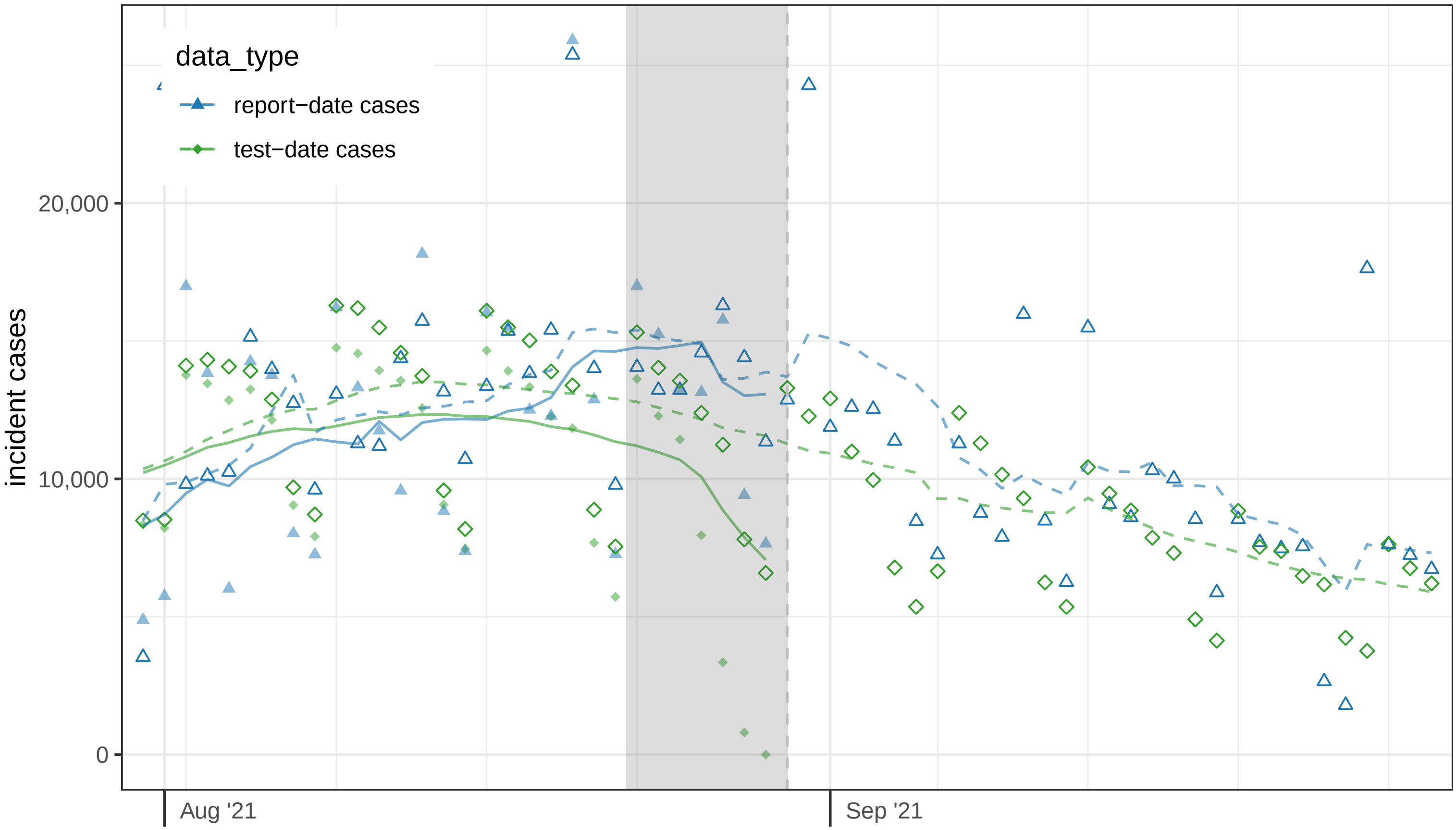

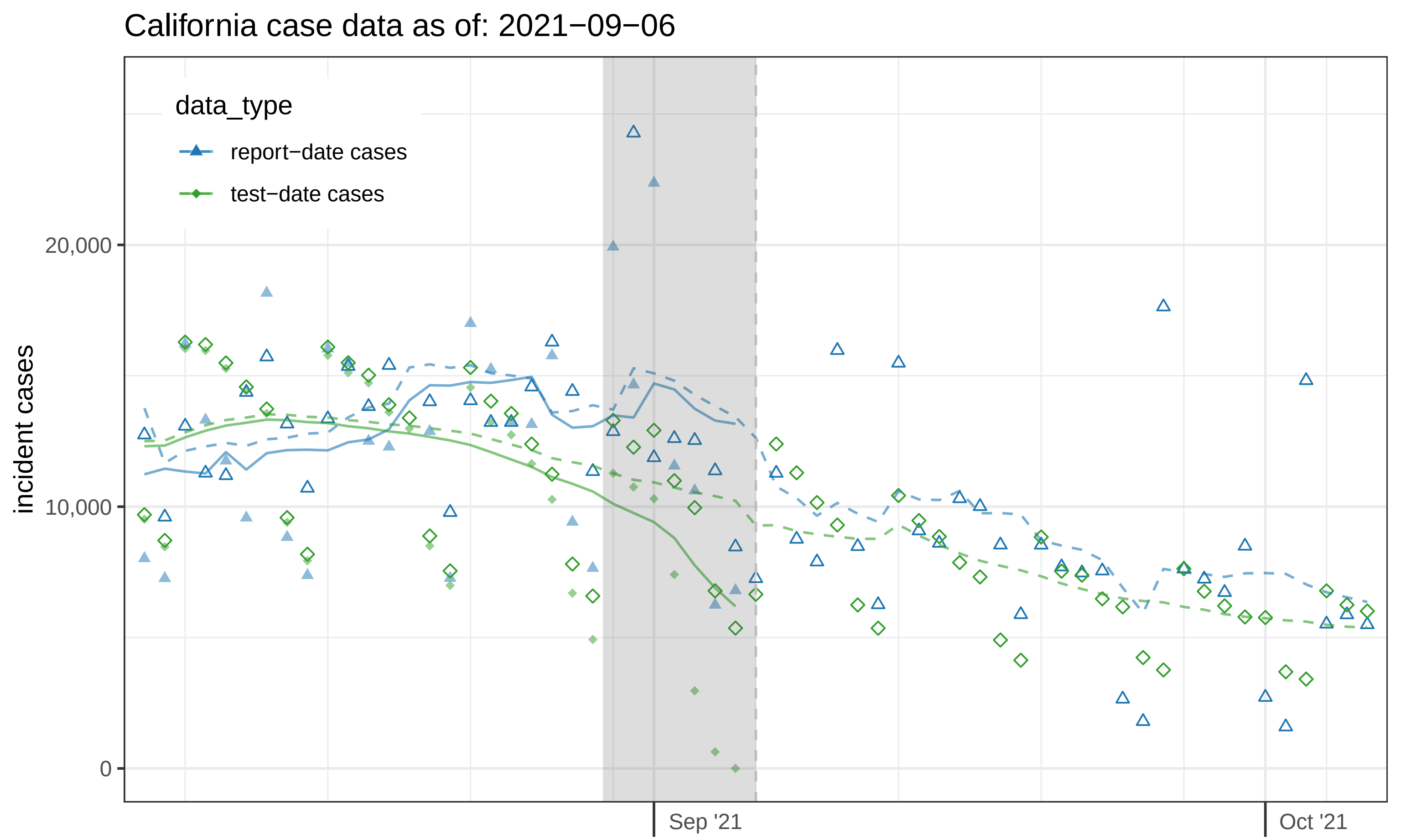
California case data as of: 2021−09−06
data_type
report−date cases
test−date cases
incident cases
20,000
10,000
0
Sep '21
Oct '21

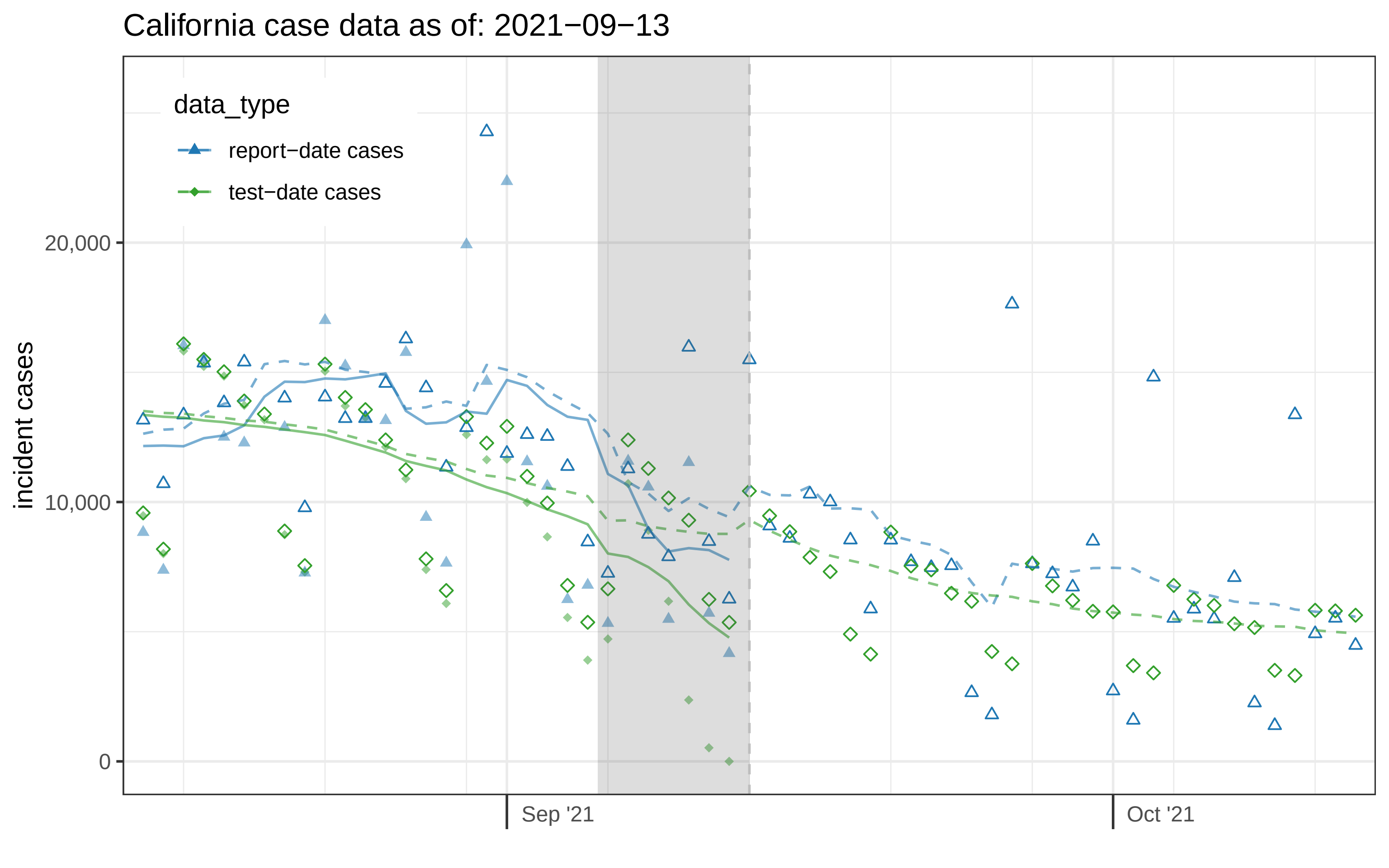
California case data as of: 2021−09−13
data_type
report−date cases
test−date cases
incident cases
20,000
10,000
0
Sep '21
Oct '21

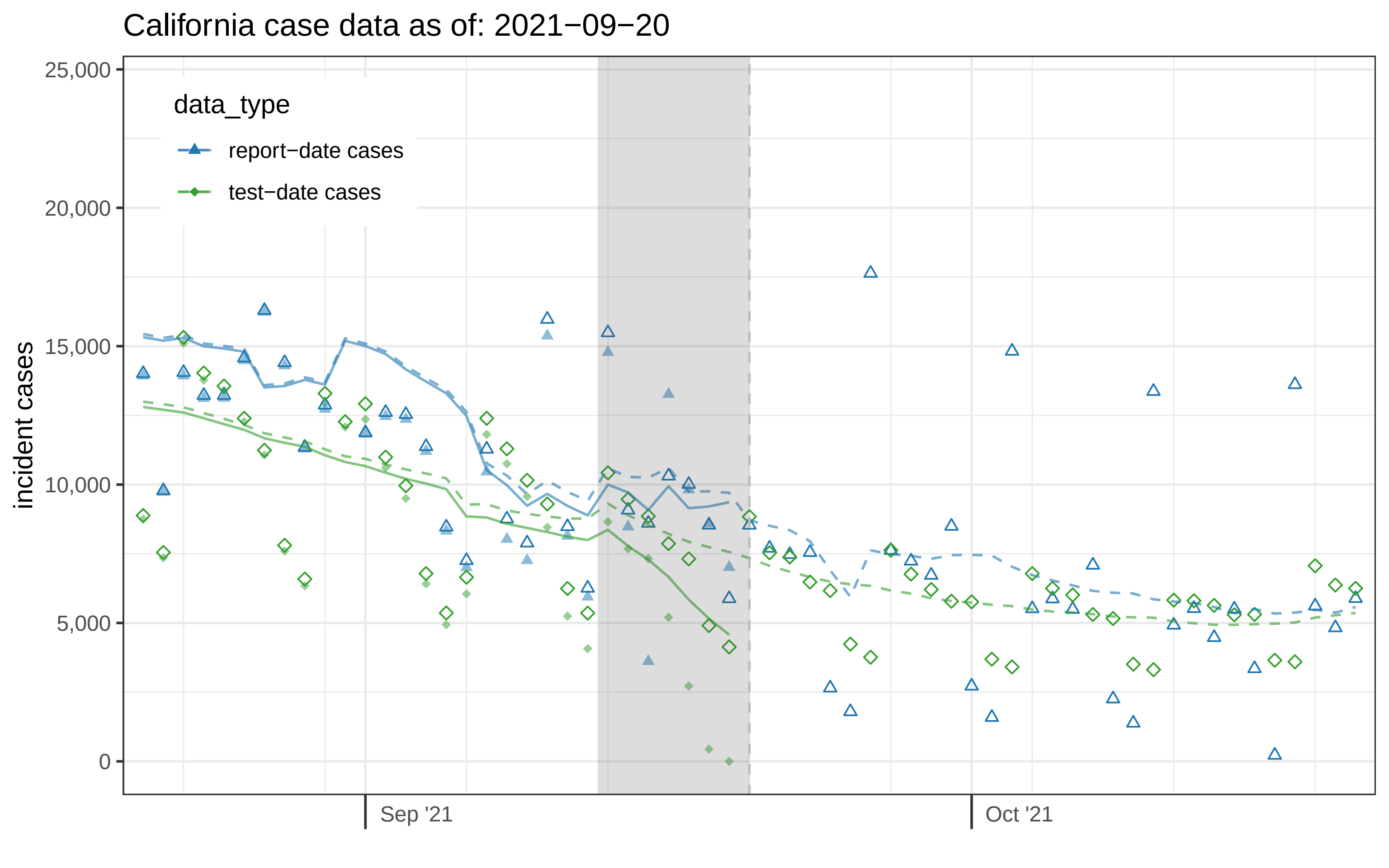

California case data as of: 2021−09−20
data_type
report−date cases
test−date cases
incident cases
25,000
20,000
15,000
10,000
5,000
0
Sep '21
Oct '21

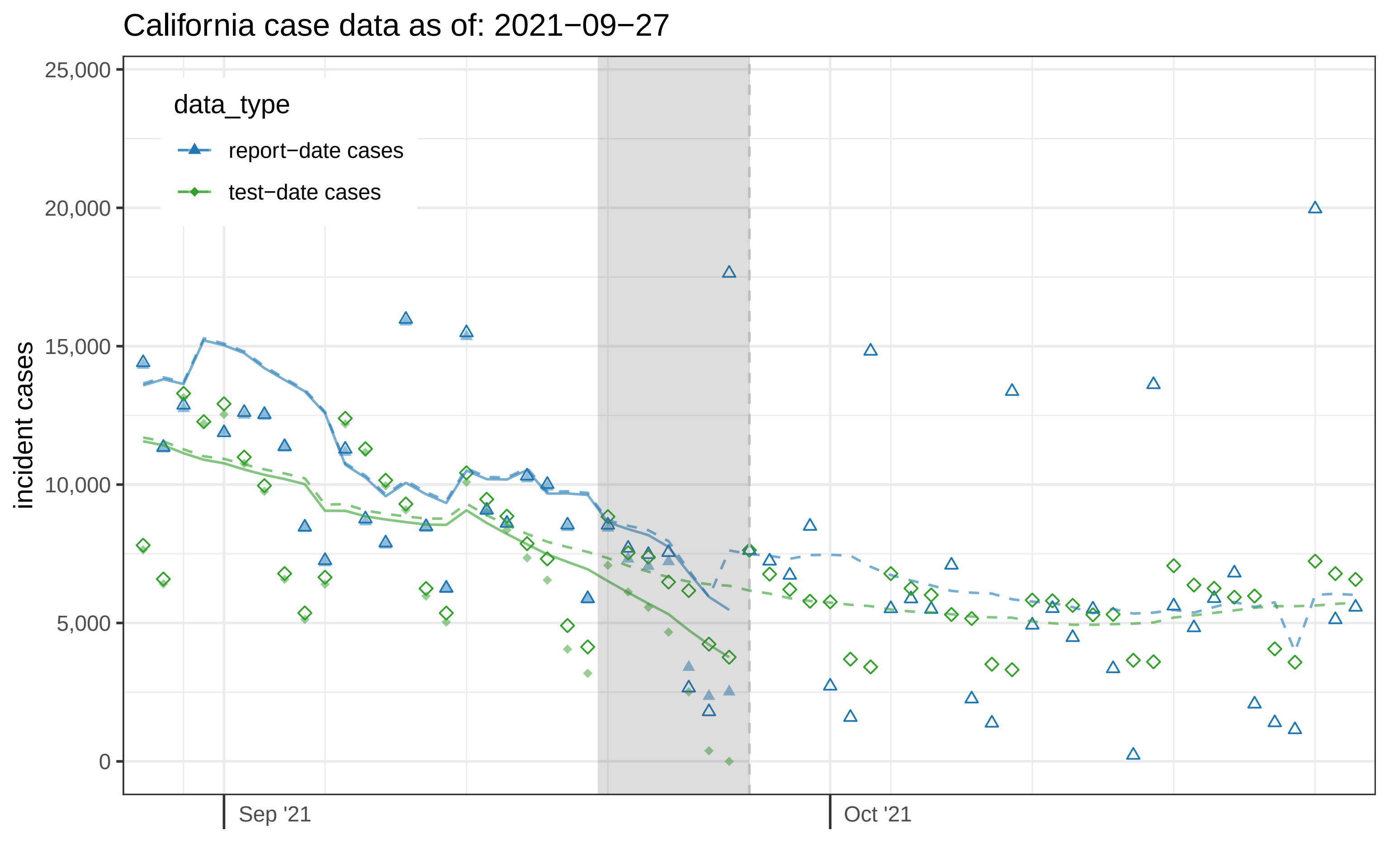

California case data as of: 2021−09−27
data_type
report−date cases
test−date cases
incident cases
25,000
20,000
15,000
10,000
5,000
0
Sep '21
Oct '21

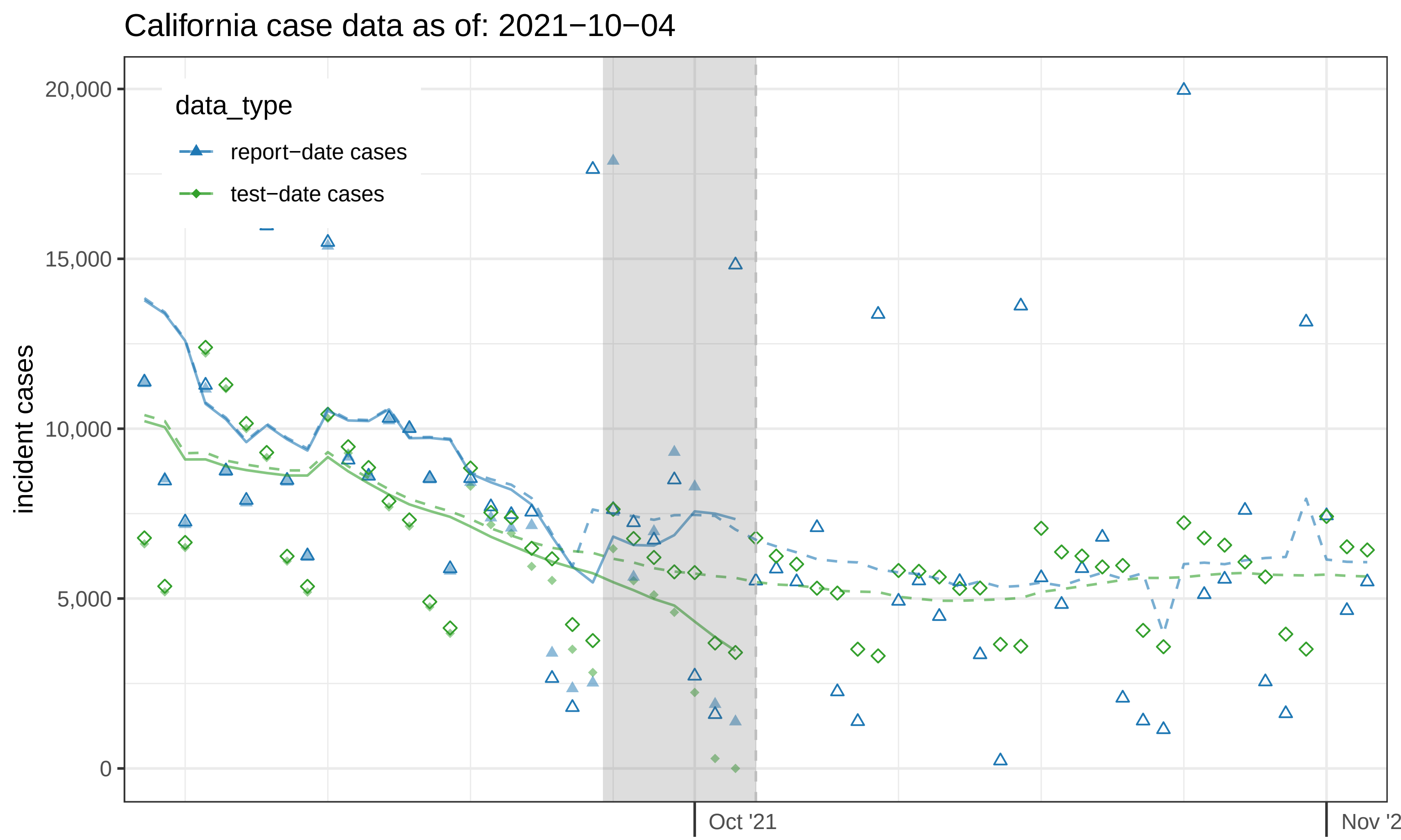

California case data as of: 2021−10−04
data_type
report−date cases
test−date cases
incident cases
20,000
15,000
10,000
5,000
0
Oct '21
Nov '2

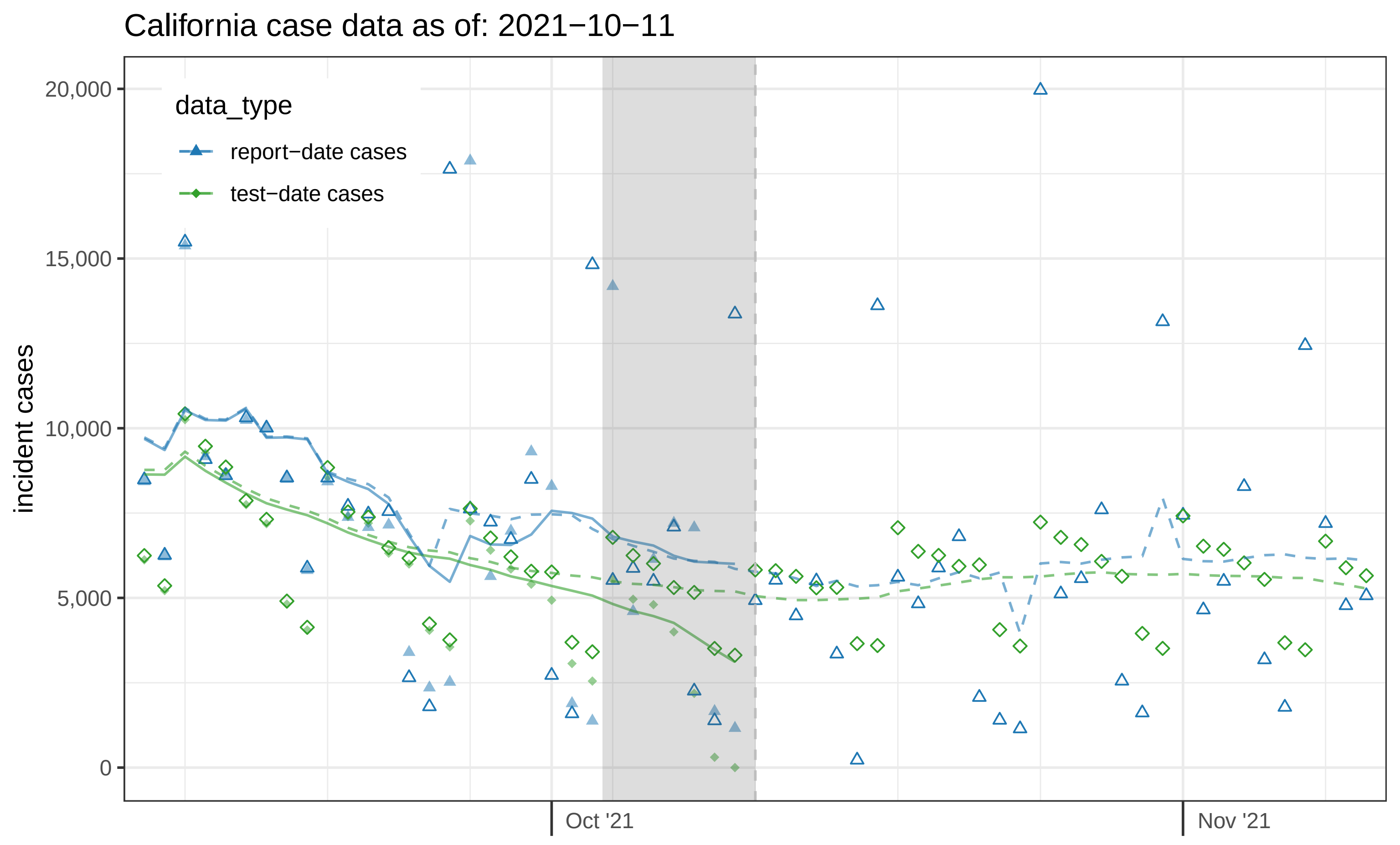

California case data as of: 2021−10−11
data_type
report−date cases
test−date cases
incident cases
20,000
15,000
10,000
5,000
0
Oct '21
Nov '21

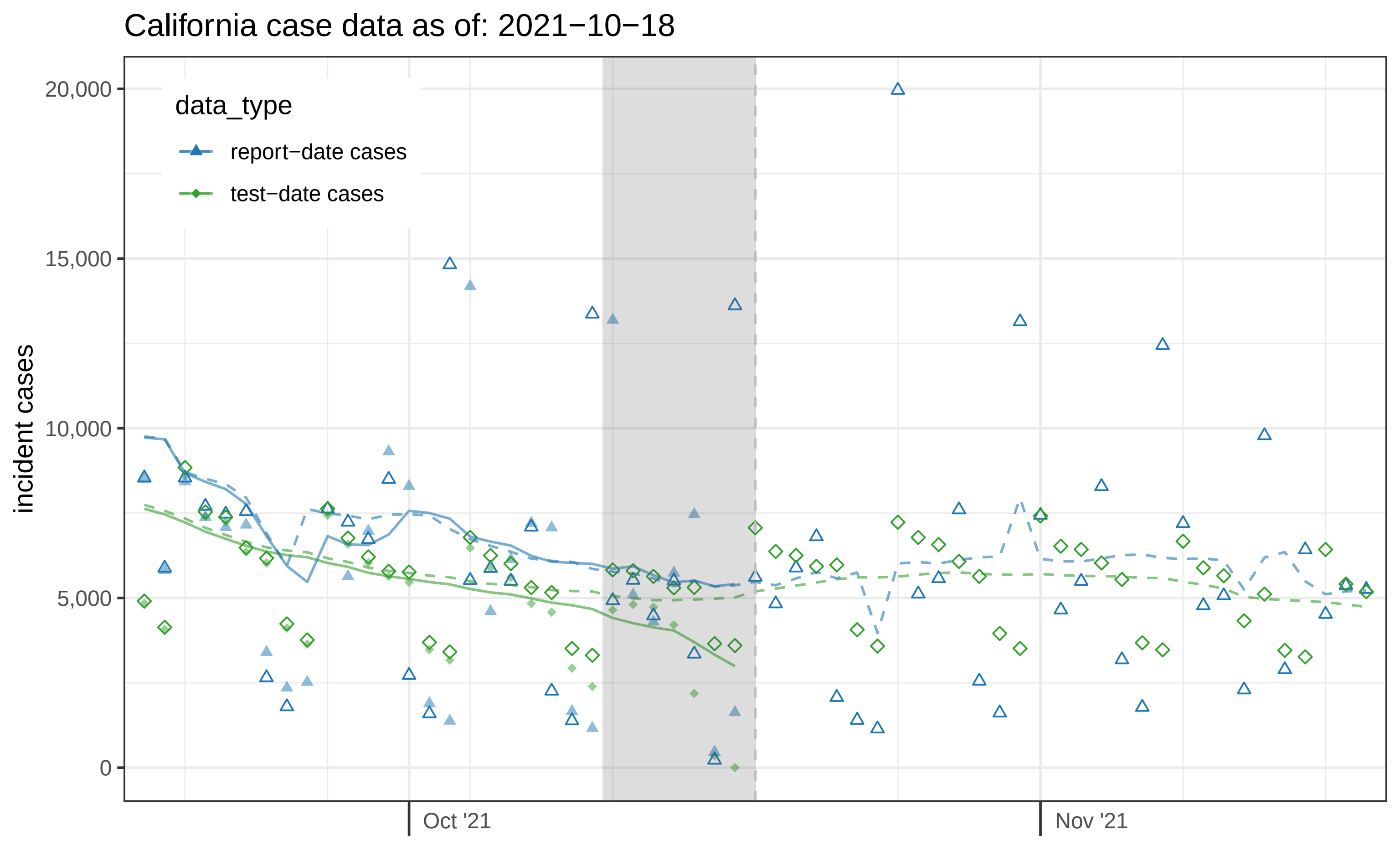

California case data as of: 2021−10−18
data_type
report−date cases
test−date cases
incident cases
20,000
15,000
10,000
5,000
0
Oct '21
Nov '21

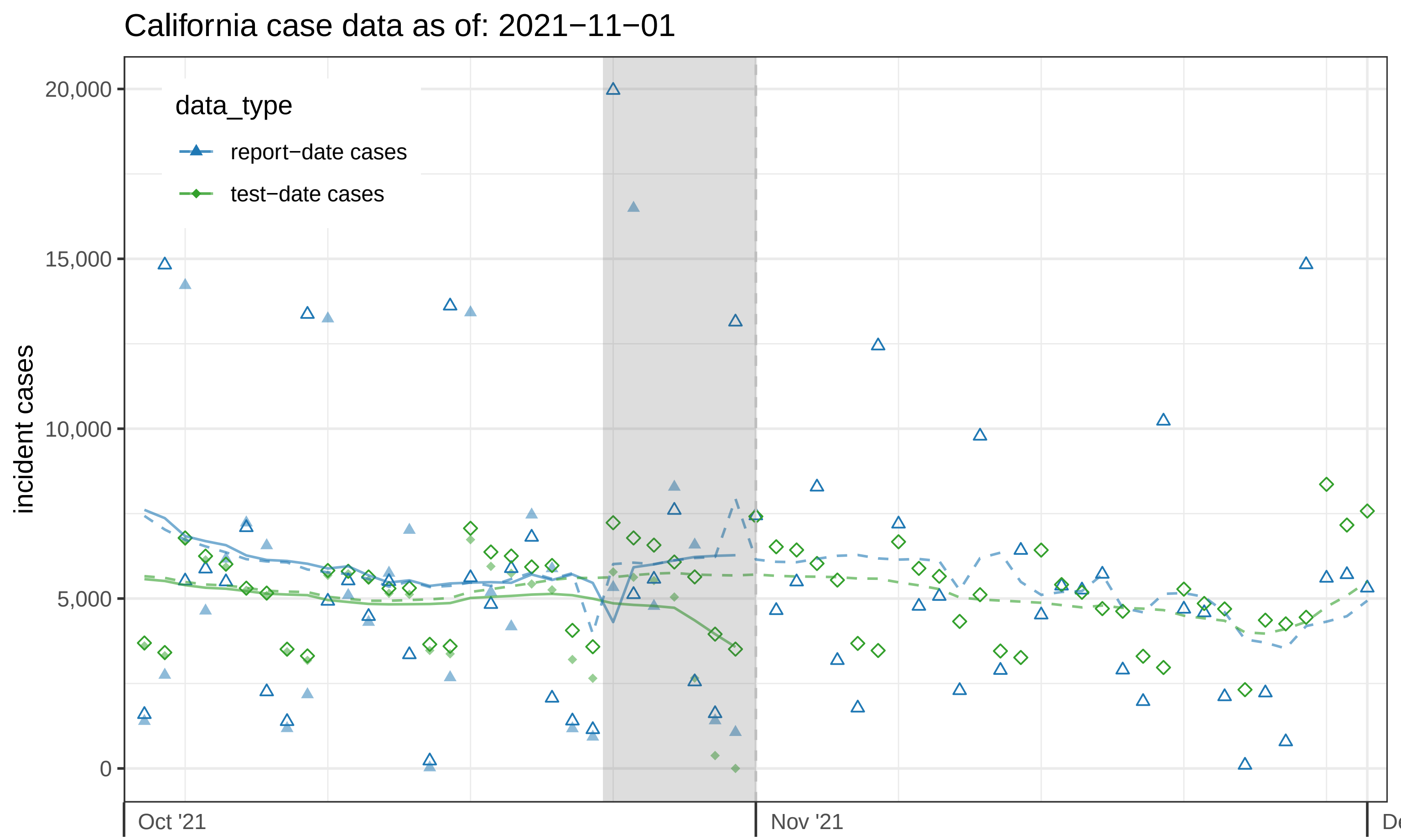

California case data as of: 2021−11−01
data_type
report−date cases
test−date cases
incident cases
20,000
15,000
10,000
5,000
0
Oct '21
Nov '21

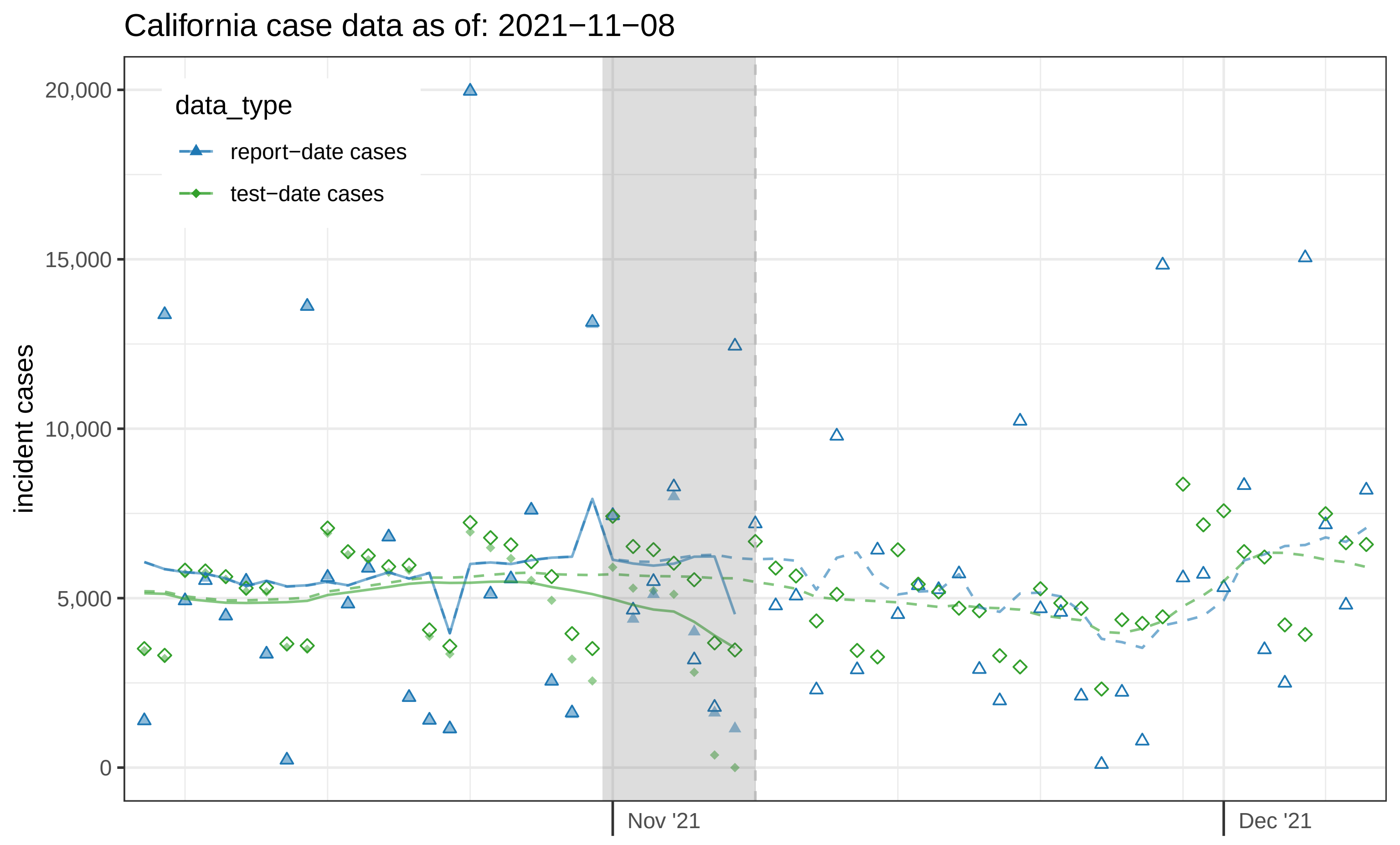
California case data as of: 2021−11−08
data_type
report−date cases
test−date cases
incident cases
20,000
15,000
10,000
5,000
0
Nov '21
Dec '21

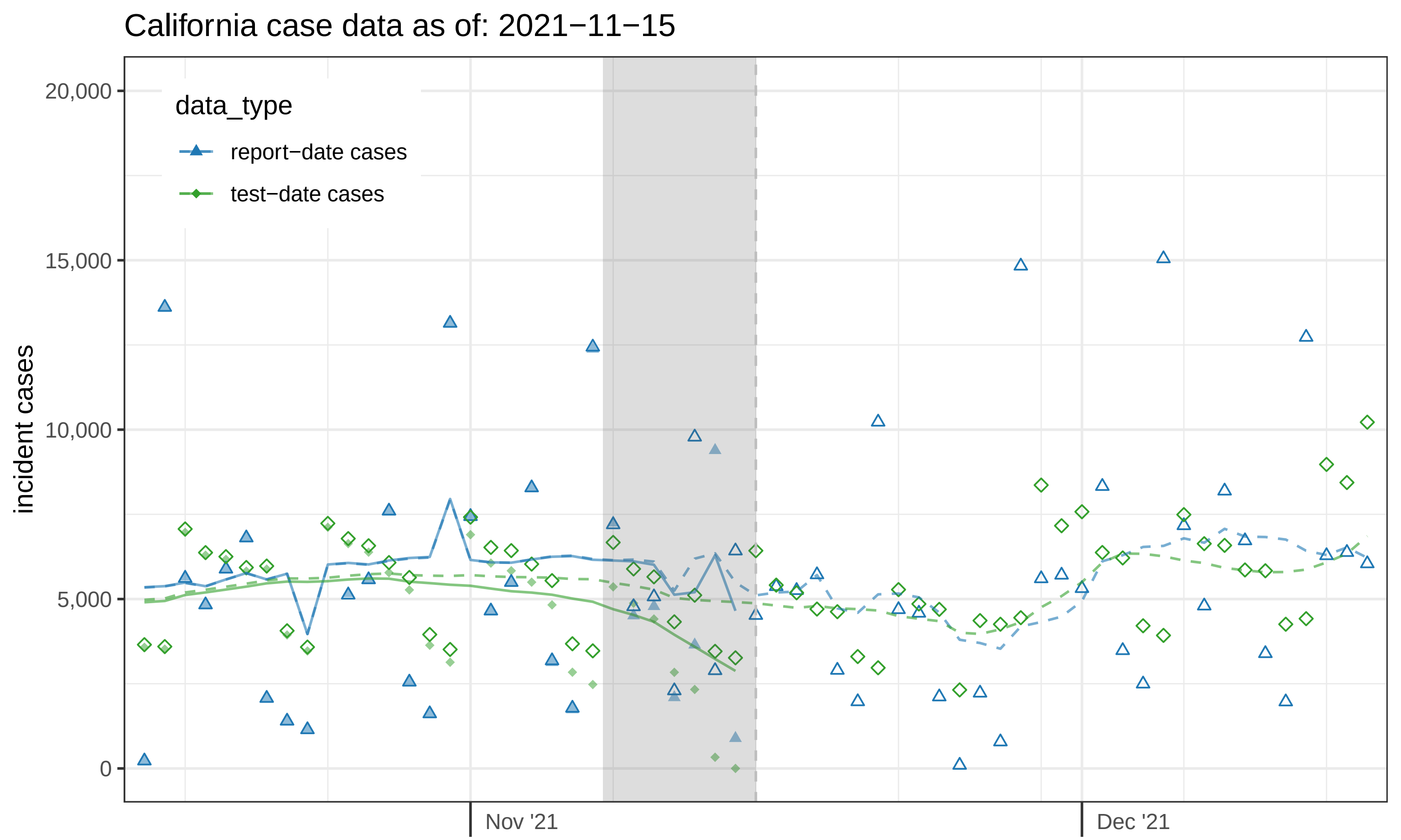
California case data as of: 2021−11−15
data_type
report−date cases
test−date cases
incident cases
20,000
15,000
10,000
5,000
0
Nov '21
Dec '21

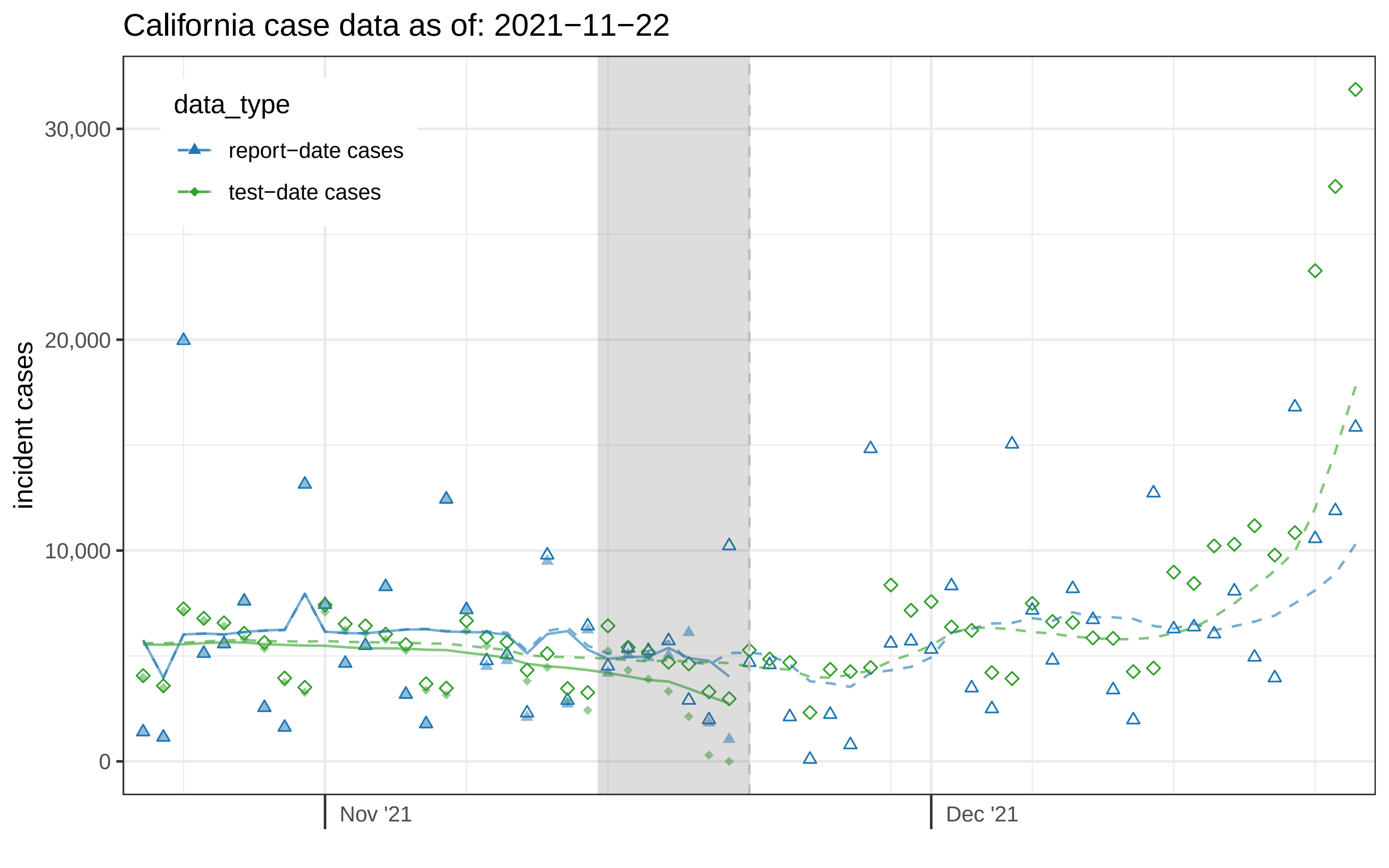
California case data as of: 2021−11−22
data_type
report−date cases
test−date cases
30,000
20,000
10,000
0
incident cases
Nov '21
Dec '21

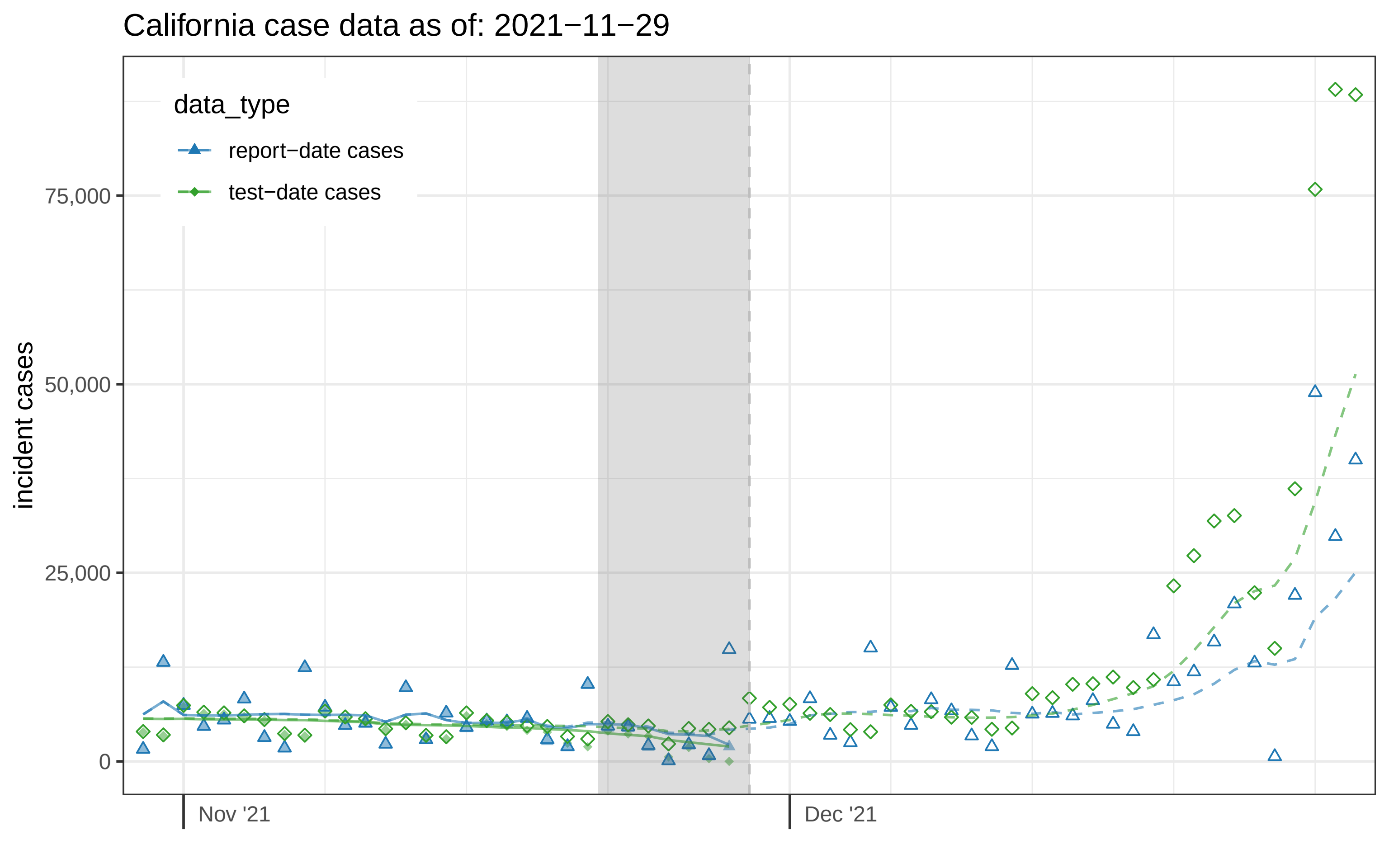

California case data as of: 2021−11−29
data_type
report−date cases
test−date cases
incident cases
75,000
50,000
25,000
0
Nov '21
Dec '21

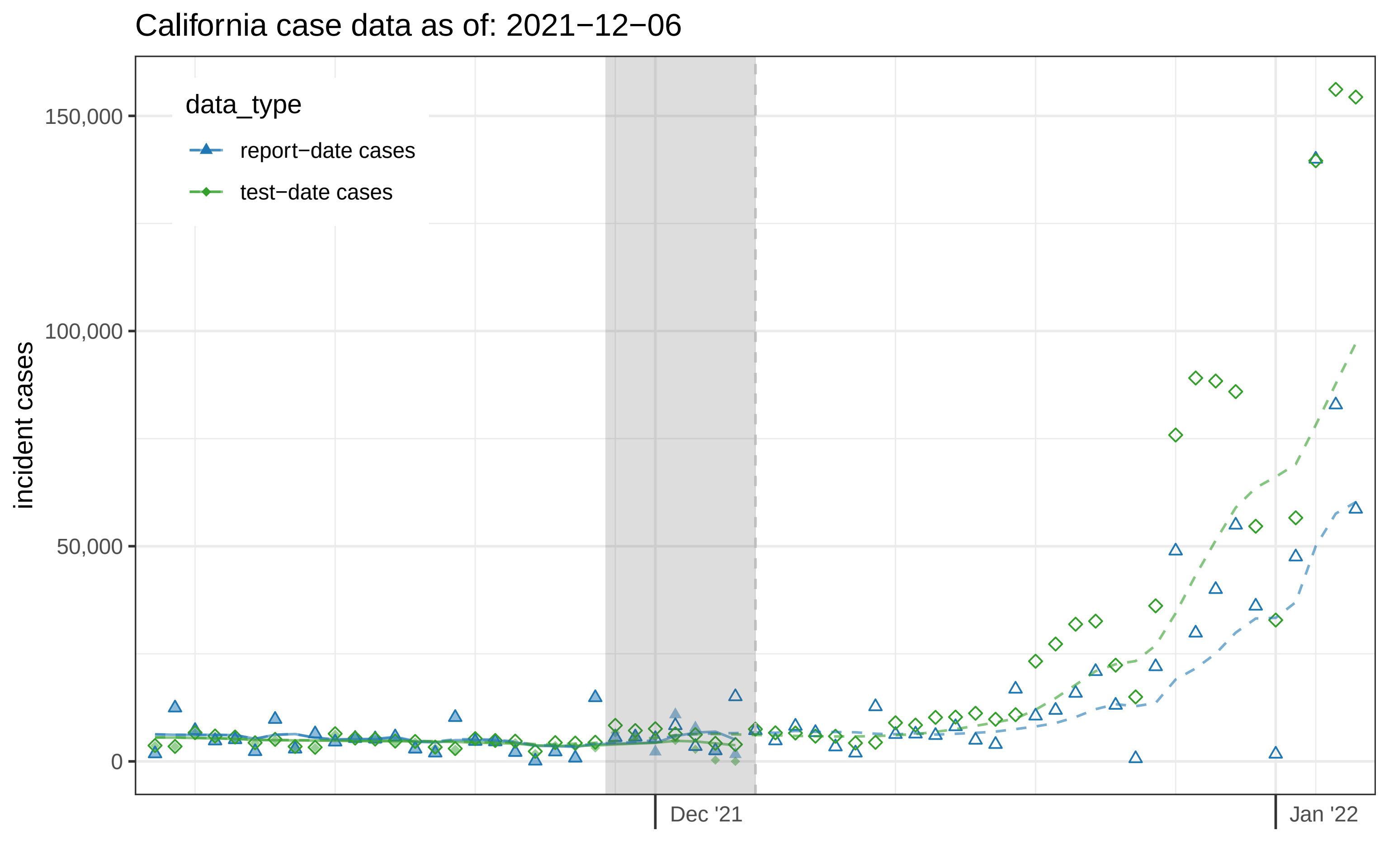

California case data as of: 2021−12−06
data_type
report−date cases
test−date cases
incident cases
150,000
100,000
50,000
0
Dec '21
Jan '22

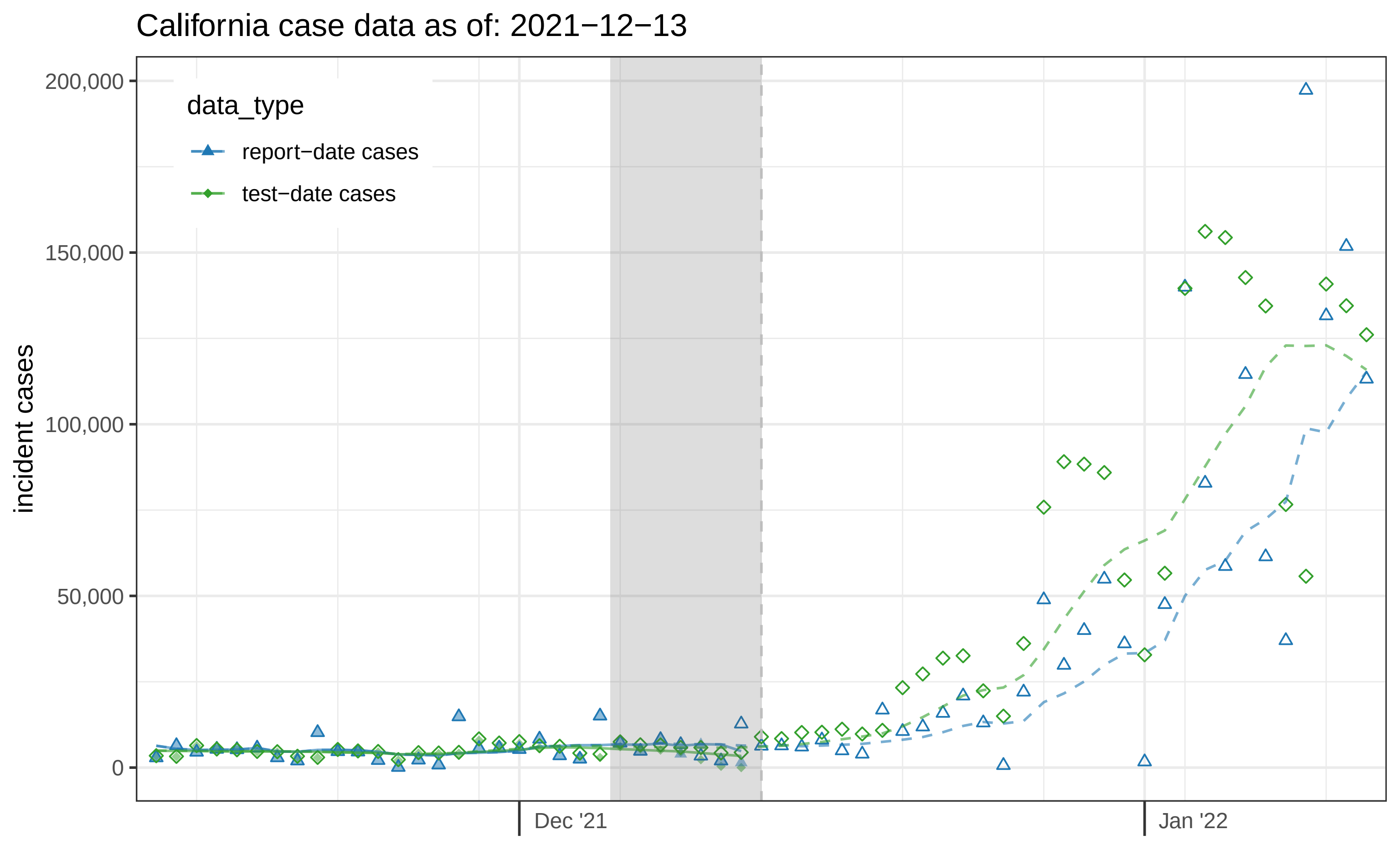

California case data as of: 2021−12−13
data_type
report−date cases
test−date cases
incident cases
200,000
150,000
100,000
50,000
0
Dec '21
Jan '22

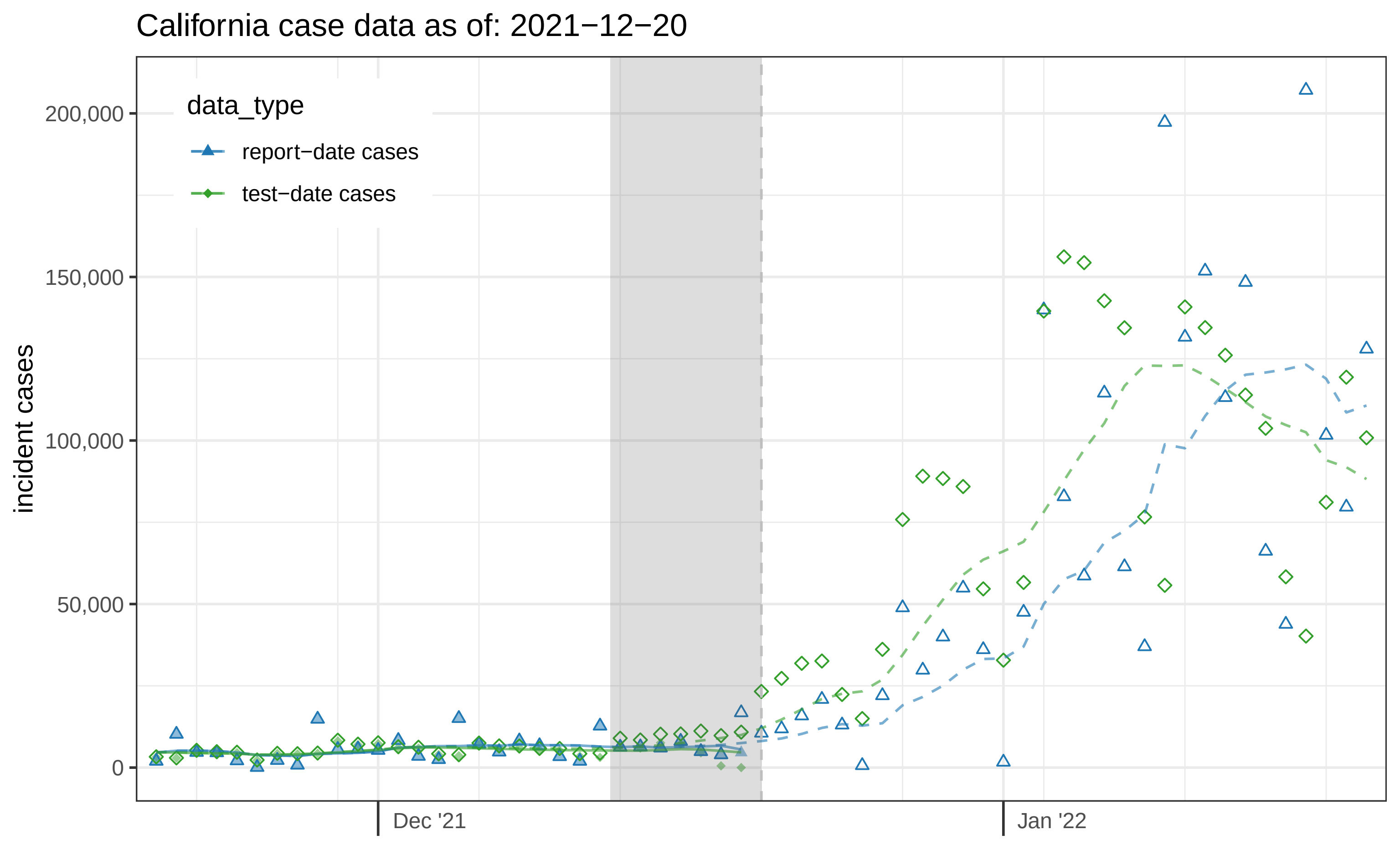

California case data as of: 2021−12−20
data_type
report−date cases
test−date cases
incident cases
200,000
150,000
100,000
50,000
0
Dec '21
Jan '22

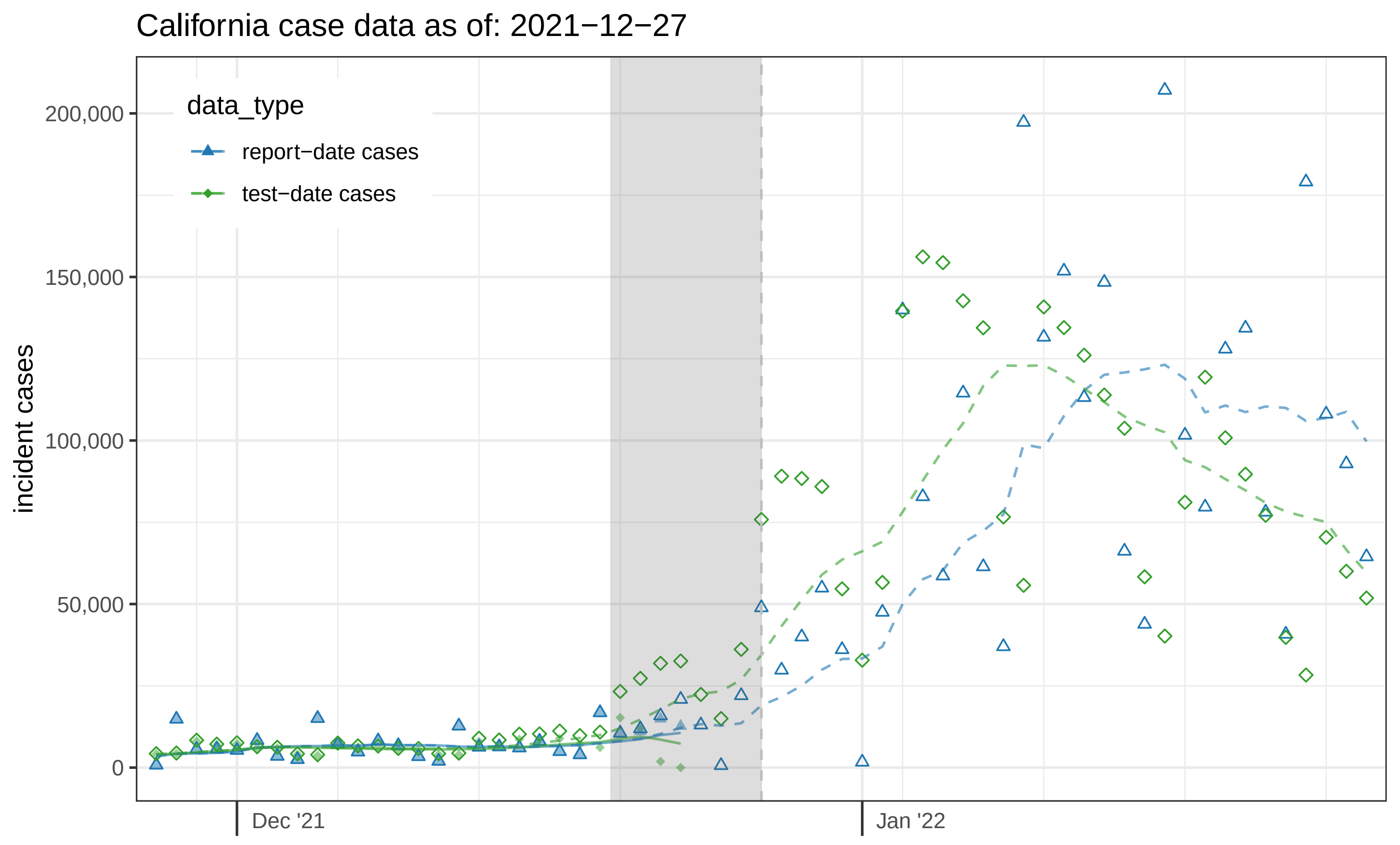

California case data as of: 2021−12−27
data_type
report−date cases
test−date cases
incident cases
200,000
150,000
100,000
50,000
0
Dec '21
Jan '22

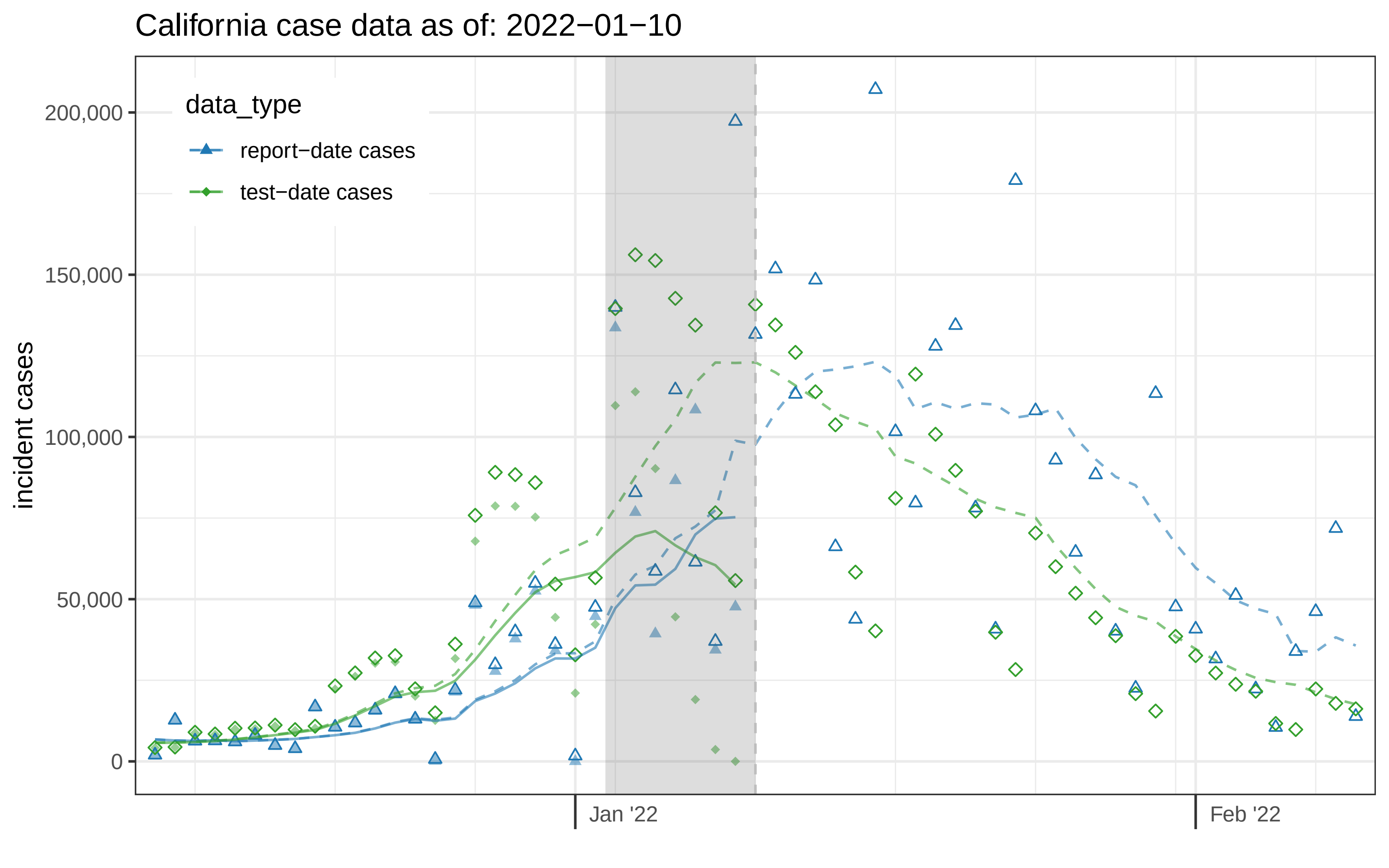
California case data as of: 2022−01−10
data_type
report−date cases
test−date cases
incident cases
200,000
150,000
100,000
50,000
0
Jan '22
Feb '22

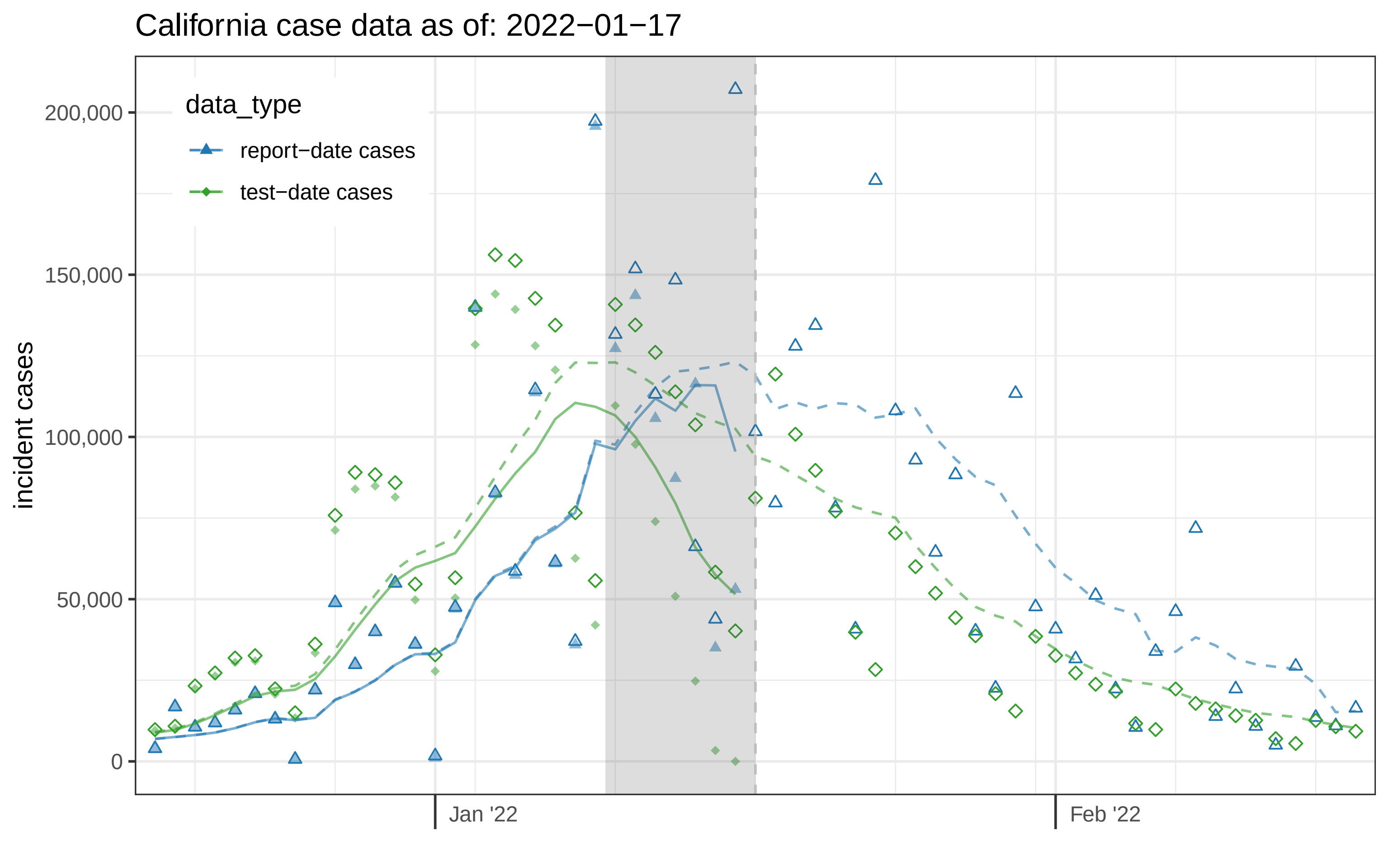

California case data as of: 2022−01−17
data_type
report−date cases
test−date cases
incident cases
200,000
150,000
100,000
50,000
0
Jan '22
Feb '22

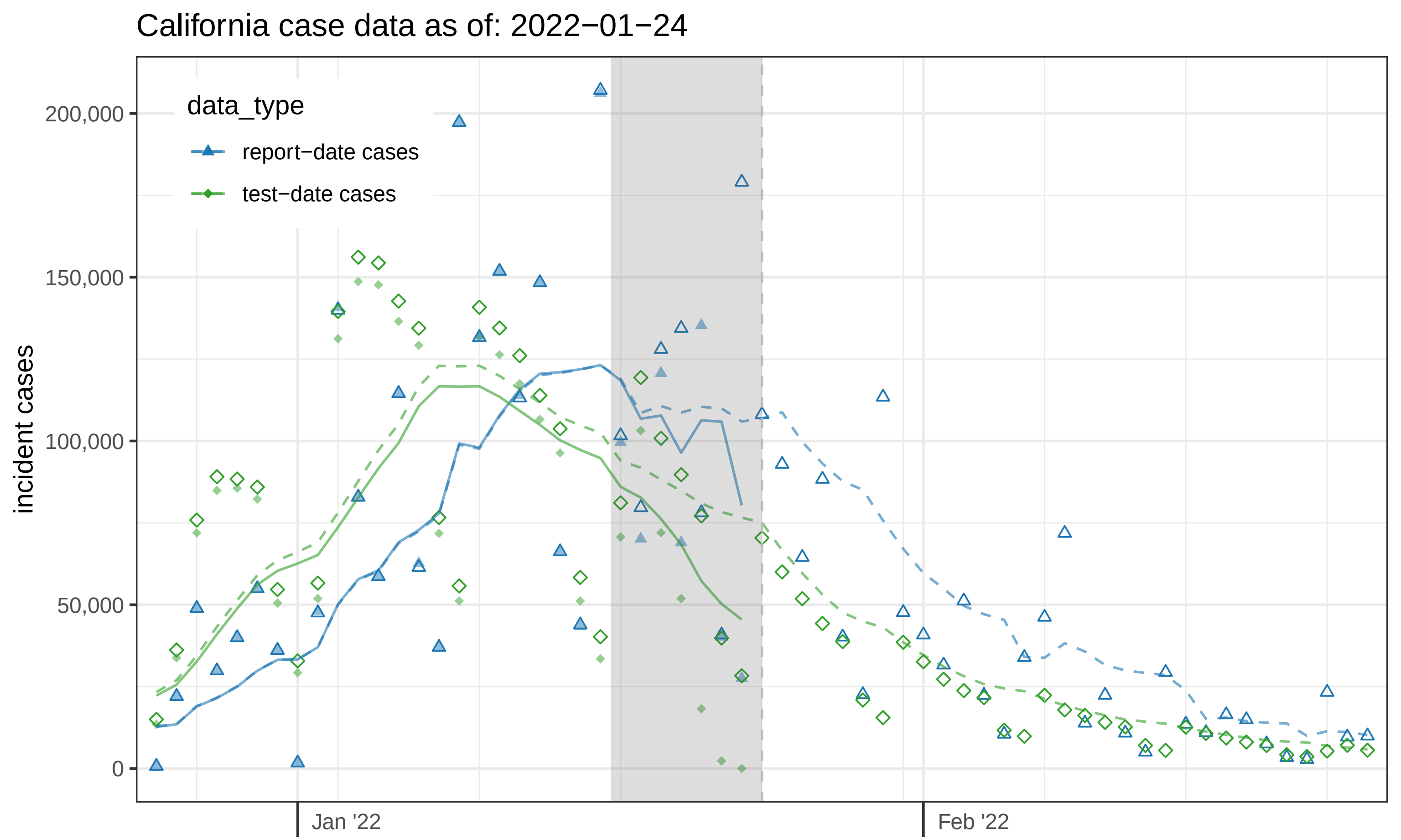
California case data as of: 2022−01−24
data_type
report−date cases
test−date cases
incident cases
200,000
150,000
100,000
50,000
0
Jan '22
Feb '22

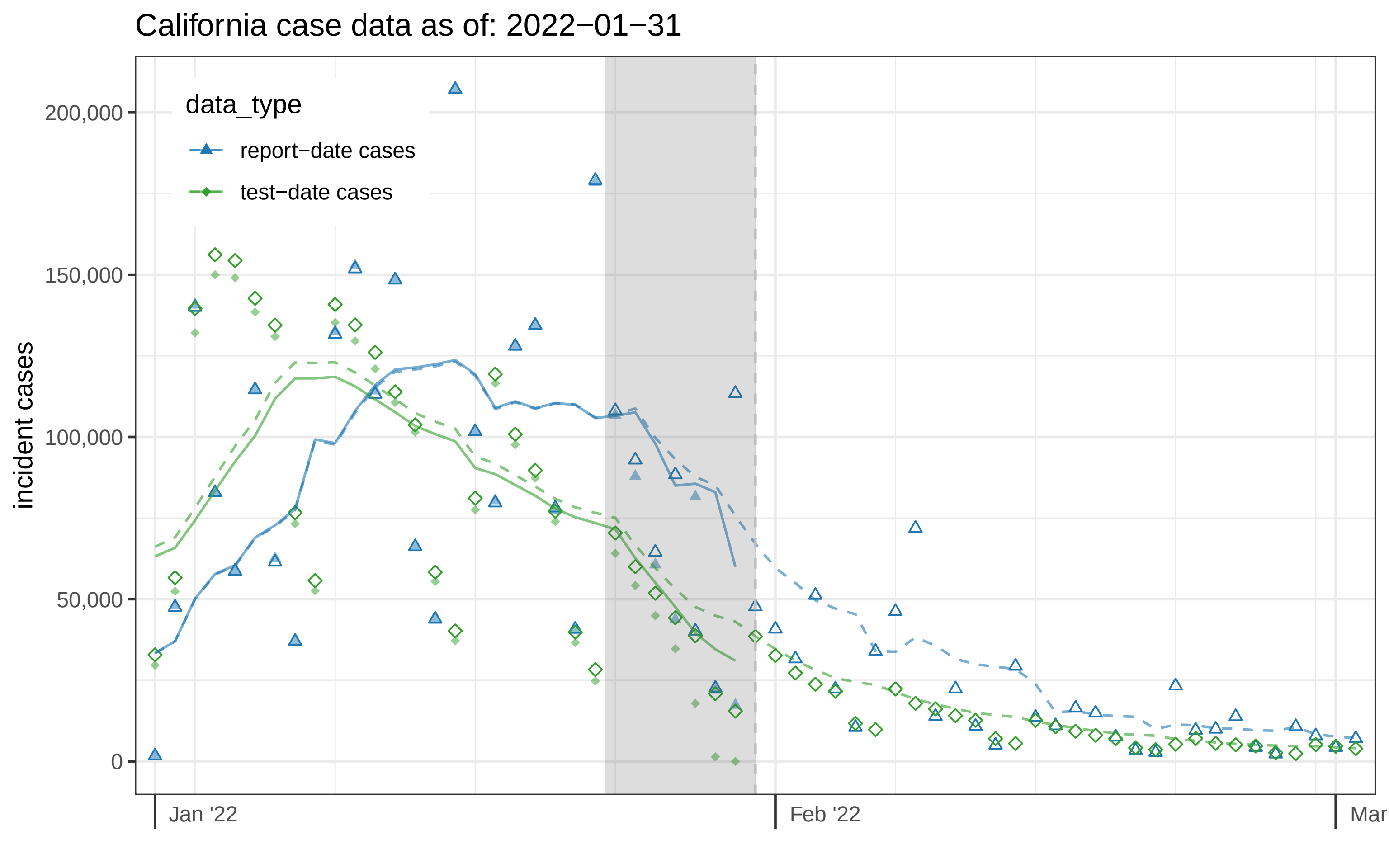
California case data as of: 2022−01−31
data_type
report−date cases
test−date cases
incident cases
200,000
150,000
100,000
50,000
0
Jan '22
Feb '22
Mar

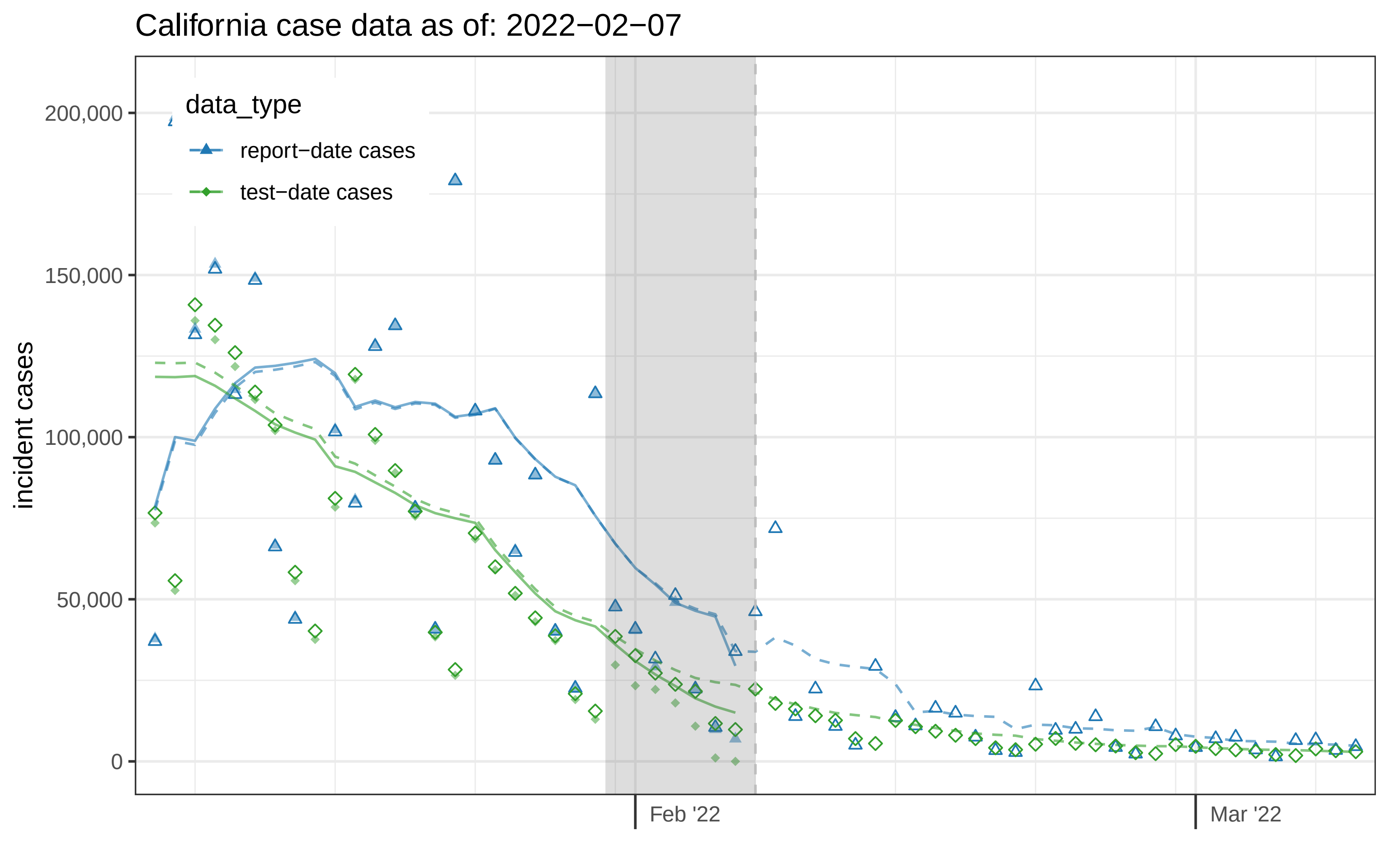

California case data as of: 2022−02−07
data_type
report−date cases
test−date cases
incident cases
200,000
150,000
100,000
50,000
0
Feb '22
Mar '22

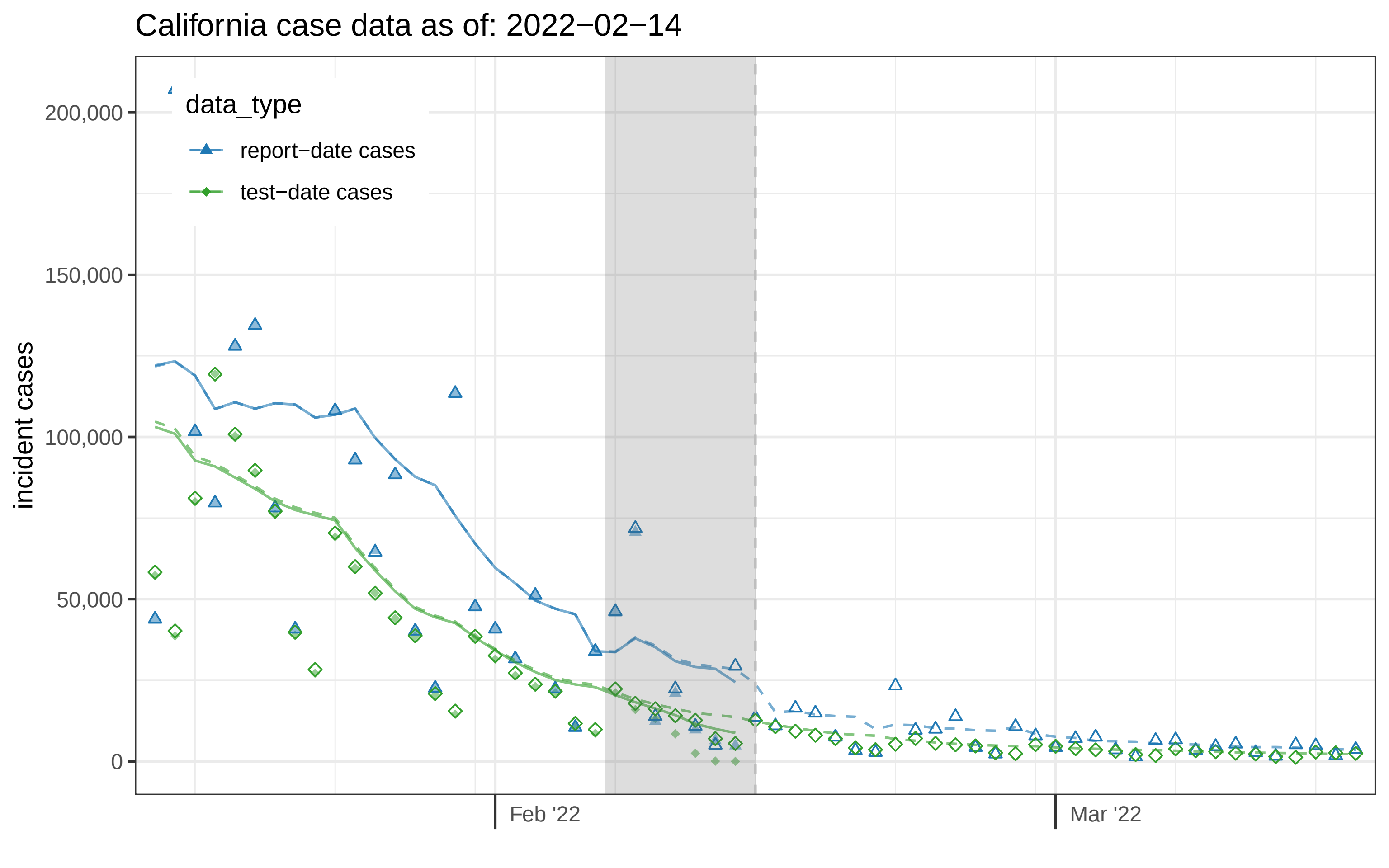
California case data as of: 2022−02−14
data_type
report−date cases
test−date cases
incident cases
200,000
150,000
100,000
50,000
0
Feb '22
Mar '22

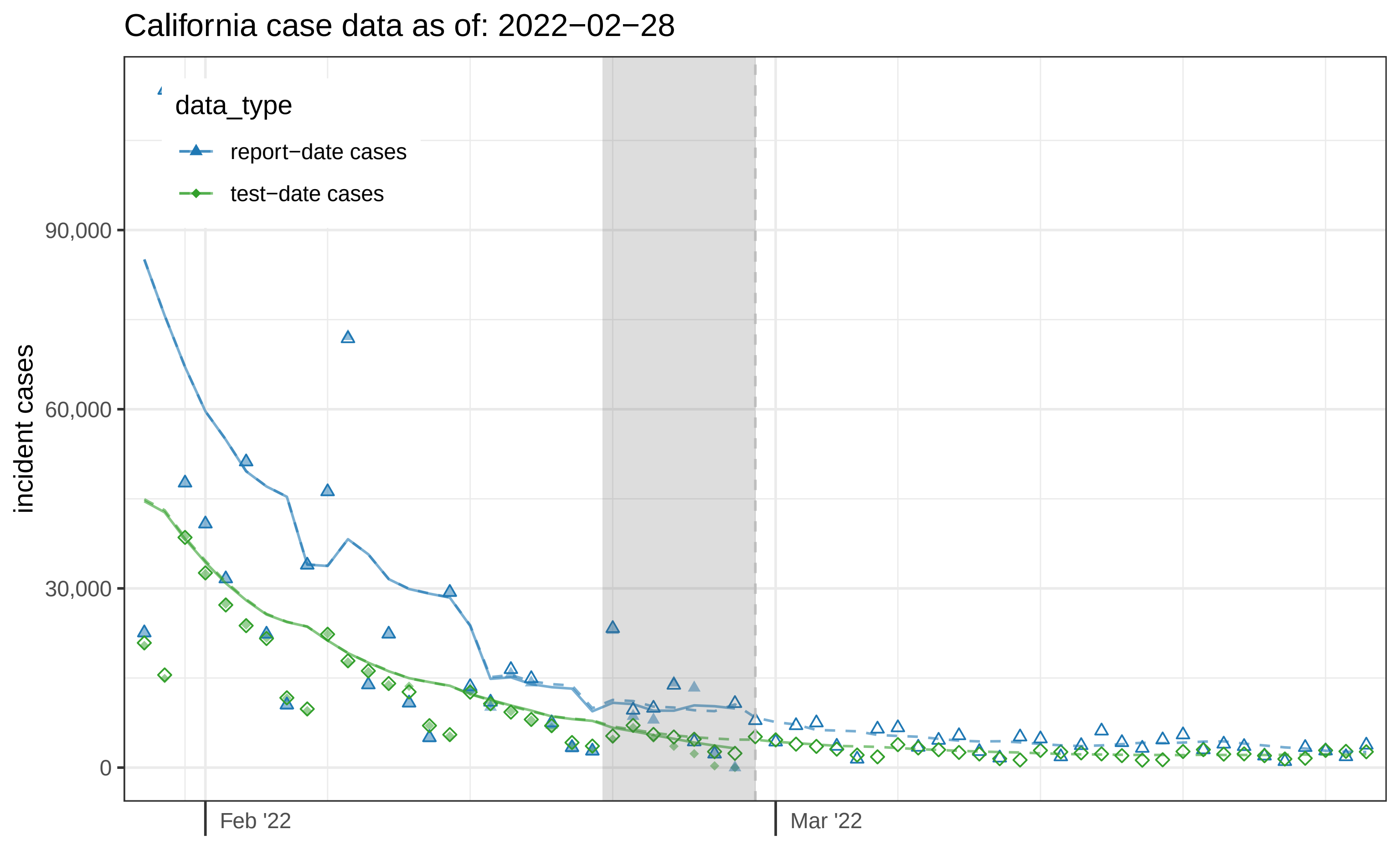
California case data as of: 2022−02−28
data_type
report−date cases
test−date cases
incident cases
90,000
60,000
30,000
0
Feb '22
Mar '22

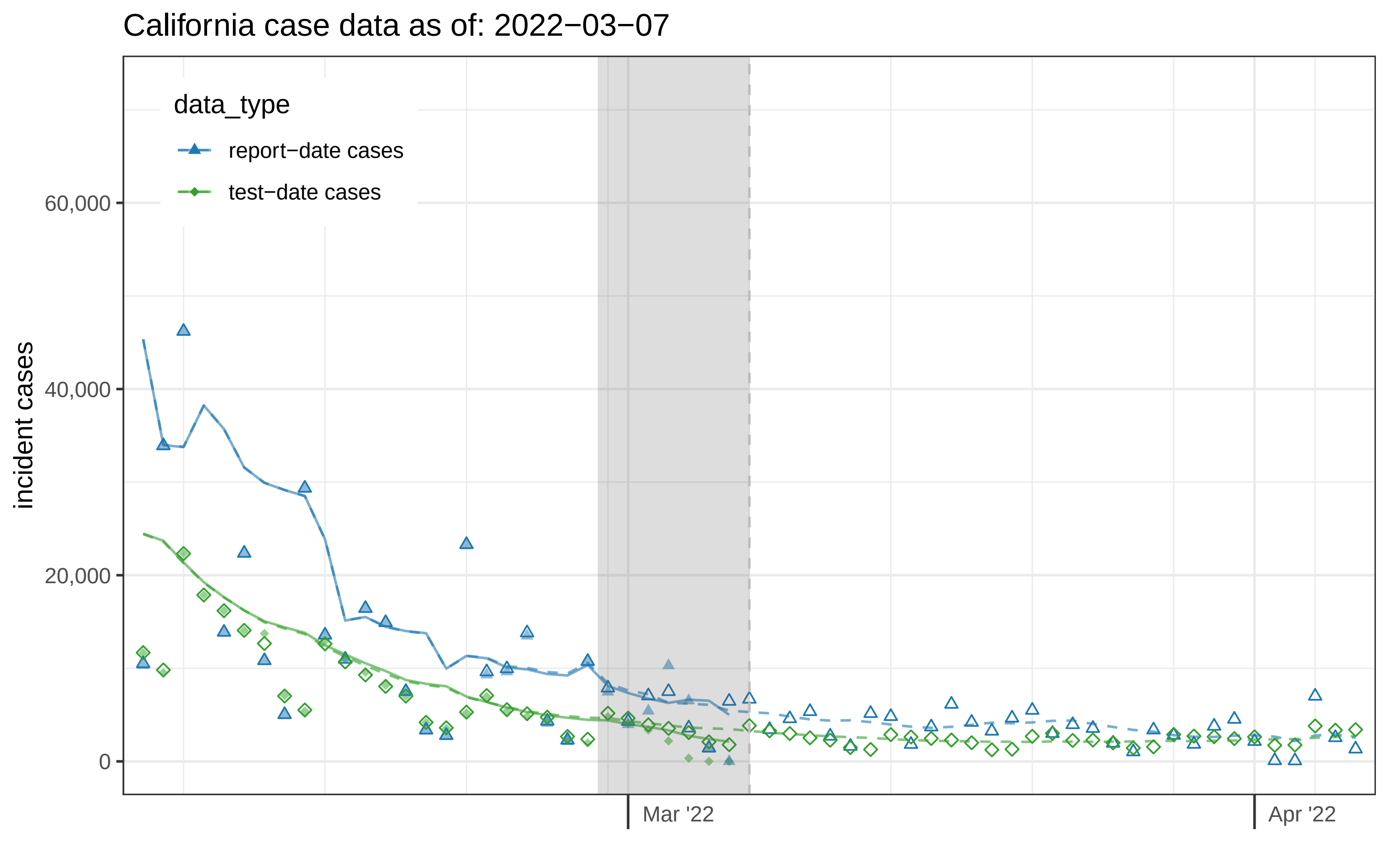
California case data as of: 2022−03−07
data_type
report−date cases
test−date cases
incident cases
60,000
40,000
20,000
0
Mar '22
Apr '22

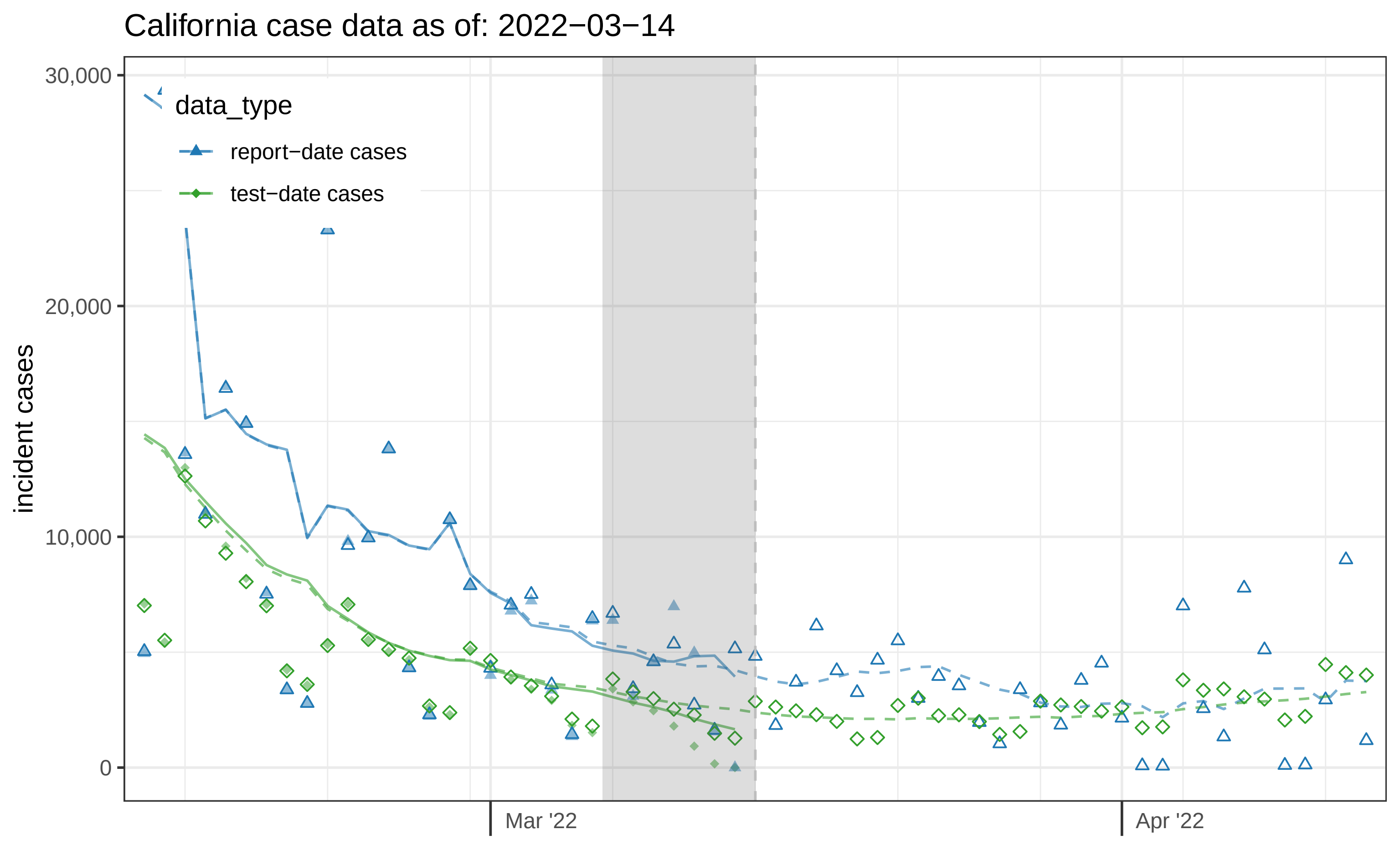

California case data as of: 2022−03−14
data_type
report−date cases
test−date cases
incident cases
30,000
20,000
10,000
0
Mar '22
Apr '22

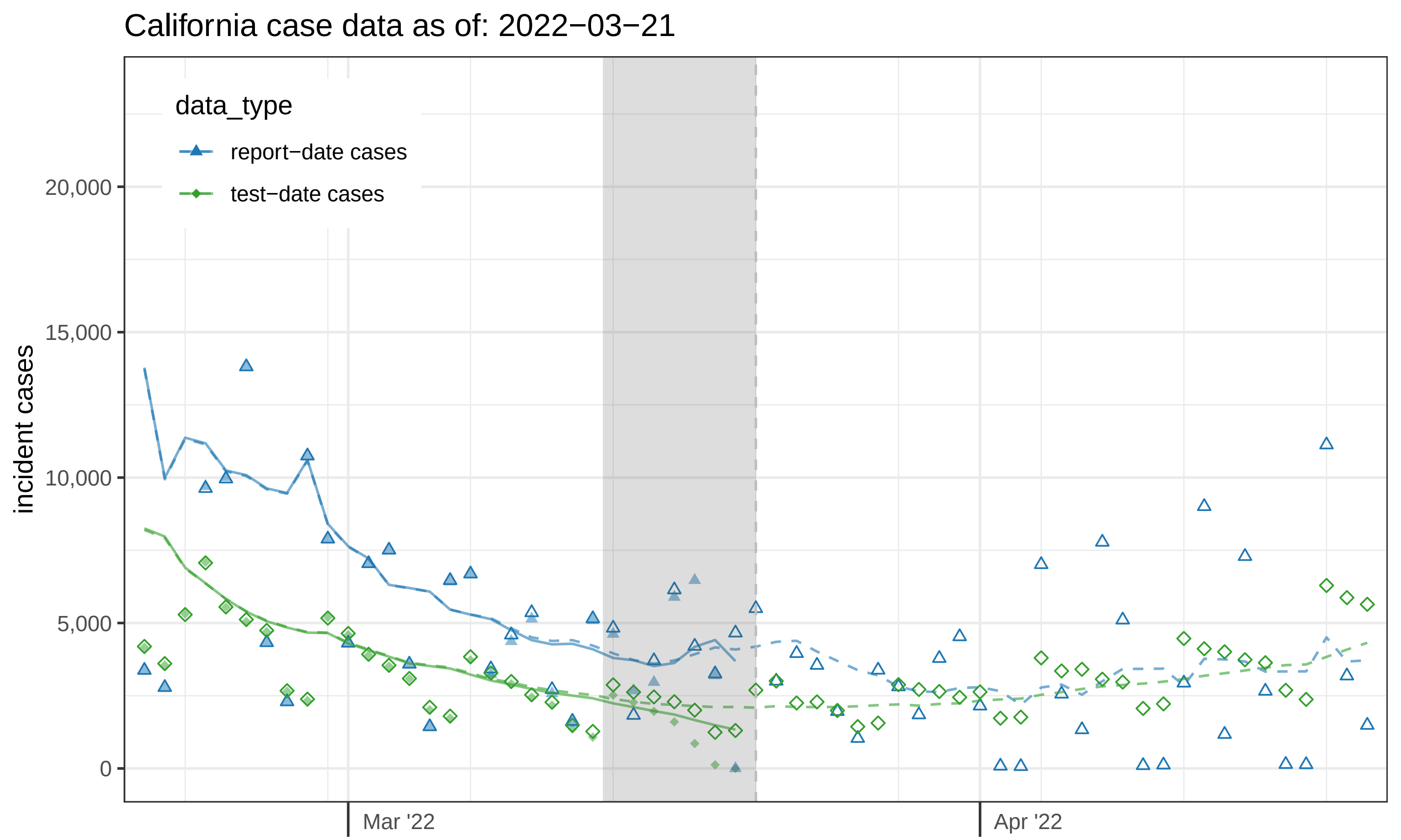

California case data as of: 2022−03−21
data_type
report−date cases
test−date cases
incident cases
20,000
15,000
10,000
5,000
0
Mar '22
Apr '22

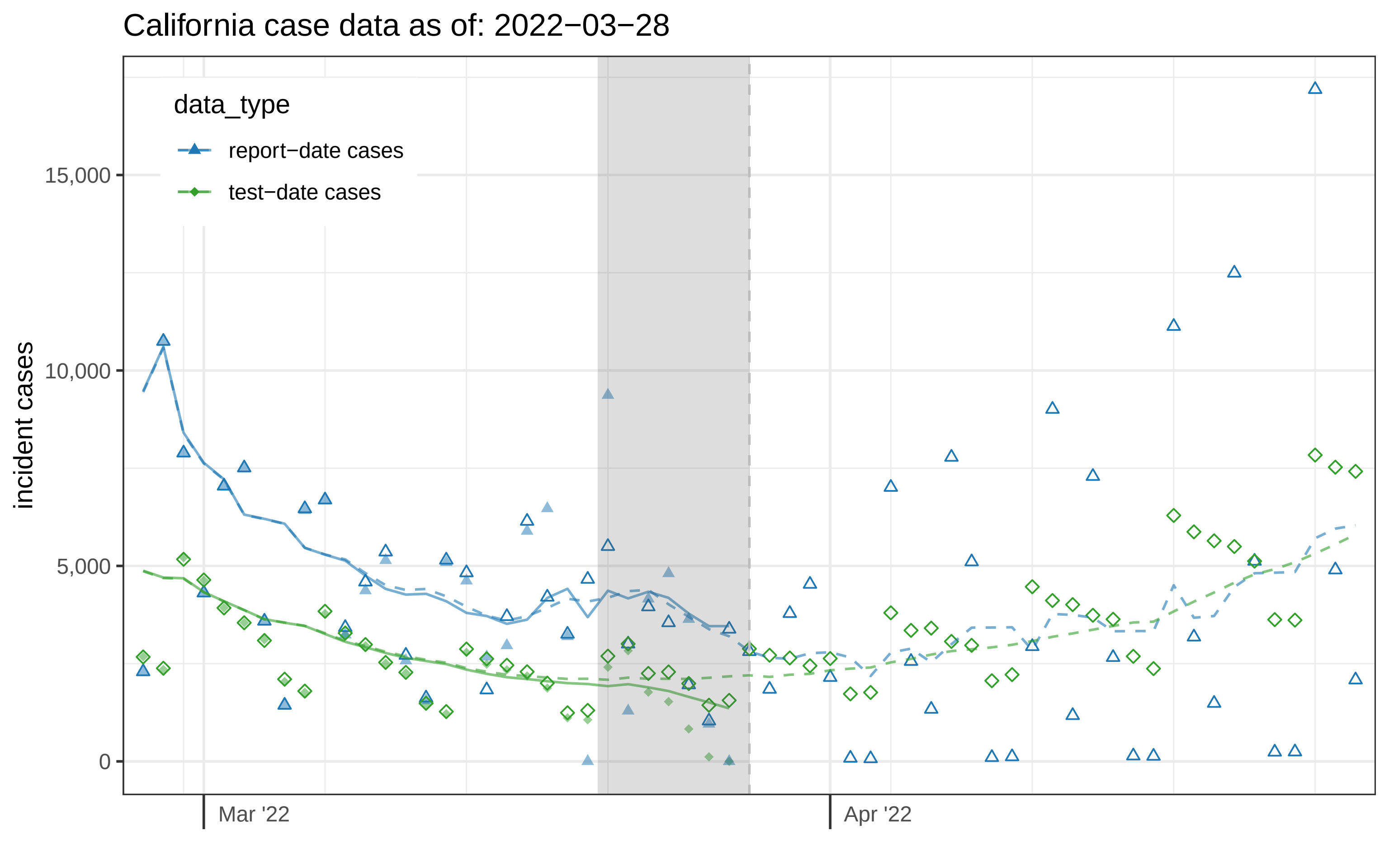

California case data as of: 2022−03−28
data_type
report−date cases
test−date cases
incident cases
15,000
10,000
5,000
0
Mar '22
Apr '22

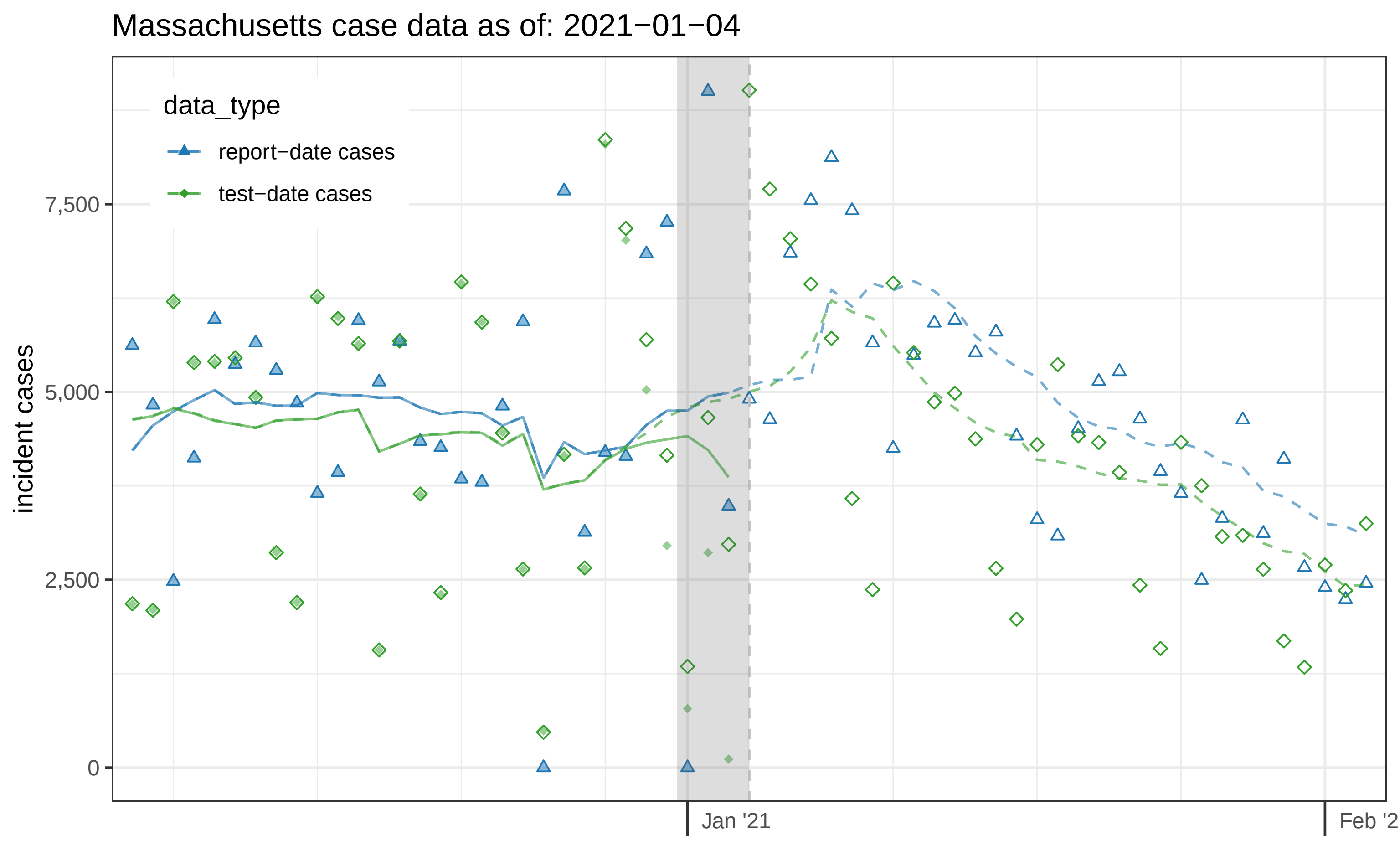
Massachusetts case data as of: 2021−01−04
data_type
report−date cases
test−date cases
incident cases
7,500
5,000
2,500
0
Jan '21
Feb '2

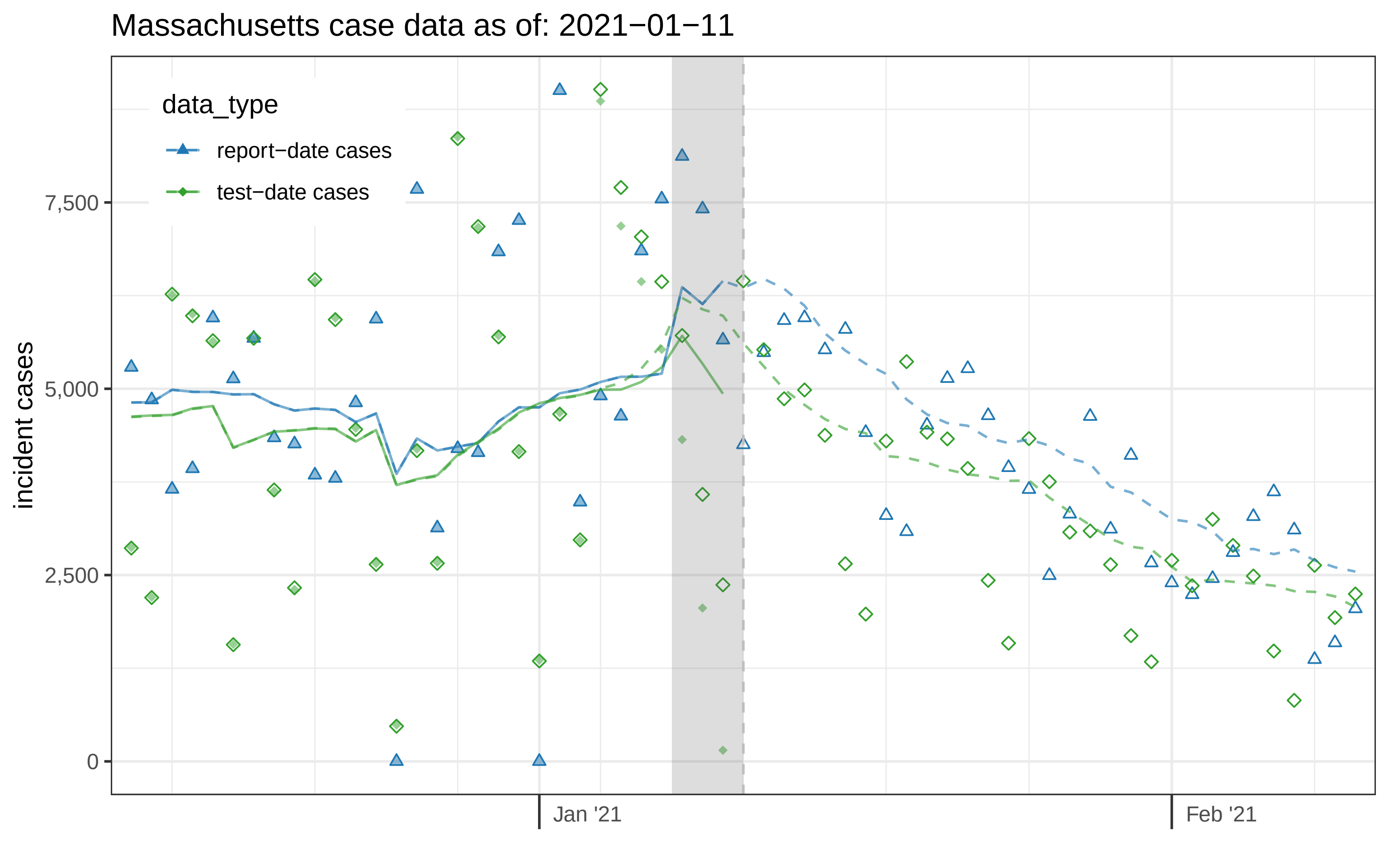

Massachusetts case data as of: 2021−01−11
data_type
report−date cases
test−date cases
incident cases
7,500
5,000
2,500
0
Jan '21
Feb '21

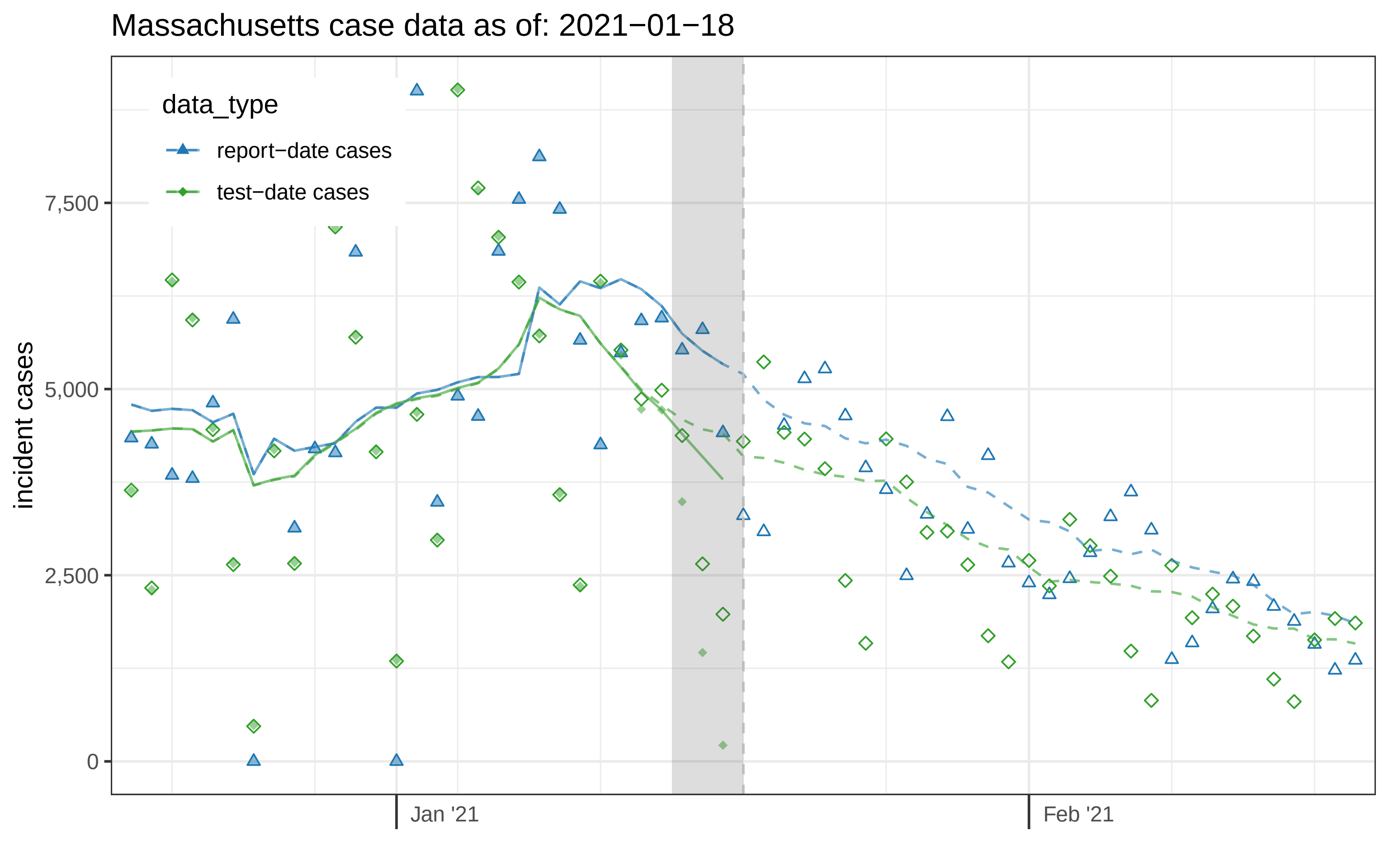

Massachusetts case data as of: 2021−01−18
data_type
report−date cases
test−date cases
incident cases
7,500
5,000
2,500
0
Jan '21
Feb '21

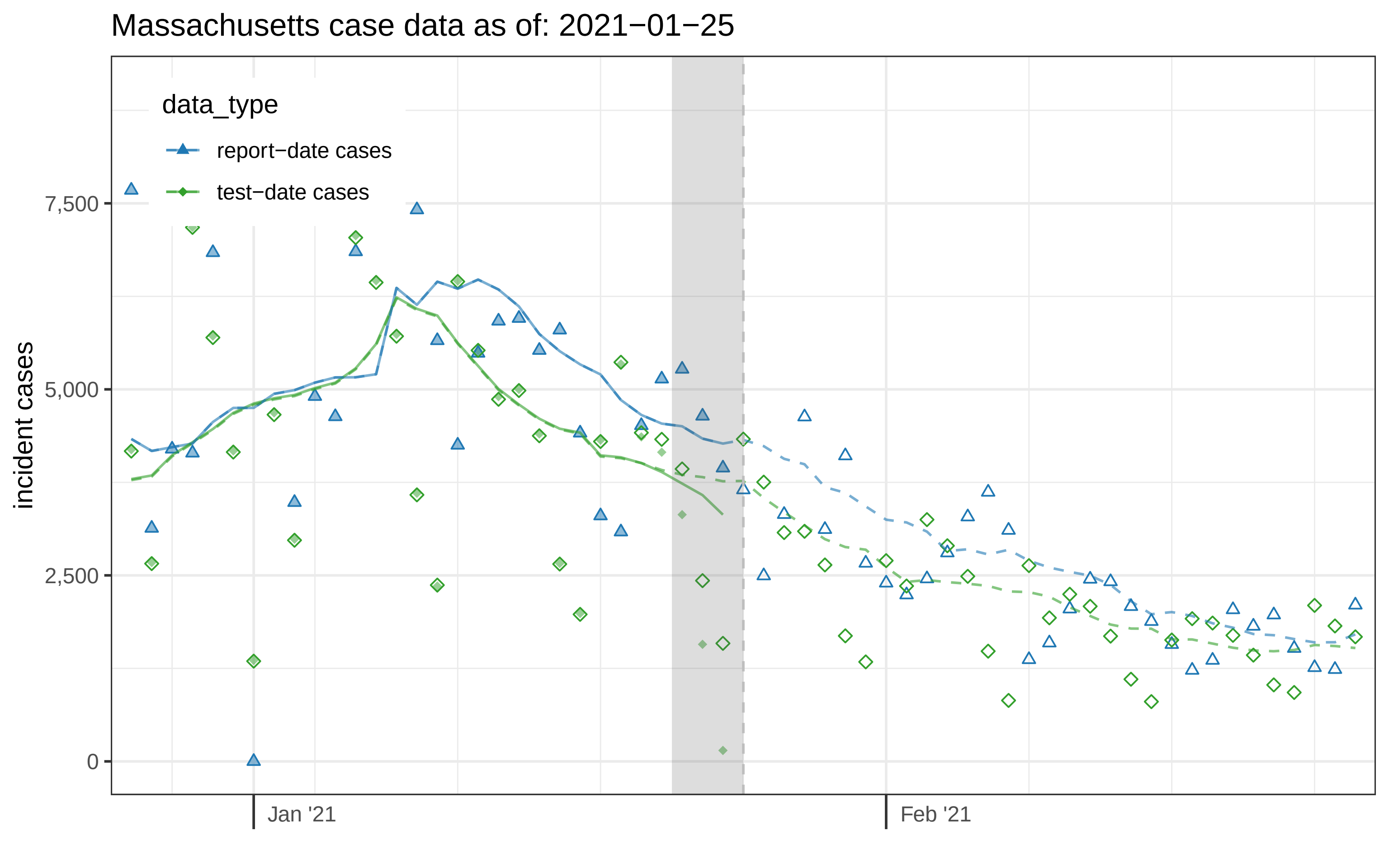

Massachusetts case data as of: 2021−01−25
data_type
report−date cases
test−date cases
incident cases
7,500
5,000
2,500
0
Jan '21
Feb '21

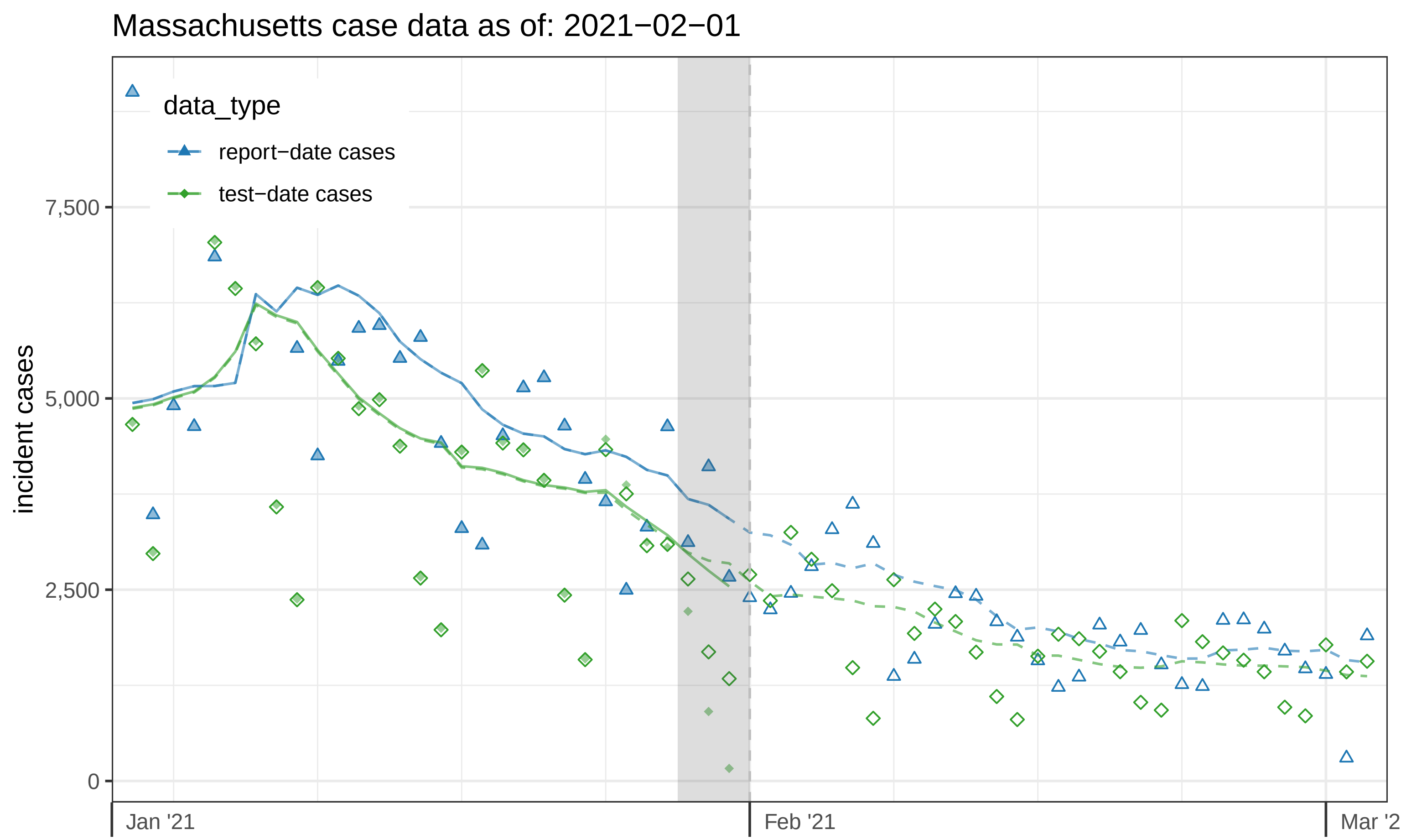

Massachusetts case data as of: 2021−02−01
data_type
report−date cases
test−date cases
incident cases
7,500
5,000
2,500
0
Jan '21
Feb '21
Mar '2

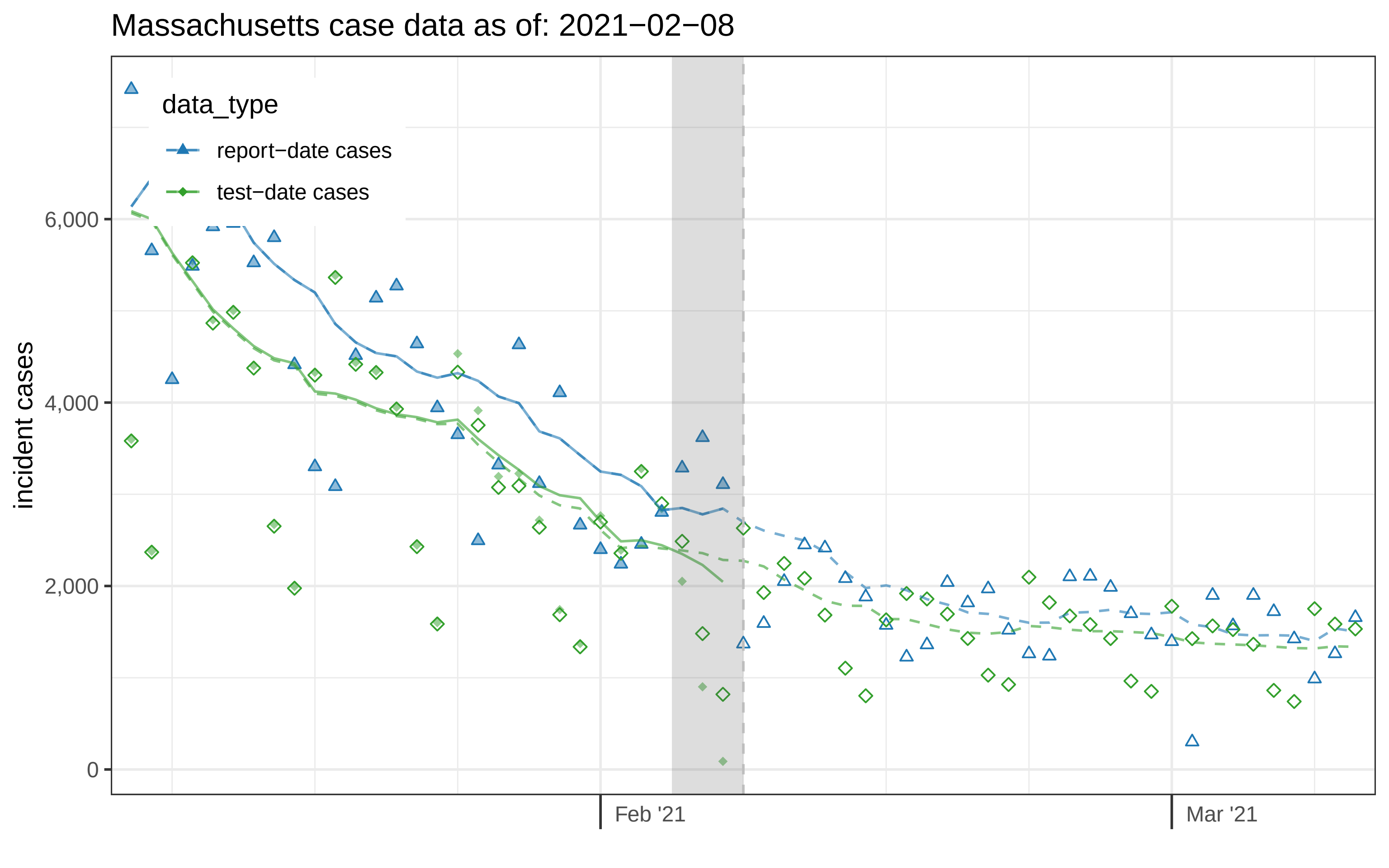

Massachusetts case data as of: 2021−02−08
data_type
report−date cases
test−date cases
incident cases
6,000
4,000
2,000
0
Feb '21
Mar '21

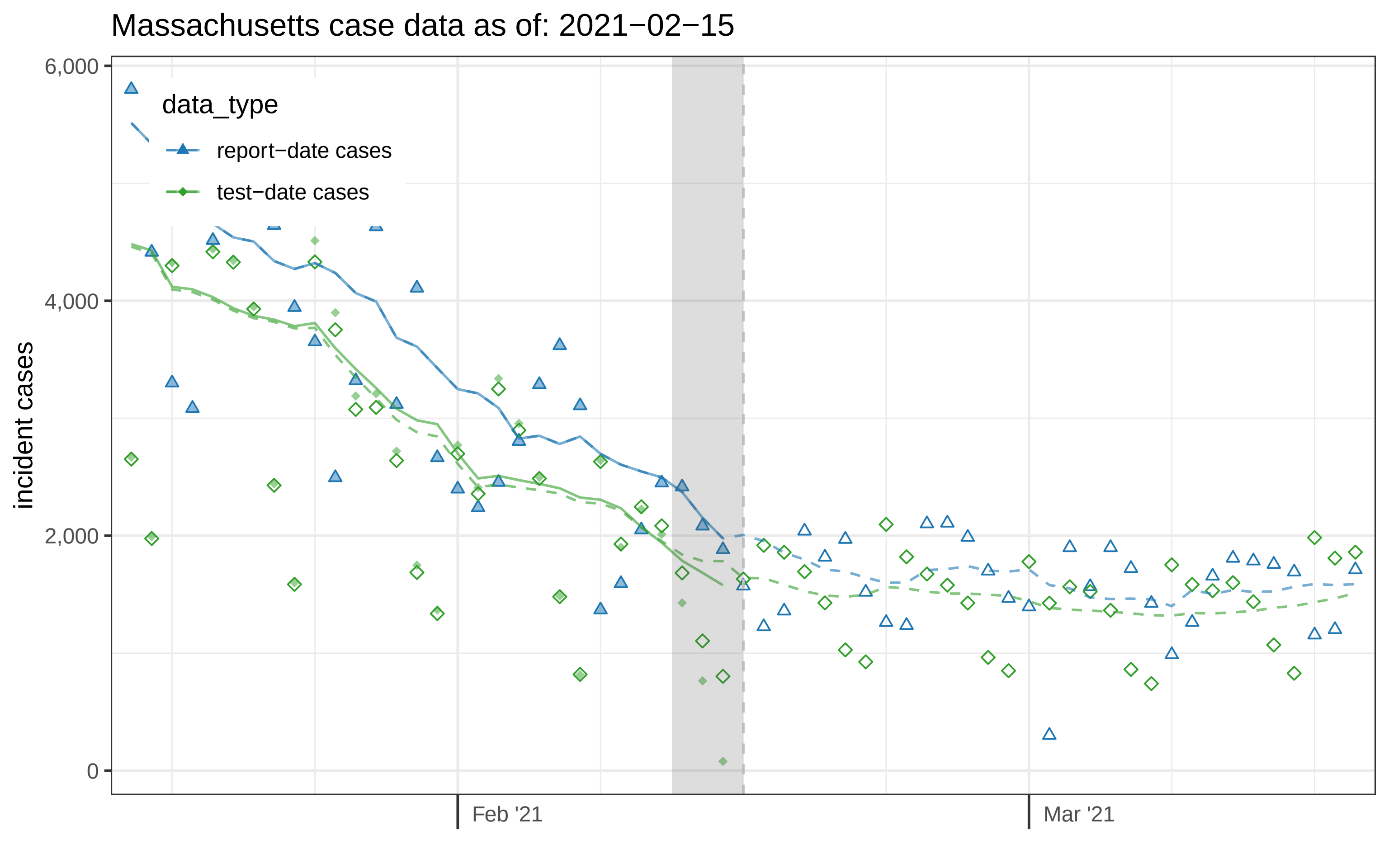
Massachusetts case data as of: 2021−02−15
6,000
4,000
2,000
0
incident cases
data_type
report−date cases
test−date cases
Feb '21
Mar '21

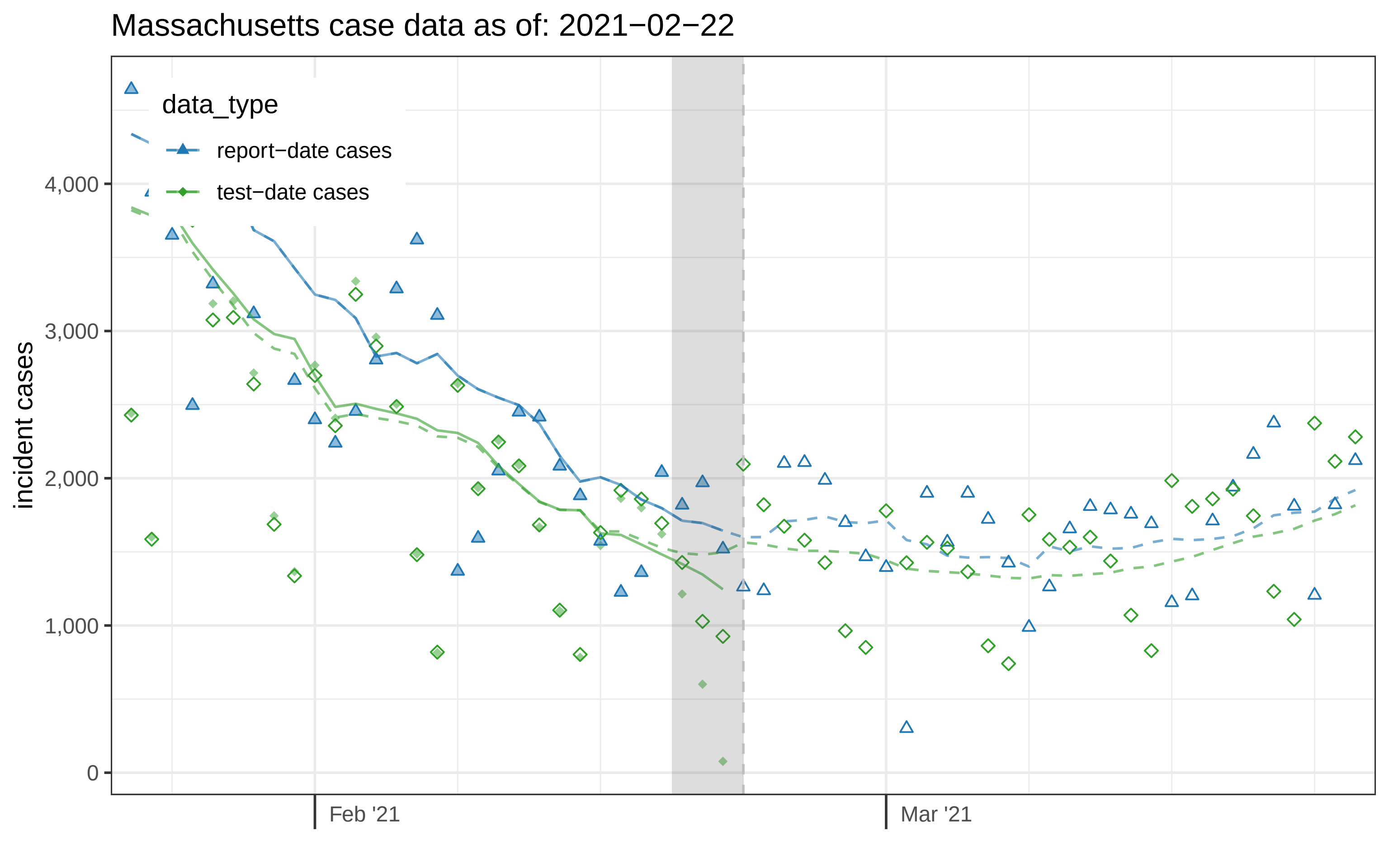

Massachusetts case data as of: 2021−02−22
data_type
report−date cases
test−date cases
incident cases
4,000
3,000
2,000
1,000
0
Feb '21
Mar '21

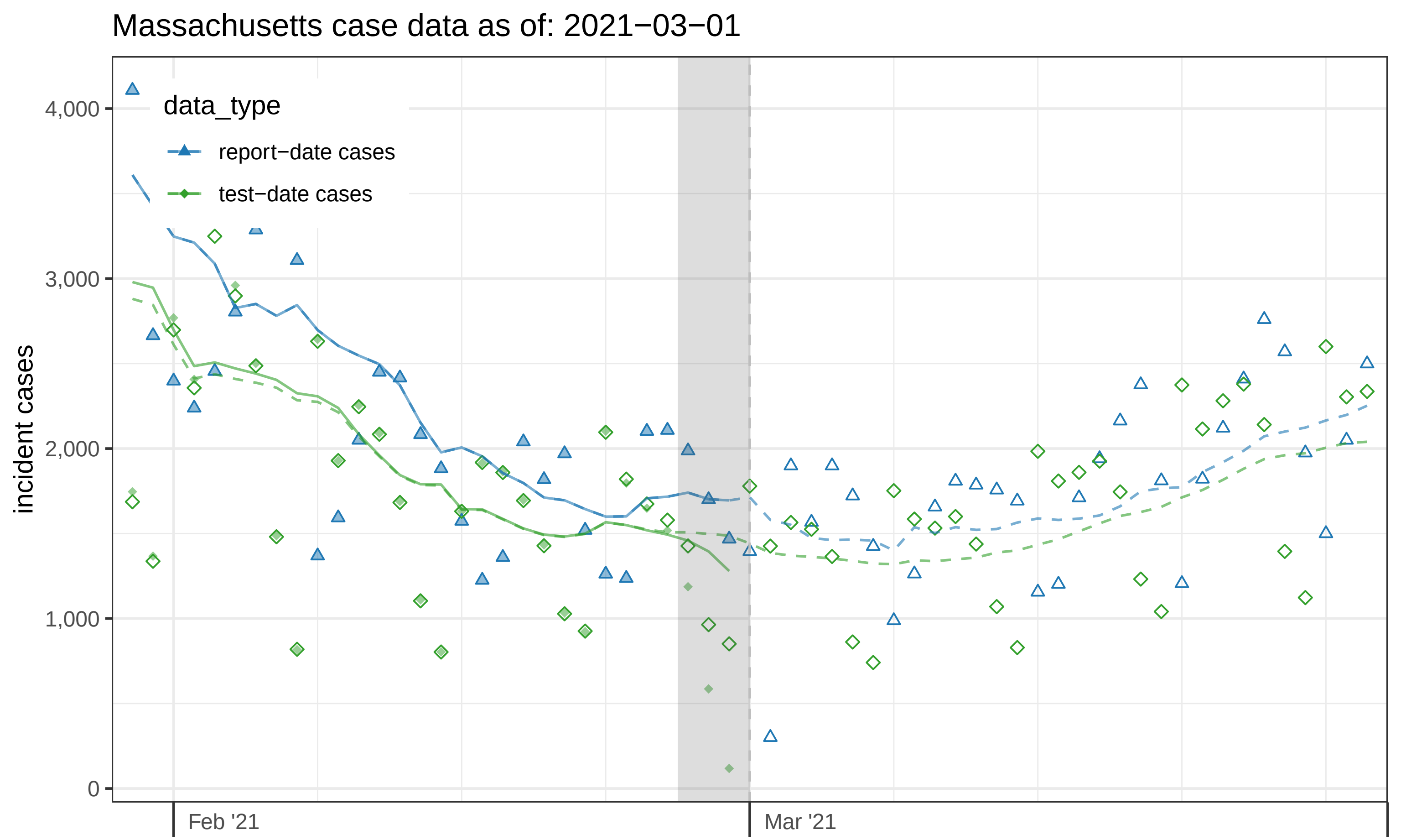
Massachusetts case data as of: 2021−03−01
data_type
report−date cases
test−date cases
incident cases
4,000
3,000
2,000
1,000
0
Feb '21
Mar '21

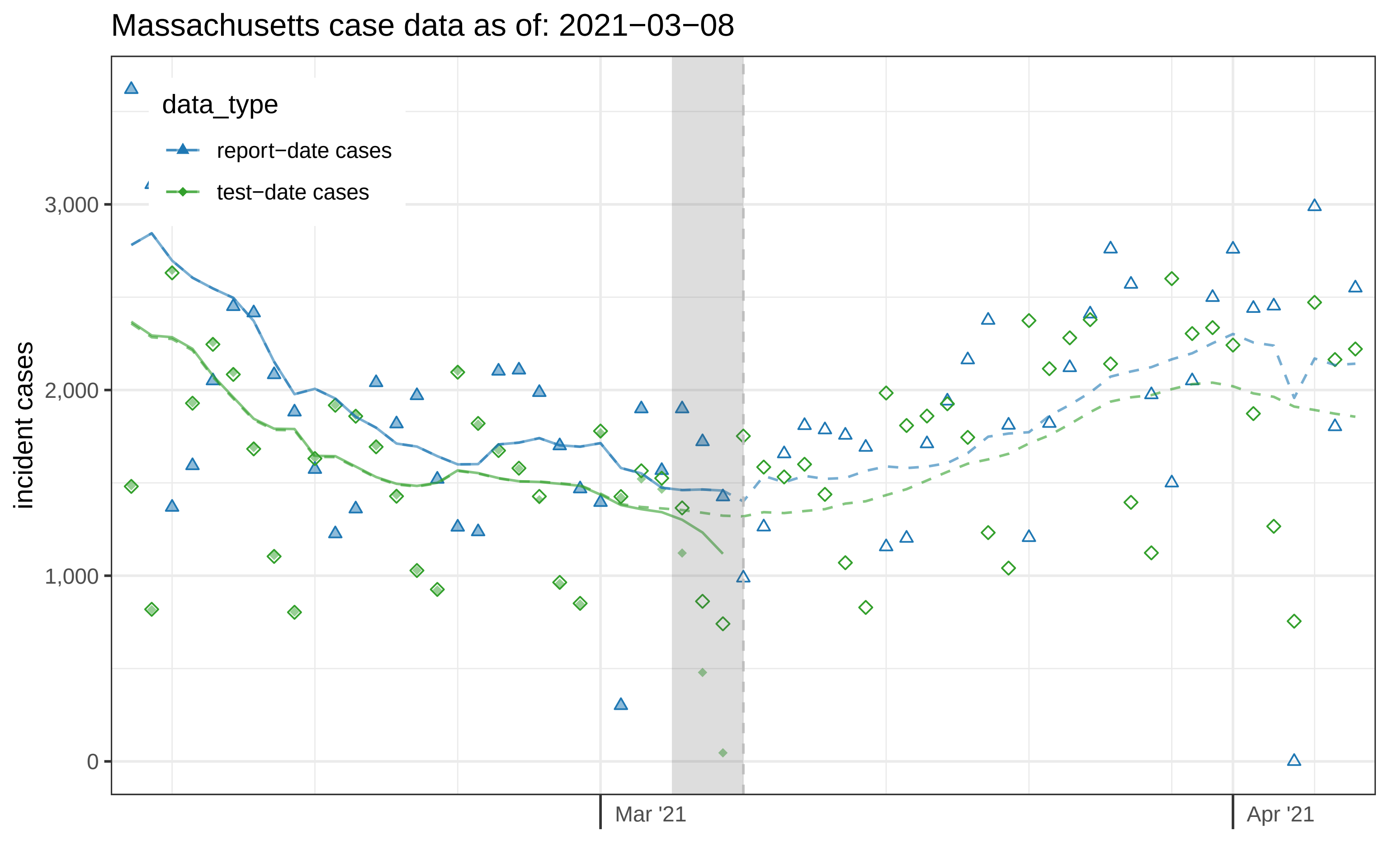
Massachusetts case data as of: 2021−03−08
data_type
report−date cases
test−date cases
incident cases
3,000
2,000
1,000
0
Mar '21
Apr '21

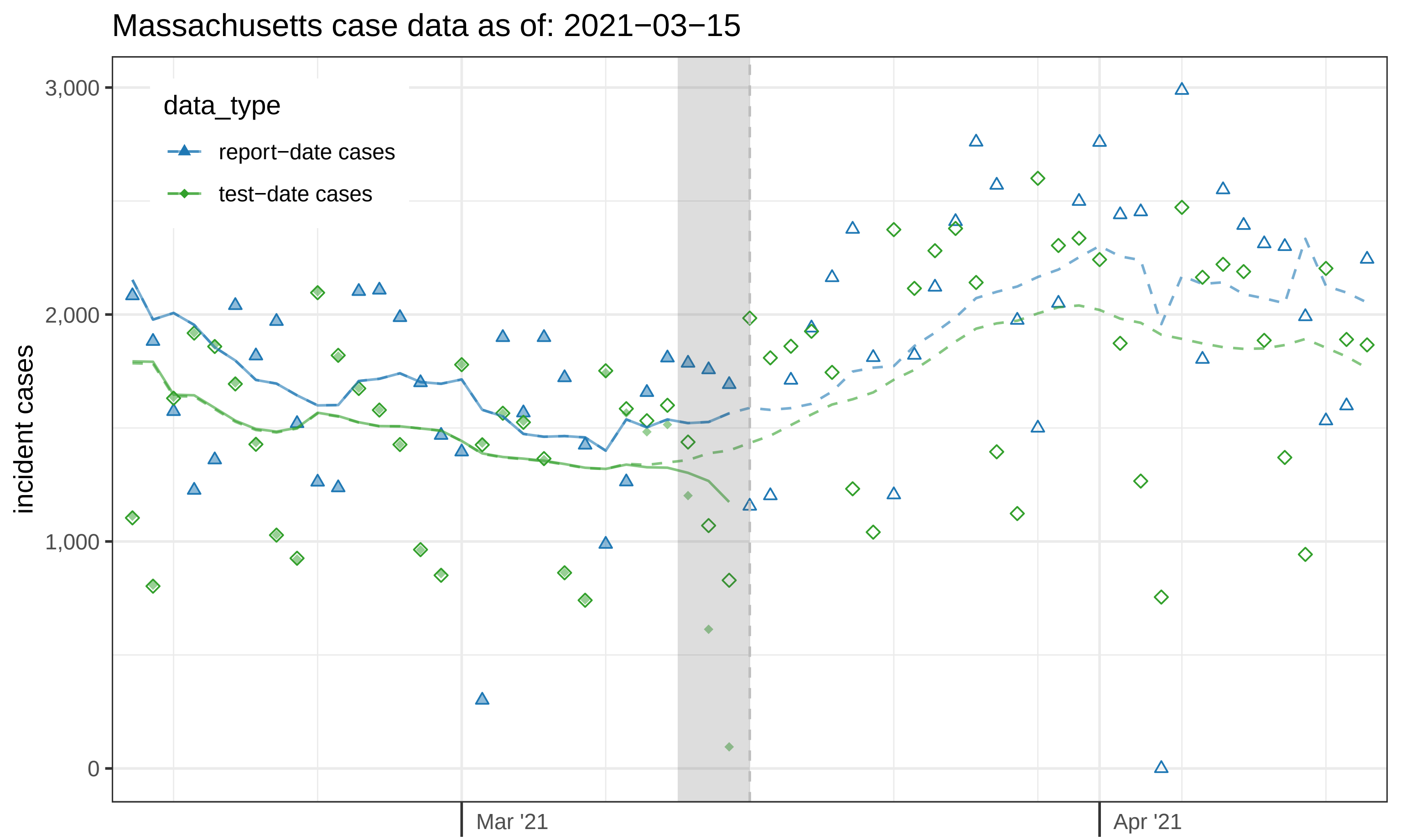
Massachusetts case data as of: 2021−03−15
data_type
report−date cases
test−date cases
incident cases
3,000
2,000
1,000
0
Mar '21
Apr '21

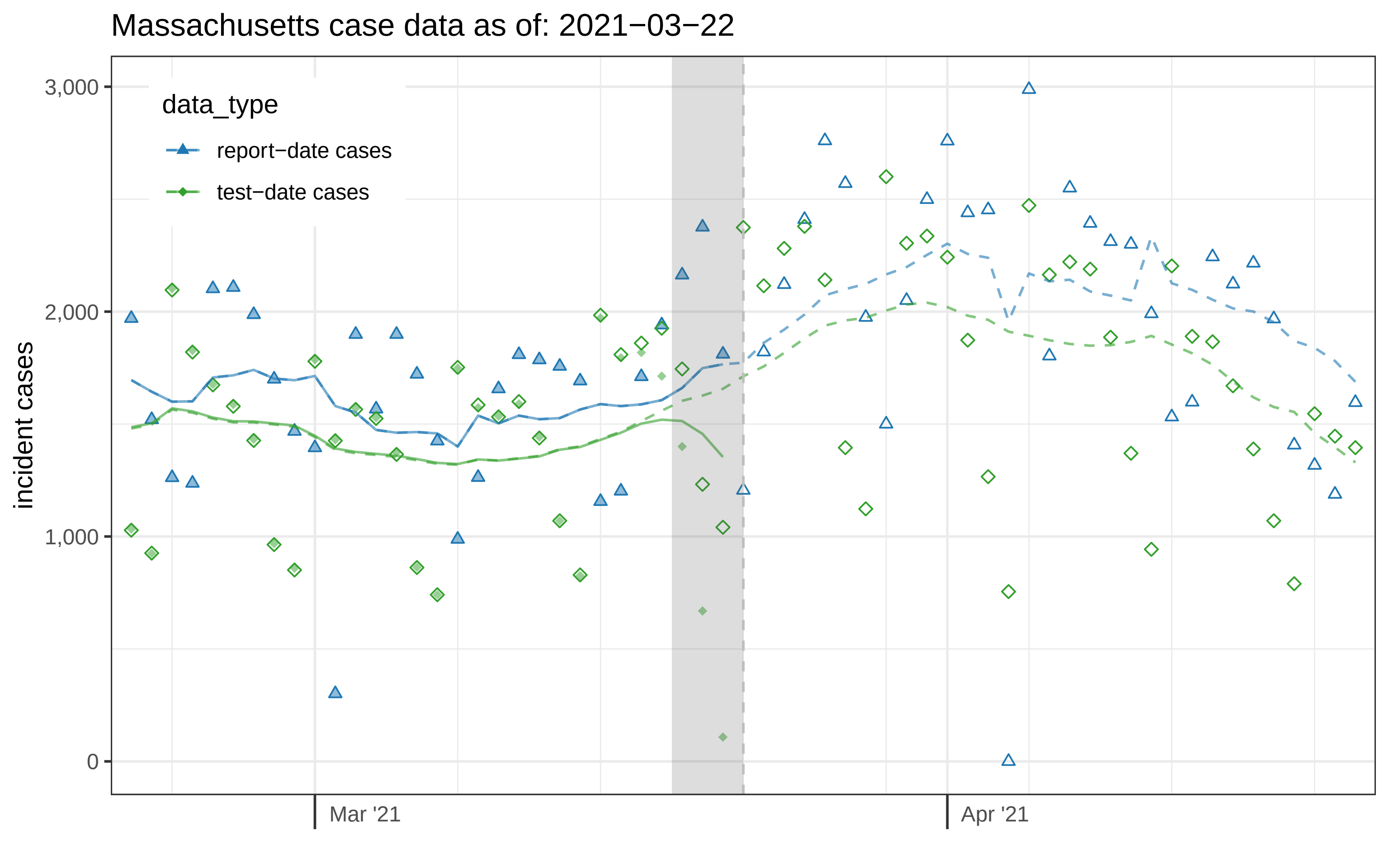

Massachusetts case data as of: 2021−03−22
data_type
report−date cases
test−date cases
incident cases
3,000
2,000
1,000
0
Mar '21
Apr '21

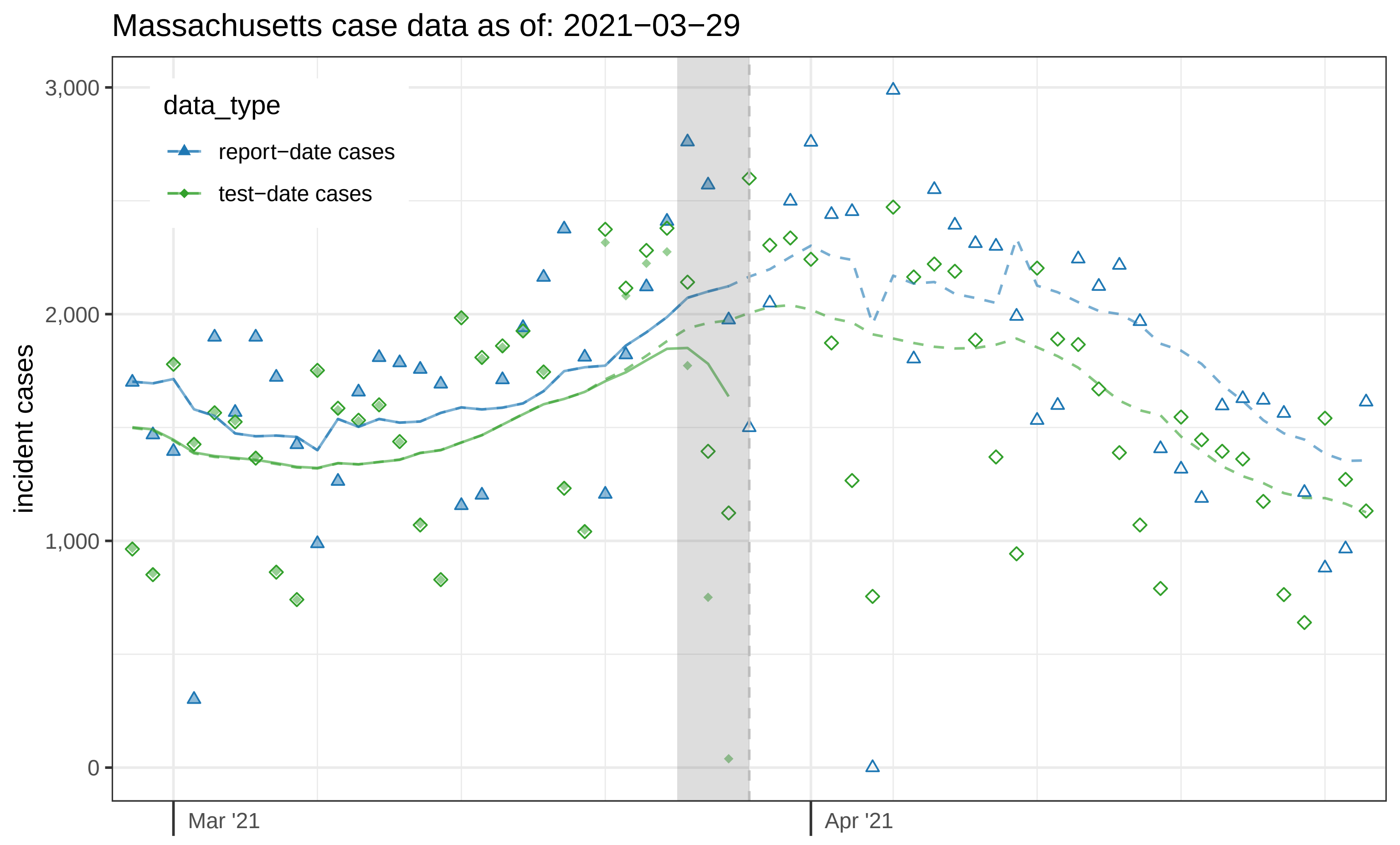

Massachusetts case data as of: 2021−03−29
data_type
report−date cases
test−date cases
incident cases
3,000
2,000
1,000
0
Mar '21
Apr '21

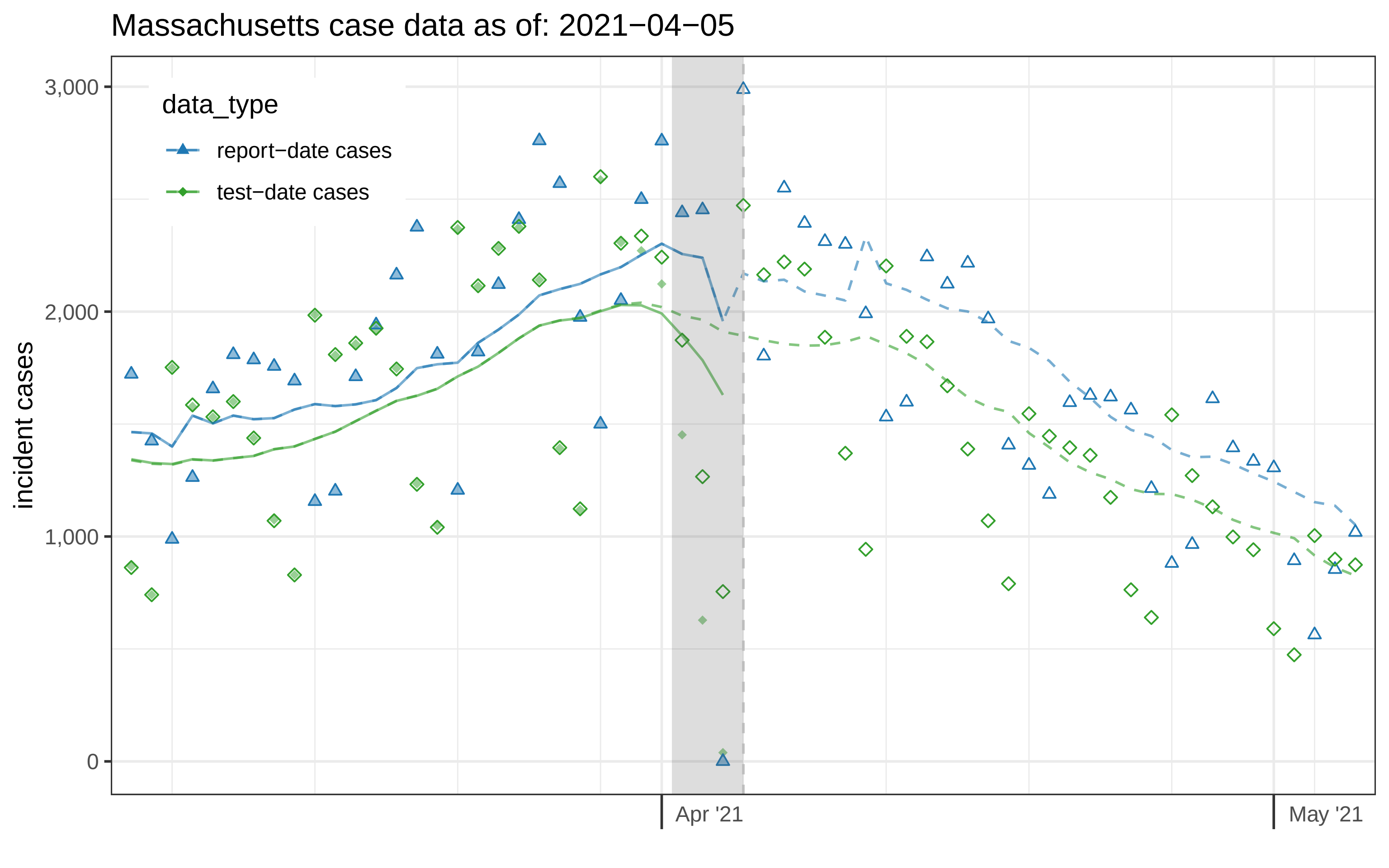

Massachusetts case data as of: 2021−04−05
data_type
report−date cases
test−date cases
incident cases
3,000
2,000
1,000
0
Apr '21
May '21

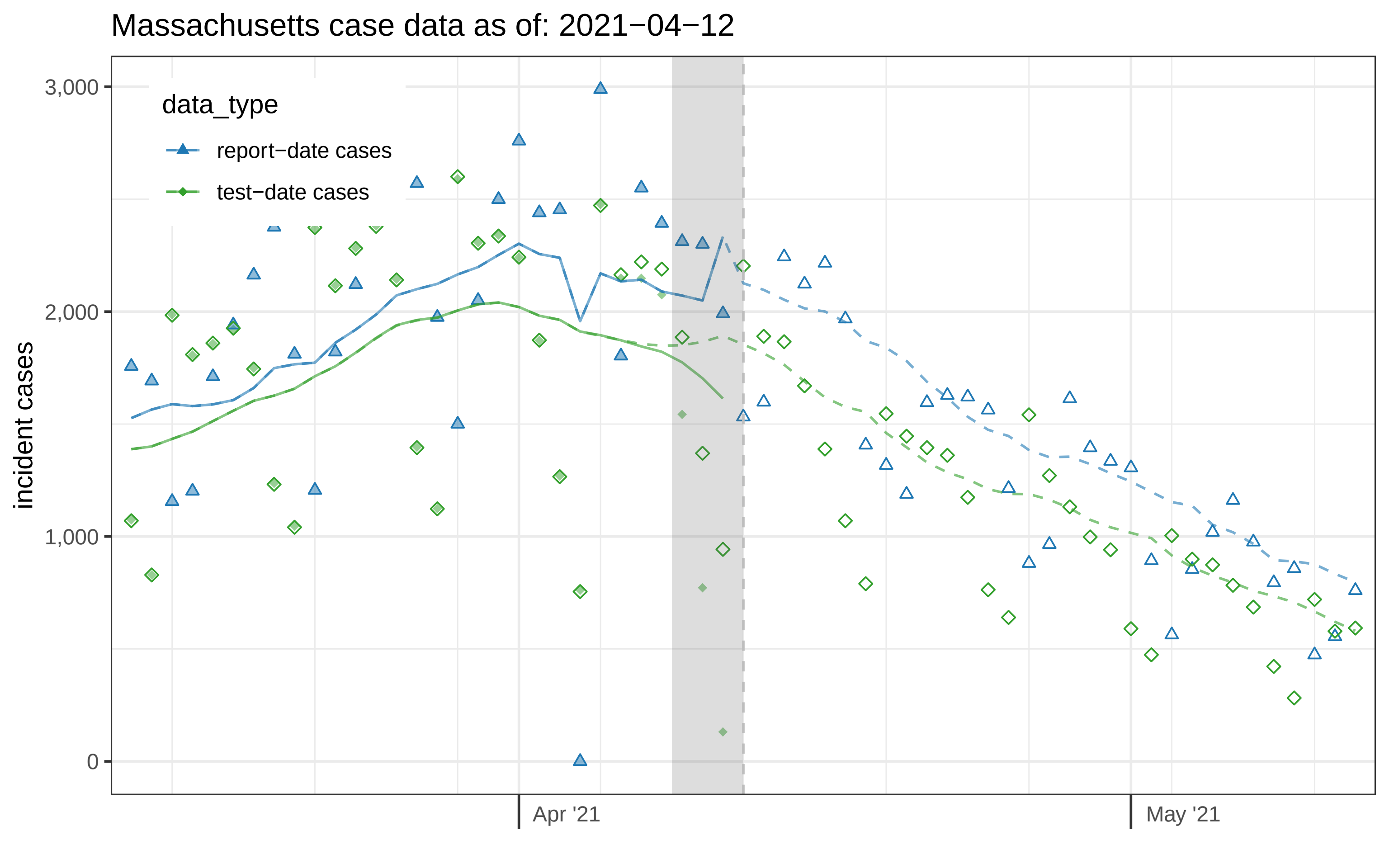

Massachusetts case data as of: 2021−04−12
data_type
report−date cases
test−date cases
incident cases
3,000
2,000
1,000
0
Apr '21
May '21

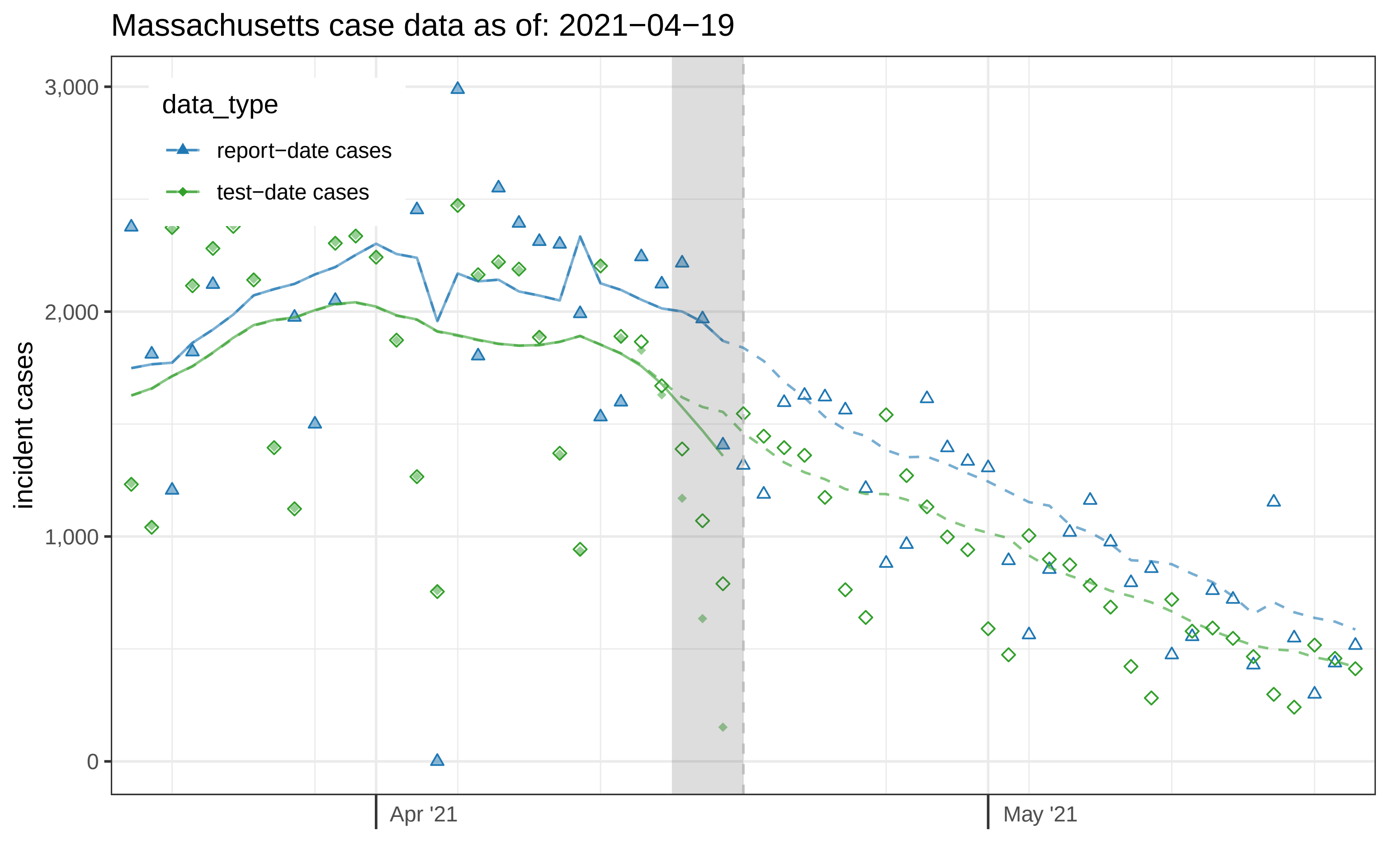

Massachusetts case data as of: 2021−04−19
data_type
report−date cases
test−date cases
incident cases
3,000
2,000
1,000
0
Apr '21
May '21

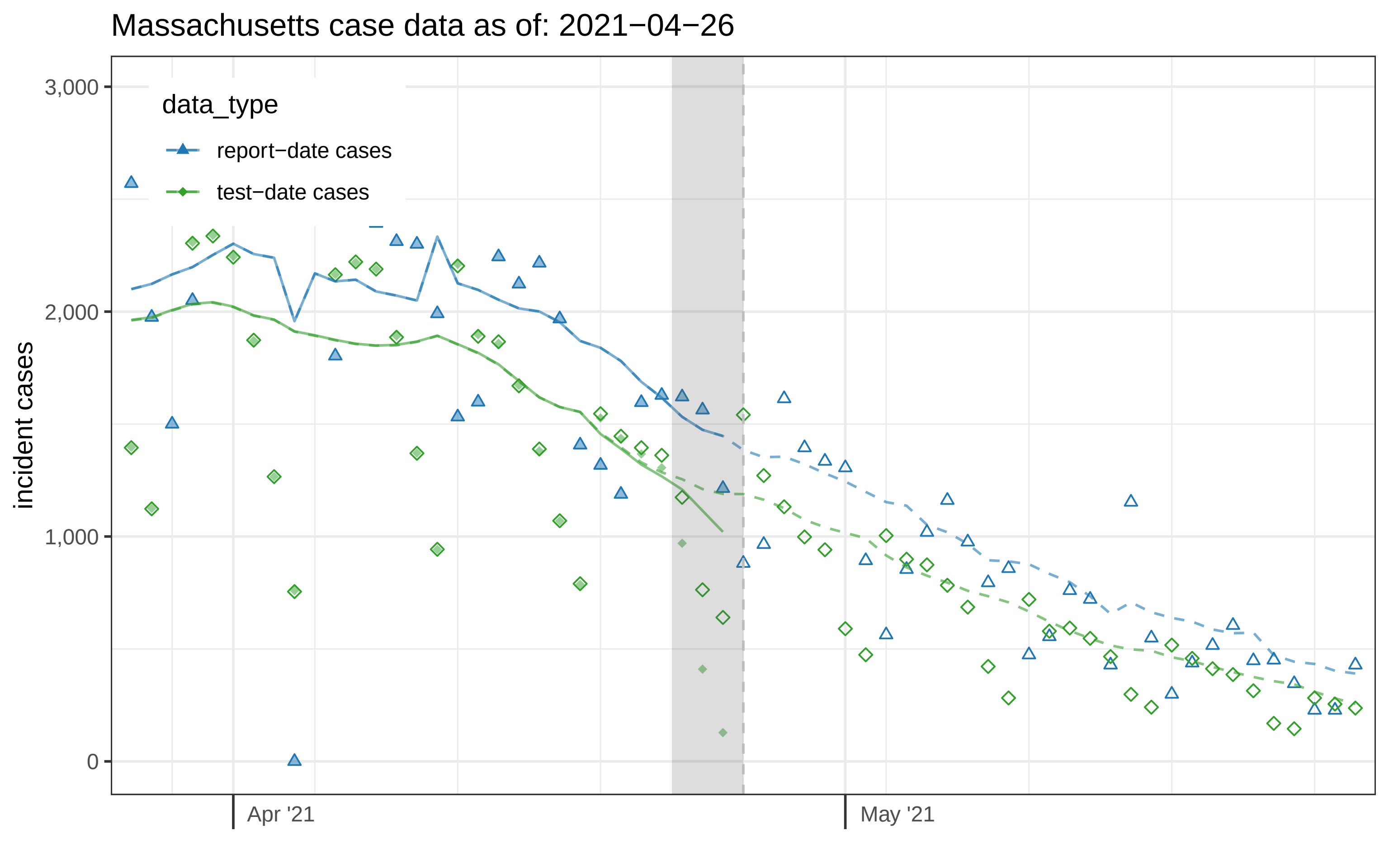

Massachusetts case data as of: 2021−04−26
data_type
report−date cases
test−date cases
incident cases
3,000
2,000
1,000
0
Apr '21
May '21

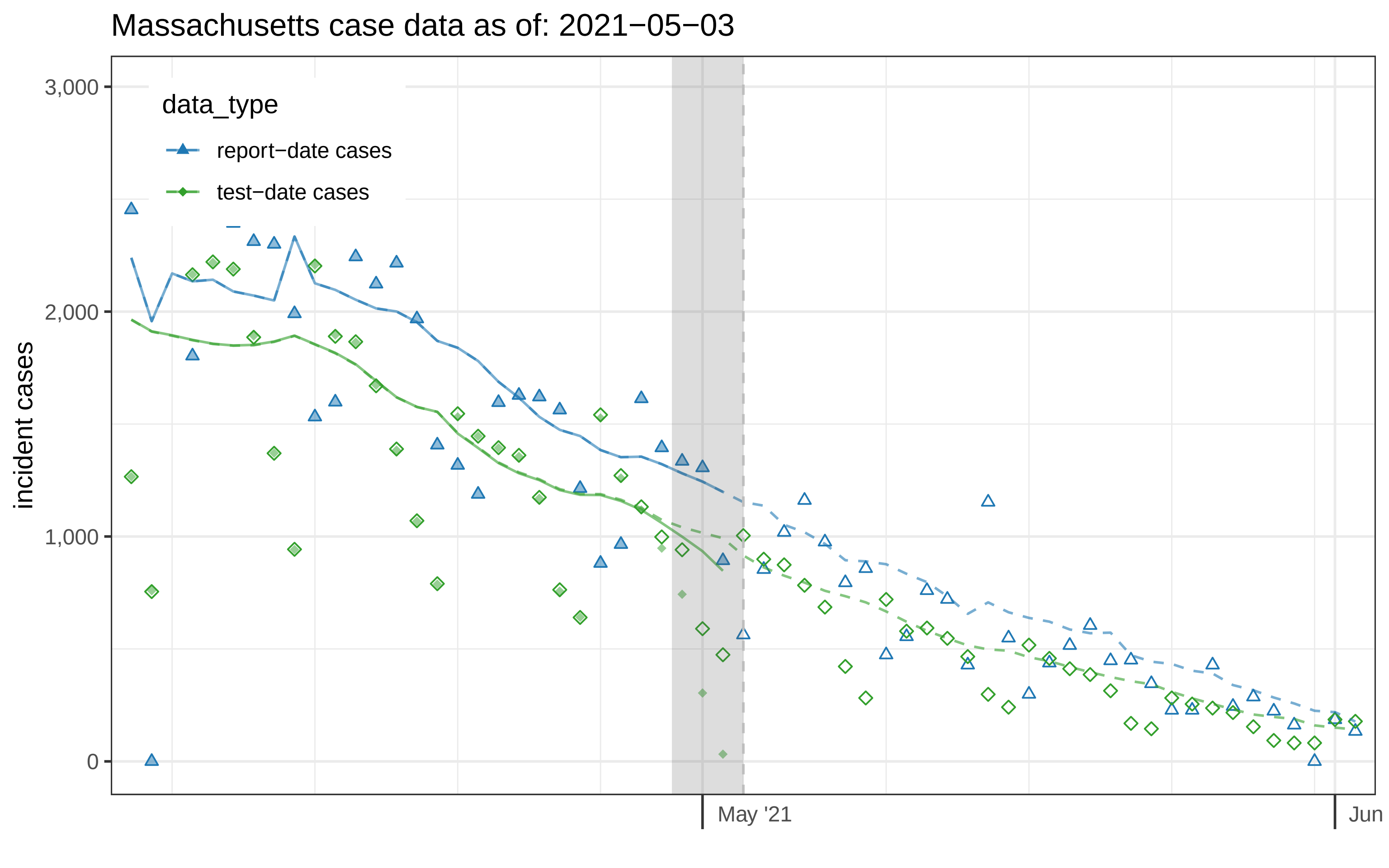
Massachusetts case data as of: 2021−05−03
incident cases
3,000
2,000
1,000
0
May '21
Jun
data_type
report−date cases
test−date cases

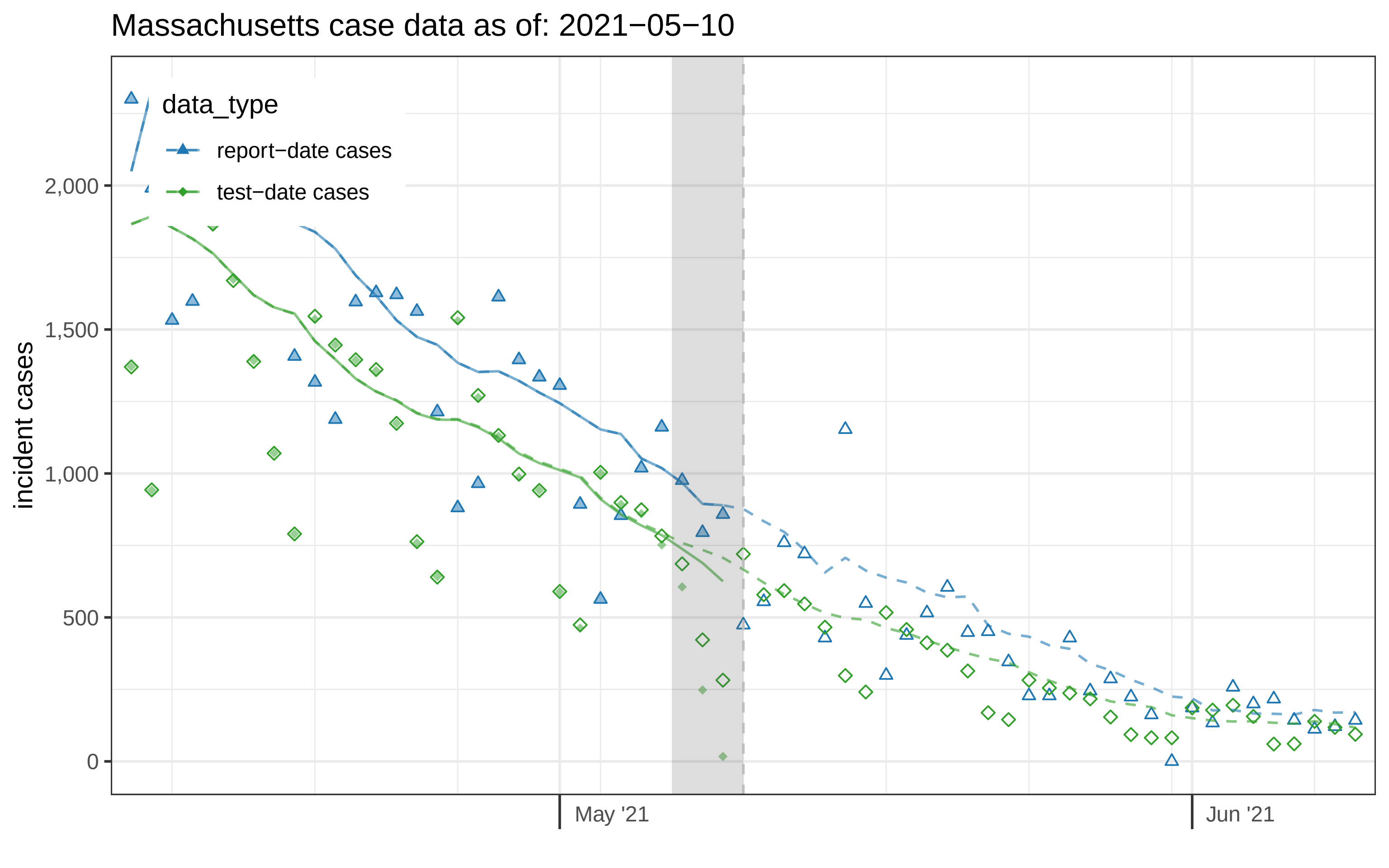
Massachusetts case data as of: 2021−05−10
data_type
report−date cases
test−date cases
incident cases
2,000
1,500
1,000
500
0
May '21
Jun '21

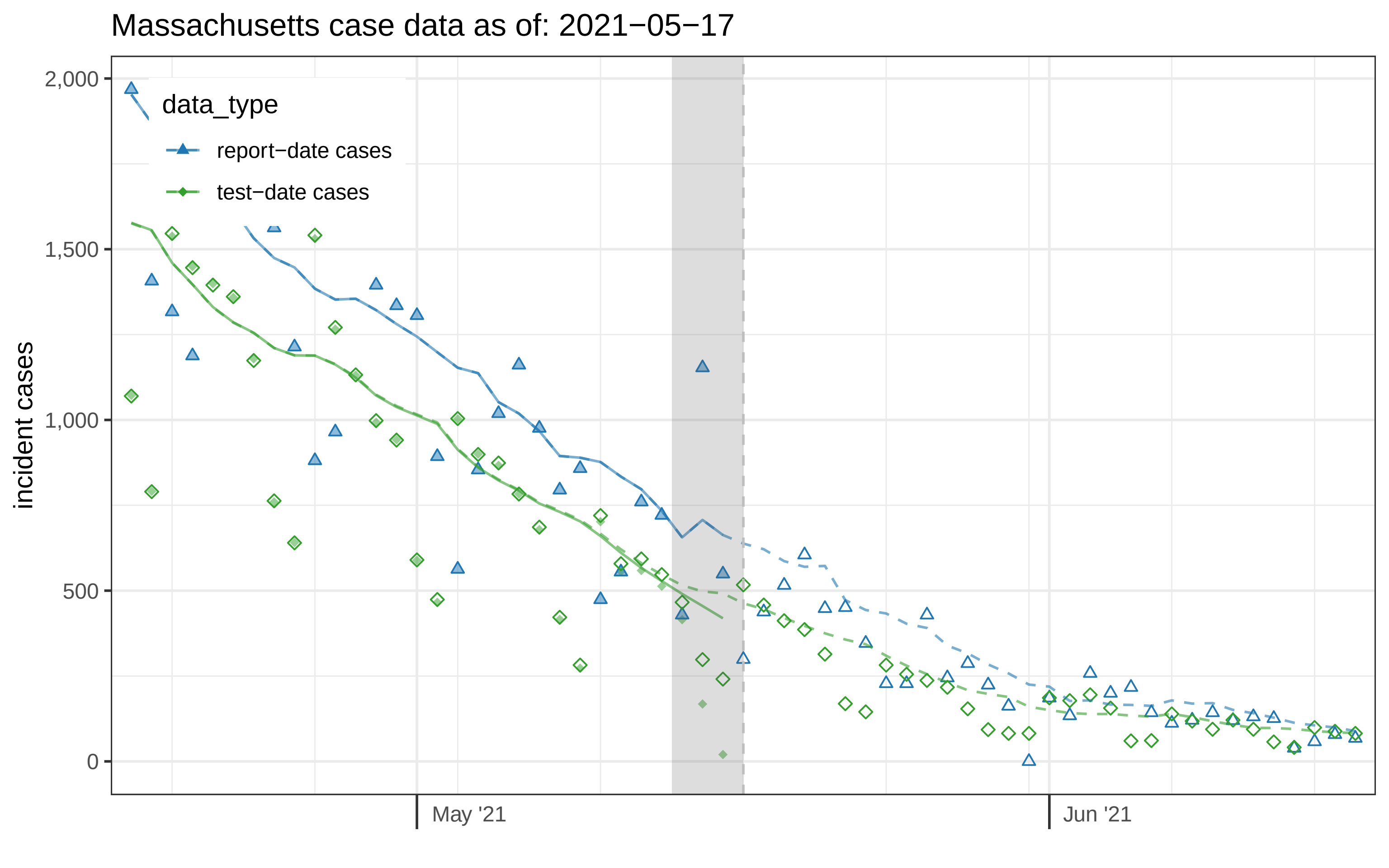

Massachusetts case data as of: 2021−05−17
incident cases
2,000
1,500
1,000
500
0
May '21
Jun '21
data_type
report−date cases
test−date cases

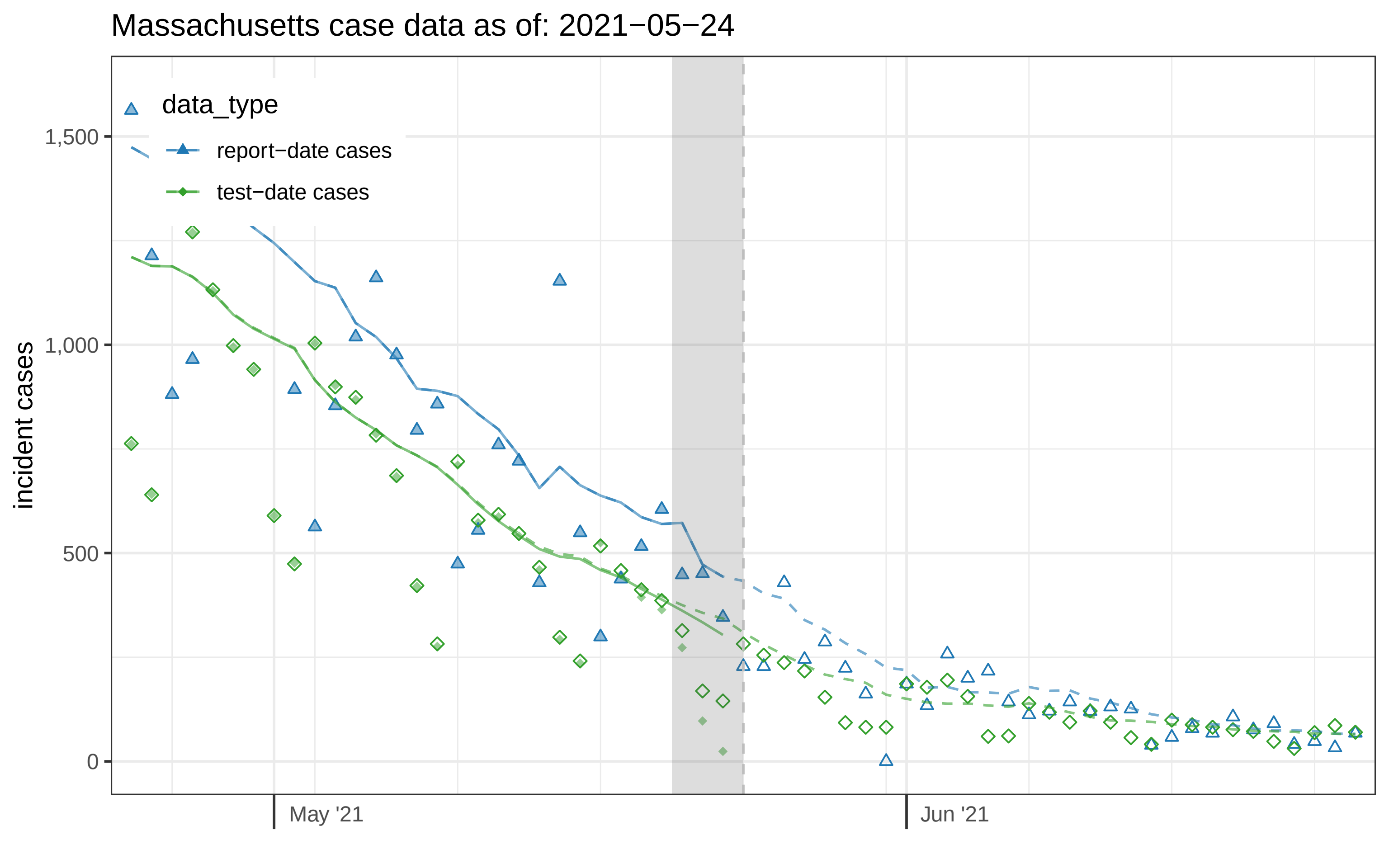
Massachusetts case data as of: 2021−05−24
data_type
report−date cases
test−date cases
incident cases
1,500
1,000
500
0
May '21
Jun '21

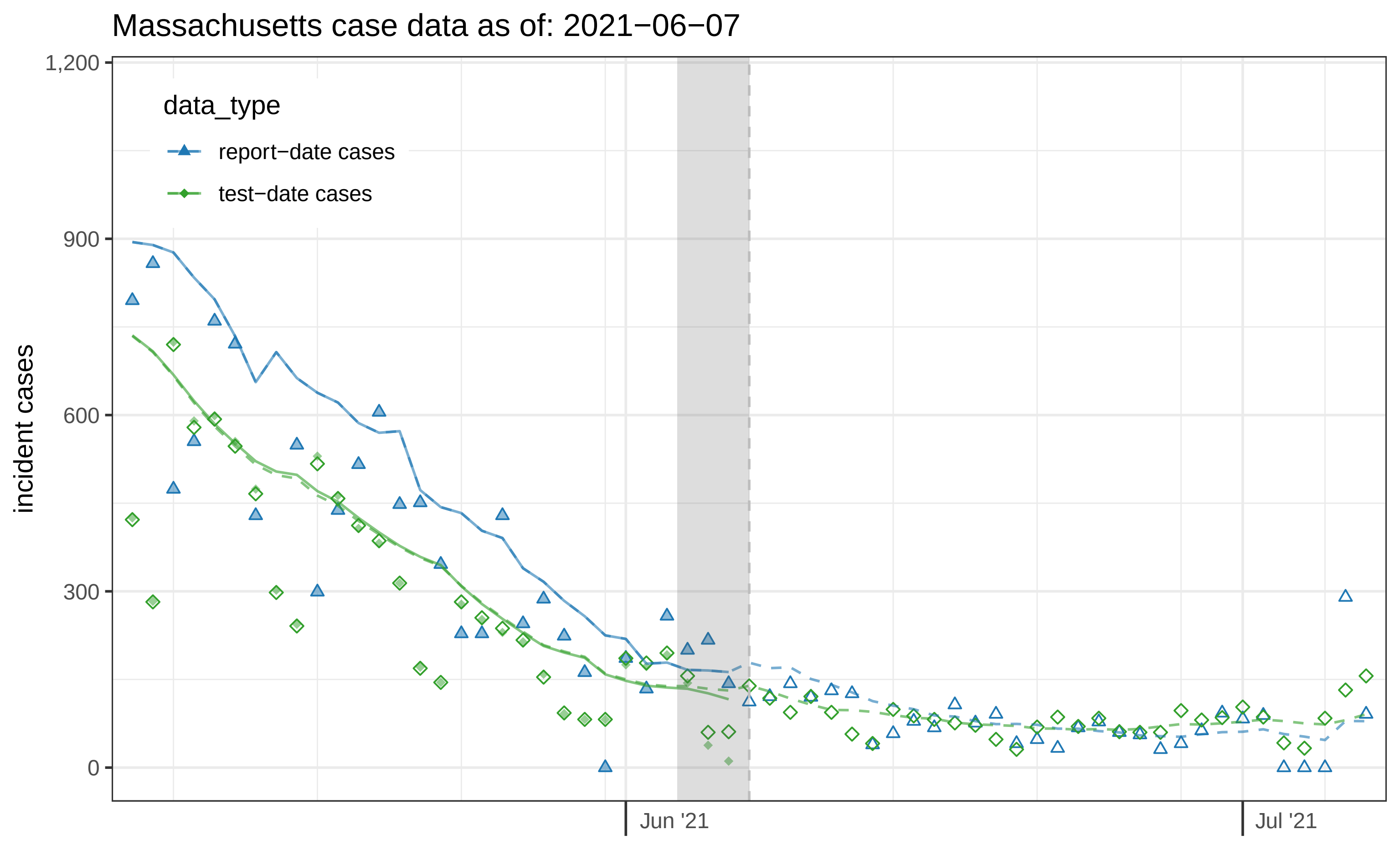

Massachusetts case data as of: 2021−06−07
1,200
900
600
300
0
incident cases
data_type
report−date cases
test−date cases
Jun '21
Jul '21

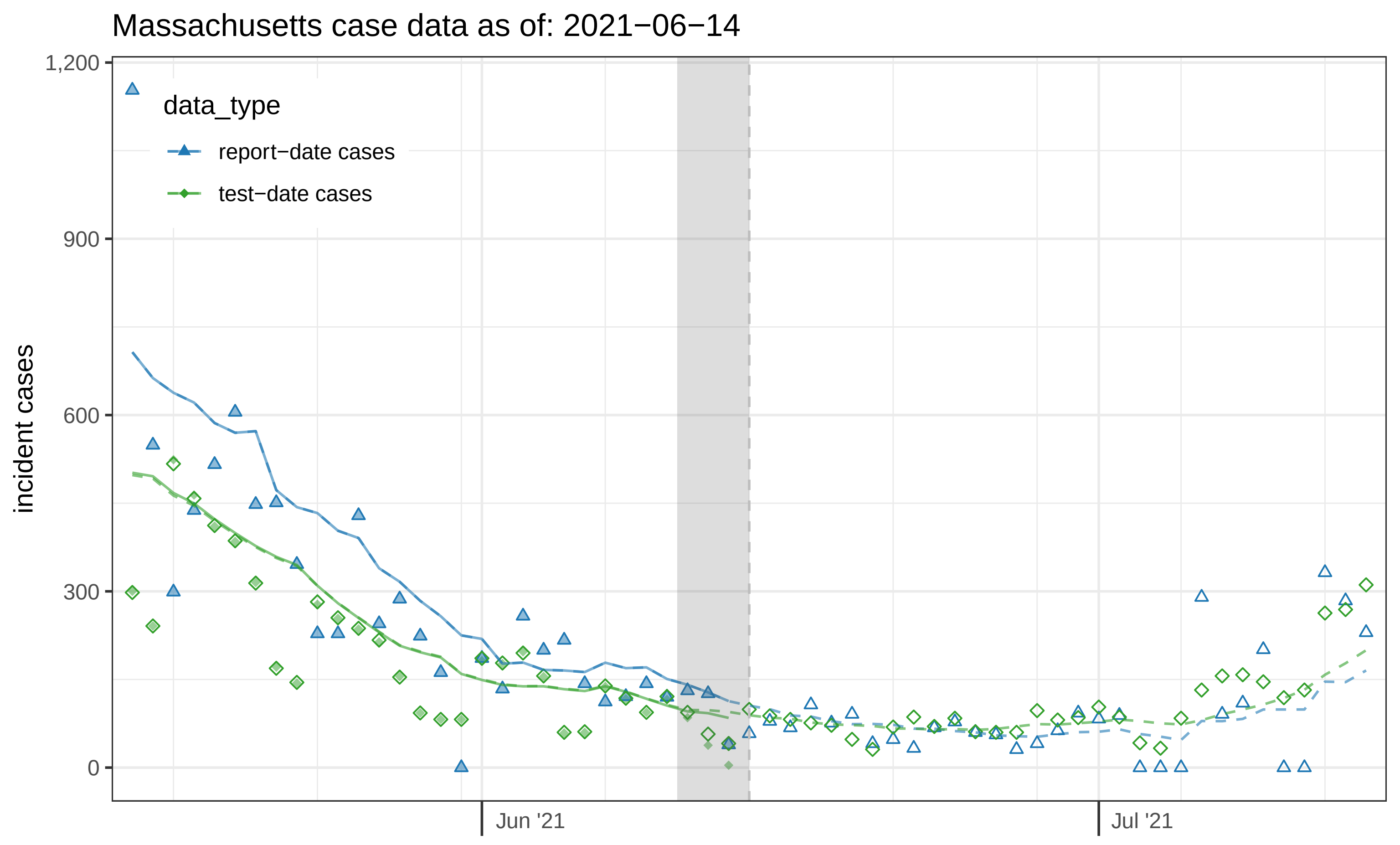

Massachusetts case data as of: 2021−06−14
1,200
900
600
300
0
incident cases
data_type
report−date cases
test−date cases
Jun '21
Jul '21

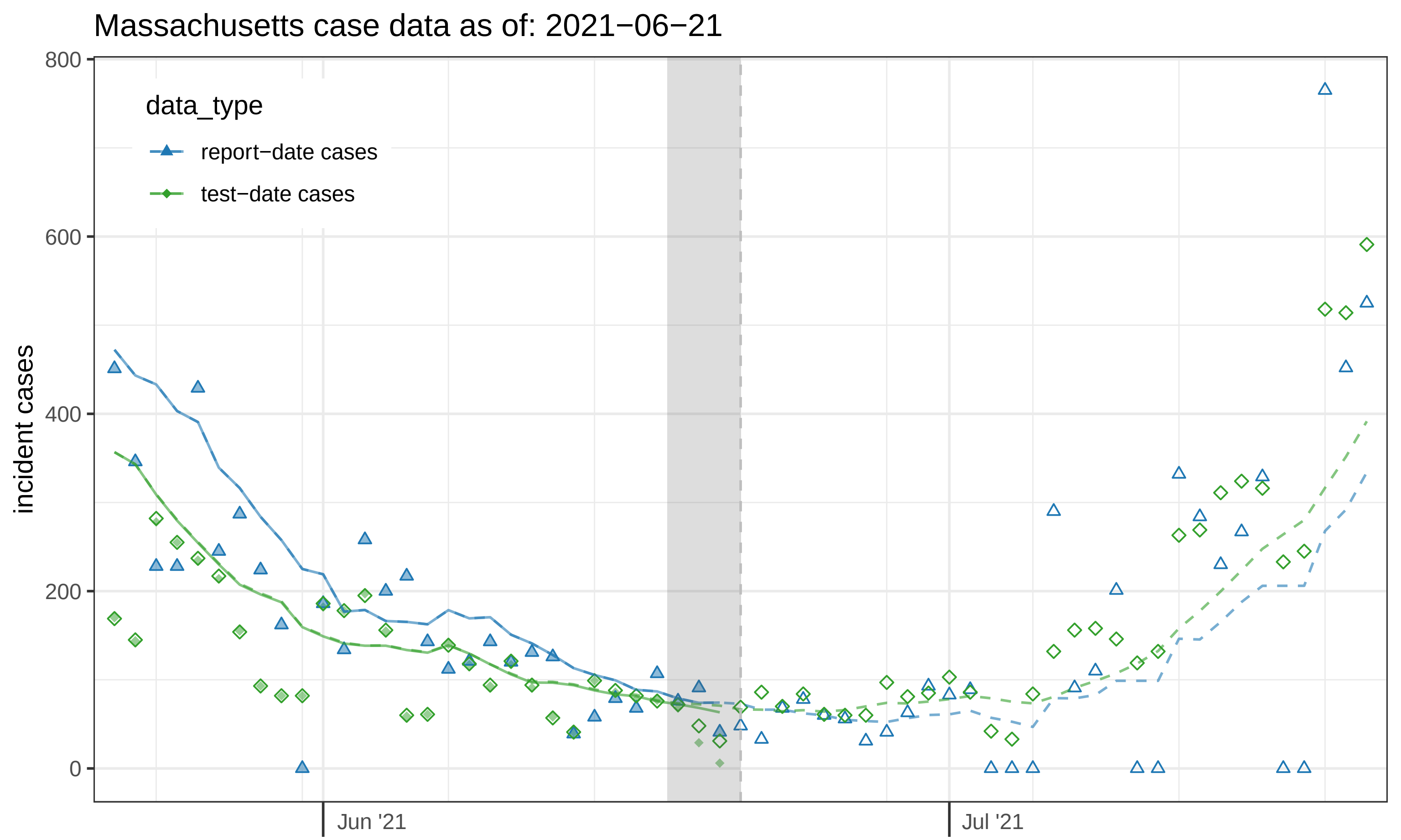
Massachusetts case data as of: 2021−06−21
data_type
report−date cases
test−date cases
incident cases
800
600
400
200
0
Jun '21
Jul '21

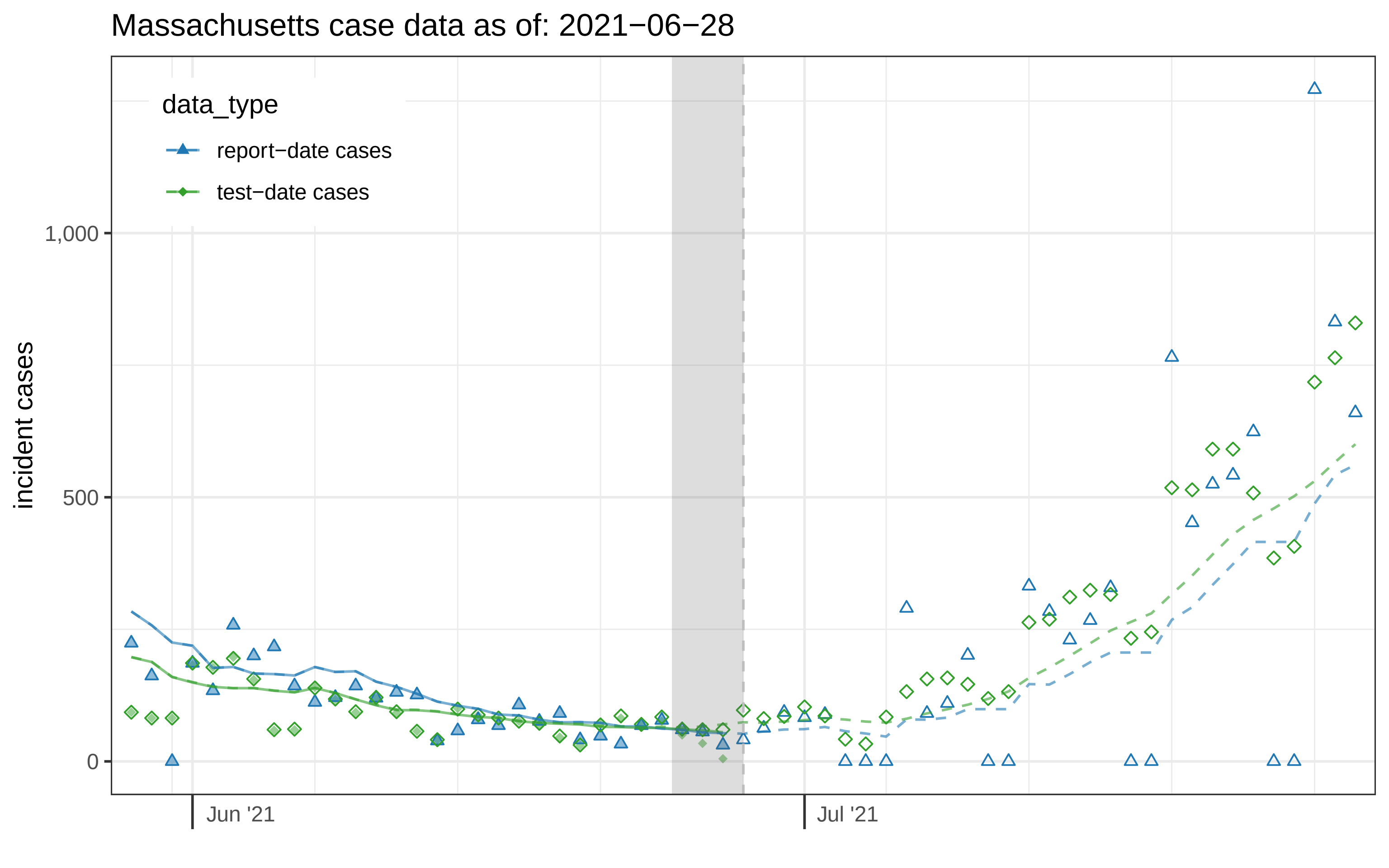

Massachusetts case data as of: 2021−06−28
data_type
report−date cases
test−date cases
incident cases
1,000
500
0
Jun '21
Jul '21

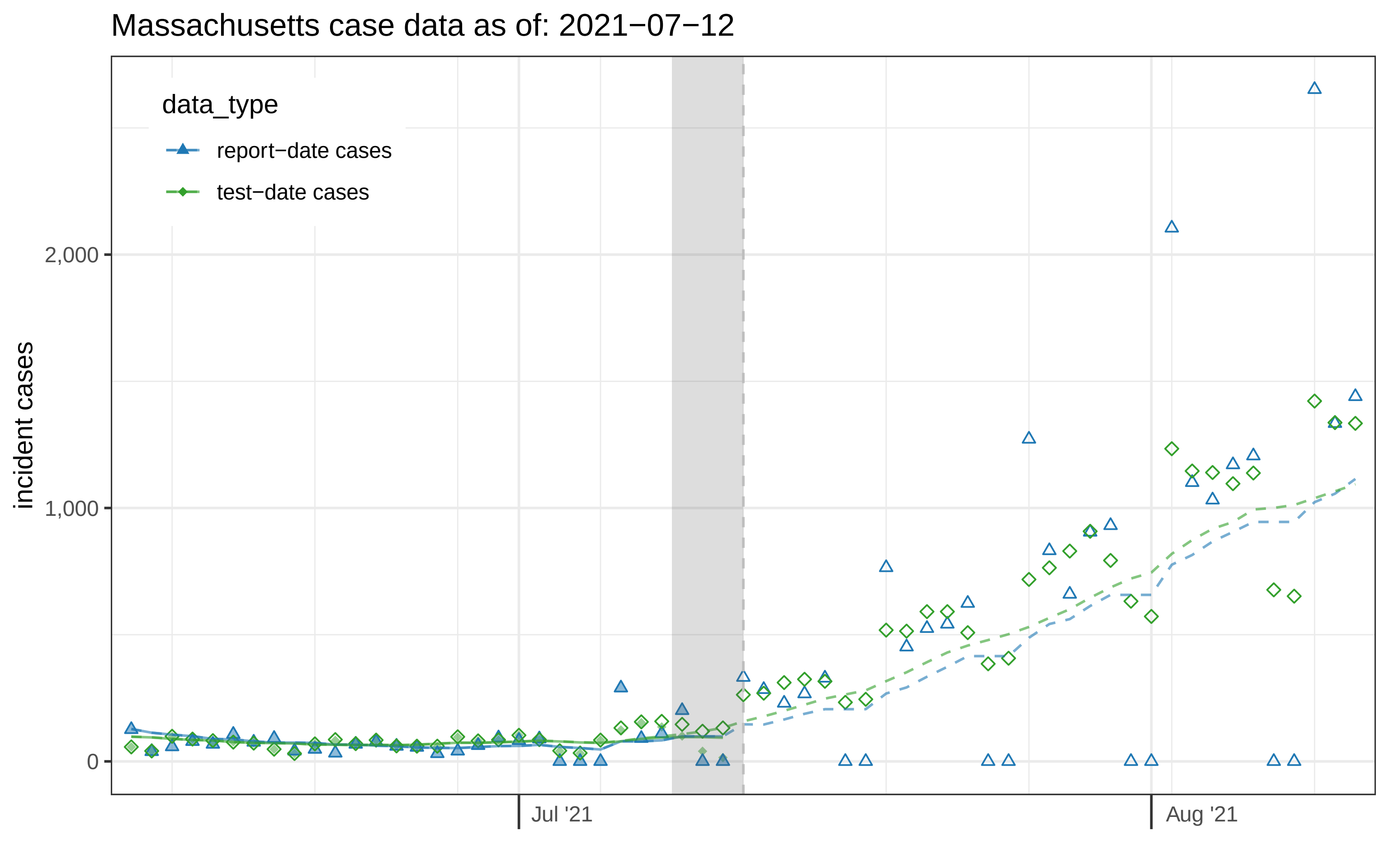
Massachusetts case data as of: 2021−07−12
data_type
report−date cases
test−date cases
incident cases
2,000
1,000
0
Jul '21
Aug '21

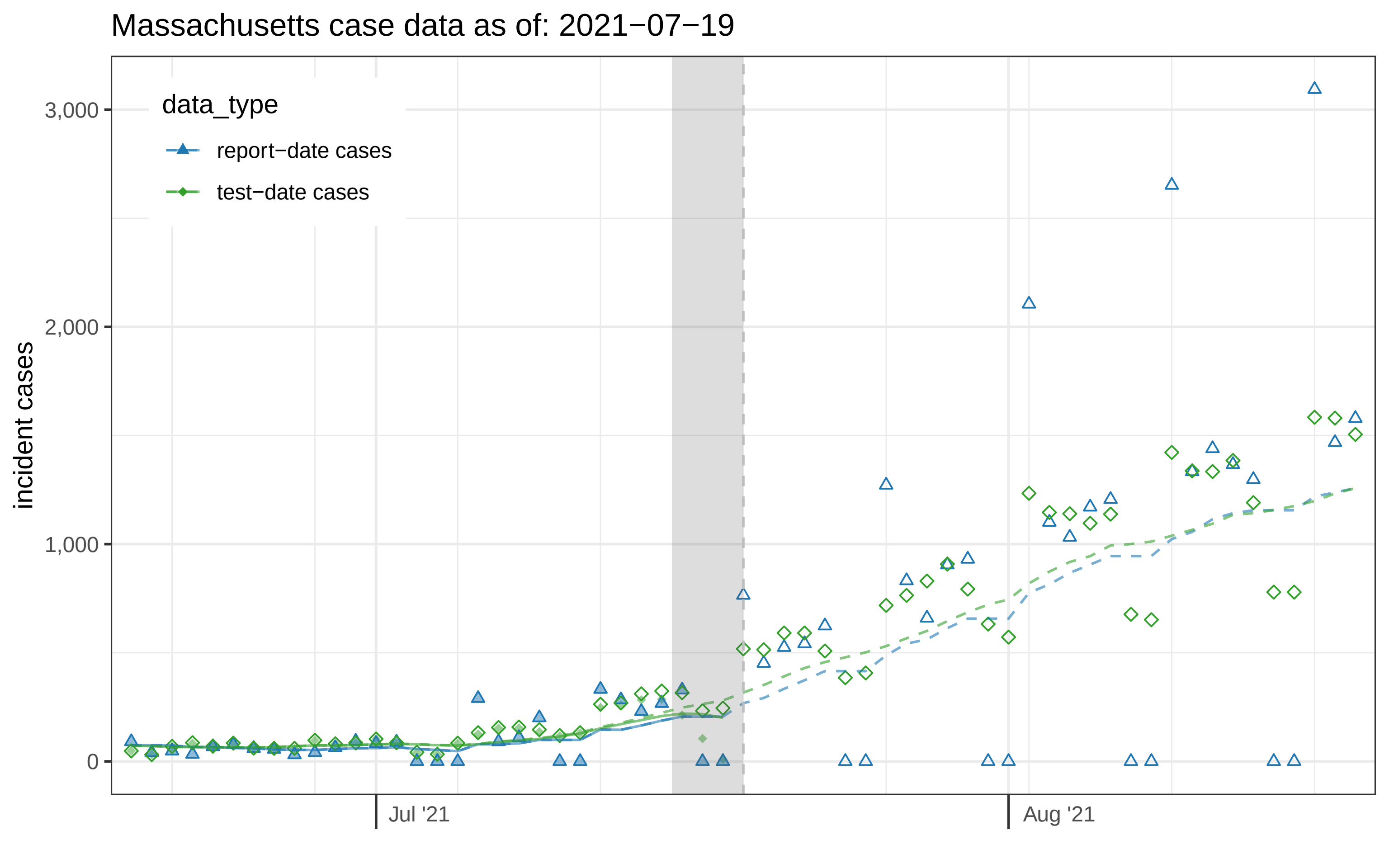

Massachusetts case data as of: 2021−07−19
data_type
report−date cases
test−date cases
incident cases
3,000
2,000
1,000
0
Jul '21
Aug '21

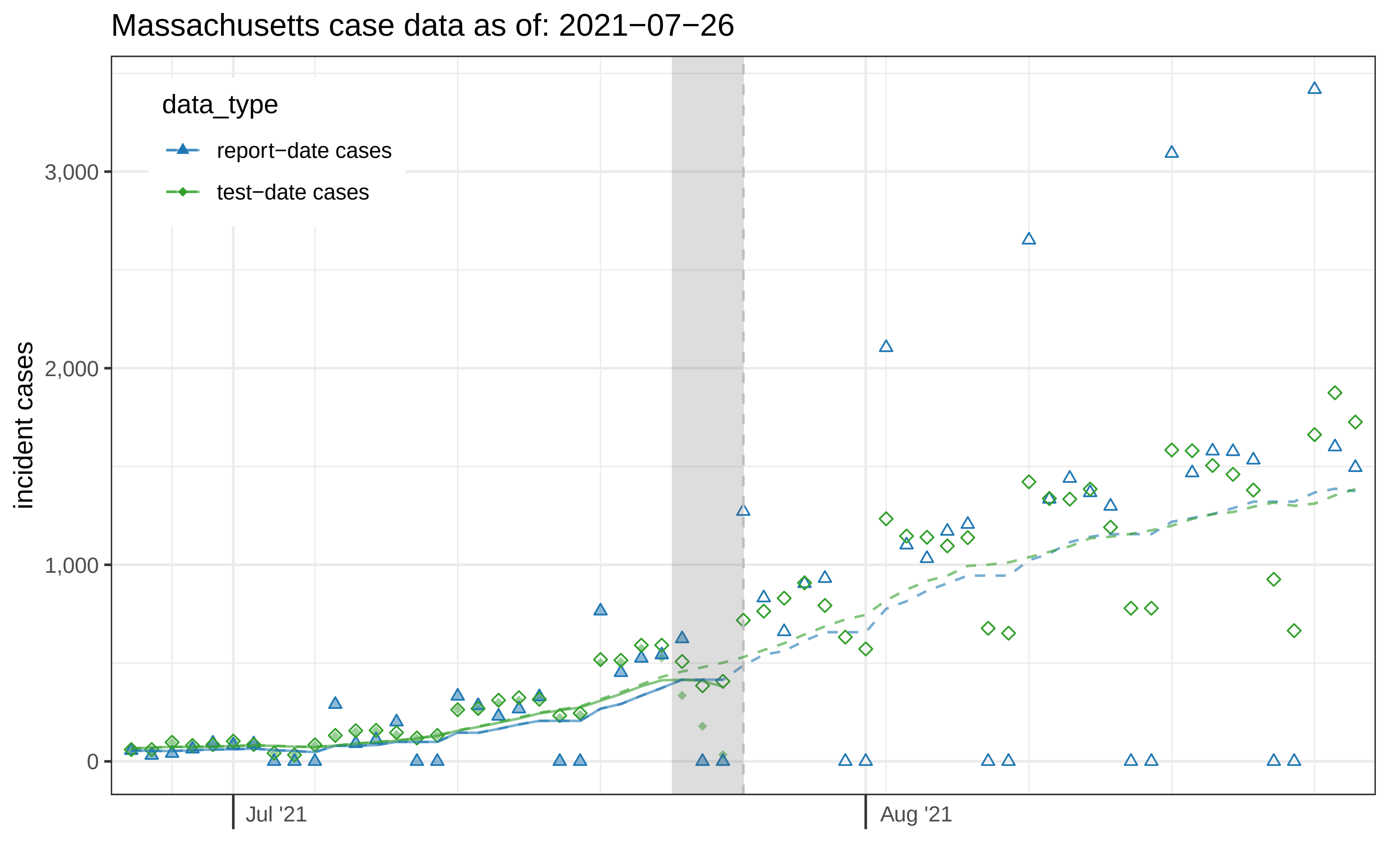

Massachusetts case data as of: 2021−07−26
data_type
report−date cases
test−date cases
incident cases
3,000
2,000
1,000
0
Jul '21
Aug '21

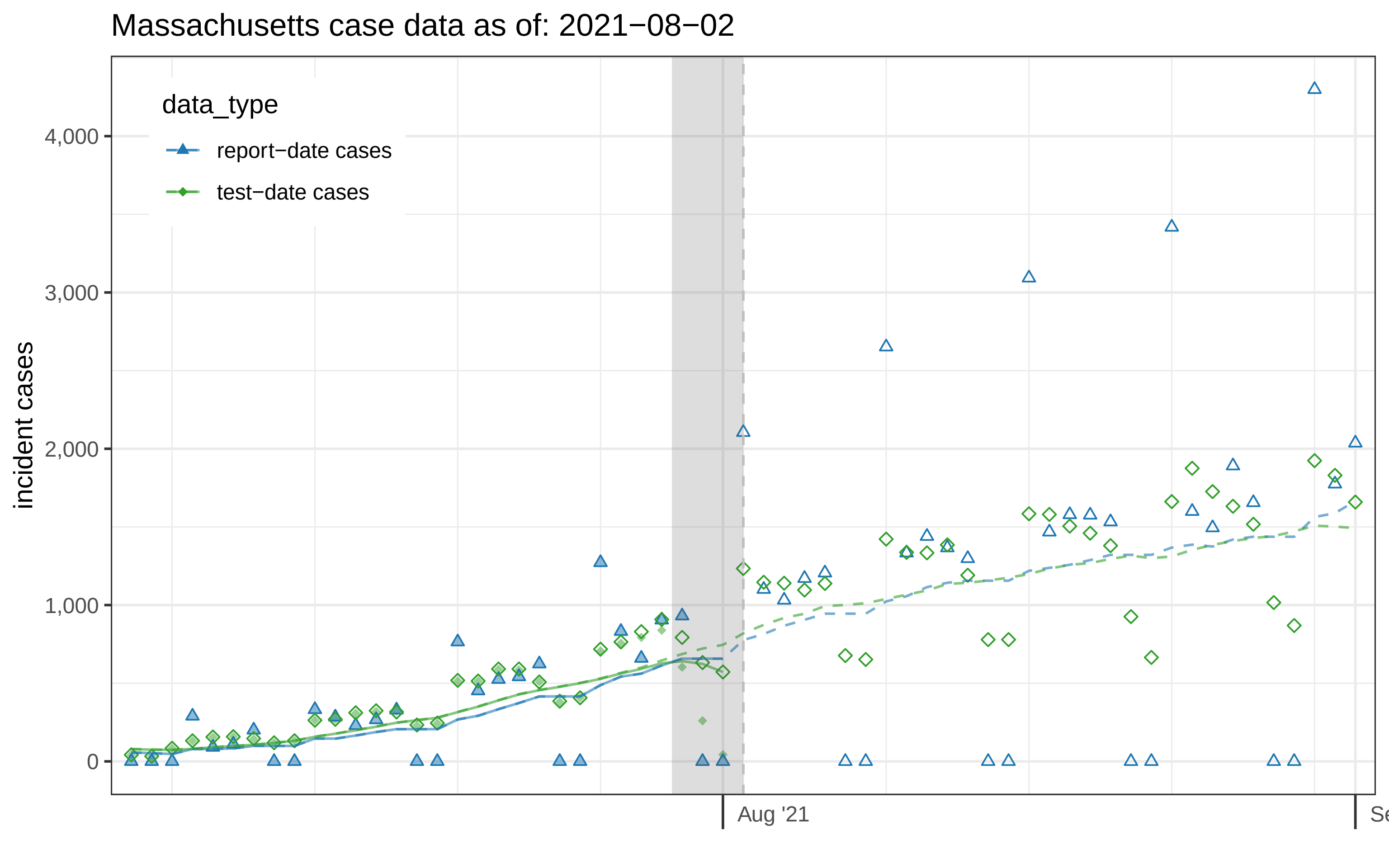
Massachusetts case data as of: 2021−08−02
data_type
report−date cases
test−date cases
incident cases
4,000
3,000
2,000
1,000
0
Aug '21
Se

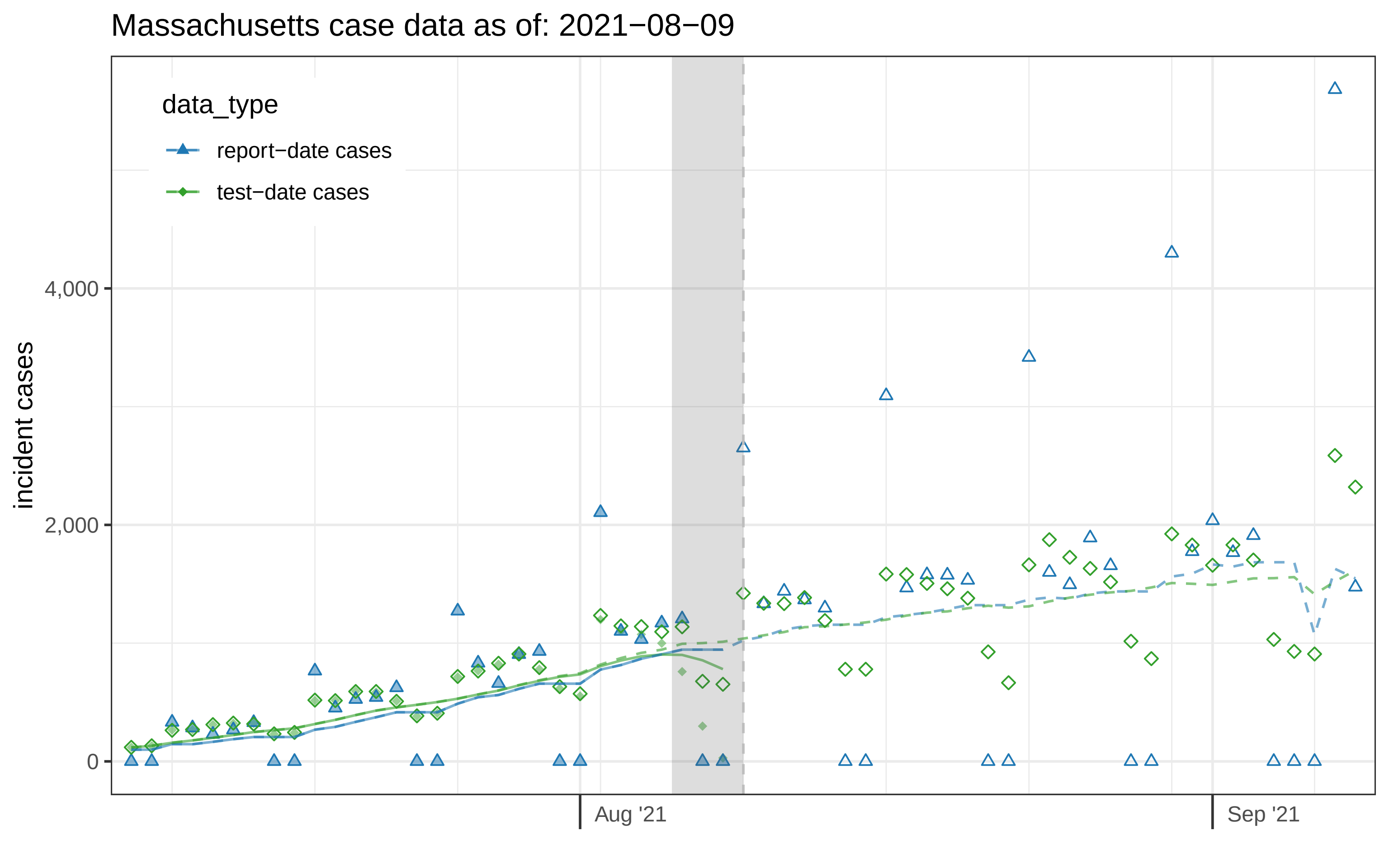

Massachusetts case data as of: 2021−08−09
data_type
report−date cases
test−date cases
incident cases
4,000
2,000
0
Aug '21
Sep '21

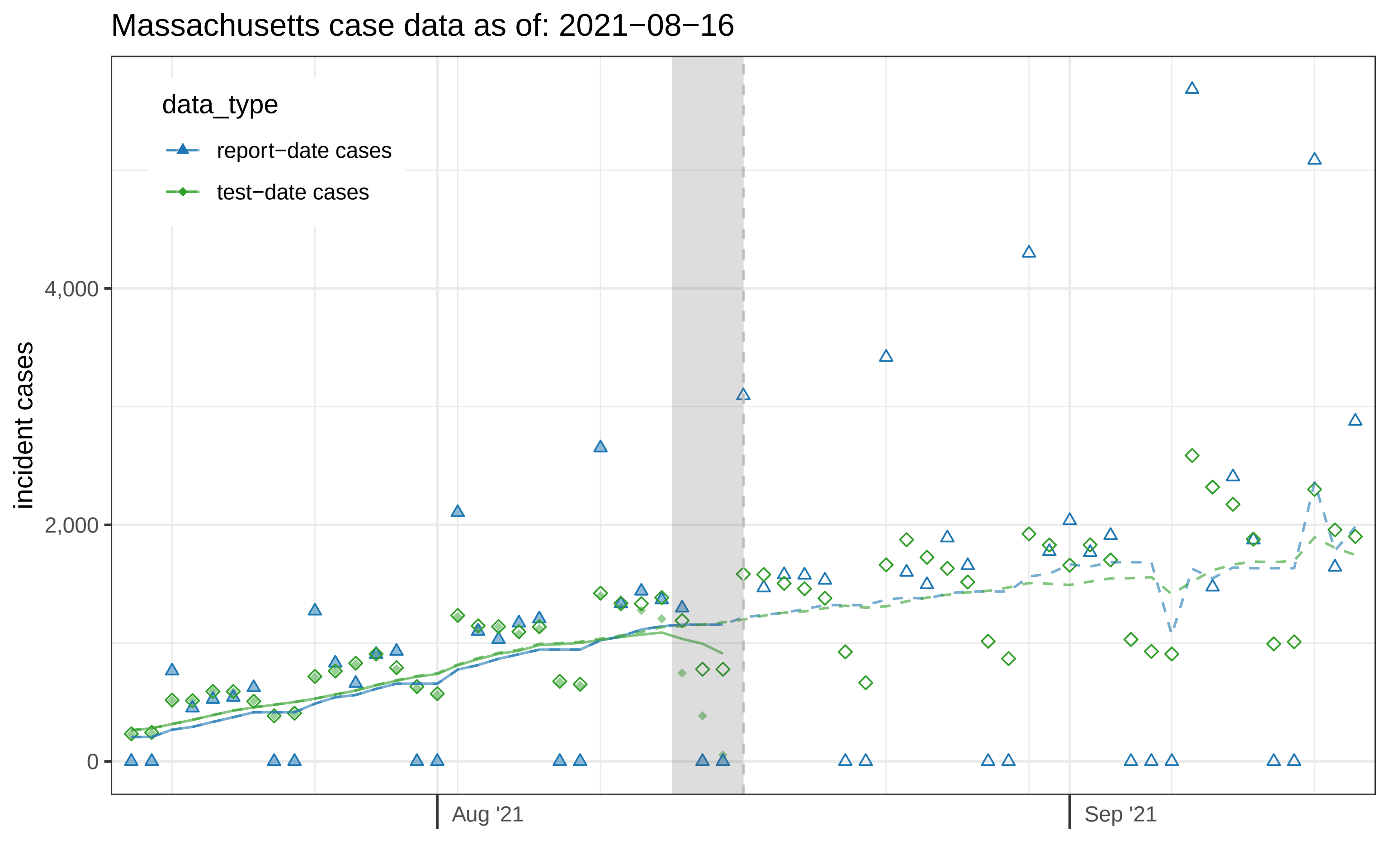

Massachusetts case data as of: 2021−08−16
data_type
report−date cases
test−date cases
incident cases
4,000
2,000
0
Aug '21
Sep '21

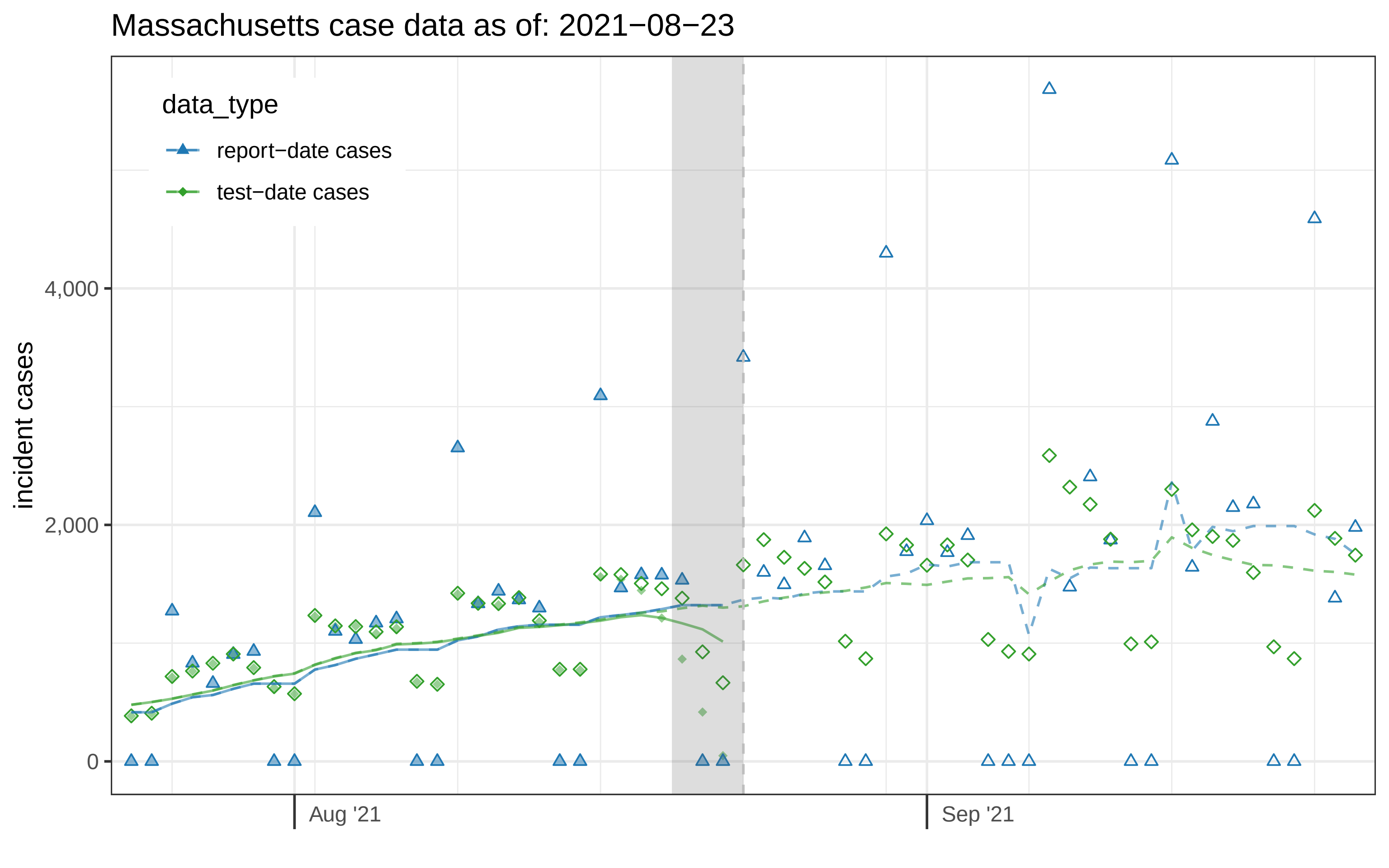

Massachusetts case data as of: 2021−08−23
data_type
report−date cases
test−date cases
incident cases
4,000
2,000
0
Aug '21
Sep '21

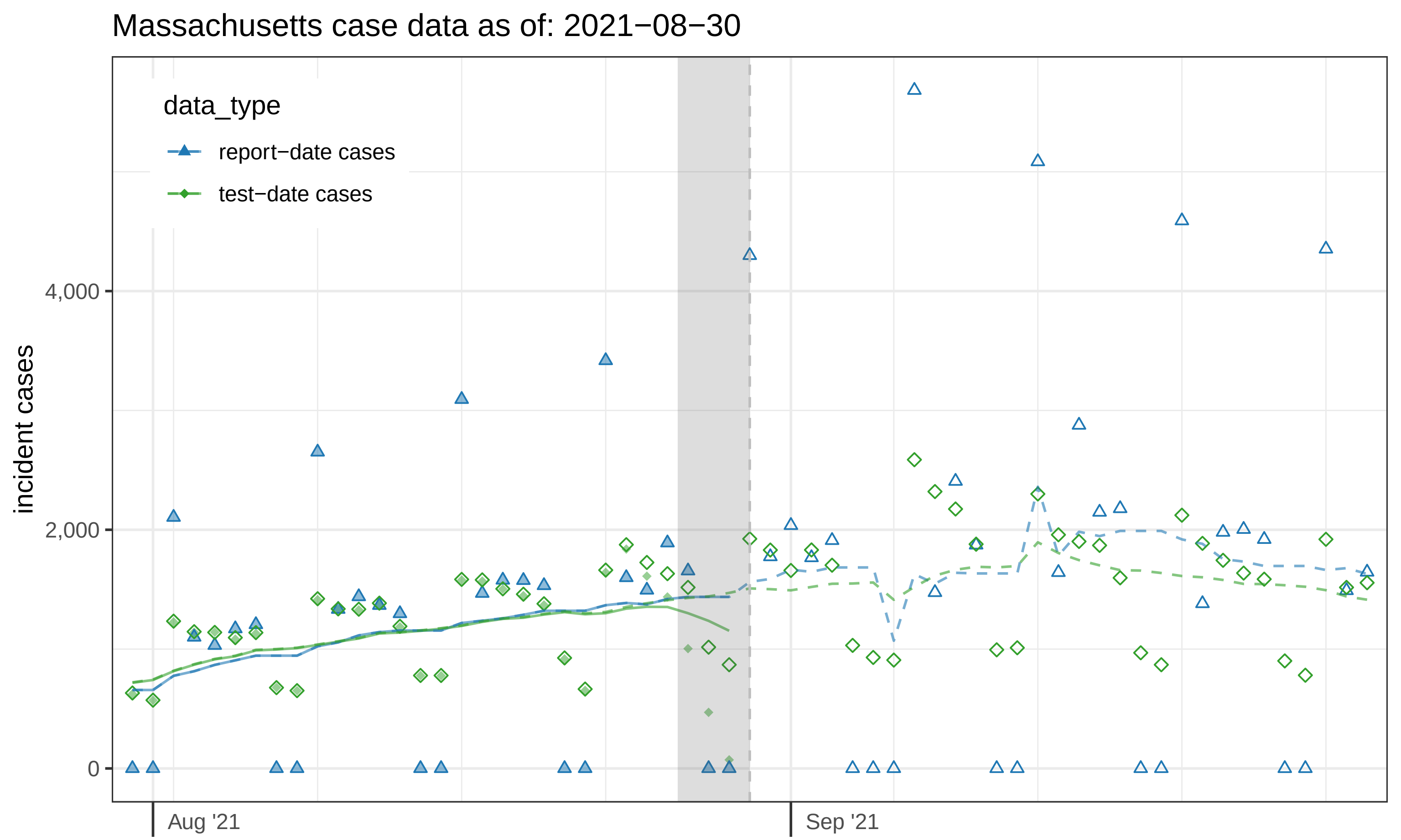
Massachusetts case data as of: 2021−08−30
data_type
report−date cases
test−date cases
incident cases
4,000
2,000
0
Aug '21
Sep '21

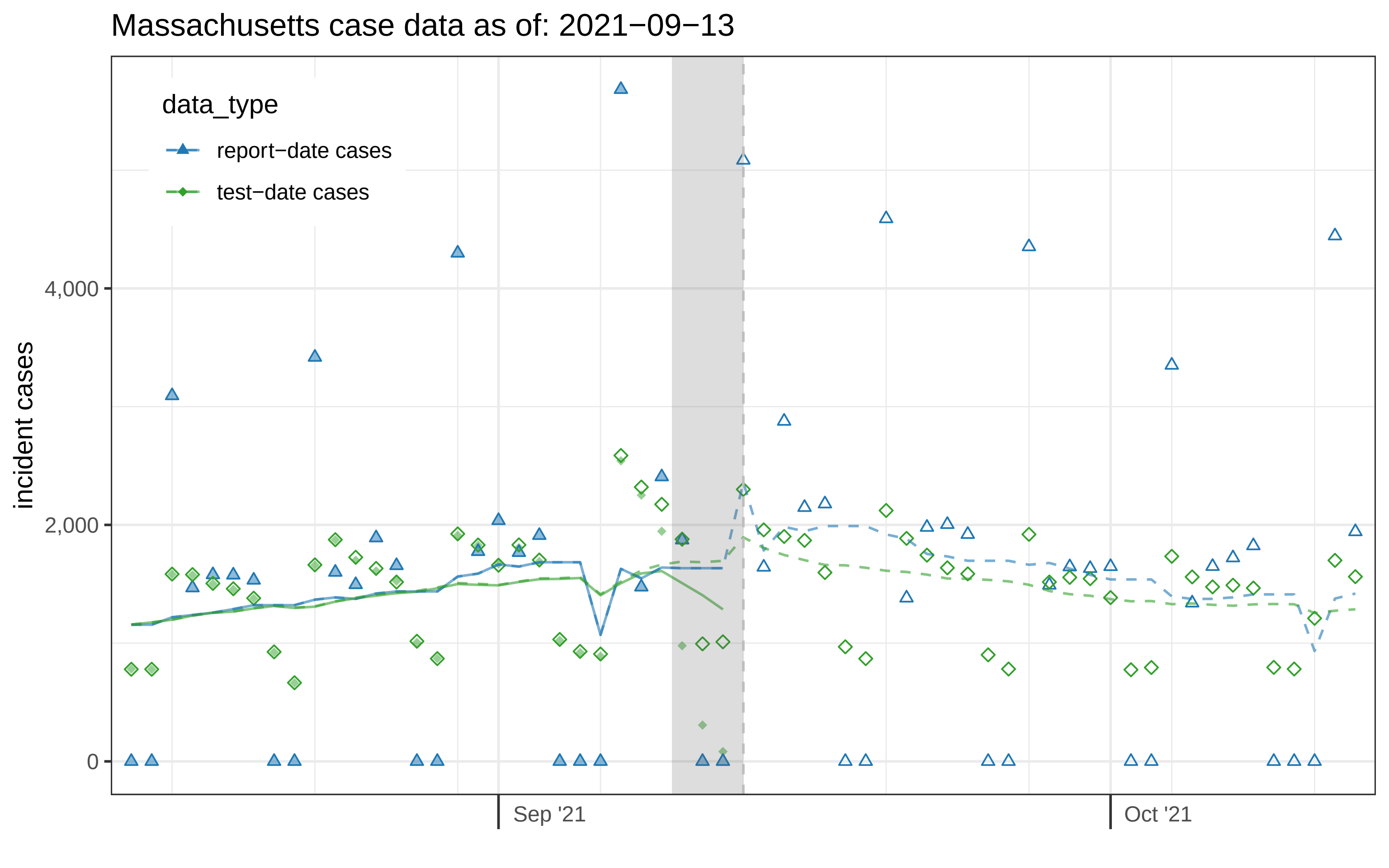

Massachusetts case data as of: 2021−09−13
data_type
report−date cases
test−date cases
incident cases
4,000
2,000
0
Sep '21
Oct '21

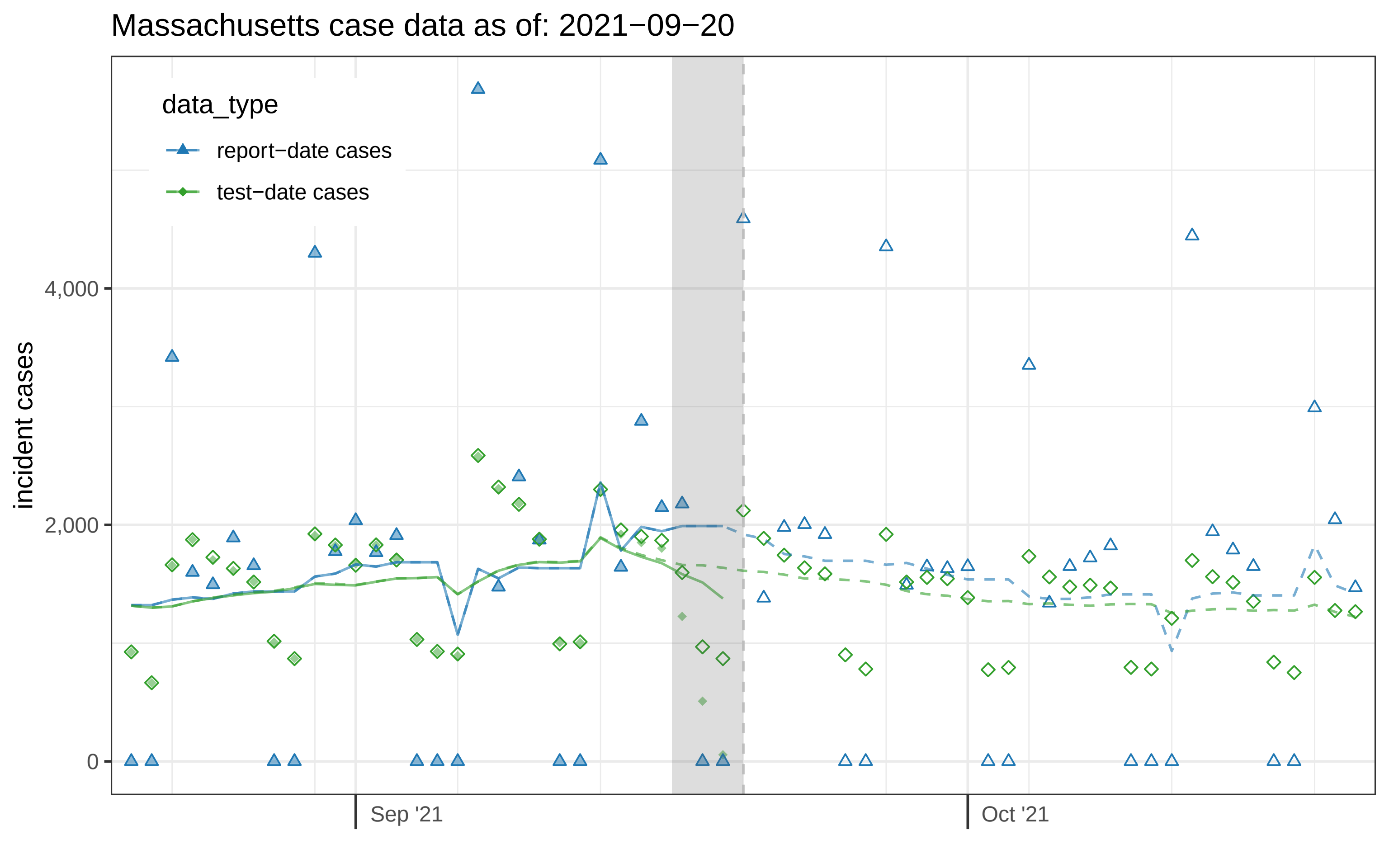

Massachusetts case data as of: 2021−09−20
data_type
report−date cases
test−date cases
incident cases
4,000
2,000
0
Sep '21
Oct '21

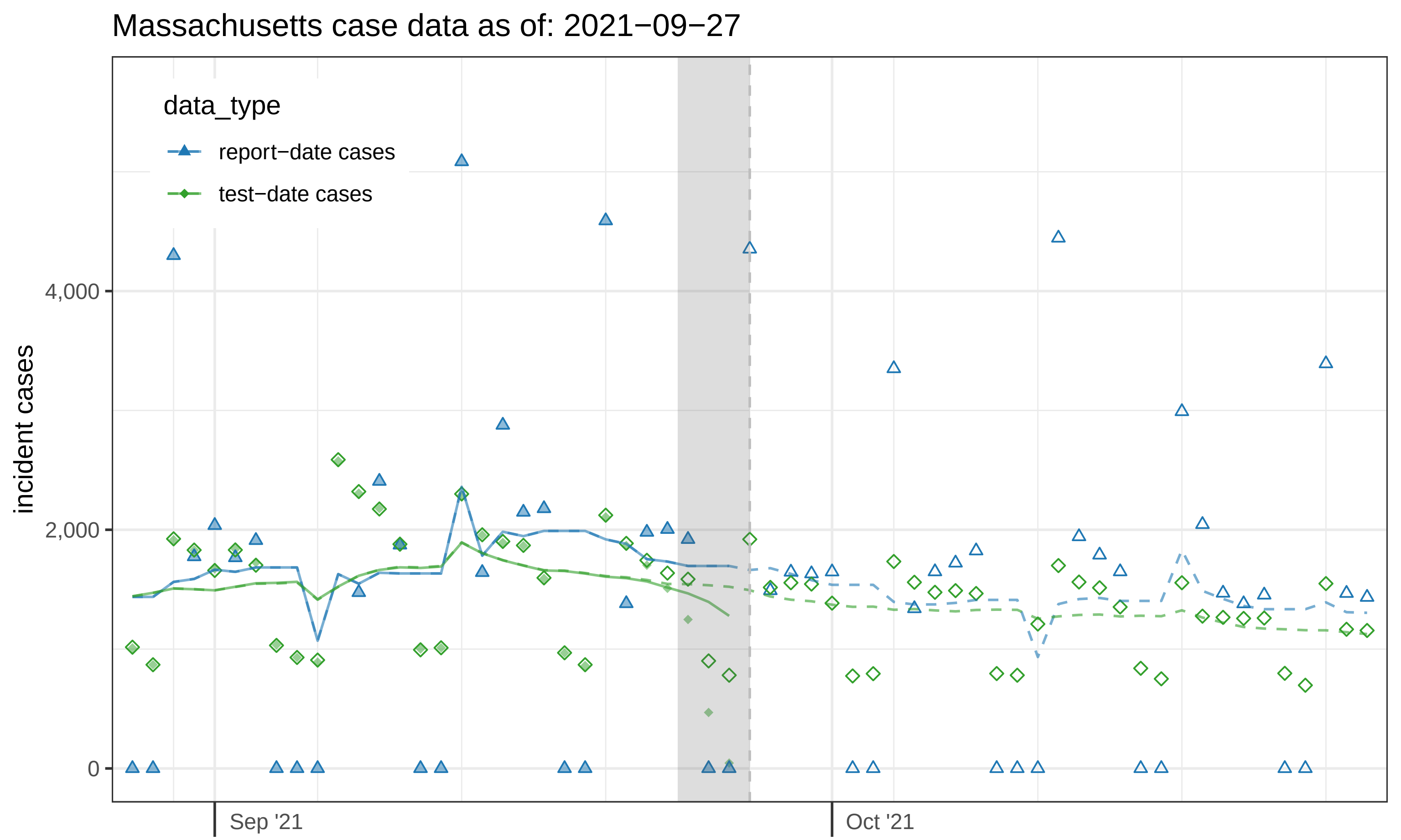

Massachusetts case data as of: 2021−09−27
data_type
report−date cases
test−date cases
incident cases
4,000
2,000
0
Sep '21
Oct '21

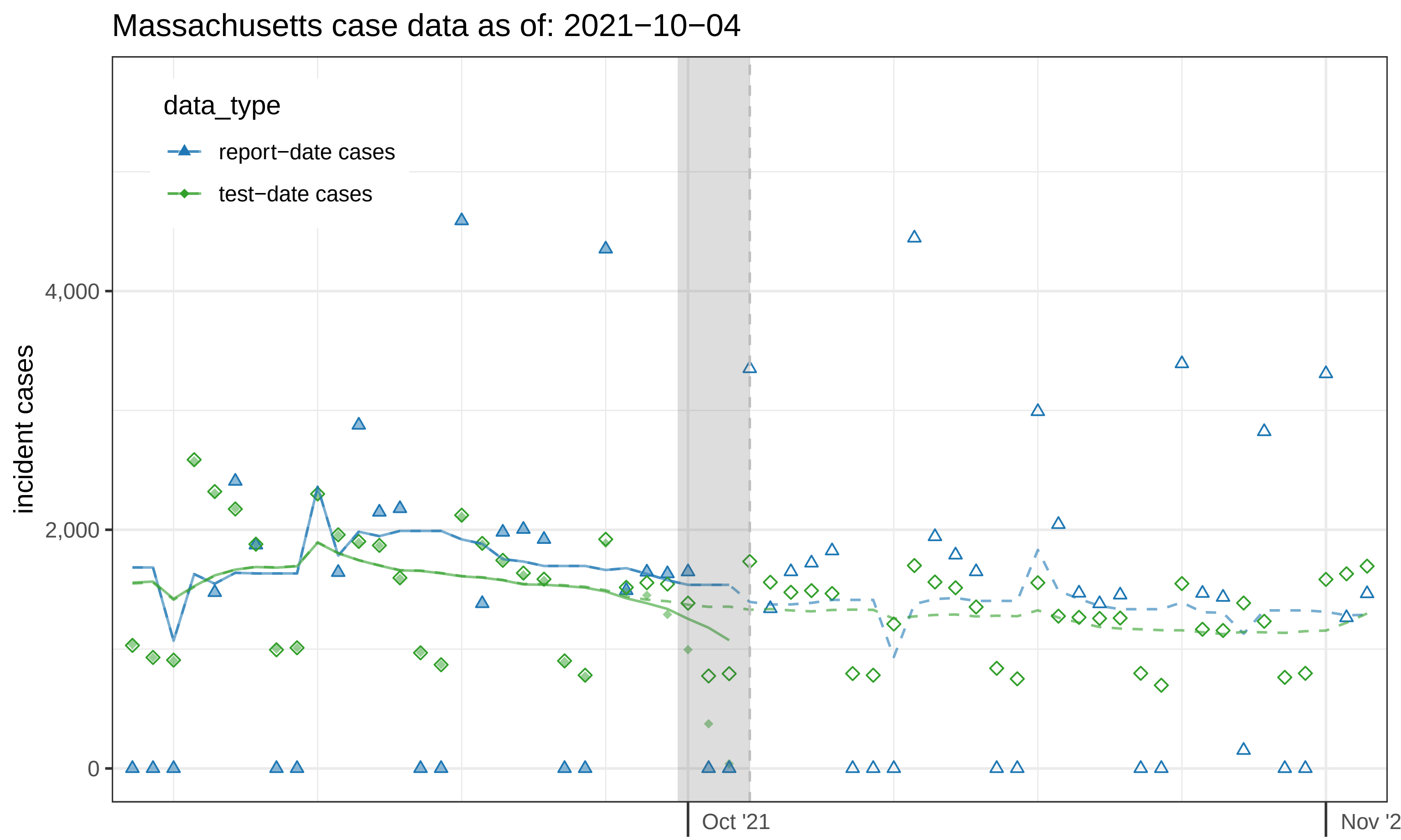
Massachusetts case data as of: 2021−10−04
data_type
report−date cases
test−date cases
incident cases
4,000
2,000
0
Oct '21
Nov '2

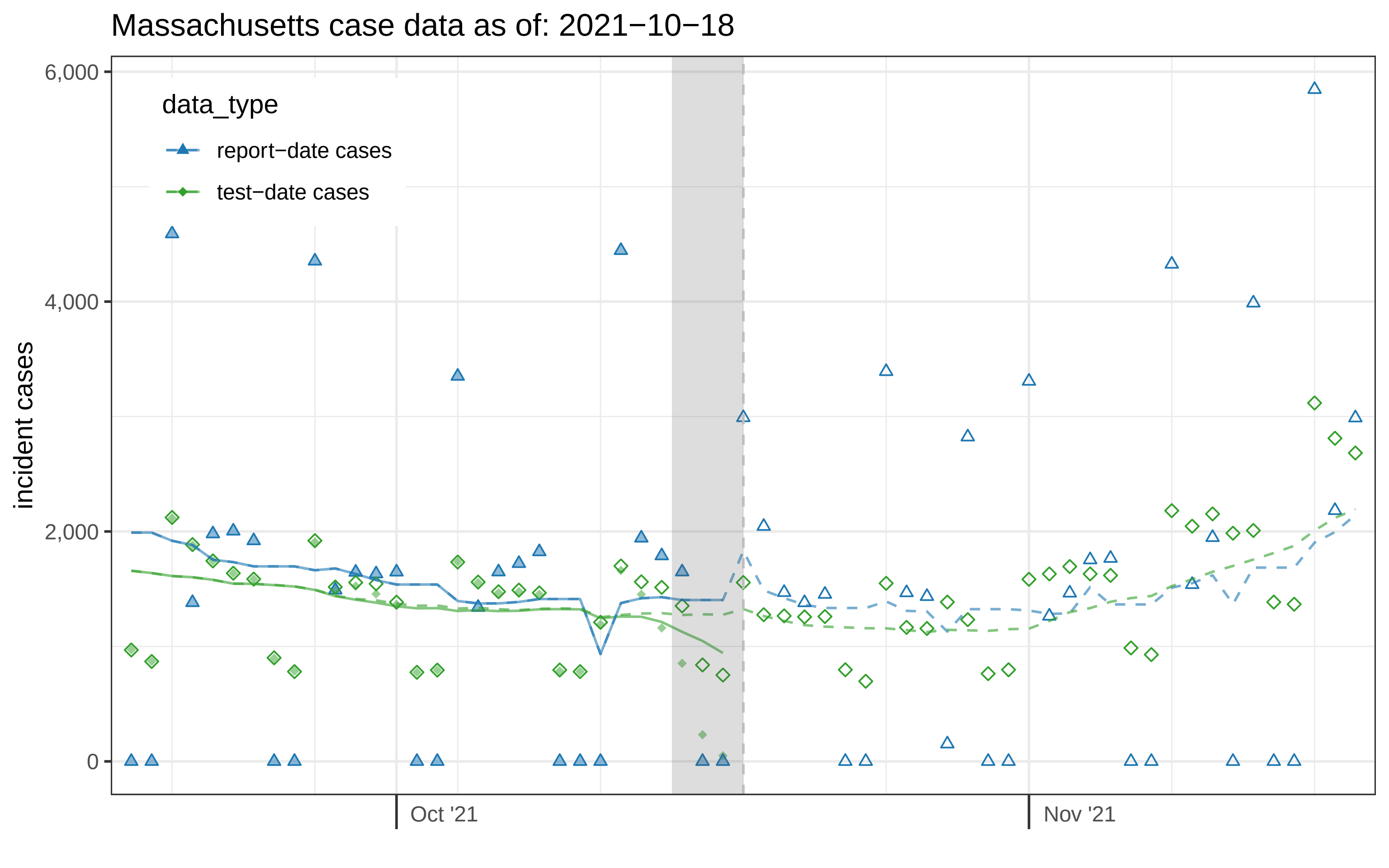

Massachusetts case data as of: 2021−10−18
data_type
report−date cases
test−date cases
incident cases
6,000
4,000
2,000
0
Oct '21
Nov '21

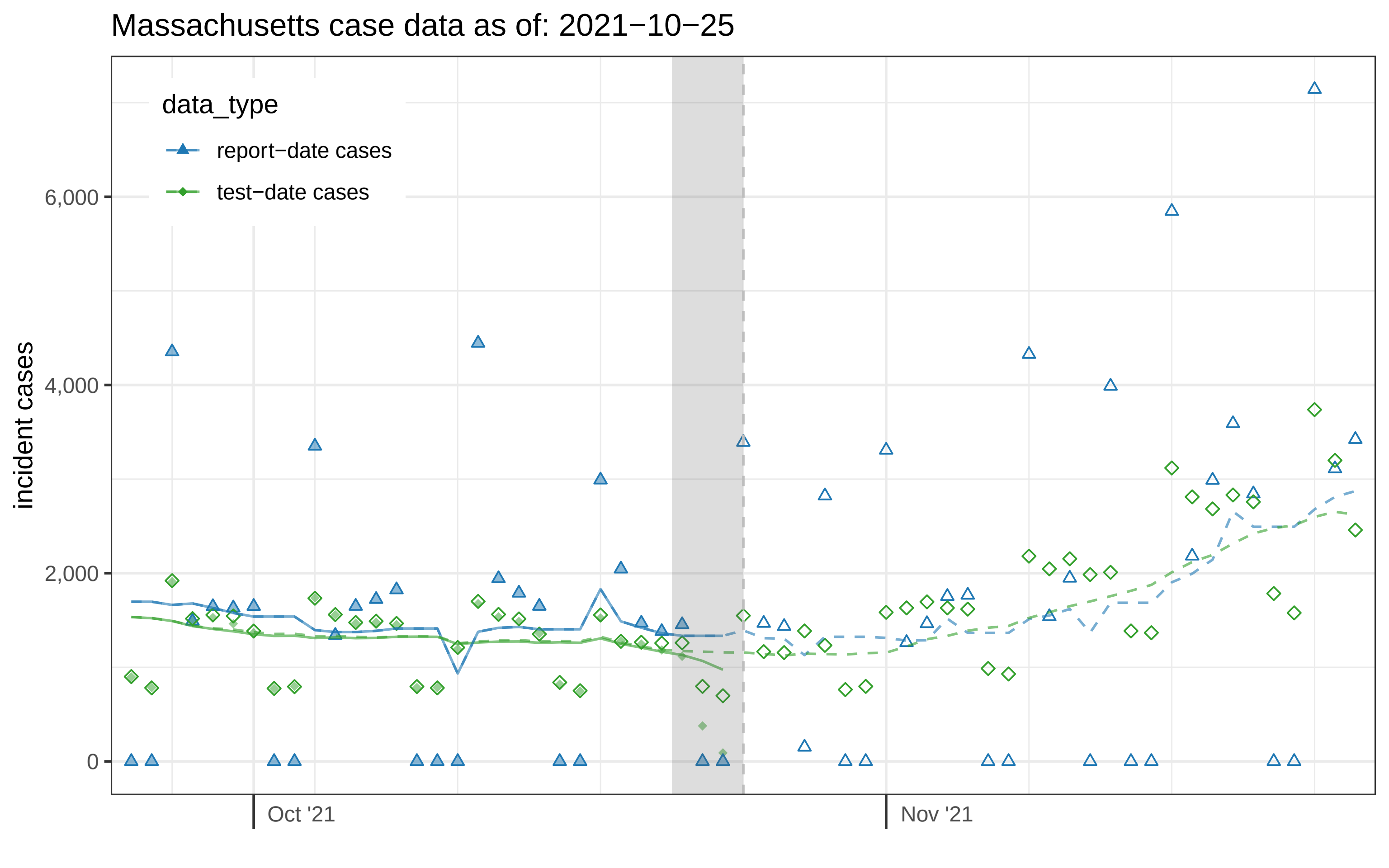
Massachusetts case data as of: 2021−10−25
data_type
report−date cases
test−date cases
incident cases
6,000
4,000
2,000
0
Oct '21
Nov '21

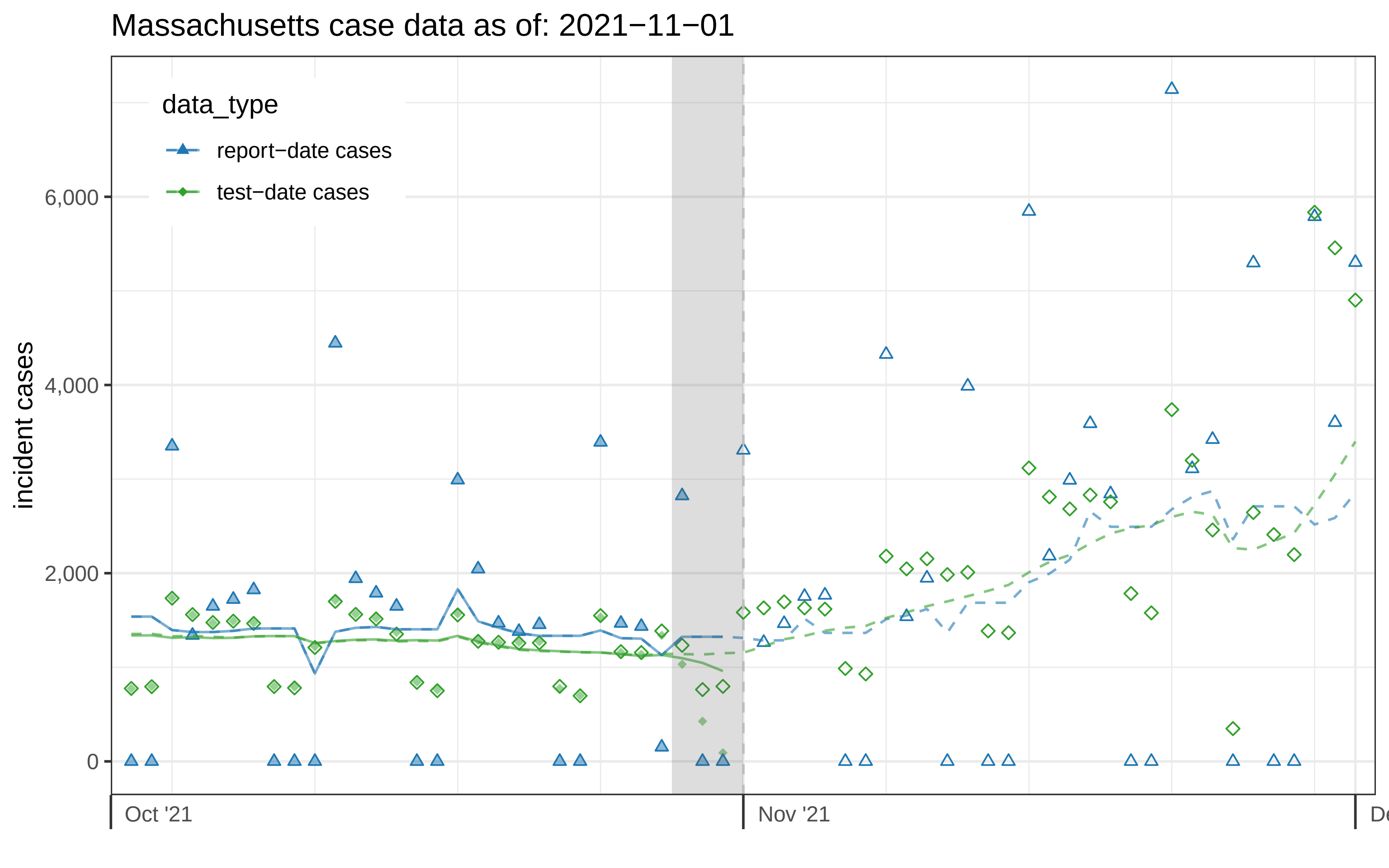

Massachusetts case data as of: 2021−11−01
data_type
report−date cases
test−date cases
incident cases
6,000
4,000
2,000
0
Oct '21
Nov '21

### Massachusetts case data as of: 2021−11−08

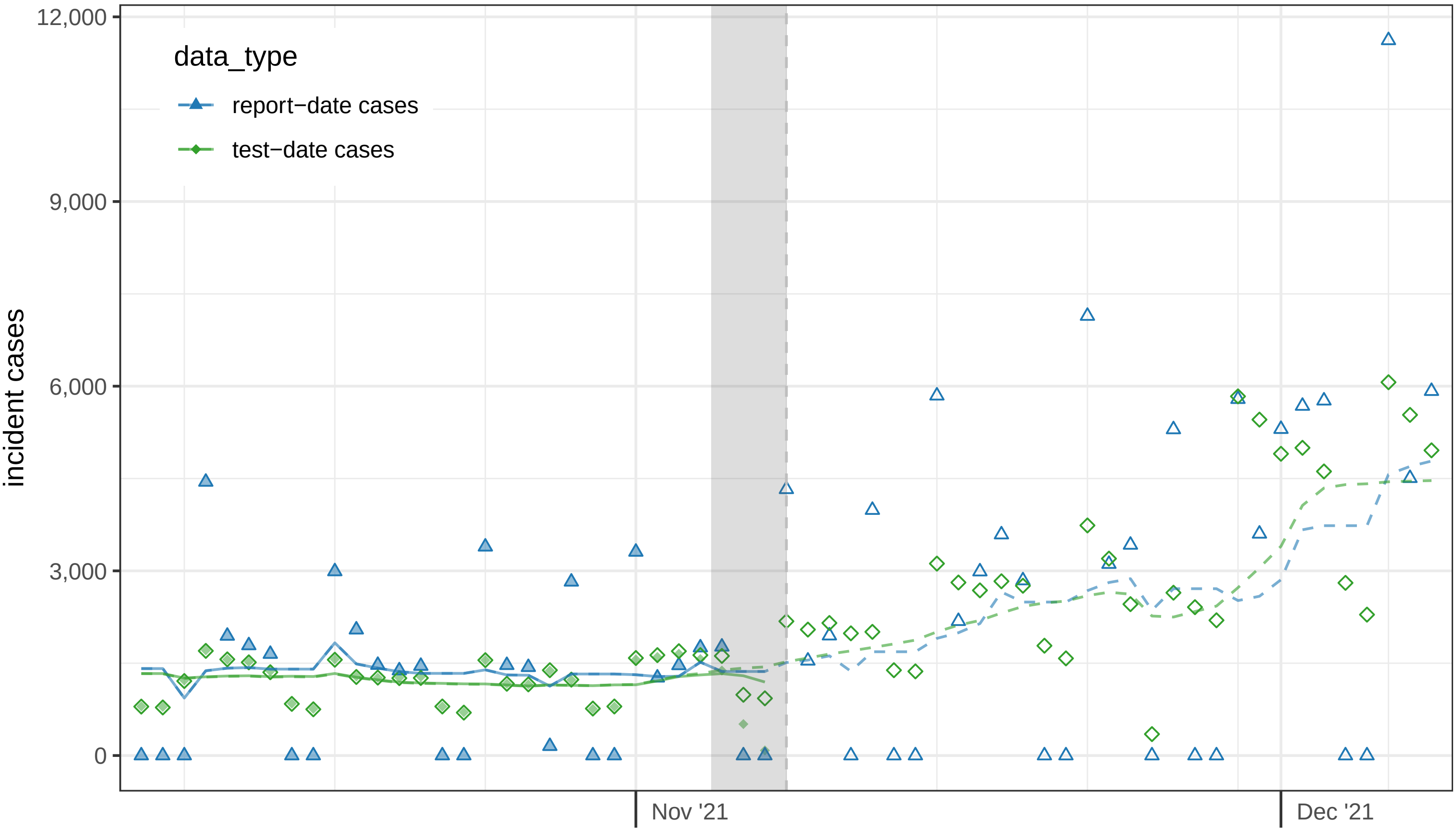

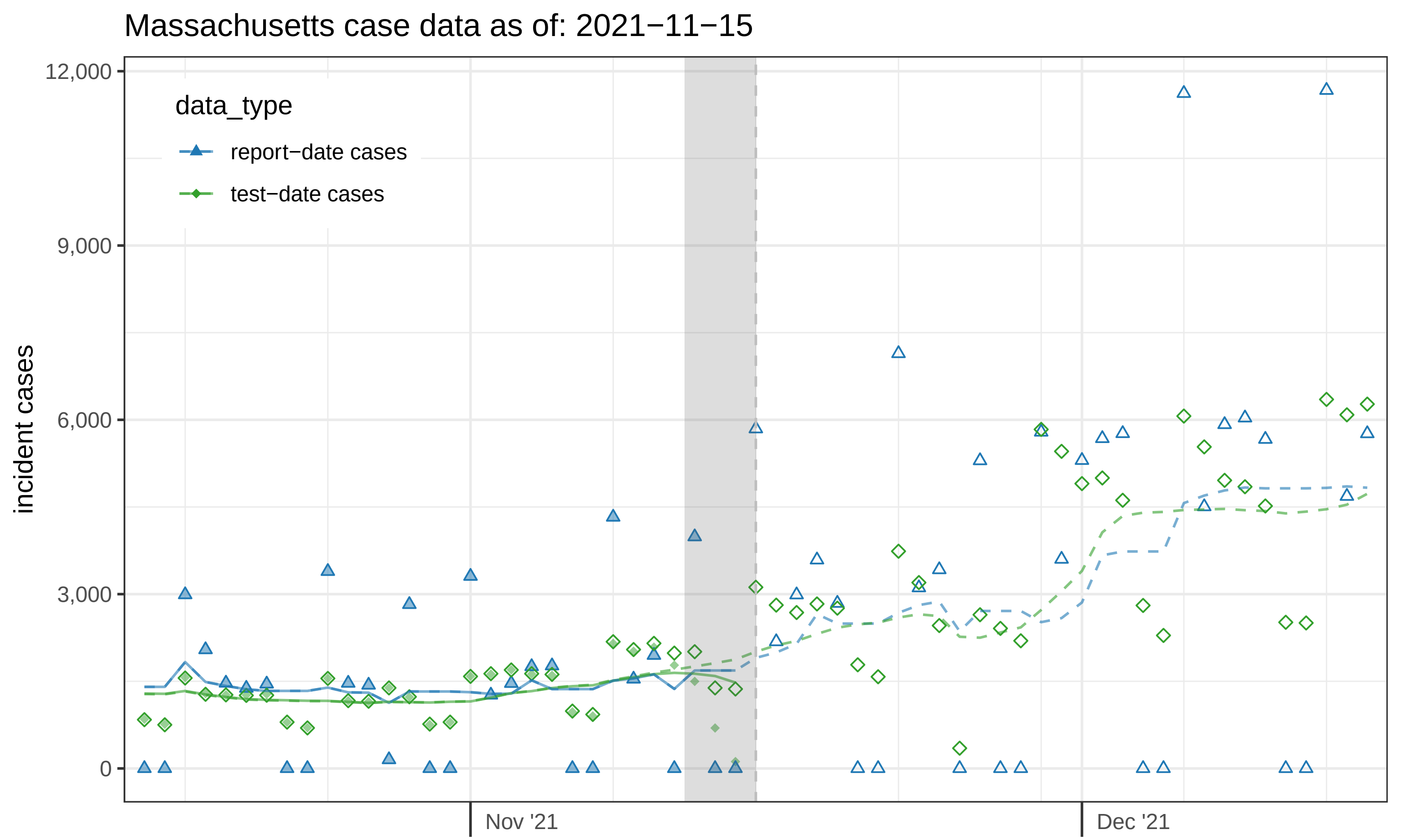
Massachusetts case data as of: 2021−11−15
data_type
report−date cases
test−date cases
incident cases
12,000
9,000
6,000
3,000
0
Nov '21
Dec '21

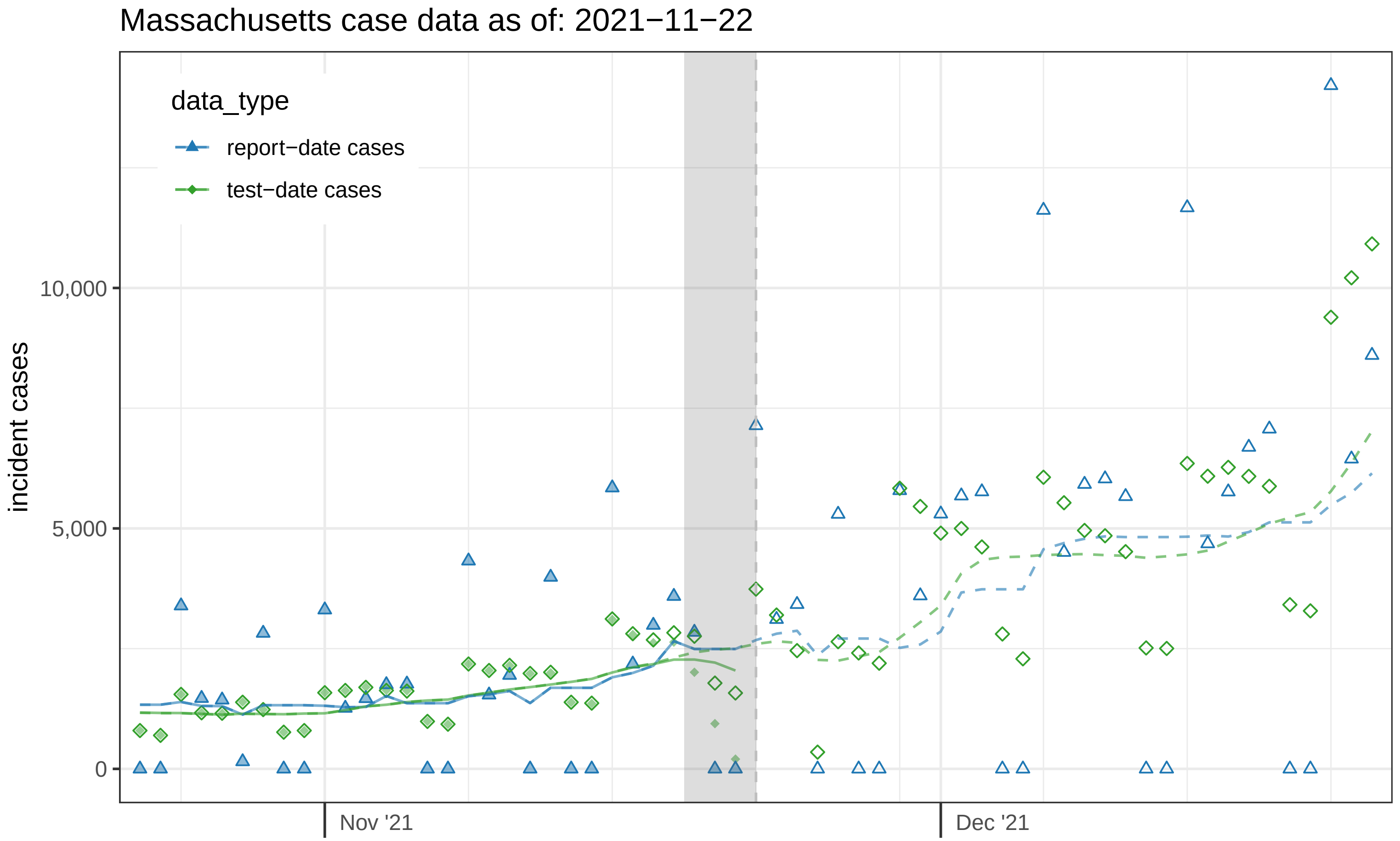

Massachusetts case data as of: 2021−11−22
data_type
report−date cases
test−date cases
incident cases
10,000
5,000
0
Nov '21
Dec '21

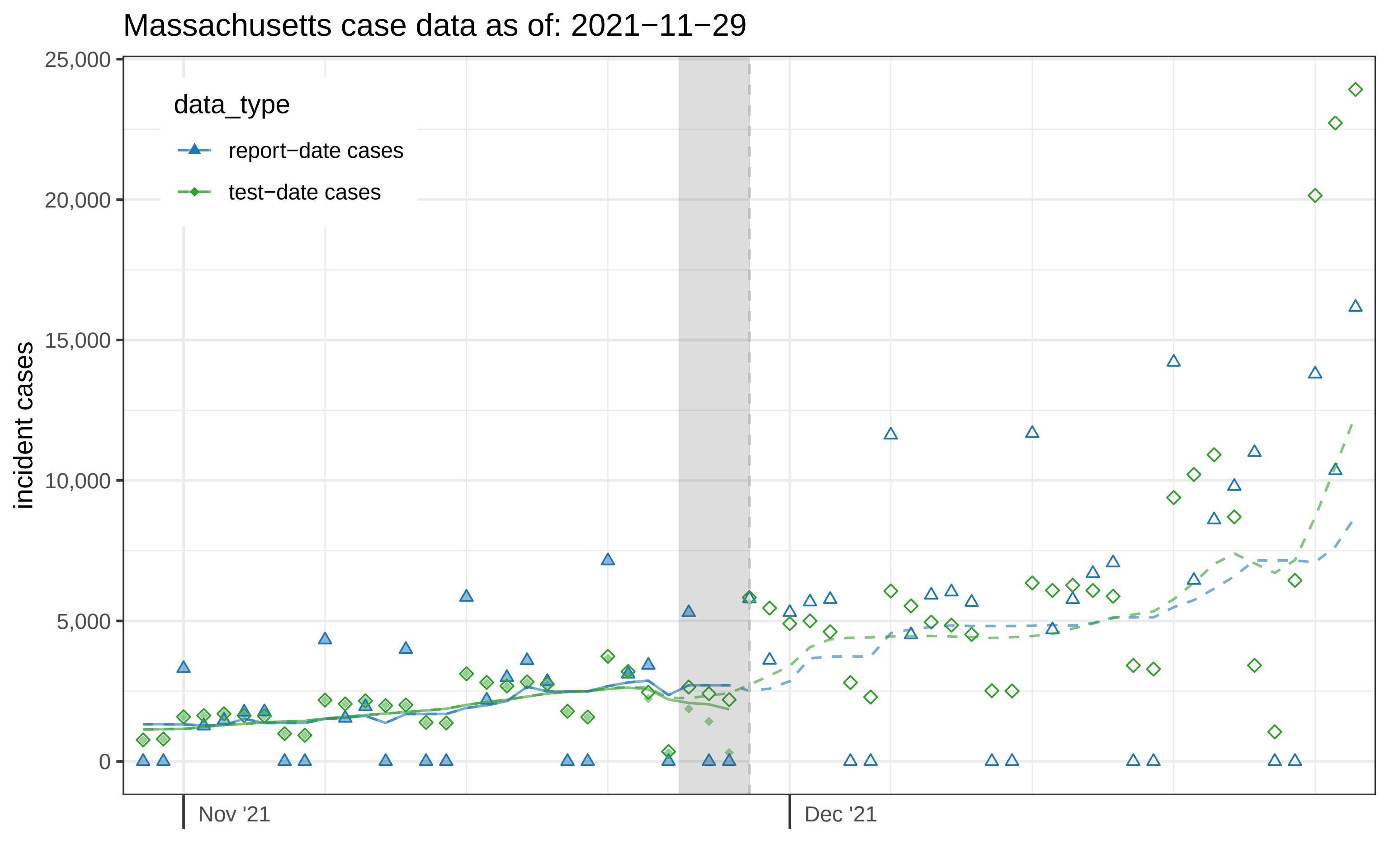

Massachusetts case data as of: 2021−11−29
data_type
report−date cases
test−date cases
incident cases
25,000
20,000
15,000
10,000
5,000
0
Nov '21
Dec '21

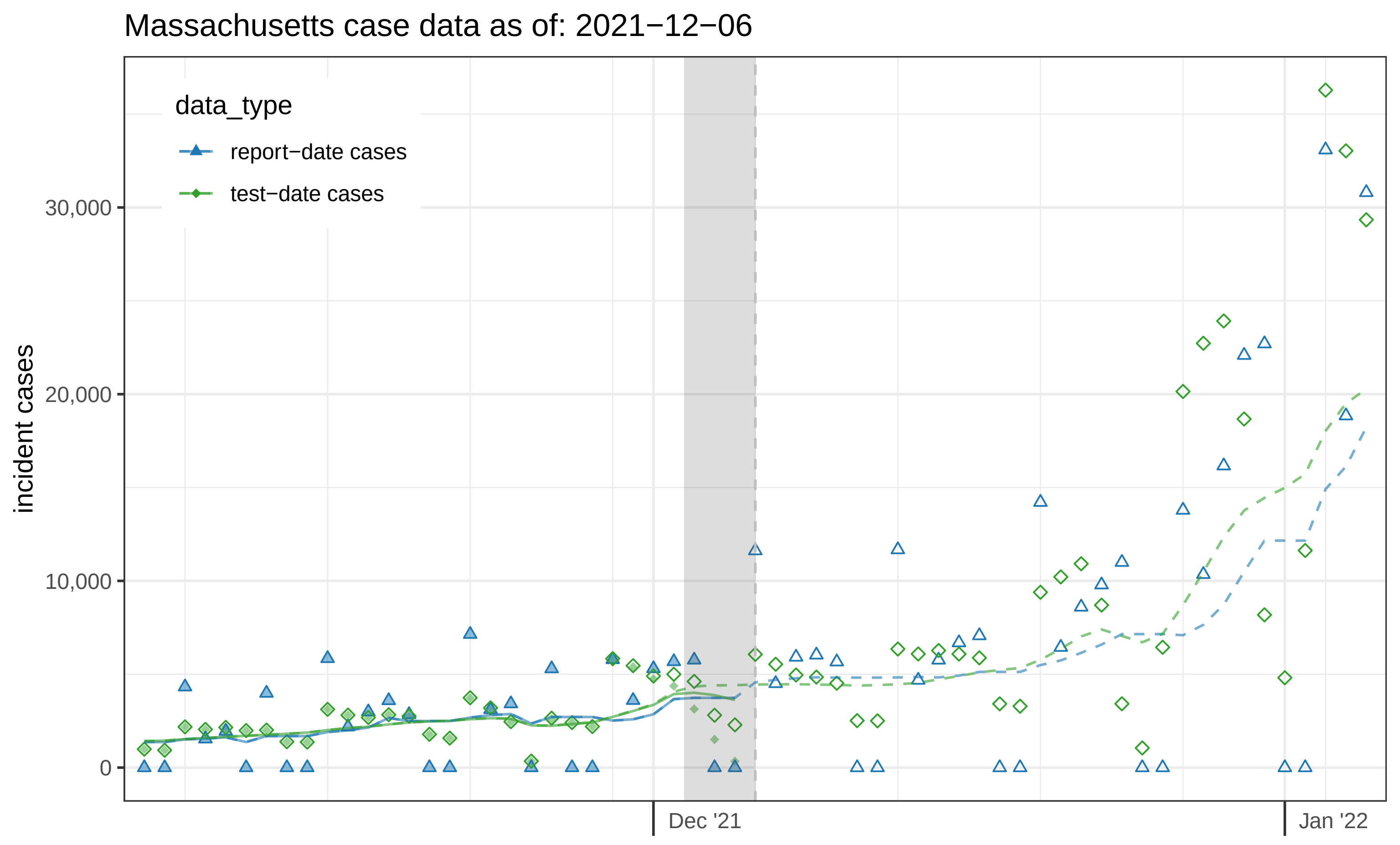
Massachusetts case data as of: 2021−12−06
data_type
report−date cases
test−date cases
incident cases
30,000
20,000
10,000
0
Dec '21
Jan '22

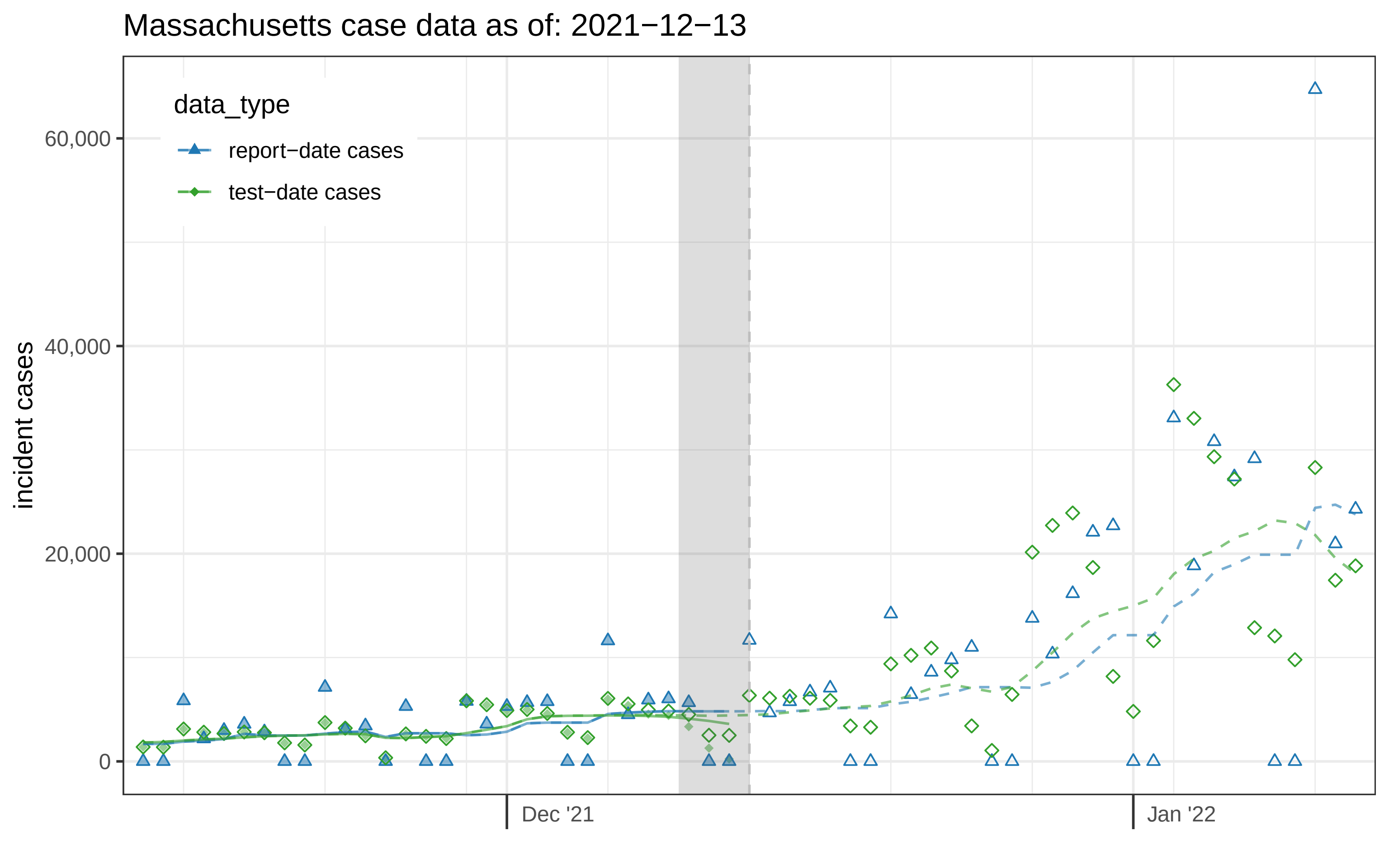

Massachusetts case data as of: 2021−12−13
data_type
report−date cases
test−date cases
incident cases
60,000
40,000
20,000
0
Dec '21
Jan '22

Massachusetts case data as of: 2021−12−20

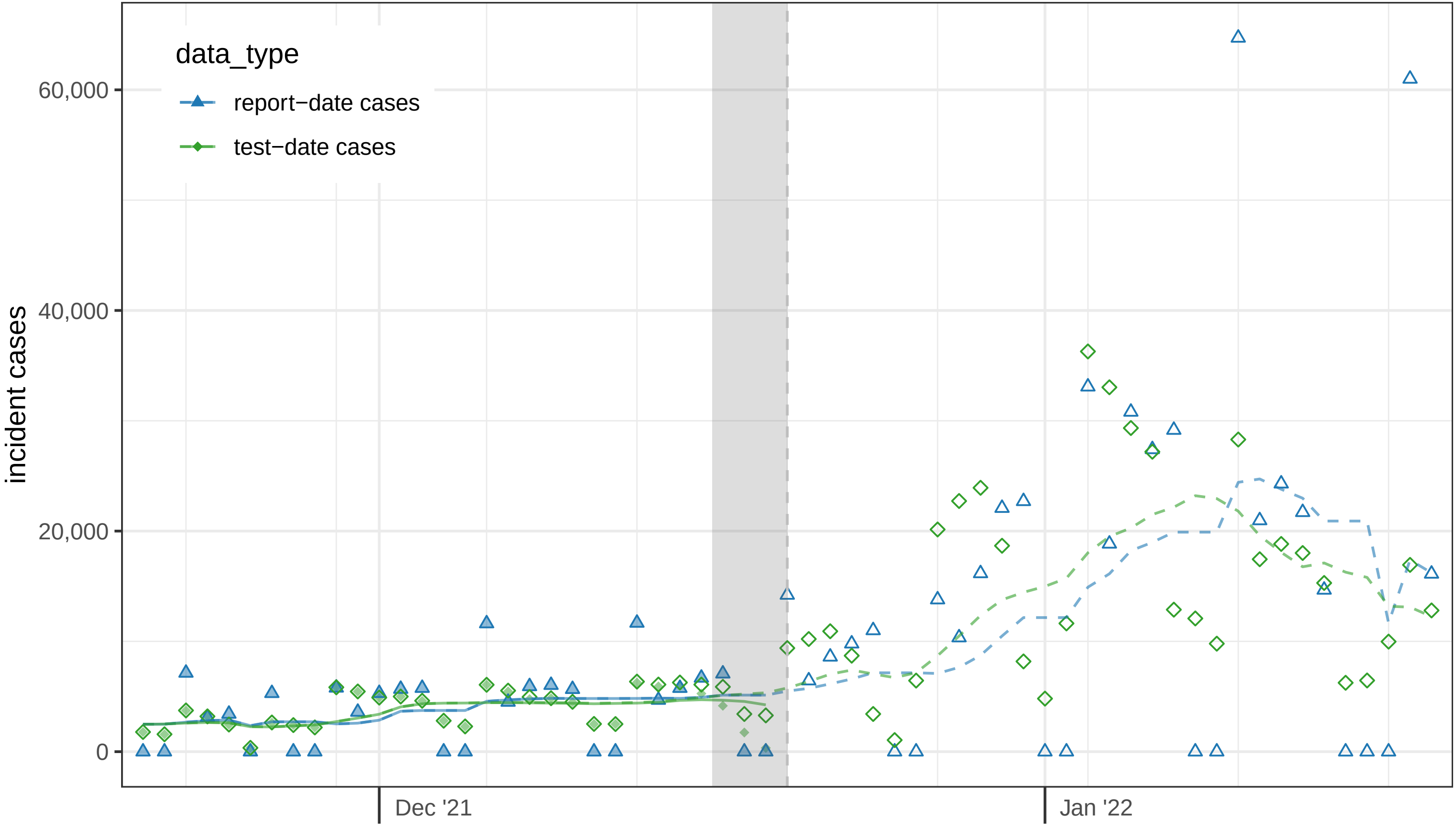

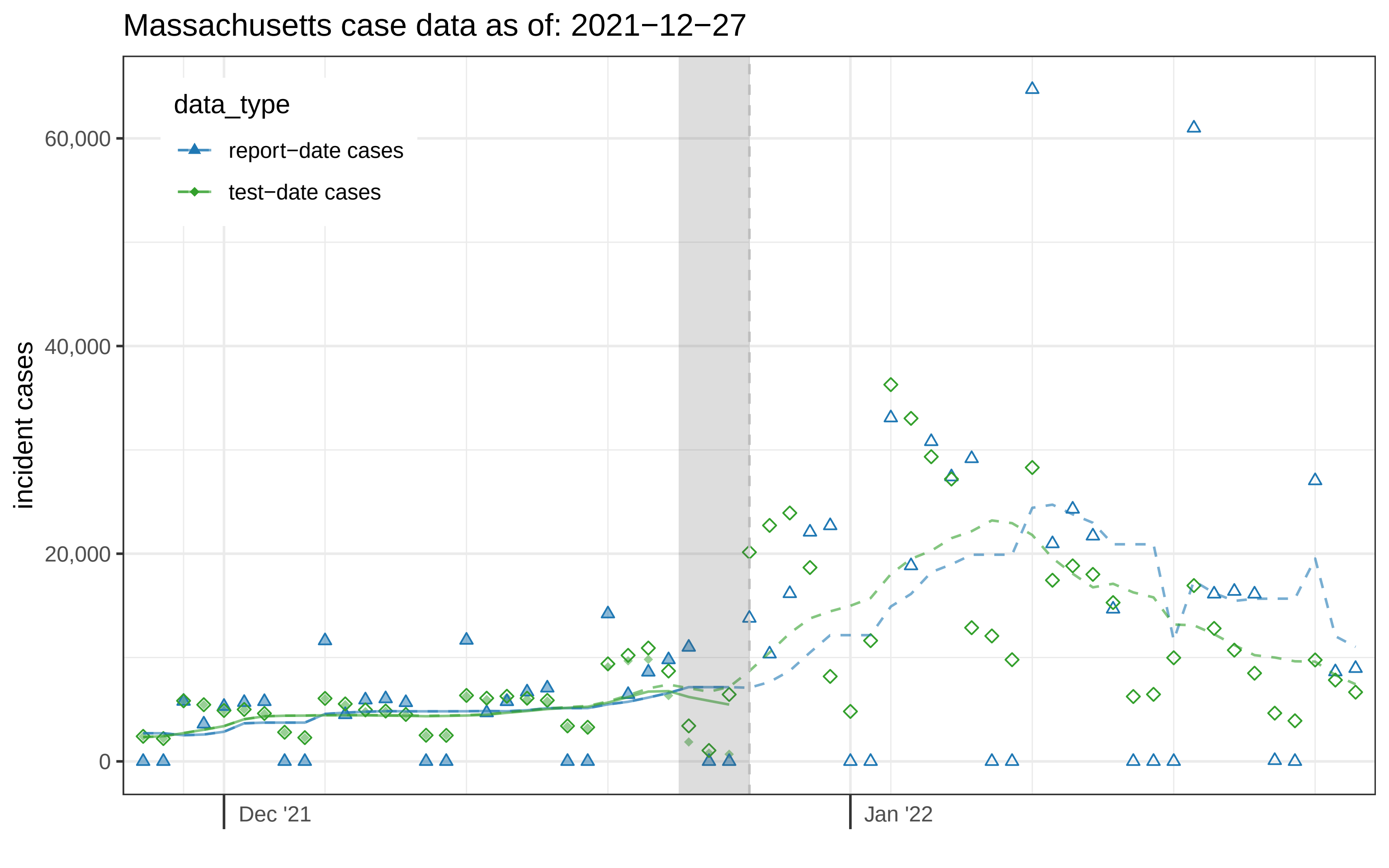

Massachusetts case data as of: 2021−12−27
data_type
report−date cases
test−date cases
60,000
40,000
20,000
0
incident cases
Dec '21
Jan '22

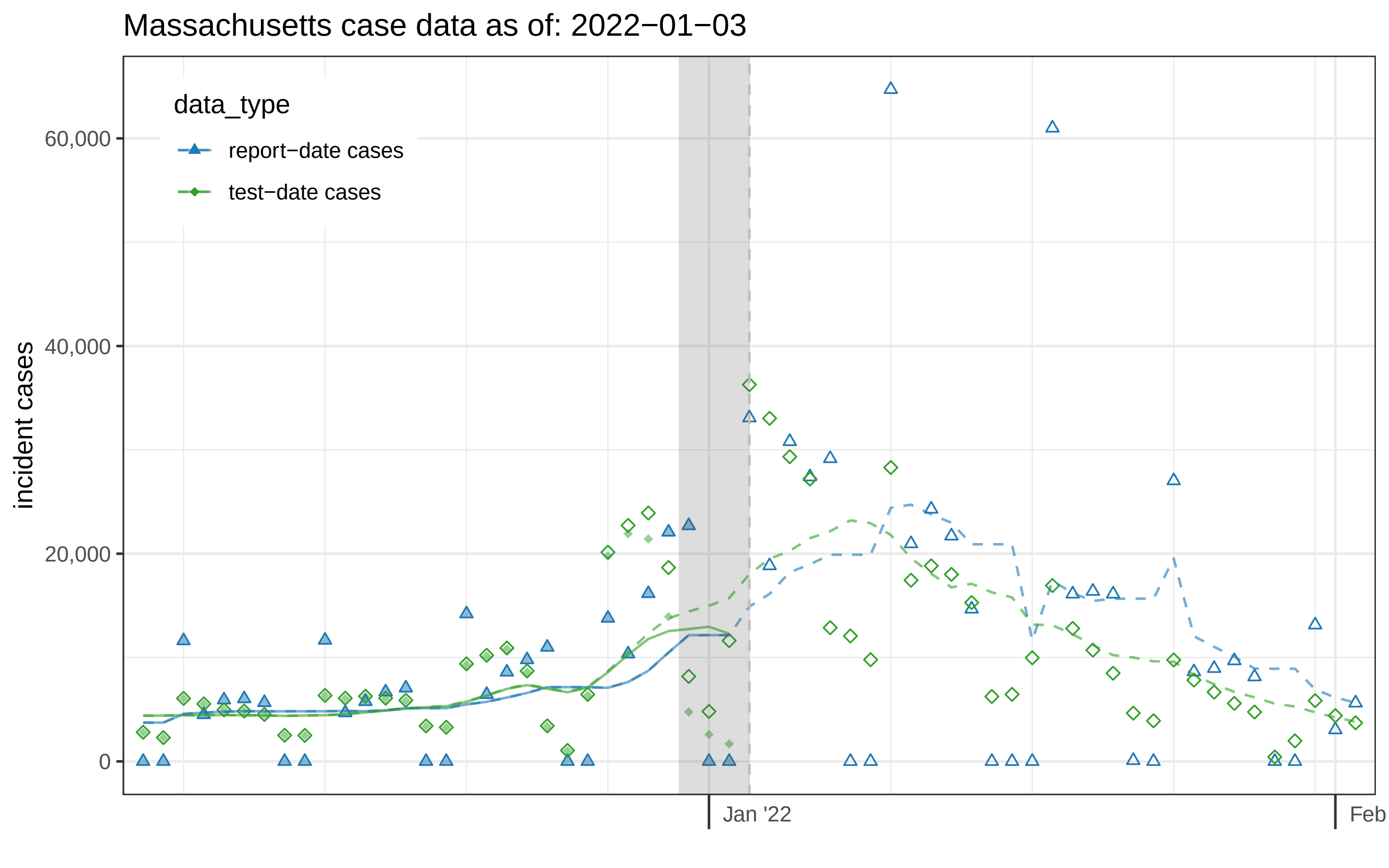

Massachusetts case data as of: 2022−01−03
data_type
report−date cases
test−date cases
incident cases
60,000
40,000
20,000
0
Jan '22
Feb

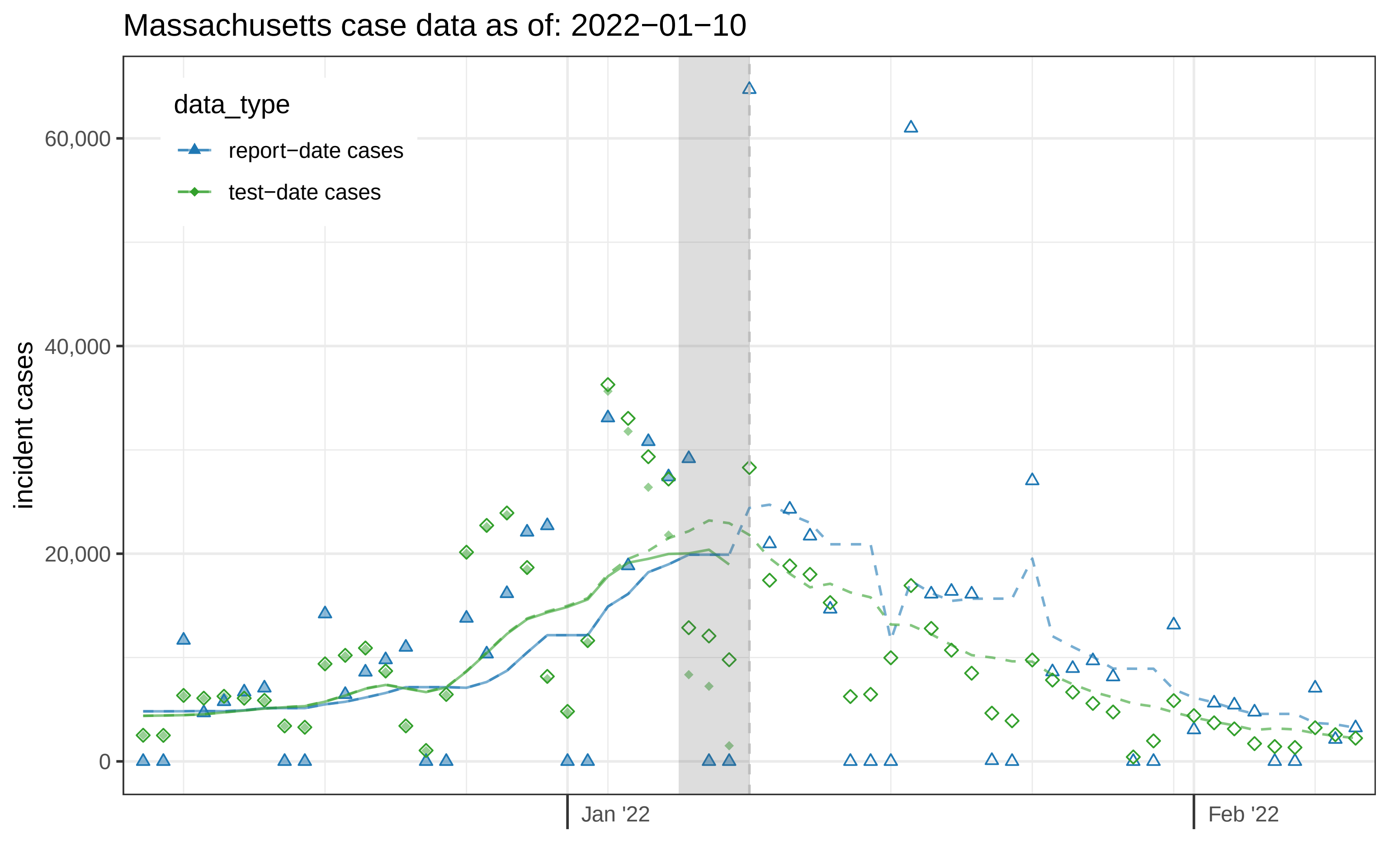

Massachusetts case data as of: 2022−01−10
data_type
report−date cases
test−date cases
incident cases
60,000
40,000
20,000
0
Jan '22
Feb '22

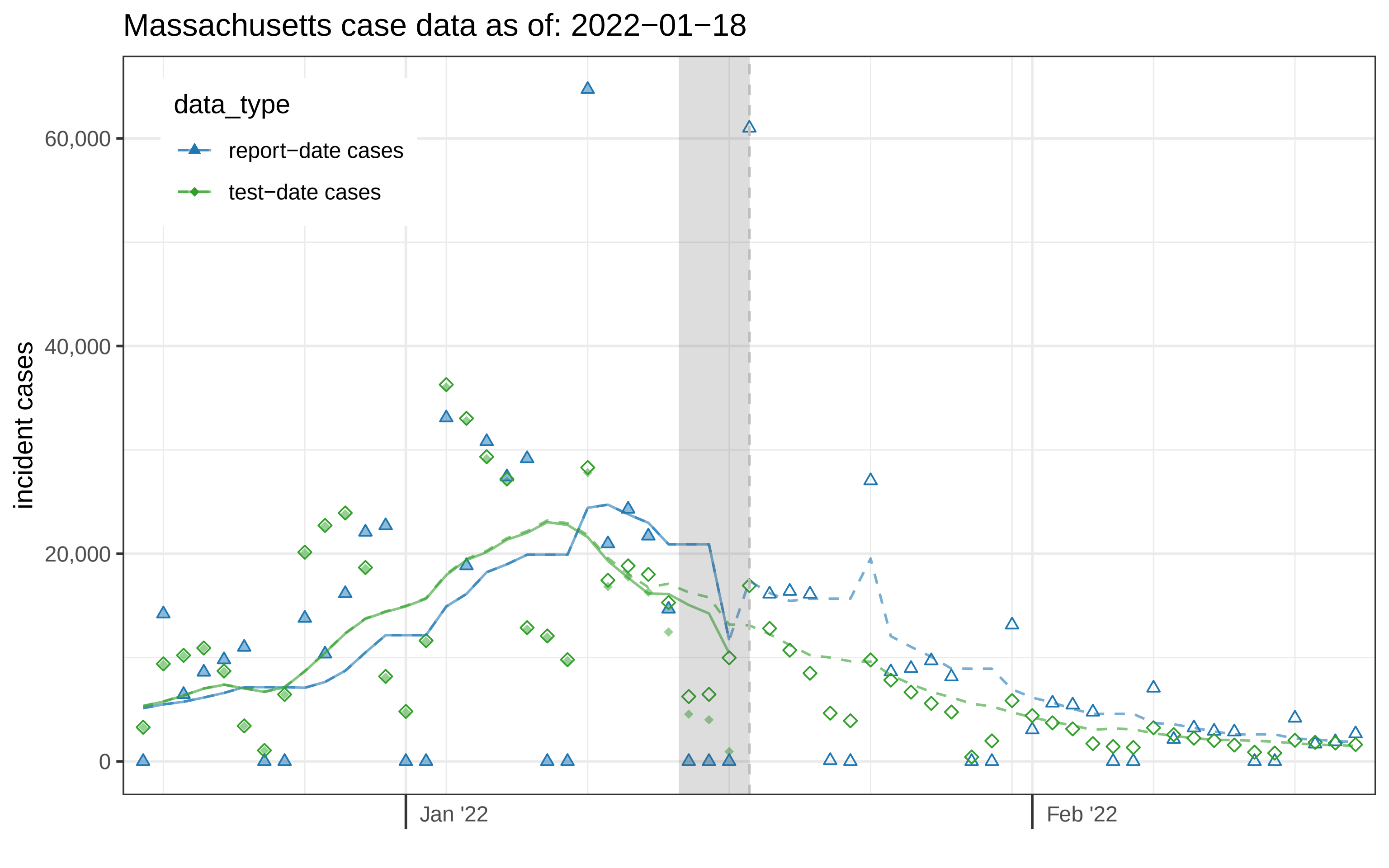

Massachusetts case data as of: 2022−01−18
data_type
report−date cases
test−date cases
incident cases
60,000
40,000
20,000
0
Jan '22
Feb '22

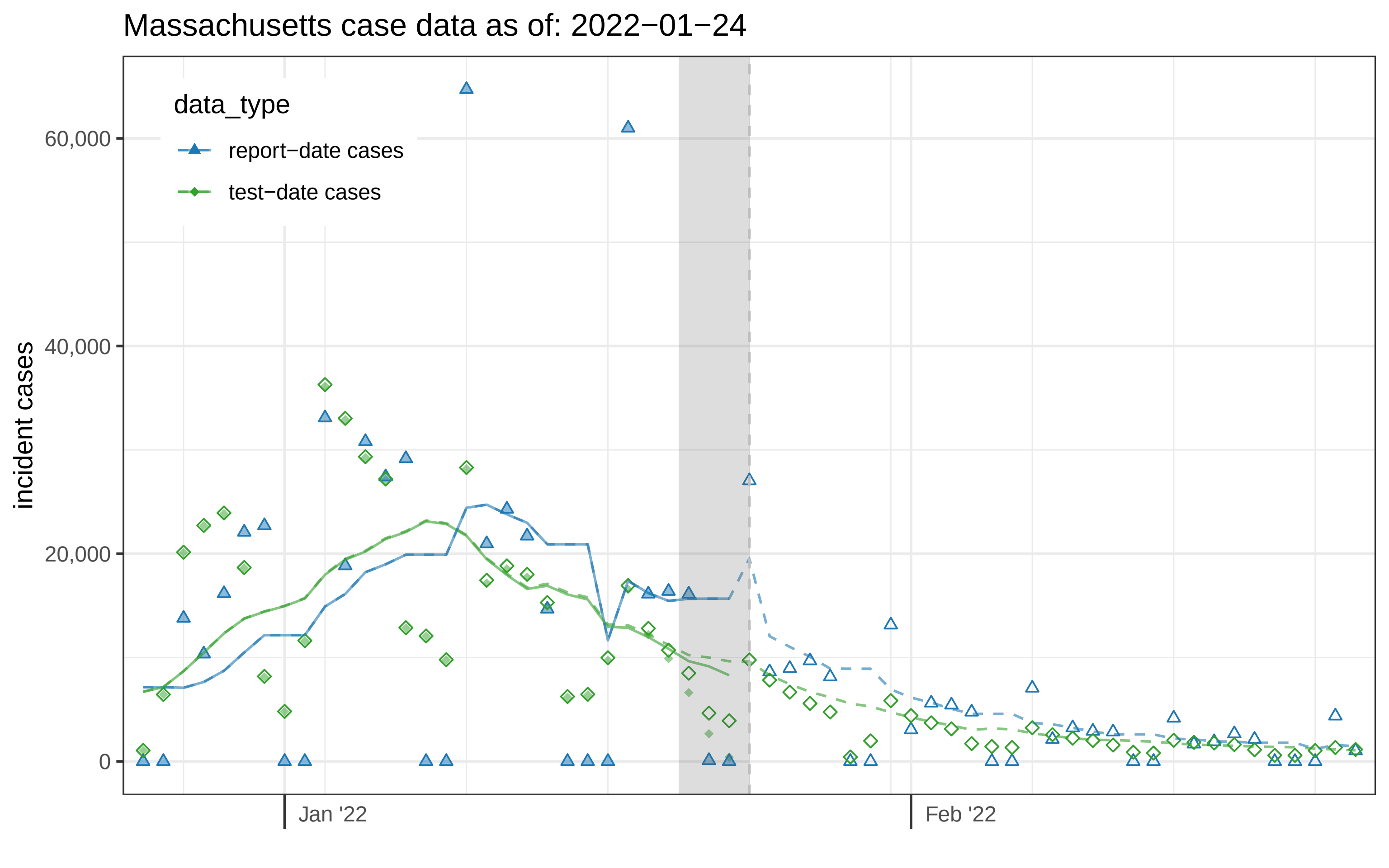
Massachusetts case data as of: 2022−01−24
data_type
report−date cases
test−date cases
incident cases
60,000
40,000
20,000
0
Jan '22
Feb '22

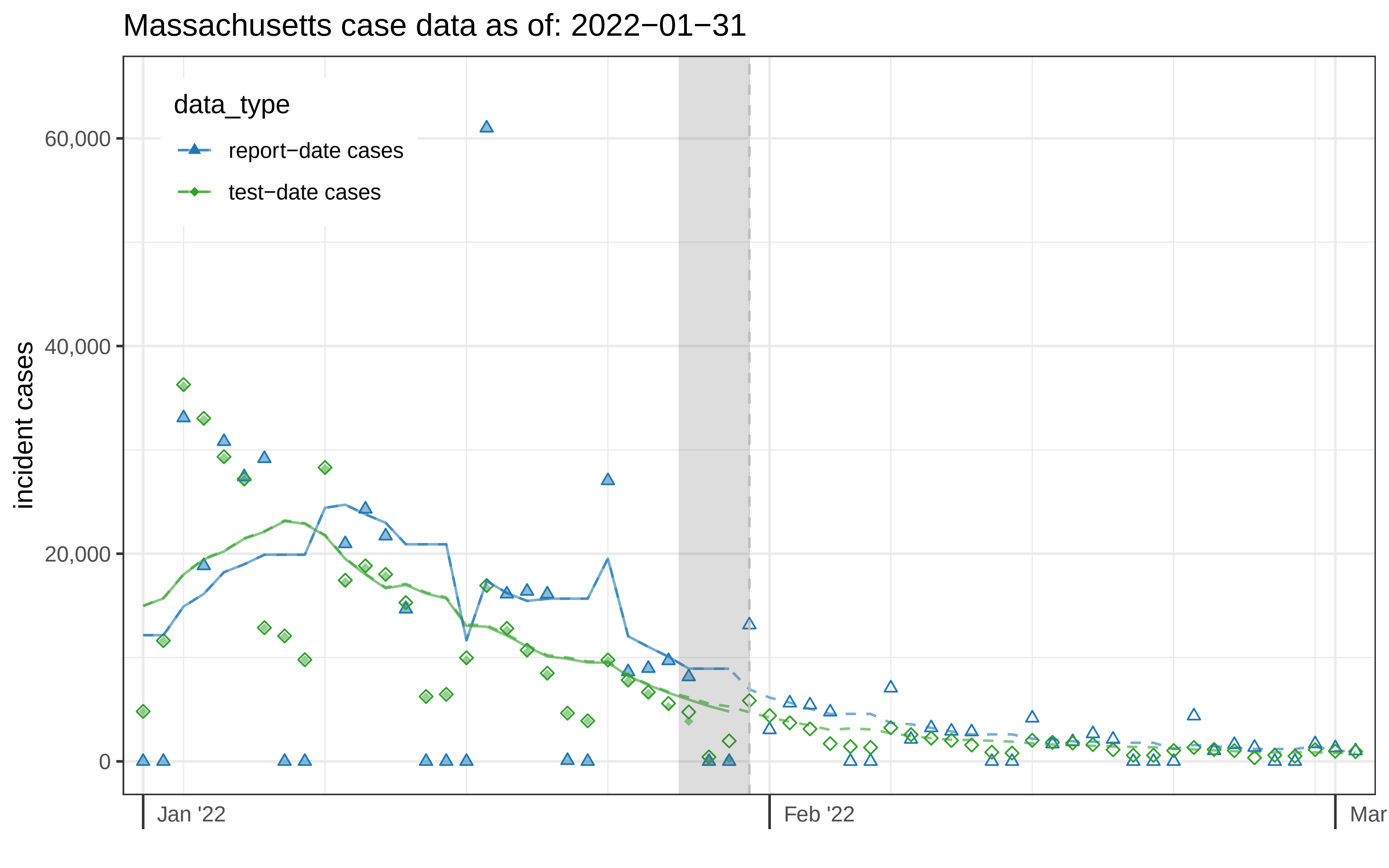

Massachusetts case data as of: 2022−01−31
data_type
report−date cases
test−date cases
incident cases
60,000
40,000
20,000
0
Jan '22
Feb '22
Mar

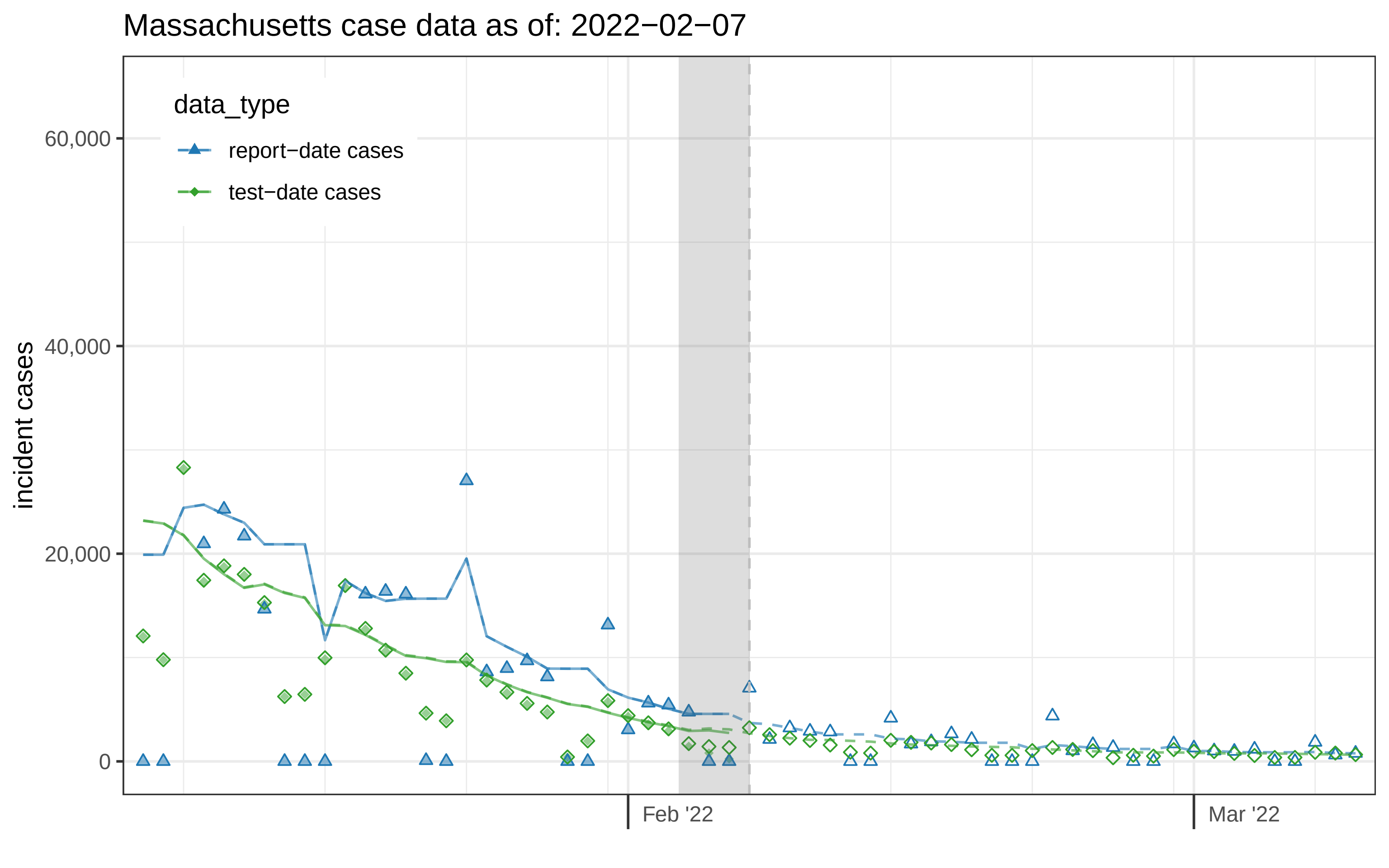
Massachusetts case data as of: 2022−02−07
data_type
report−date cases
test−date cases
incident cases
60,000
40,000
20,000
0
Feb '22
Mar '22

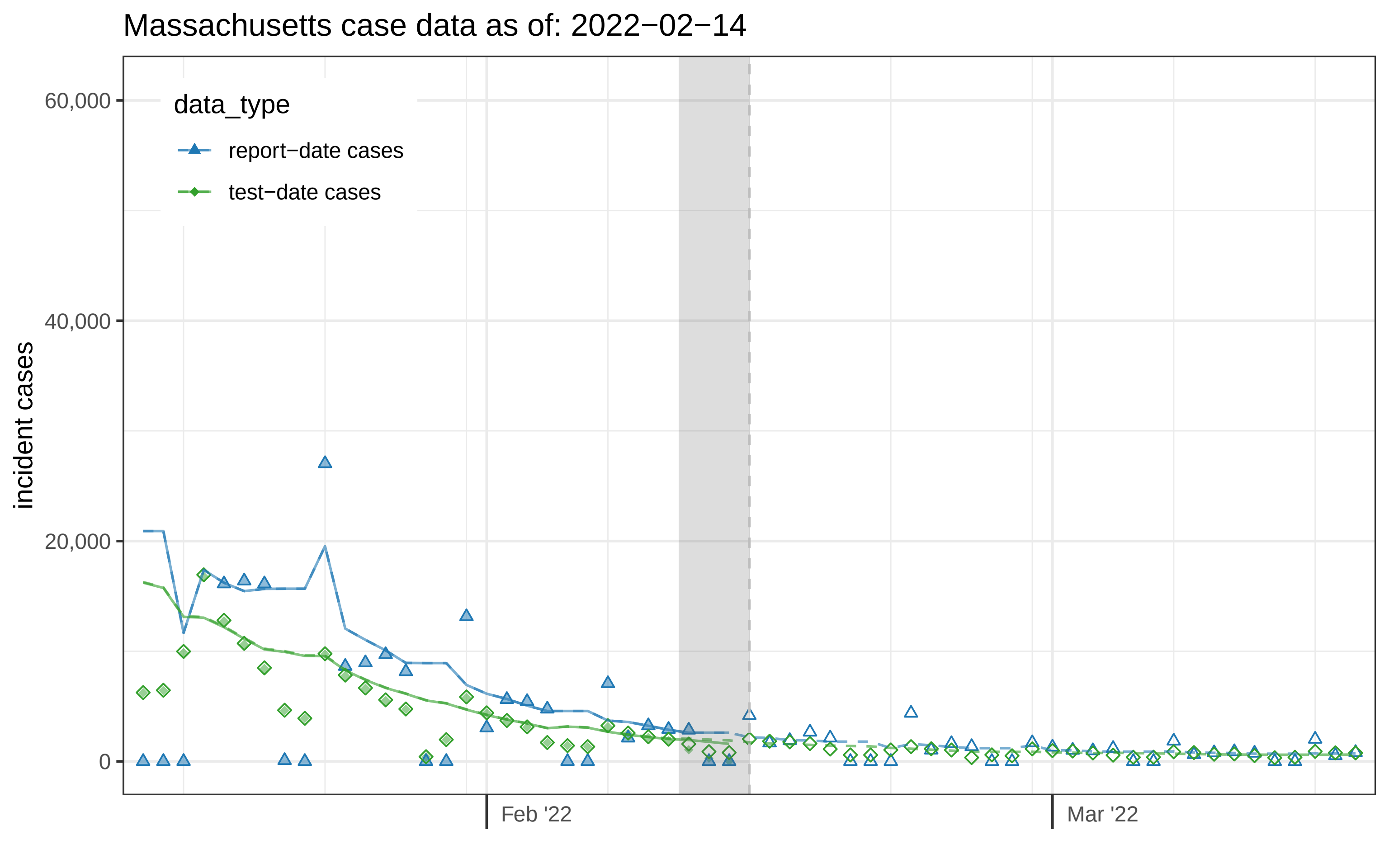
Massachusetts case data as of: 2022−02−14
incident cases
60,000
40,000
20,000
0
data_type
report−date cases
test−date cases
Feb '22
Mar '22

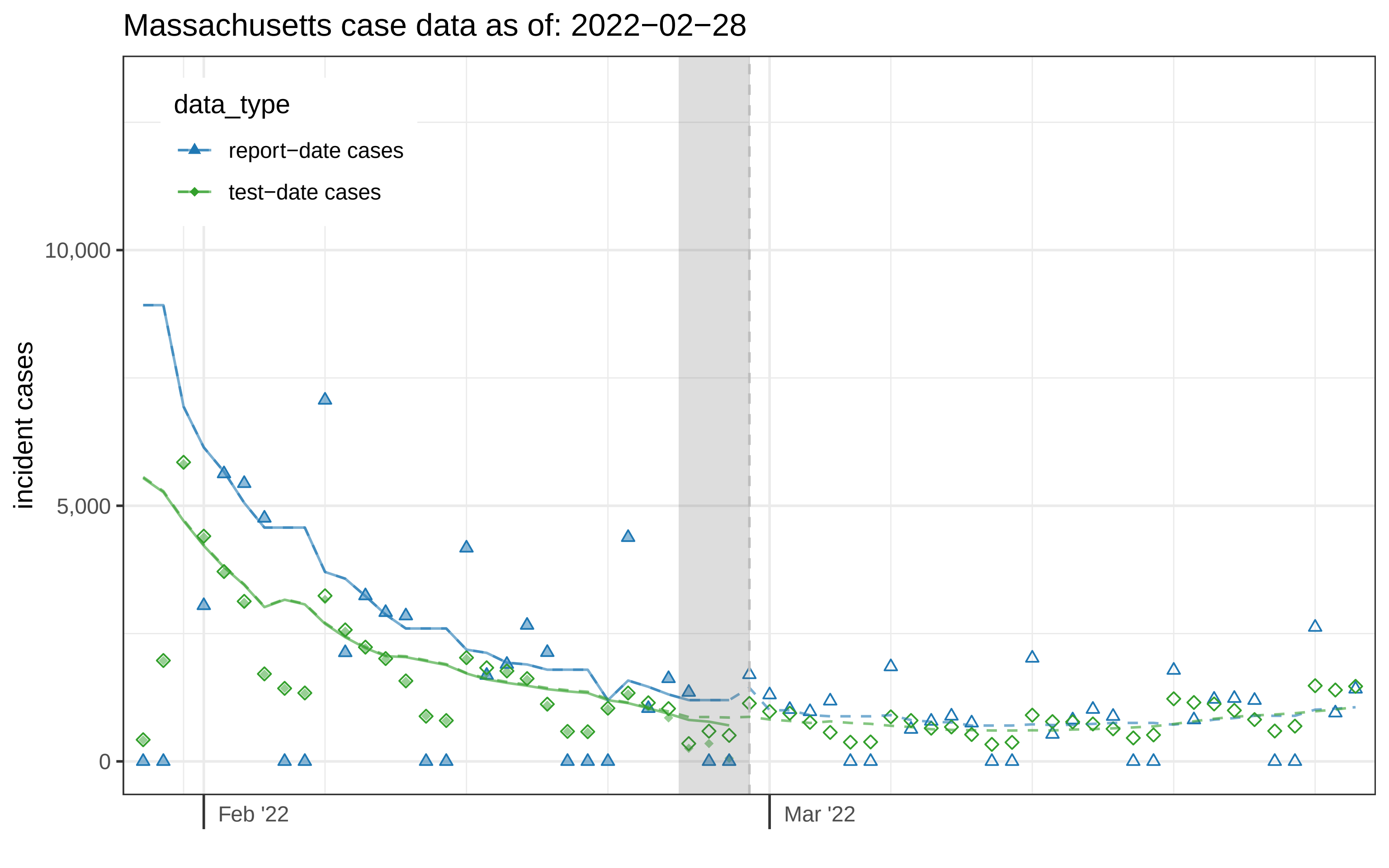
Massachusetts case data as of: 2022−02−28
data_type
report−date cases
test−date cases
incident cases
10,000
5,000
0
Feb '22
Mar '22

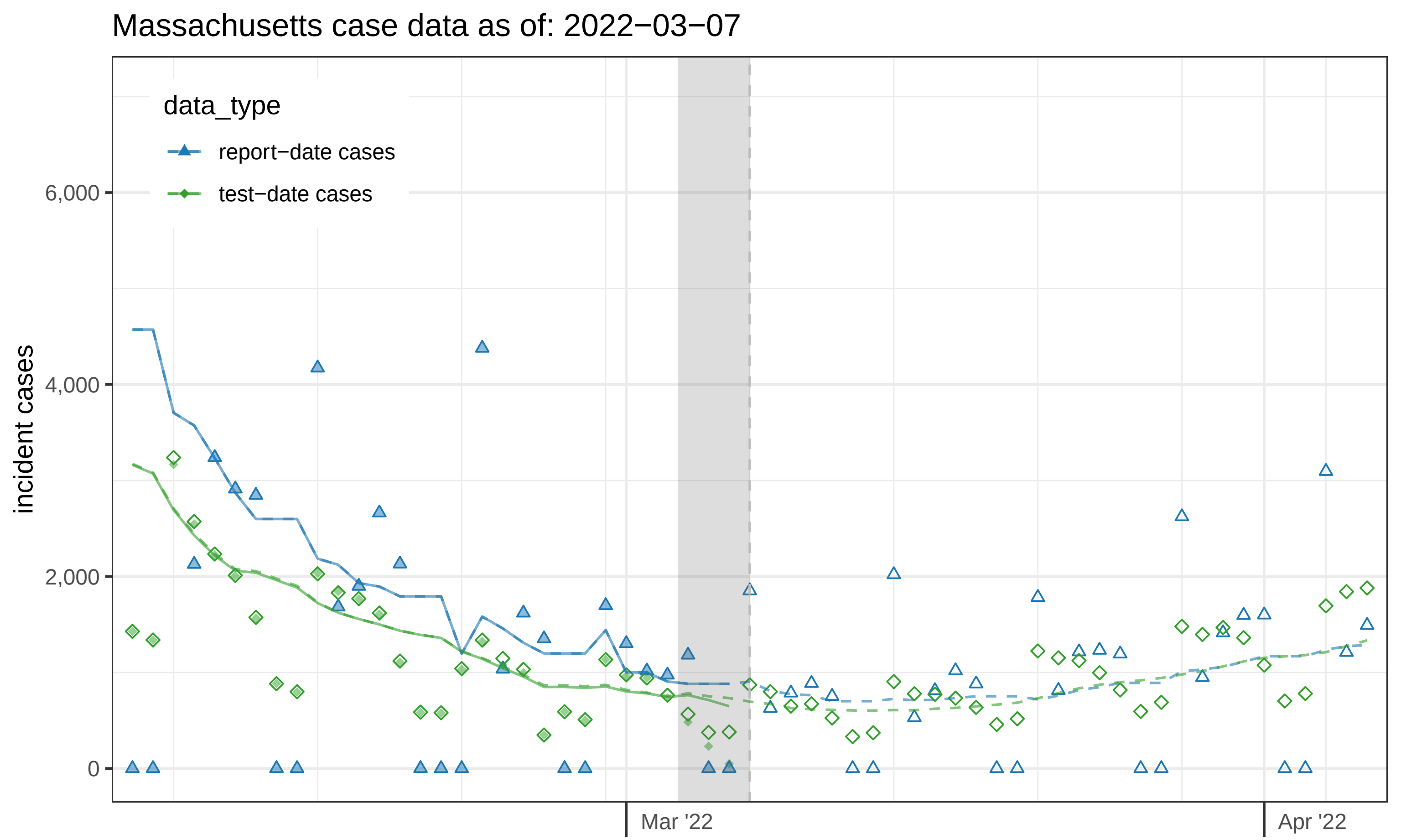

Massachusetts case data as of: 2022−03−07
data_type
report−date cases
test−date cases
incident cases
6,000
4,000
2,000
0
Mar '22
Apr '22

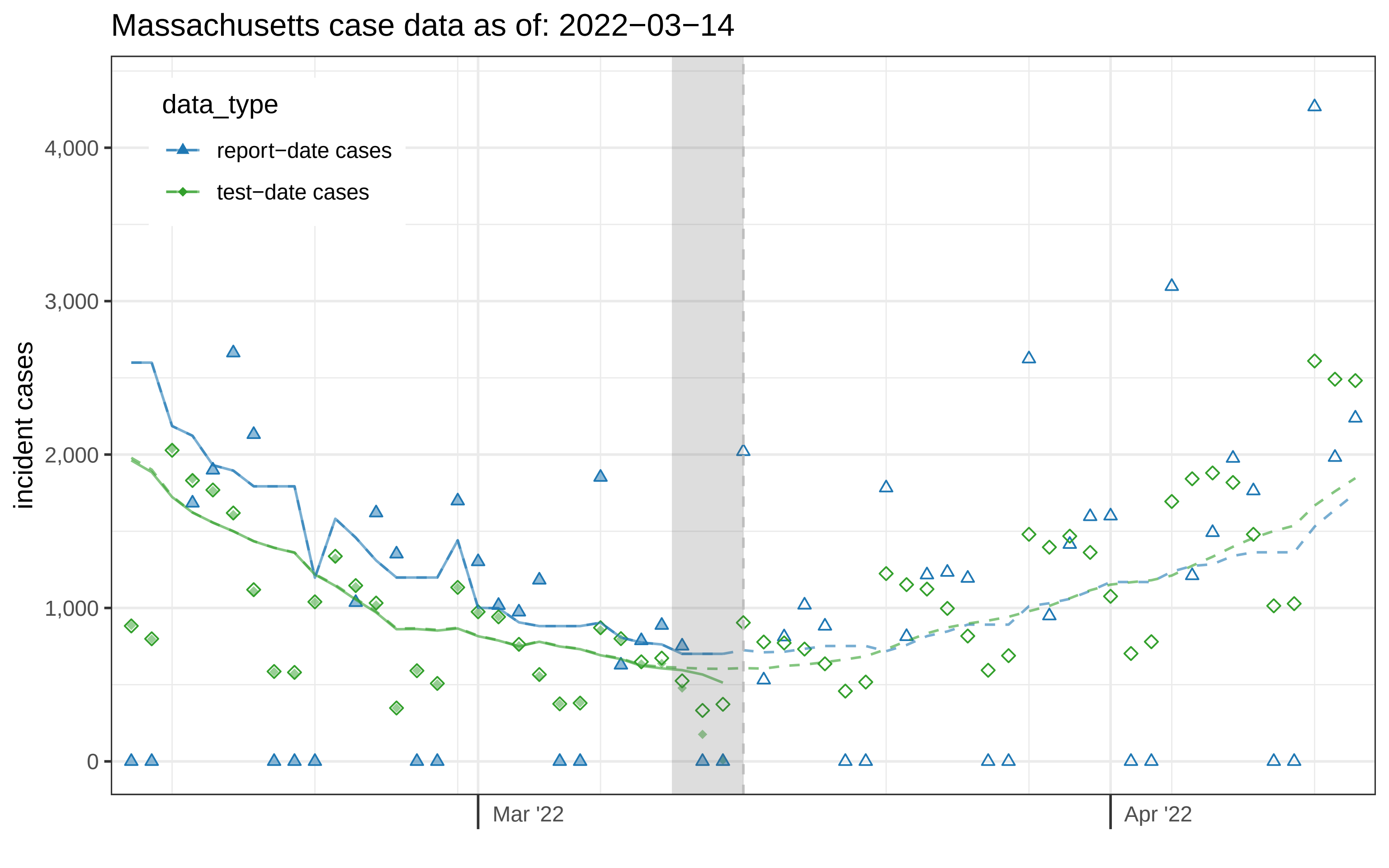

Massachusetts case data as of: 2022−03−14
data_type
report−date cases
test−date cases
incident cases
4,000
3,000
2,000
1,000
0
Mar '22
Apr '22

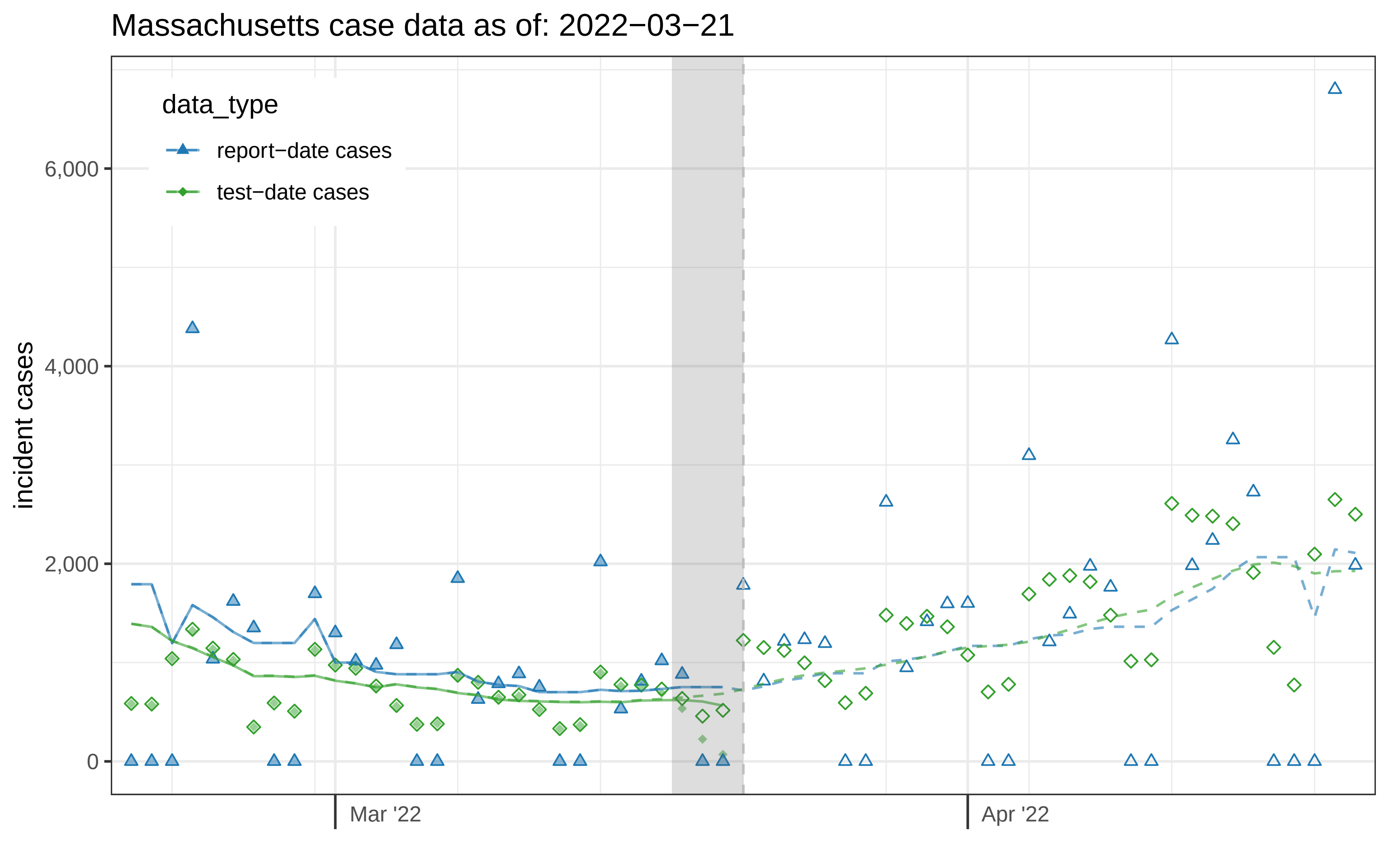

Massachusetts case data as of: 2022−03−21
data_type
report−date cases
test−date cases
incident cases
6,000
4,000
2,000
0
Mar '22
Apr '22

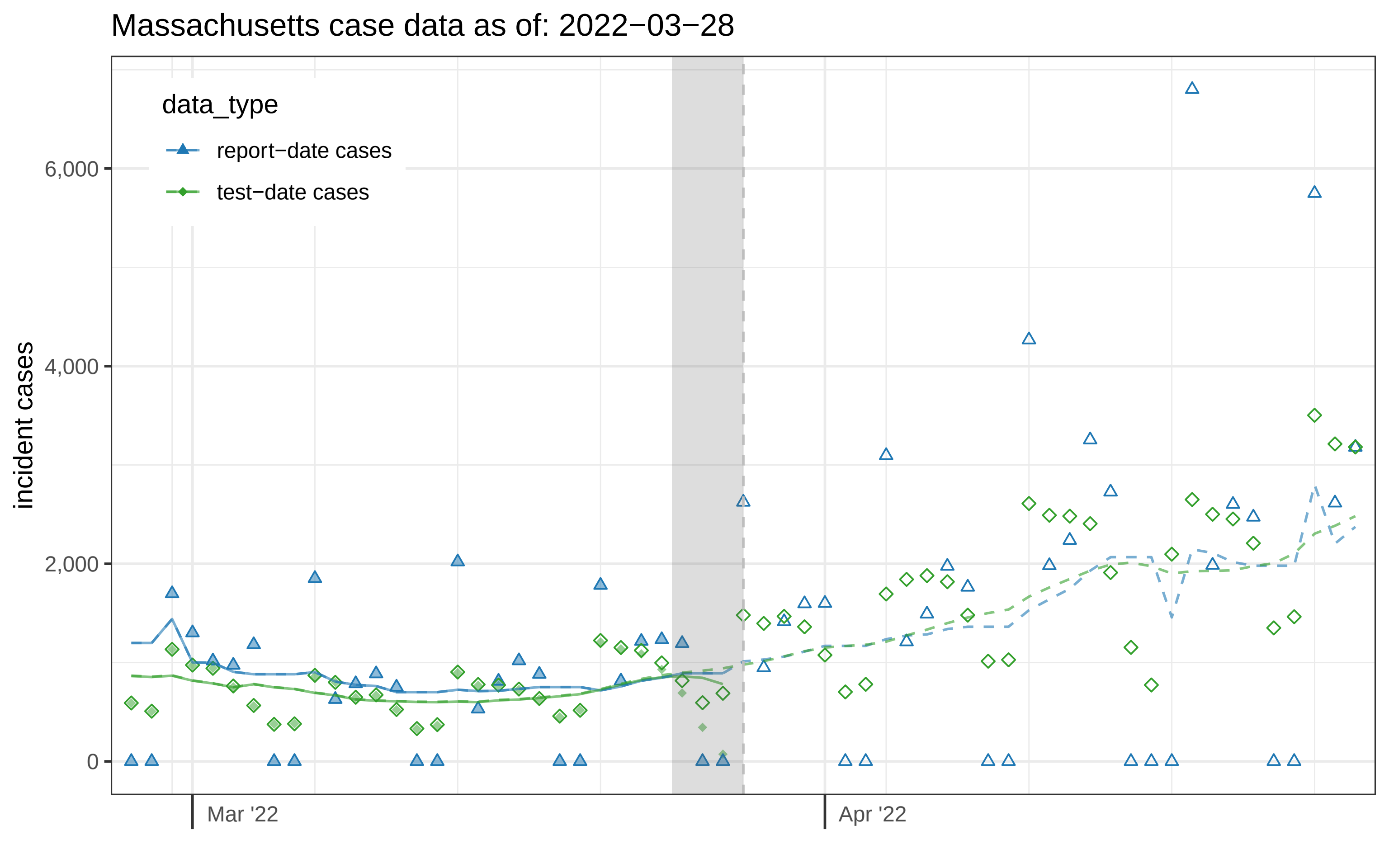
Massachusetts case data as of: 2022−03−28
data_type
report−date cases
test−date cases
incident cases
6,000
4,000
2,000
0
Mar '22
Apr '22

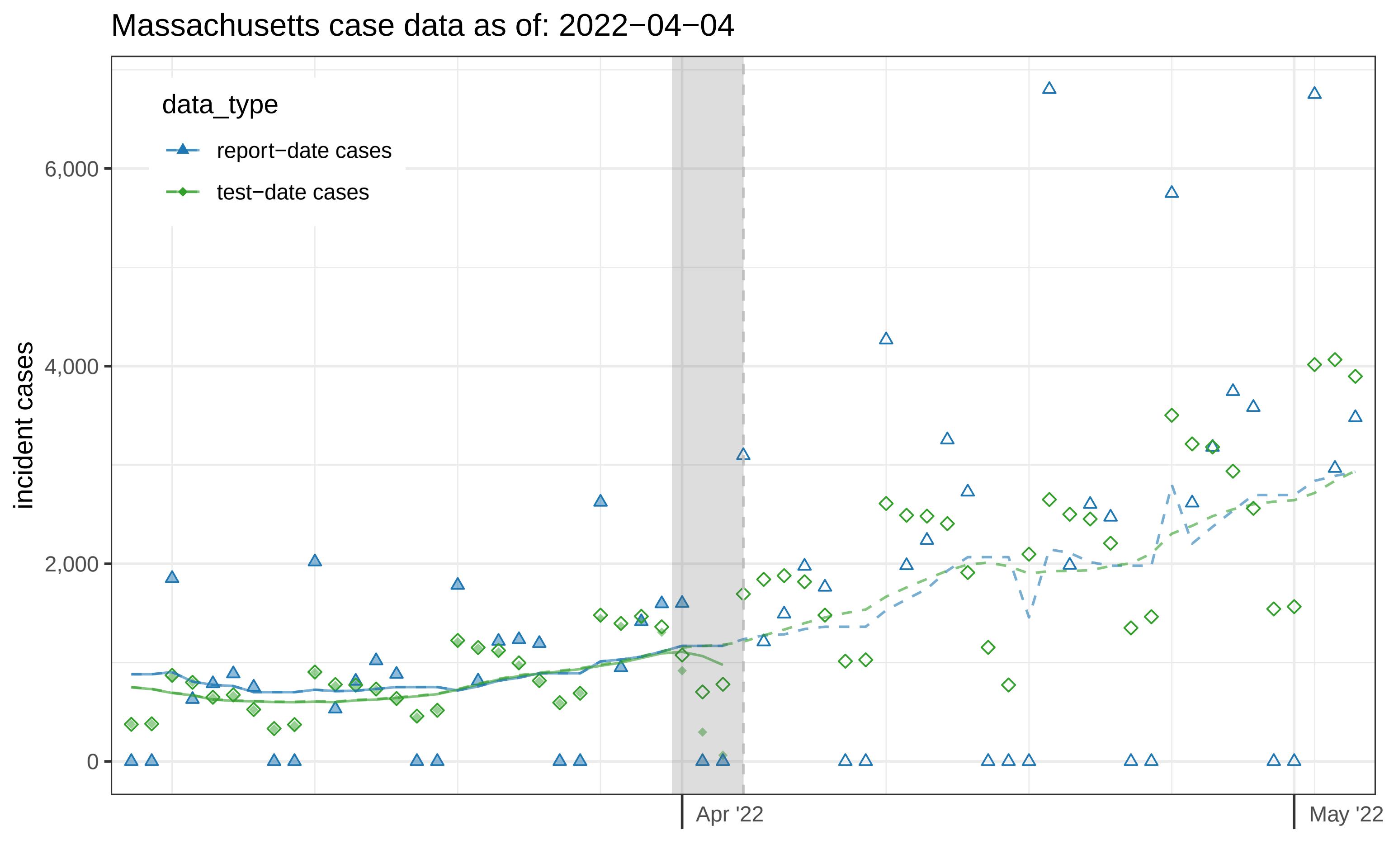

Massachusetts case data as of: 2022−04−04
data_type
report−date cases
test−date cases
incident cases
6,000
4,000
2,000
0
Apr '22
May '22

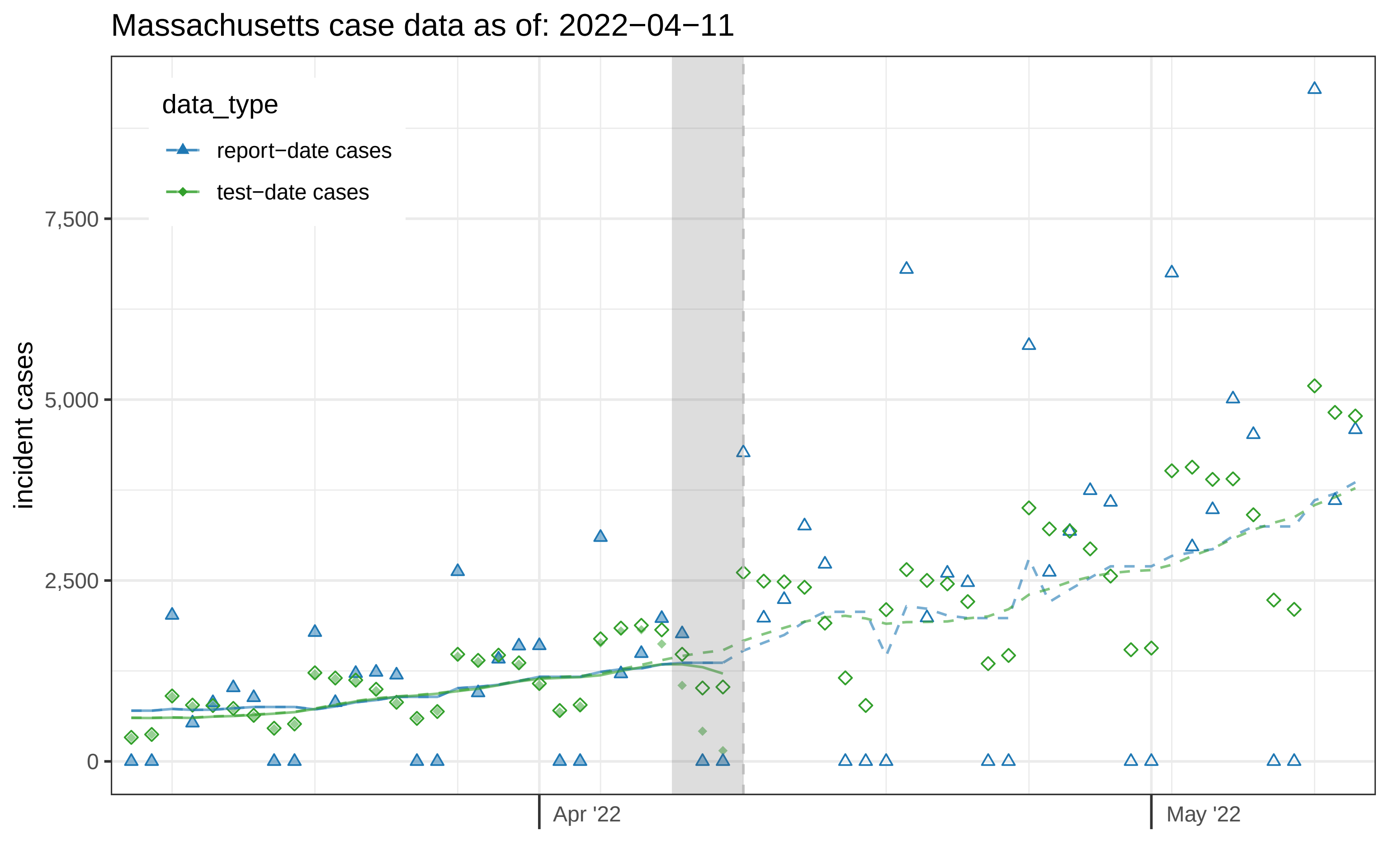
Massachusetts case data as of: 2022−04−11
data_type
report−date cases
test−date cases
incident cases
7,500
5,000
2,500
0
Apr '22
May '22

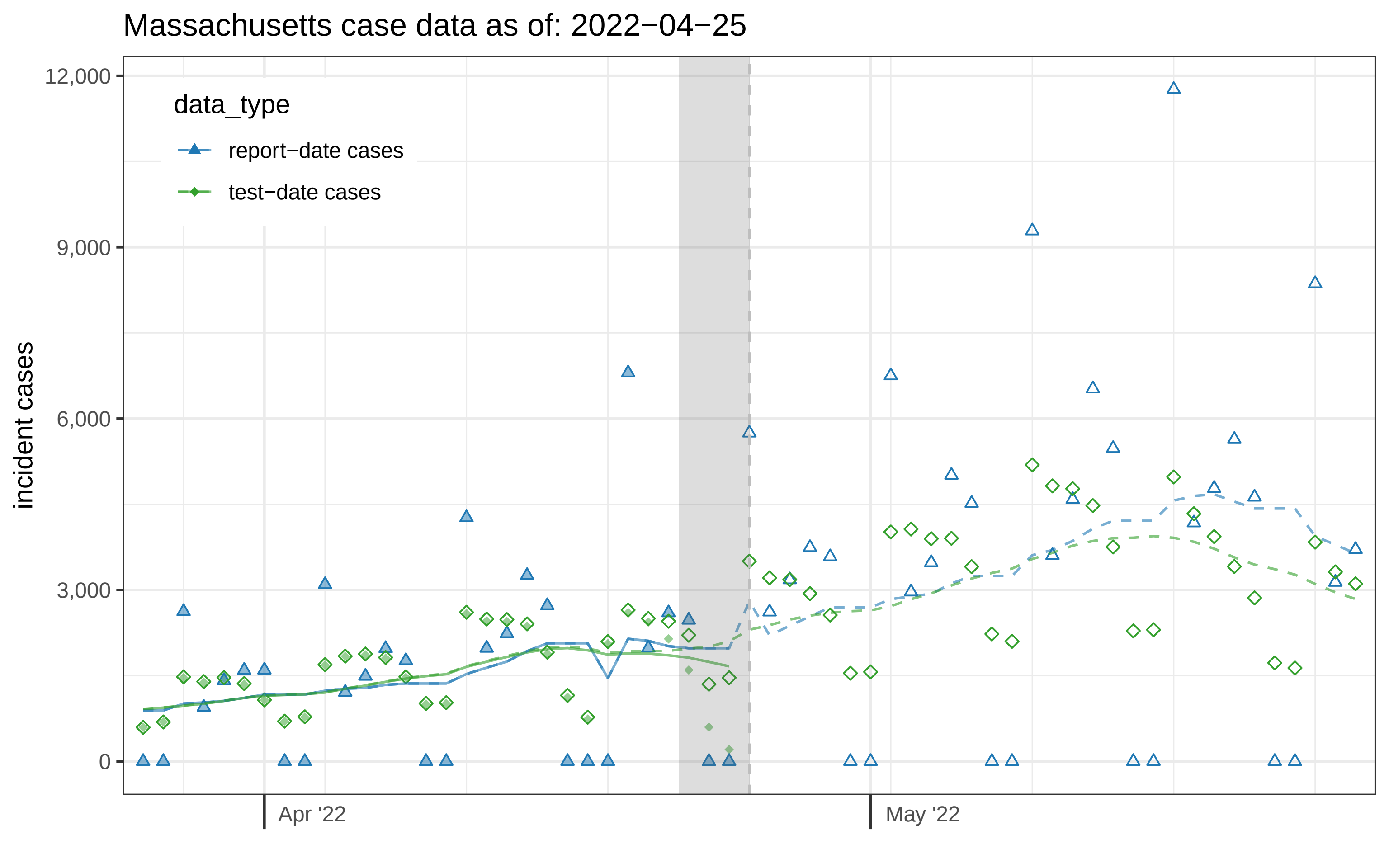

Massachusetts case data as of: 2022−04−25
data_type
report−date cases
test−date cases
incident cases
12,000
9,000
6,000
3,000
0
Apr '22
May '22

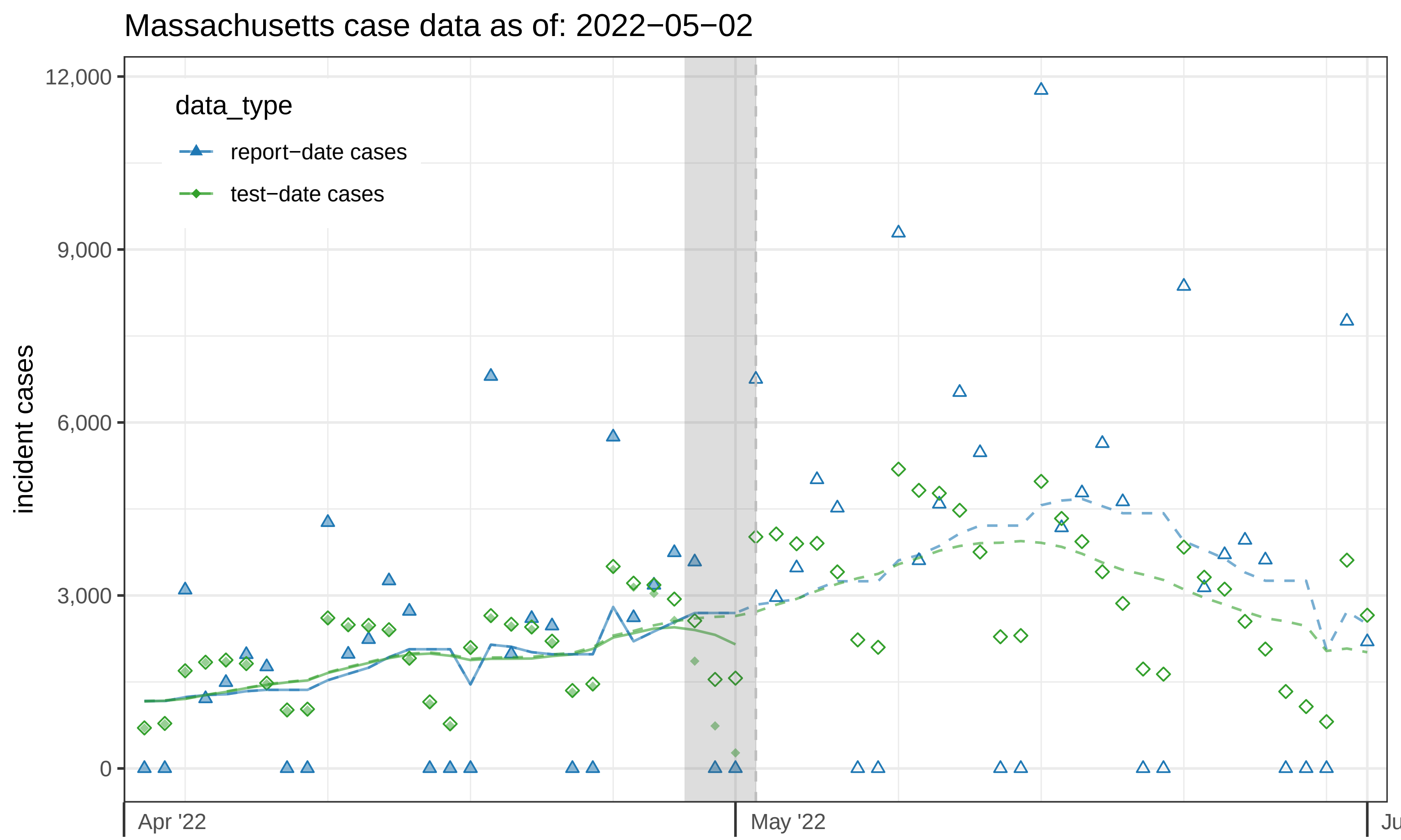

Massachusetts case data as of: 2022−05−02
data_type
report−date cases
test−date cases
incident cases
12,000
9,000
6,000
3,000
0
Apr '22
May '22
Ju

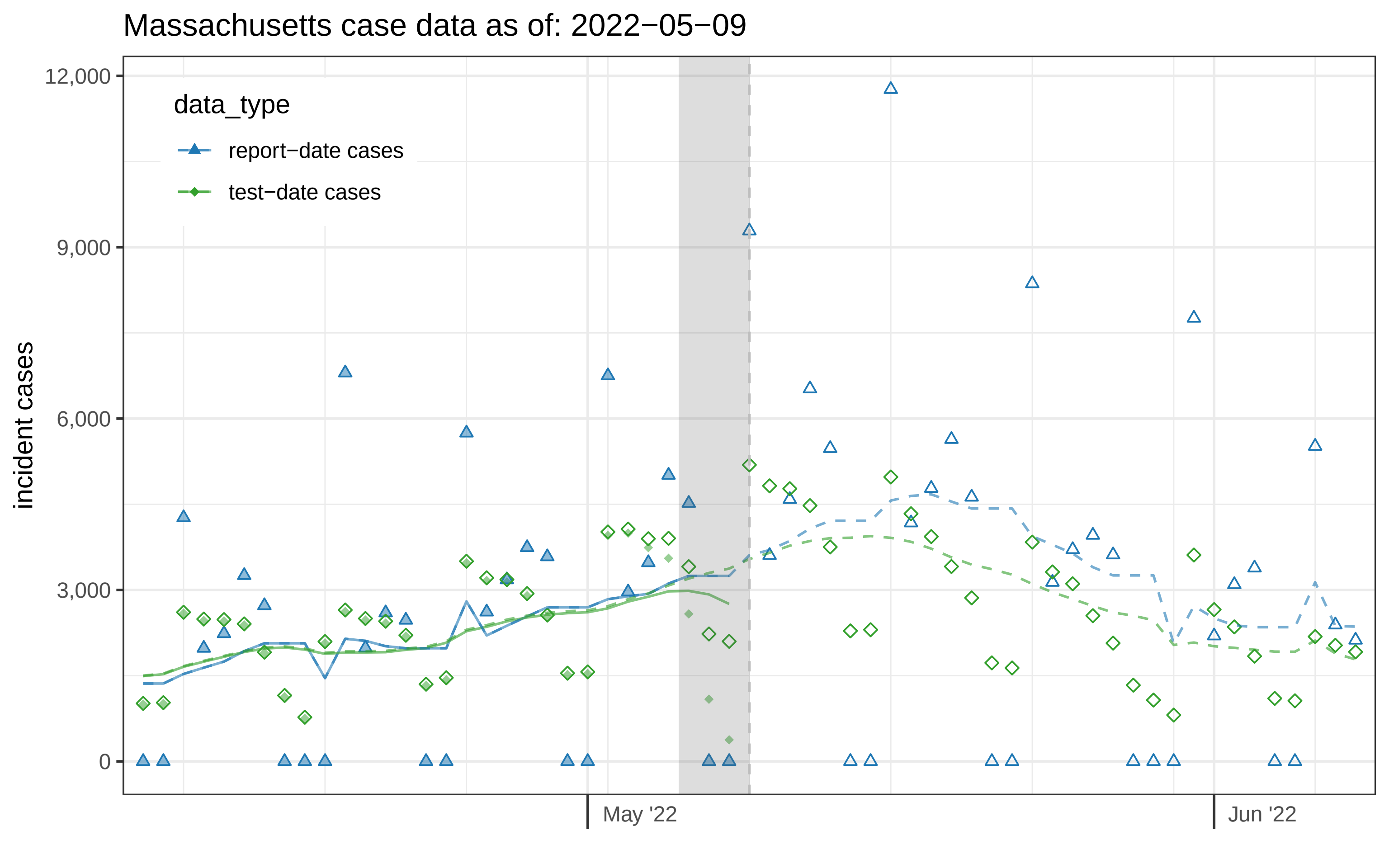

Massachusetts case data as of: 2022−05−09
data_type
report−date cases
test−date cases
incident cases
12,000
9,000
6,000
3,000
0
May '22
Jun '22

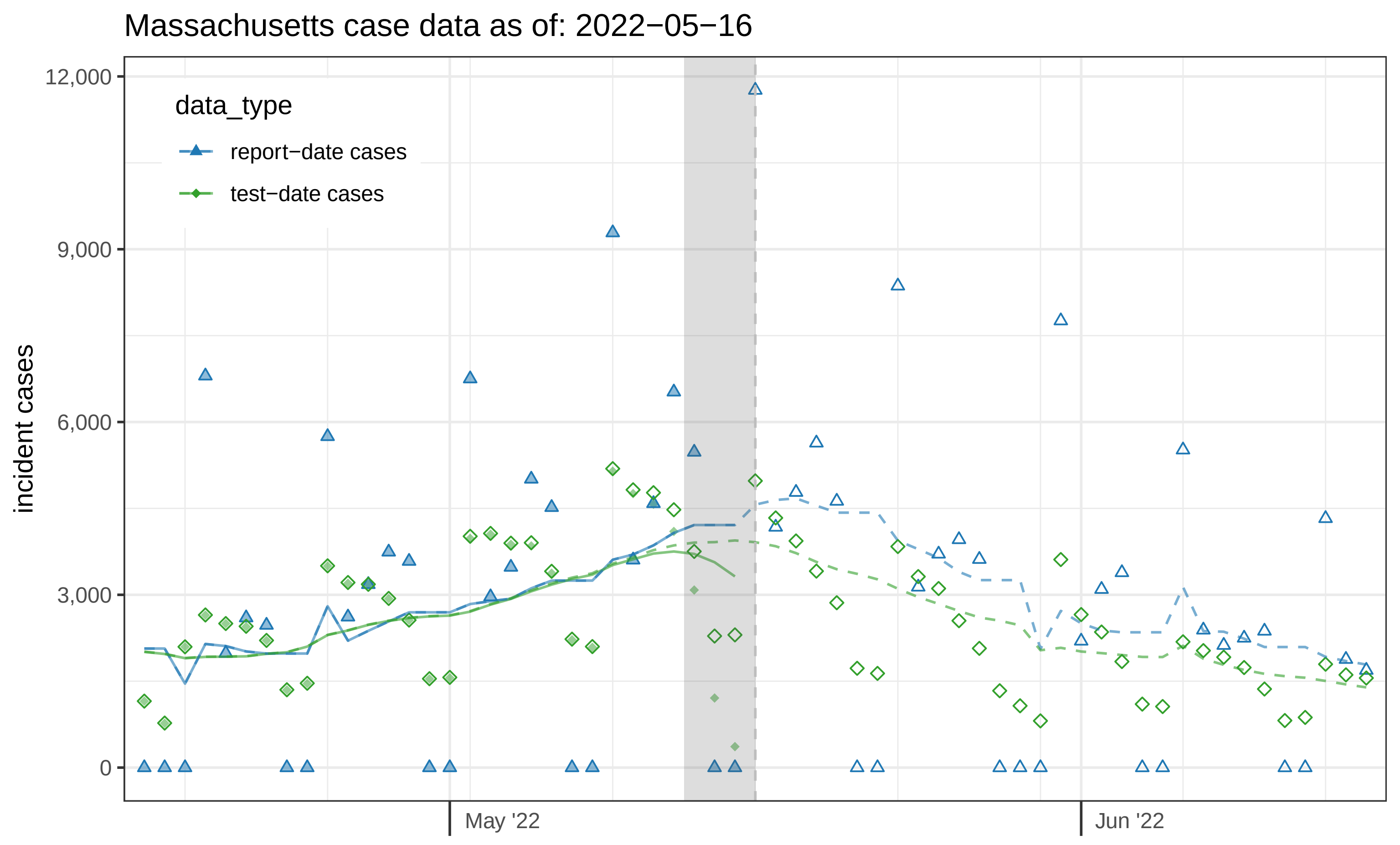
Massachusetts case data as of: 2022−05−16
data_type
report−date cases
test−date cases
incident cases
12,000
9,000
6,000
3,000
0
May '22
Jun '22

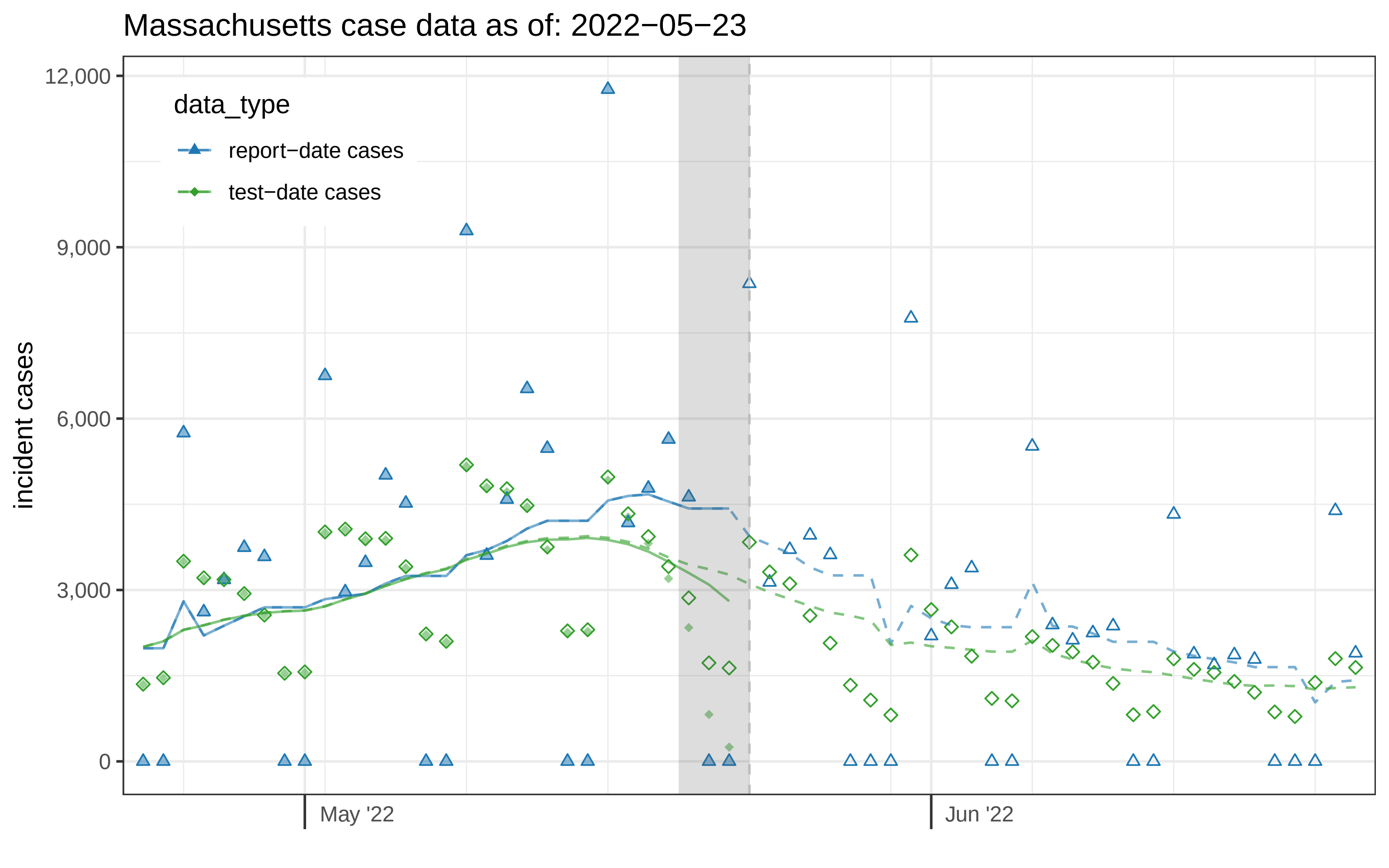

Massachusetts case data as of: 2022−05−23
data_type
report−date cases
test−date cases
incident cases
12,000
9,000
6,000
3,000
0
May '22
Jun '22

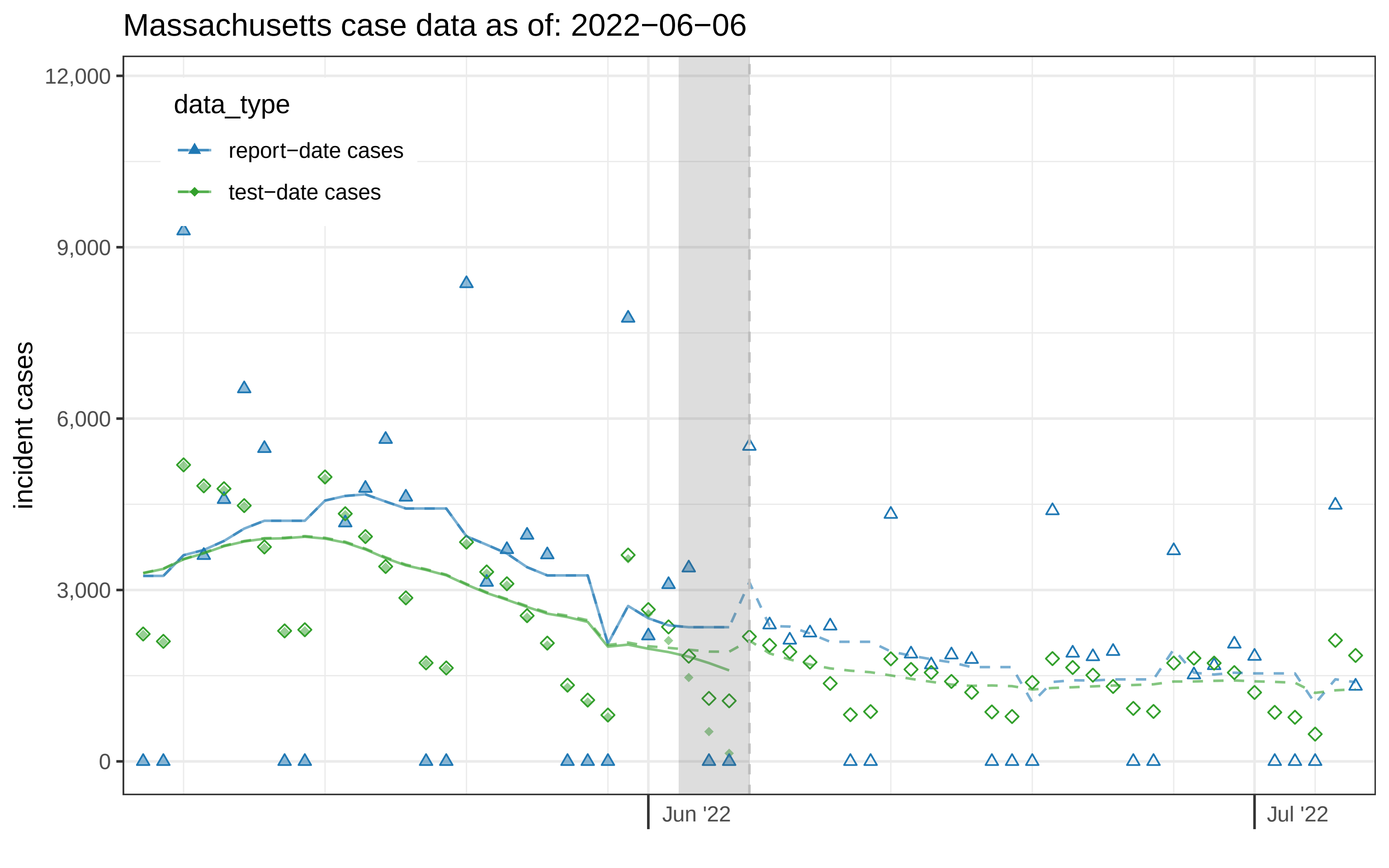
Massachusetts case data as of: 2022−06−06
data_type
report−date cases
test−date cases
incident cases
12,000
9,000
6,000
3,000
0
Jun '22
Jul '22

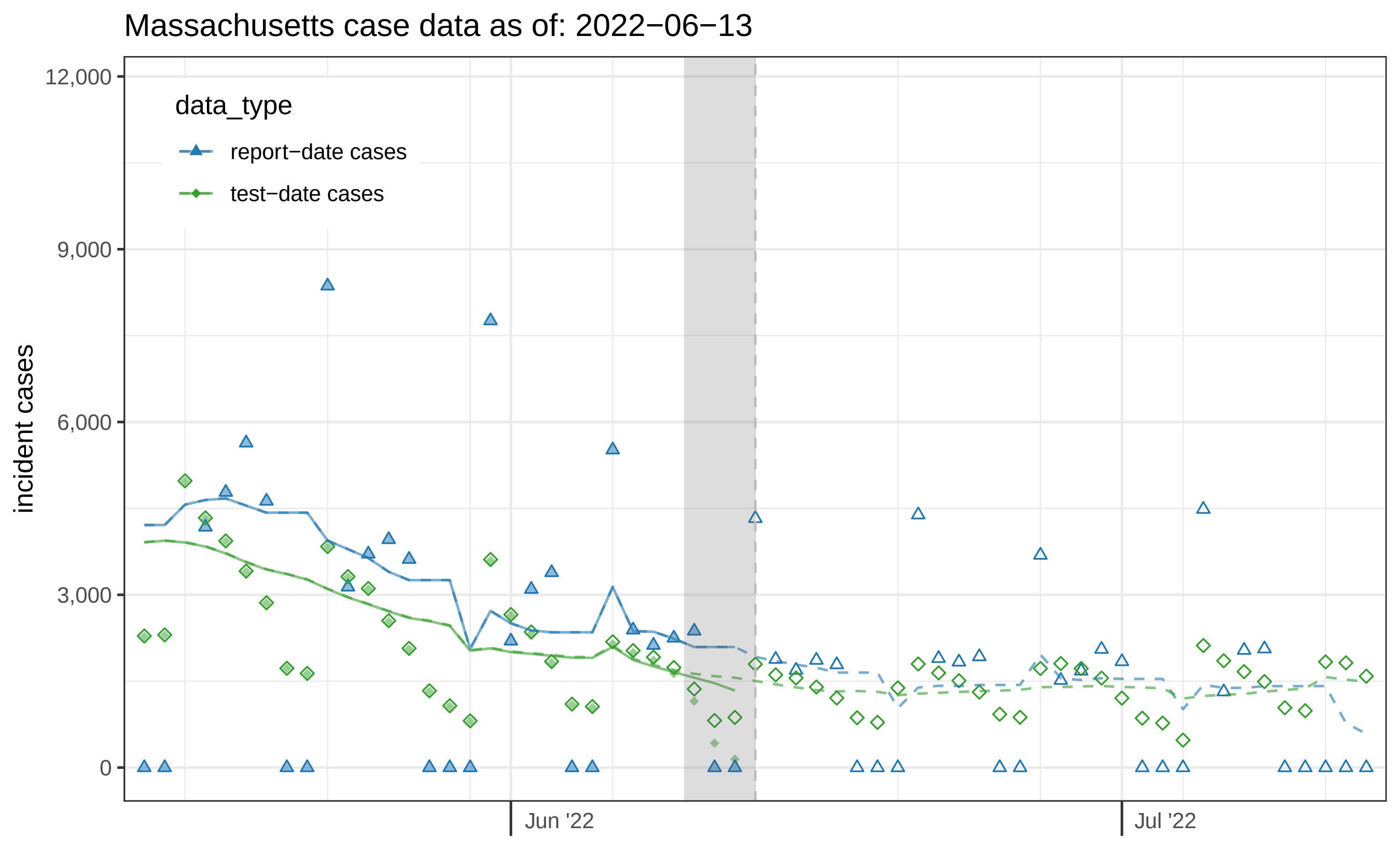
Massachusetts case data as of: 2022−06−13
data_type
report−date cases
test−date cases
incident cases
12,000
9,000
6,000
3,000
0
Jun '22
Jul '22

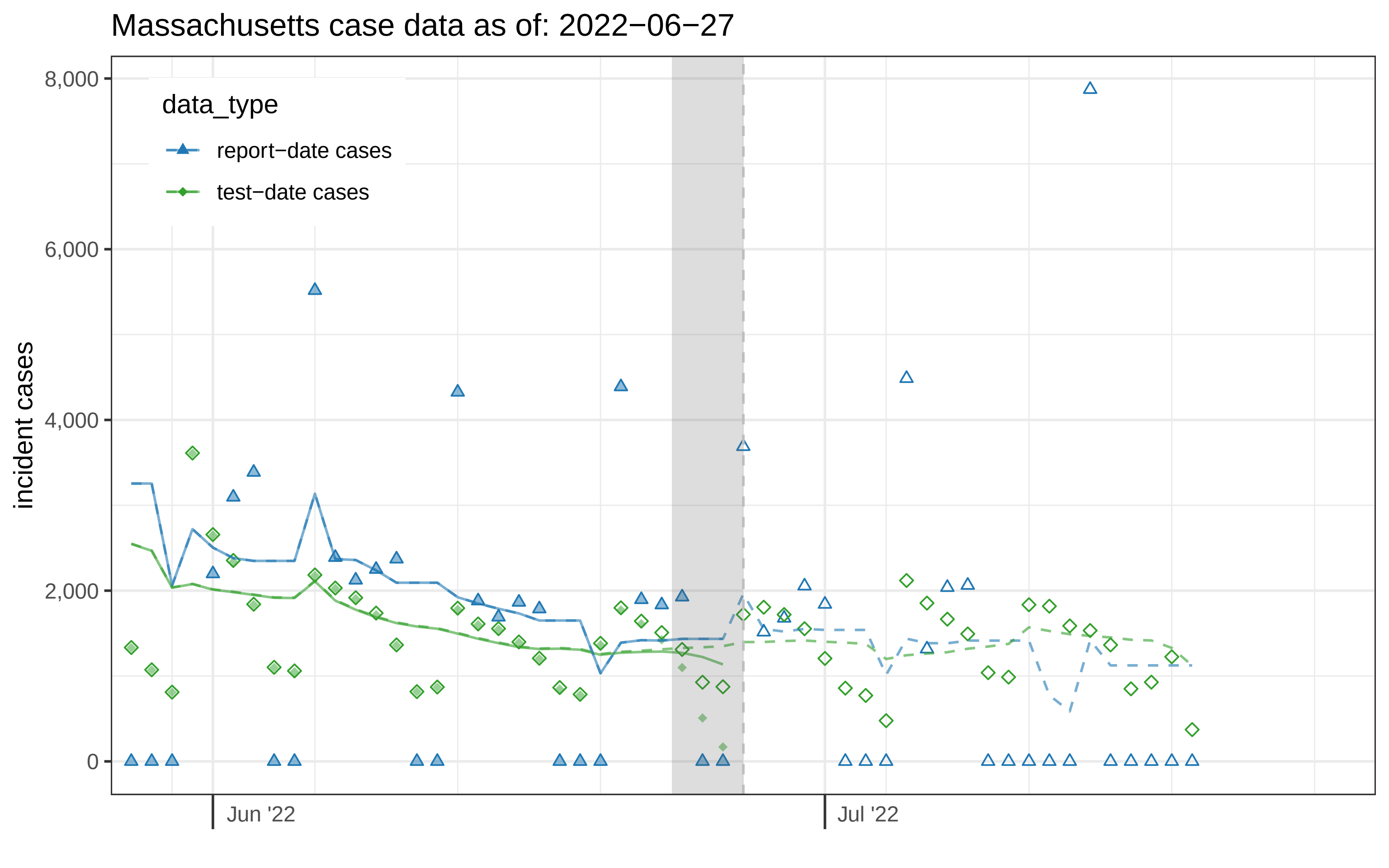

Massachusetts case data as of: 2022−06−27
data_type
report−date cases
test−date cases
incident cases
8,000
6,000
4,000
2,000
0
Jun '22
Jul '22
