## Supplemental File 2 for "Assessing the utility of COVID-19 case reports as a leading indicator for hospitalization forecasting in the United States"

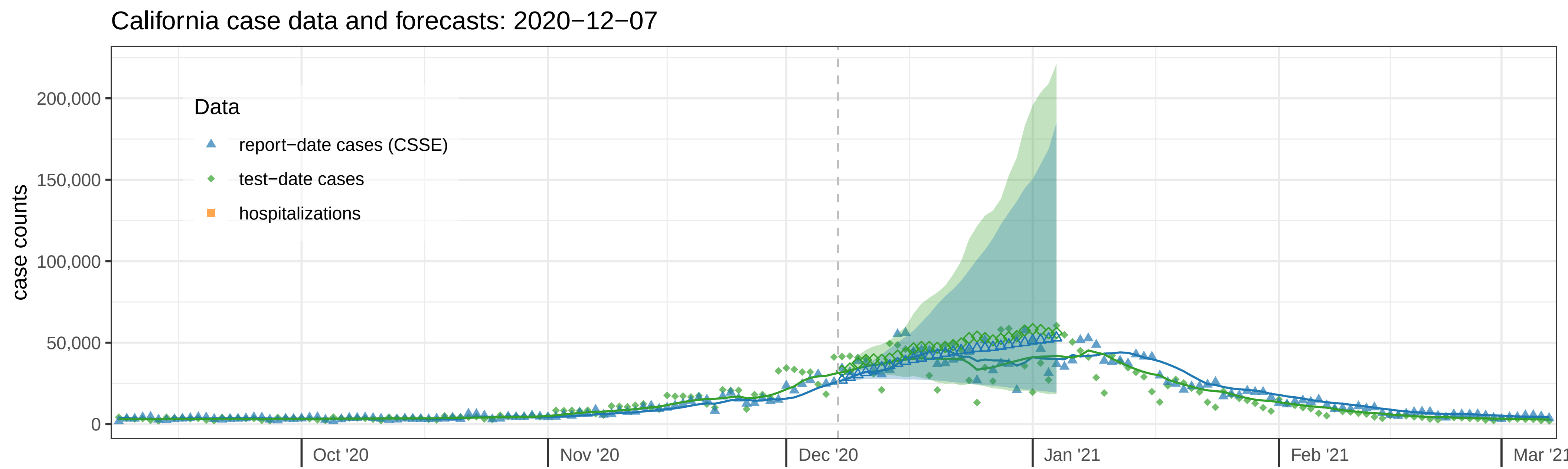
California case data and forecasts: 2020−12−07
Data
report−date cases (CSSE)
test−date cases
hospitalizations
case counts
200,000
150,000
100,000
50,000
0
Oct '20
Nov '20
Dec '20
Jan '21
Feb '21
Mar '21

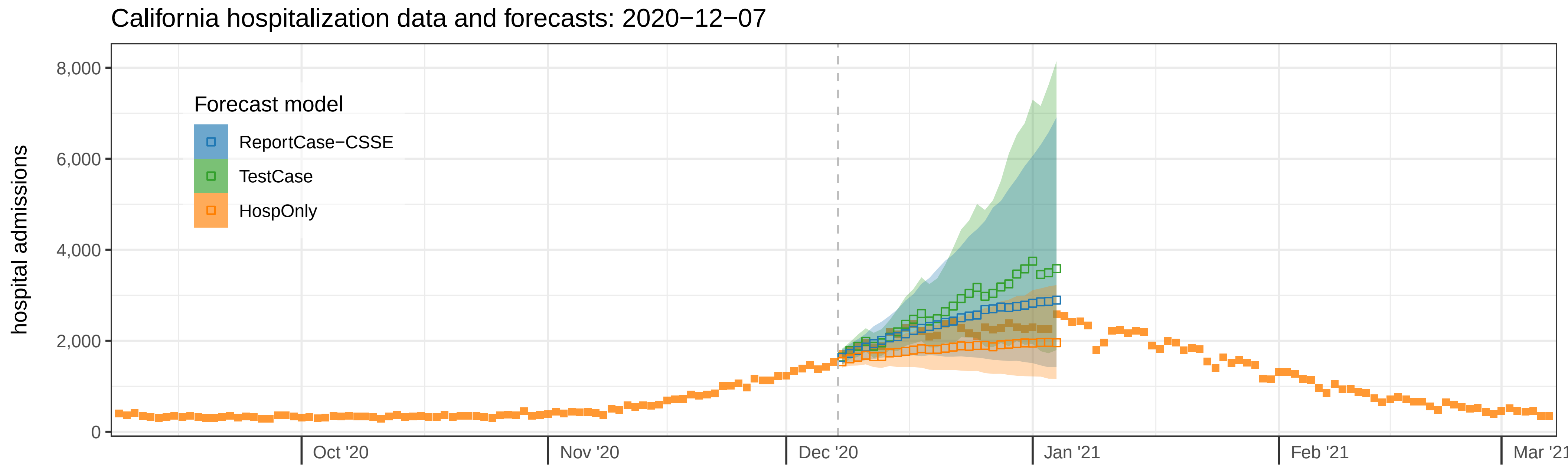
California hospitalization data and forecasts: 2020−12−07
Forecast model
ReportCase−CSSE
TestCase
HospOnly
hospital admissions
8,000
6,000
4,000
2,000
0
Oct '20
Nov '20
Dec '20
Jan '21
Feb '21
Mar '21

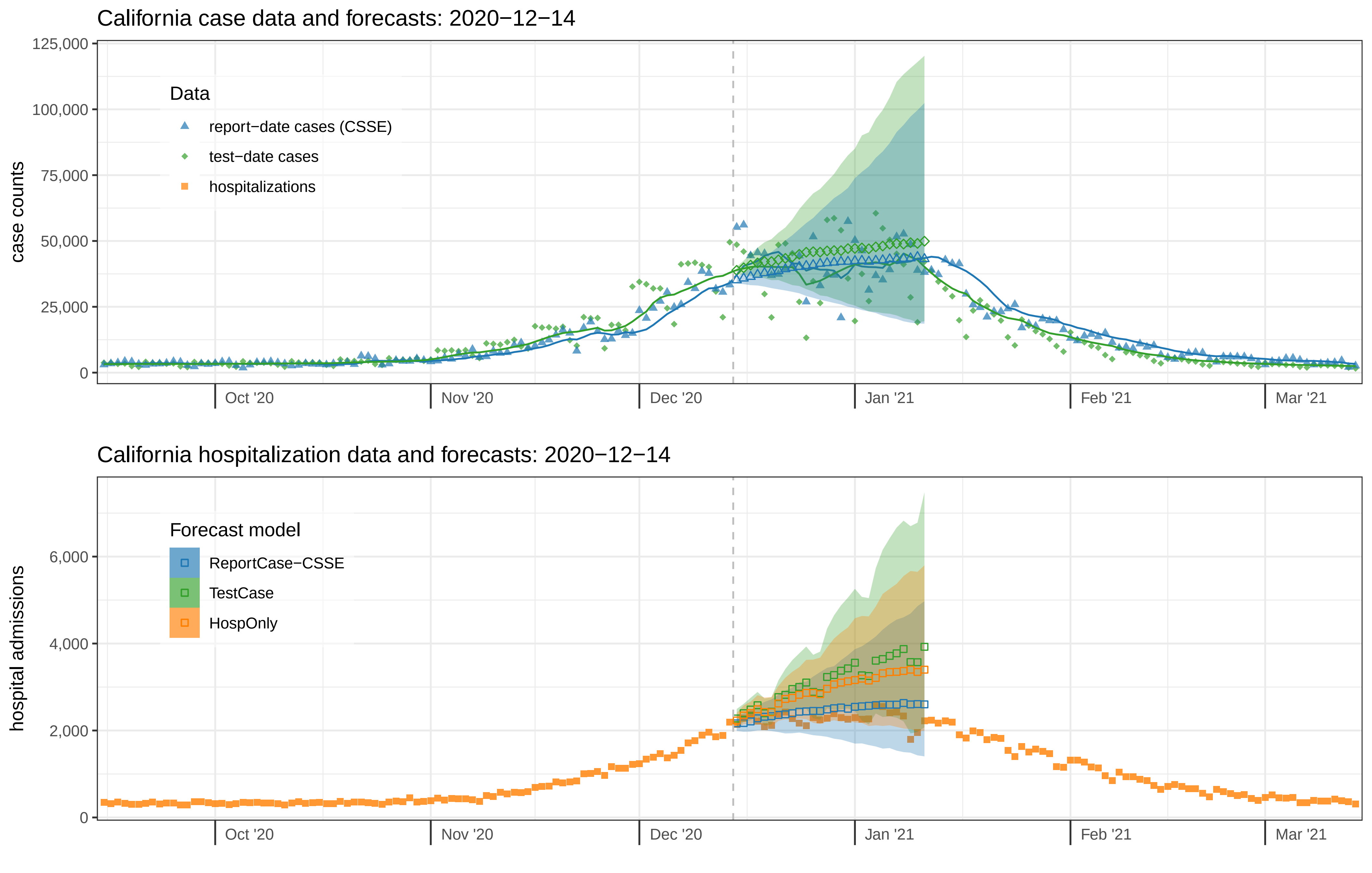

California case data and forecasts: 2020−12−14
Data
report−date cases (CSSE)
test−date cases
hospitalizations
case counts
125,000
100,000
75,000
50,000
25,000
0
Oct '20
Nov '20
Dec '20
Jan '21
Feb '21
Mar '21
California hospitalization data and forecasts: 2020−12−14
Forecast model
ReportCase−CSSE
TestCase
HospOnly
hospital admissions
6,000
4,000
2,000
0
Oct '20
Nov '20
Dec '20
Jan '21
Feb '21
Mar '21

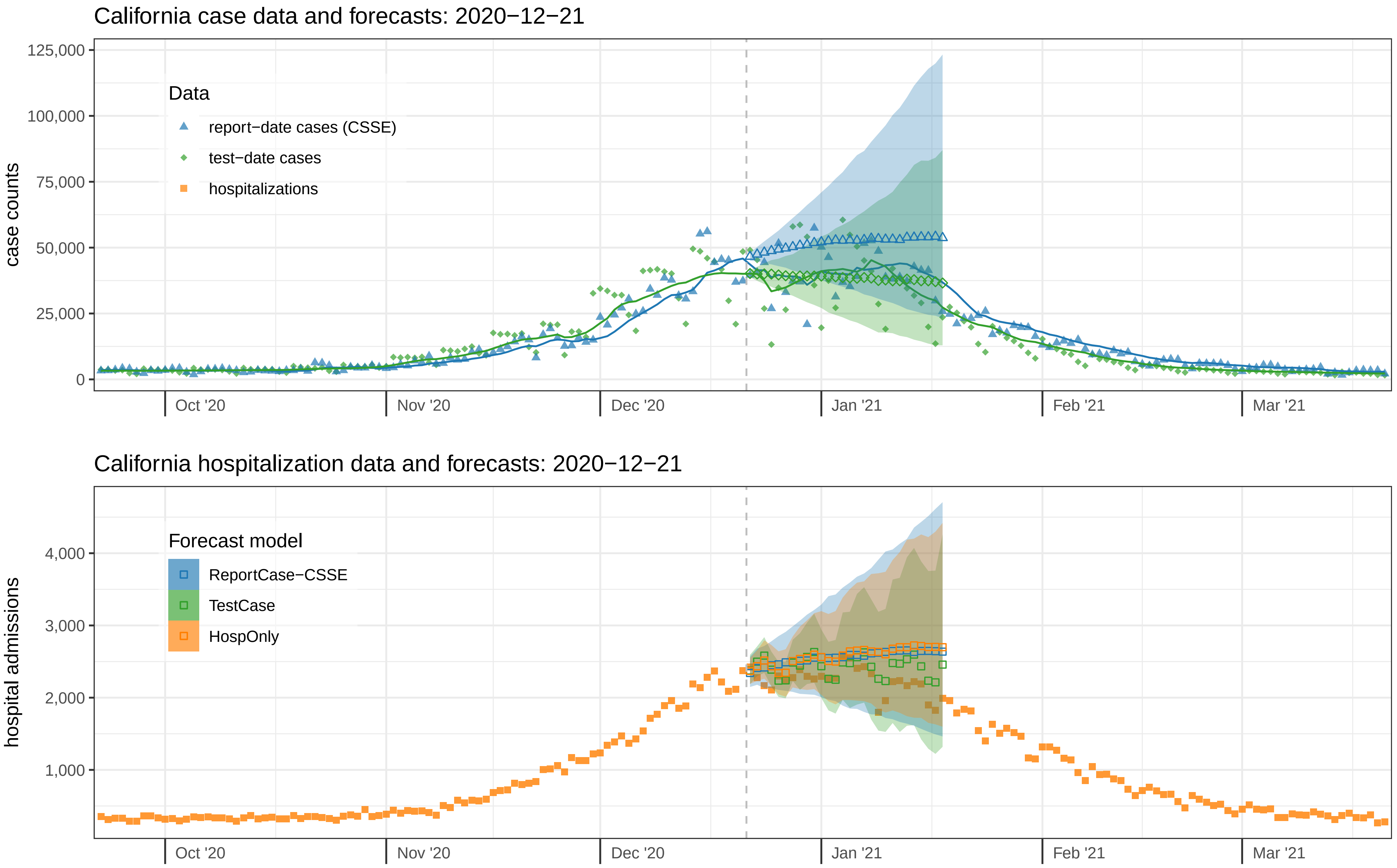

California case data and forecasts: 2020−12−21
case counts
125,000
100,000
75,000
50,000
25,000
0
Data
report−date cases (CSSE)
test−date cases
hospitalizations
Oct '20
Nov '20
Dec '20
Jan '21
Feb '21
Mar '21
California hospitalization data and forecasts: 2020−12−21
hospital admissions
4,000
3,000
2,000
1,000
Forecast model
ReportCase−CSSE
TestCase
HospOnly
Oct '20
Nov '20
Dec '20
Jan '21
Feb '21
Mar '21

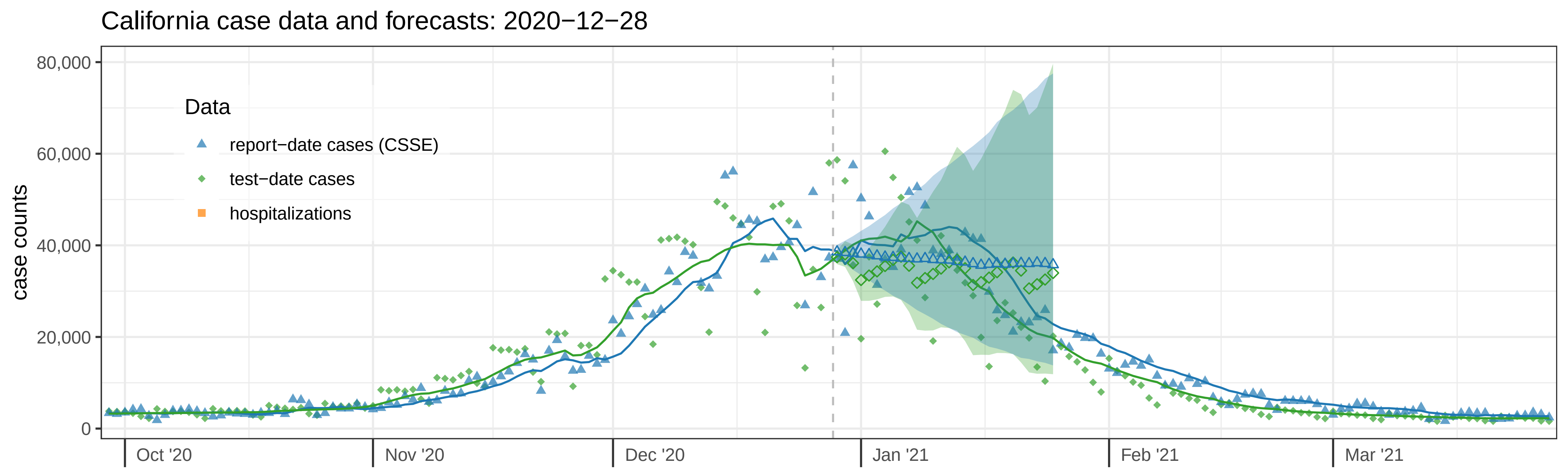
California case data and forecasts: 2020−12−28
case counts
80,000
60,000
40,000
20,000
0
Data
report−date cases (CSSE)
test−date cases
hospitalizations
Oct '20
Nov '20
Dec '20
Jan '21
Feb '21
Mar '21

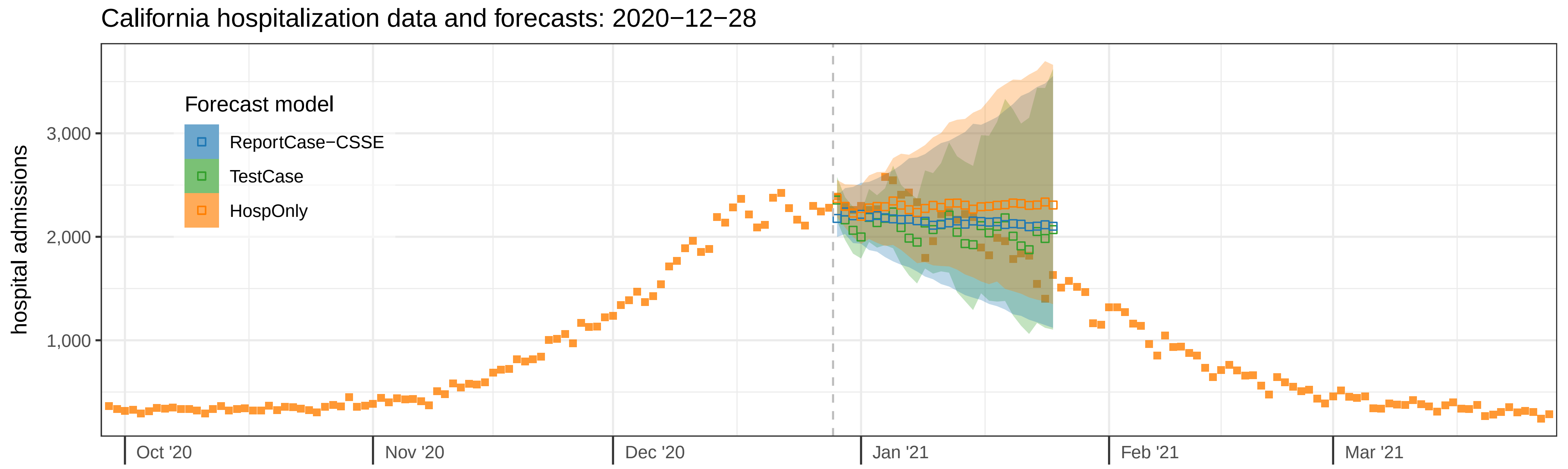
California hospitalization data and forecasts: 2020−12−28
hospital admissions
3,000
2,000
1,000
Forecast model
ReportCase−CSSE
TestCase
HospOnly
Oct '20
Nov '20
Dec '20
Jan '21
Feb '21
Mar '21

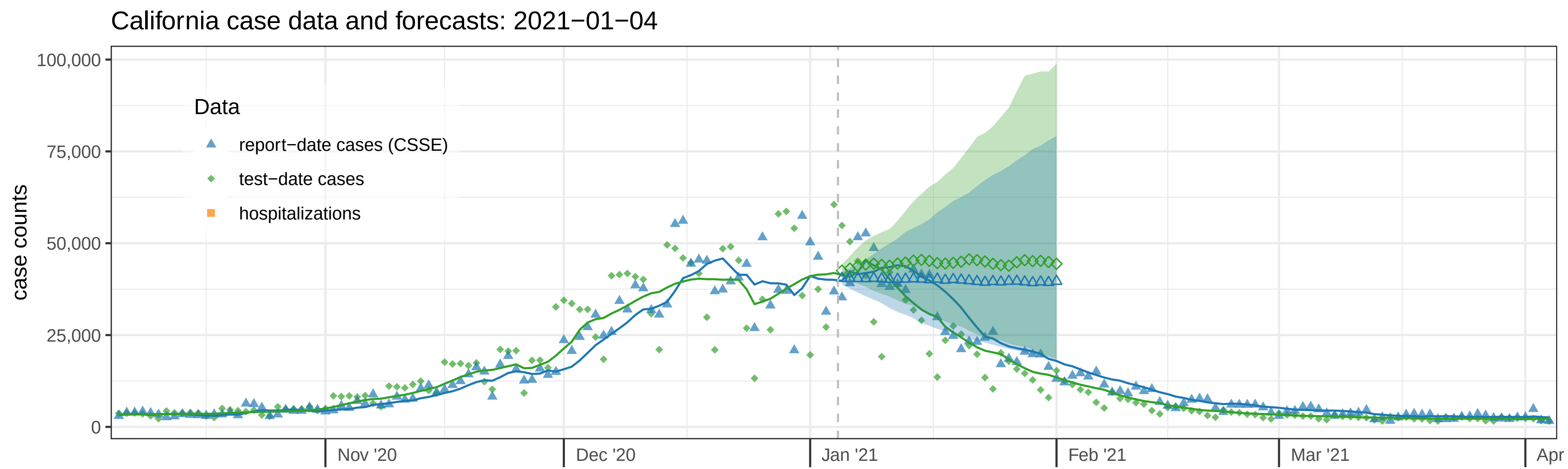

California case data and forecasts: 2021−01−04
case counts
100,000
75,000
50,000
25,000
0
Data
report−date cases (CSSE)
test−date cases
hospitalizations
Nov '20
Dec '20
Jan '21
Feb '21
Mar '21
Apr


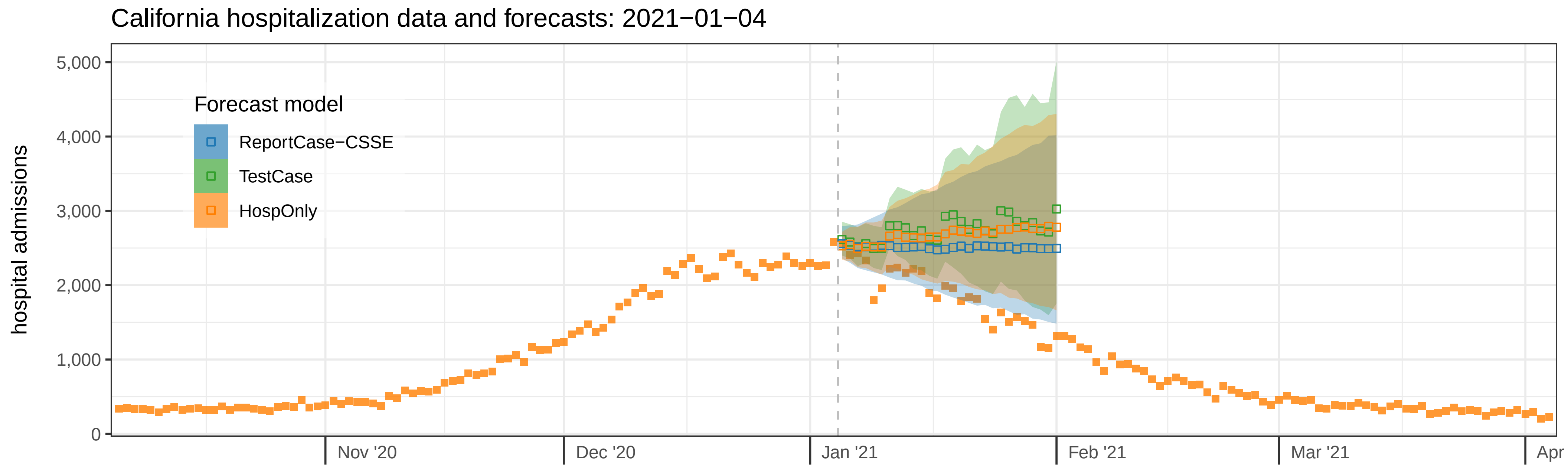

California hospitalization data and forecasts: 2021−01−04
hospital admissions
5,000
4,000
3,000
2,000
1,000
0
Forecast model
ReportCase−CSSE
TestCase
HospOnly
Nov '20
Dec '20
Jan '21
Feb '21
Mar '21
Apr

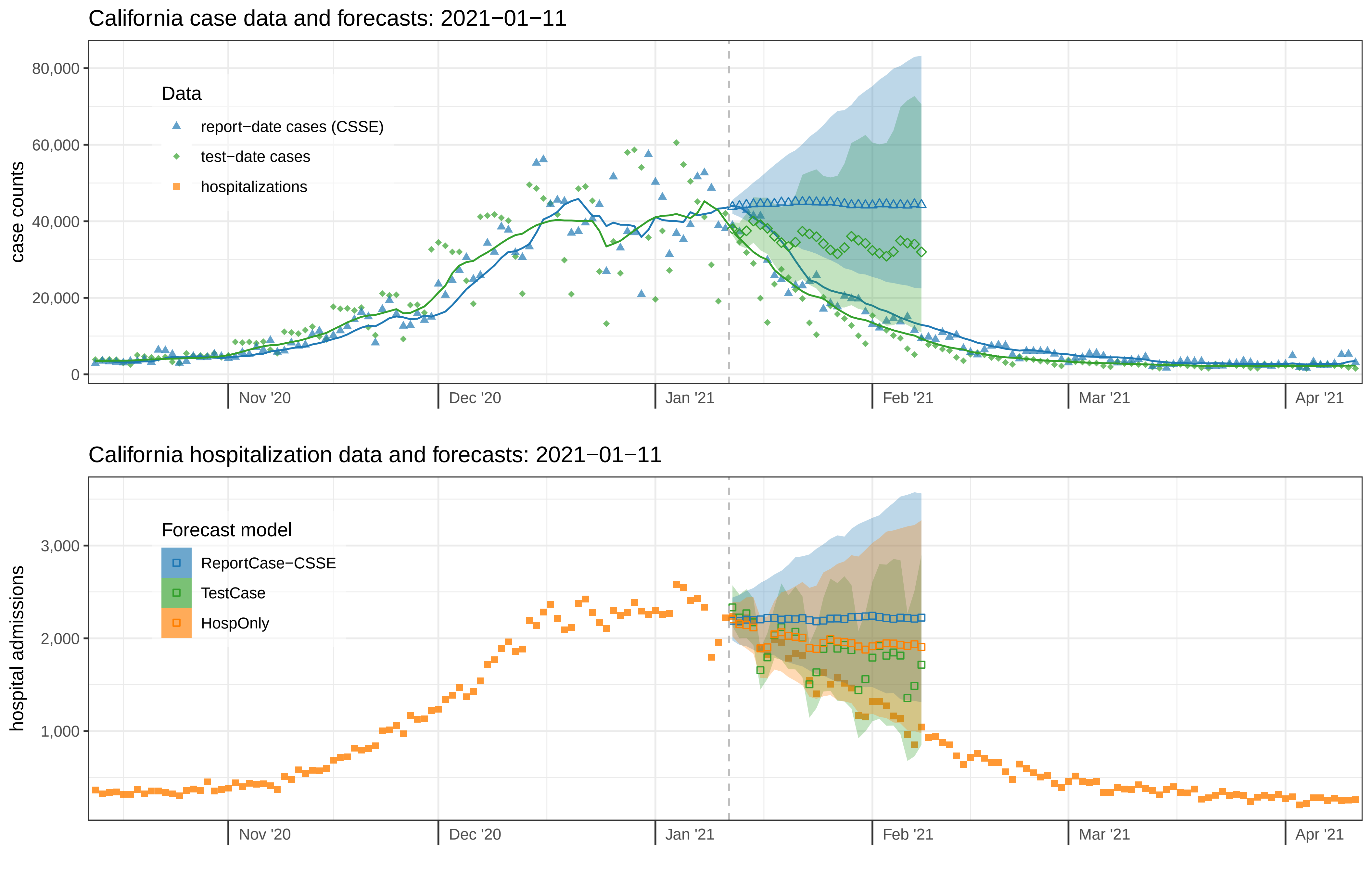

California case data and forecasts: 2021−01−11
case counts
80,000
60,000
40,000
20,000
0
Data
report−date cases (CSSE)
test−date cases
hospitalizations
Nov '20
Dec '20
Jan '21
Feb '21
Mar '21
Apr '21
California hospitalization data and forecasts: 2021−01−11
hospital admissions
3,000
2,000
1,000
Forecast model
ReportCase−CSSE
TestCase
HospOnly
Nov '20
Dec '20
Jan '21
Feb '21
Mar '21
Apr '21

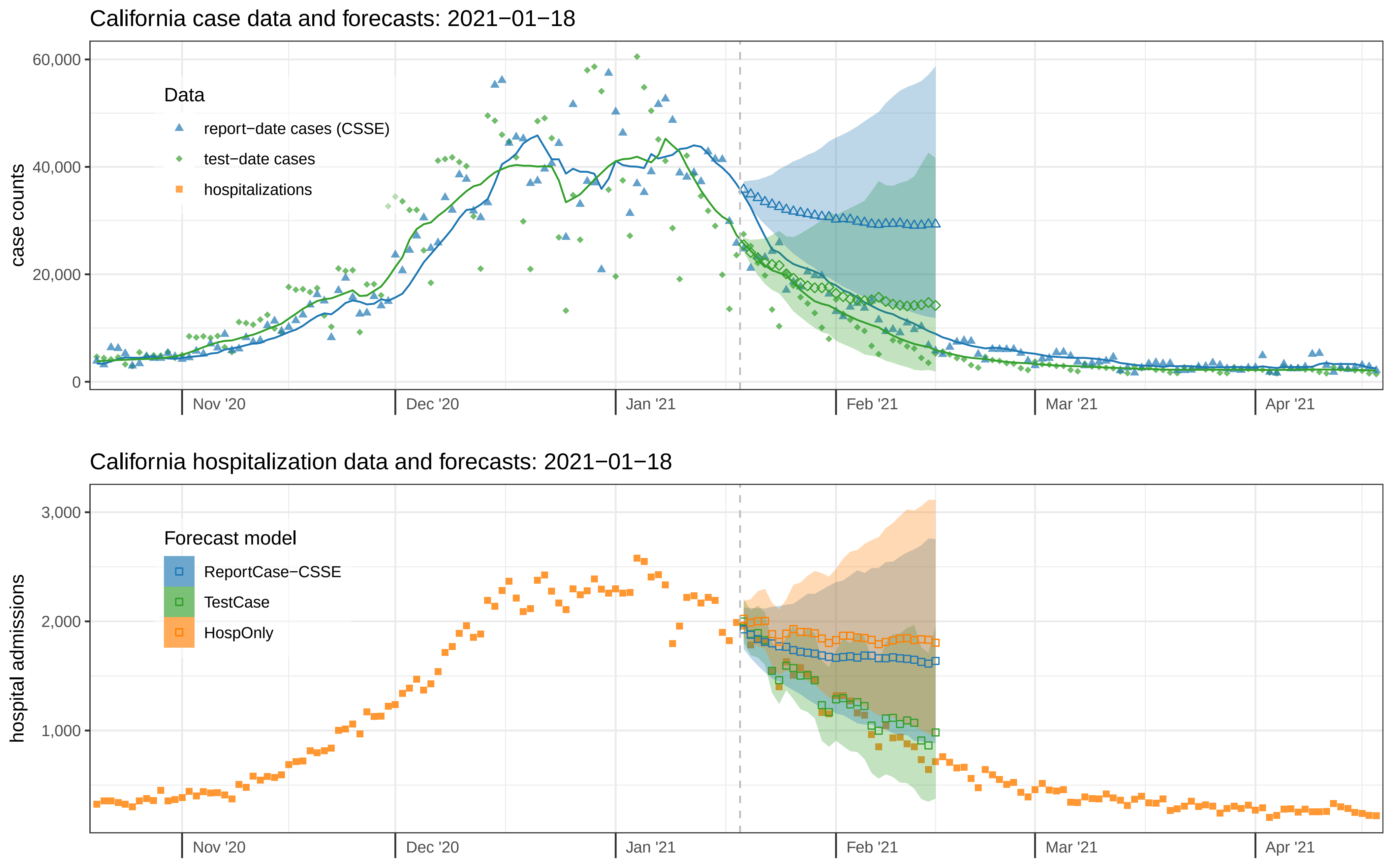

California case data and forecasts: 2021−01−18
case counts
60,000
40,000
20,000
0
Data
report−date cases (CSSE)
test−date cases
hospitalizations
Nov '20
Dec '20
Jan '21
Feb '21
Mar '21
Apr '21
California hospitalization data and forecasts: 2021−01−18
hospital admissions
3,000
2,000
1,000
Forecast model
ReportCase−CSSE
TestCase
HospOnly
Nov '20
Dec '20
Jan '21
Feb '21
Mar '21
Apr '21

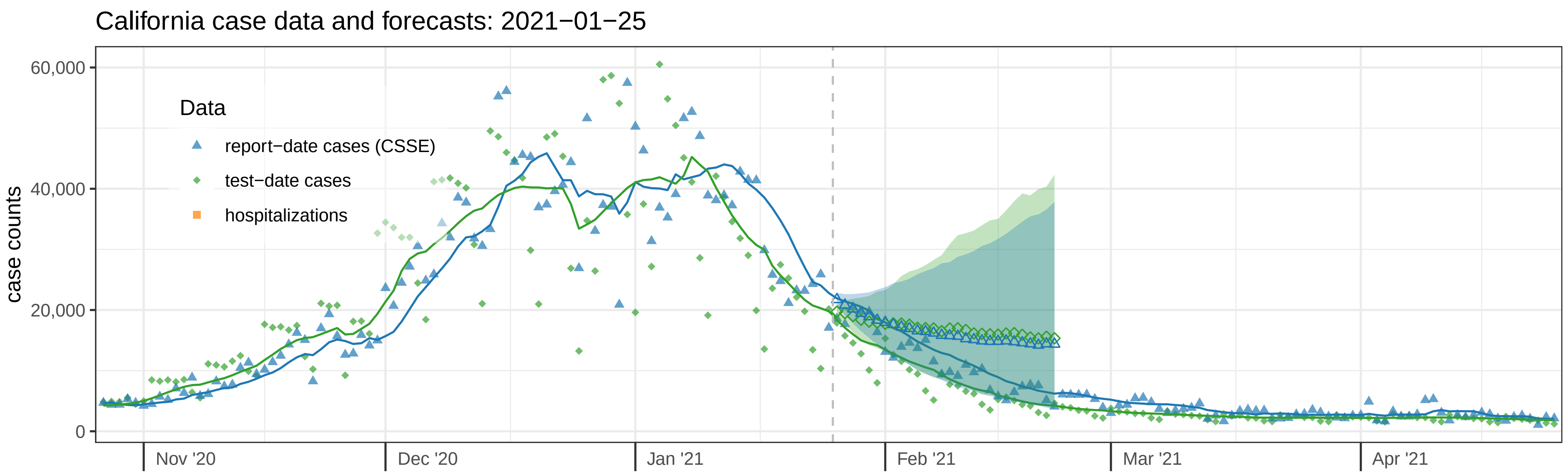
California case data and forecasts: 2021−01−25
case counts
60,000
40,000
20,000
0
Data
report−date cases (CSSE)
test−date cases
hospitalizations
Nov '20
Dec '20
Jan '21
Feb '21
Mar '21
Apr '21

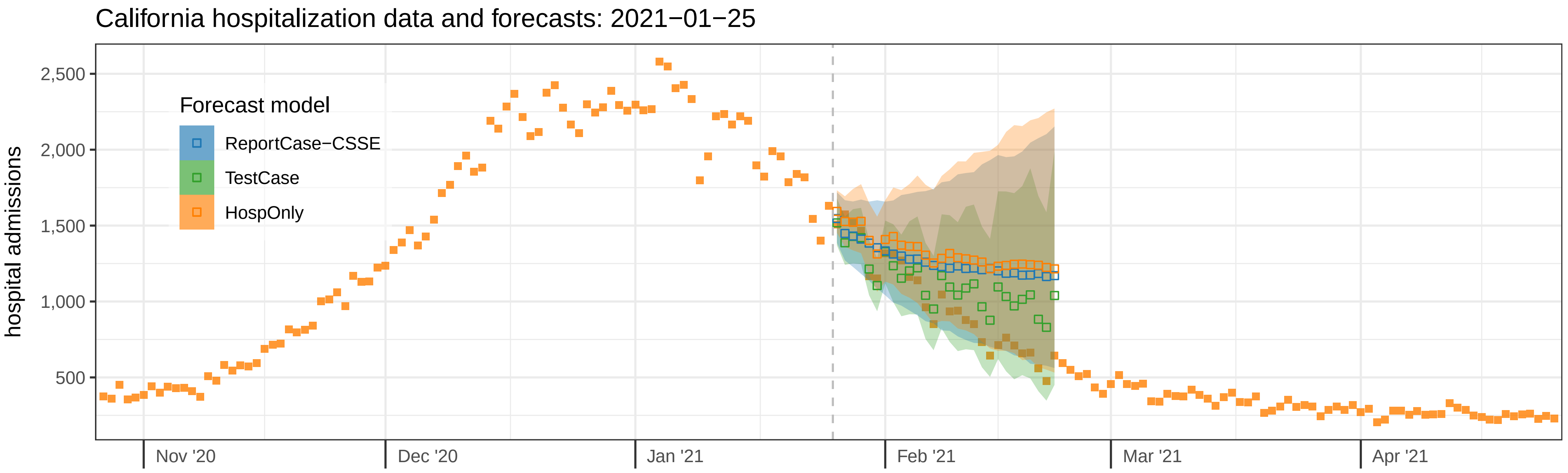
California hospitalization data and forecasts: 2021−01−25
hospital admissions
2,500
2,000
1,500
1,000
500
Forecast model
ReportCase−CSSE
TestCase
HospOnly
Nov '20
Dec '20
Jan '21
Feb '21
Mar '21
Apr '21

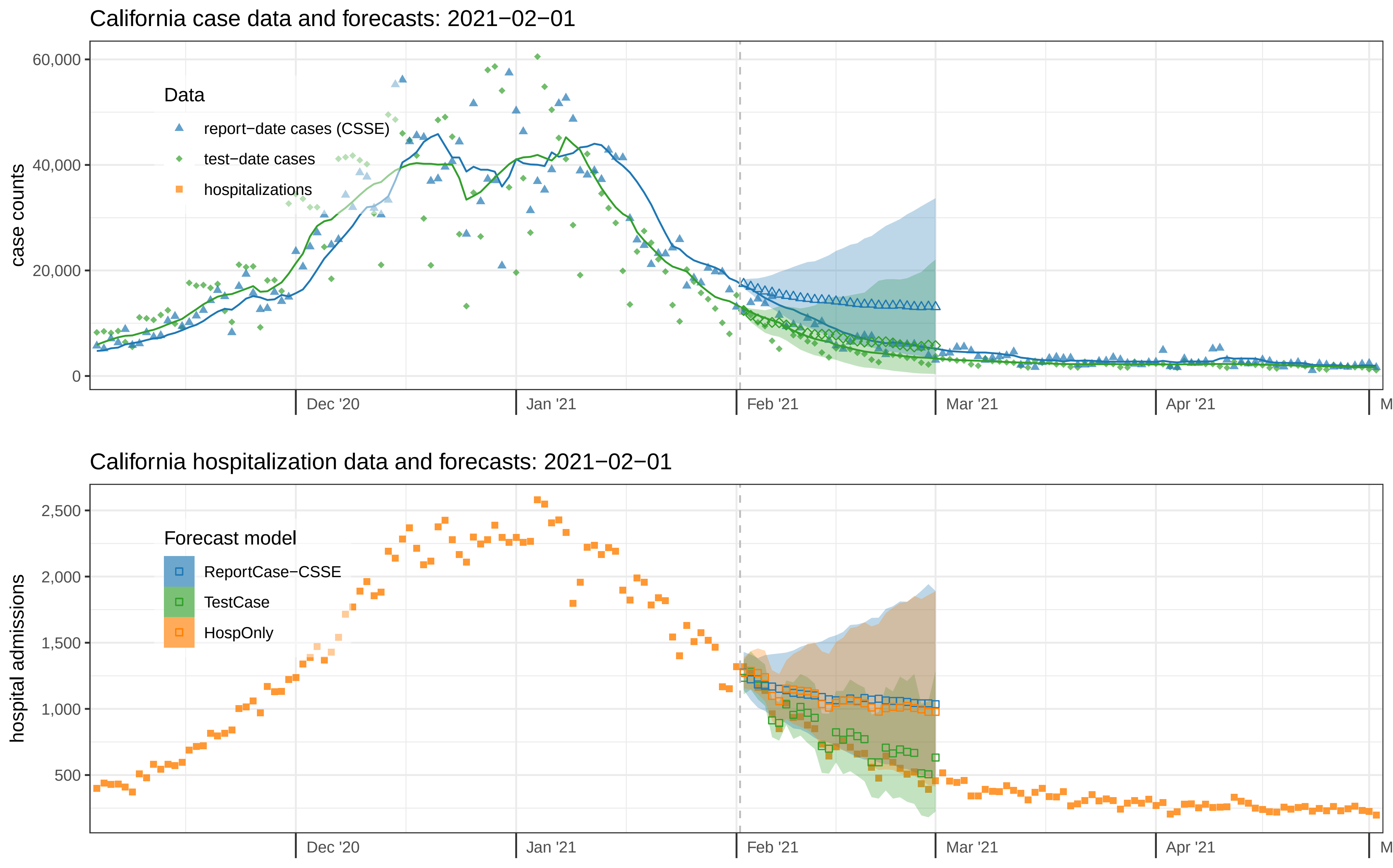
California case data and forecasts: 2021−02−01
case counts
60,000
40,000
20,000
0
Data
report−date cases (CSSE)
test−date cases
hospitalizations
Dec '20
Jan '21
Feb '21
Mar '21
Apr '21
California hospitalization data and forecasts: 2021−02−01
hospital admissions
2,500
2,000
1,500
1,000
500
Forecast model
ReportCase−CSSE
TestCase
HospOnly
Dec '20
Jan '21
Feb '21
Mar '21
Apr '21

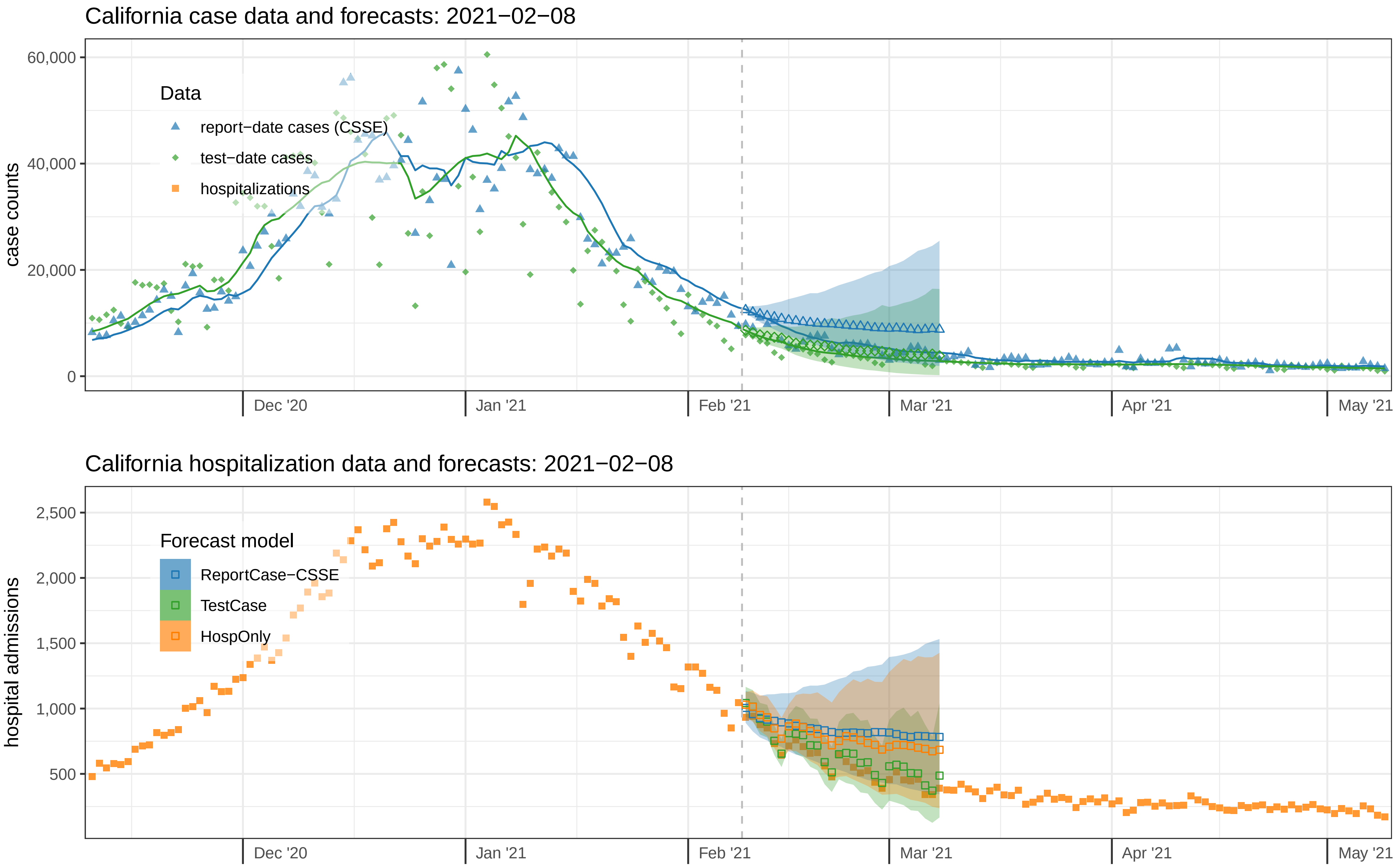

California case data and forecasts: 2021−02−08
case counts
60,000
40,000
20,000
0
Data
report−date cases (CSSE)
test−date cases
hospitalizations
Dec '20
Jan '21
Feb '21
Mar '21
Apr '21
May '21
California hospitalization data and forecasts: 2021−02−08
hospital admissions
2,500
2,000
1,500
1,000
500
Forecast model
ReportCase−CSSE
TestCase
HospOnly
Dec '20
Jan '21
Feb '21
Mar '21
Apr '21
May '21

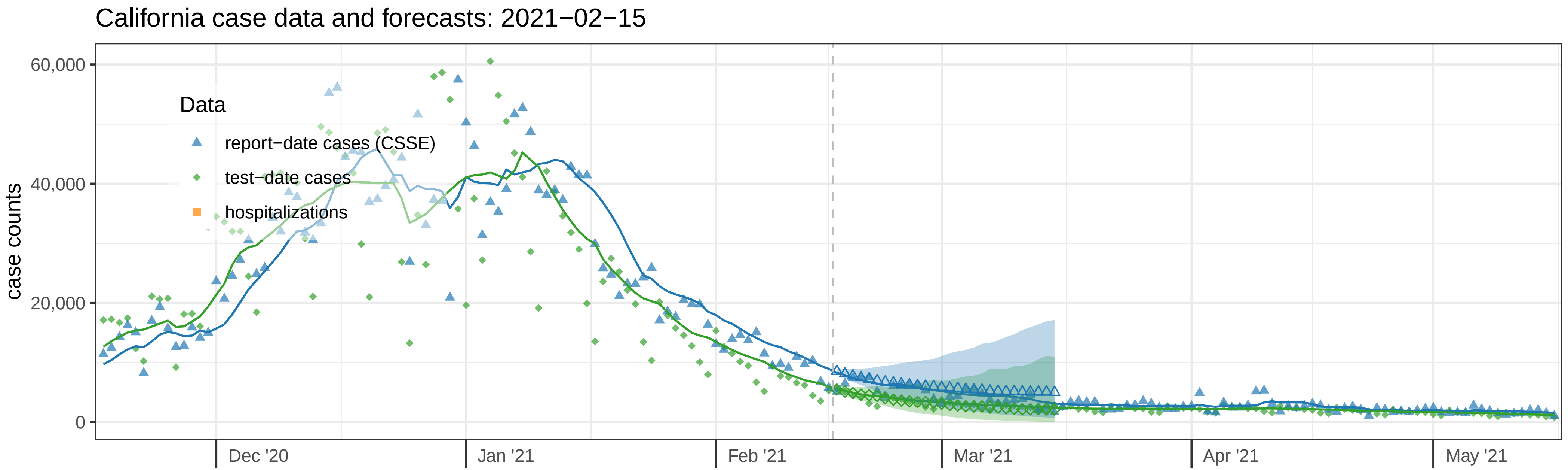

California case data and forecasts: 2021−02−15
case counts
60,000
40,000
20,000
0
Data
report−date cases (CSSE)
test−date cases
hospitalizations
Dec '20
Jan '21
Feb '21
Mar '21
Apr '21
May '21


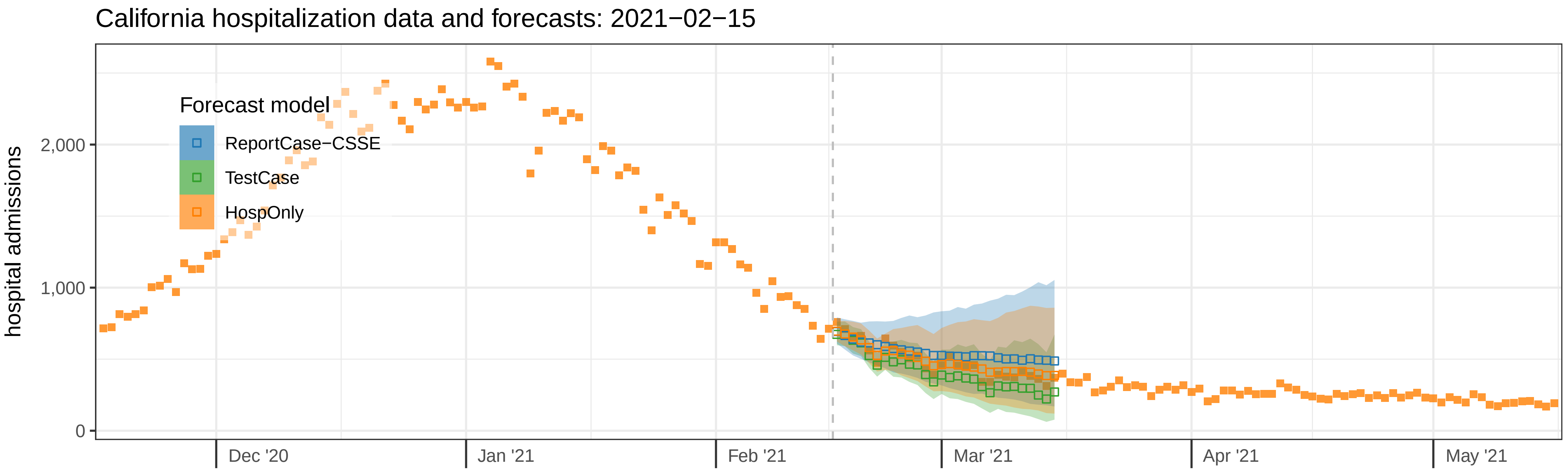

California hospitalization data and forecasts: 2021−02−15
hospital admissions
2,000
1,000
0
Forecast model
ReportCase−CSSE
TestCase
HospOnly
Dec '20
Jan '21
Feb '21
Mar '21
Apr '21
May '21

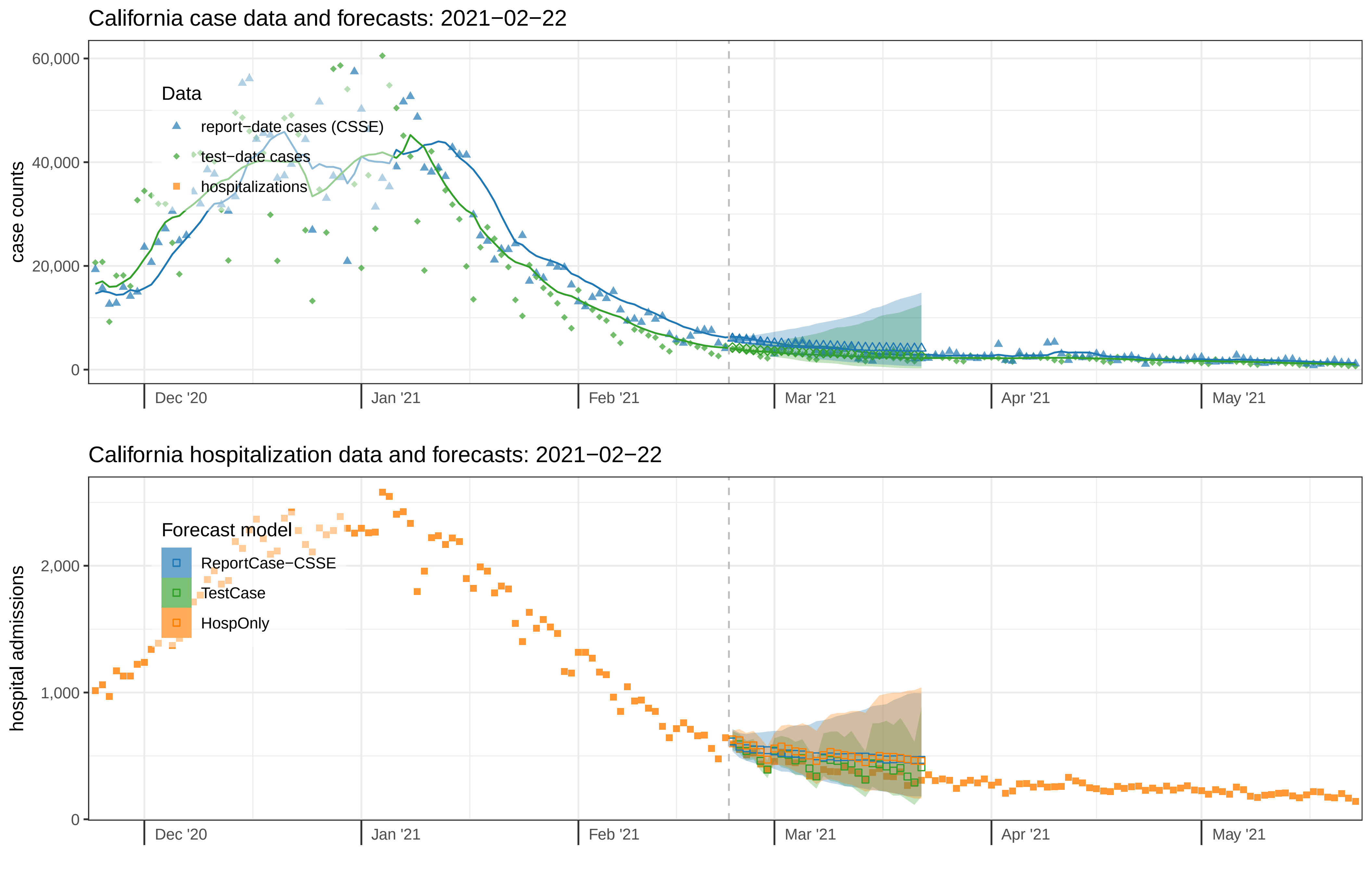

California case data and forecasts: 2021−02−22
case counts
60,000
40,000
20,000
0
Data
report−date cases (CSSE)
test−date cases
hospitalizations
Dec '20
Jan '21
Feb '21
Mar '21
Apr '21
May '21
California hospitalization data and forecasts: 2021−02−22
hospital admissions
2,000
1,000
0
Forecast model
ReportCase−CSSE
TestCase
HospOnly
Dec '20
Jan '21
Feb '21
Mar '21
Apr '21
May '21

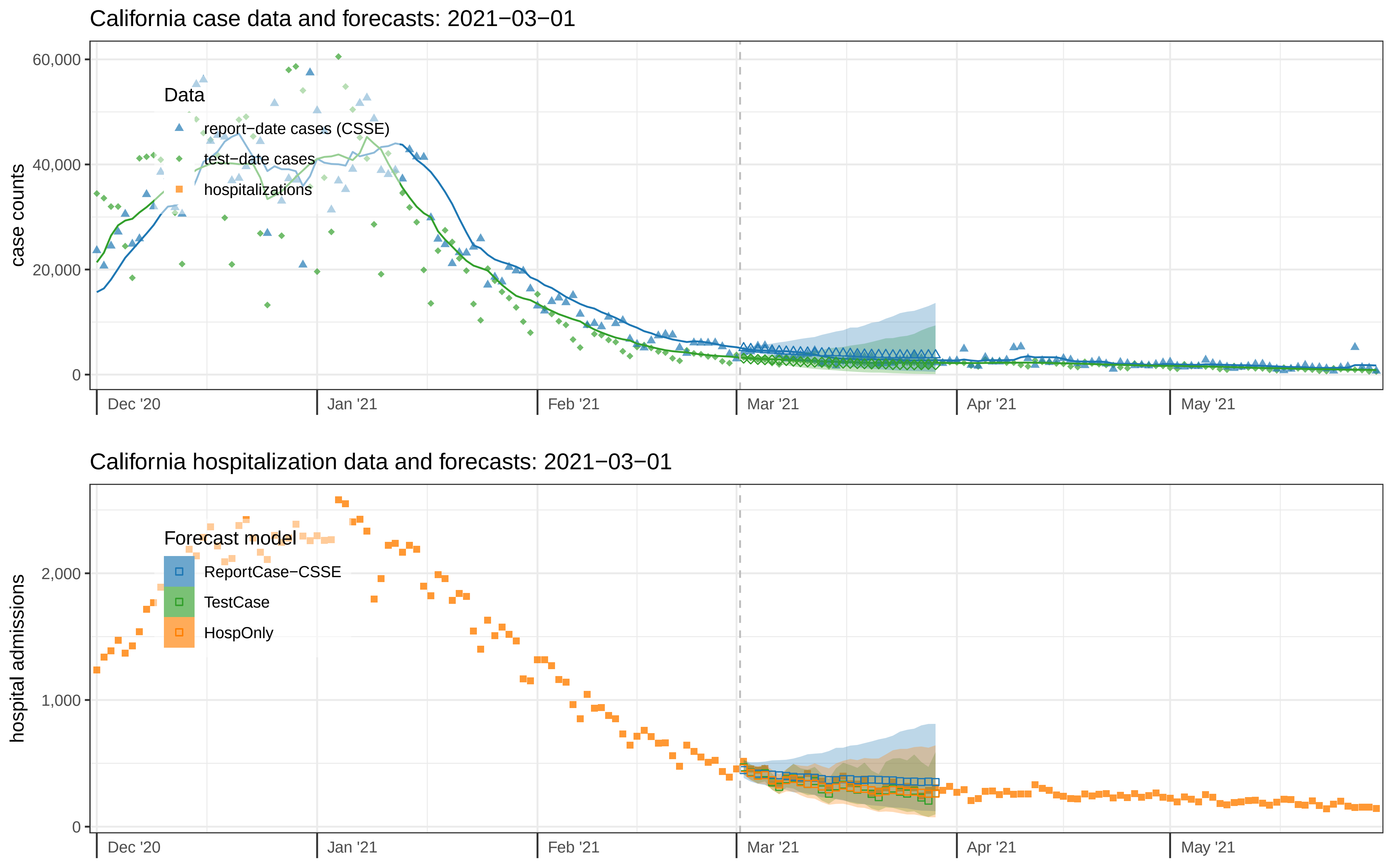

California case data and forecasts: 2021−03−01
case counts
60,000
40,000
20,000
0
Data
report−date cases (CSSE)
test−date cases
hospitalizations
Dec '20
Jan '21
Feb '21
Mar '21
Apr '21
May '21
California hospitalization data and forecasts: 2021−03−01
hospital admissions
2,000
1,000
0
Forecast model
ReportCase−CSSE
TestCase
HospOnly
Dec '20
Jan '21
Feb '21
Mar '21
Apr '21
May '21

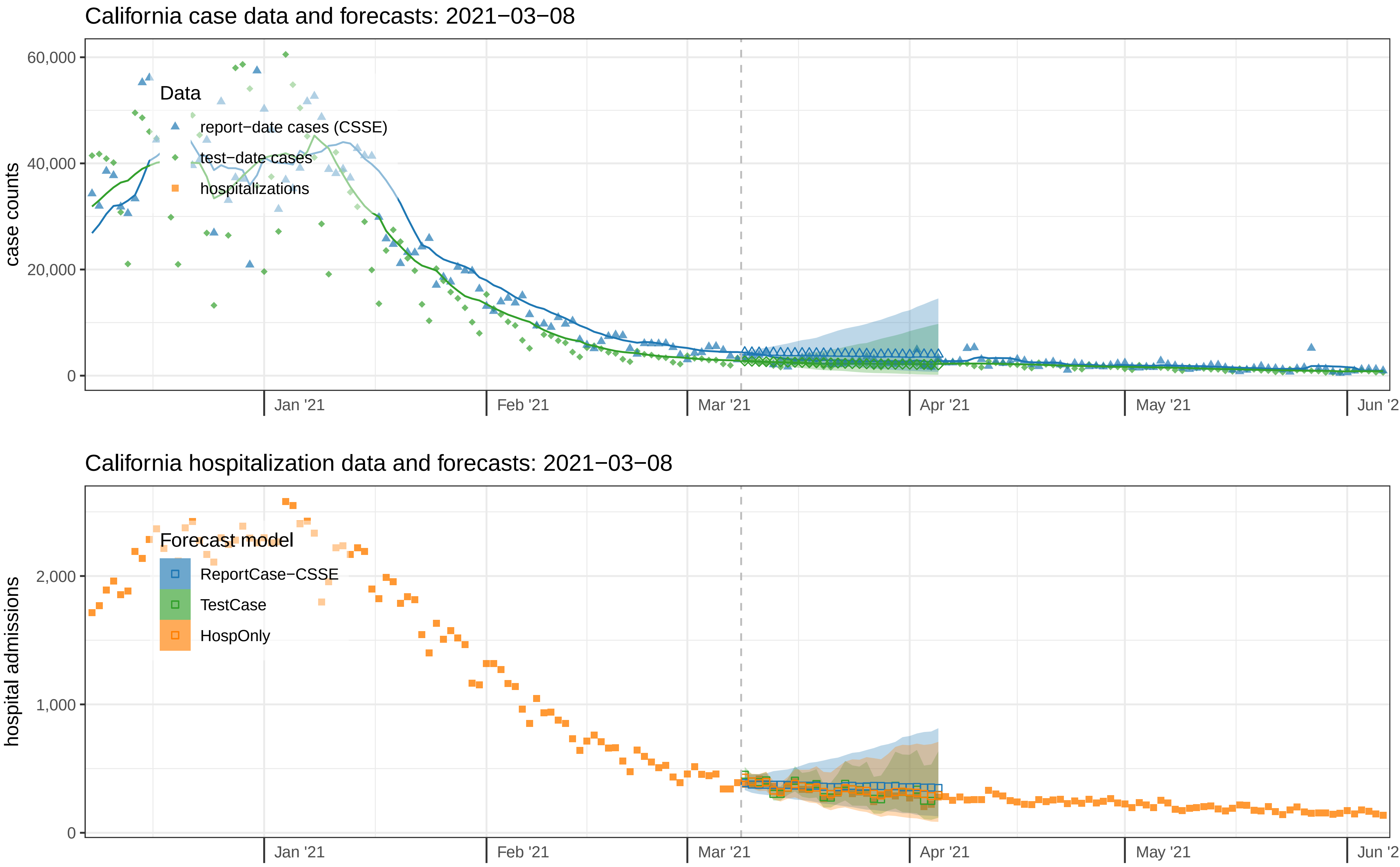

California case data and forecasts: 2021−03−08
case counts
60,000
40,000
20,000
0
Data
report−date cases (CSSE)
test−date cases
hospitalizations
Jan '21
Feb '21
Mar '21
Apr '21
May '21
California hospitalization data and forecasts: 2021−03−08
hospital admissions
2,000
1,000
0
Forecast model
ReportCase−CSSE
TestCase
HospOnly
Jan '21
Feb '21
Mar '21
Apr '21
May '21

California case data and forecasts: 2021−03−15

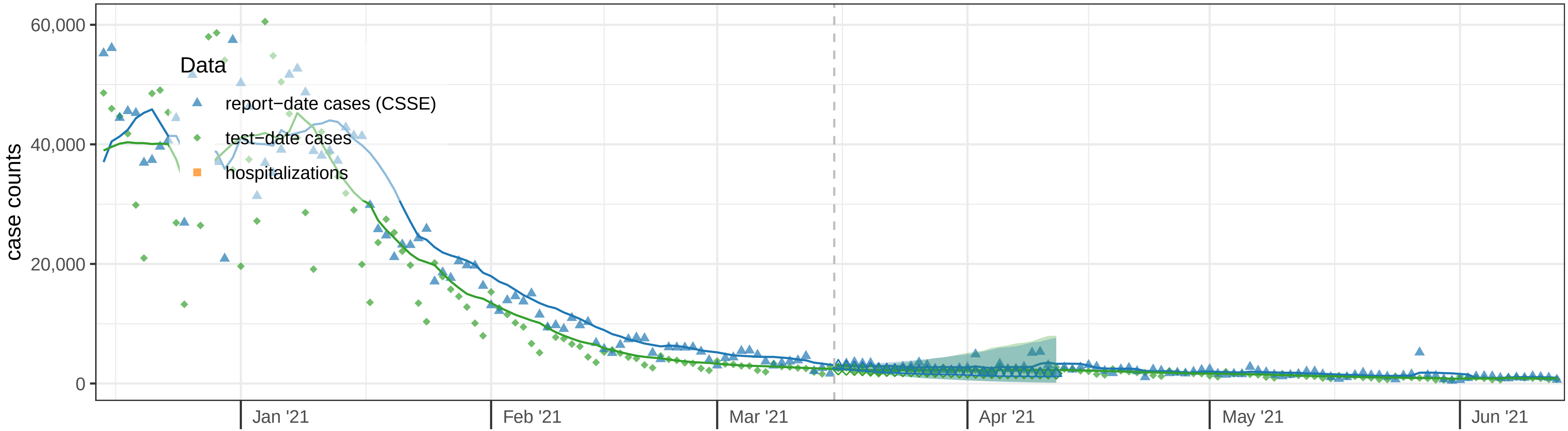


California hospitalization data and forecasts: 2021−03−15

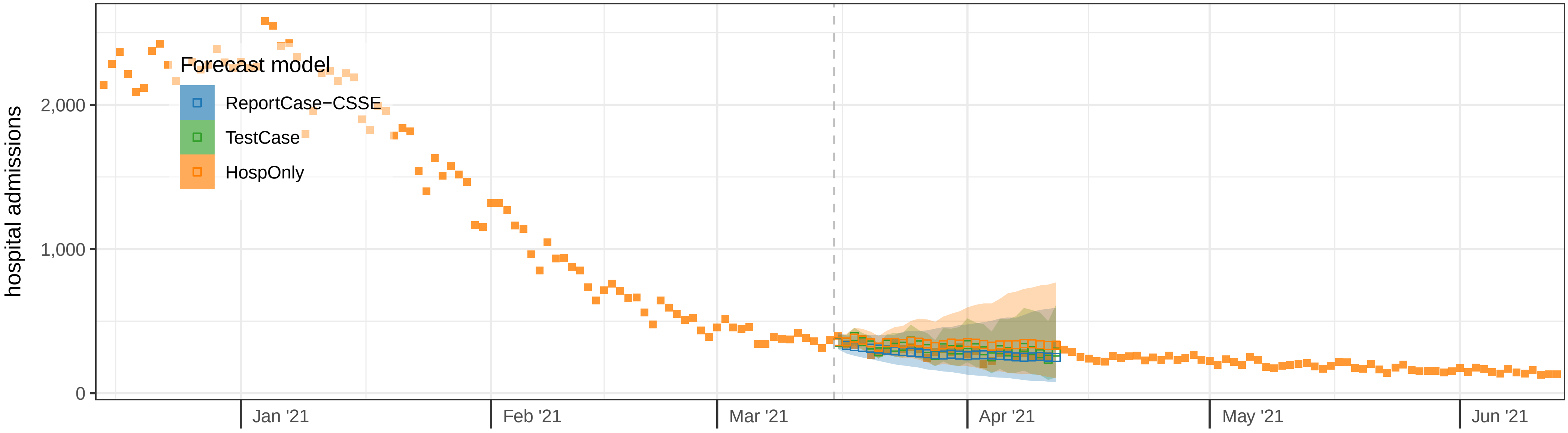

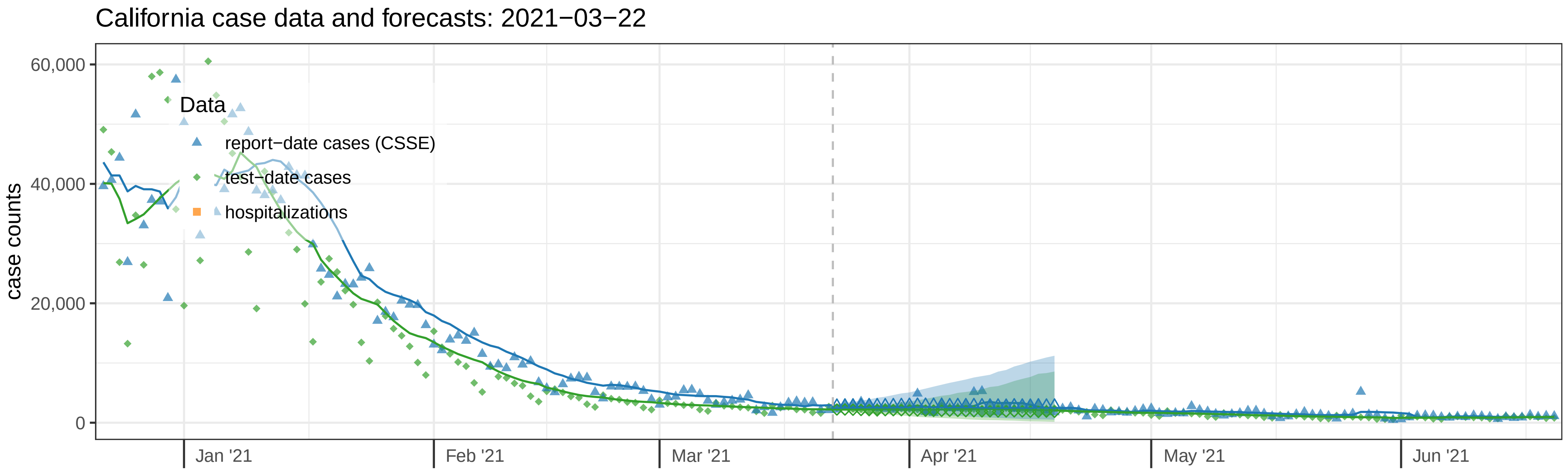

California case data and forecasts: 2021−03−22
case counts
60,000
40,000
20,000
0
Data
report−date cases (CSSE)
test−date cases
hospitalizations
Jan '21
Feb '21
Mar '21
Apr '21
May '21
Jun '21


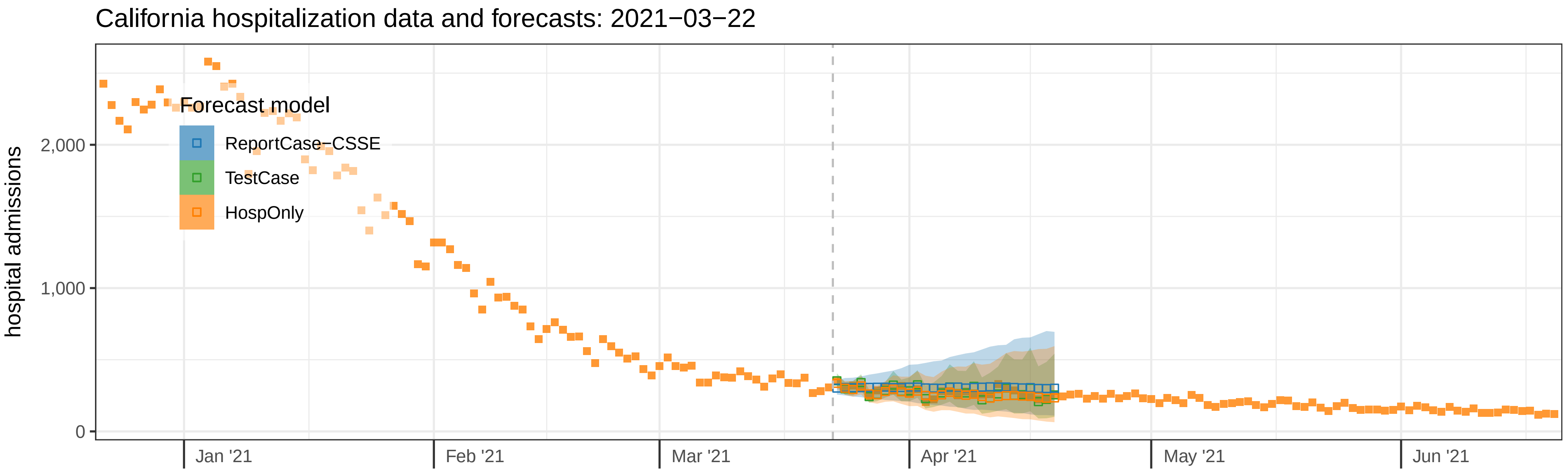

California hospitalization data and forecasts: 2021−03−22
hospital admissions
2,000
1,000
0
Forecast model
ReportCase−CSSE
TestCase
HospOnly
Jan '21
Feb '21
Mar '21
Apr '21
May '21
Jun '21

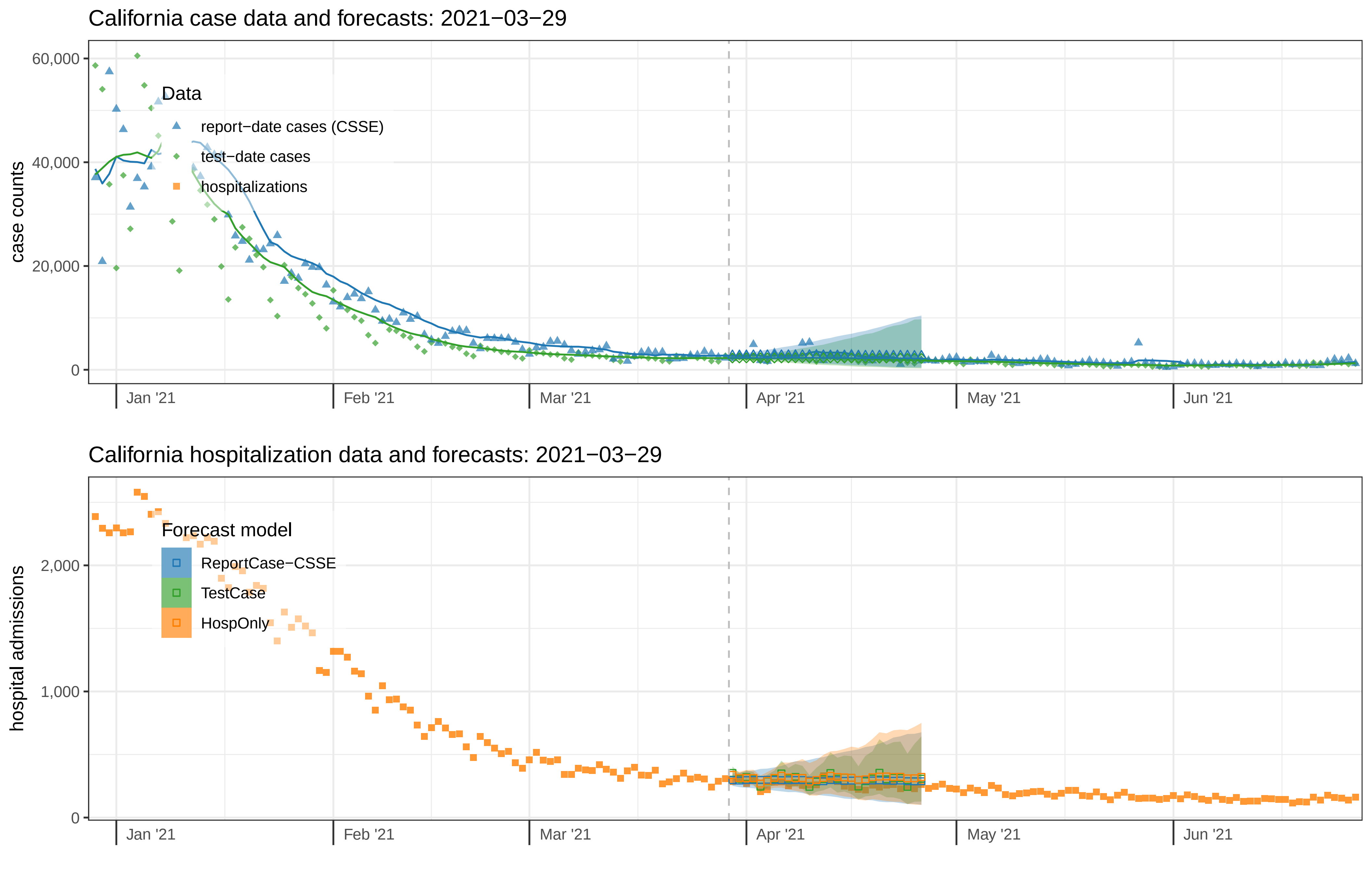

California case data and forecasts: 2021−03−29
case counts
60,000
40,000
20,000
0
Data
report−date cases (CSSE)
test−date cases
hospitalizations
Jan '21
Feb '21
Mar '21
Apr '21
May '21
Jun '21
California hospitalization data and forecasts: 2021−03−29
hospital admissions
2,000
1,000
0
Forecast model
ReportCase−CSSE
TestCase
HospOnly
Jan '21
Feb '21
Mar '21
Apr '21
May '21
Jun '21

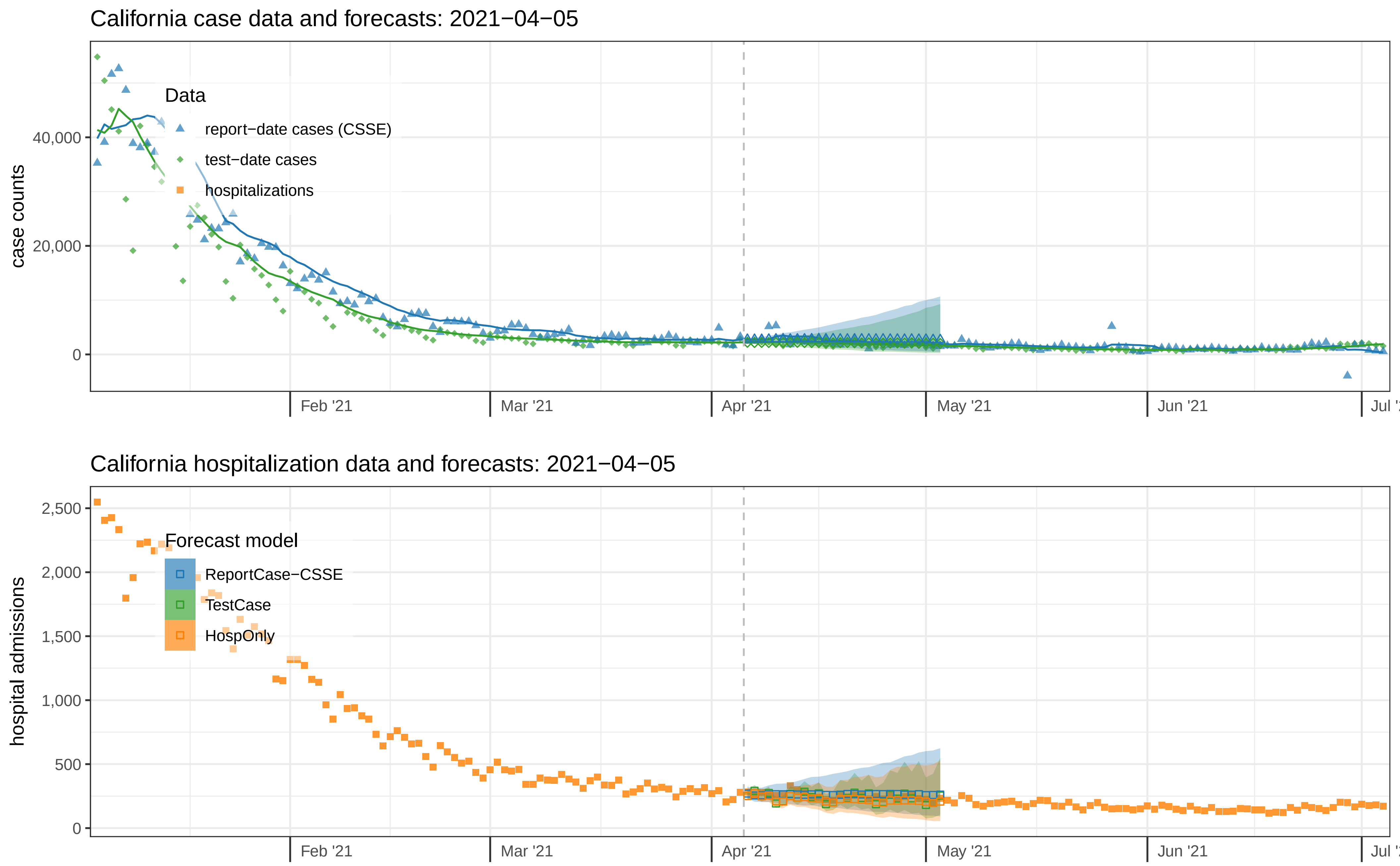

California case data and forecasts: 2021−04−05
case counts
40,000
20,000
0
Data
report−date cases (CSSE)
test−date cases
hospitalizations
Feb '21
Mar '21
Apr '21
May '21
Jun '21
California hospitalization data and forecasts: 2021−04−05
hospital admissions
2,500
2,000
1,500
1,000
500
0
Forecast model
ReportCase−CSSE
TestCase
HospOnly
Feb '21
Mar '21
Apr '21
May '21
Jun '21

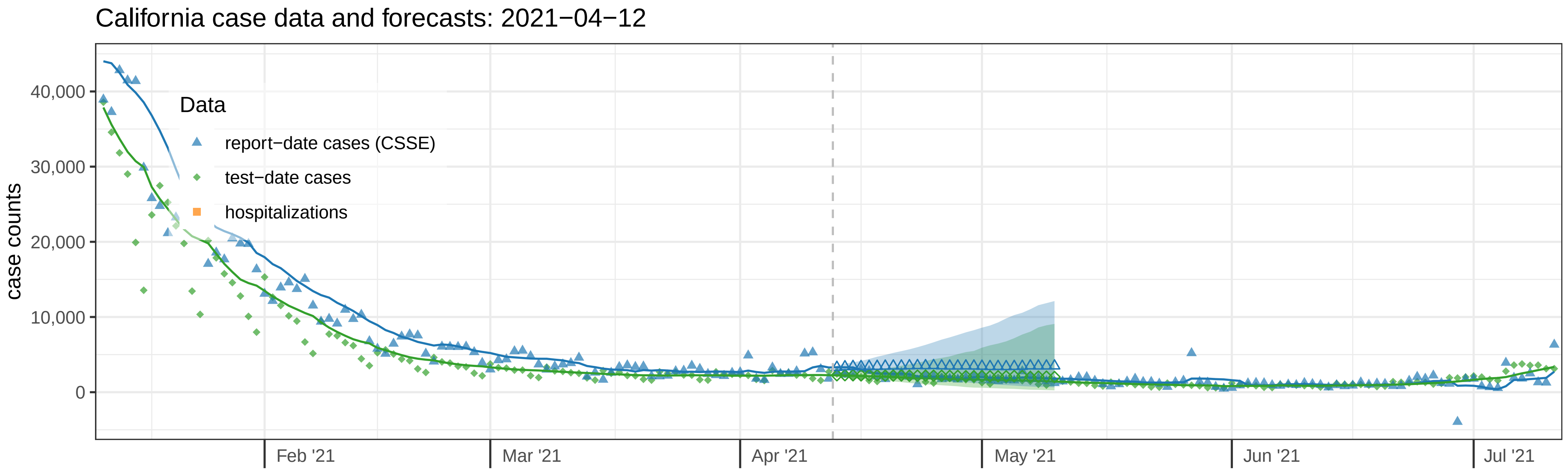
California case data and forecasts: 2021−04−12
case counts
40,000
30,000
20,000
10,000
0
Data
report−date cases (CSSE)
test−date cases
hospitalizations
Feb '21
Mar '21
Apr '21
May '21
Jun '21
Jul '21

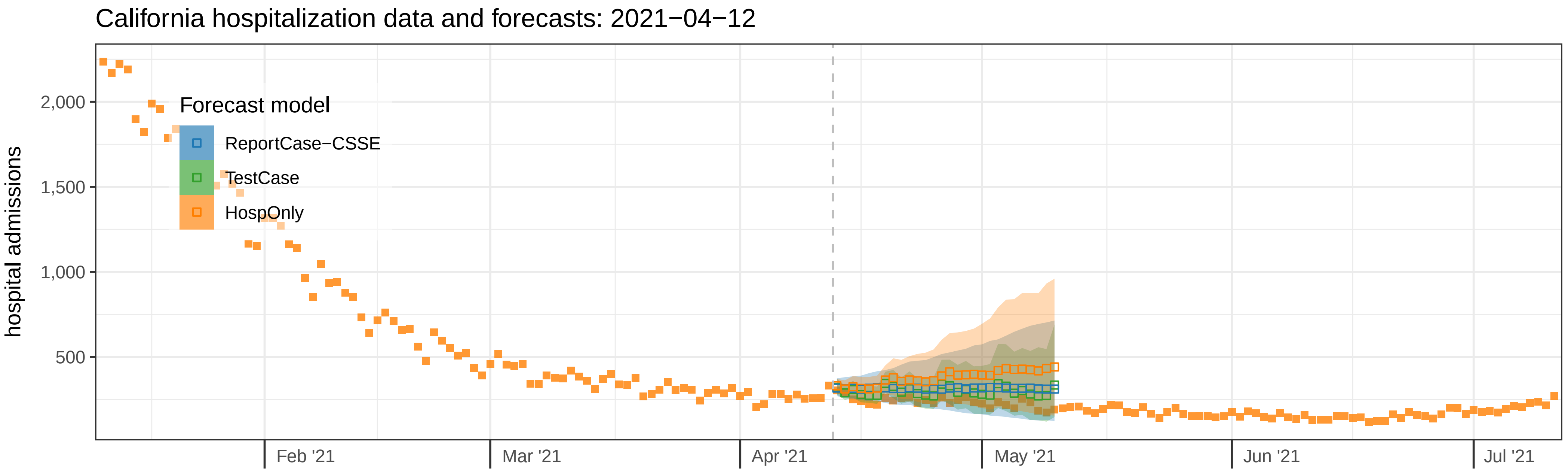
California hospitalization data and forecasts: 2021−04−12
hospital admissions
2,000
1,500
1,000
500
Forecast model
ReportCase−CSSE
TestCase
HospOnly
Feb '21
Mar '21
Apr '21
May '21
Jun '21
Jul '21

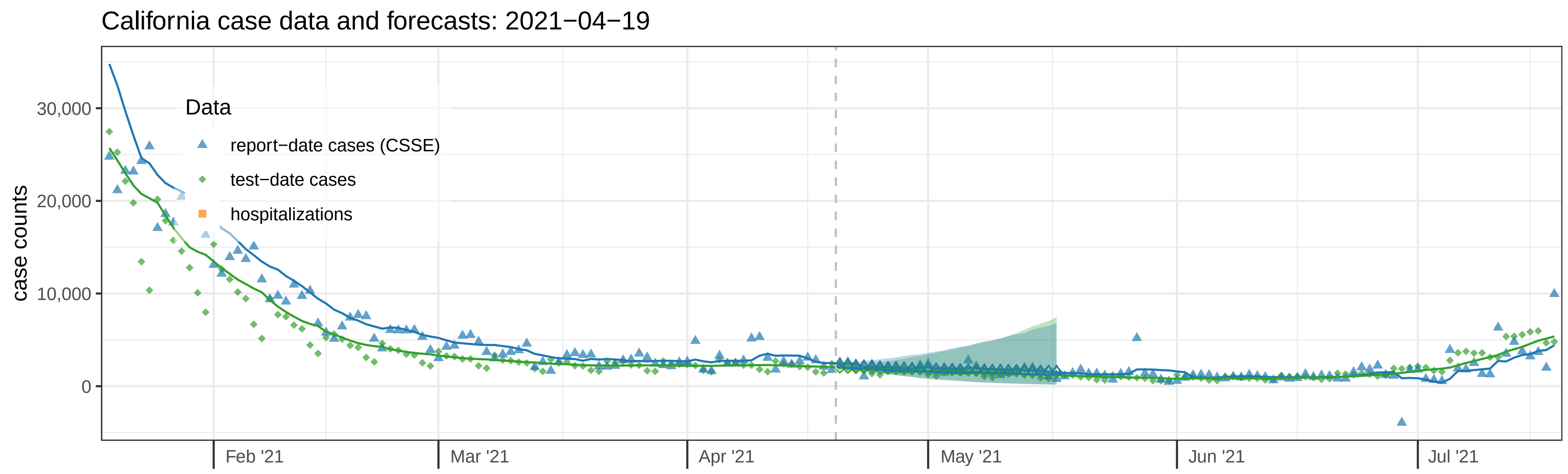
California case data and forecasts: 2021−04−19
case counts
30,000
20,000
10,000
0
Data
report−date cases (CSSE)
test−date cases
hospitalizations
Feb '21
Mar '21
Apr '21
May '21
Jun '21
Jul '21

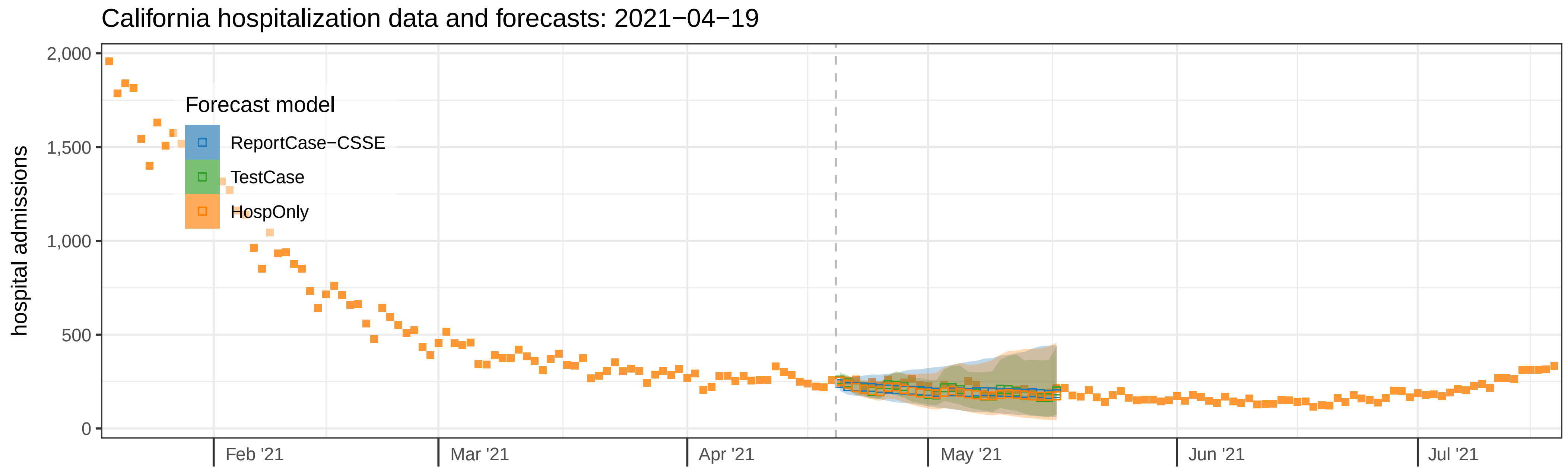
California hospitalization data and forecasts: 2021−04−19
hospital admissions
2,000
1,500
1,000
500
0
Forecast model
ReportCase−CSSE
TestCase
HospOnly
Feb '21
Mar '21
Apr '21
May '21
Jun '21
Jul '21

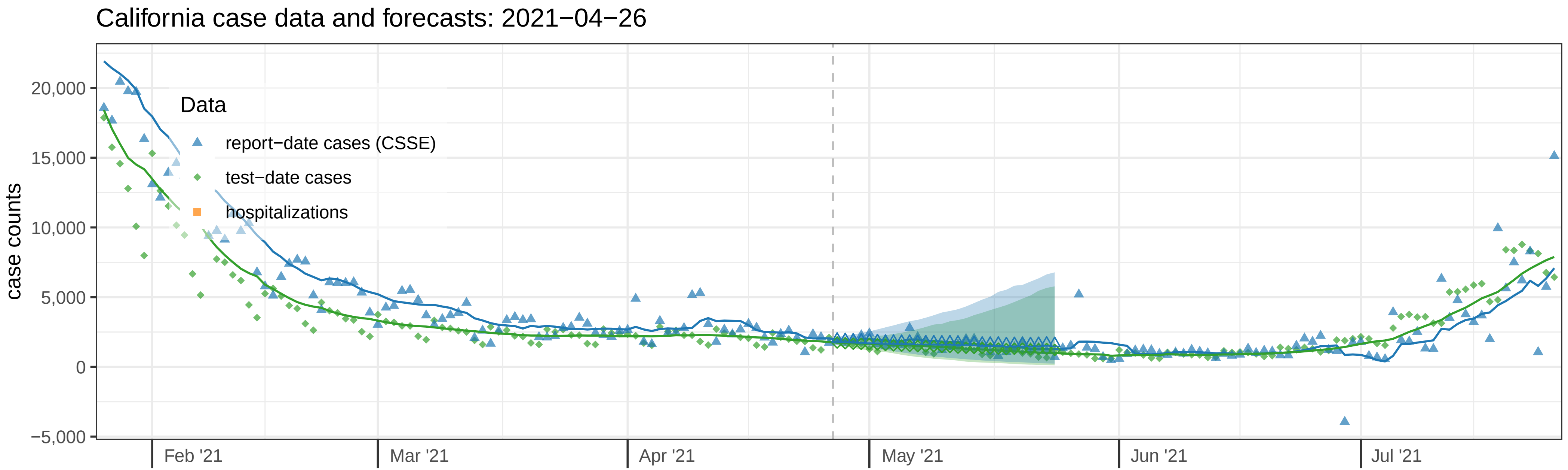

California case data and forecasts: 2021−04−26
case counts
20,000
15,000
10,000
5,000
0
−5,000
Data
report−date cases (CSSE)
test−date cases
hospitalizations
Feb '21
Mar '21
Apr '21
May '21
Jun '21
Jul '21


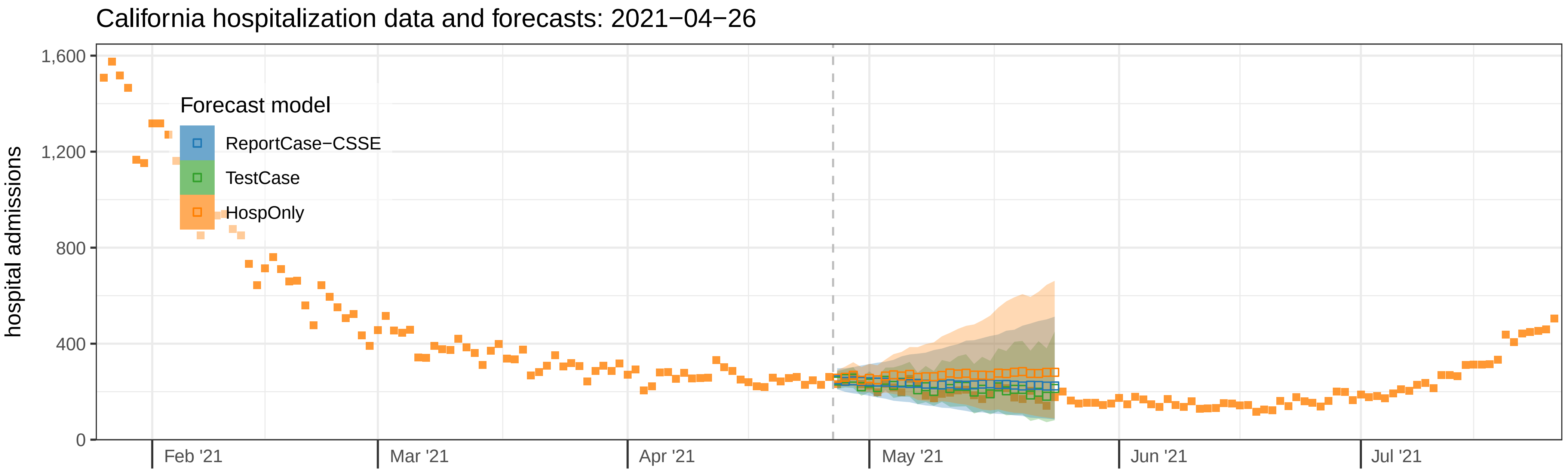

California hospitalization data and forecasts: 2021−04−26
hospital admissions
1,600
1,200
800
400
0
Forecast model
ReportCase−CSSE
TestCase
HospOnly
Feb '21
Mar '21
Apr '21
May '21
Jun '21
Jul '21

California case data and forecasts: 2021−05−03

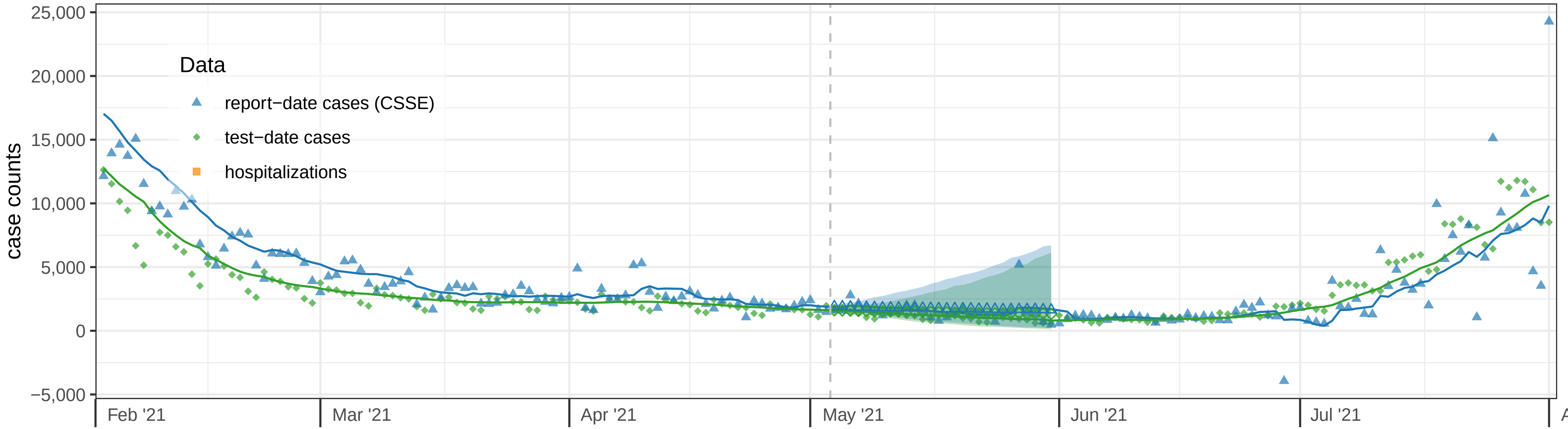


California hospitalization data and forecasts: 2021−05−03

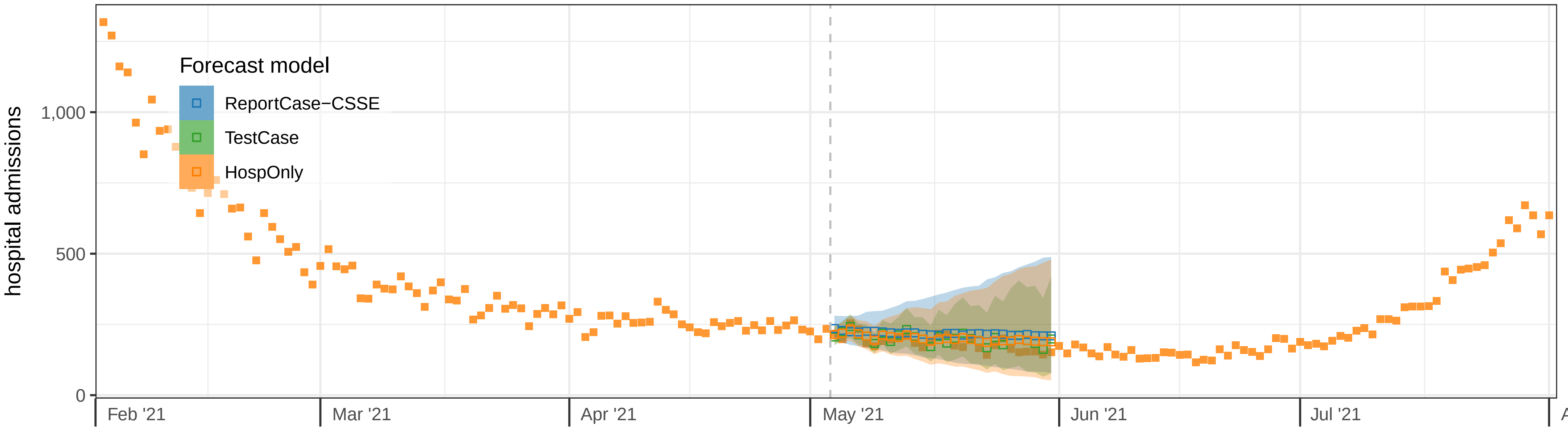

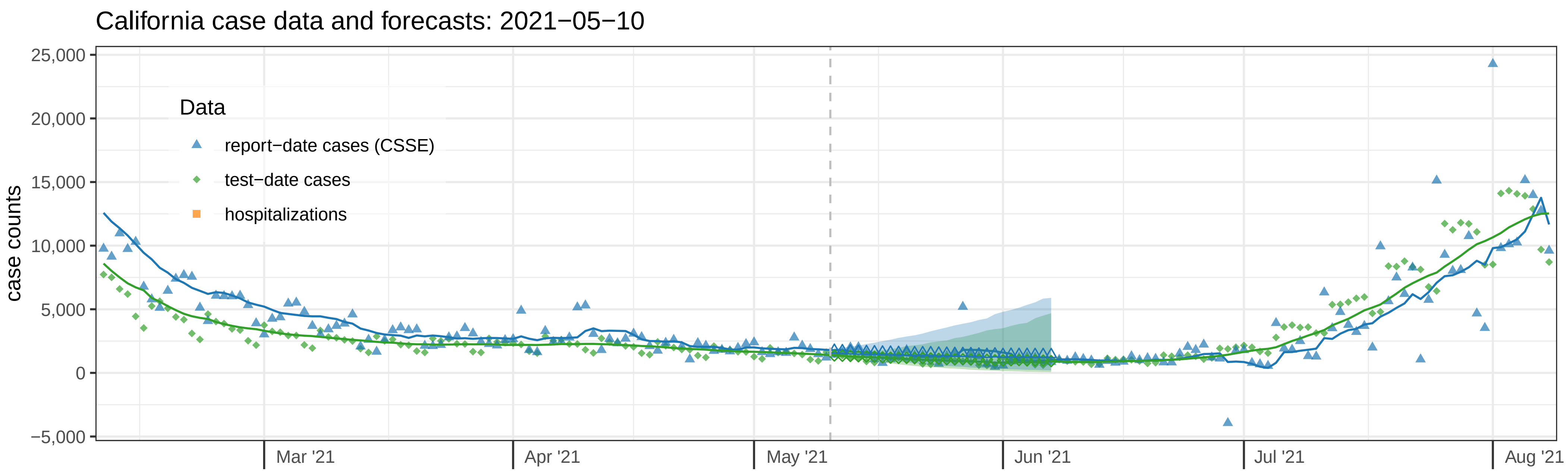
California case data and forecasts: 2021−05−10
case counts
25,000
20,000
15,000
10,000
5,000
0
−5,000
Data
report−date cases (CSSE)
test−date cases
hospitalizations
Mar '21
Apr '21
May '21
Jun '21
Jul '21
Aug '21

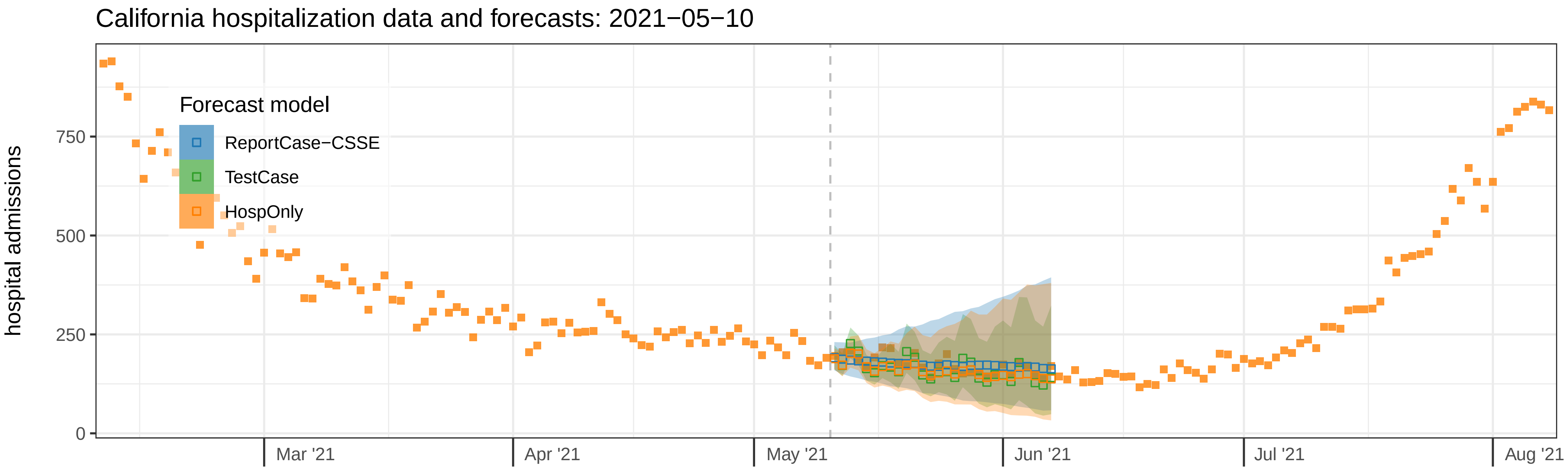
California hospitalization data and forecasts: 2021−05−10
hospital admissions
750
500
250
0
Forecast model
ReportCase−CSSE
TestCase
HospOnly
Mar '21
Apr '21
May '21
Jun '21
Jul '21
Aug '21

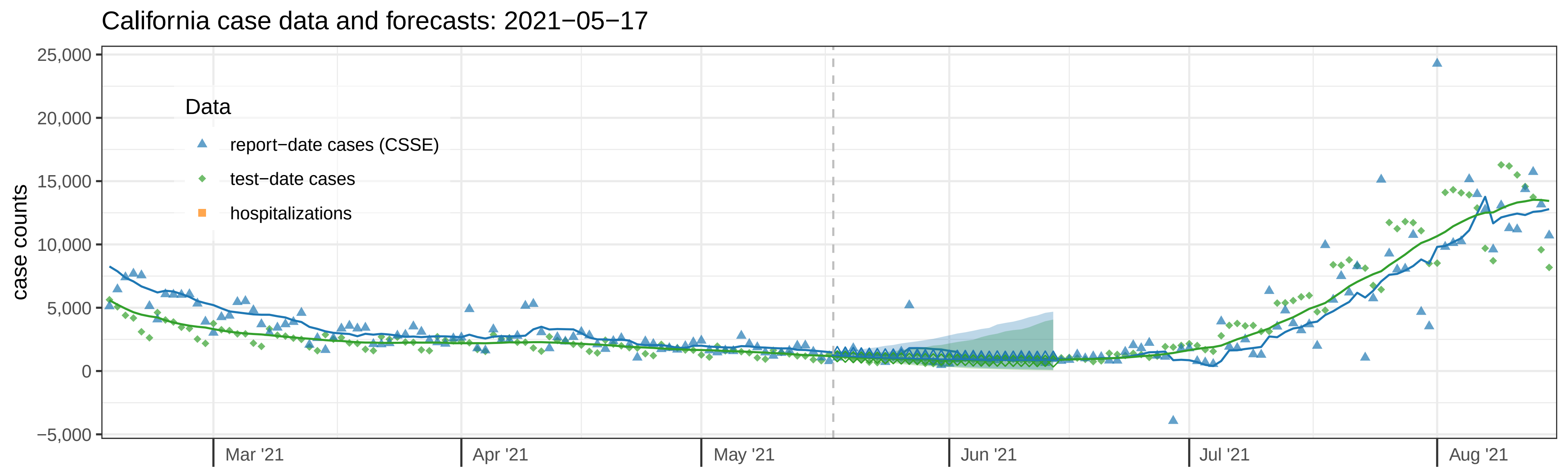
California case data and forecasts: 2021−05−17
case counts
25,000
20,000
15,000
10,000
5,000
0
−5,000
Data
report−date cases (CSSE)
test−date cases
hospitalizations
Mar '21
Apr '21
May '21
Jun '21
Jul '21
Aug '21

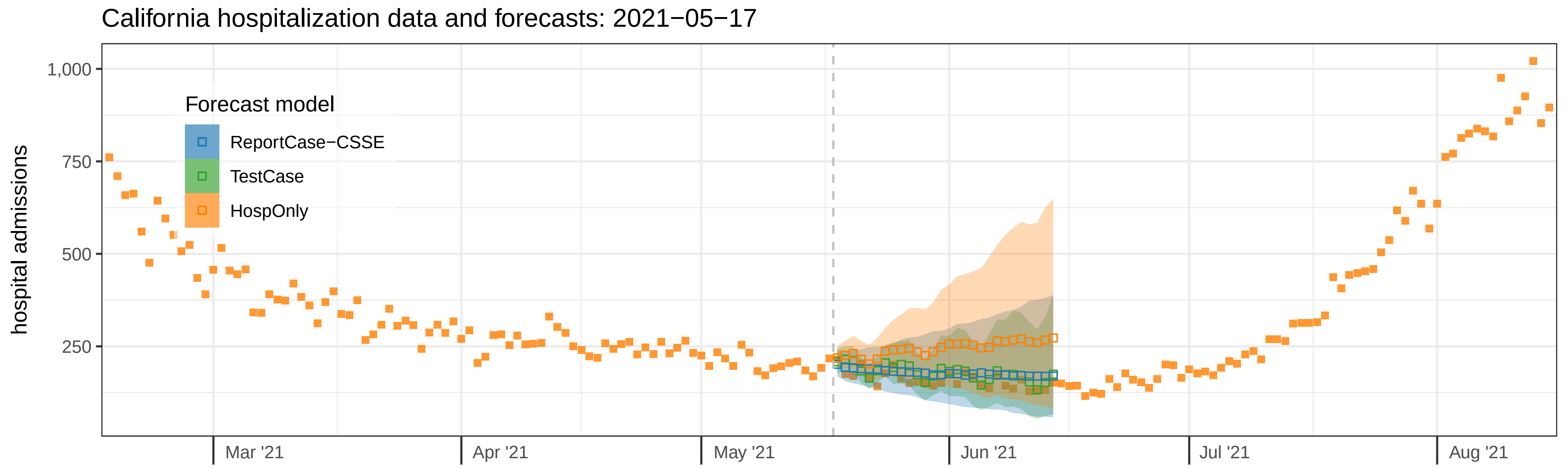
California hospitalization data and forecasts: 2021−05−17
hospital admissions
1,000
750
500
250
Forecast model
ReportCase−CSSE
TestCase
HospOnly
Mar '21
Apr '21
May '21
Jun '21
Jul '21
Aug '21

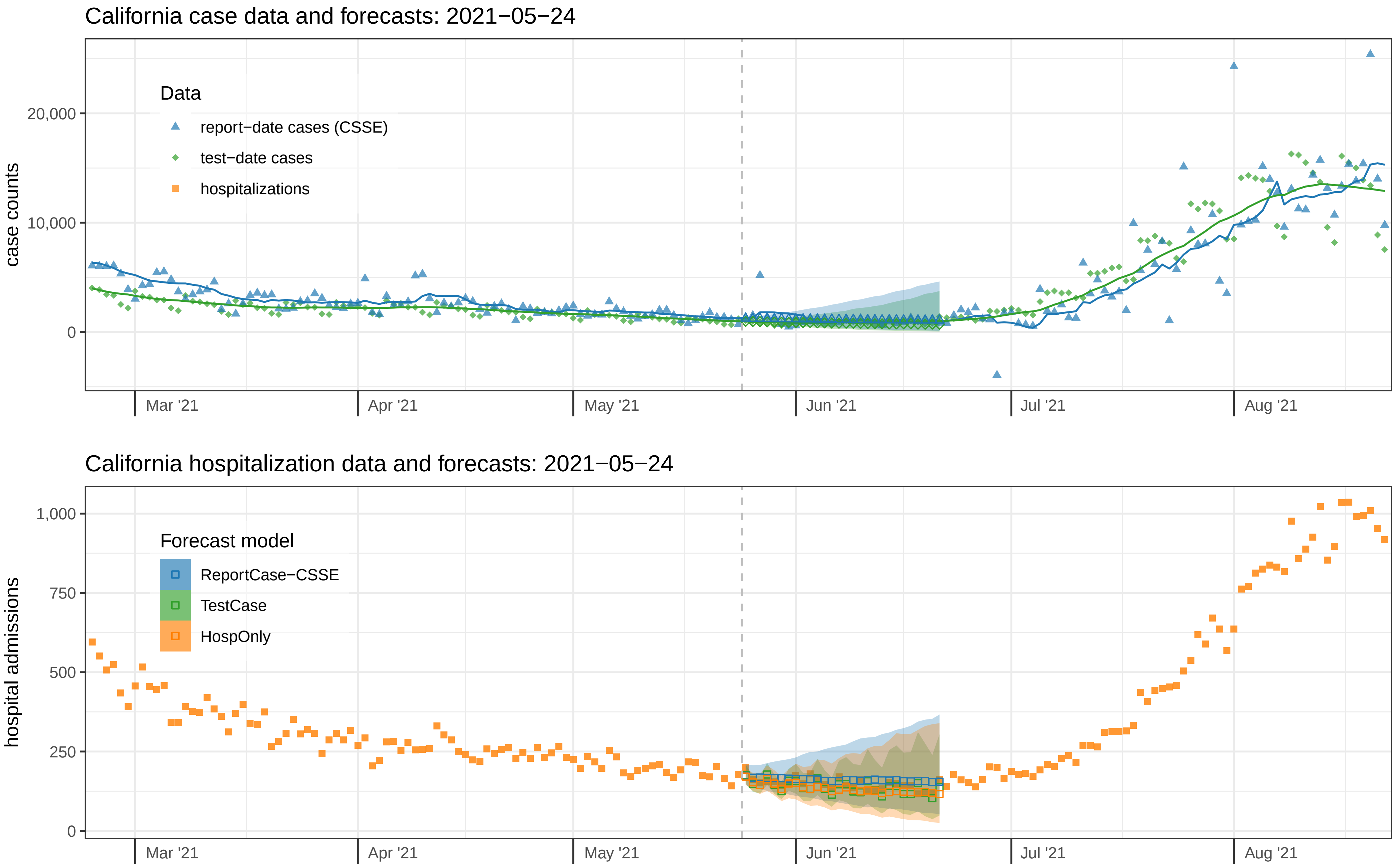
California case data and forecasts: 2021−05−24
case counts
20,000
10,000
0
Data
report−date cases (CSSE)
test−date cases
hospitalizations
Mar '21
Apr '21
May '21
Jun '21
Jul '21
Aug '21
California hospitalization data and forecasts: 2021−05−24
hospital admissions
1,000
750
500
250
0
Forecast model
ReportCase−CSSE
TestCase
HospOnly
Mar '21
Apr '21
May '21
Jun '21
Jul '21
Aug '21

California case data and forecasts: 2021−05−31

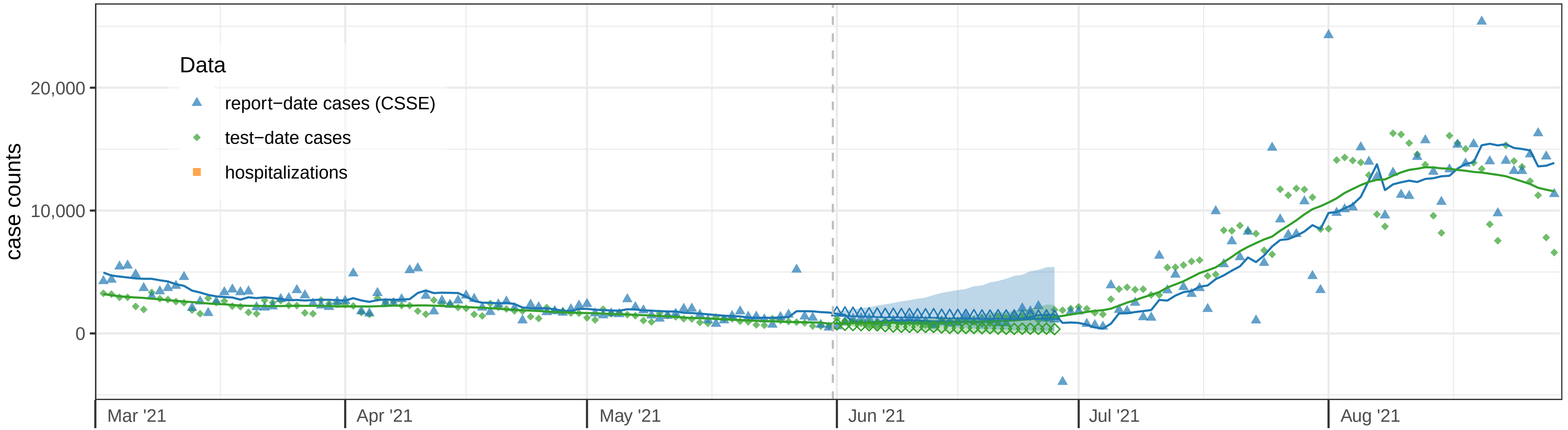


California hospitalization data and forecasts: 2021−05−31

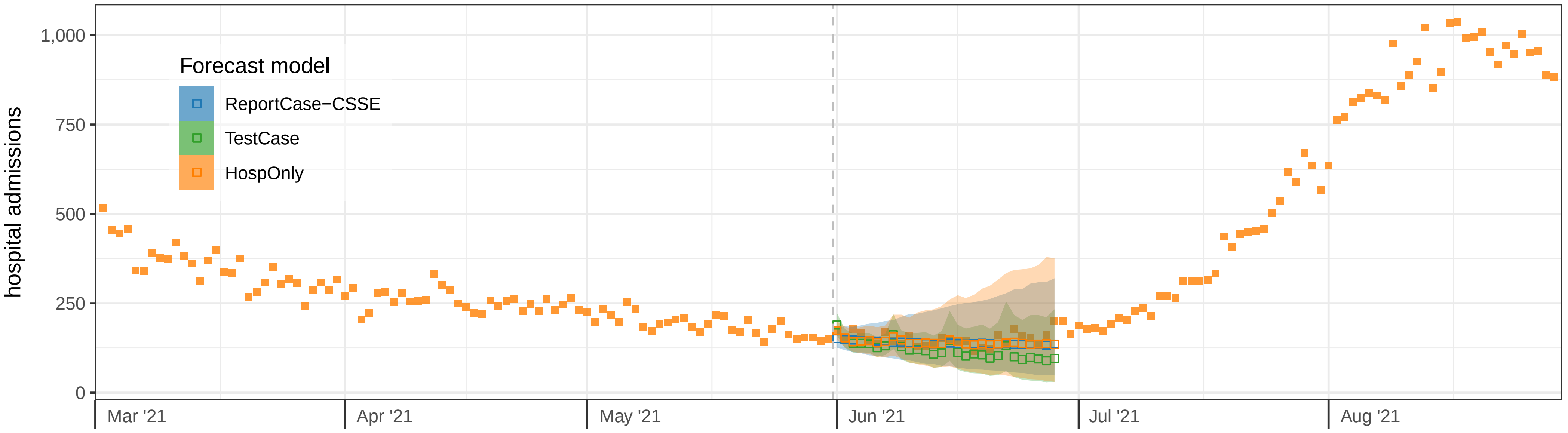

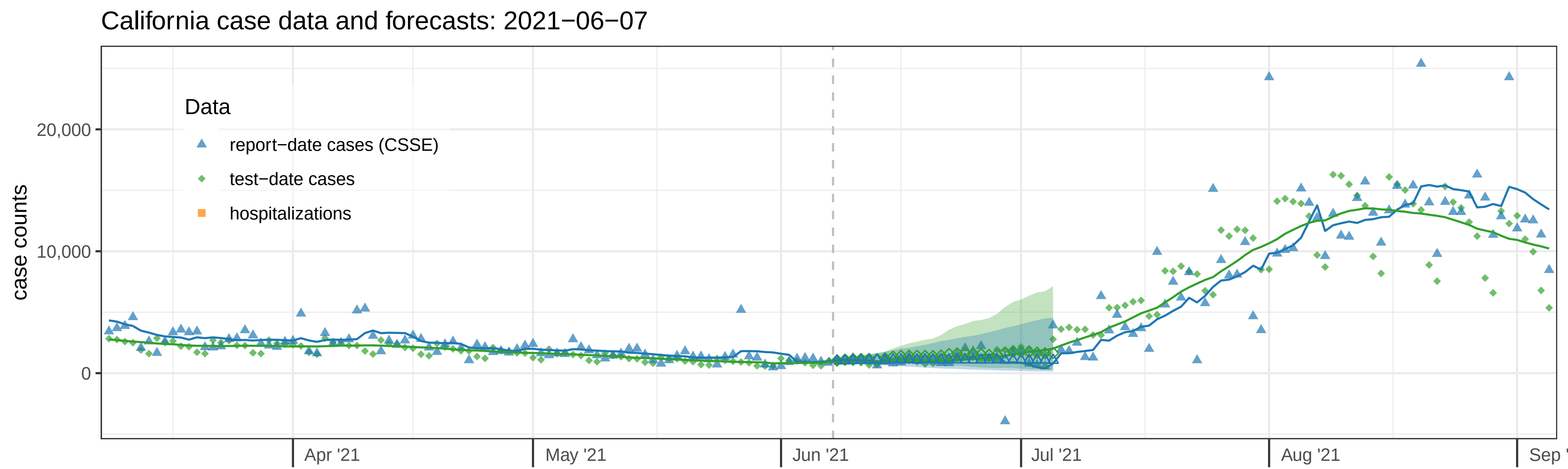
California case data and forecasts: 2021−06−07
case counts
20,000
10,000
0
Data
report−date cases (CSSE)
test−date cases
hospitalizations
Apr '21
May '21
Jun '21
Jul '21
Aug '21
Sep '

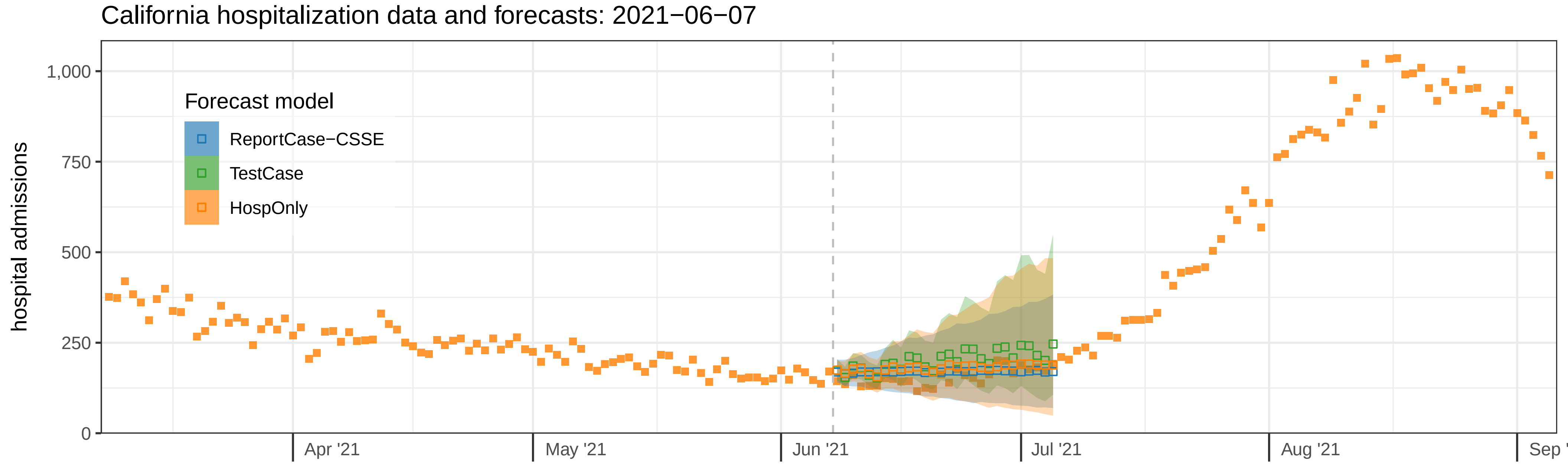
California hospitalization data and forecasts: 2021−06−07
hospital admissions
1,000
750
500
250
0
Forecast model
ReportCase−CSSE
TestCase
HospOnly
Apr '21
May '21
Jun '21
Jul '21
Aug '21
Sep '

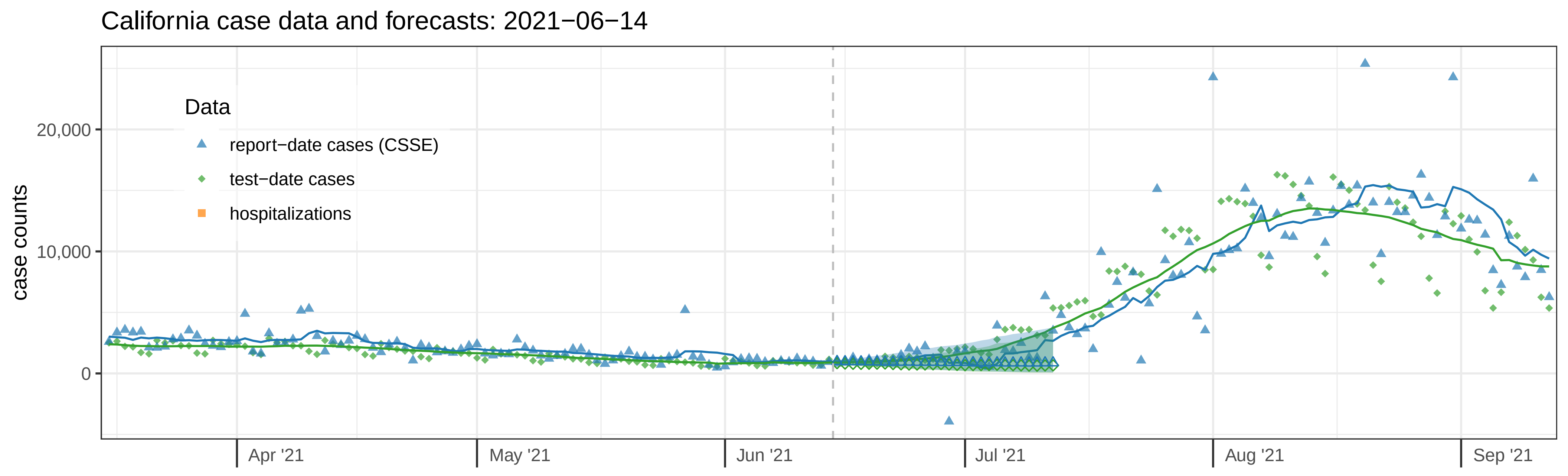
California case data and forecasts: 2021−06−14
case counts
20,000
10,000
0
Data
report−date cases (CSSE)
test−date cases
hospitalizations
Apr '21
May '21
Jun '21
Jul '21
Aug '21
Sep '21

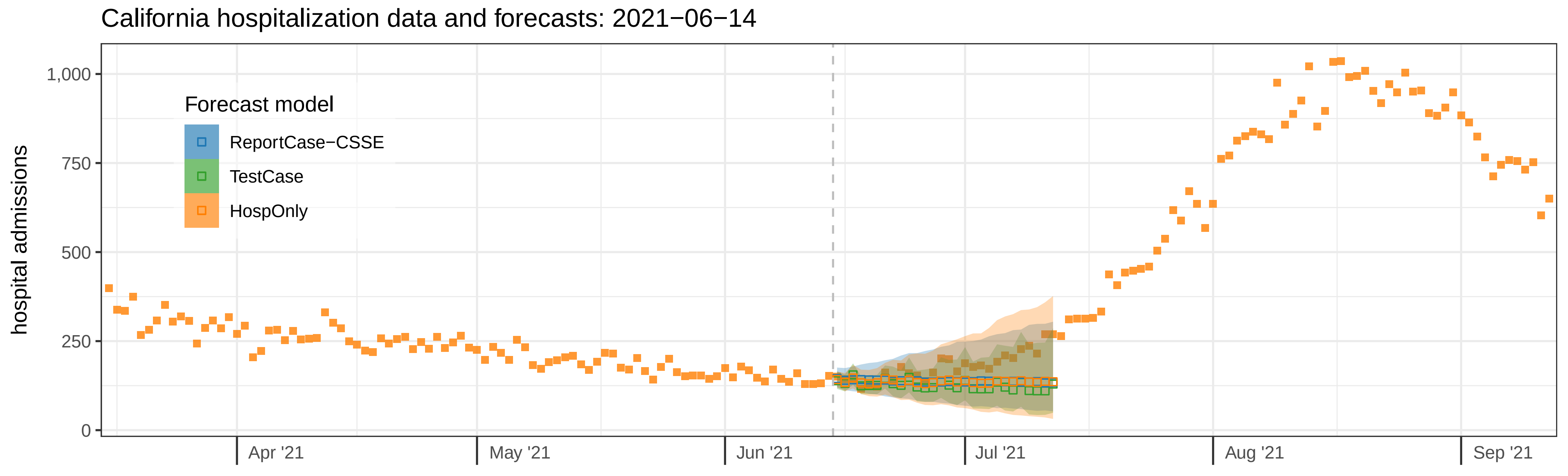
California hospitalization data and forecasts: 2021−06−14
hospital admissions
1,000
750
500
250
0
Forecast model
ReportCase−CSSE
TestCase
HospOnly
Apr '21
May '21
Jun '21
Jul '21
Aug '21
Sep '21

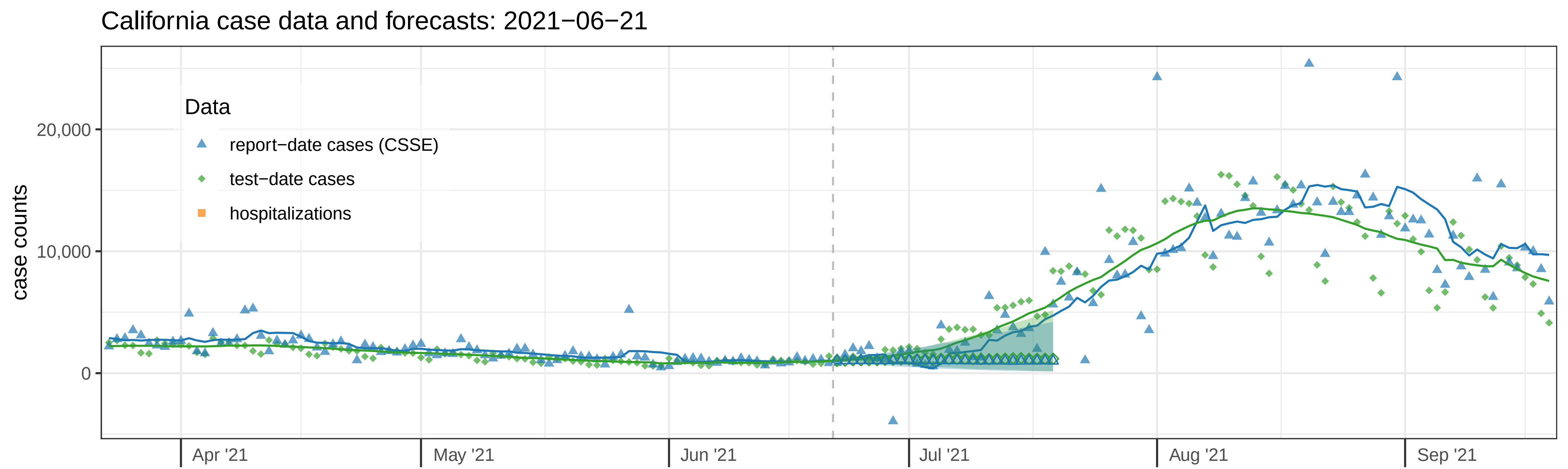

California case data and forecasts: 2021−06−21
Data
report−date cases (CSSE)
test−date cases
hospitalizations
case counts
20,000
10,000
0
Apr '21
May '21
Jun '21
Jul '21
Aug '21
Sep '21


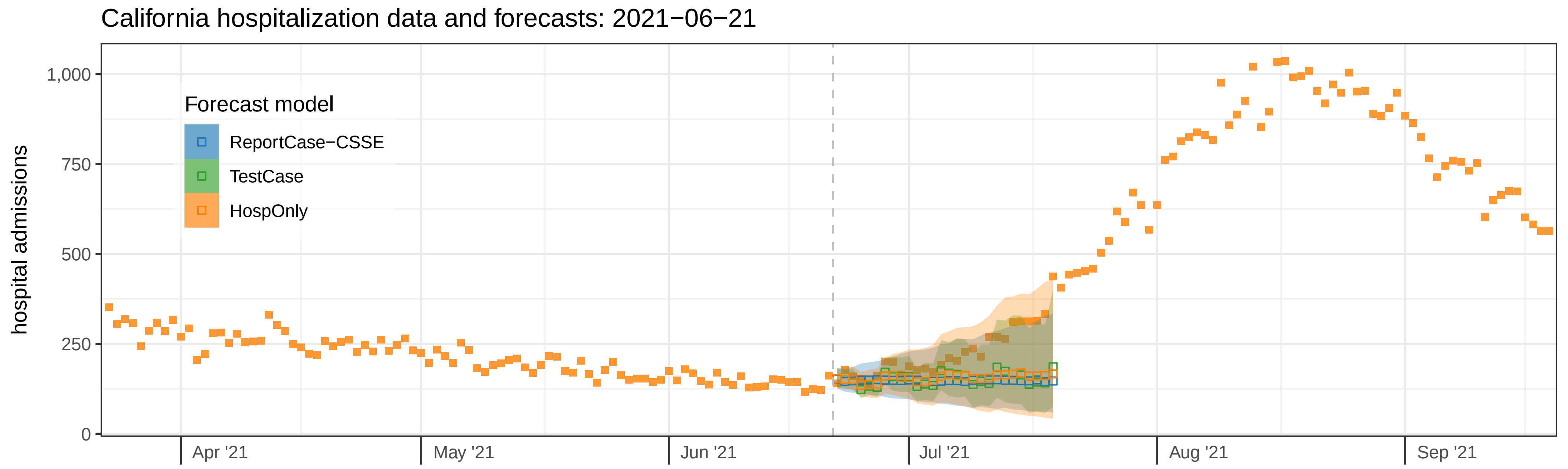

California hospitalization data and forecasts: 2021−06−21
Forecast model
ReportCase−CSSE
TestCase
HospOnly
hospital admissions
1,000
750
500
250
0
Apr '21
May '21
Jun '21
Jul '21
Aug '21
Sep '21

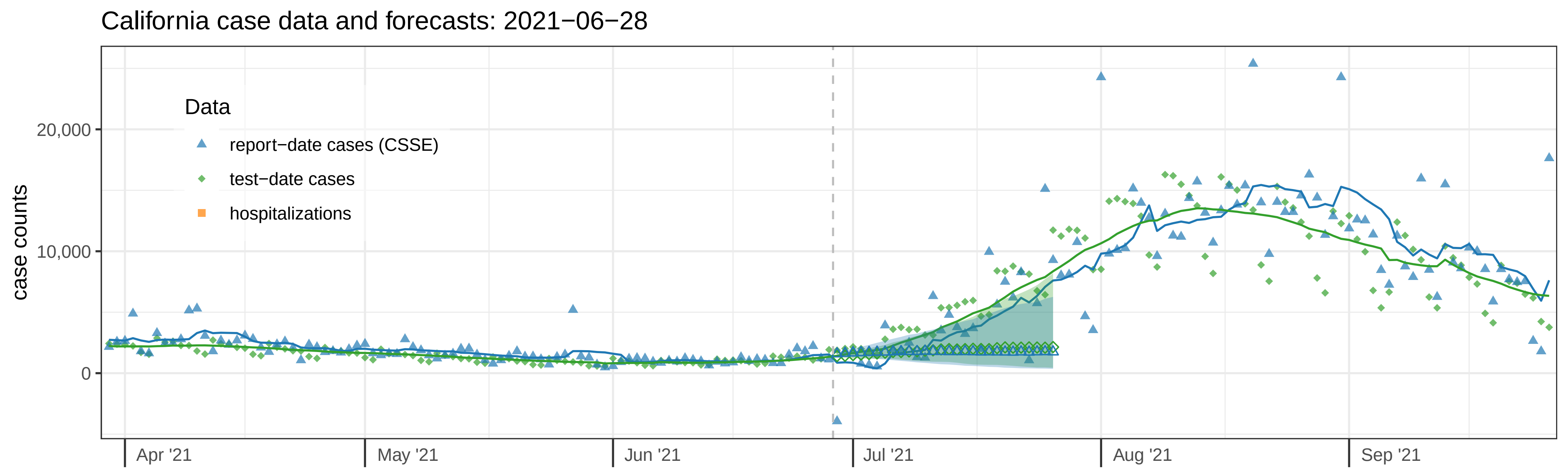
California case data and forecasts: 2021−06−28
Data
report−date cases (CSSE)
test−date cases
hospitalizations
case counts
20,000
10,000
0
Apr '21
May '21
Jun '21
Jul '21
Aug '21
Sep '21

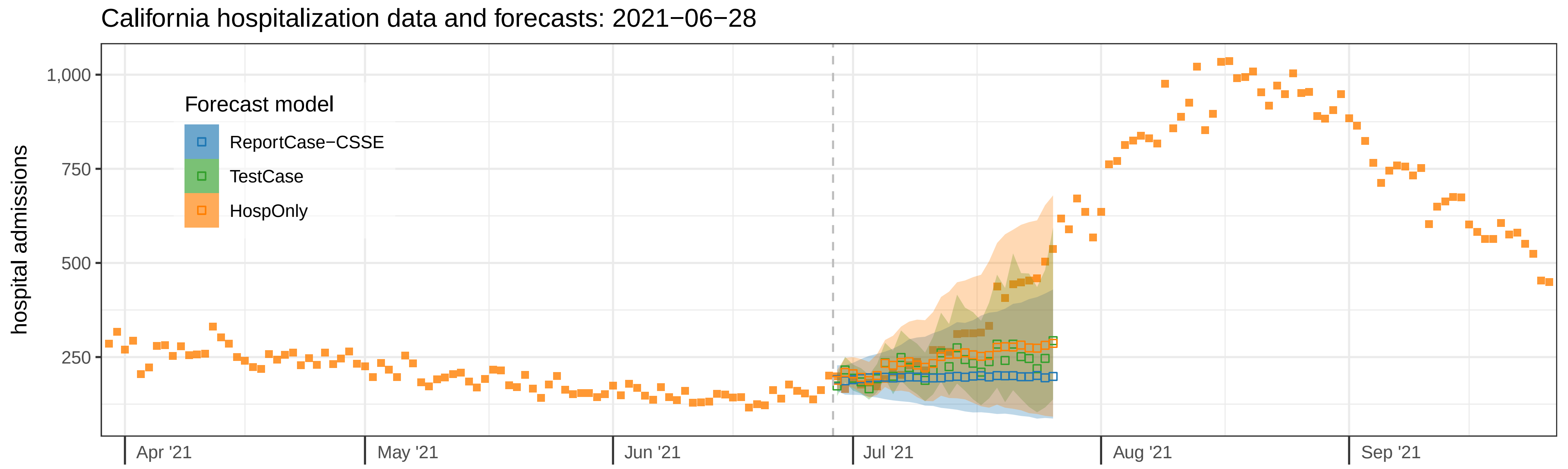
California hospitalization data and forecasts: 2021−06−28
Forecast model
ReportCase−CSSE
TestCase
HospOnly
hospital admissions
1,000
750
500
250
Apr '21
May '21
Jun '21
Jul '21
Aug '21
Sep '21

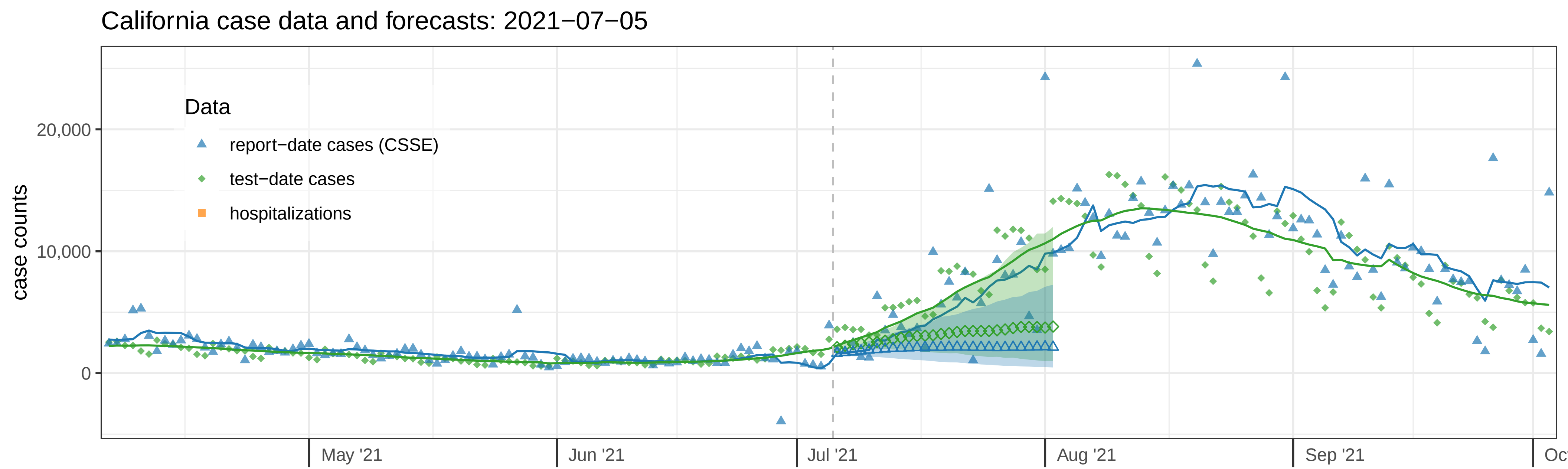
California case data and forecasts: 2021−07−05
case counts
20,000
10,000
0
Data
report−date cases (CSSE)
test−date cases
hospitalizations
May '21
Jun '21
Jul '21
Aug '21
Sep '21

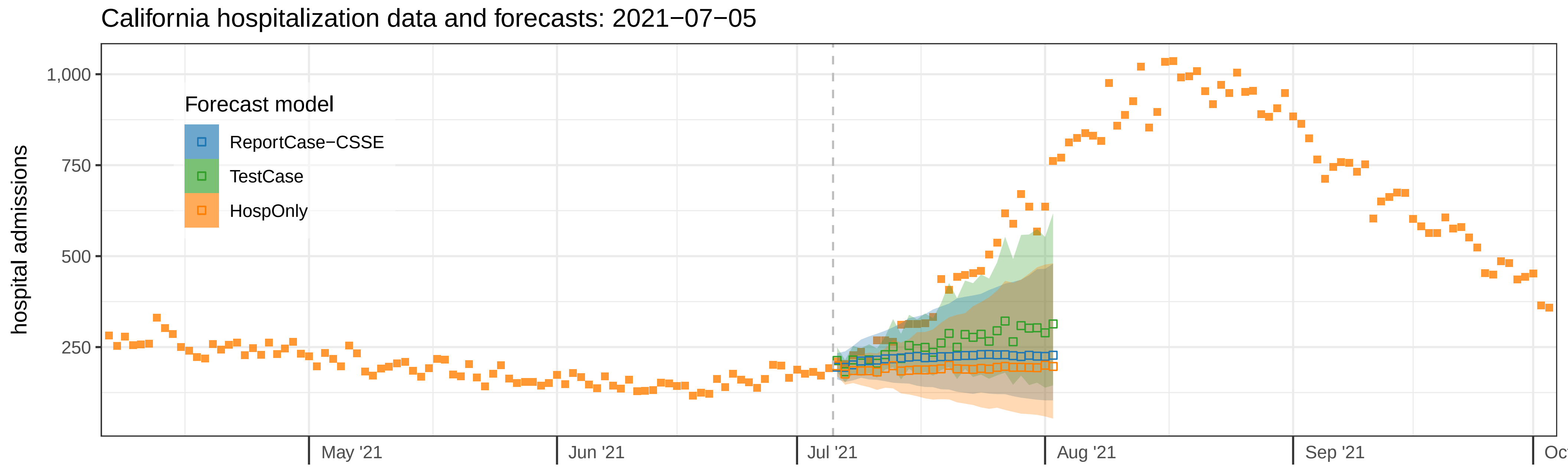
California hospitalization data and forecasts: 2021−07−05
hospital admissions
1,000
750
500
250
Forecast model
ReportCase−CSSE
TestCase
HospOnly
May '21
Jun '21
Jul '21
Aug '21
Sep '21

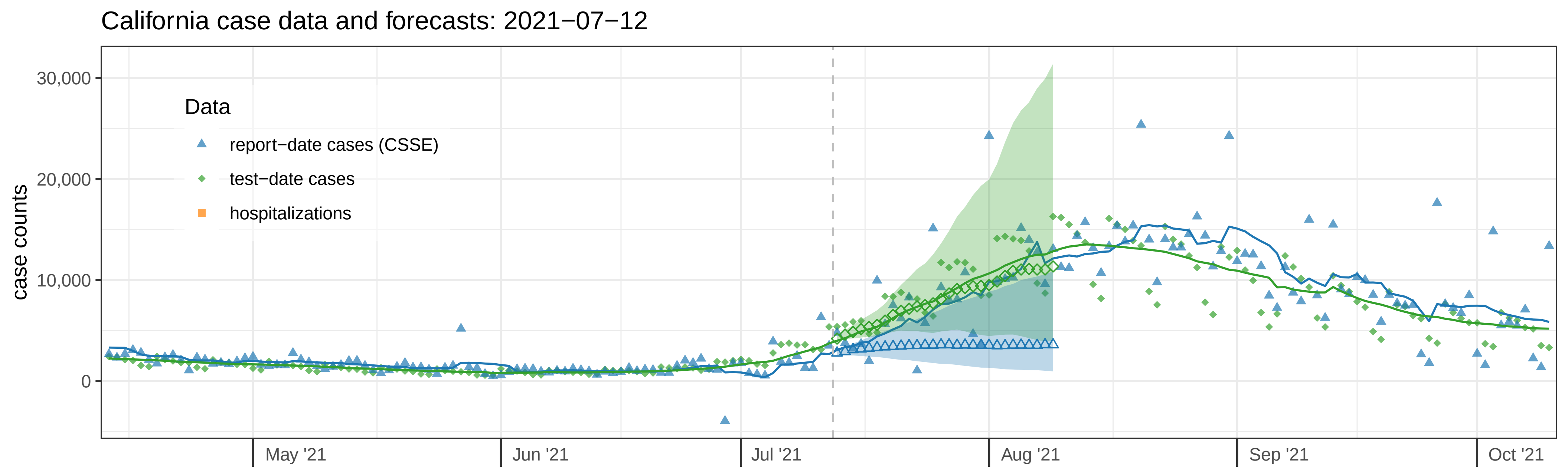

California case data and forecasts: 2021−07−12
case counts
30,000
20,000
10,000
0
Data
report−date cases (CSSE)
test−date cases
hospitalizations
May '21
Jun '21
Jul '21
Aug '21
Sep '21
Oct '21


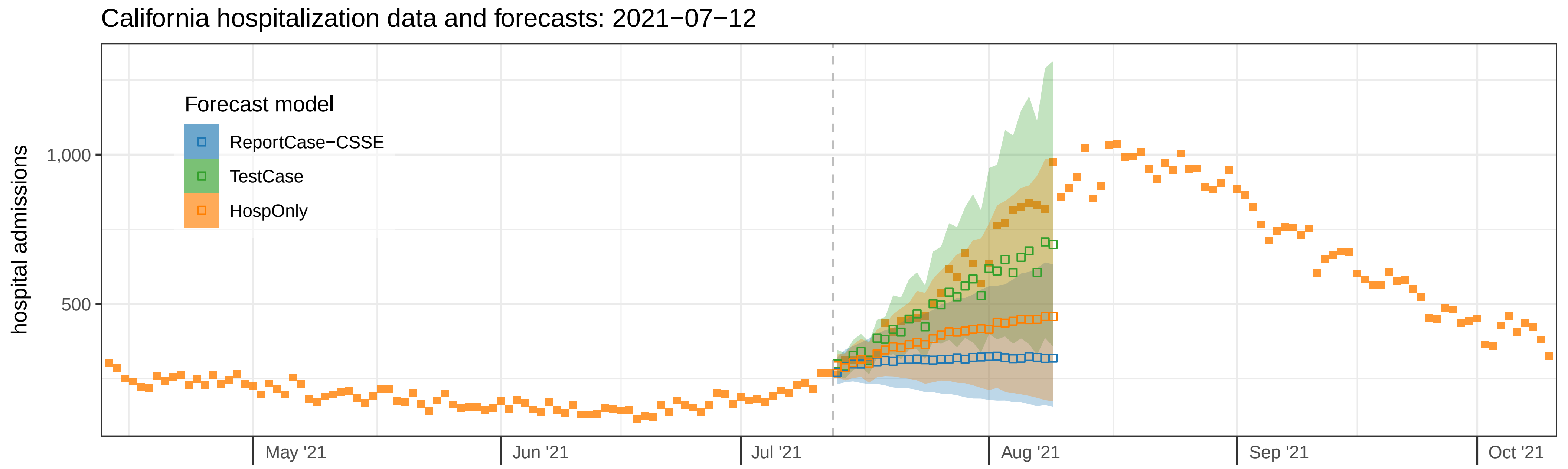

California hospitalization data and forecasts: 2021−07−12
hospital admissions
1,000
500
Forecast model
ReportCase−CSSE
TestCase
HospOnly
May '21
Jun '21
Jul '21
Aug '21
Sep '21
Oct '21

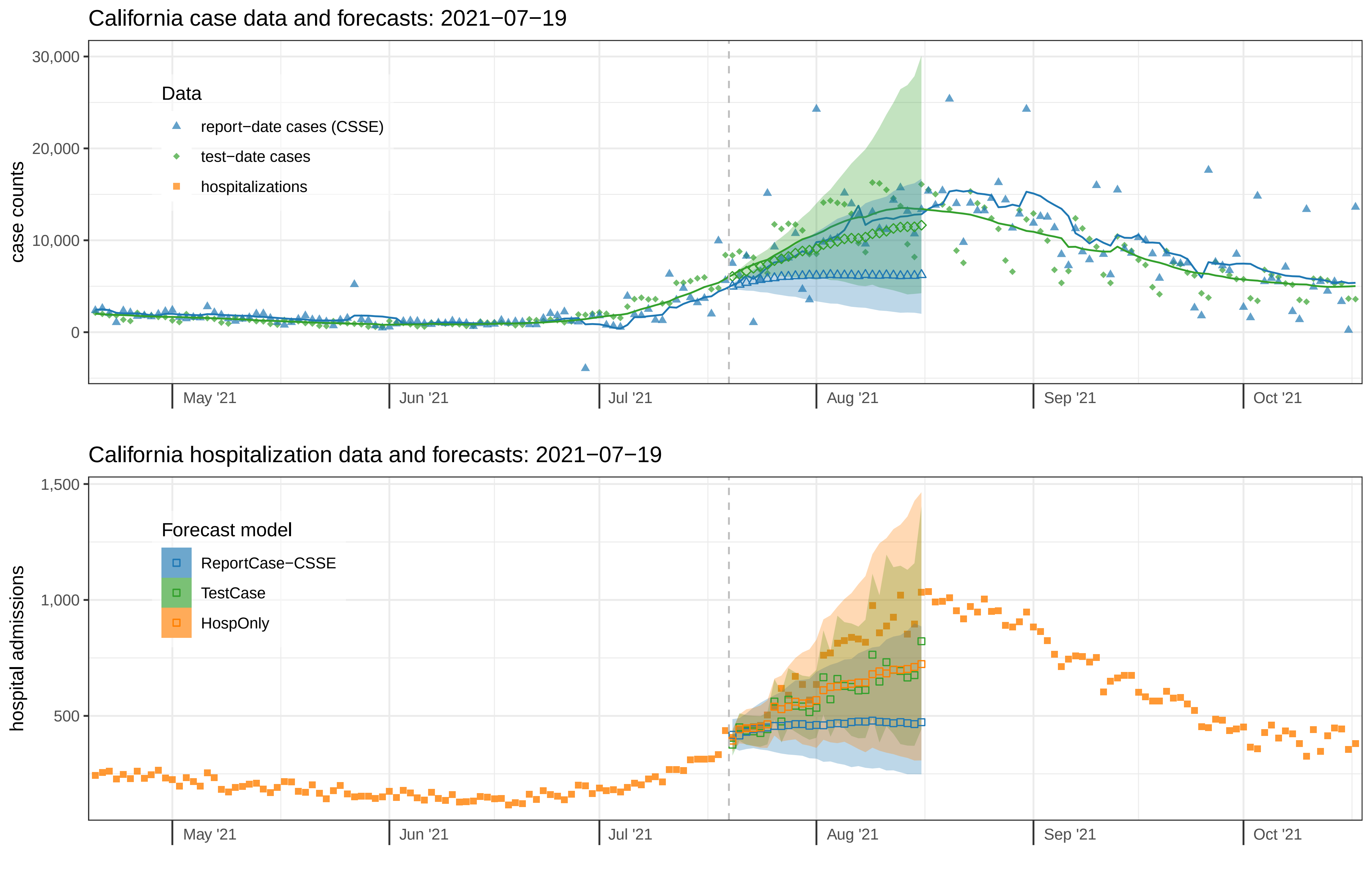
California case data and forecasts: 2021−07−19
case counts
30,000
20,000
10,000
0
Data
report−date cases (CSSE)
test−date cases
hospitalizations
May '21
Jun '21
Jul '21
Aug '21
Sep '21
Oct '21
California hospitalization data and forecasts: 2021−07−19
hospital admissions
1,500
1,000
500
Forecast model
ReportCase−CSSE
TestCase
HospOnly
May '21
Jun '21
Jul '21
Aug '21
Sep '21
Oct '21

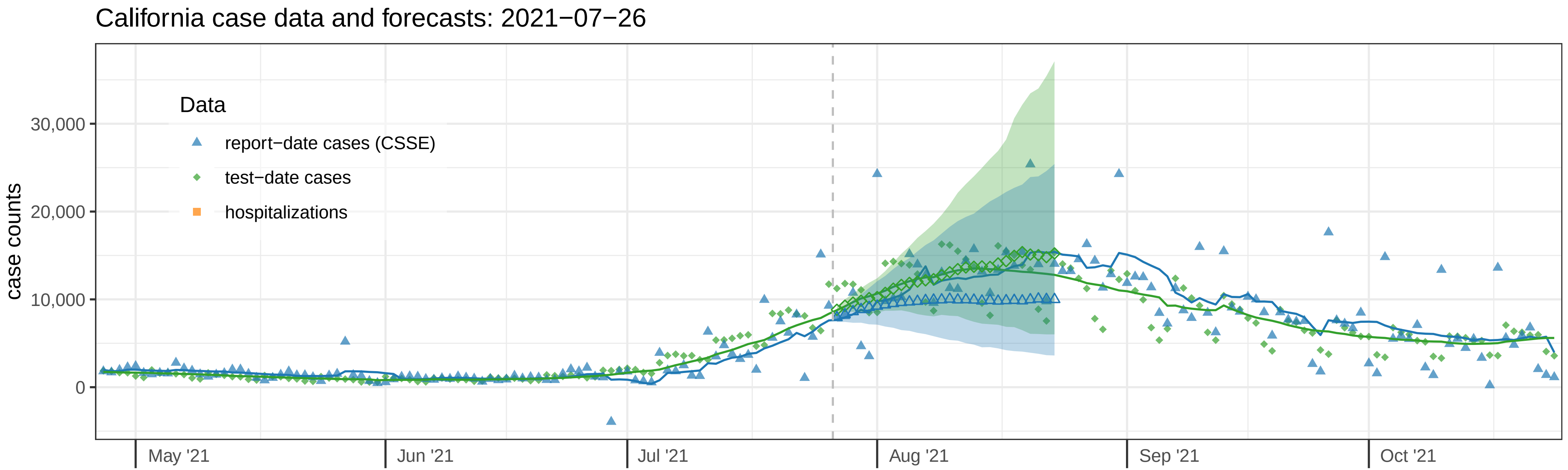
California case data and forecasts: 2021−07−26
case counts
30,000
20,000
10,000
0
Data
report−date cases (CSSE)
test−date cases
hospitalizations
May '21
Jun '21
Jul '21
Aug '21
Sep '21
Oct '21

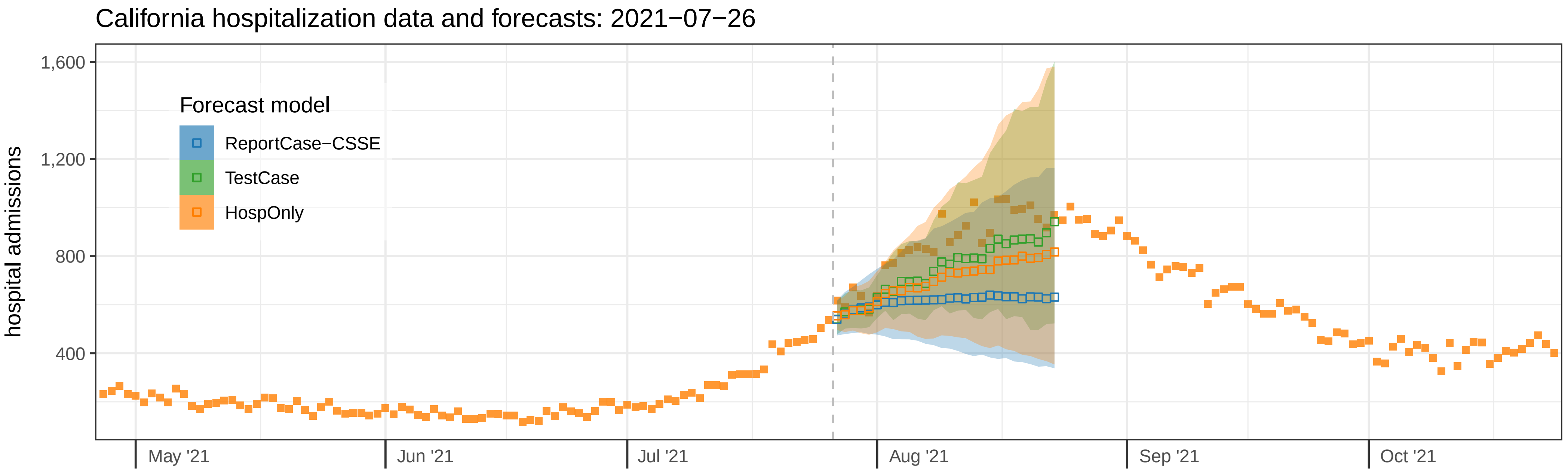
California hospitalization data and forecasts: 2021−07−26
hospital admissions
1,600
1,200
800
400
Forecast model
ReportCase−CSSE
TestCase
HospOnly
May '21
Jun '21
Jul '21
Aug '21
Sep '21
Oct '21

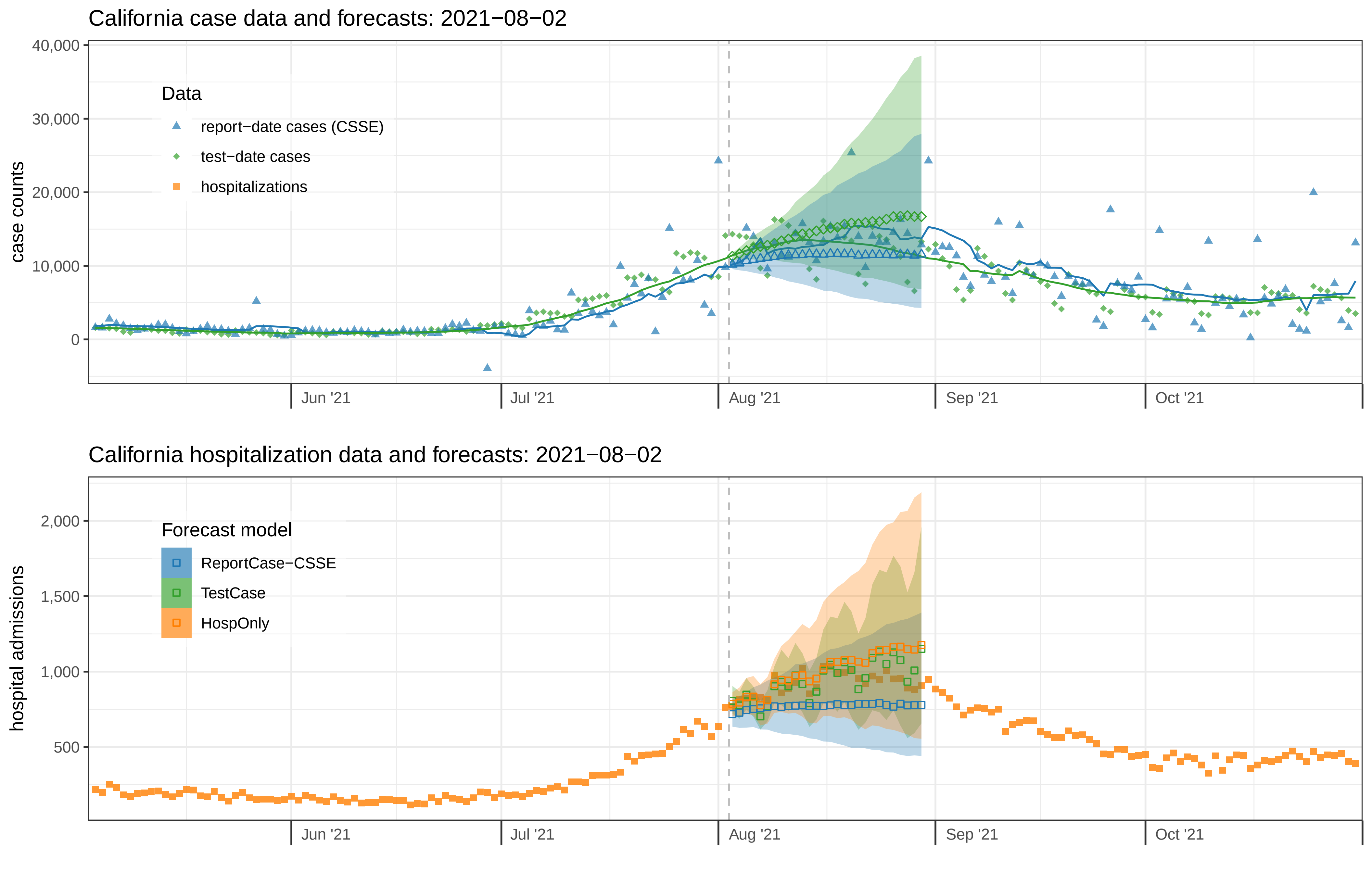
California case data and forecasts: 2021−08−02
case counts
40,000
30,000
20,000
10,000
0
Data
report−date cases (CSSE)
test−date cases
hospitalizations
Jun '21
Jul '21
Aug '21
Sep '21
Oct '21
California hospitalization data and forecasts: 2021−08−02
hospital admissions
2,000
1,500
1,000
500
Forecast model
ReportCase−CSSE
TestCase
HospOnly
Jun '21
Jul '21
Aug '21
Sep '21
Oct '21

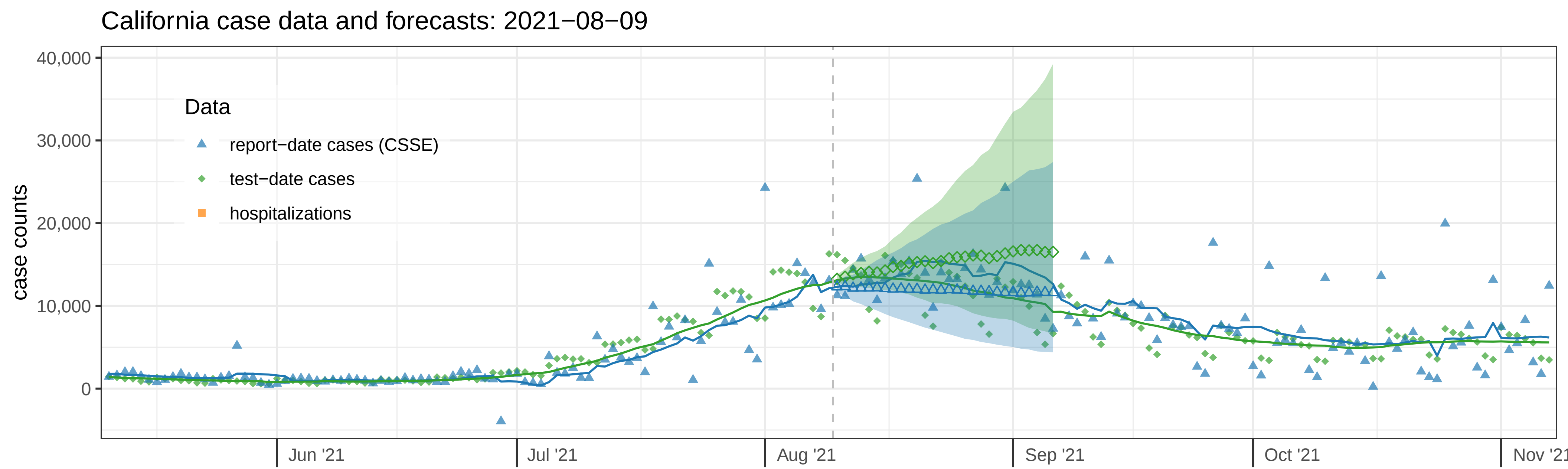
California case data and forecasts: 2021−08−09
case counts
40,000
30,000
20,000
10,000
0
Data
report−date cases (CSSE)
test−date cases
hospitalizations
Jun '21
Jul '21
Aug '21
Sep '21
Oct '21
Nov '21

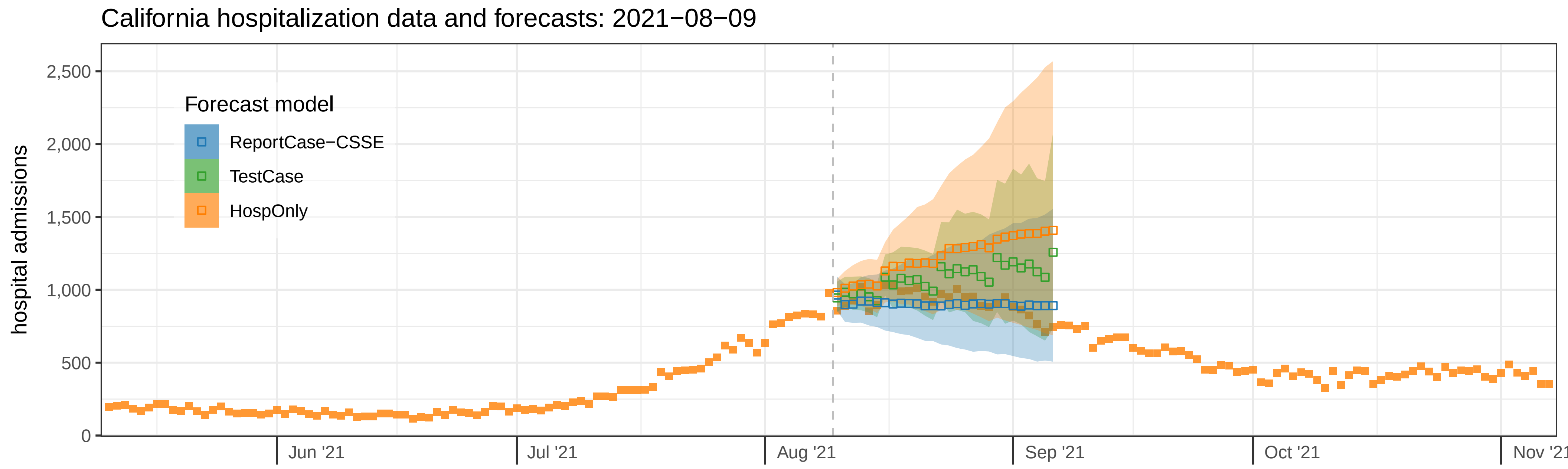
California hospitalization data and forecasts: 2021−08−09
hospital admissions
2,500
2,000
1,500
1,000
500
0
Forecast model
ReportCase−CSSE
TestCase
HospOnly
Jun '21
Jul '21
Aug '21
Sep '21
Oct '21
Nov '21

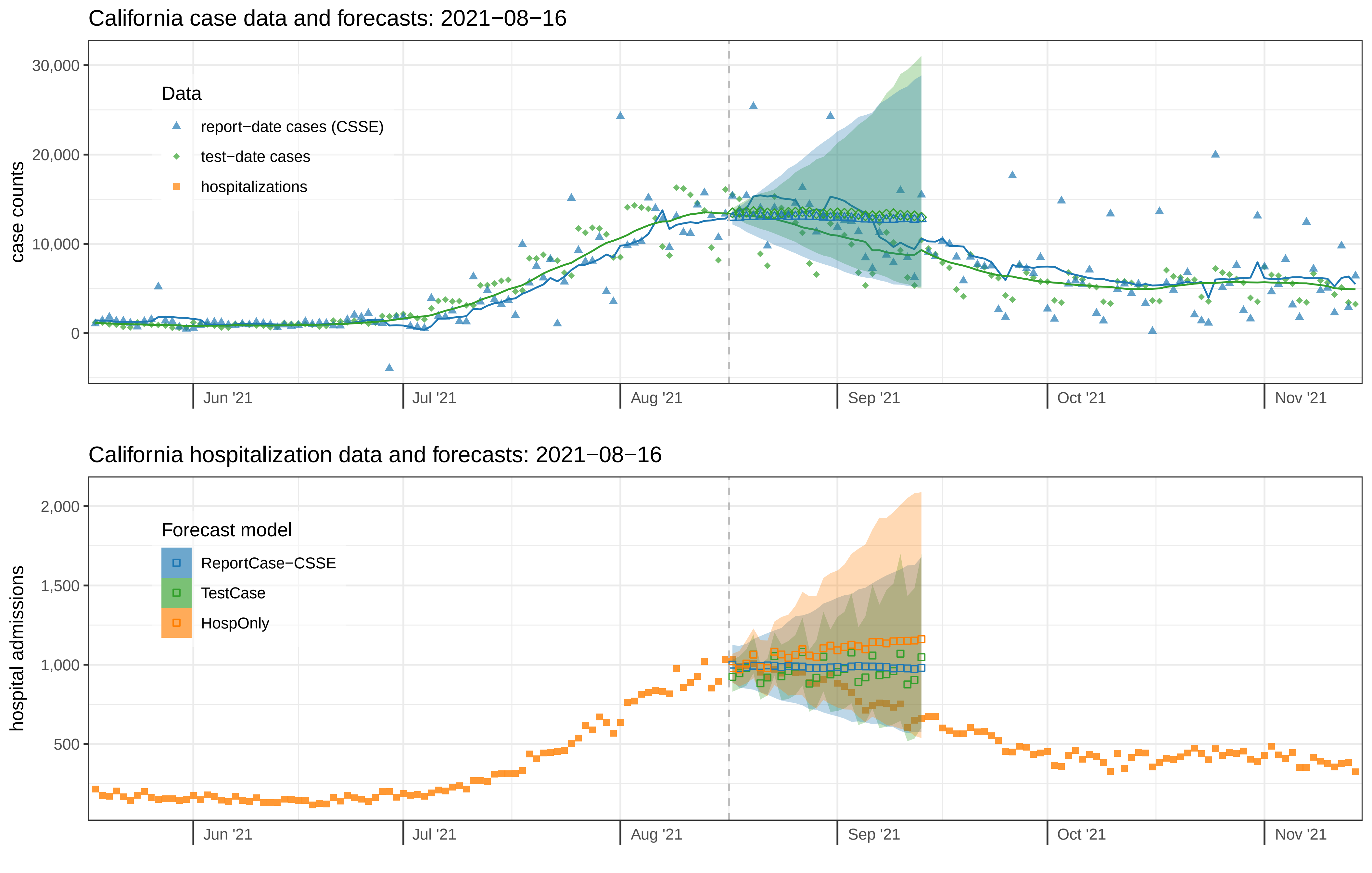
California case data and forecasts: 2021−08−16
case counts
30,000
20,000
10,000
0
Data
report−date cases (CSSE)
test−date cases
hospitalizations
Jun '21
Jul '21
Aug '21
Sep '21
Oct '21
Nov '21
California hospitalization data and forecasts: 2021−08−16
hospital admissions
2,000
1,500
1,000
500
Forecast model
ReportCase−CSSE
TestCase
HospOnly
Jun '21
Jul '21
Aug '21
Sep '21
Oct '21
Nov '21

California case data and forecasts: 2021−08−23

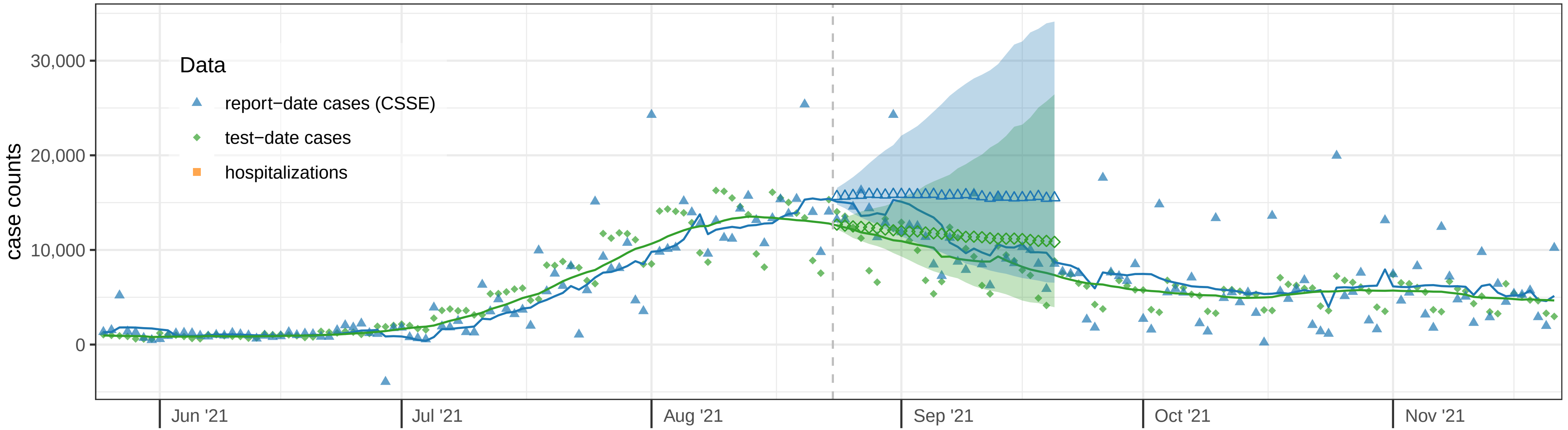


California hospitalization data and forecasts: 2021−08−23

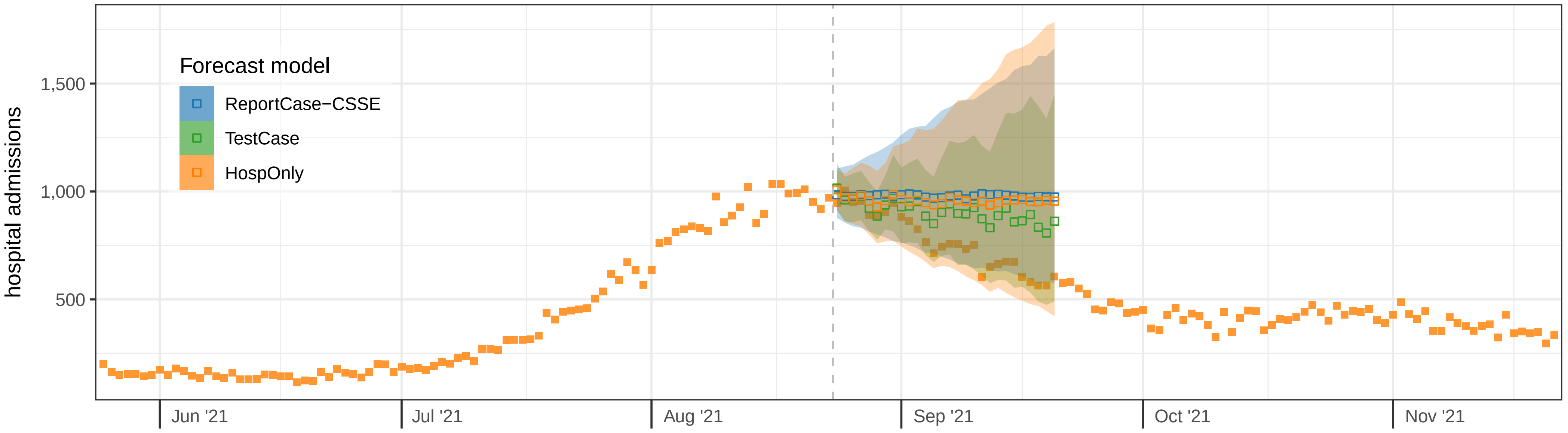

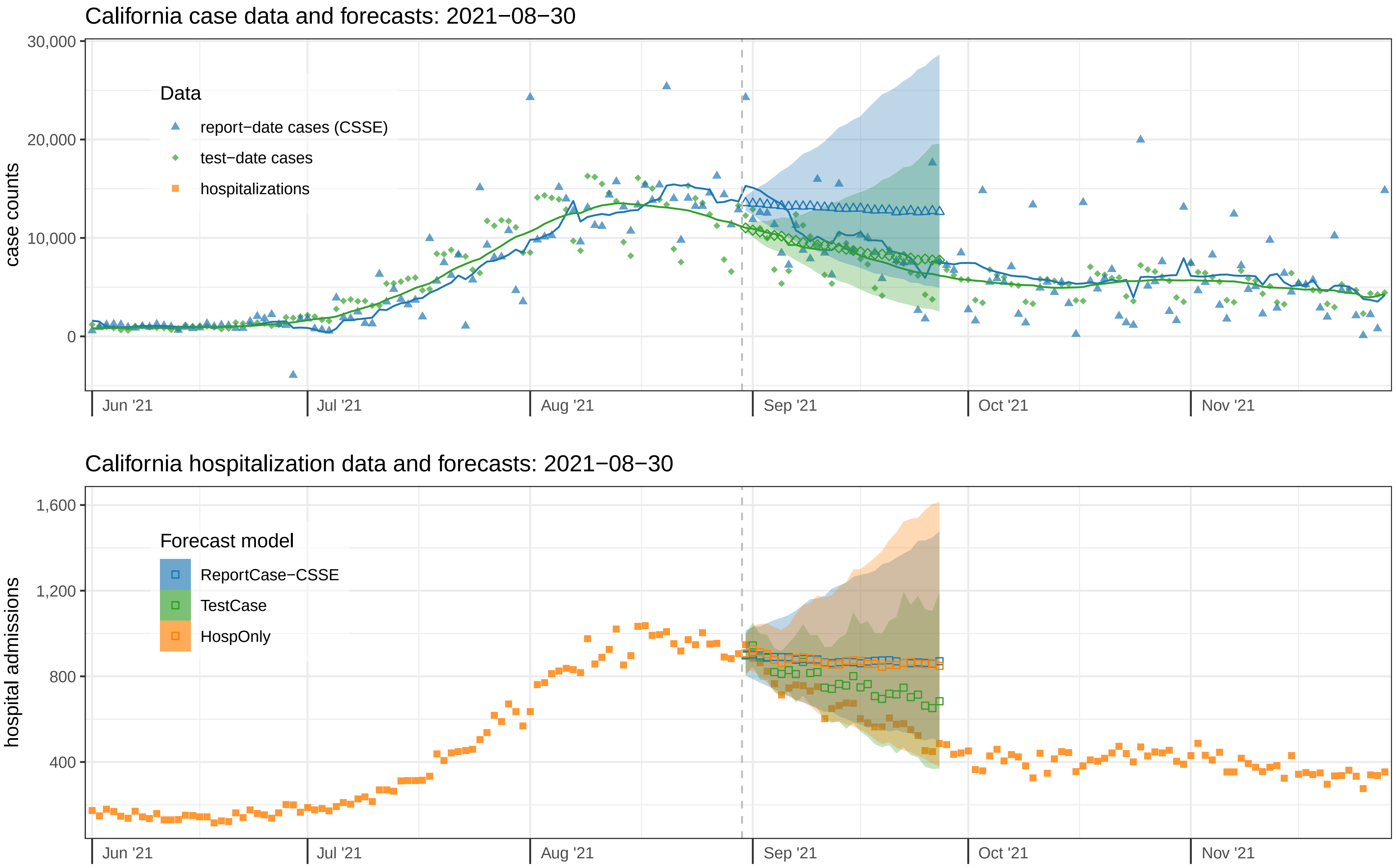
California case data and forecasts: 2021−08−30
case counts
30,000
20,000
10,000
0
Data
report−date cases (CSSE)
test−date cases
hospitalizations
Jun '21
Jul '21
Aug '21
Sep '21
Oct '21
Nov '21
California hospitalization data and forecasts: 2021−08−30
hospital admissions
1,600
1,200
800
400
Forecast model
ReportCase−CSSE
TestCase
HospOnly
Jun '21
Jul '21
Aug '21
Sep '21
Oct '21
Nov '21

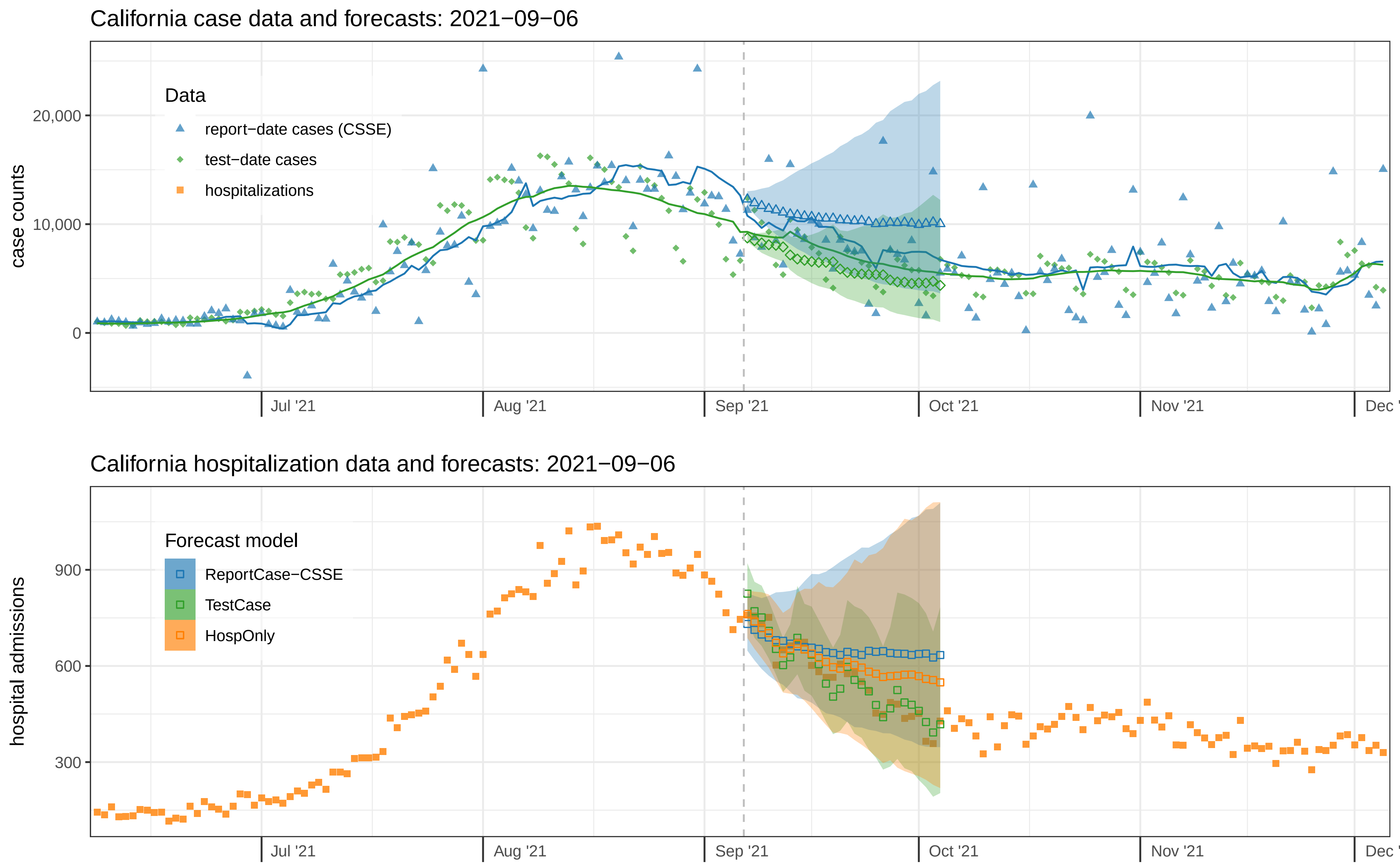

California case data and forecasts: 2021−09−06
case counts
20,000
10,000
0
Data
report−date cases (CSSE)
test−date cases
hospitalizations
Jul '21
Aug '21
Sep '21
Oct '21
Nov '21
Dec '
California hospitalization data and forecasts: 2021−09−06
hospital admissions
900
600
300
Forecast model
ReportCase−CSSE
TestCase
HospOnly
Jul '21
Aug '21
Sep '21
Oct '21
Nov '21
Dec '

### California case data and forecasts: 2021−09−13

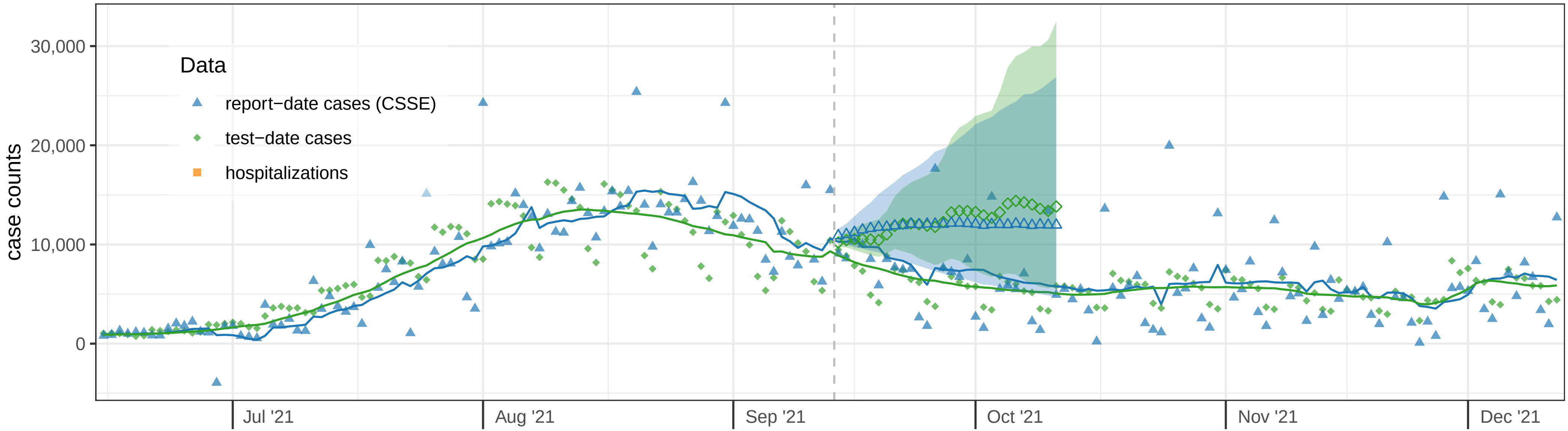


### California hospitalization data and forecasts: 2021−09−13

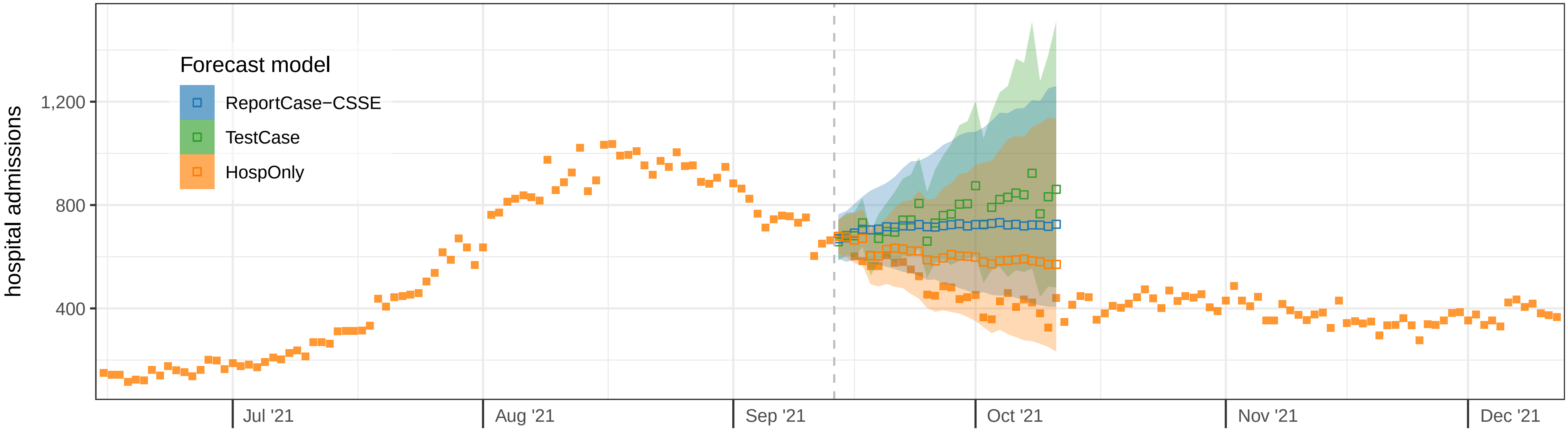

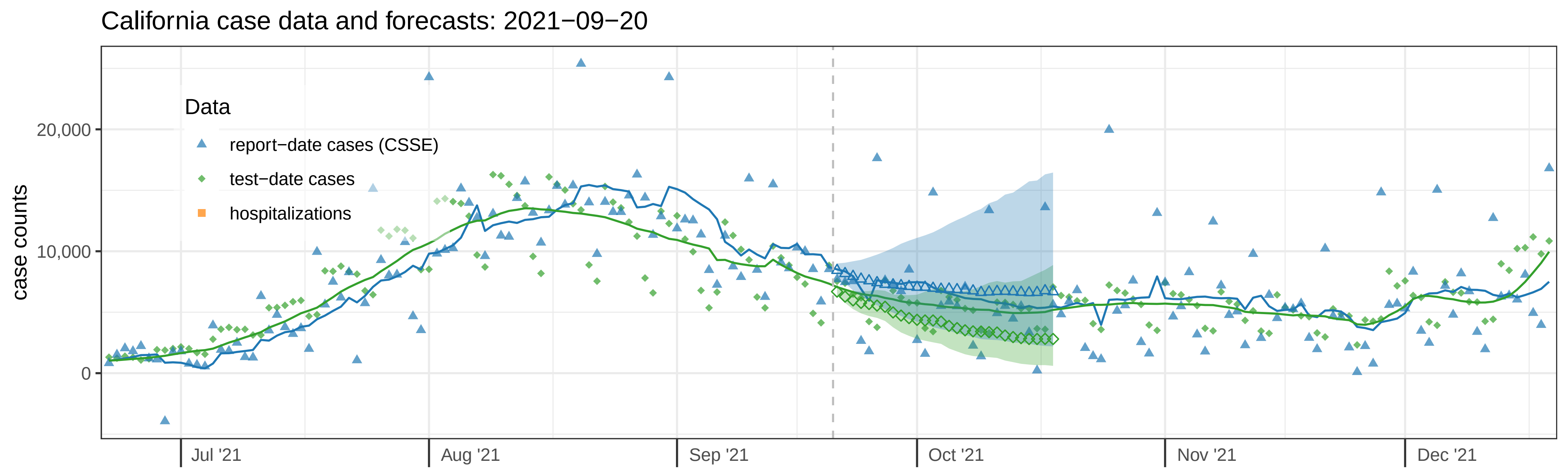
California case data and forecasts: 2021−09−20
case counts
20,000
10,000
0
Data
report−date cases (CSSE)
test−date cases
hospitalizations
Jul '21
Aug '21
Sep '21
Oct '21
Nov '21
Dec '21

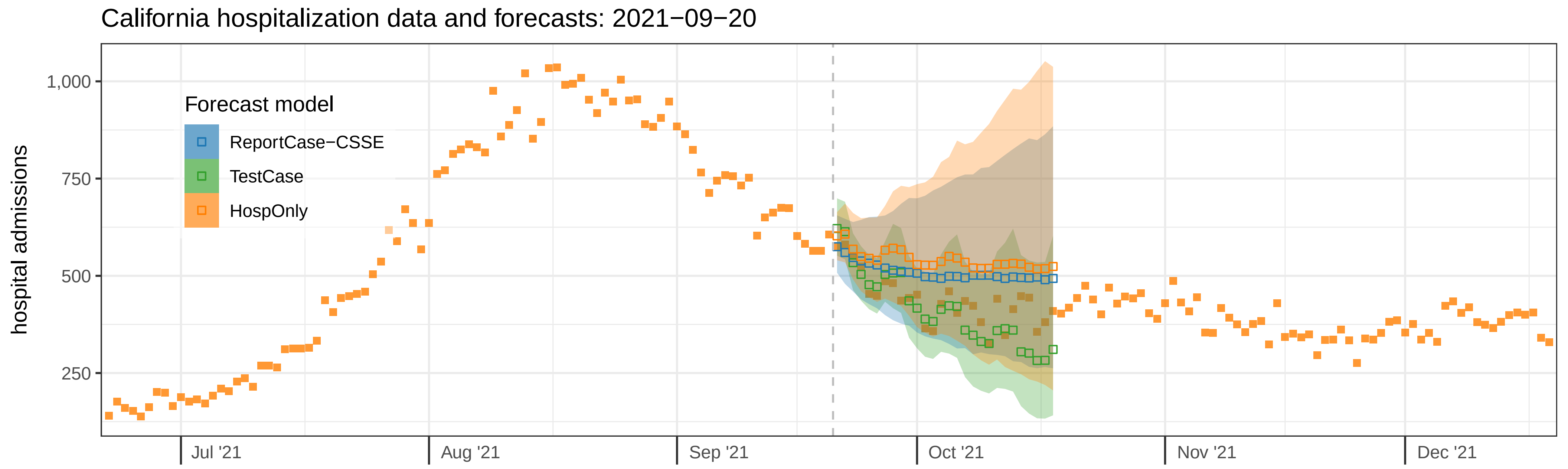
California hospitalization data and forecasts: 2021−09−20
hospital admissions
1,000
750
500
250
Forecast model
ReportCase−CSSE
TestCase
HospOnly
Jul '21
Aug '21
Sep '21
Oct '21
Nov '21
Dec '21

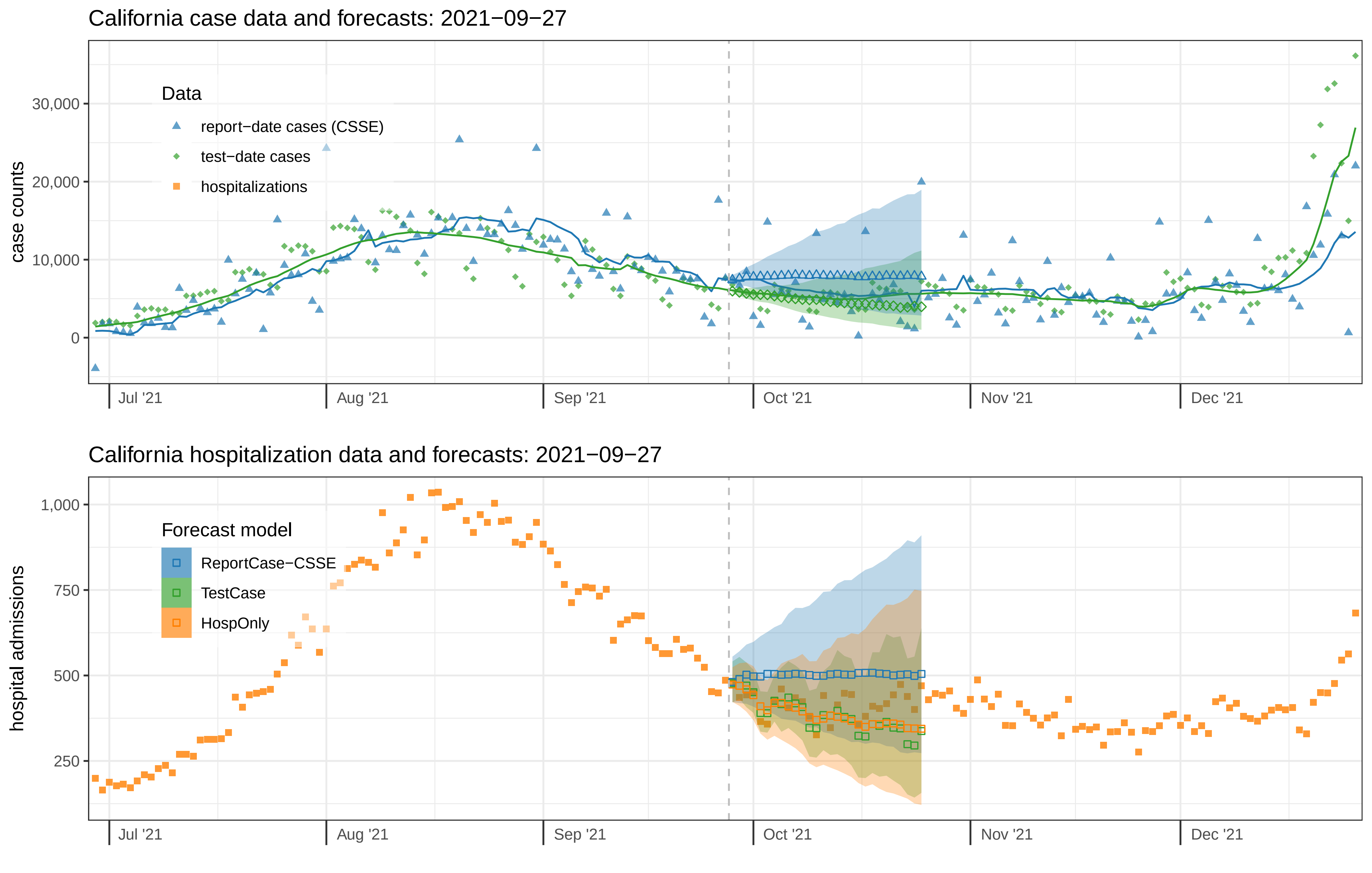
California case data and forecasts: 2021−09−27
case counts
30,000
20,000
10,000
0
Data
report−date cases (CSSE)
test−date cases
hospitalizations
Jul '21
Aug '21
Sep '21
Oct '21
Nov '21
Dec '21
California hospitalization data and forecasts: 2021−09−27
hospital admissions
1,000
750
500
250
Forecast model
ReportCase−CSSE
TestCase
HospOnly
Jul '21
Aug '21
Sep '21
Oct '21
Nov '21
Dec '21

### California case data and forecasts: 2021−10−04

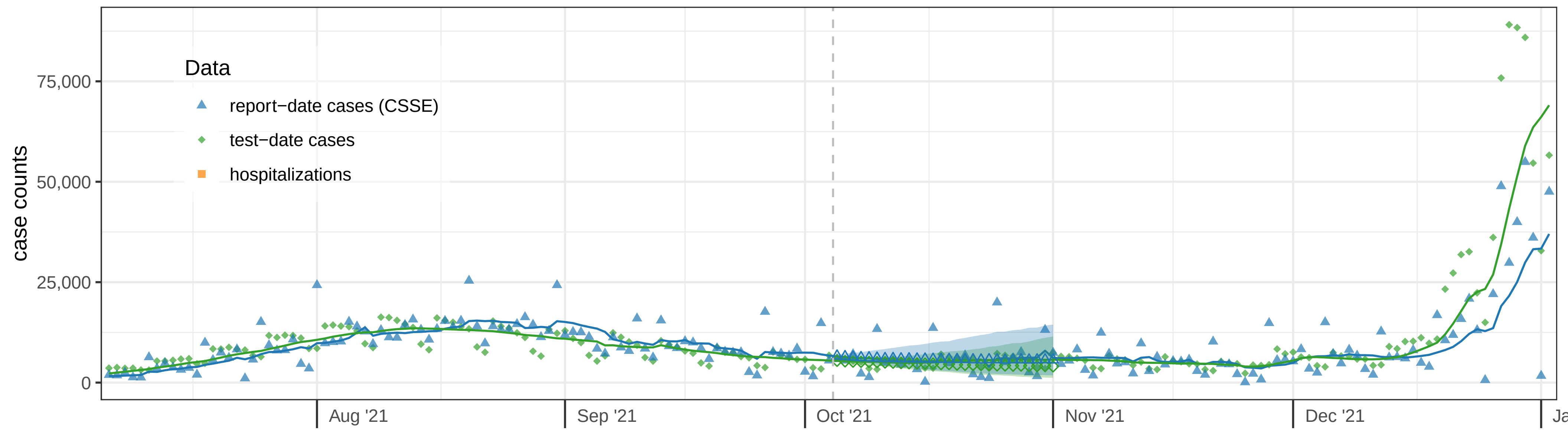


### California hospitalization data and forecasts: 2021−10−04

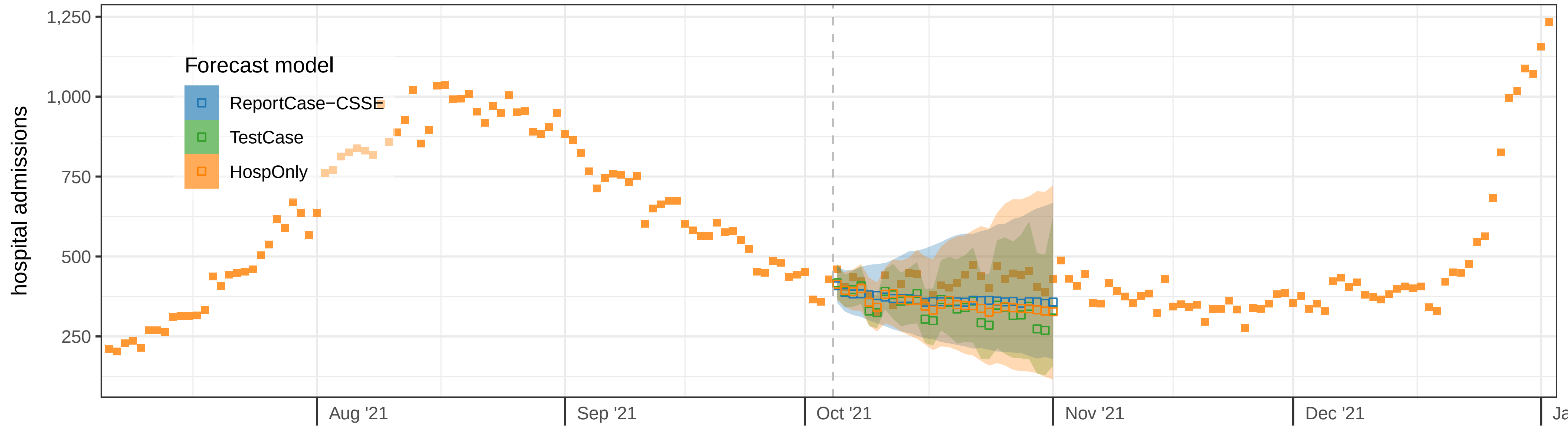

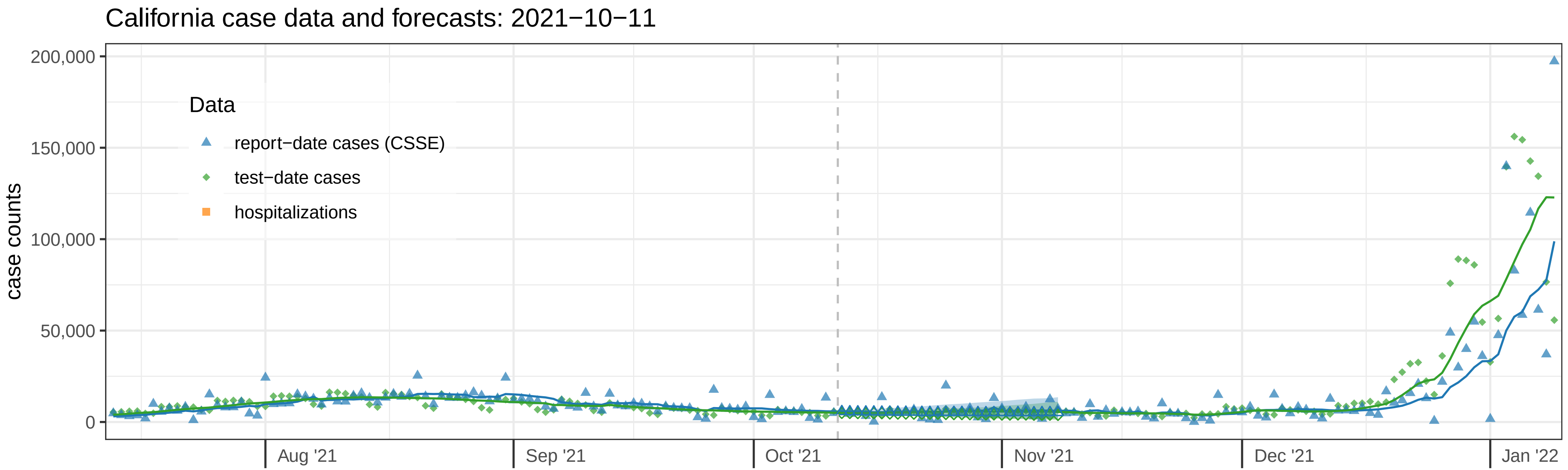

California case data and forecasts: 2021−10−11
case counts
200,000
150,000
100,000
50,000
0
Data
report−date cases (CSSE)
test−date cases
hospitalizations
Aug '21
Sep '21
Oct '21
Nov '21
Dec '21
Jan '22


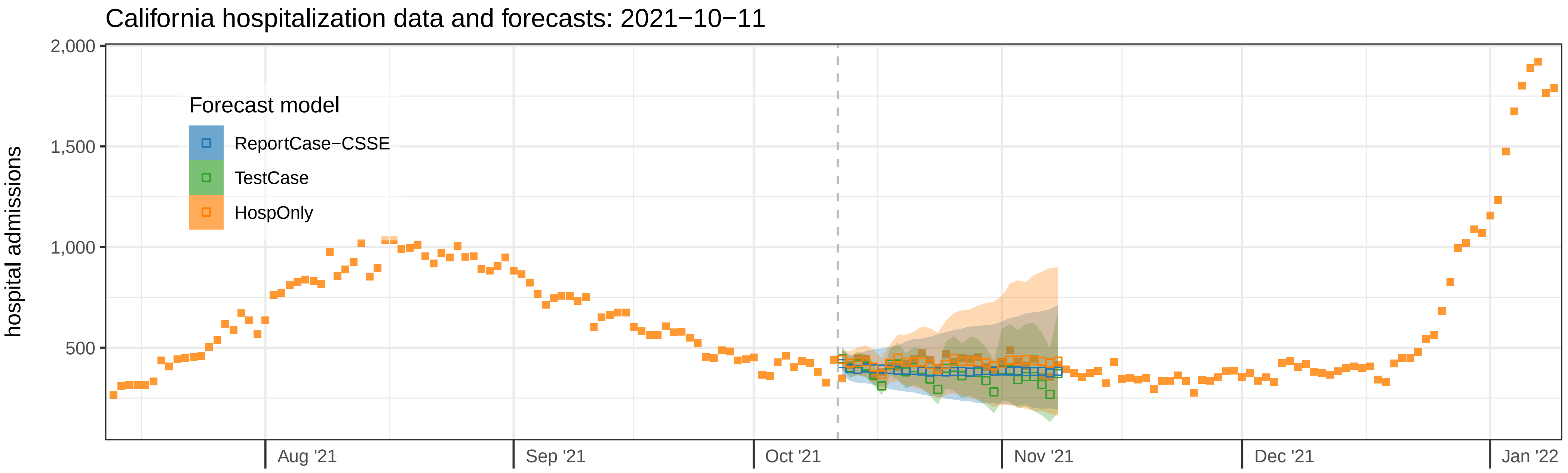

California hospitalization data and forecasts: 2021−10−11
hospital admissions
2,000
1,500
1,000
500
Forecast model
ReportCase−CSSE
TestCase
HospOnly
Aug '21
Sep '21
Oct '21
Nov '21
Dec '21
Jan '22

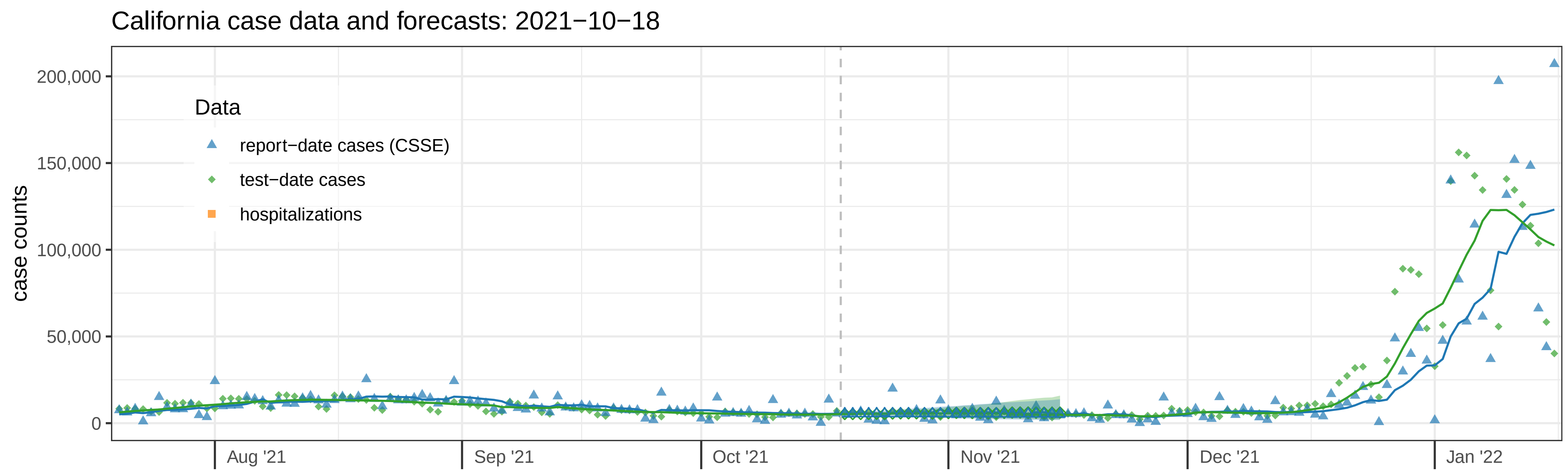
California case data and forecasts: 2021−10−18
case counts
200,000
150,000
100,000
50,000
0
Data
report−date cases (CSSE)
test−date cases
hospitalizations
Aug '21
Sep '21
Oct '21
Nov '21
Dec '21
Jan '22

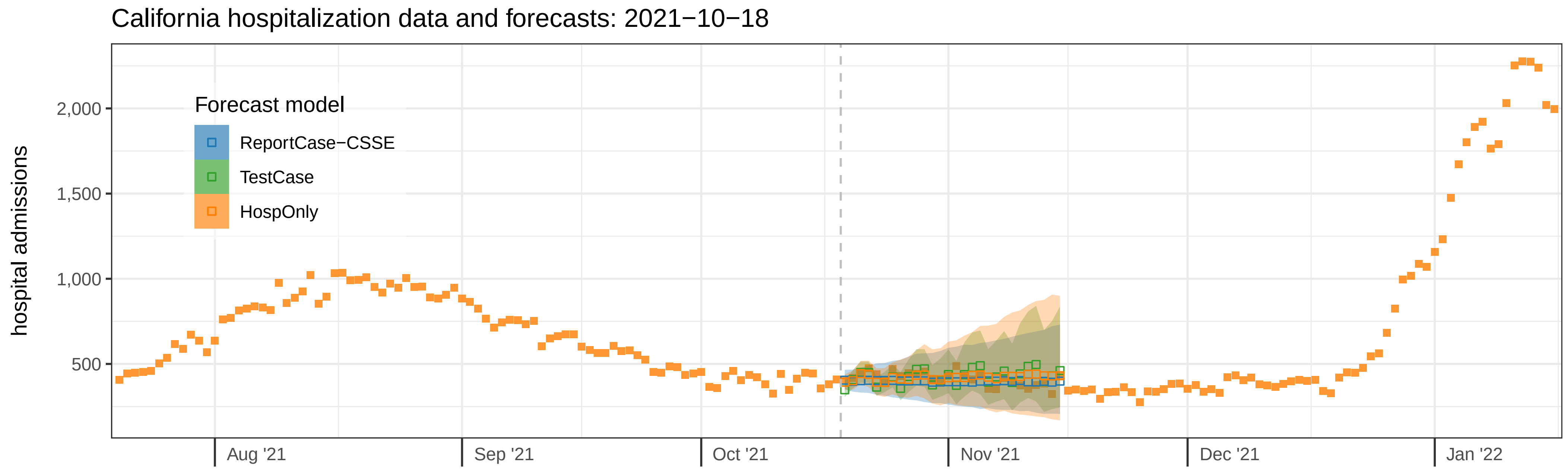
California hospitalization data and forecasts: 2021−10−18
hospital admissions
2,000
1,500
1,000
500
Forecast model
ReportCase−CSSE
TestCase
HospOnly
Aug '21
Sep '21
Oct '21
Nov '21
Dec '21
Jan '22

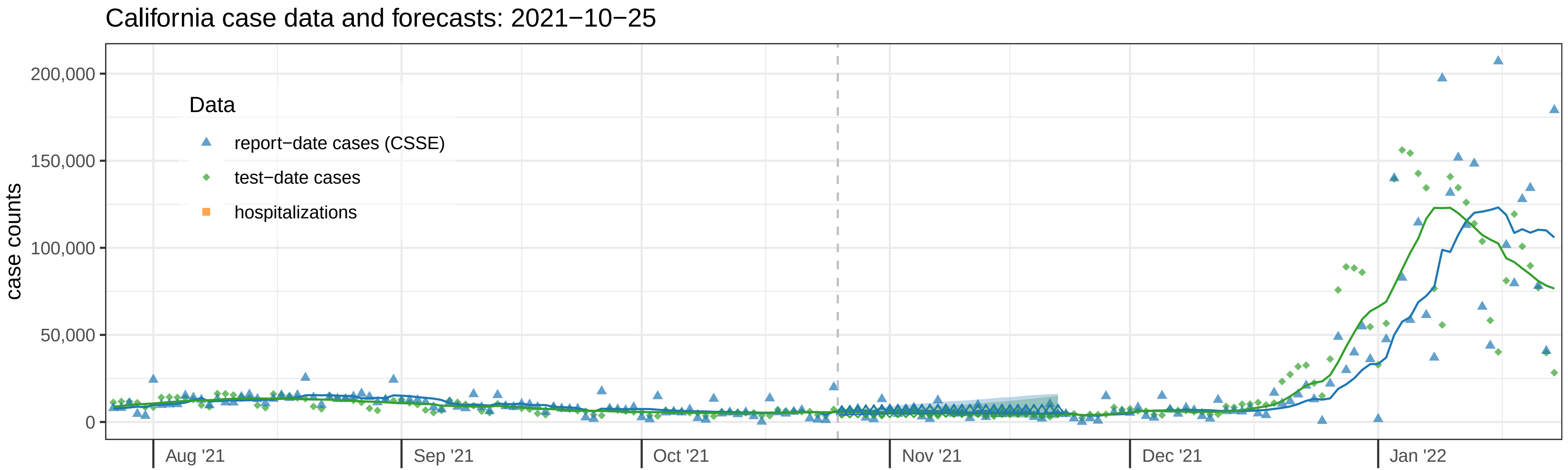
California case data and forecasts: 2021−10−25
case counts
200,000
150,000
100,000
50,000
0
Data
report−date cases (CSSE)
test−date cases
hospitalizations
Aug '21
Sep '21
Oct '21
Nov '21
Dec '21
Jan '22

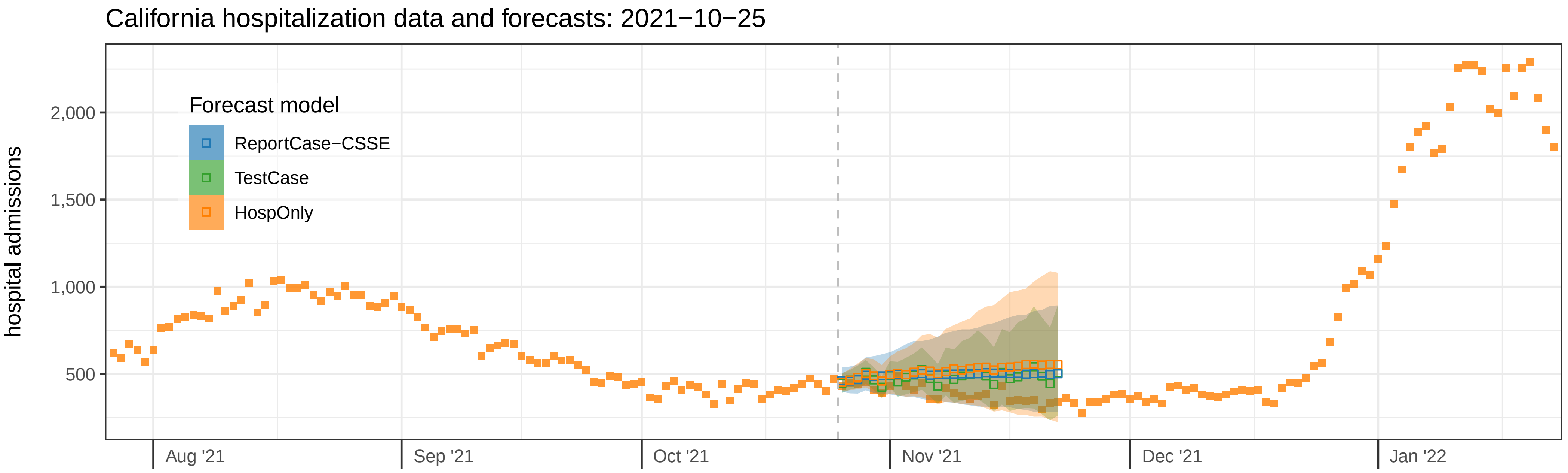
California hospitalization data and forecasts: 2021−10−25
hospital admissions
2,000
1,500
1,000
500
Forecast model
ReportCase−CSSE
TestCase
HospOnly
Aug '21
Sep '21
Oct '21
Nov '21
Dec '21
Jan '22

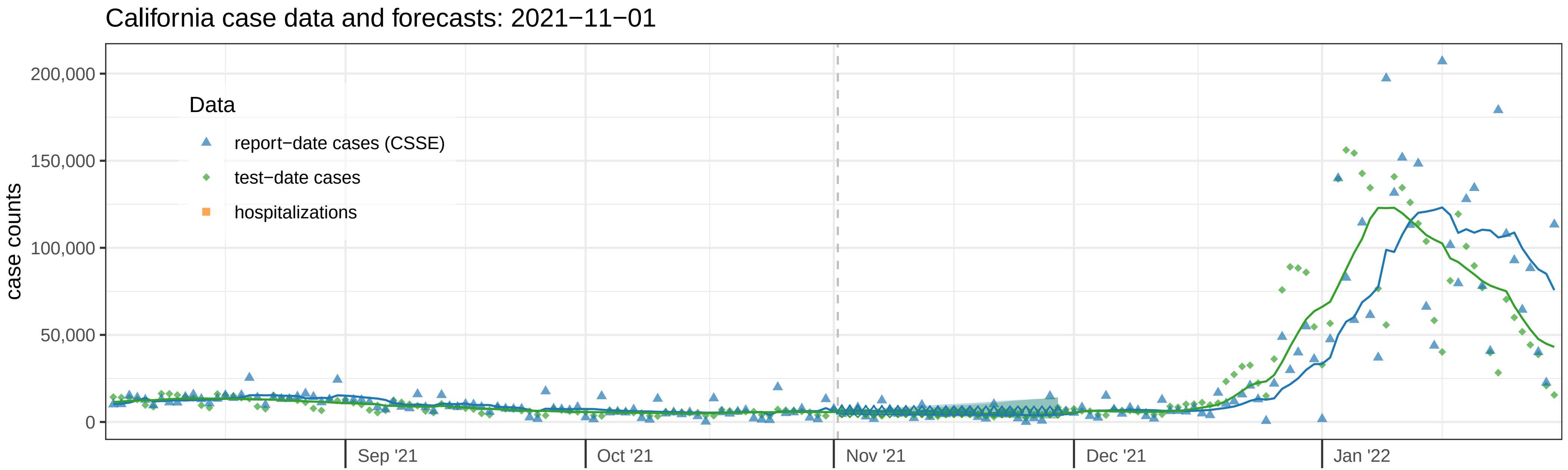
California case data and forecasts: 2021−11−01
case counts
200,000
150,000
100,000
50,000
0
Data
report−date cases (CSSE)
test−date cases
hospitalizations
Sep '21
Oct '21
Nov '21
Dec '21
Jan '22

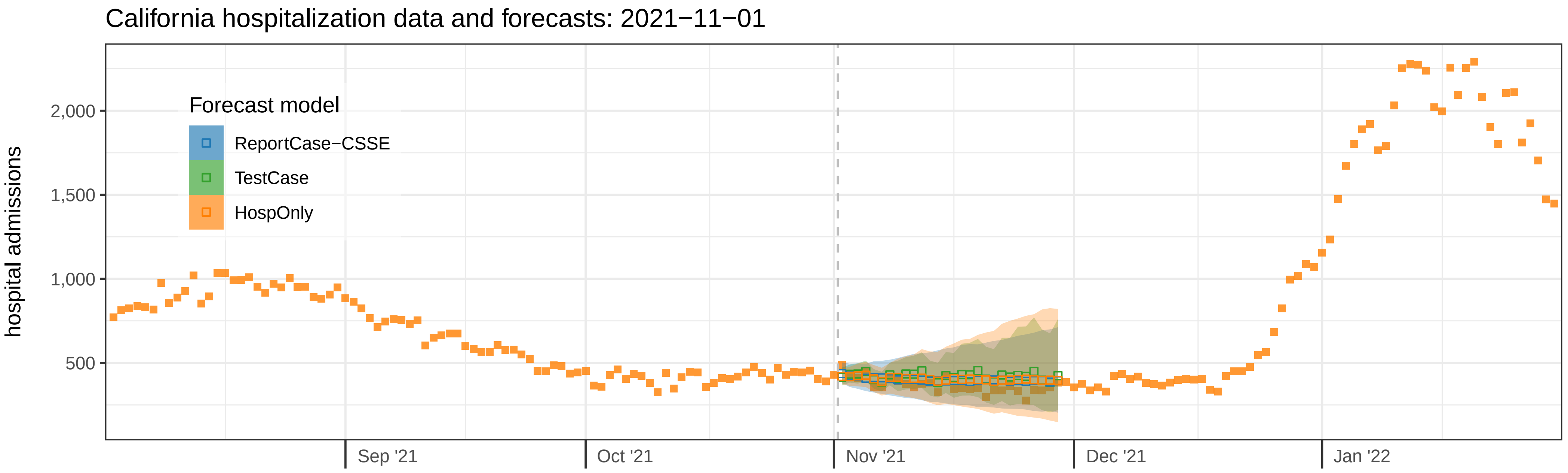
California hospitalization data and forecasts: 2021−11−01
hospital admissions
2,000
1,500
1,000
500
Forecast model
ReportCase−CSSE
TestCase
HospOnly
Sep '21
Oct '21
Nov '21
Dec '21
Jan '22

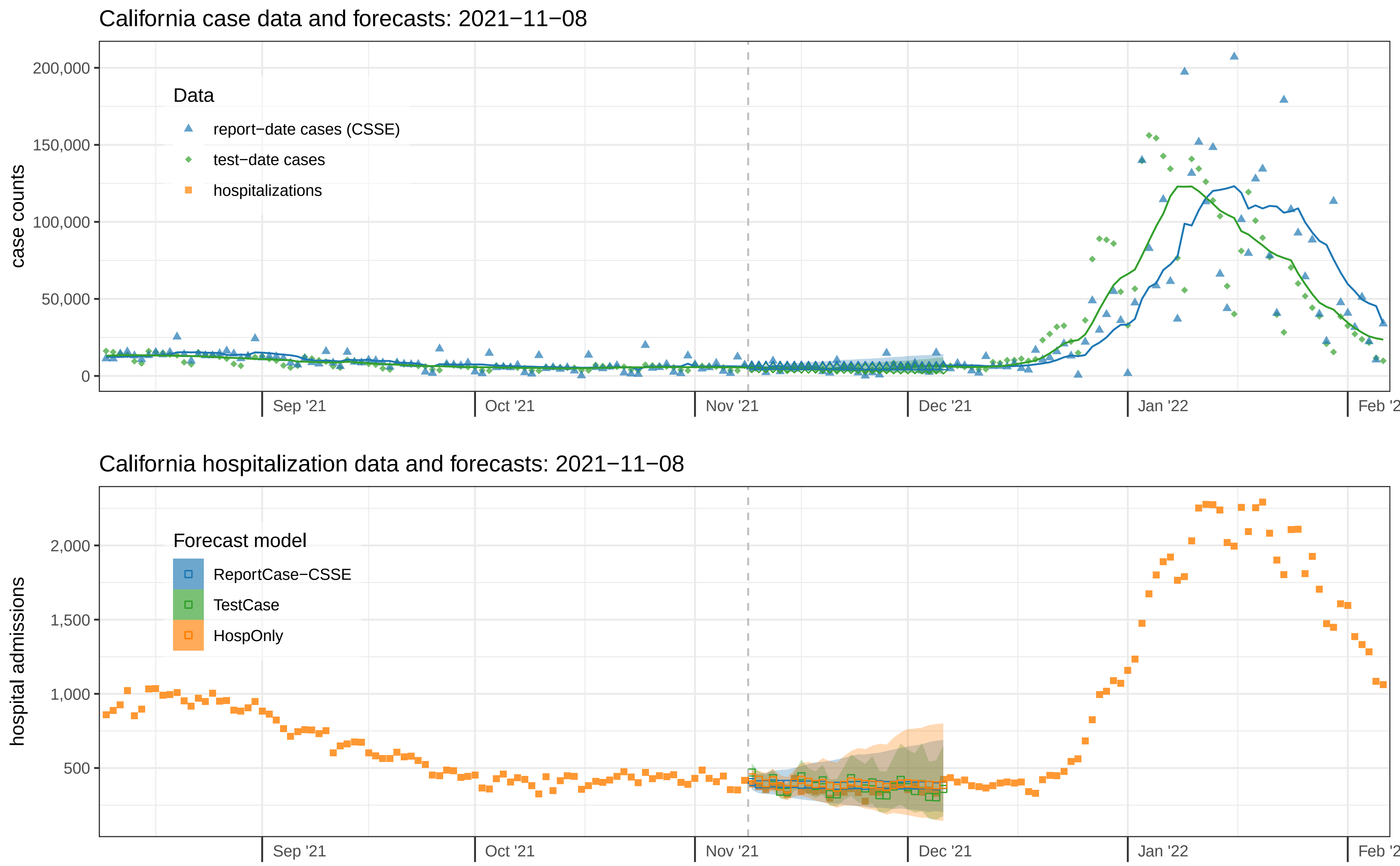
California case data and forecasts: 2021−11−08
case counts
200,000
150,000
100,000
50,000
0
Data
report−date cases (CSSE)
test−date cases
hospitalizations
Sep '21
Oct '21
Nov '21
Dec '21
Jan '22
California hospitalization data and forecasts: 2021−11−08
hospital admissions
2,000
1,500
1,000
500
Forecast model
ReportCase−CSSE
TestCase
HospOnly
Sep '21
Oct '21
Nov '21
Dec '21
Jan '22

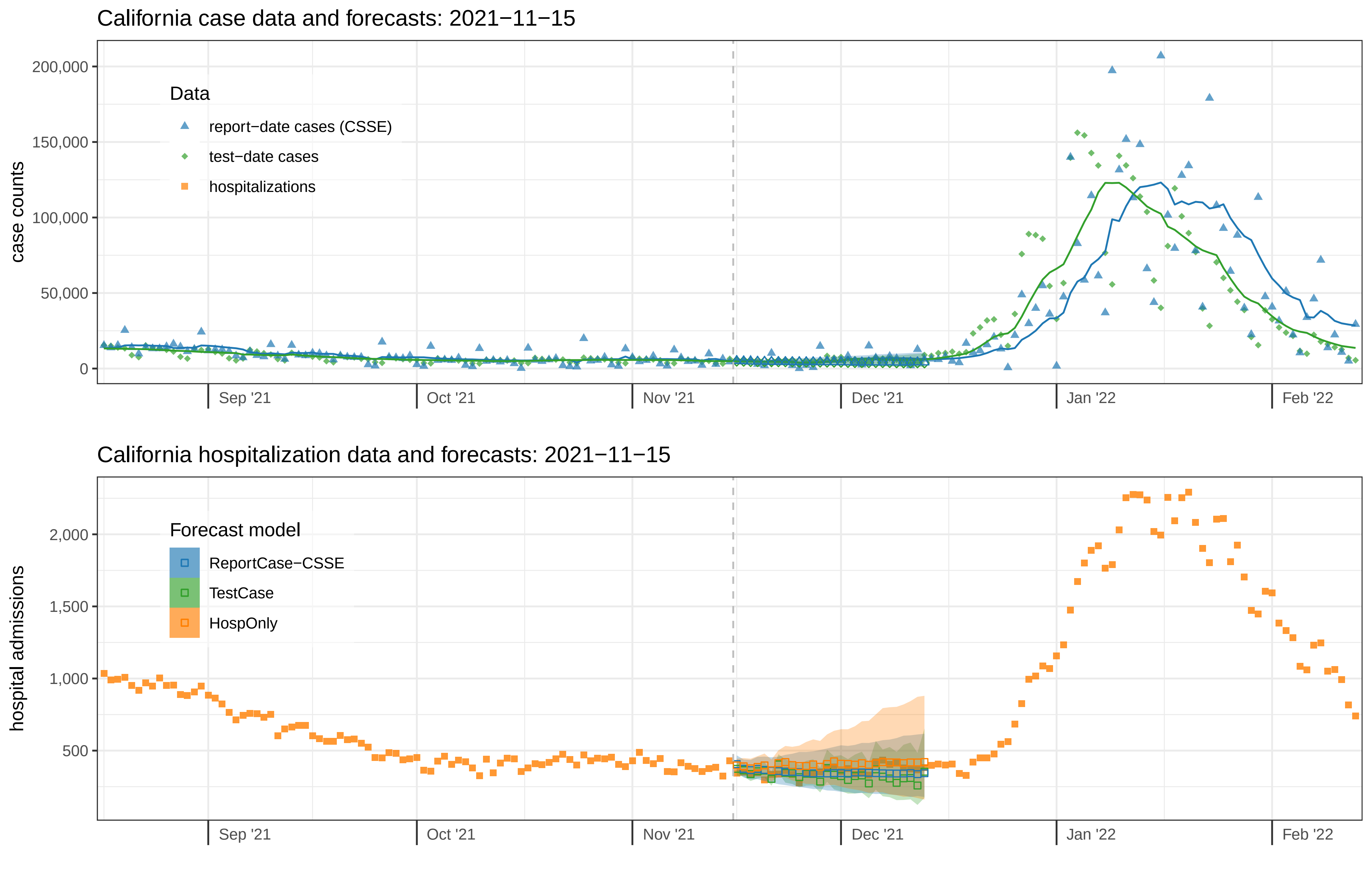

California case data and forecasts: 2021−11−15
case counts
200,000
150,000
100,000
50,000
0
Data
report−date cases (CSSE)
test−date cases
hospitalizations
Sep '21
Oct '21
Nov '21
Dec '21
Jan '22
Feb '22
California hospitalization data and forecasts: 2021−11−15
hospital admissions
2,000
1,500
1,000
500
Forecast model
ReportCase−CSSE
TestCase
HospOnly

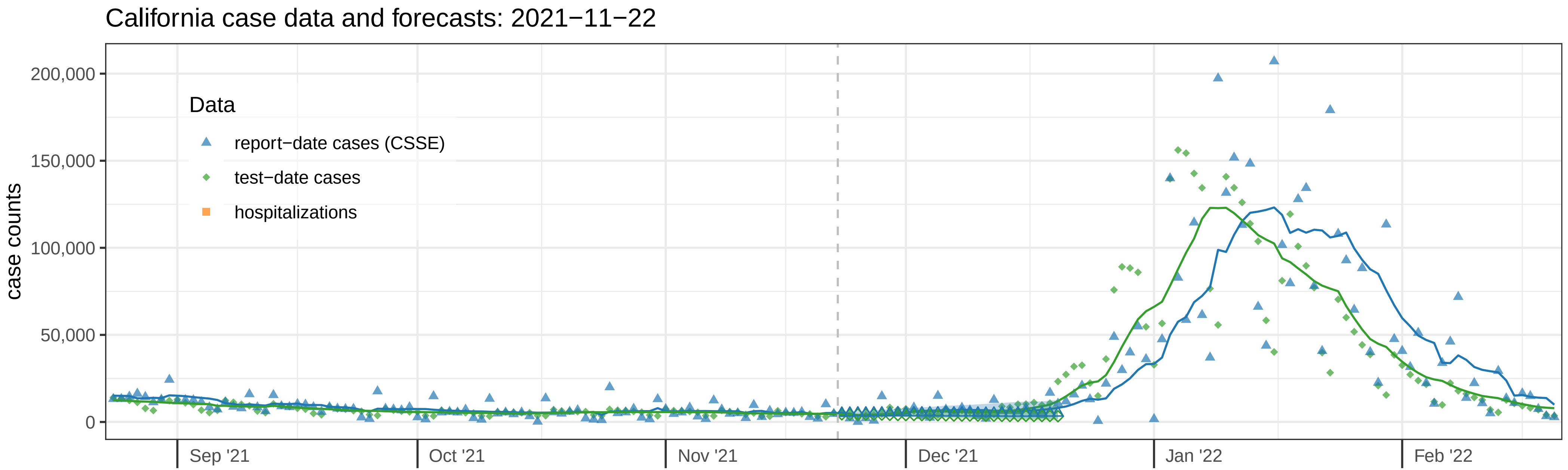
California case data and forecasts: 2021−11−22
case counts
200,000
150,000
100,000
50,000
0
Data
report−date cases (CSSE)
test−date cases
hospitalizations
Sep '21
Oct '21
Nov '21
Dec '21
Jan '22
Feb '22

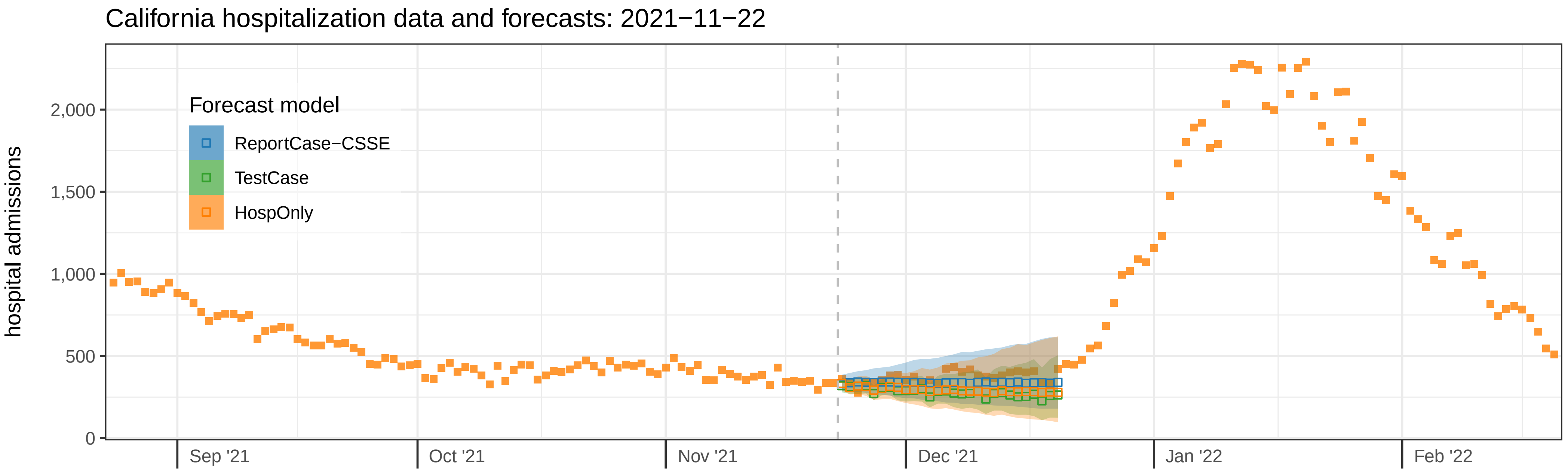
California hospitalization data and forecasts: 2021−11−22
hospital admissions
2,000
1,500
1,000
500
0
Forecast model
ReportCase−CSSE
TestCase
HospOnly
Sep '21
Oct '21
Nov '21
Dec '21
Jan '22
Feb '22

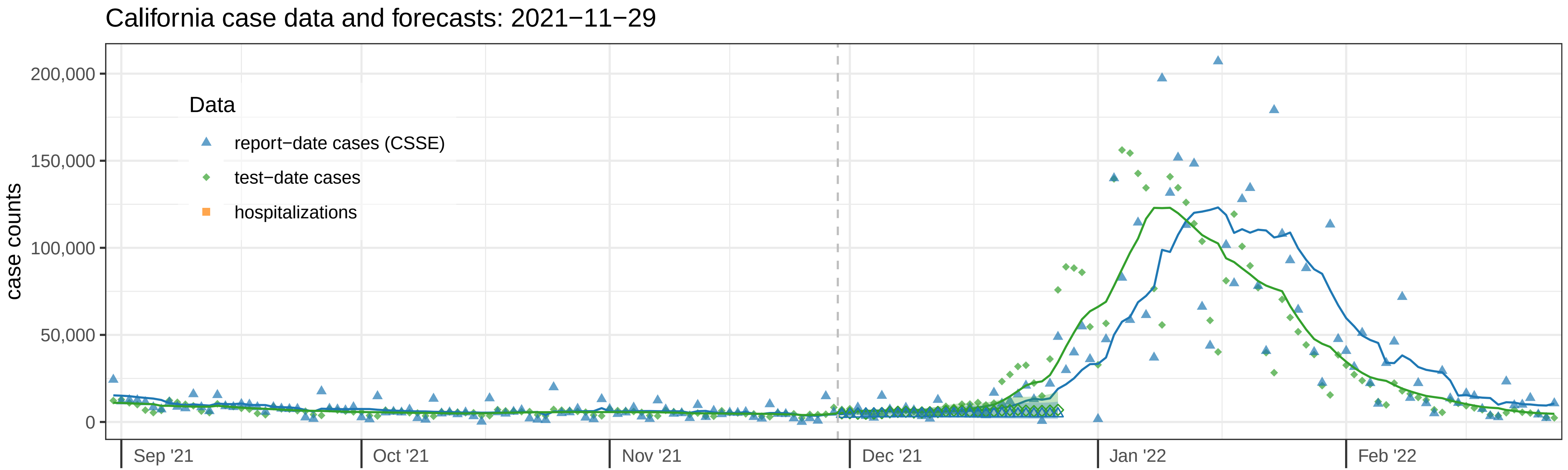
California case data and forecasts: 2021−11−29
case counts
200,000
150,000
100,000
50,000
0
Data
report−date cases (CSSE)
test−date cases
hospitalizations
Sep '21
Oct '21
Nov '21
Dec '21
Jan '22
Feb '22

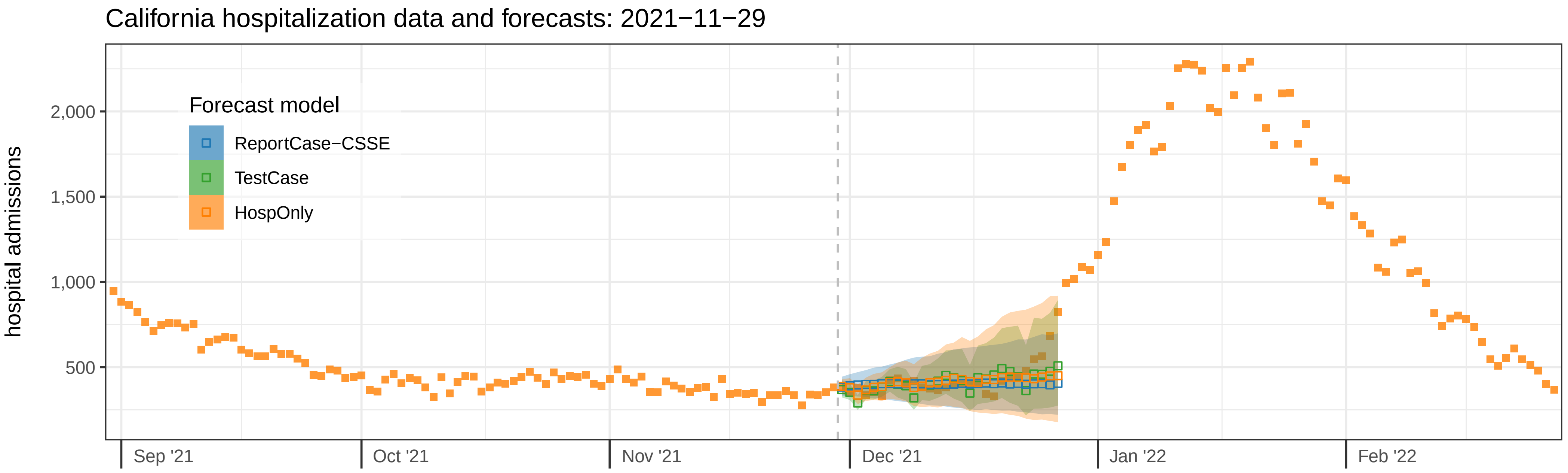
California hospitalization data and forecasts: 2021−11−29
hospital admissions
2,000
1,500
1,000
500
Forecast model
ReportCase−CSSE
TestCase
HospOnly
Sep '21
Oct '21
Nov '21
Dec '21
Jan '22
Feb '22

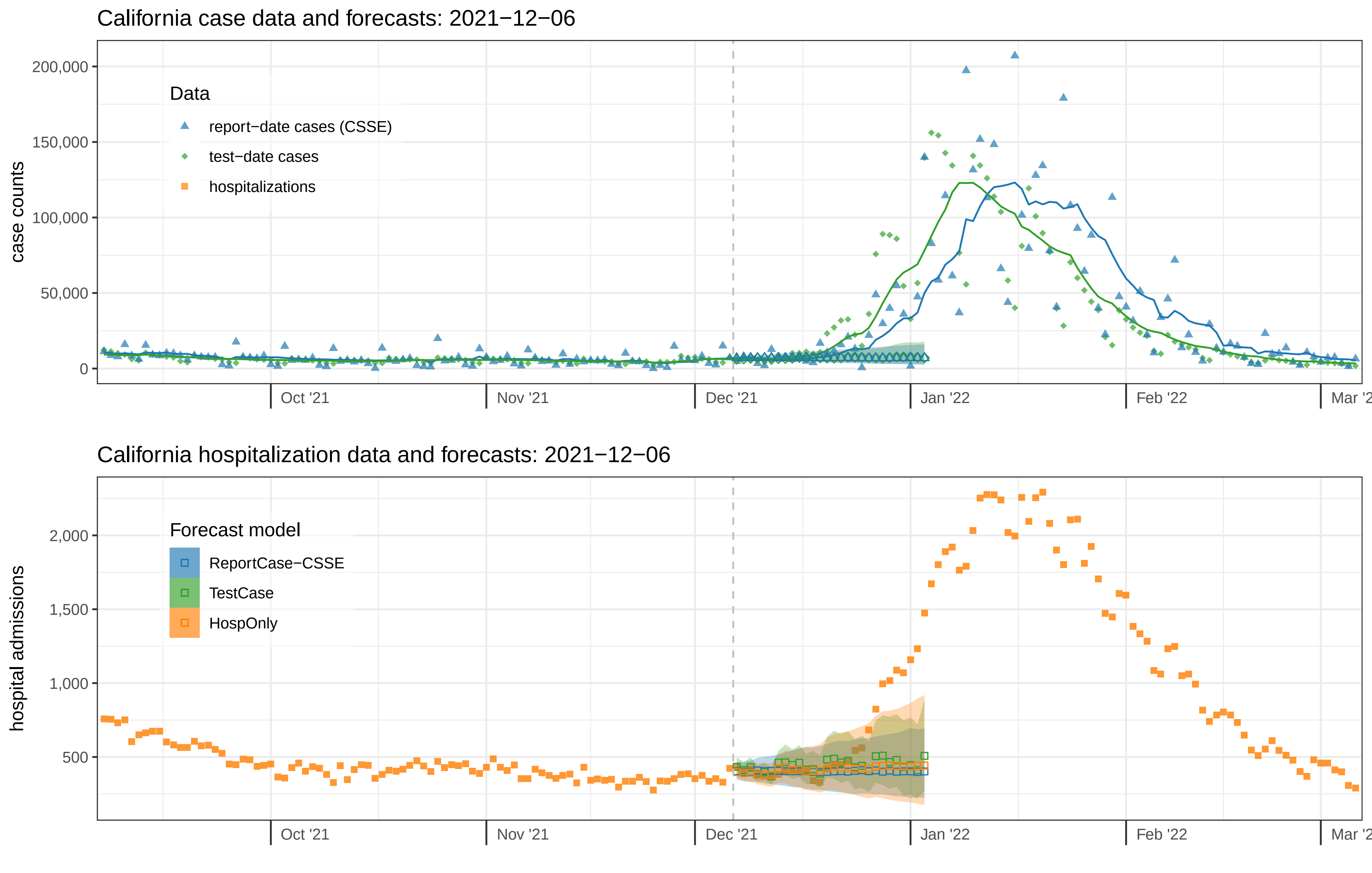
California case data and forecasts: 2021−12−06
case counts
200,000
150,000
100,000
50,000
0
Data
report−date cases (CSSE)
test−date cases
hospitalizations
Oct '21
Nov '21
Dec '21
Jan '22
Feb '22
California hospitalization data and forecasts: 2021−12−06
hospital admissions
2,000
1,500
1,000
500
Forecast model
ReportCase−CSSE
TestCase
HospOnly
Oct '21
Nov '21
Dec '21
Jan '22
Feb '22

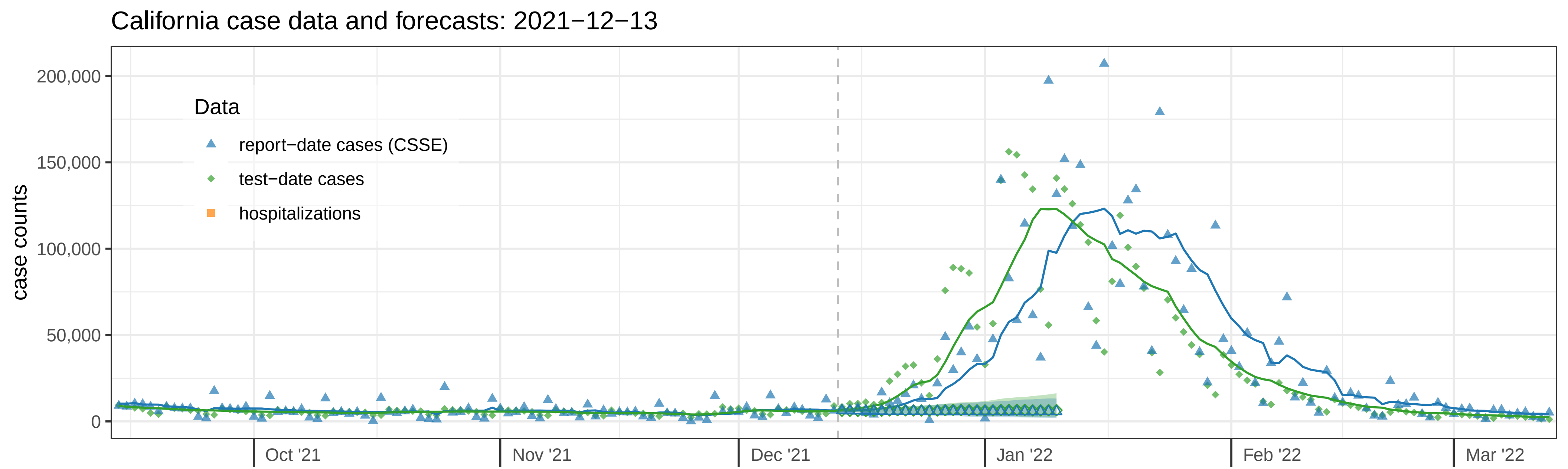

California case data and forecasts: 2021−12−13
case counts
200,000
150,000
100,000
50,000
0
Data
report−date cases (CSSE)
test−date cases
hospitalizations
Oct '21
Nov '21
Dec '21
Jan '22
Feb '22
Mar '22


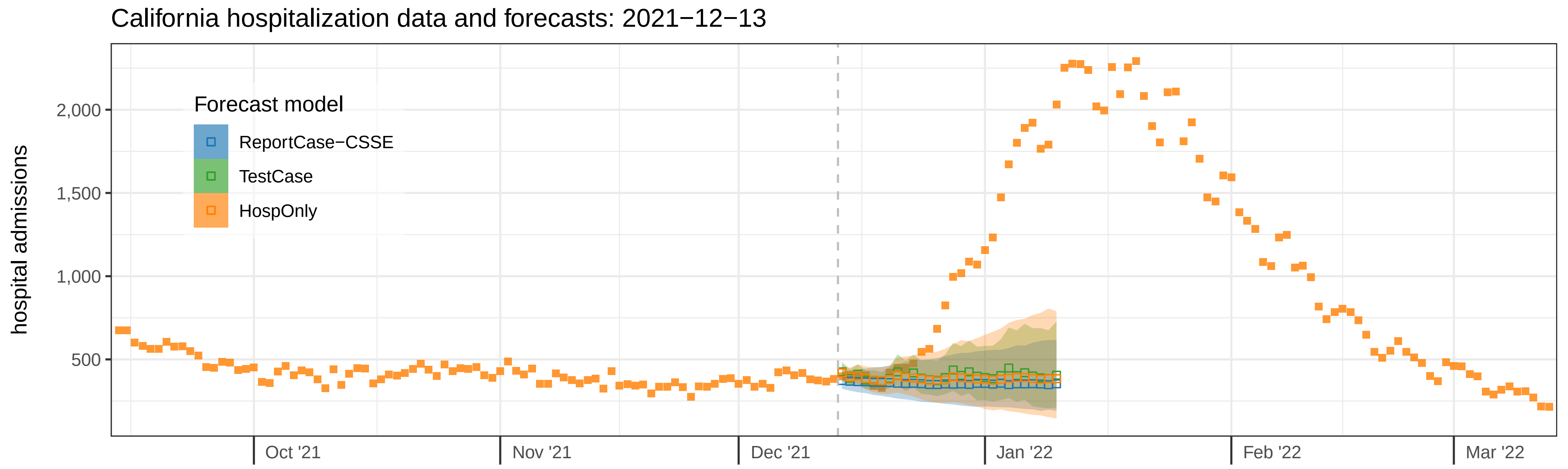

California hospitalization data and forecasts: 2021−12−13
hospital admissions
2,000
1,500
1,000
500
Forecast model
ReportCase−CSSE
TestCase
HospOnly
Oct '21
Nov '21
Dec '21
Jan '22
Feb '22
Mar '22

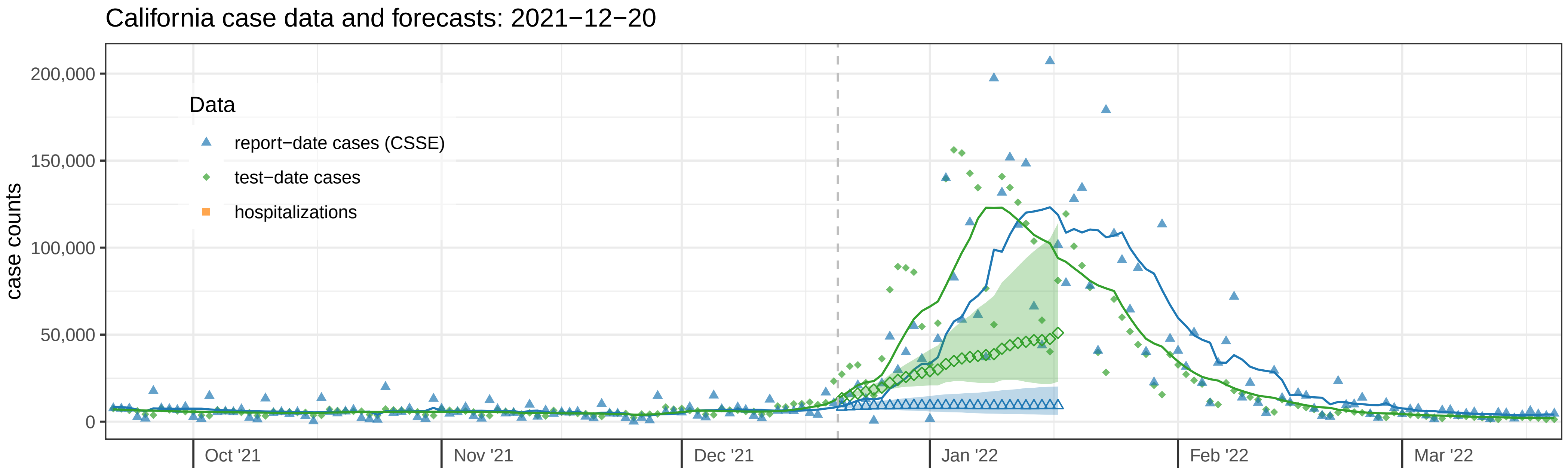
California case data and forecasts: 2021−12−20
case counts
200,000
150,000
100,000
50,000
0
Data
report−date cases (CSSE)
test−date cases
hospitalizations
Oct '21
Nov '21
Dec '21
Jan '22
Feb '22
Mar '22

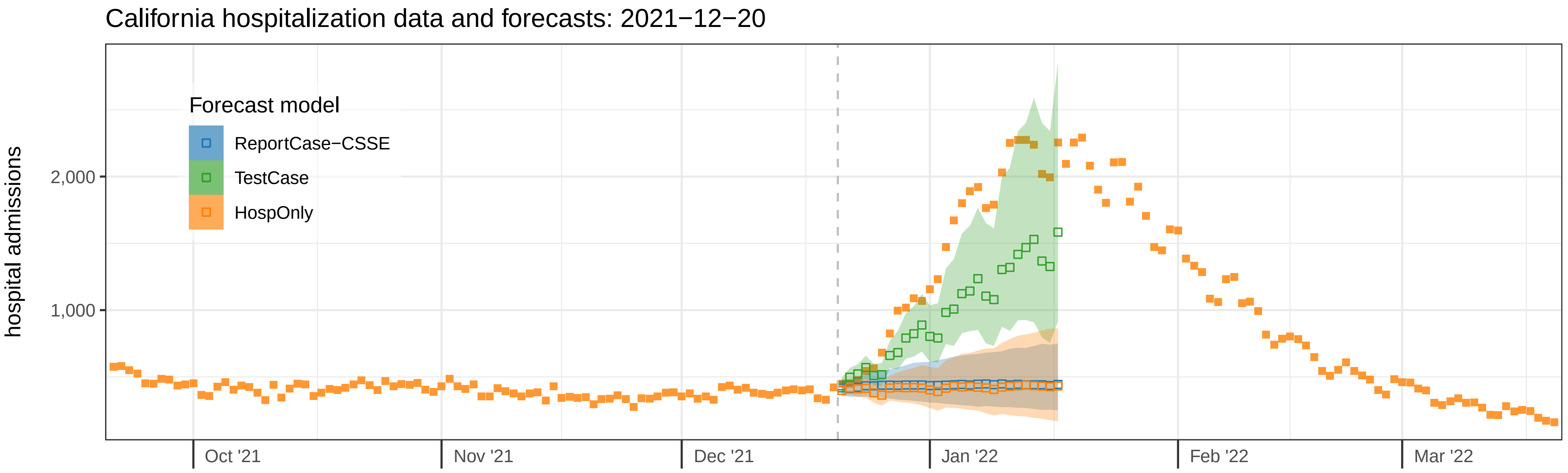
California hospitalization data and forecasts: 2021−12−20
hospital admissions
2,000
1,000
Forecast model
ReportCase−CSSE
TestCase
HospOnly
Oct '21
Nov '21
Dec '21
Jan '22
Feb '22
Mar '22

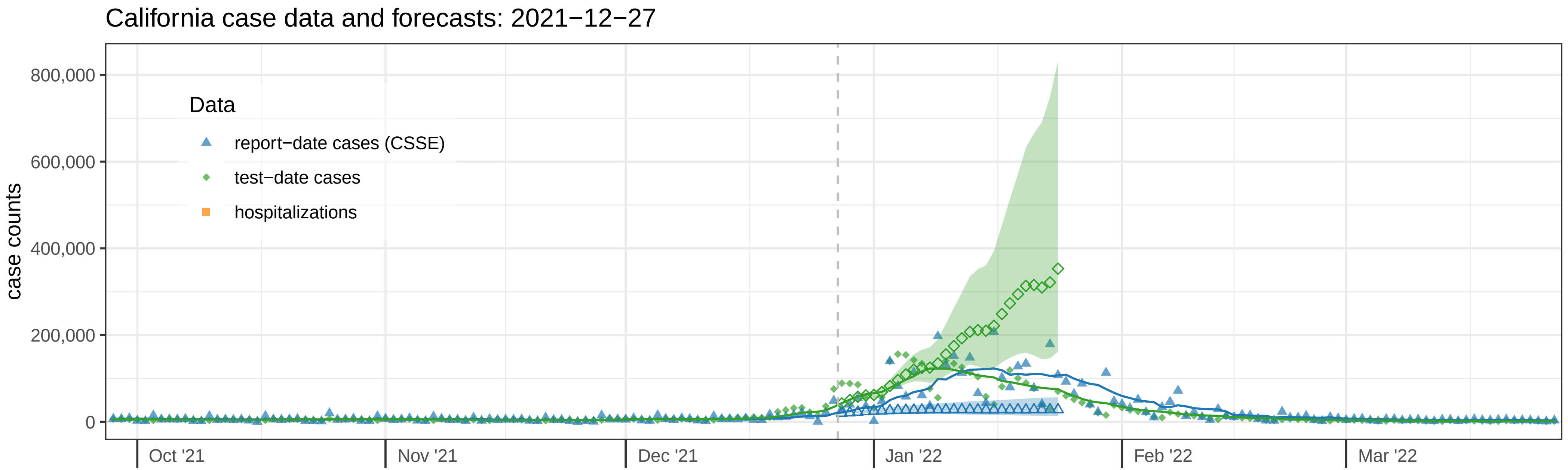

California case data and forecasts: 2021−12−27
case counts
800,000
600,000
400,000
200,000
0
Data
report−date cases (CSSE)
test−date cases
hospitalizations
Oct '21
Nov '21
Dec '21
Jan '22
Feb '22
Mar '22


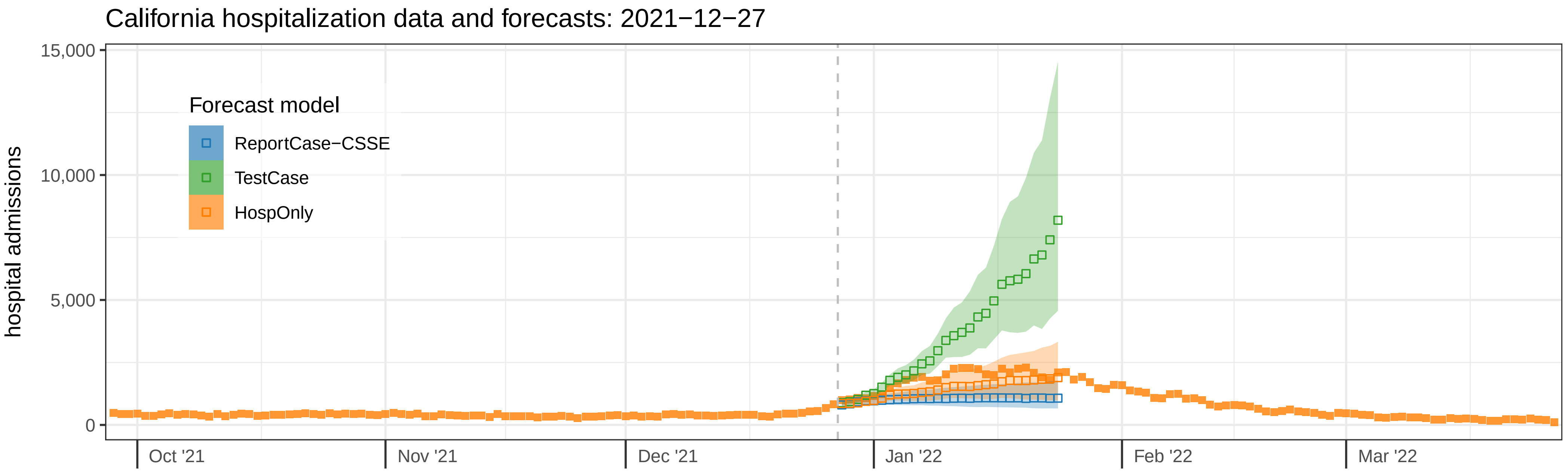

California hospitalization data and forecasts: 2021−12−27
hospital admissions
15,000
10,000
5,000
0
Forecast model
ReportCase−CSSE
TestCase
HospOnly
Oct '21
Nov '21
Dec '21
Jan '22
Feb '22
Mar '22

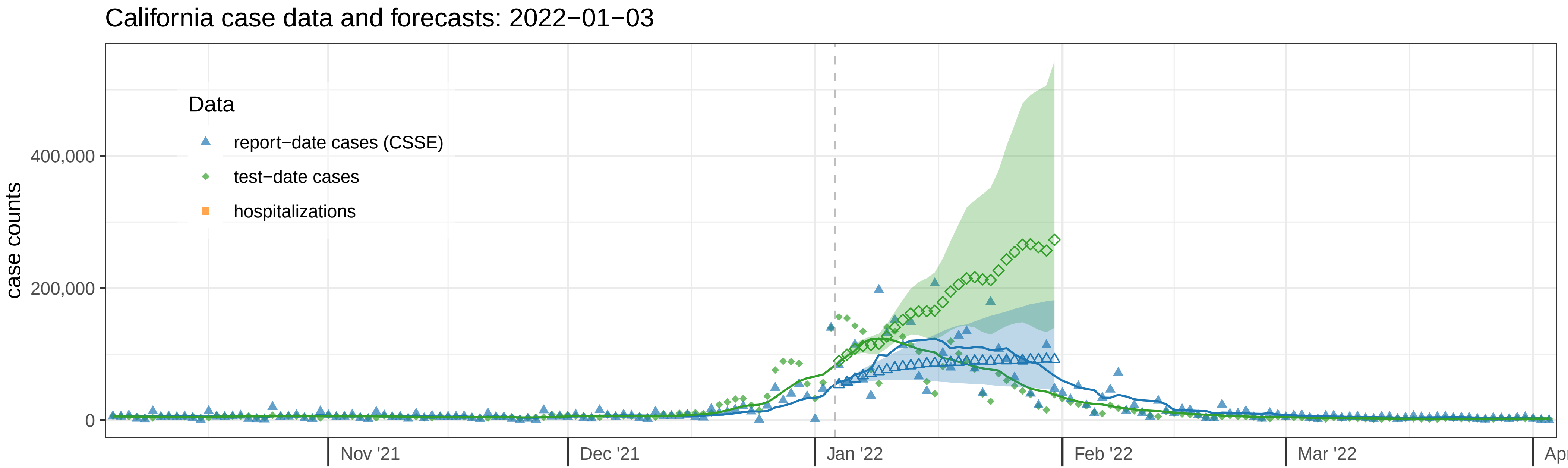
California case data and forecasts: 2022−01−03
Data
report−date cases (CSSE)
test−date cases
hospitalizations
case counts
400,000
200,000
0
Nov '21
Dec '21
Jan '22
Feb '22
Mar '22

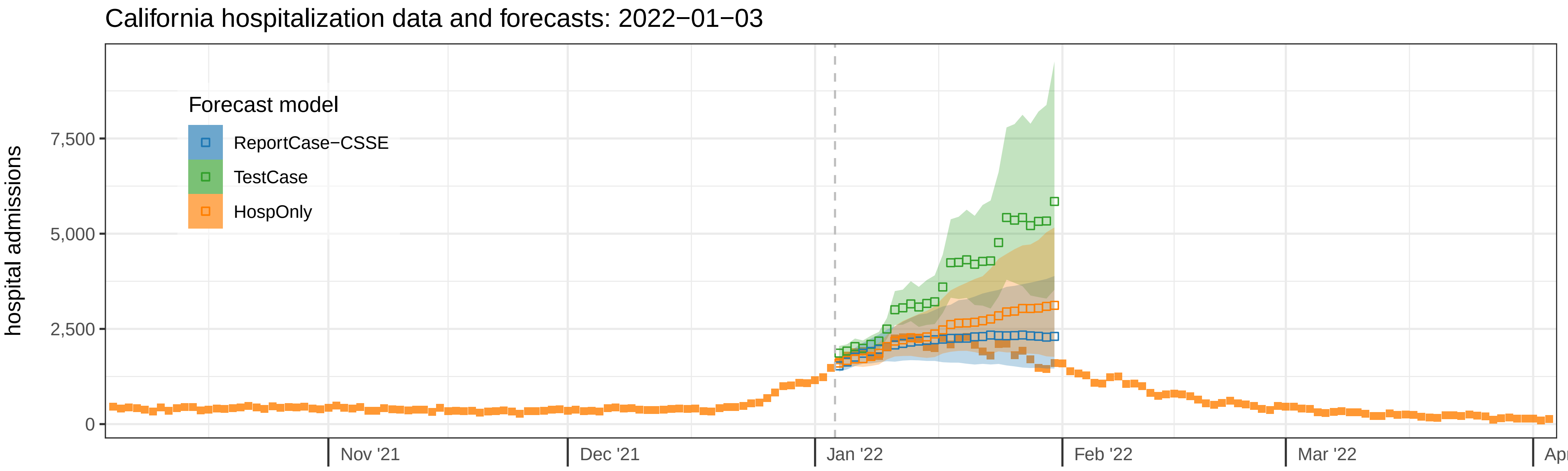
California hospitalization data and forecasts: 2022−01−03
Forecast model
ReportCase−CSSE
TestCase
HospOnly
hospital admissions
7,500
5,000
2,500
0
Nov '21
Dec '21
Jan '22
Feb '22
Mar '22

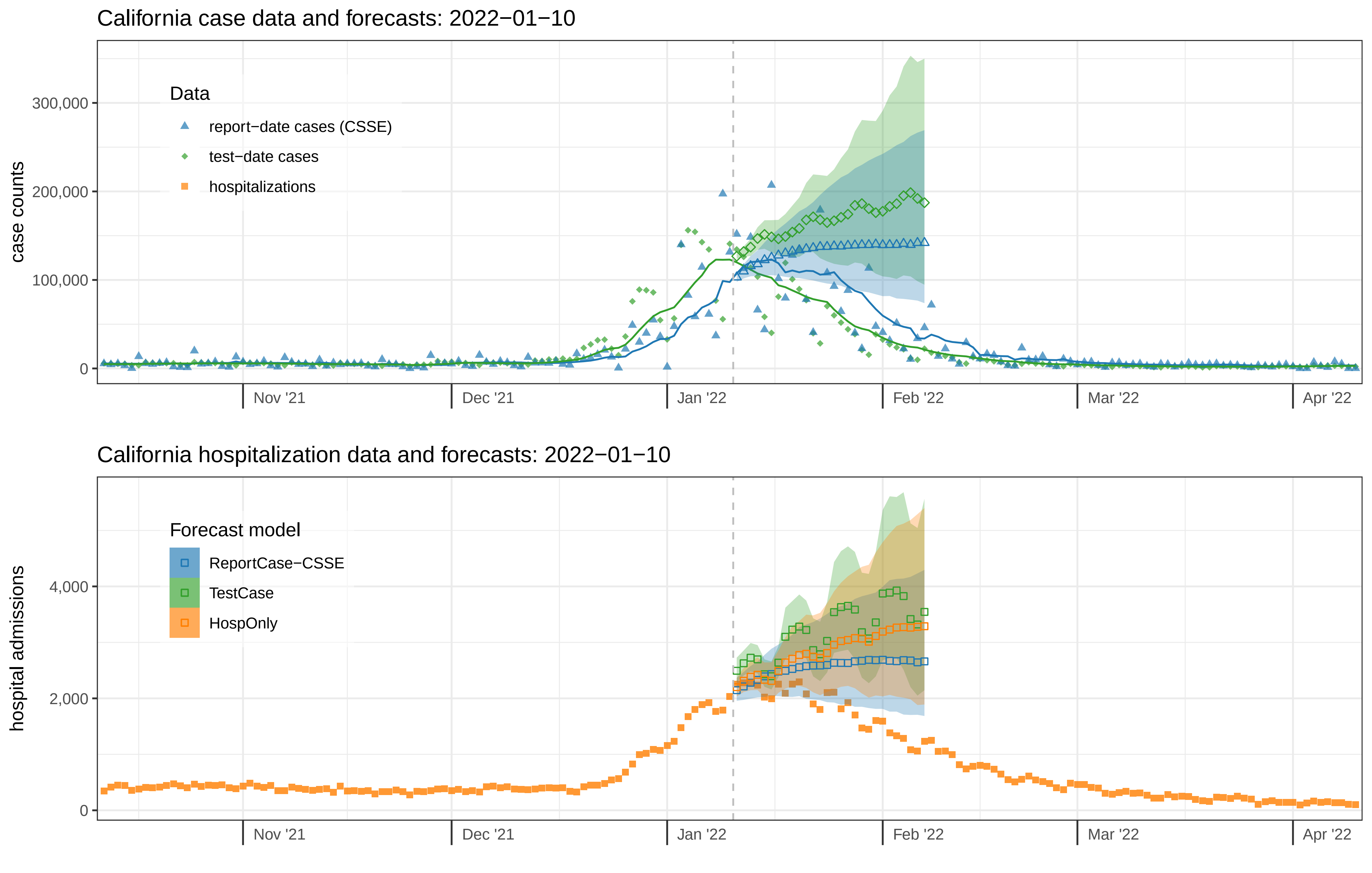

California case data and forecasts: 2022−01−10
case counts
300,000
200,000
100,000
0
Data
report−date cases (CSSE)
test−date cases
hospitalizations
Nov '21
Dec '21
Jan '22
Feb '22
Mar '22
Apr '22
California hospitalization data and forecasts: 2022−01−10
hospital admissions
4,000
2,000
0
Forecast model
ReportCase−CSSE
TestCase
HospOnly
Nov '21
Dec '21
Jan '22
Feb '22
Mar '22
Apr '22

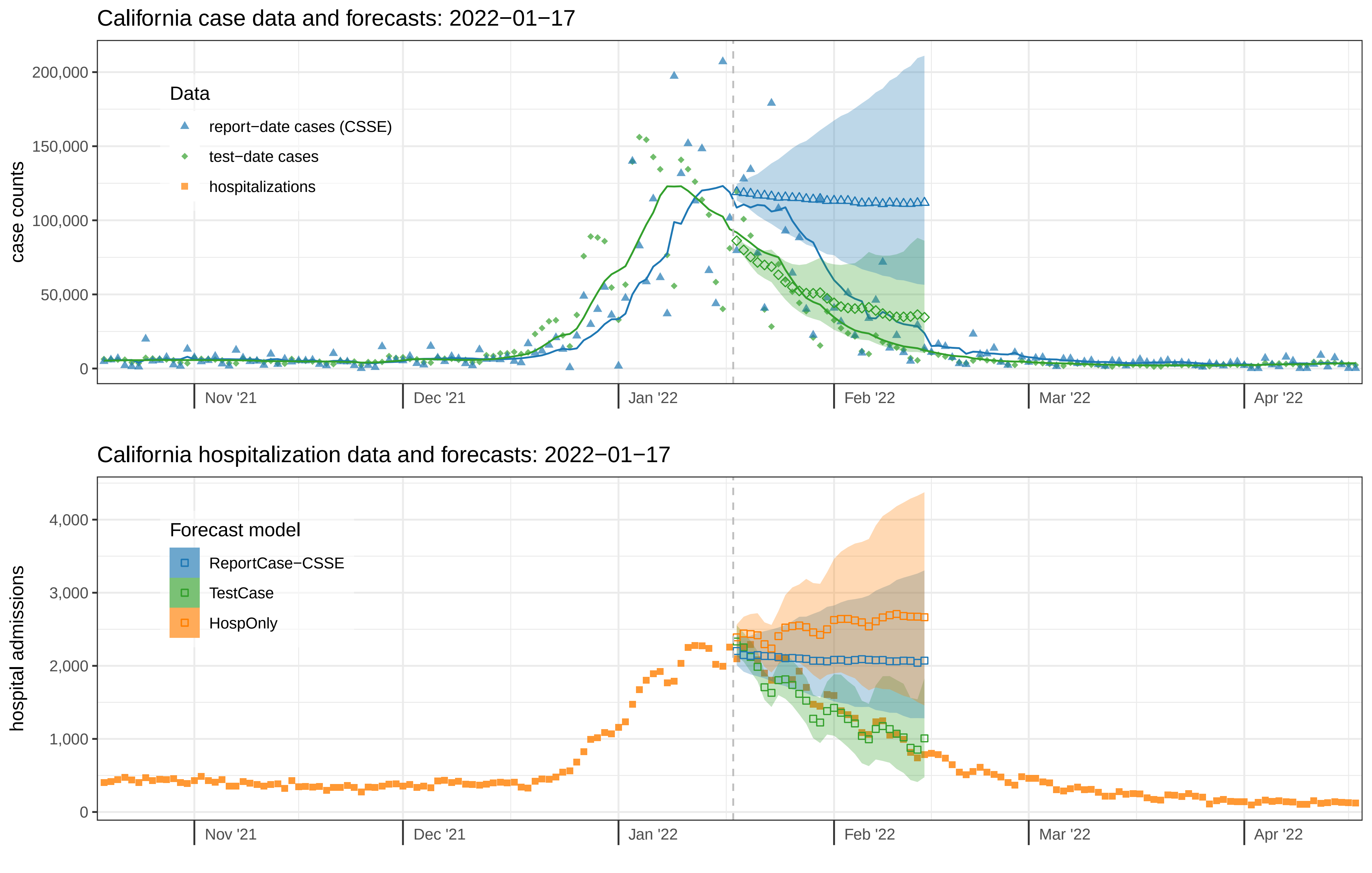

California case data and forecasts: 2022−01−17
case counts
200,000
150,000
100,000
50,000
0
Data
report−date cases (CSSE)
test−date cases
hospitalizations
Nov '21
Dec '21
Jan '22
Feb '22
Mar '22
Apr '22
California hospitalization data and forecasts: 2022−01−17
hospital admissions
4,000
3,000
2,000
1,000
0
Forecast model
ReportCase−CSSE
TestCase
HospOnly
Nov '21
Dec '21
Jan '22
Feb '22
Mar '22
Apr '22

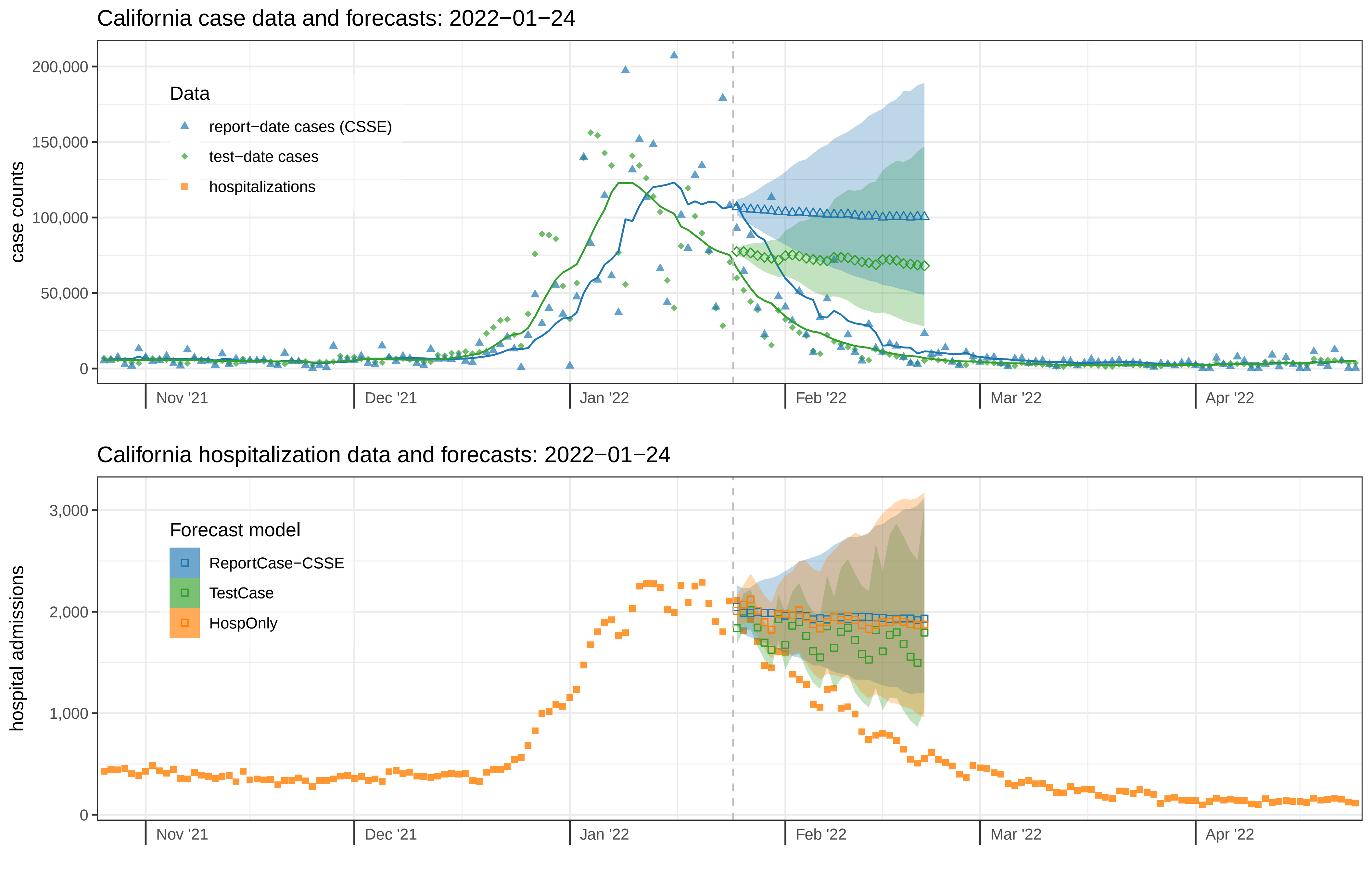
California case data and forecasts: 2022−01−24
case counts
200,000
150,000
100,000
50,000
0
Data
report−date cases (CSSE)
test−date cases
hospitalizations
Nov '21
Dec '21
Jan '22
Feb '22
Mar '22
Apr '22
California hospitalization data and forecasts: 2022−01−24
hospital admissions
3,000
2,000
1,000
0
Forecast model
ReportCase−CSSE
TestCase
HospOnly
Nov '21
Dec '21
Jan '22
Feb '22
Mar '22
Apr '22

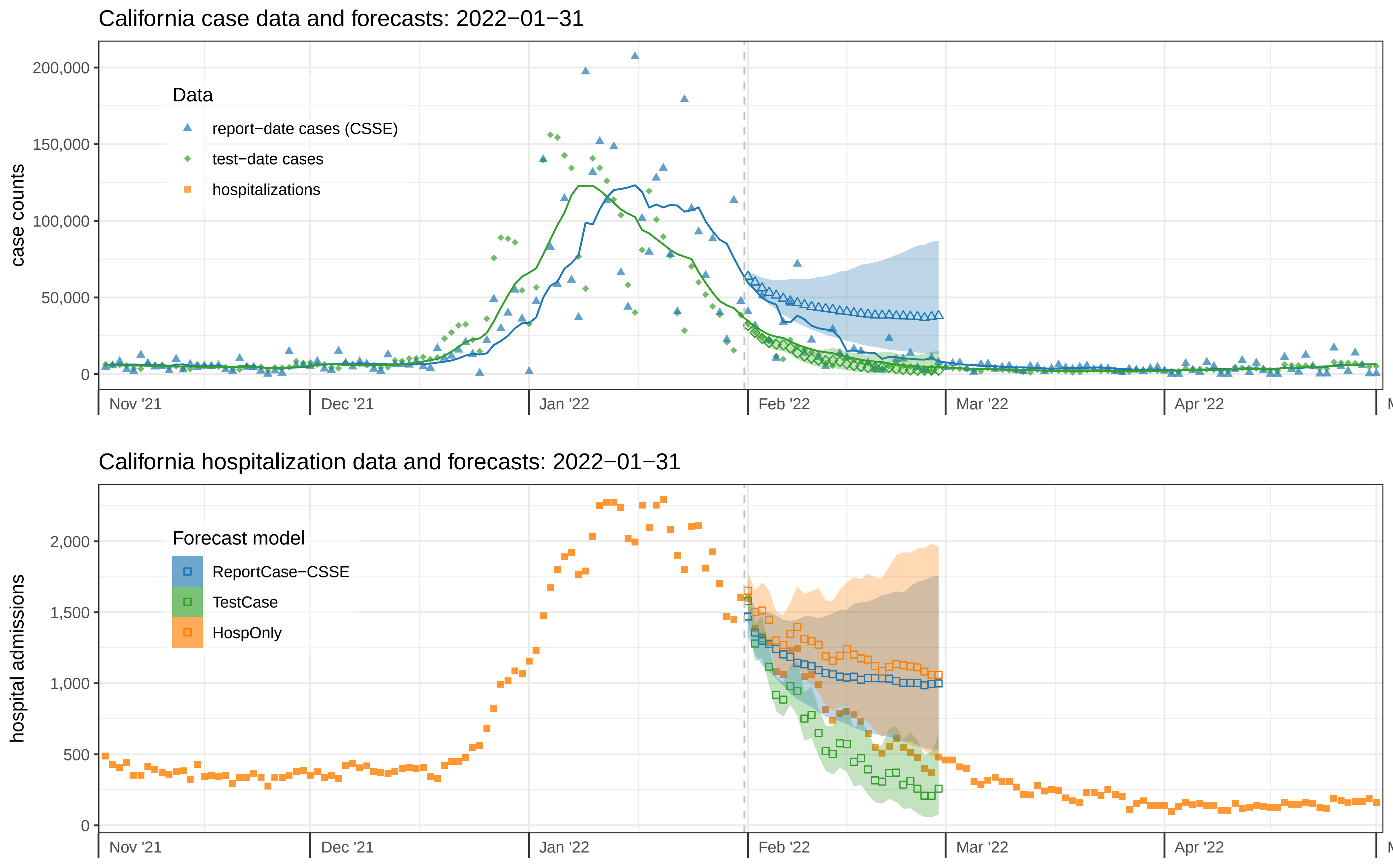
California case data and forecasts: 2022−01−31
case counts
200,000
150,000
100,000
50,000
0
Data
report−date cases (CSSE)
test−date cases
hospitalizations
Nov '21
Dec '21
Jan '22
Feb '22
Mar '22
Apr '22
California hospitalization data and forecasts: 2022−01−31
hospital admissions
2,000
1,500
1,000
500
0
Forecast model
ReportCase−CSSE
TestCase
HospOnly
Nov '21
Dec '21
Jan '22
Feb '22
Mar '22
Apr '22

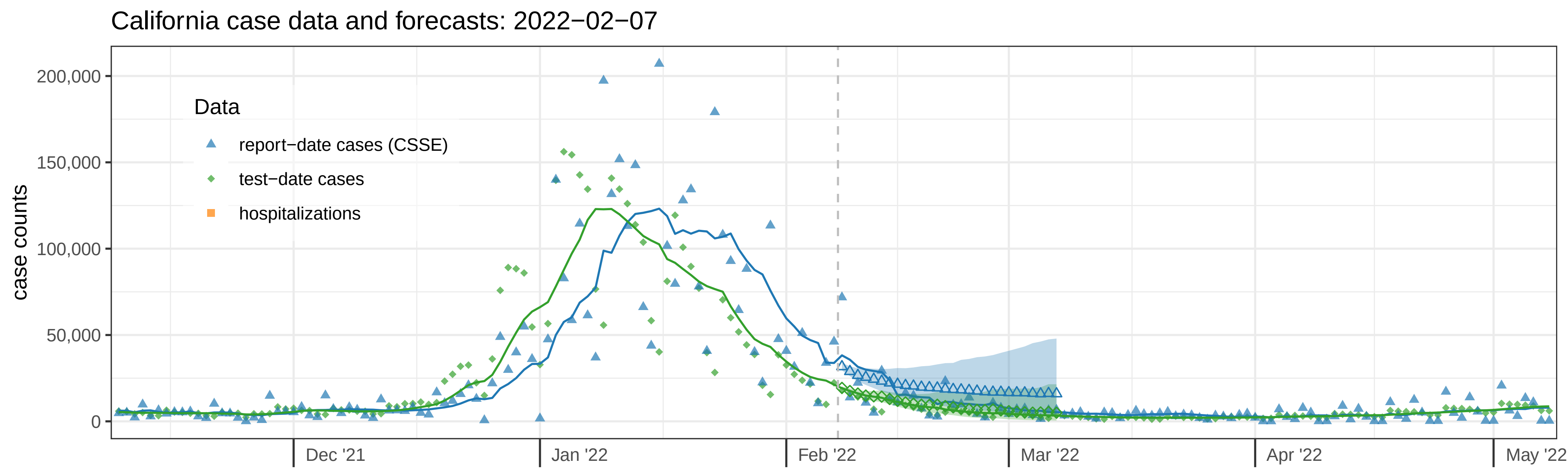
California case data and forecasts: 2022−02−07
case counts
200,000
150,000
100,000
50,000
0
Data
report−date cases (CSSE)
test−date cases
hospitalizations
Dec '21
Jan '22
Feb '22
Mar '22
Apr '22
May '22

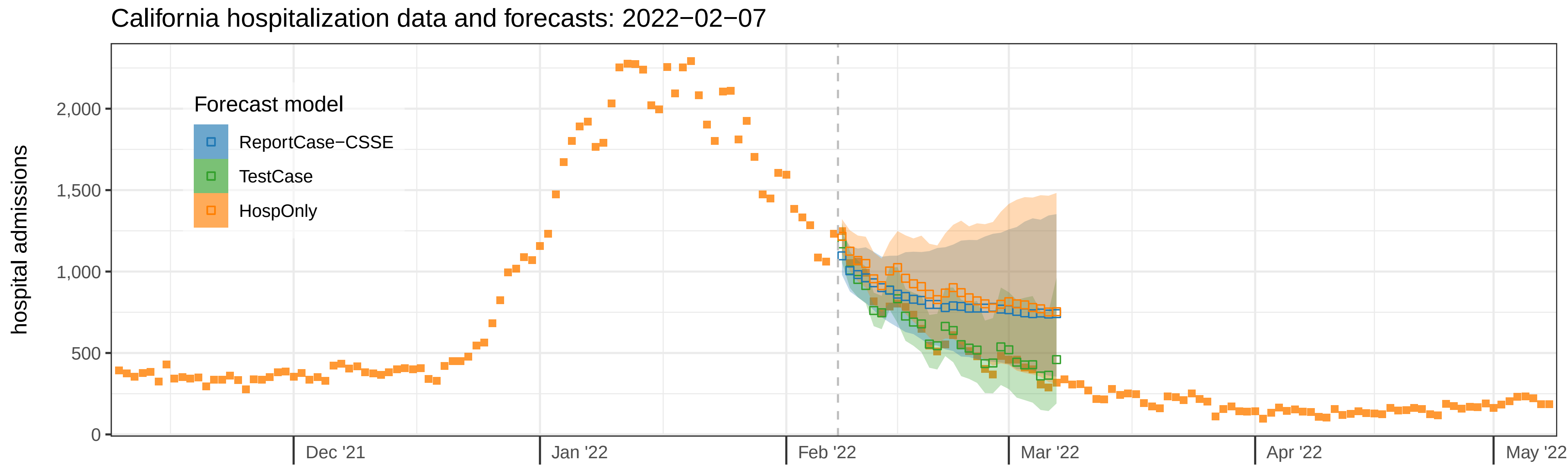
California hospitalization data and forecasts: 2022−02−07
hospital admissions
2,000
1,500
1,000
500
0
Forecast model
ReportCase−CSSE
TestCase
HospOnly
Dec '21
Jan '22
Feb '22
Mar '22
Apr '22
May '22

California case data and forecasts: 2022−02−14

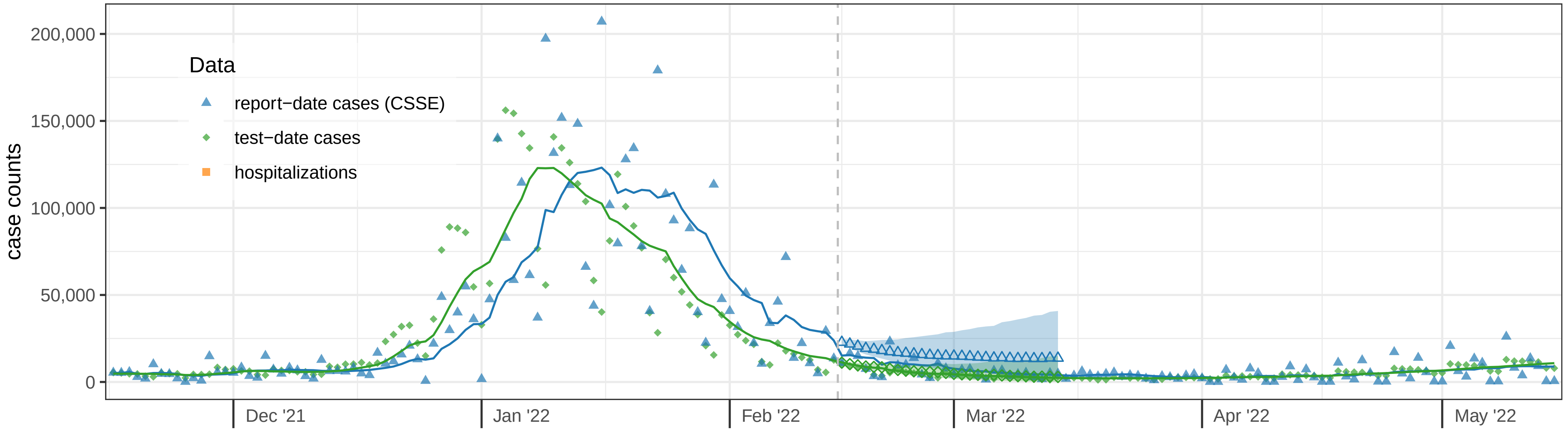


California hospitalization data and forecasts: 2022−02−14

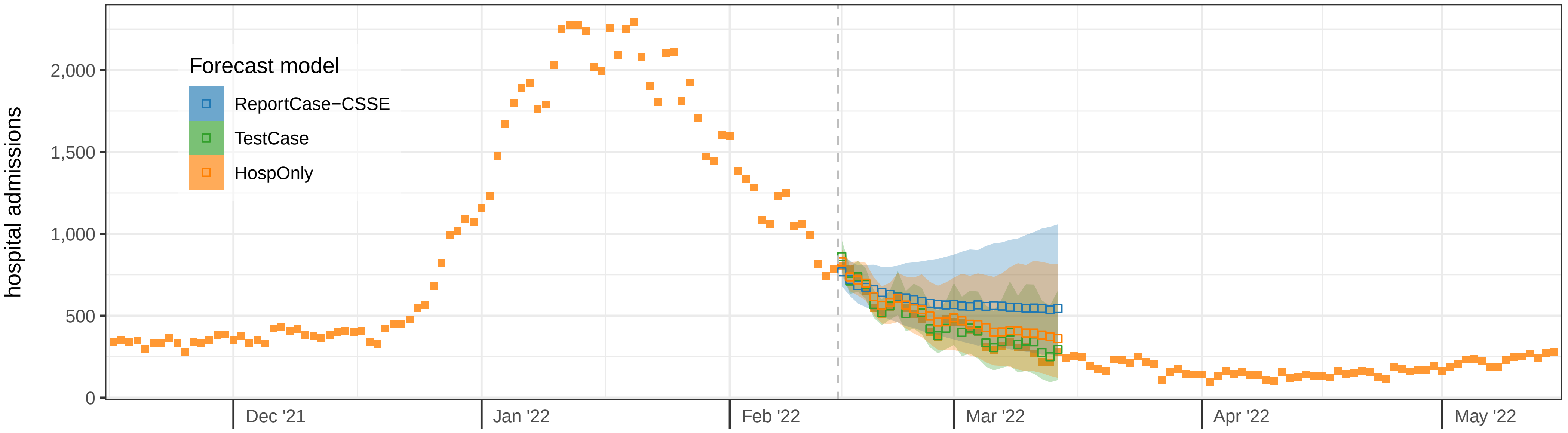

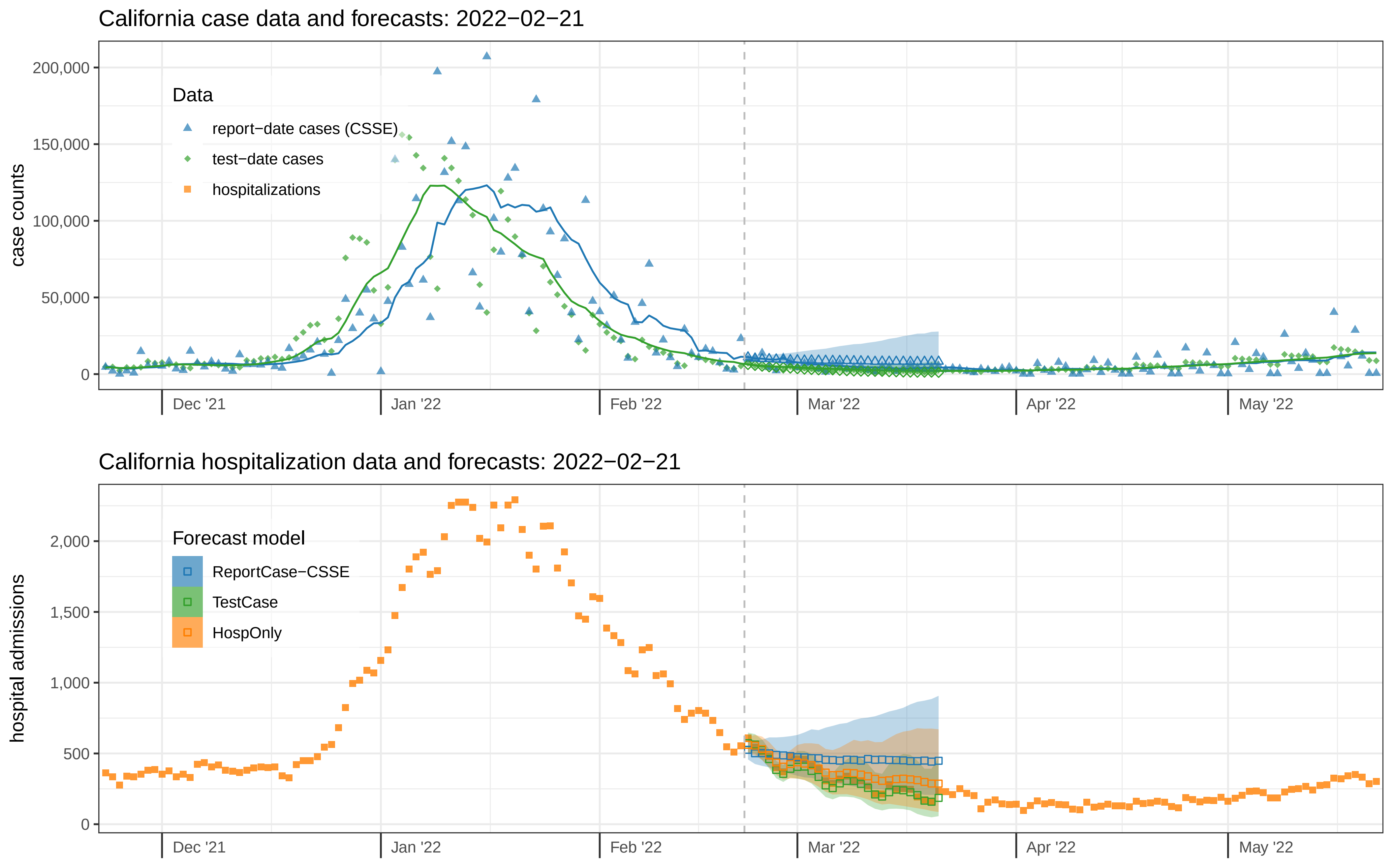

California case data and forecasts: 2022−02−21
case counts
200,000
150,000
100,000
50,000
0
Data
report−date cases (CSSE)
test−date cases
hospitalizations
Dec '21
Jan '22
Feb '22
Mar '22
Apr '22
May '22
California hospitalization data and forecasts: 2022−02−21
hospital admissions
2,000
1,500
1,000
500
0
Forecast model
ReportCase−CSSE
TestCase
HospOnly
Dec '21
Jan '22
Feb '22
Mar '22
Apr '22
May '22

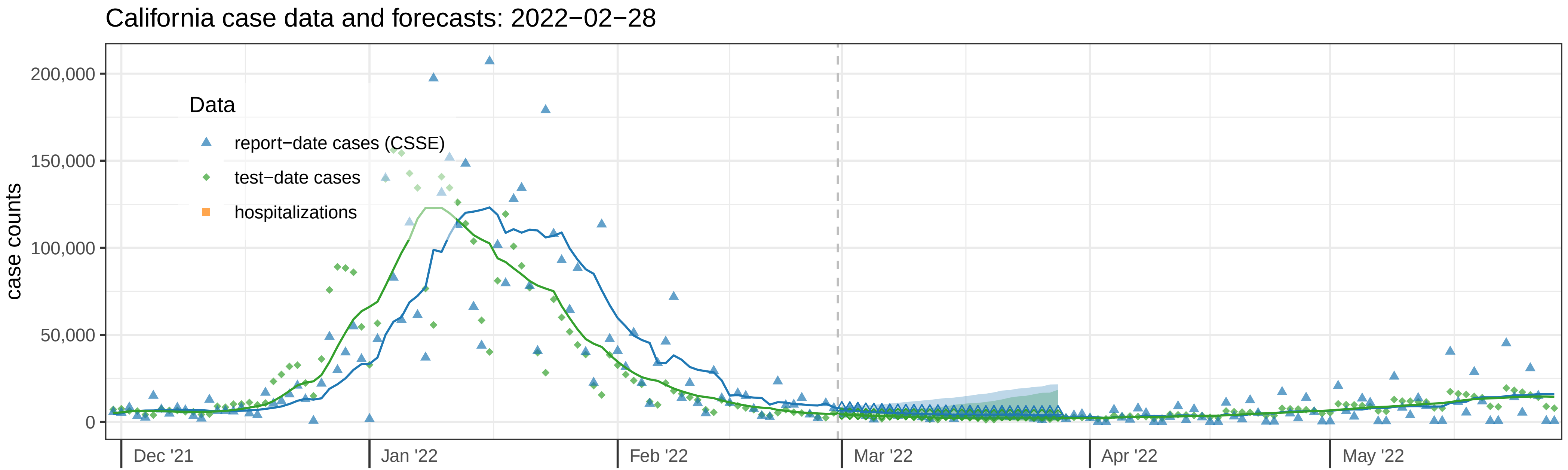
California case data and forecasts: 2022−02−28
case counts
200,000
150,000
100,000
50,000
0
Data
report−date cases (CSSE)
test−date cases
hospitalizations
Dec '21
Jan '22
Feb '22
Mar '22
Apr '22
May '22

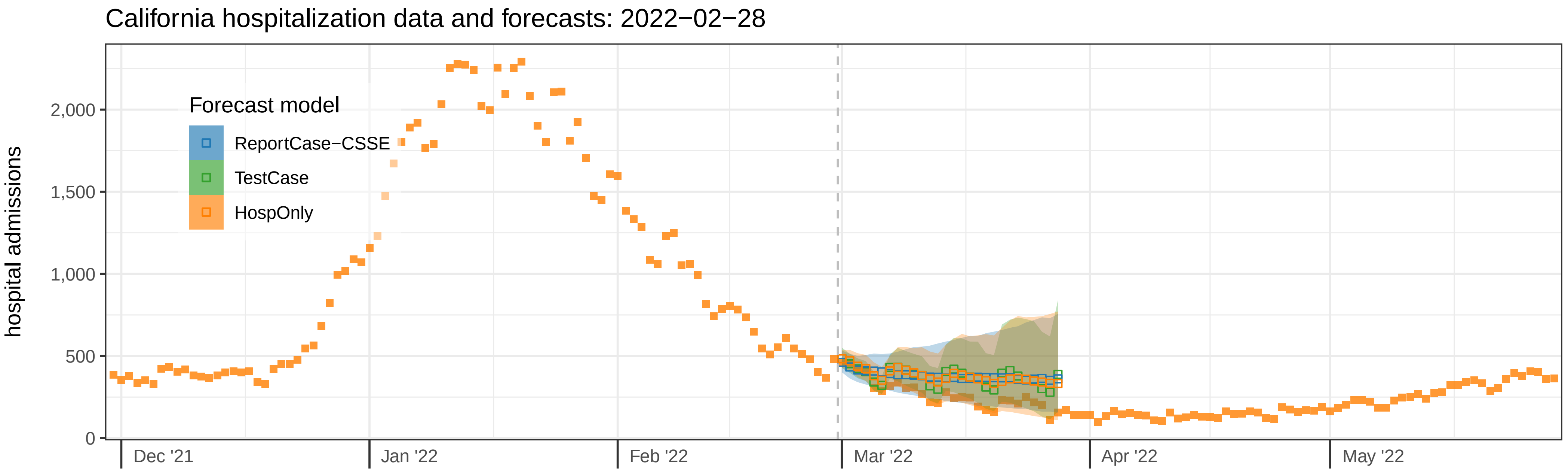
California hospitalization data and forecasts: 2022−02−28
hospital admissions
2,000
1,500
1,000
500
0
Forecast model
ReportCase−CSSE
TestCase
HospOnly
Dec '21
Jan '22
Feb '22
Mar '22
Apr '22
May '22

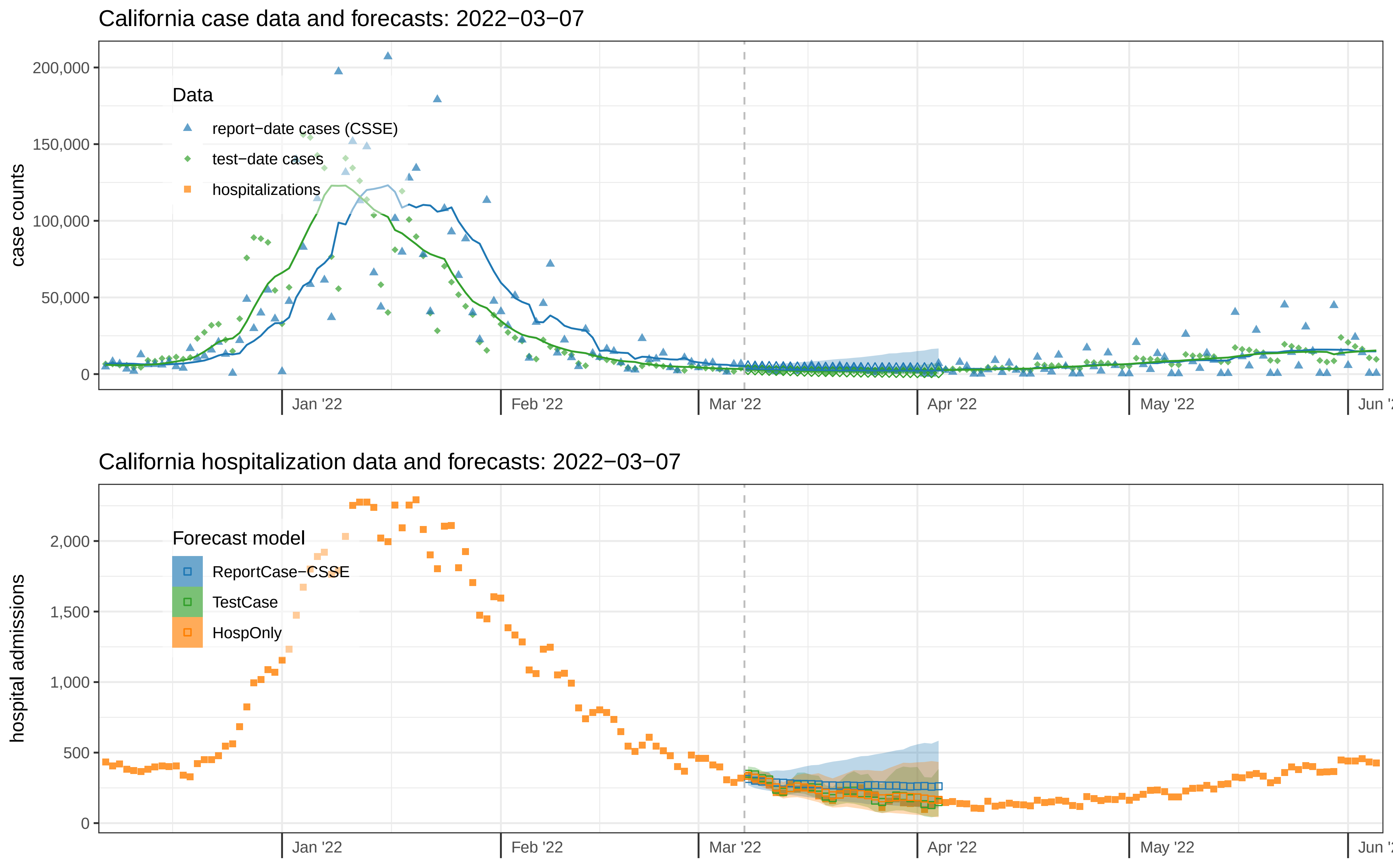

California case data and forecasts: 2022−03−07
case counts
200,000
150,000
100,000
50,000
0
Data
report−date cases (CSSE)
test−date cases
hospitalizations
Jan '22
Feb '22
Mar '22
Apr '22
May '22
California hospitalization data and forecasts: 2022−03−07
hospital admissions
2,000
1,500
1,000
500
0
Forecast model
ReportCase−CSSE
TestCase
HospOnly
Jan '22
Feb '22
Mar '22
Apr '22
May '22

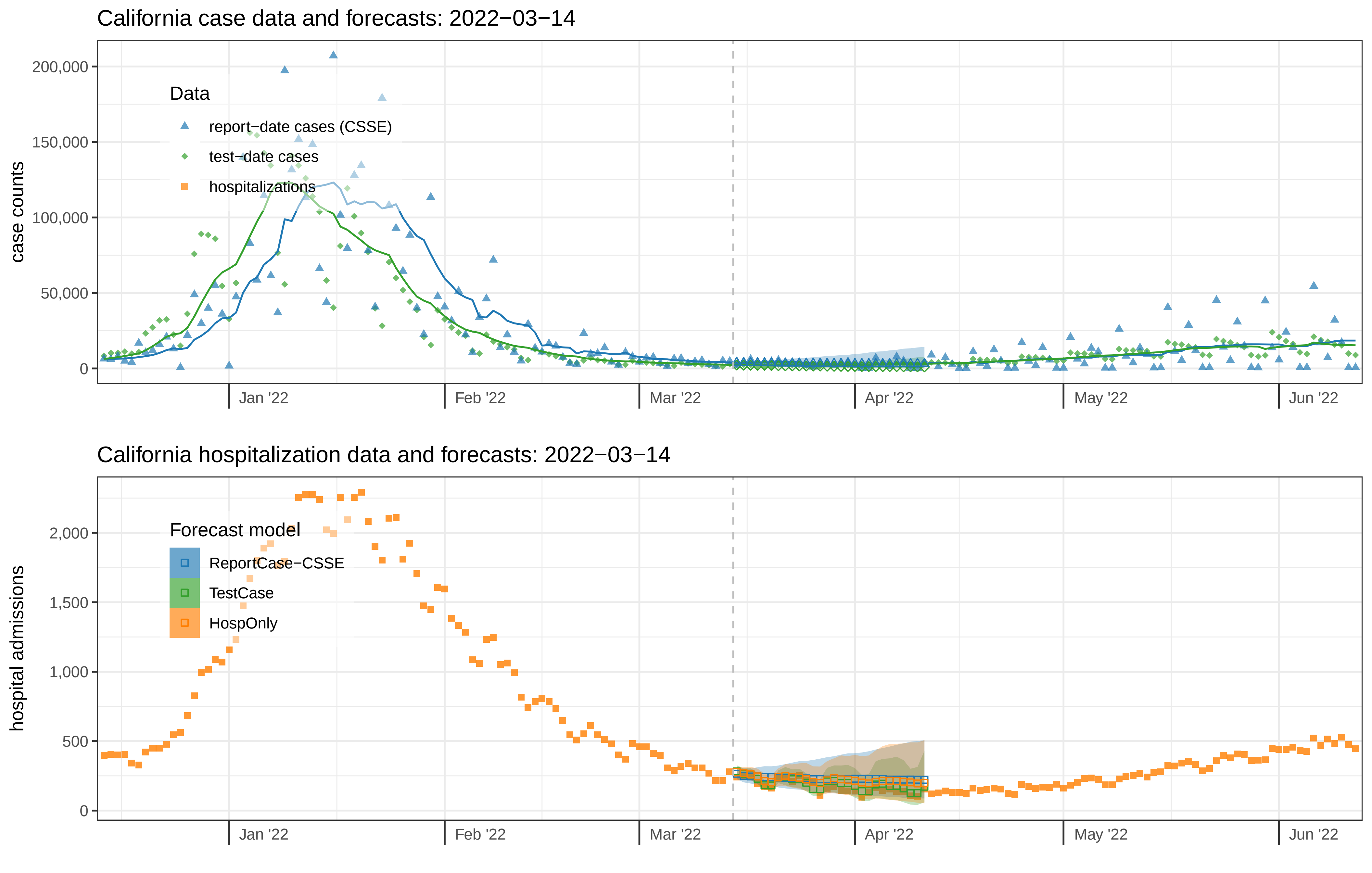
California case data and forecasts: 2022−03−14
case counts
200,000
150,000
100,000
50,000
0
Data
report−date cases (CSSE)
test−date cases
hospitalizations
Jan '22
Feb '22
Mar '22
Apr '22
May '22
Jun '22
California hospitalization data and forecasts: 2022−03−14
hospital admissions
2,000
1,500
1,000
500
0
Forecast model
ReportCase−CSSE
TestCase
HospOnly
Jan '22
Feb '22
Mar '22
Apr '22
May '22
Jun '22

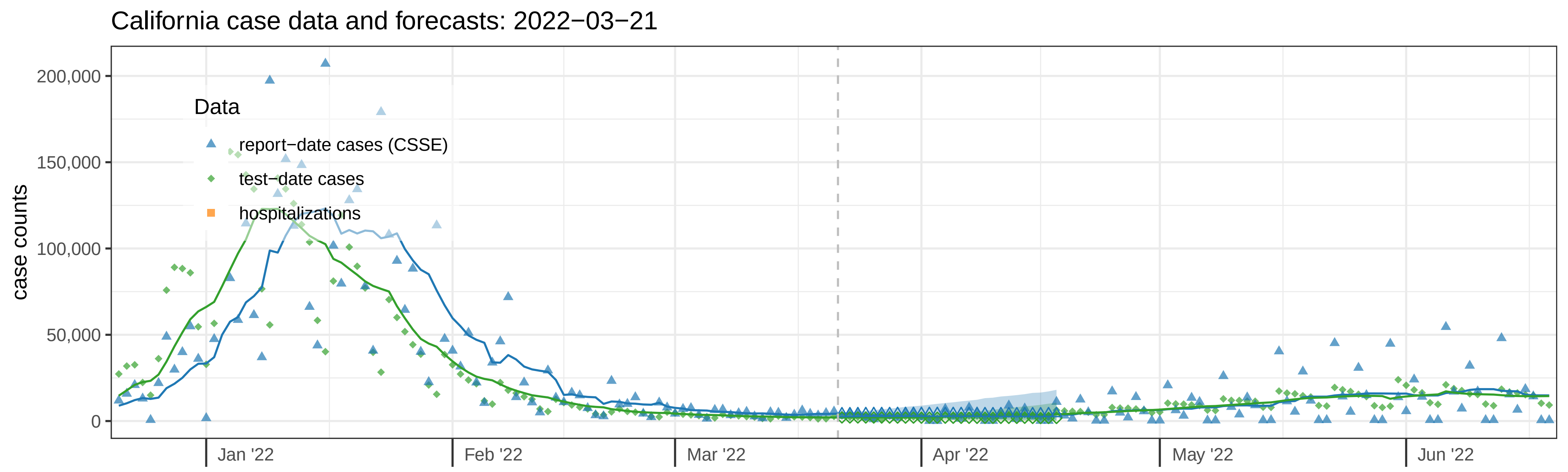
California case data and forecasts: 2022−03−21
case counts
200,000
150,000
100,000
50,000
0
Data
report−date cases (CSSE)
test−date cases
hospitalizations
Jan '22
Feb '22
Mar '22
Apr '22
May '22
Jun '22

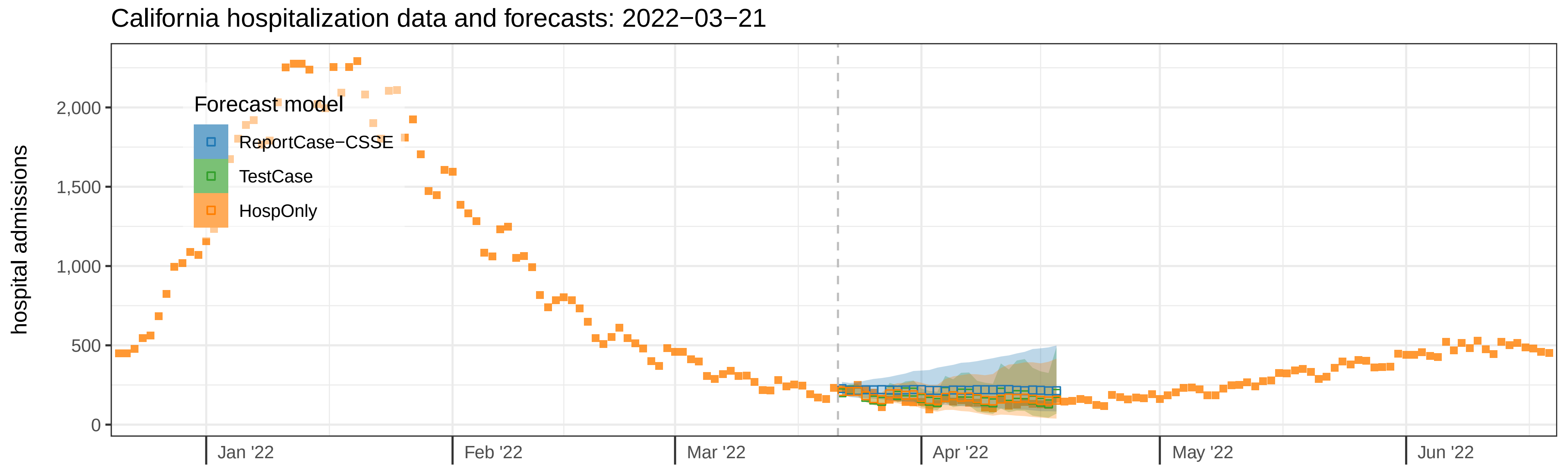
California hospitalization data and forecasts: 2022−03−21
hospital admissions
2,000
1,500
1,000
500
0
Forecast model
ReportCase−CSSE
TestCase
HospOnly
Jan '22
Feb '22
Mar '22
Apr '22
May '22
Jun '22

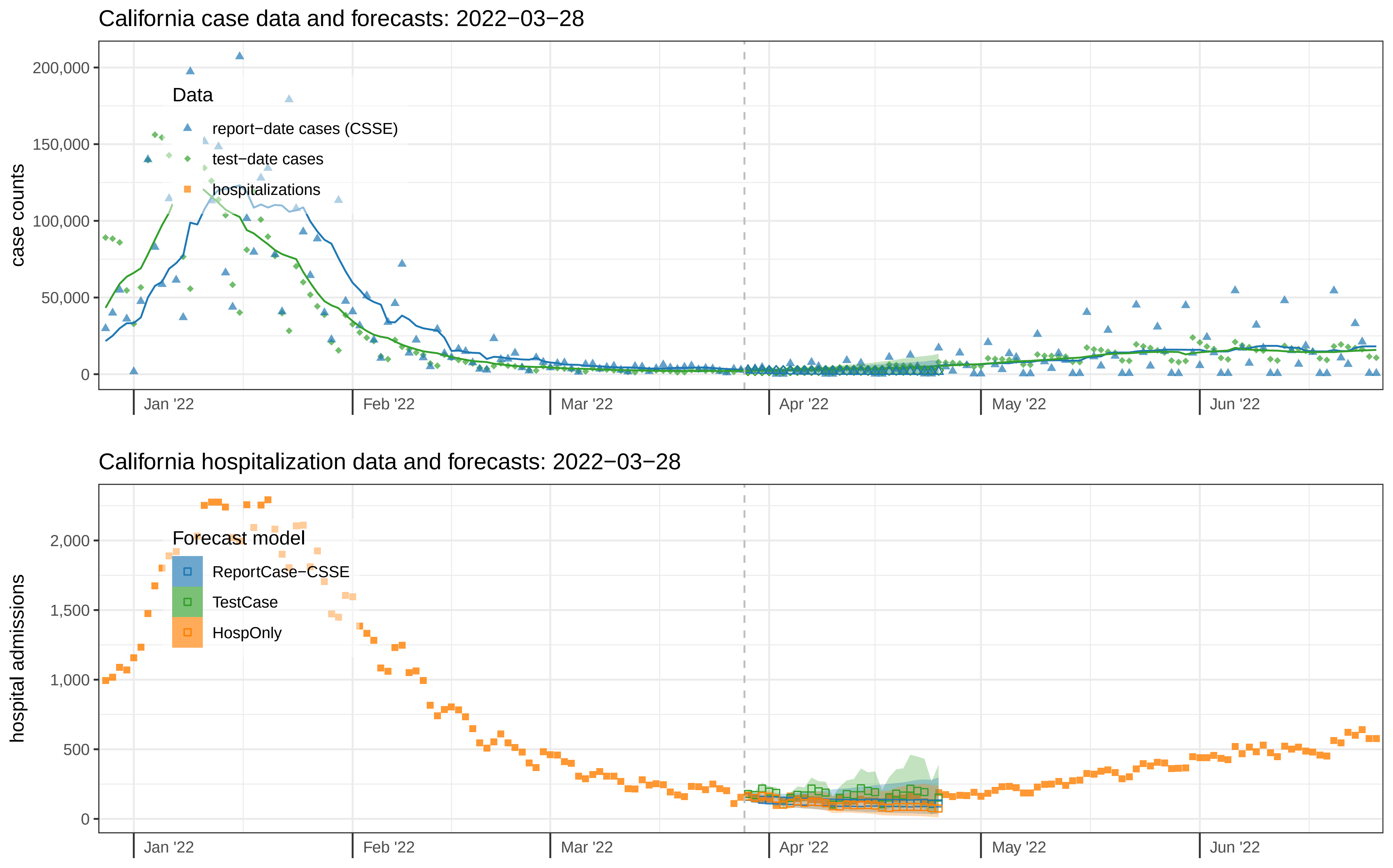
California case data and forecasts: 2022−03−28
case counts
200,000
150,000
100,000
50,000
0
Data
report−date cases (CSSE)
test−date cases
hospitalizations
Jan '22
Feb '22
Mar '22
Apr '22
May '22
Jun '22
California hospitalization data and forecasts: 2022−03−28
hospital admissions
2,000
1,500
1,000
500
0
Forecast model
ReportCase−CSSE
TestCase
HospOnly
Jan '22
Feb '22
Mar '22
Apr '22
May '22
Jun '22

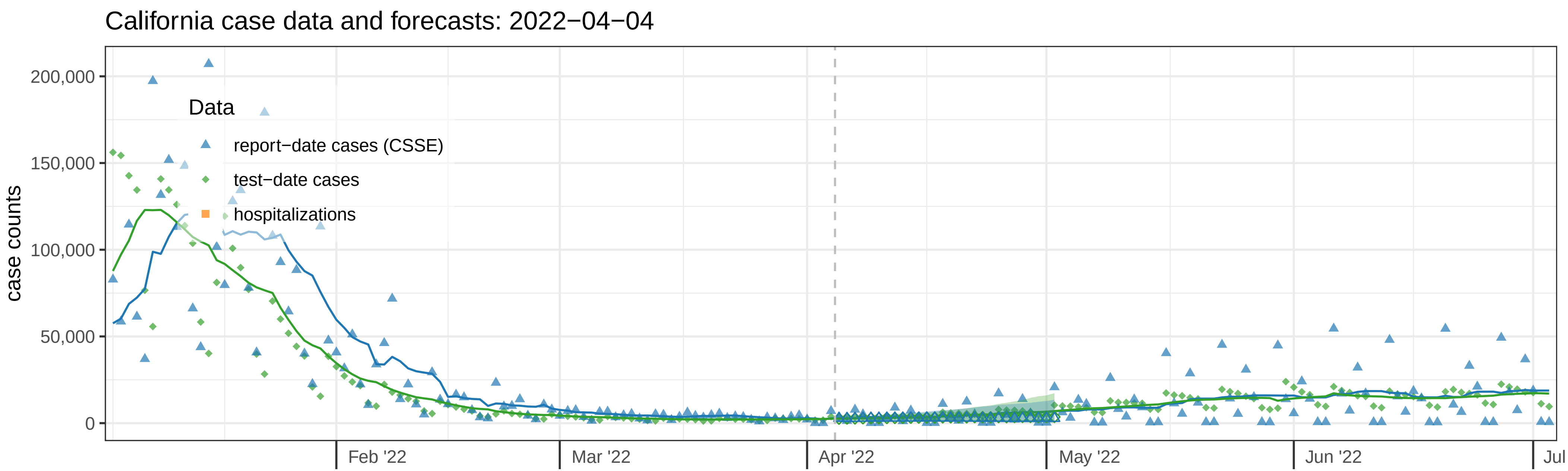

California case data and forecasts: 2022−04−04
case counts
200,000
150,000
100,000
50,000
0
Data
report−date cases (CSSE)
test−date cases
hospitalizations
Feb '22
Mar '22
Apr '22
May '22
Jun '22
Jul


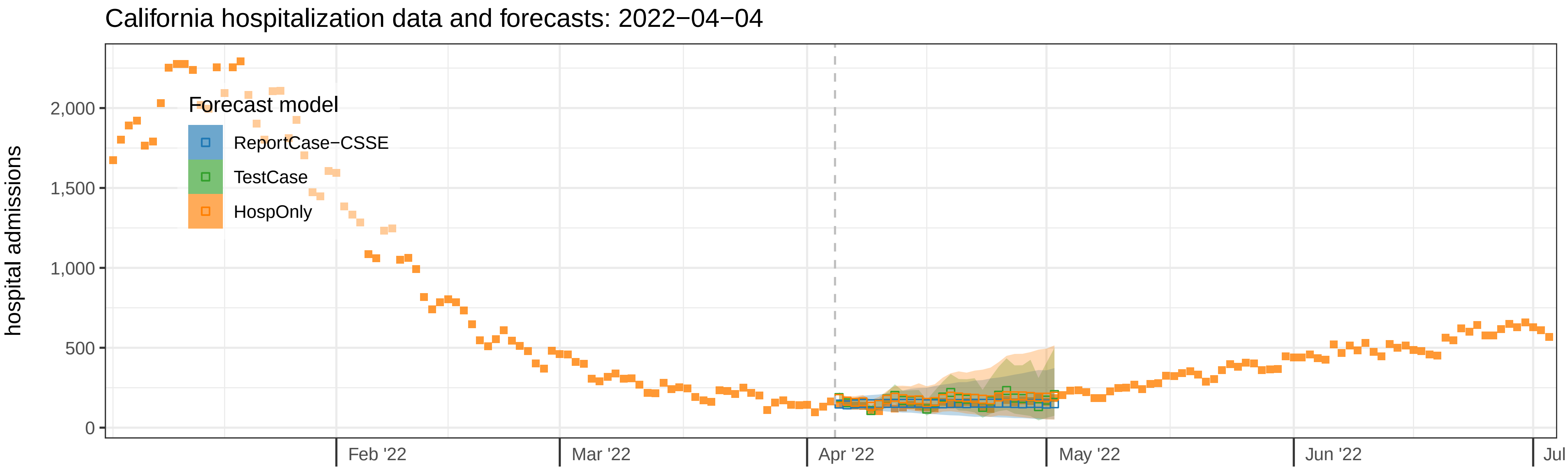

California hospitalization data and forecasts: 2022−04−04
hospital admissions
2,000
1,500
1,000
500
0
Forecast model
ReportCase−CSSE
TestCase
HospOnly
Feb '22
Mar '22
Apr '22
May '22
Jun '22
Jul

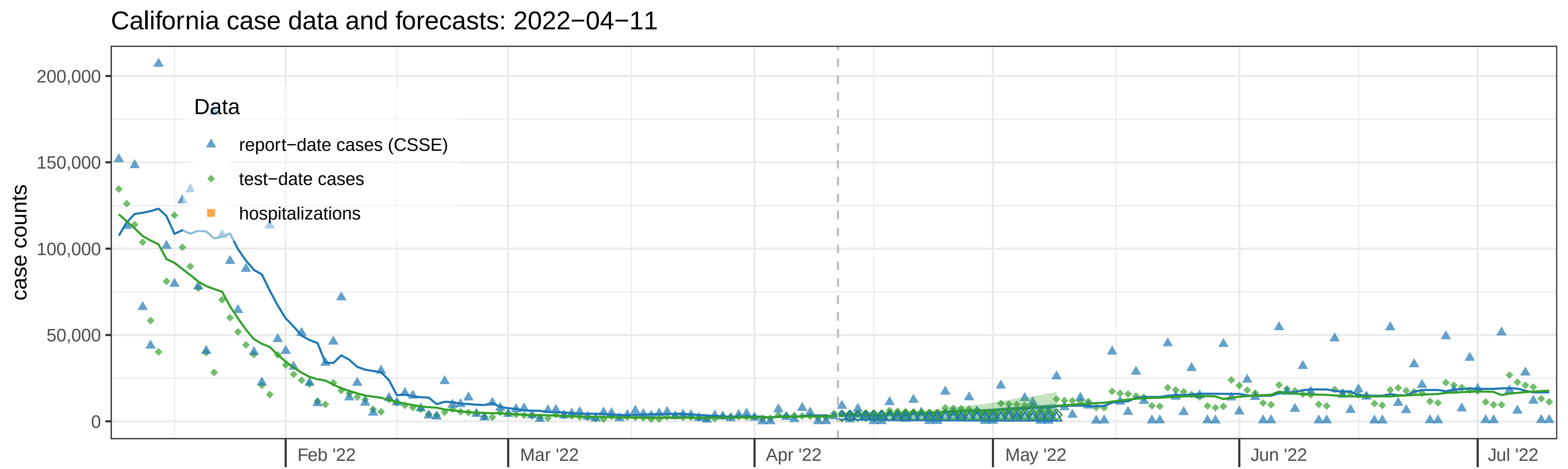
California case data and forecasts: 2022−04−11
case counts
200,000
150,000
100,000
50,000
0
Data
report−date cases (CSSE)
test−date cases
hospitalizations
Feb '22
Mar '22
Apr '22
May '22
Jun '22
Jul '22

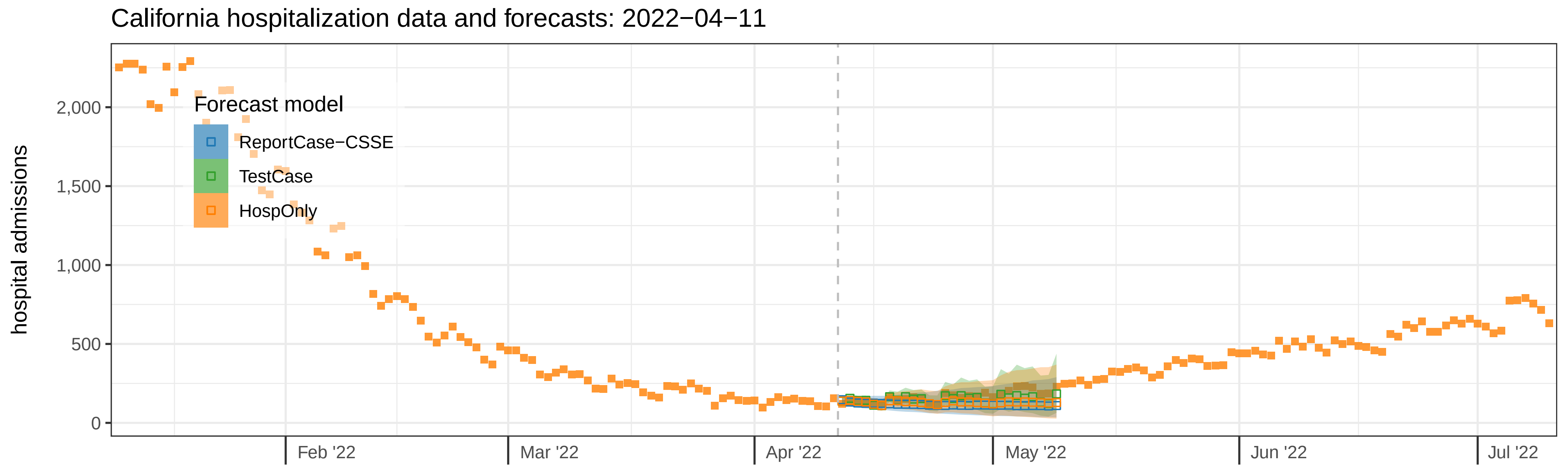
California hospitalization data and forecasts: 2022−04−11
hospital admissions
2,000
1,500
1,000
500
0
Forecast model
ReportCase−CSSE
TestCase
HospOnly
Feb '22
Mar '22
Apr '22
May '22
Jun '22
Jul '22

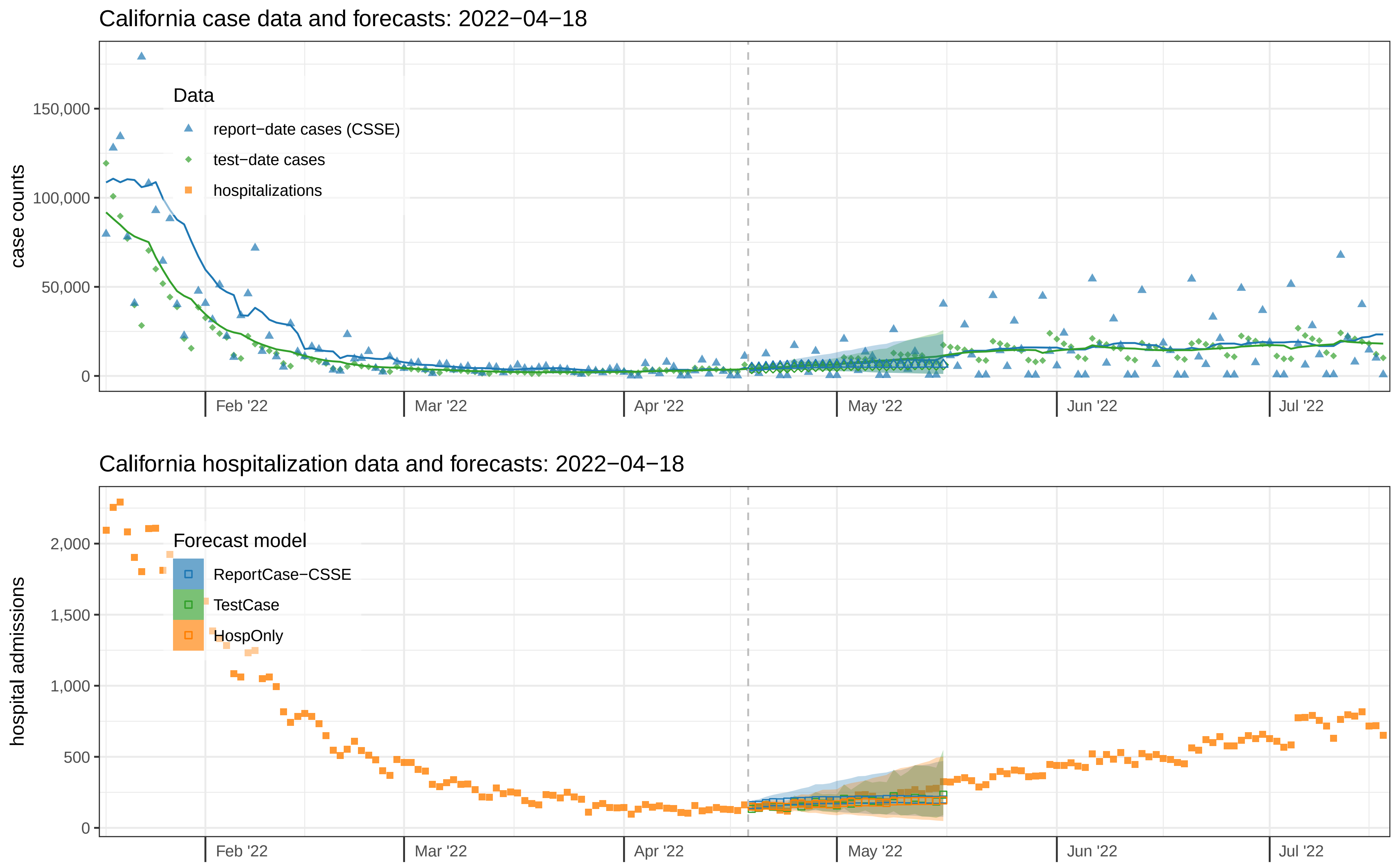
California case data and forecasts: 2022−04−18
case counts
150,000
100,000
50,000
0
Data
report−date cases (CSSE)
test−date cases
hospitalizations
Feb '22
Mar '22
Apr '22
May '22
Jun '22
Jul '22
California hospitalization data and forecasts: 2022−04−18
hospital admissions
2,000
1,500
1,000
500
0
Forecast model
ReportCase−CSSE
TestCase
HospOnly
Feb '22
Mar '22
Apr '22
May '22
Jun '22
Jul '22

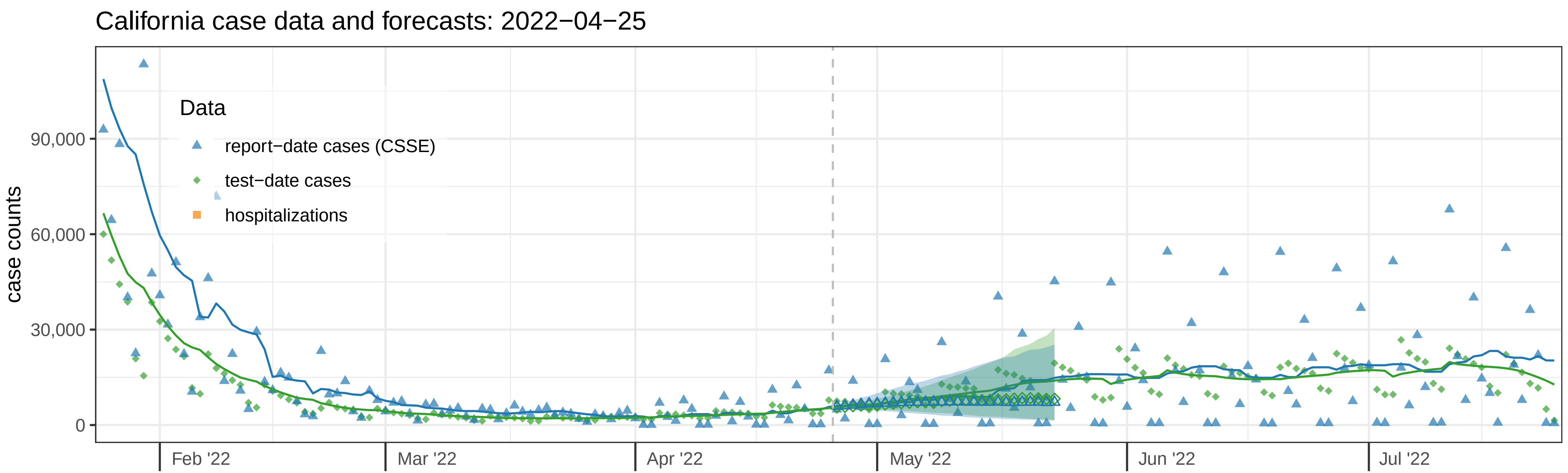
California case data and forecasts: 2022−04−25
case counts
90,000
60,000
30,000
0
Data
report−date cases (CSSE)
test−date cases
hospitalizations
Feb '22
Mar '22
Apr '22
May '22
Jun '22
Jul '22

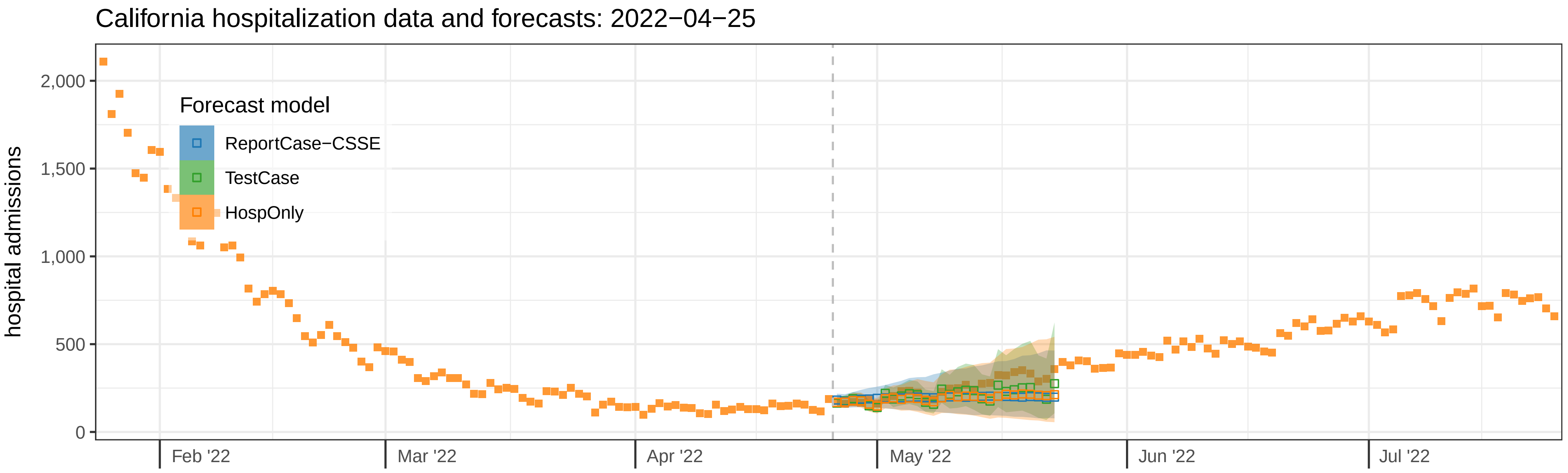
California hospitalization data and forecasts: 2022−04−25
hospital admissions
2,000
1,500
1,000
500
0
Forecast model
ReportCase−CSSE
TestCase
HospOnly
Feb '22
Mar '22
Apr '22
May '22
Jun '22
Jul '22

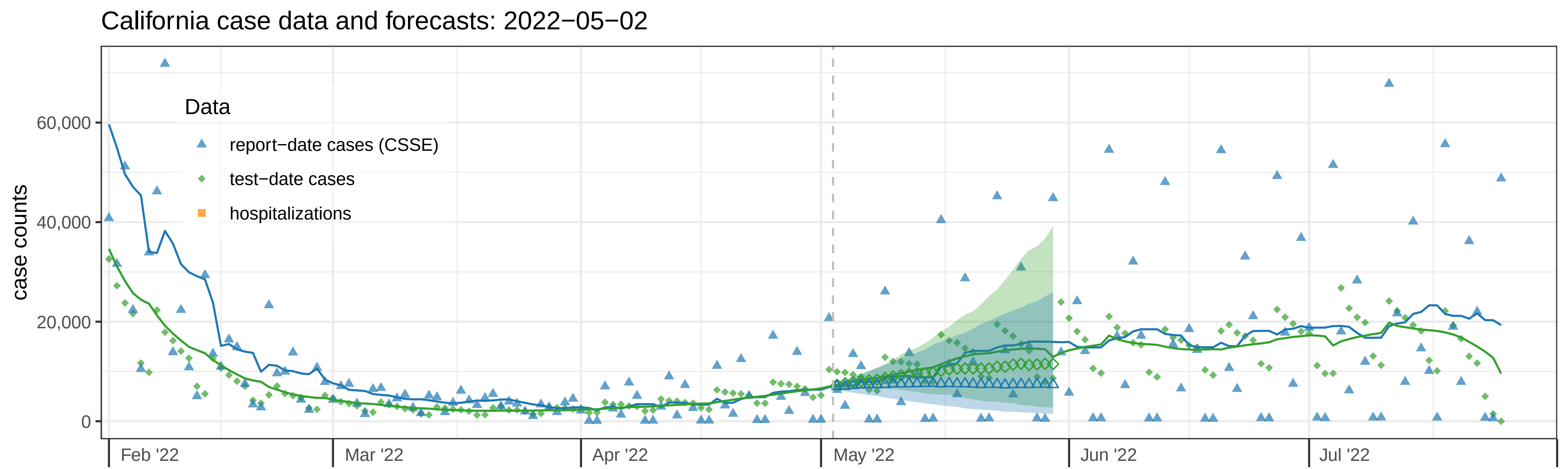
California case data and forecasts: 2022−05−02
case counts
0
20,000
40,000
60,000
Data
report−date cases (CSSE)
test−date cases
hospitalizations
Feb '22
Mar '22
Apr '22
May '22
Jun '22
Jul '22

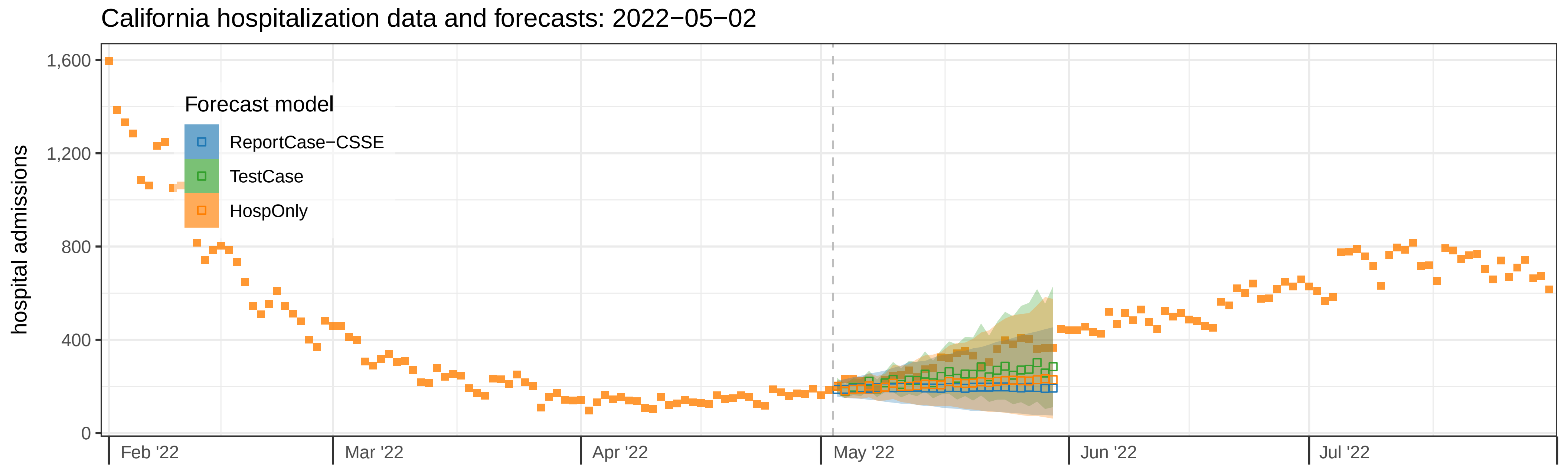
California hospitalization data and forecasts: 2022−05−02
hospital admissions
0
400
800
1,200
1,600
Forecast model
ReportCase−CSSE
TestCase
HospOnly
Feb '22
Mar '22
Apr '22
May '22
Jun '22
Jul '22

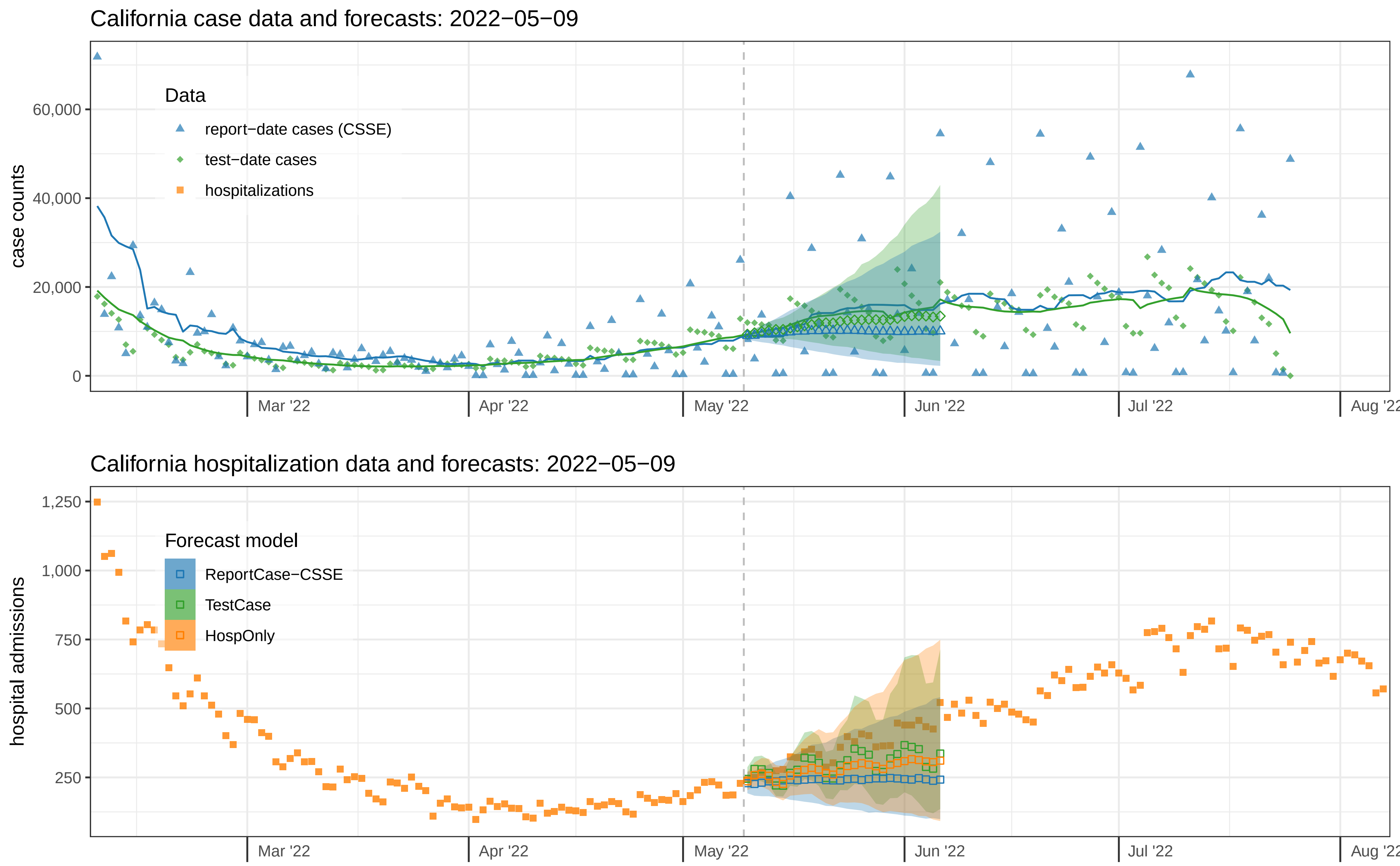

California case data and forecasts: 2022−05−09
case counts
0
20,000
40,000
60,000
Data
report−date cases (CSSE)
test−date cases
hospitalizations
Mar '22
Apr '22
May '22
Jun '22
Jul '22
Aug '22
California hospitalization data and forecasts: 2022−05−09
hospital admissions
250
500
750
1,000
1,250
Forecast model
ReportCase−CSSE
TestCase
HospOnly
Mar '22
Apr '22
May '22
Jun '22
Jul '22
Aug '22

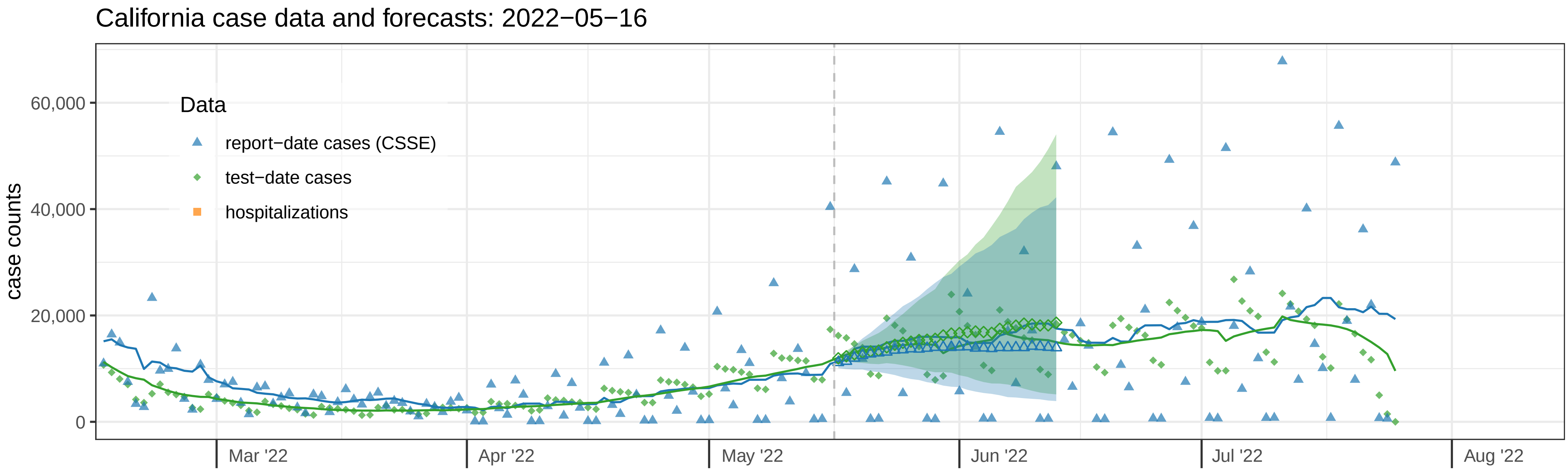

California case data and forecasts: 2022−05−16
case counts
60,000
40,000
20,000
0
Data
report−date cases (CSSE)
test−date cases
hospitalizations
Mar '22
Apr '22
May '22
Jun '22
Jul '22
Aug '22


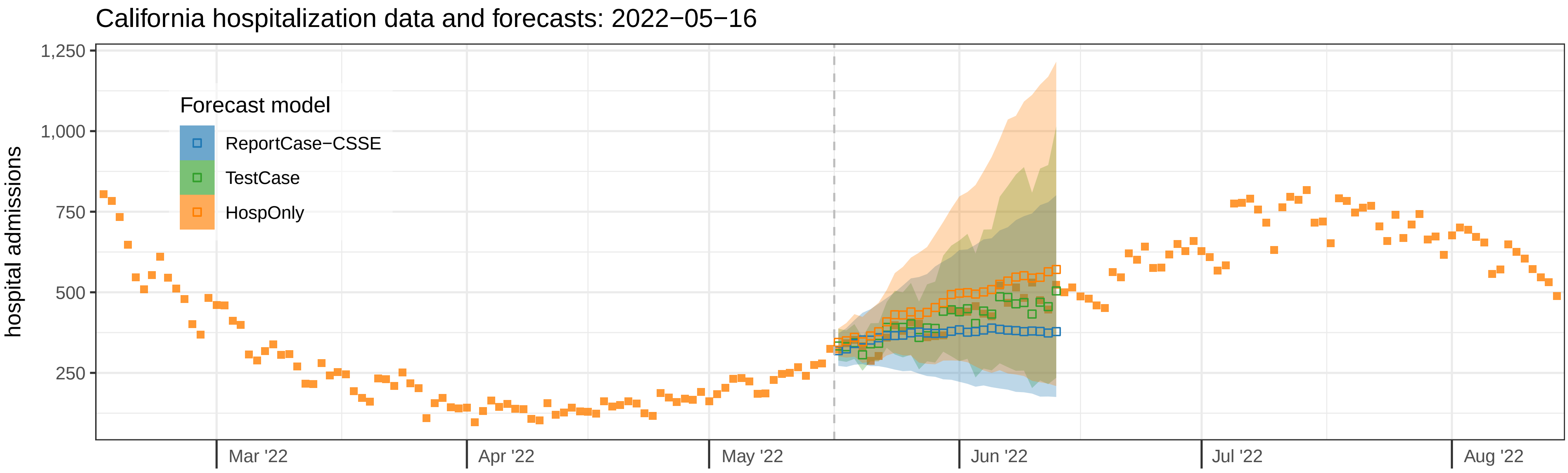

California hospitalization data and forecasts: 2022−05−16
hospital admissions
1,250
1,000
750
500
250
Forecast model
ReportCase−CSSE
TestCase
HospOnly
Mar '22
Apr '22
May '22
Jun '22
Jul '22
Aug '22

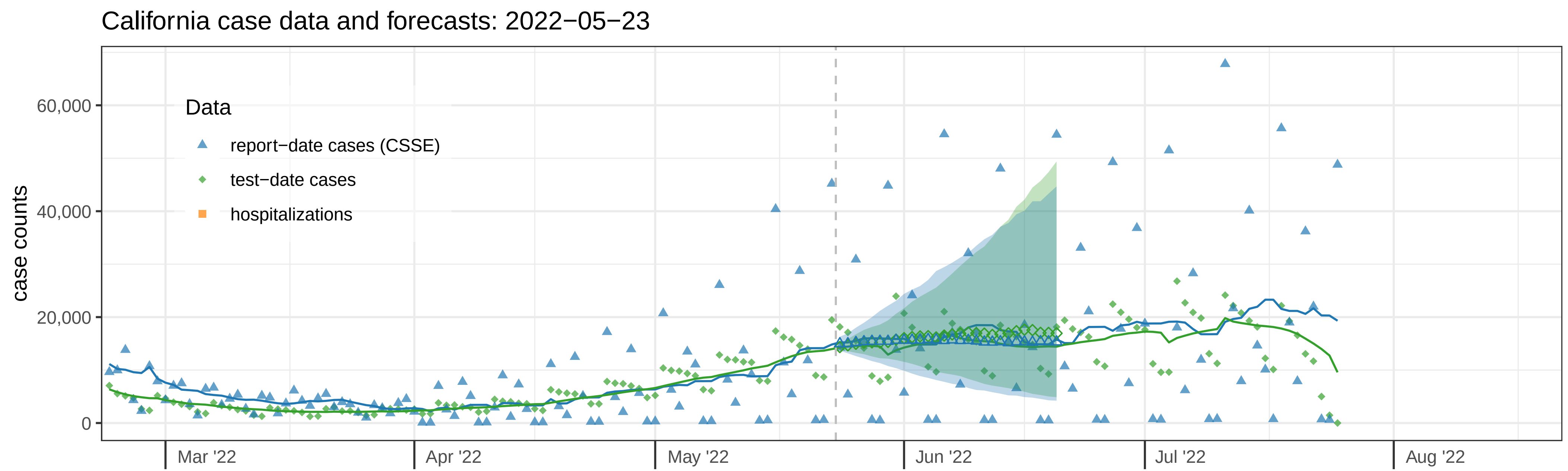
California case data and forecasts: 2022−05−23
case counts
60,000
40,000
20,000
0
Data
report−date cases (CSSE)
test−date cases
hospitalizations
Mar '22
Apr '22
May '22
Jun '22
Jul '22
Aug '22

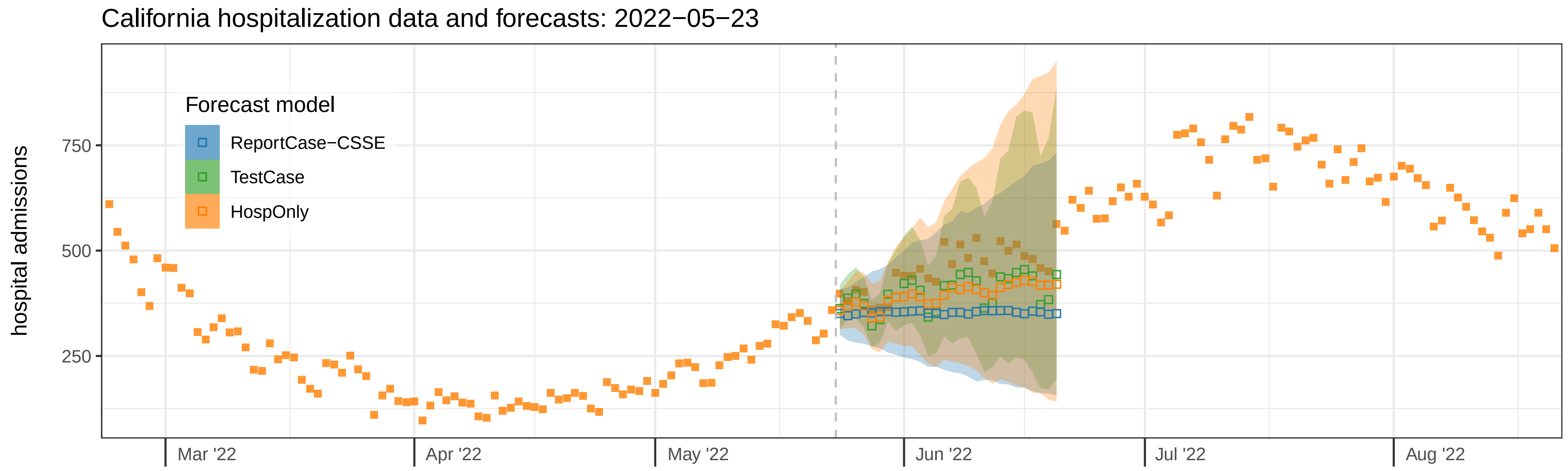
California hospitalization data and forecasts: 2022−05−23
hospital admissions
750
500
250
Forecast model
ReportCase−CSSE
TestCase
HospOnly
Mar '22
Apr '22
May '22
Jun '22
Jul '22
Aug '22

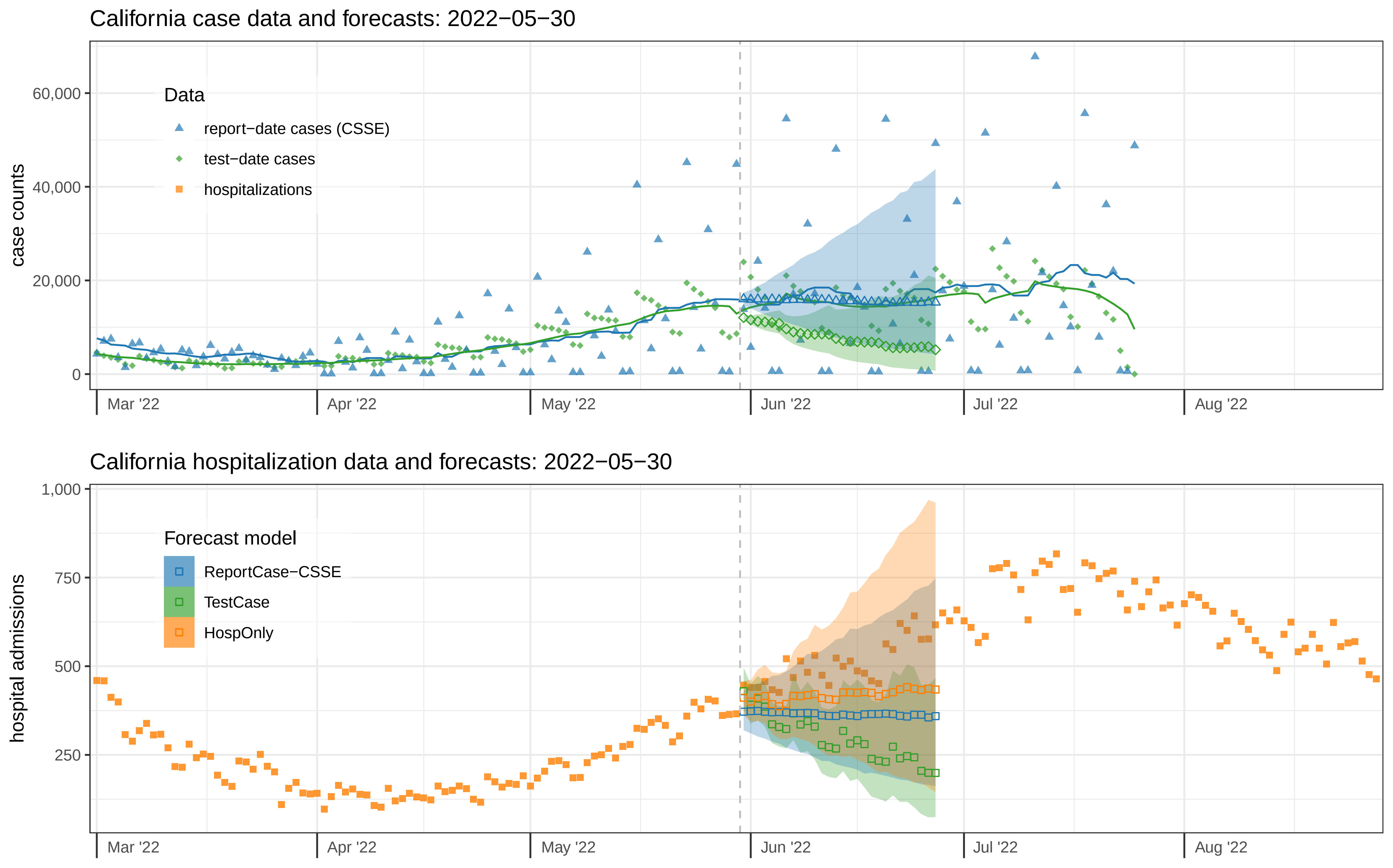

California case data and forecasts: 2022−05−30
case counts
60,000
40,000
20,000
0
Data
report−date cases (CSSE)
test−date cases
hospitalizations
Mar '22
Apr '22
May '22
Jun '22
Jul '22
Aug '22
California hospitalization data and forecasts: 2022−05−30
hospital admissions
1,000
750
500
250
Forecast model
ReportCase−CSSE
TestCase
HospOnly
Mar '22
Apr '22
May '22
Jun '22
Jul '22
Aug '22

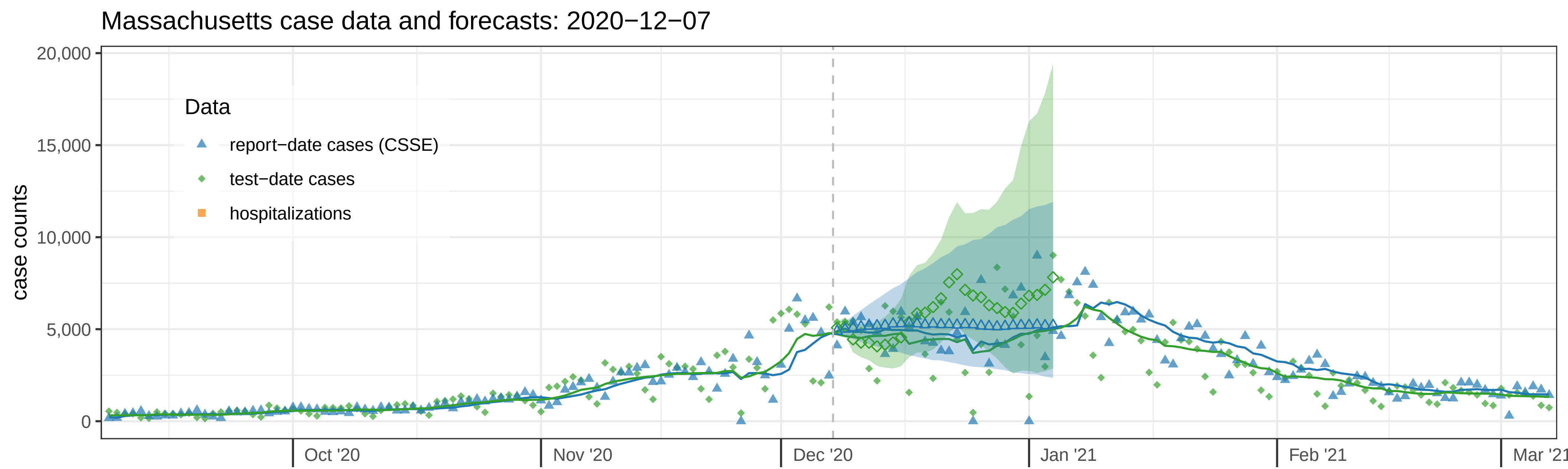

Massachusetts case data and forecasts: 2020−12−07
case counts
20,000
15,000
10,000
5,000
0
Data
report−date cases (CSSE)
test−date cases
hospitalizations
Oct '20
Nov '20
Dec '20
Jan '21
Feb '21
Mar '21


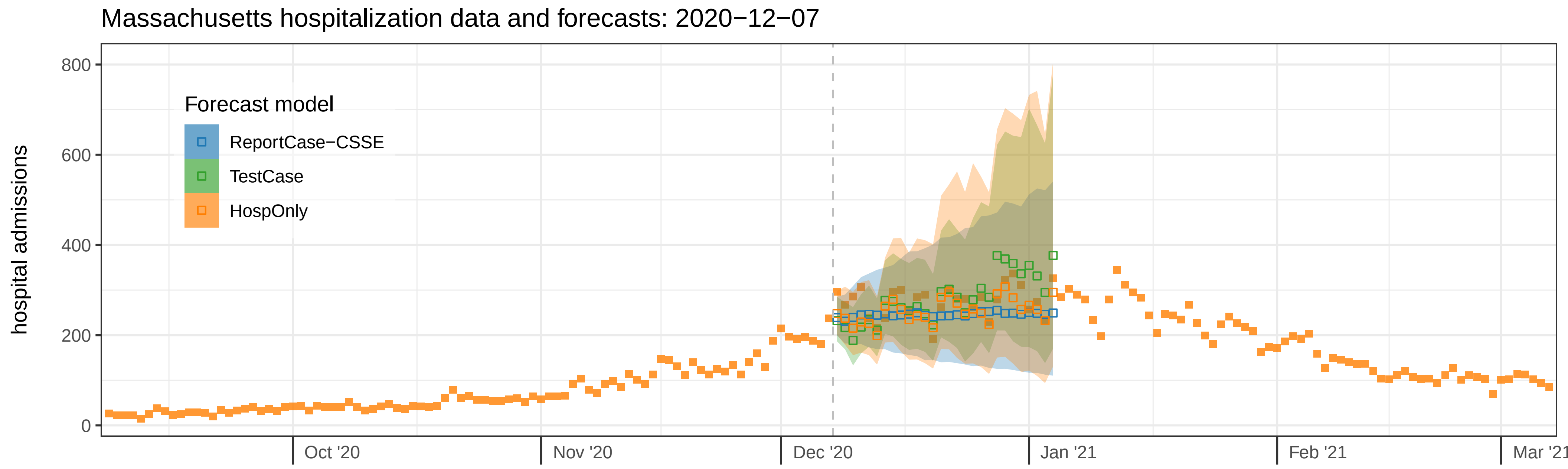

Massachusetts hospitalization data and forecasts: 2020−12−07
hospital admissions
800
600
400
200
0
Forecast model
ReportCase−CSSE
TestCase
HospOnly
Oct '20
Nov '20
Dec '20
Jan '21
Feb '21
Mar '21

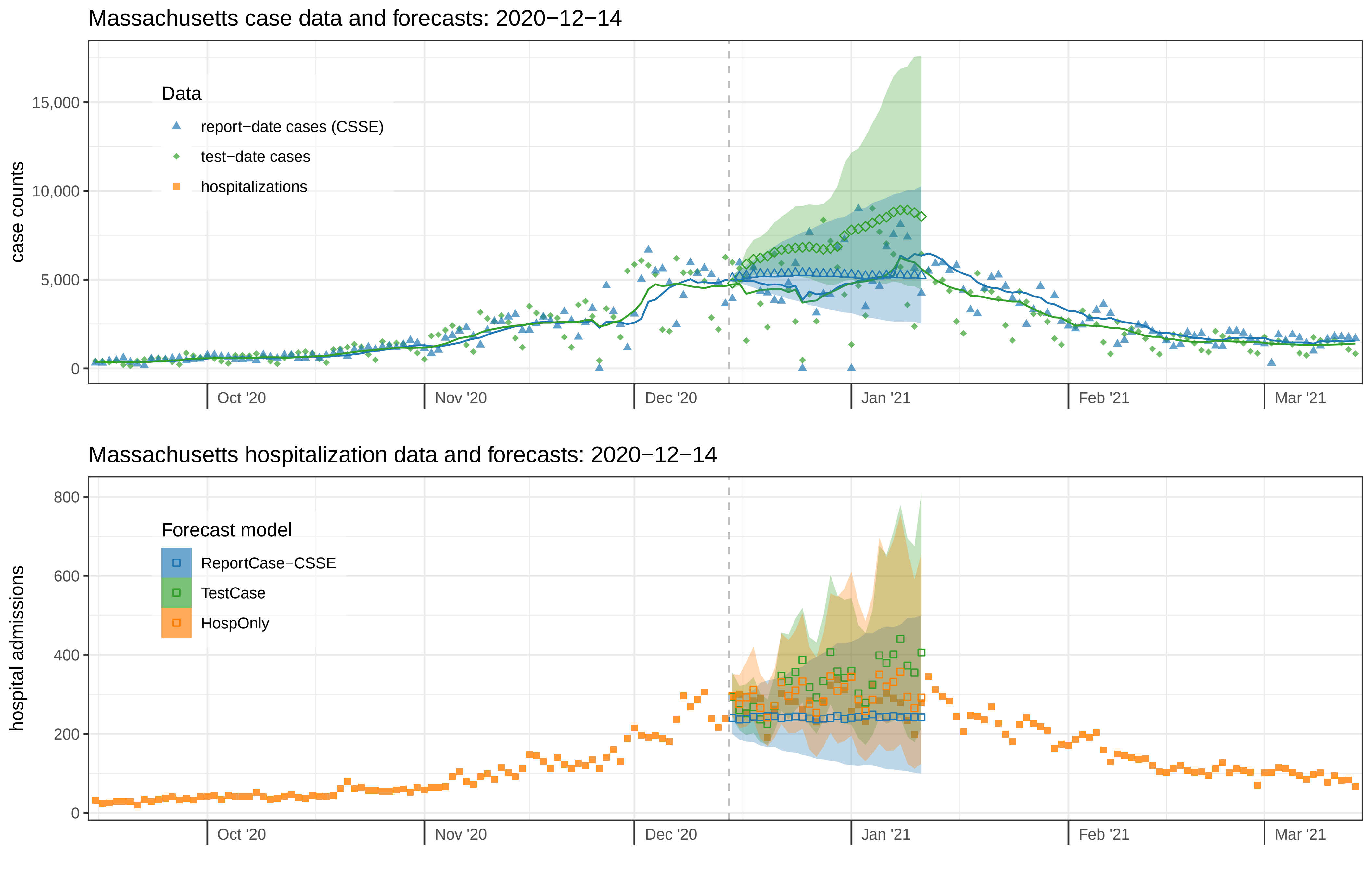

Massachusetts case data and forecasts: 2020−12−14
case counts
15,000
10,000
5,000
0
Data
report−date cases (CSSE)
test−date cases
hospitalizations
Oct '20
Nov '20
Dec '20
Jan '21
Feb '21
Mar '21
Massachusetts hospitalization data and forecasts: 2020−12−14
hospital admissions
800
600
400
200
0
Forecast model
ReportCase−CSSE
TestCase
HospOnly
Oct '20
Nov '20
Dec '20
Jan '21
Feb '21
Mar '21

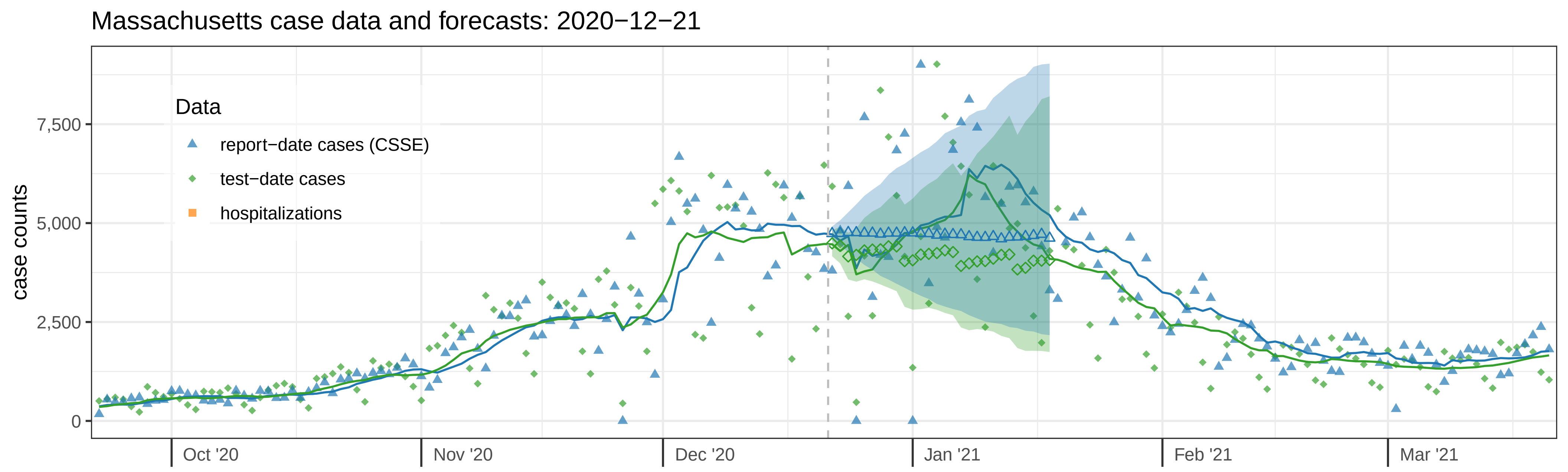

Massachusetts case data and forecasts: 2020−12−21
Data
report−date cases (CSSE)
test−date cases
hospitalizations
case counts
7,500
5,000
2,500
0
Oct '20
Nov '20
Dec '20
Jan '21
Feb '21
Mar '21


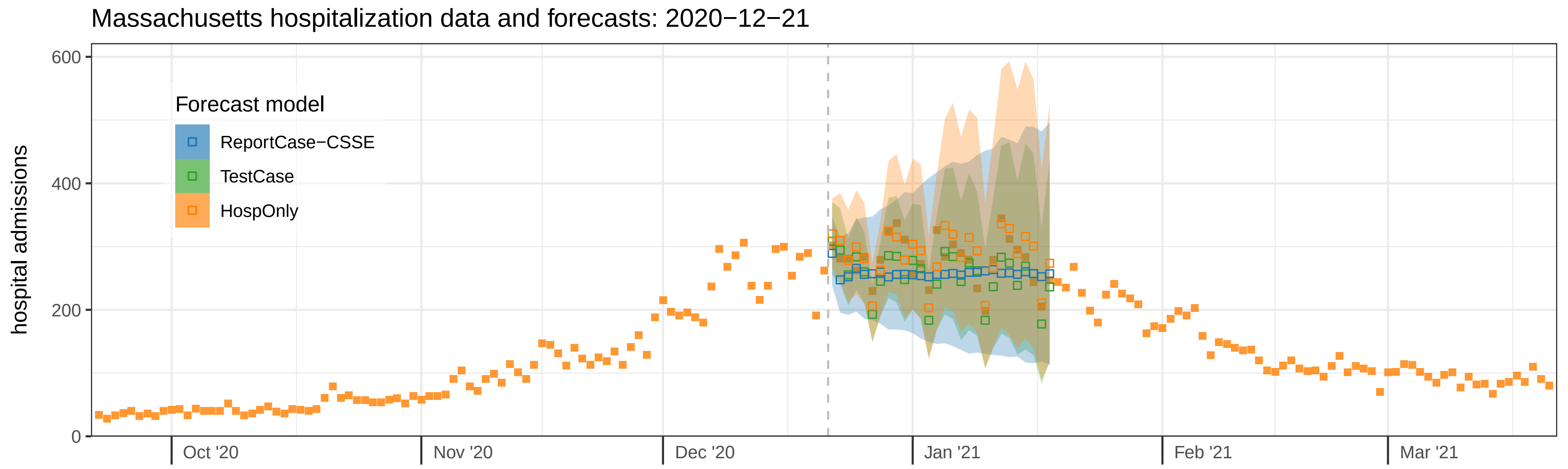

Massachusetts hospitalization data and forecasts: 2020−12−21
Forecast model
ReportCase−CSSE
TestCase
HospOnly
hospital admissions
600
400
200
0
Oct '20
Nov '20
Dec '20
Jan '21
Feb '21
Mar '21

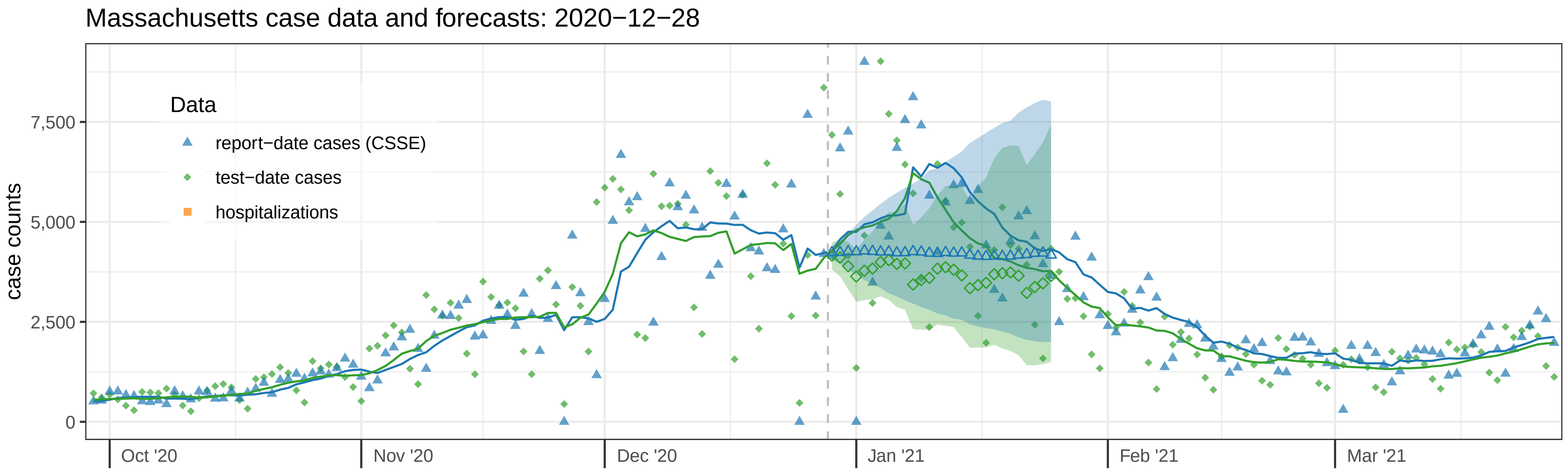

Massachusetts case data and forecasts: 2020−12−28
case counts
7,500
5,000
2,500
0
Data
report−date cases (CSSE)
test−date cases
hospitalizations
Oct '20
Nov '20
Dec '20
Jan '21
Feb '21
Mar '21


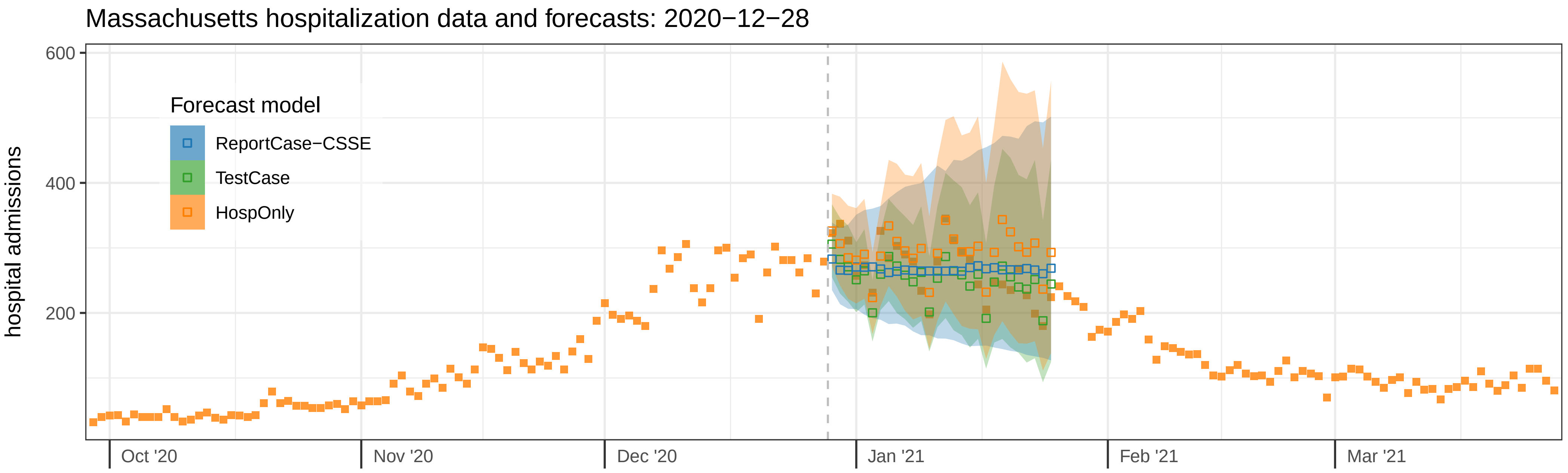

Massachusetts hospitalization data and forecasts: 2020−12−28
hospital admissions
600
400
200
Forecast model
ReportCase−CSSE
TestCase
HospOnly
Oct '20
Nov '20
Dec '20
Jan '21
Feb '21
Mar '21

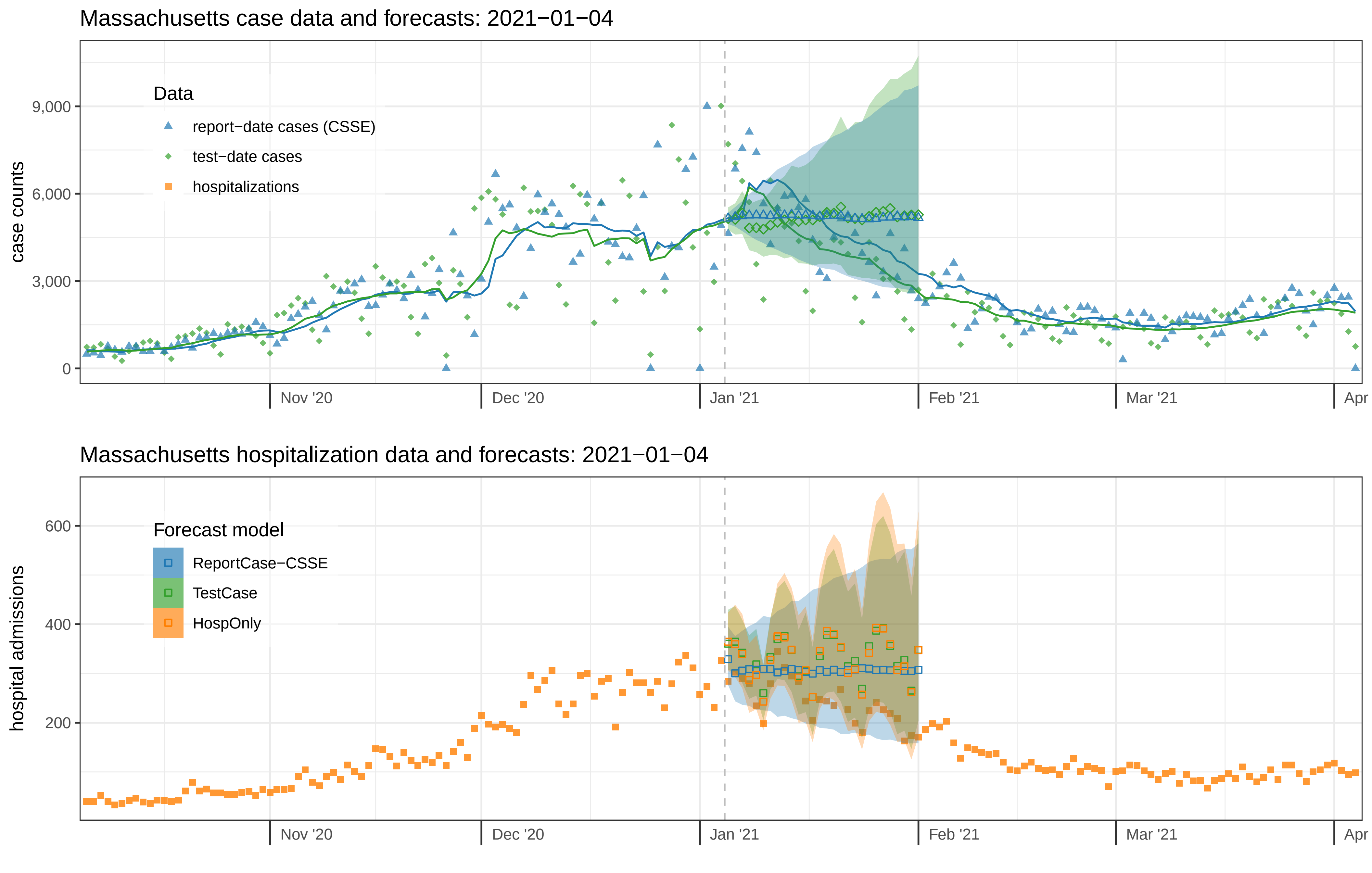

Massachusetts case data and forecasts: 2021−01−04
case counts
9,000
6,000
3,000
0
Data
report−date cases (CSSE)
test−date cases
hospitalizations
Nov '20
Dec '20
Jan '21
Feb '21
Mar '21
Apr
Massachusetts hospitalization data and forecasts: 2021−01−04
hospital admissions
600
400
200
Forecast model
ReportCase−CSSE
TestCase
HospOnly
Nov '20
Dec '20
Jan '21
Feb '21
Mar '21
Apr

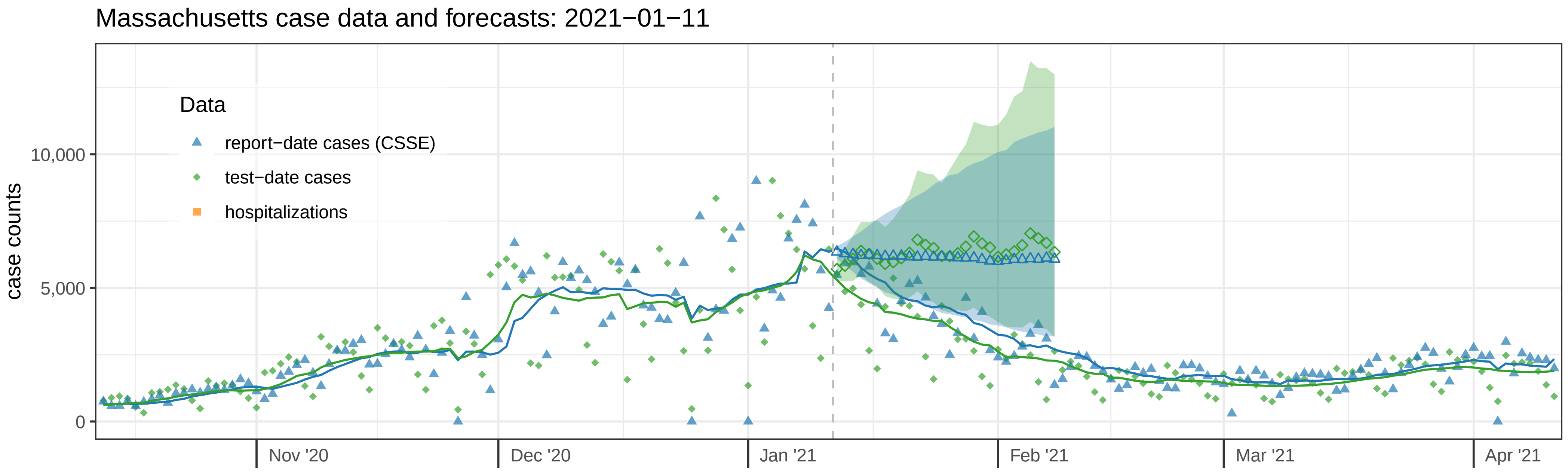

Massachusetts case data and forecasts: 2021−01−11
case counts
10,000
5,000
0
Data
report−date cases (CSSE)
test−date cases
hospitalizations
Nov '20
Dec '20
Jan '21
Feb '21
Mar '21
Apr '21


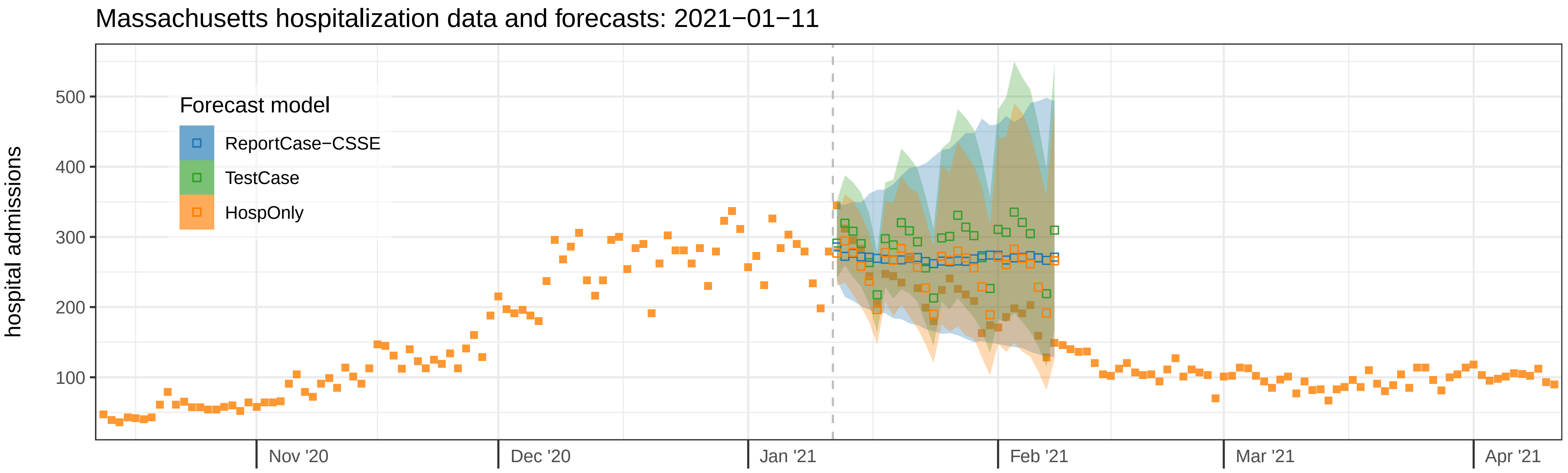

Massachusetts hospitalization data and forecasts: 2021−01−11
hospital admissions
500
400
300
200
100
Forecast model
ReportCase−CSSE
TestCase
HospOnly
Nov '20
Dec '20
Jan '21
Feb '21
Mar '21
Apr '21

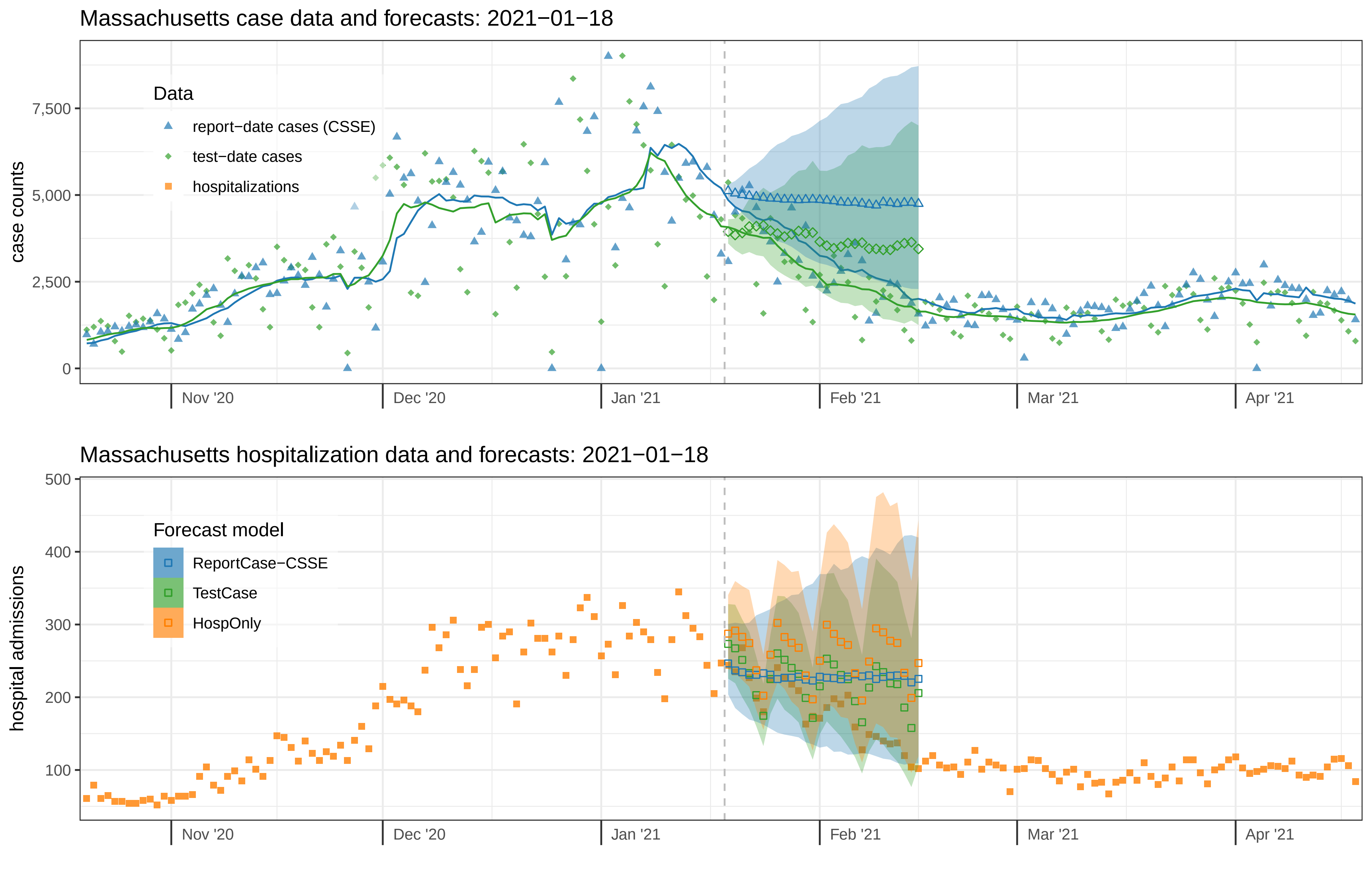
Massachusetts case data and forecasts: 2021−01−18
case counts
7,500
5,000
2,500
0
Data
report−date cases (CSSE)
test−date cases
hospitalizations
Nov '20
Dec '20
Jan '21
Feb '21
Mar '21
Apr '21
Massachusetts hospitalization data and forecasts: 2021−01−18
hospital admissions
500
400
300
200
100
Forecast model
ReportCase−CSSE
TestCase
HospOnly
Nov '20
Dec '20
Jan '21
Feb '21
Mar '21
Apr '21

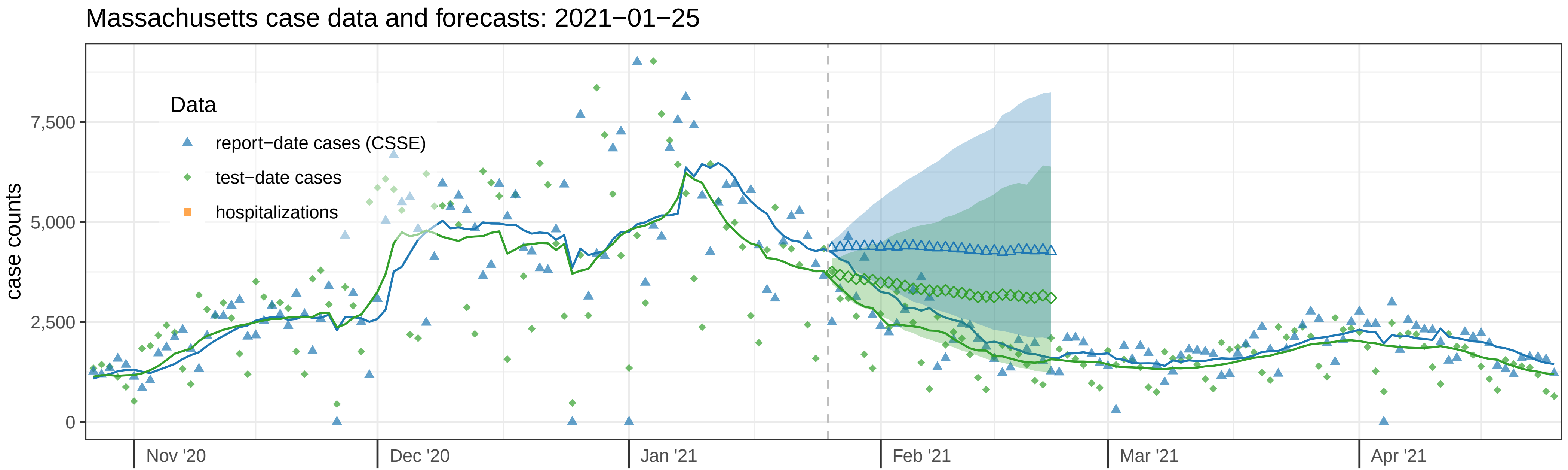

Massachusetts case data and forecasts: 2021−01−25
Data
report−date cases (CSSE)
test−date cases
hospitalizations
case counts
7,500
5,000
2,500
0
Nov '20
Dec '20
Jan '21
Feb '21
Mar '21
Apr '21


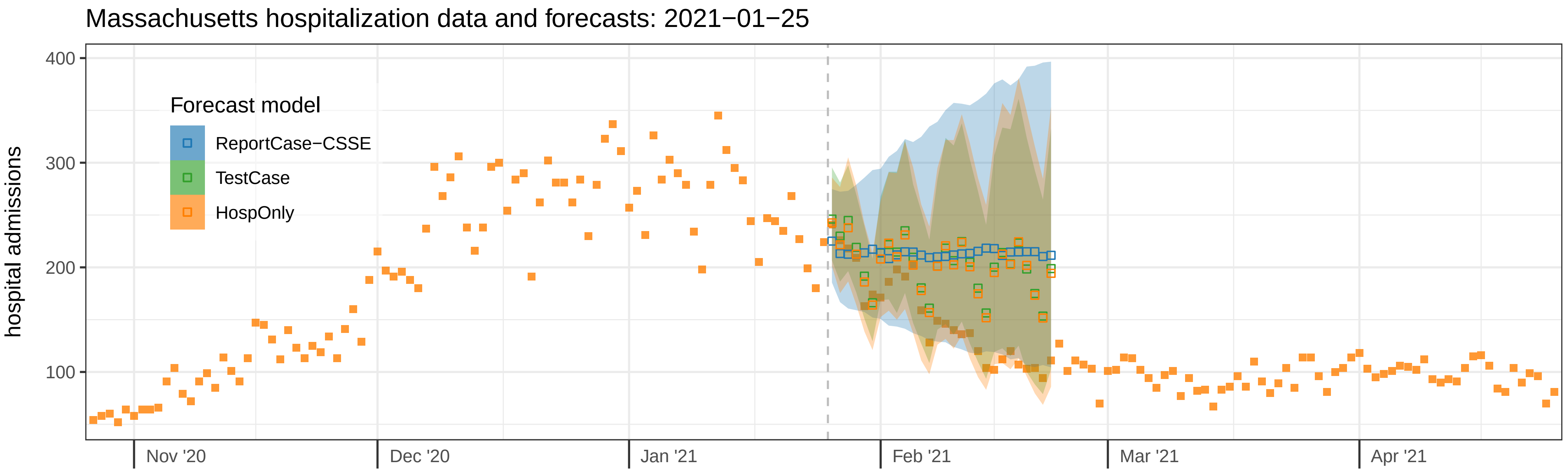

Massachusetts hospitalization data and forecasts: 2021−01−25
Forecast model
ReportCase−CSSE
TestCase
HospOnly
hospital admissions
400
300
200
100
Nov '20
Dec '20
Jan '21
Feb '21
Mar '21
Apr '21

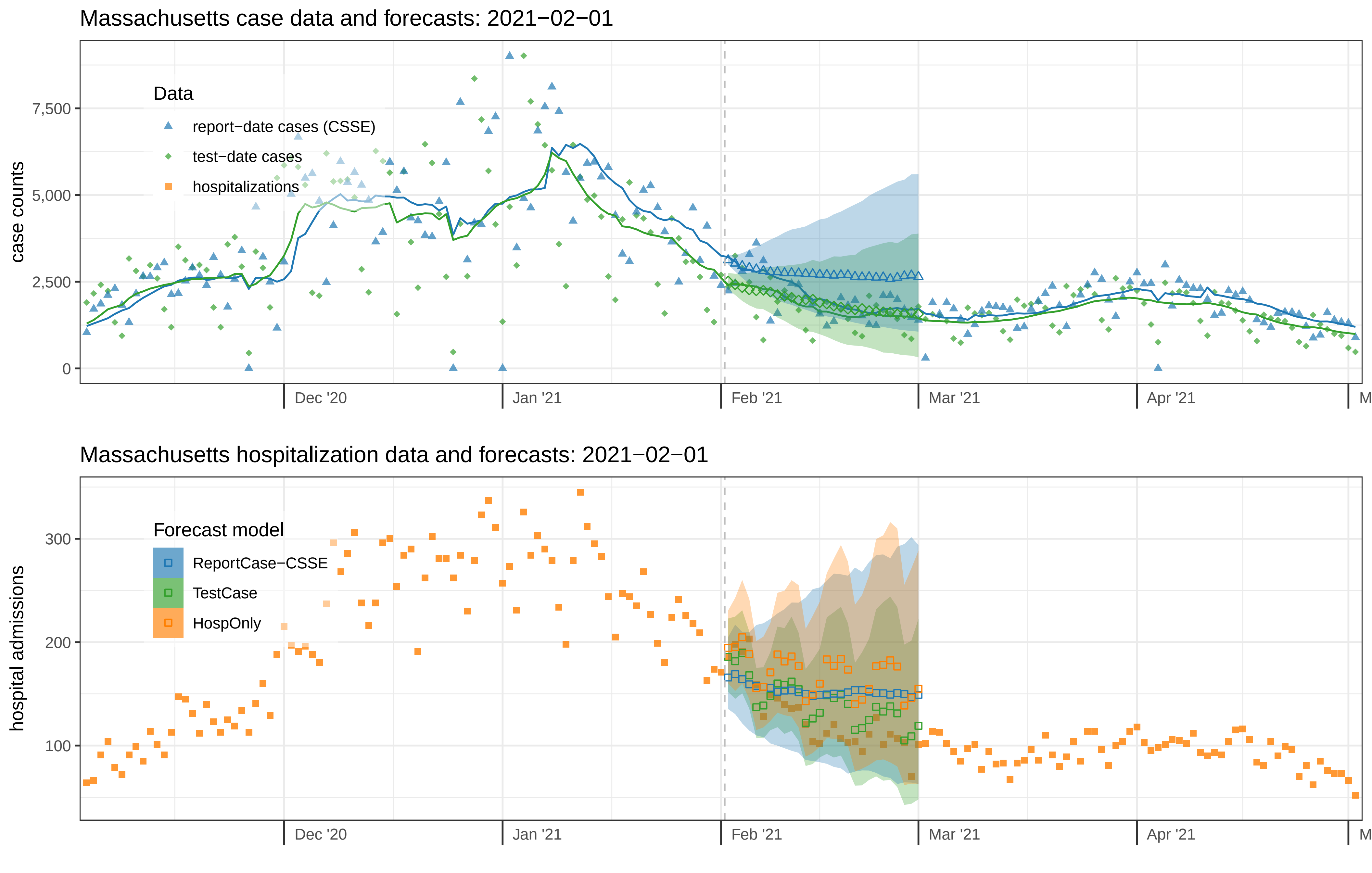
Massachusetts case data and forecasts: 2021−02−01
case counts
7,500
5,000
2,500
0
Data
report−date cases (CSSE)
test−date cases
hospitalizations
Dec '20
Jan '21
Feb '21
Mar '21
Apr '21
Massachusetts hospitalization data and forecasts: 2021−02−01
hospital admissions
300
200
100
Forecast model
ReportCase−CSSE
TestCase
HospOnly
Dec '20
Jan '21
Feb '21
Mar '21
Apr '21

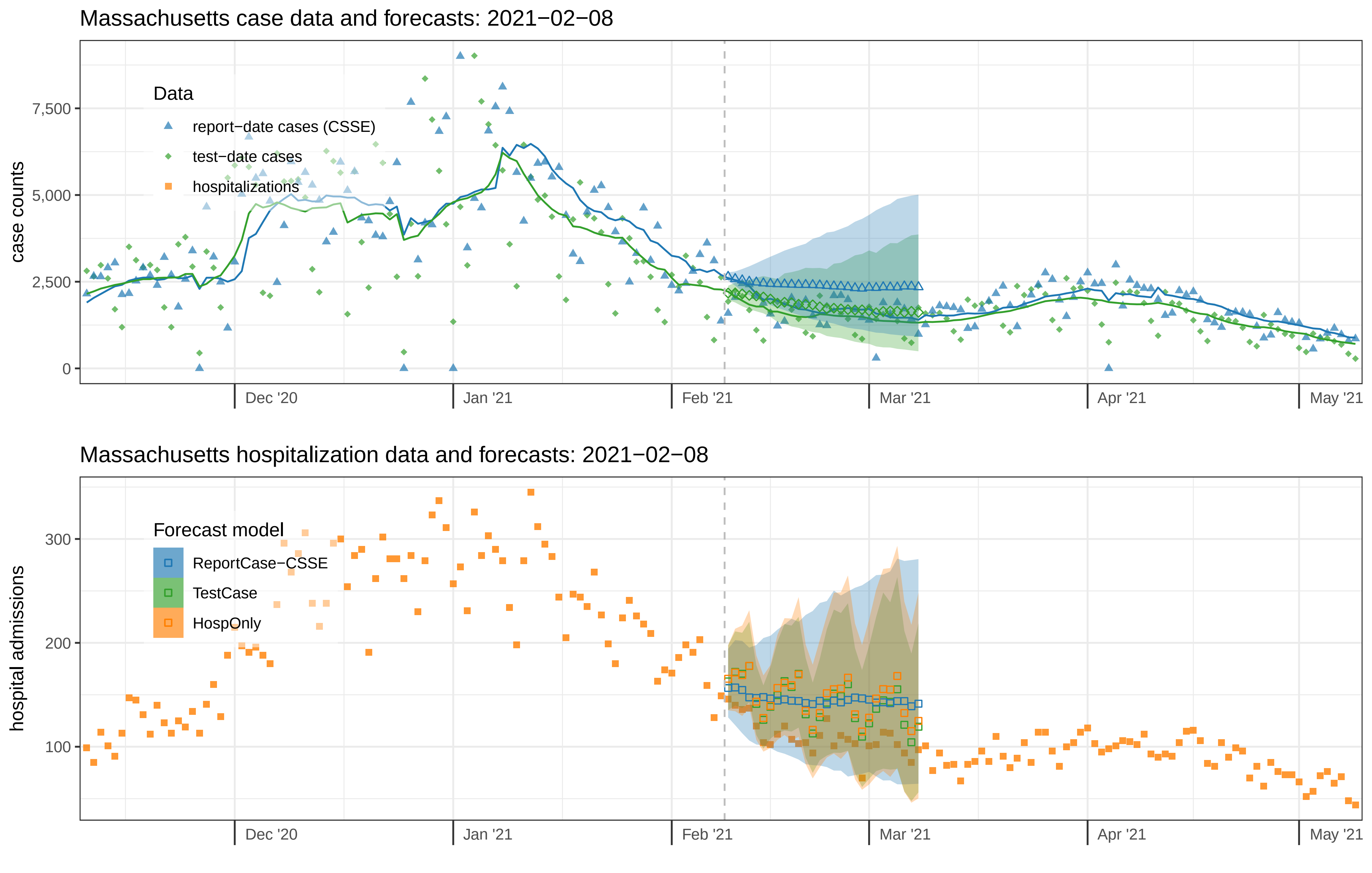

Massachusetts case data and forecasts: 2021−02−08
case counts
0
2,500
5,000
7,500
Data
report−date cases (CSSE)
test−date cases
hospitalizations
Dec '20
Jan '21
Feb '21
Mar '21
Apr '21
May '21
Massachusetts hospitalization data and forecasts: 2021−02−08
hospital admissions
100
200
300
Forecast model
ReportCase−CSSE
TestCase
HospOnly
Dec '20
Jan '21
Feb '21
Mar '21
Apr '21
May '21

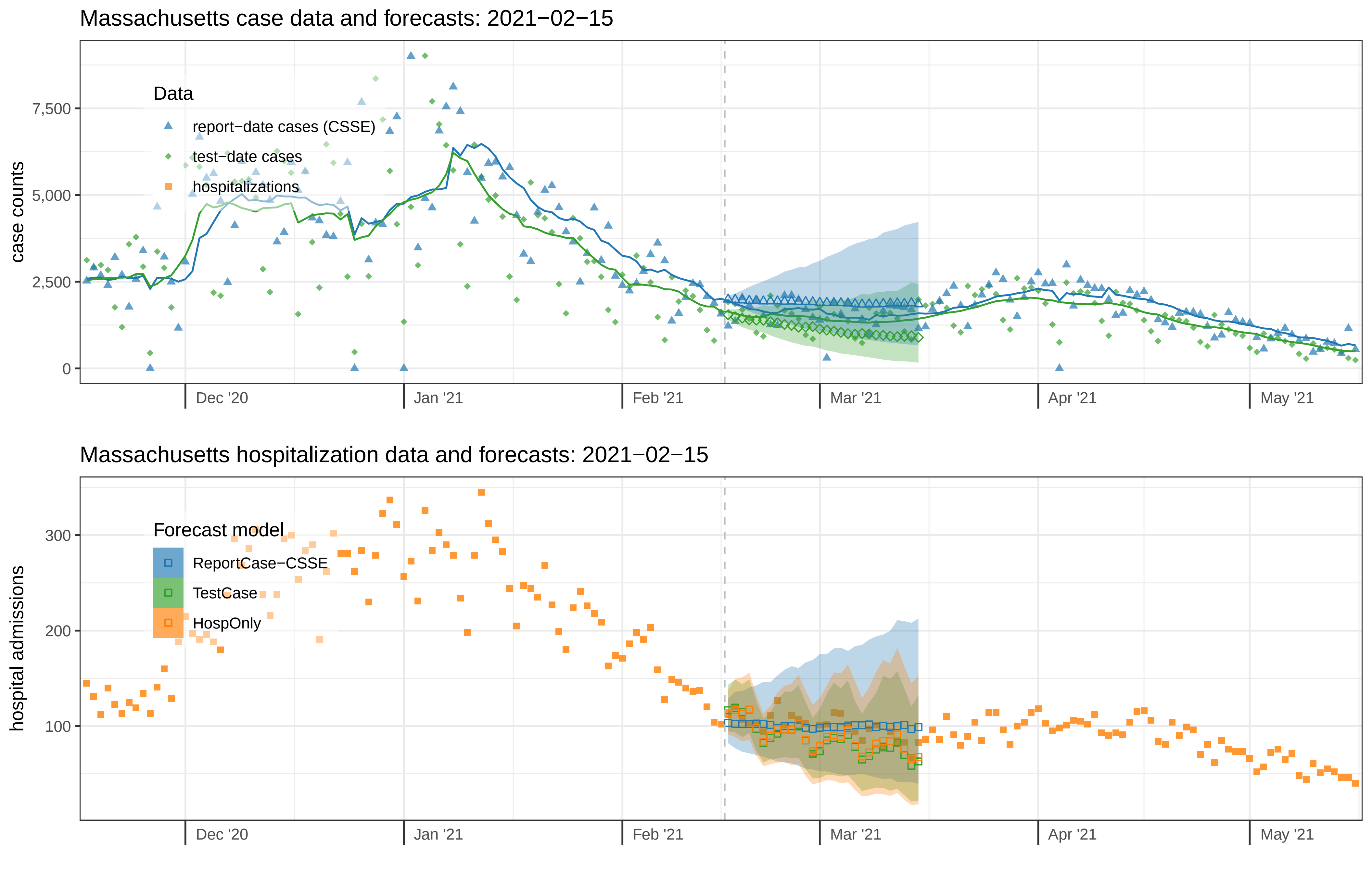

Massachusetts case data and forecasts: 2021−02−15
case counts
7,500
5,000
2,500
0
Data
report−date cases (CSSE)
test−date cases
hospitalizations
Dec '20
Jan '21
Feb '21
Mar '21
Apr '21
May '21
Massachusetts hospitalization data and forecasts: 2021−02−15
hospital admissions
300
200
100
Forecast model
ReportCase−CSSE
TestCase
HospOnly
Dec '20
Jan '21
Feb '21
Mar '21
Apr '21
May '21

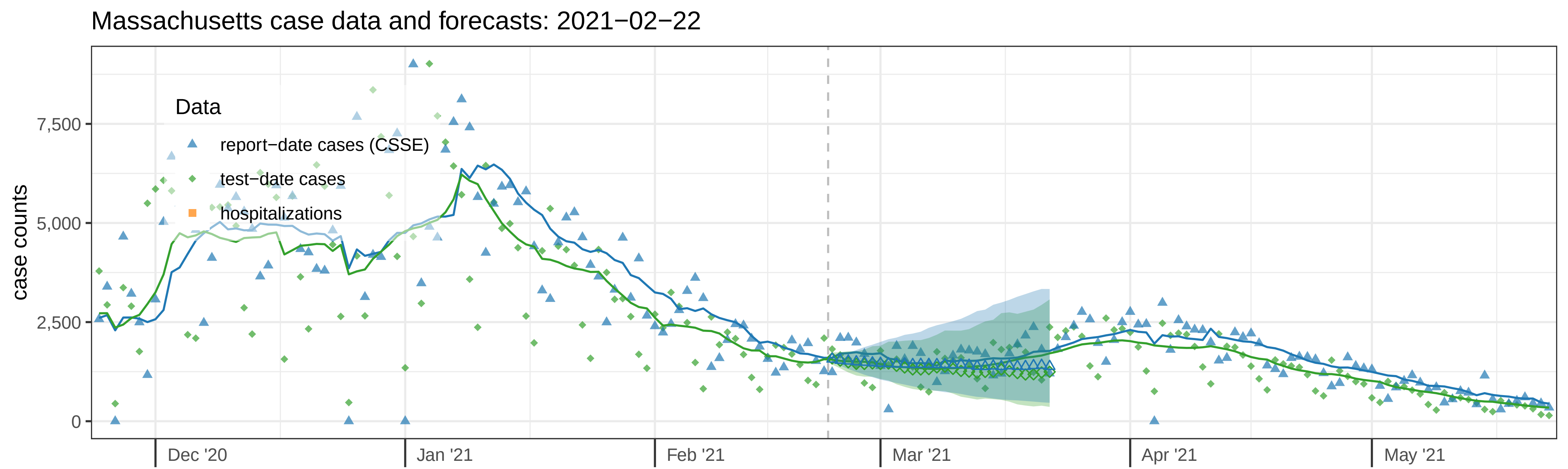

Massachusetts case data and forecasts: 2021−02−22
case counts
7,500
5,000
2,500
0
Data
report−date cases (CSSE)
test−date cases
hospitalizations
Dec '20
Jan '21
Feb '21
Mar '21
Apr '21
May '21


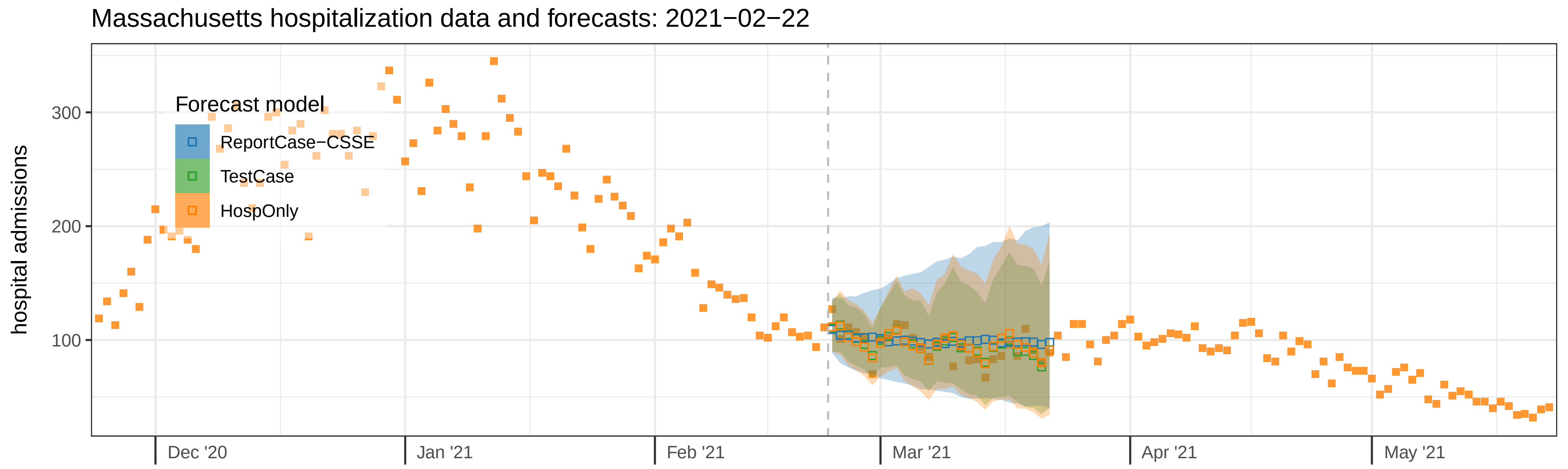

Massachusetts hospitalization data and forecasts: 2021−02−22
hospital admissions
300
200
100
Forecast model
ReportCase−CSSE
TestCase
HospOnly
Dec '20
Jan '21
Feb '21
Mar '21
Apr '21
May '21

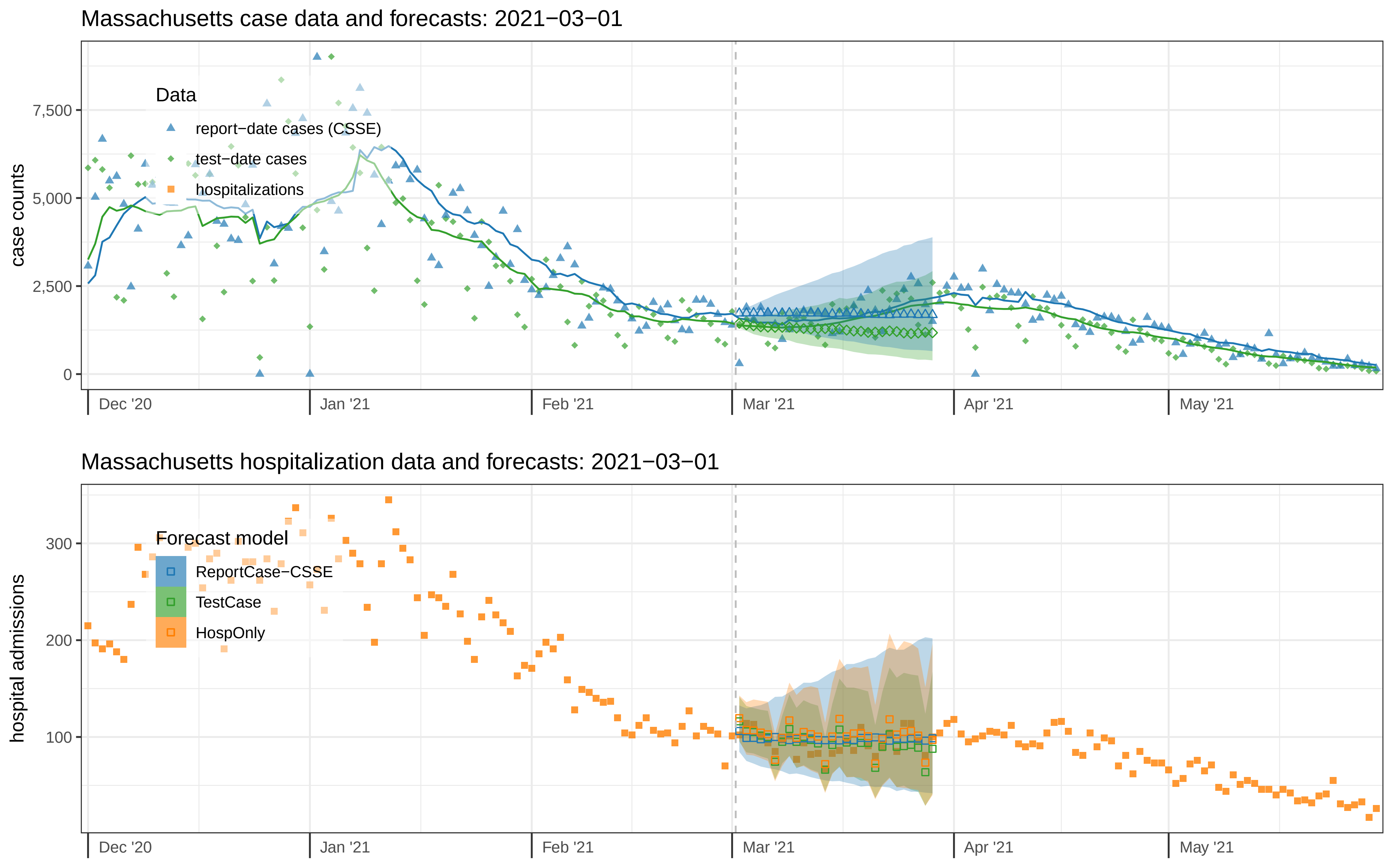

Massachusetts case data and forecasts: 2021−03−01
case counts
0
2,500
5,000
7,500
Data
report−date cases (CSSE)
test−date cases
hospitalizations
Dec '20
Jan '21
Feb '21
Mar '21
Apr '21
May '21
Massachusetts hospitalization data and forecasts: 2021−03−01
hospital admissions
100
200
300
Forecast model
ReportCase−CSSE
TestCase
HospOnly
Dec '20
Jan '21
Feb '21
Mar '21
Apr '21
May '21

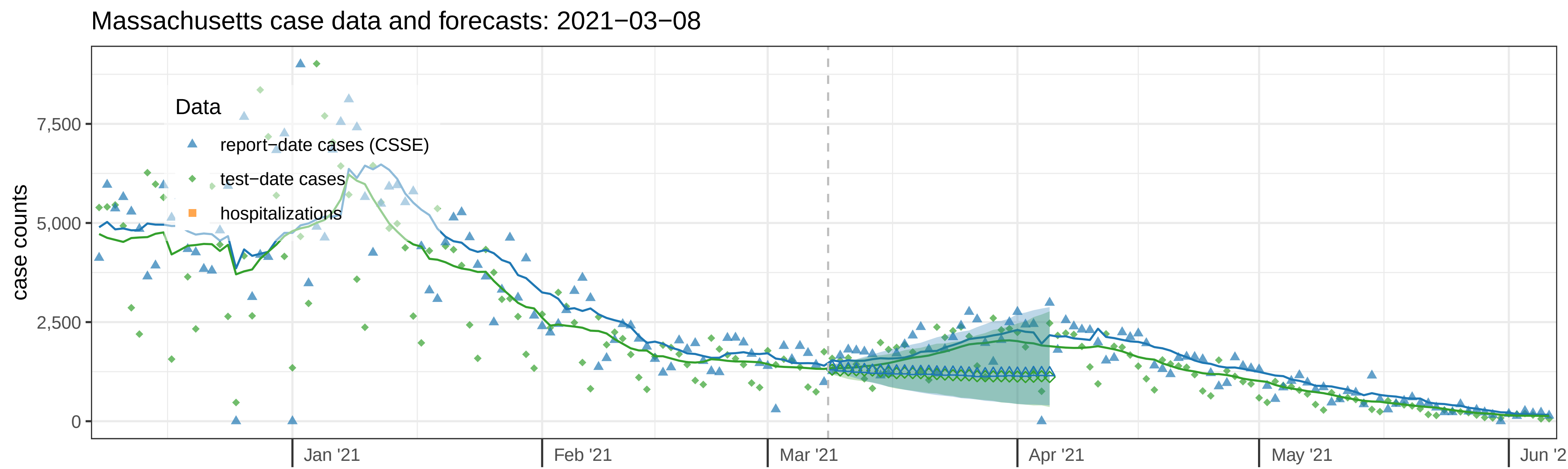

Massachusetts case data and forecasts: 2021−03−08
case counts
7,500
5,000
2,500
0
Data
report−date cases (CSSE)
test−date cases
hospitalizations
Jan '21
Feb '21
Mar '21
Apr '21
May '21
Jun '2


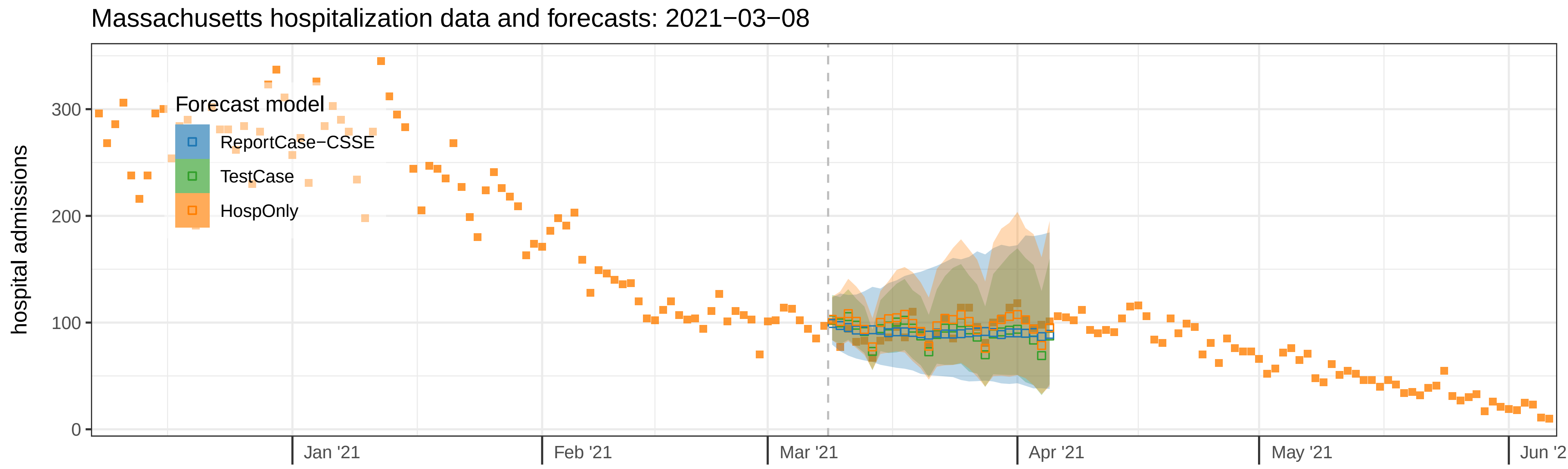

Massachusetts hospitalization data and forecasts: 2021−03−08
hospital admissions
300
200
100
0
Forecast model
ReportCase−CSSE
TestCase
HospOnly
Jan '21
Feb '21
Mar '21
Apr '21
May '21
Jun '2

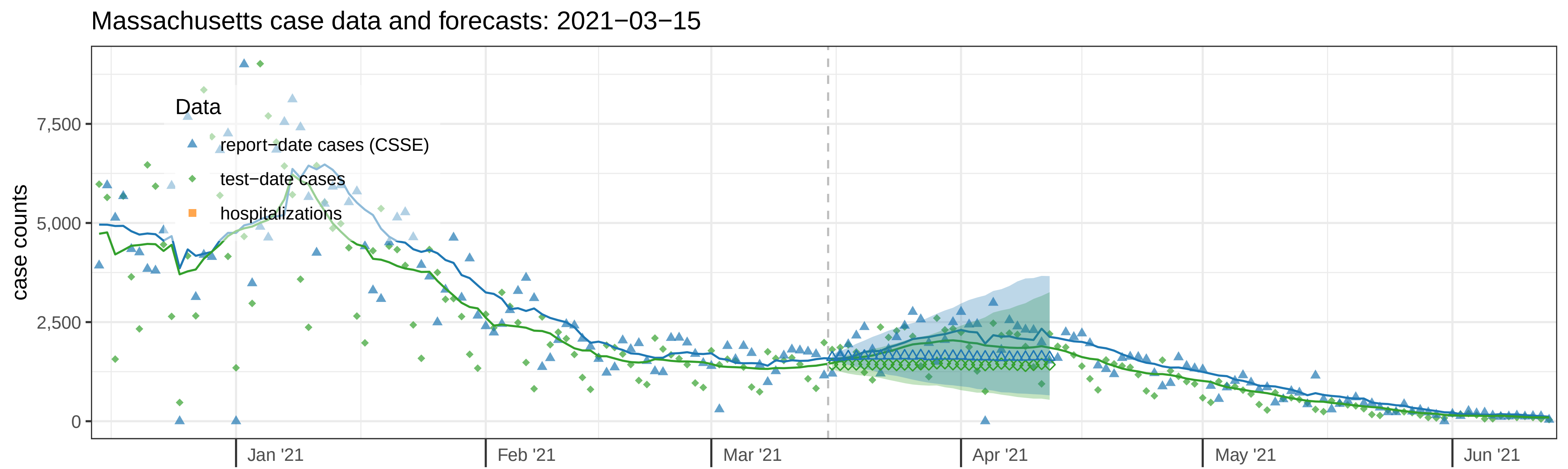
Massachusetts case data and forecasts: 2021−03−15
Data
report−date cases (CSSE)
test−date cases
hospitalizations
case counts
7,500
5,000
2,500
0
Jan '21
Feb '21
Mar '21
Apr '21
May '21
Jun '21

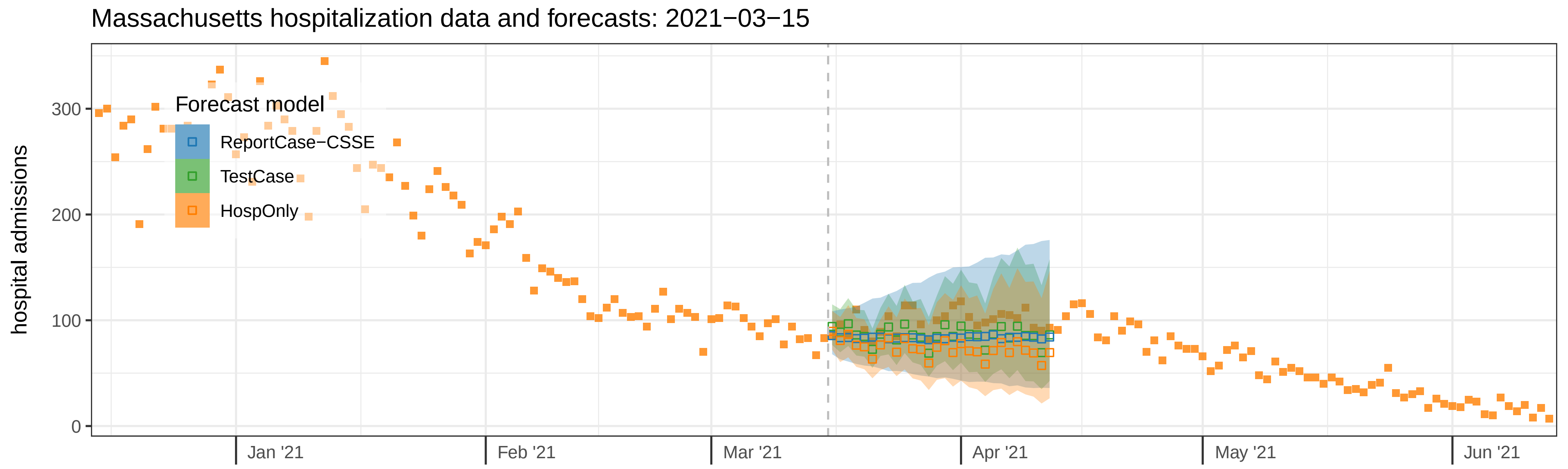
Massachusetts hospitalization data and forecasts: 2021−03−15
Forecast model
ReportCase−CSSE
TestCase
HospOnly
hospital admissions
300
200
100
0
Jan '21
Feb '21
Mar '21
Apr '21
May '21
Jun '21

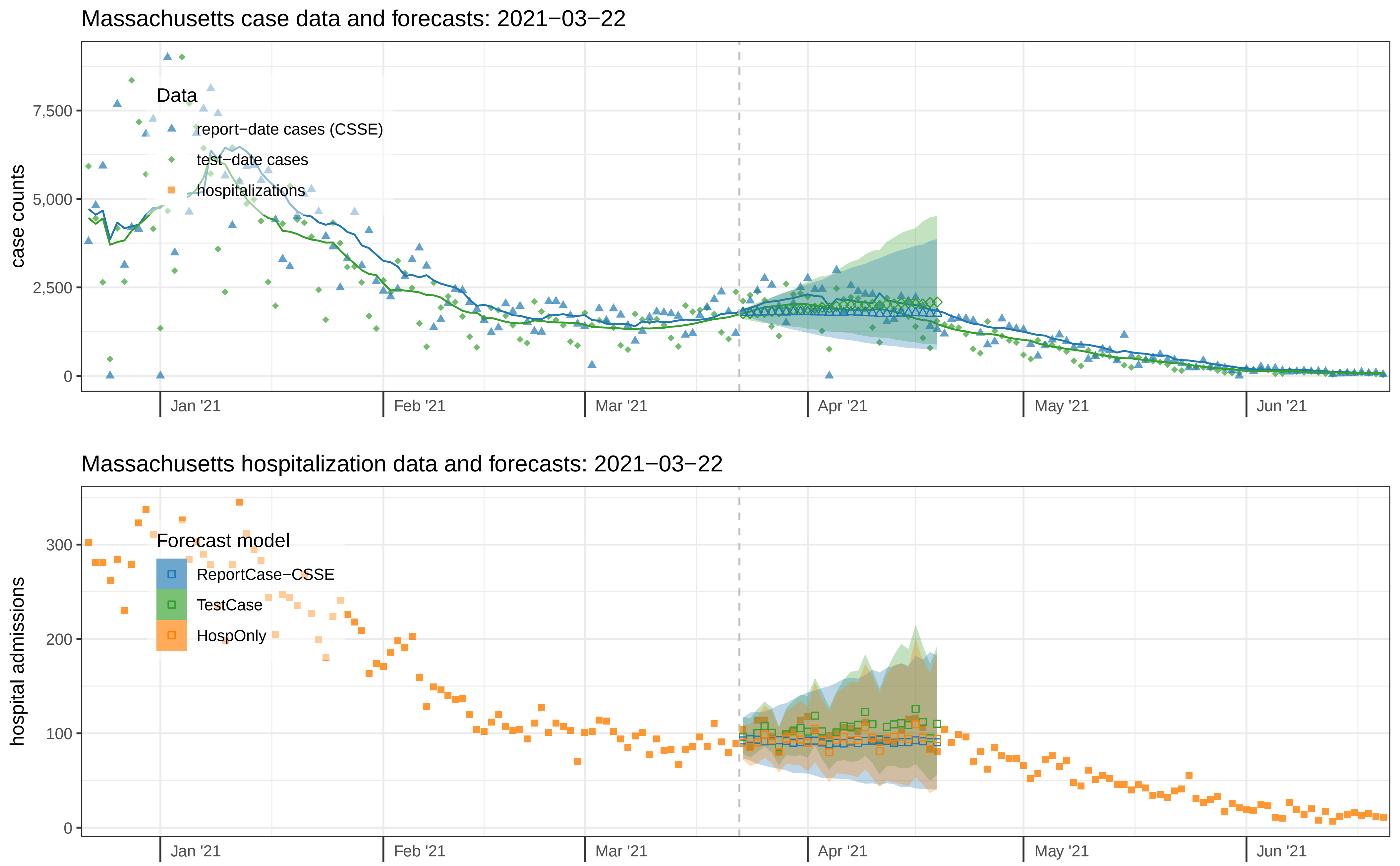

Massachusetts case data and forecasts: 2021−03−22
case counts
0
2,500
5,000
7,500
Data
report−date cases (CSSE)
test−date cases
hospitalizations
Jan '21
Feb '21
Mar '21
Apr '21
May '21
Jun '21
Massachusetts hospitalization data and forecasts: 2021−03−22
hospital admissions
0
100
200
300
Forecast model
ReportCase−CSSE
TestCase
HospOnly
Jan '21
Feb '21
Mar '21
Apr '21
May '21
Jun '21

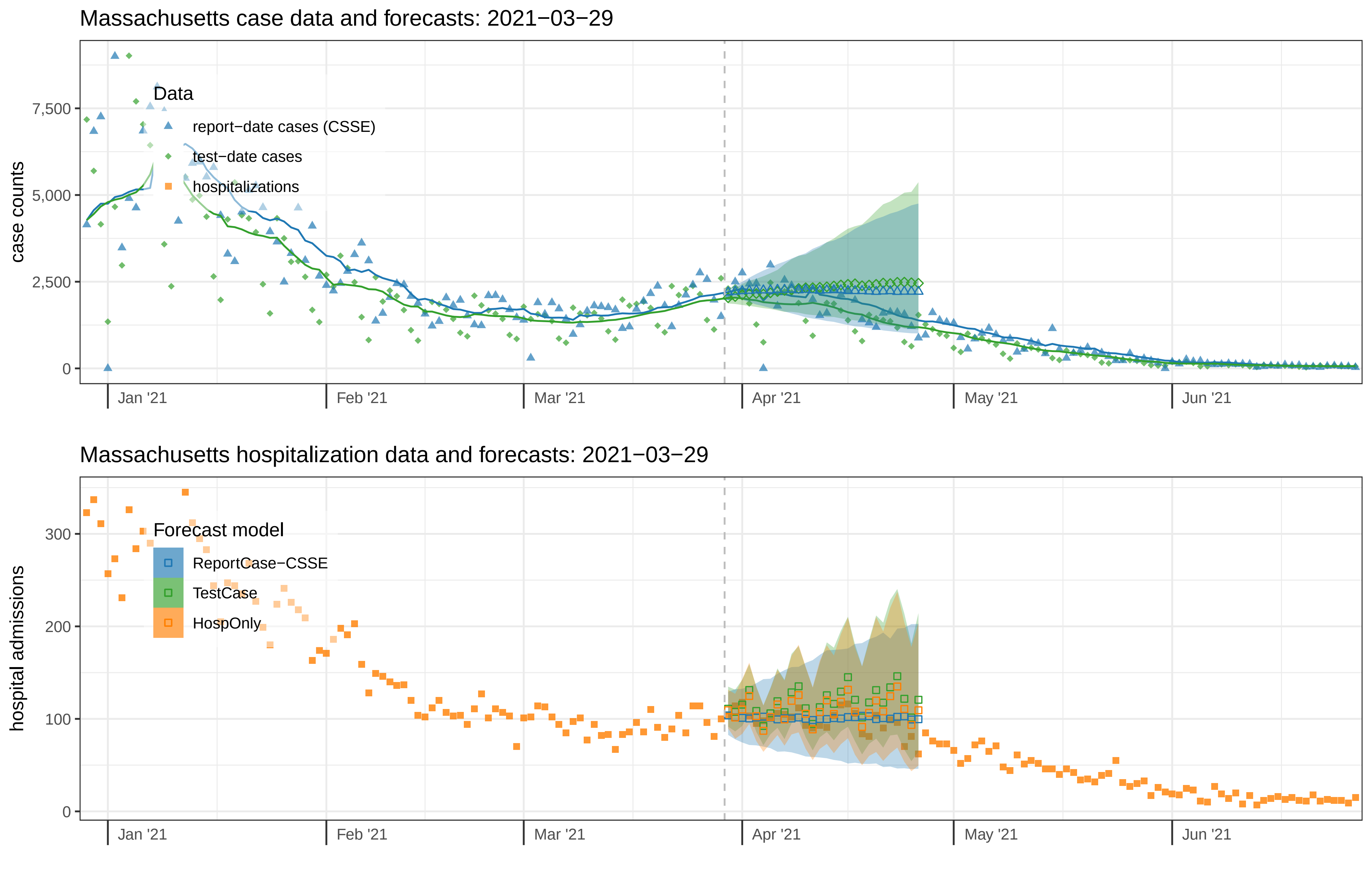

Massachusetts case data and forecasts: 2021−03−29
case counts
0
2,500
5,000
7,500
Data
report−date cases (CSSE)
test−date cases
hospitalizations
Jan '21
Feb '21
Mar '21
Apr '21
May '21
Jun '21
Massachusetts hospitalization data and forecasts: 2021−03−29
hospital admissions
0
100
200
300
Forecast model
ReportCase−CSSE
TestCase
HospOnly
Jan '21
Feb '21
Mar '21
Apr '21
May '21
Jun '21

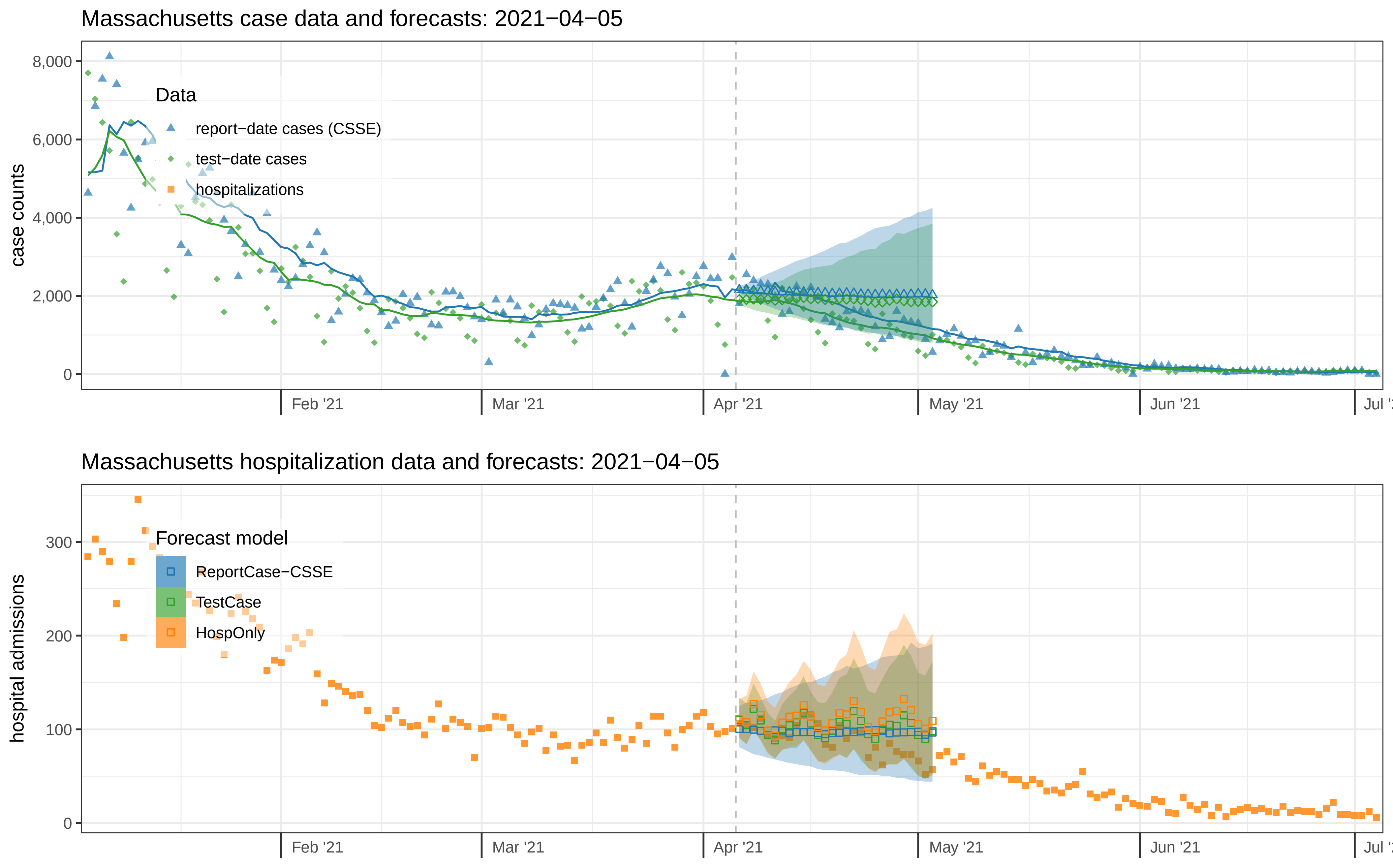
Massachusetts case data and forecasts: 2021−04−05
case counts
8,000
6,000
4,000
2,000
0
Data
report−date cases (CSSE)
test−date cases
hospitalizations
Feb '21
Mar '21
Apr '21
May '21
Jun '21
Massachusetts hospitalization data and forecasts: 2021−04−05
hospital admissions
300
200
100
0
Forecast model
ReportCase−CSSE
TestCase
HospOnly
Feb '21
Mar '21
Apr '21
May '21
Jun '21

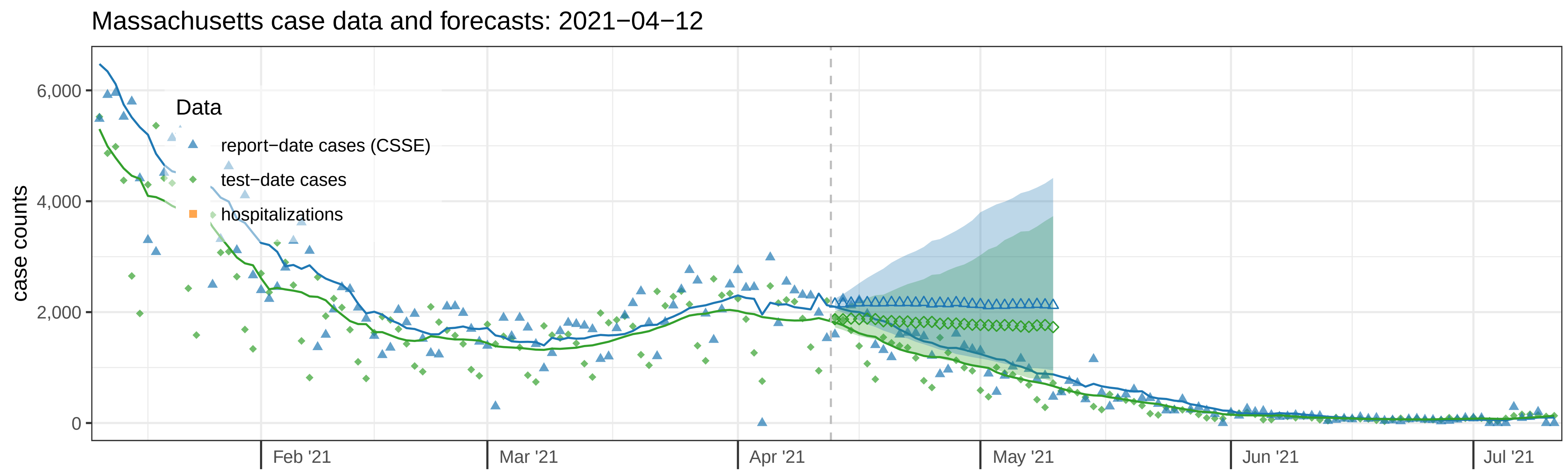
Massachusetts case data and forecasts: 2021−04−12
case counts
6,000
4,000
2,000
0
Data
report−date cases (CSSE)
test−date cases
hospitalizations
Feb '21
Mar '21
Apr '21
May '21
Jun '21
Jul '21

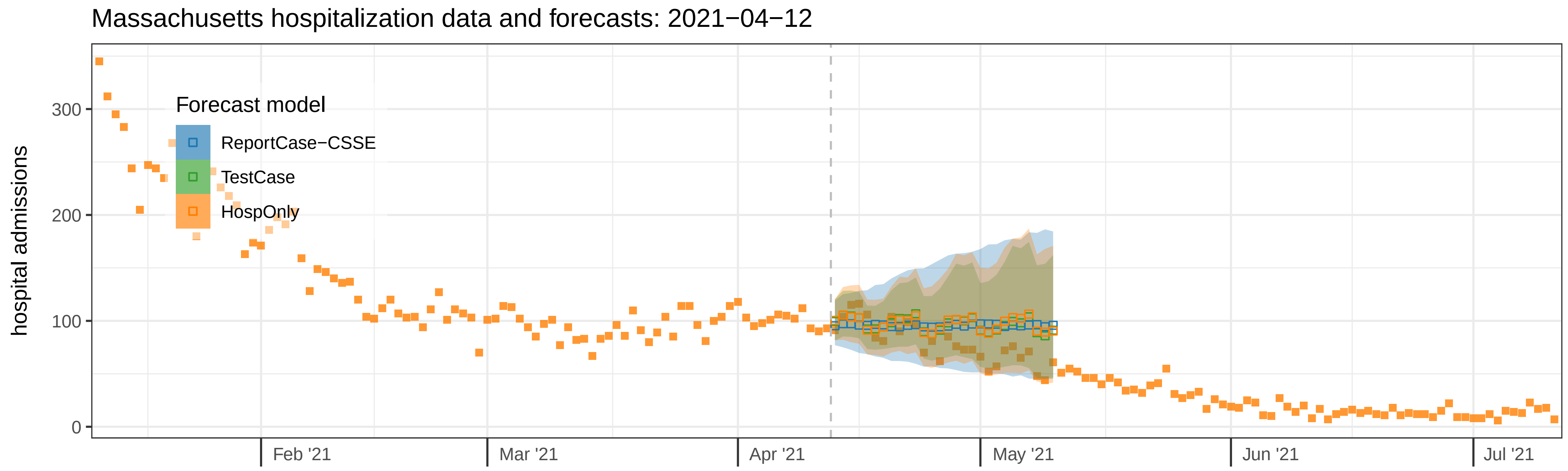
Massachusetts hospitalization data and forecasts: 2021−04−12
hospital admissions
300
200
100
0
Forecast model
ReportCase−CSSE
TestCase
HospOnly
Feb '21
Mar '21
Apr '21
May '21
Jun '21
Jul '21

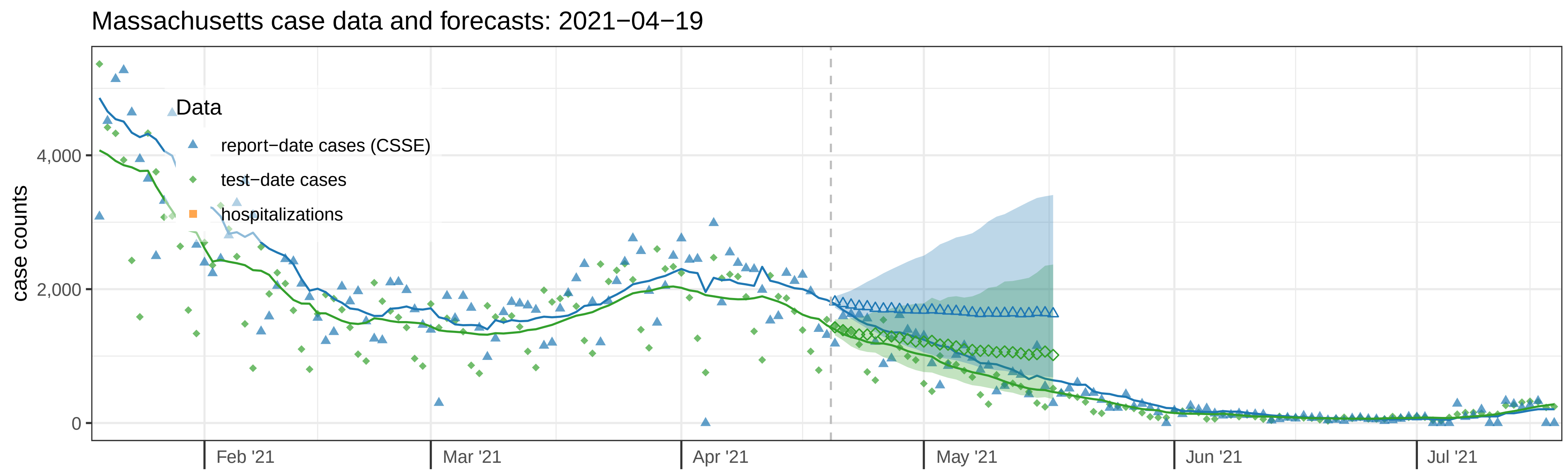

Massachusetts case data and forecasts: 2021−04−19
case counts
4,000
2,000
0
Data
report−date cases (CSSE)
test−date cases
hospitalizations
Feb '21
Mar '21
Apr '21
May '21
Jun '21
Jul '21


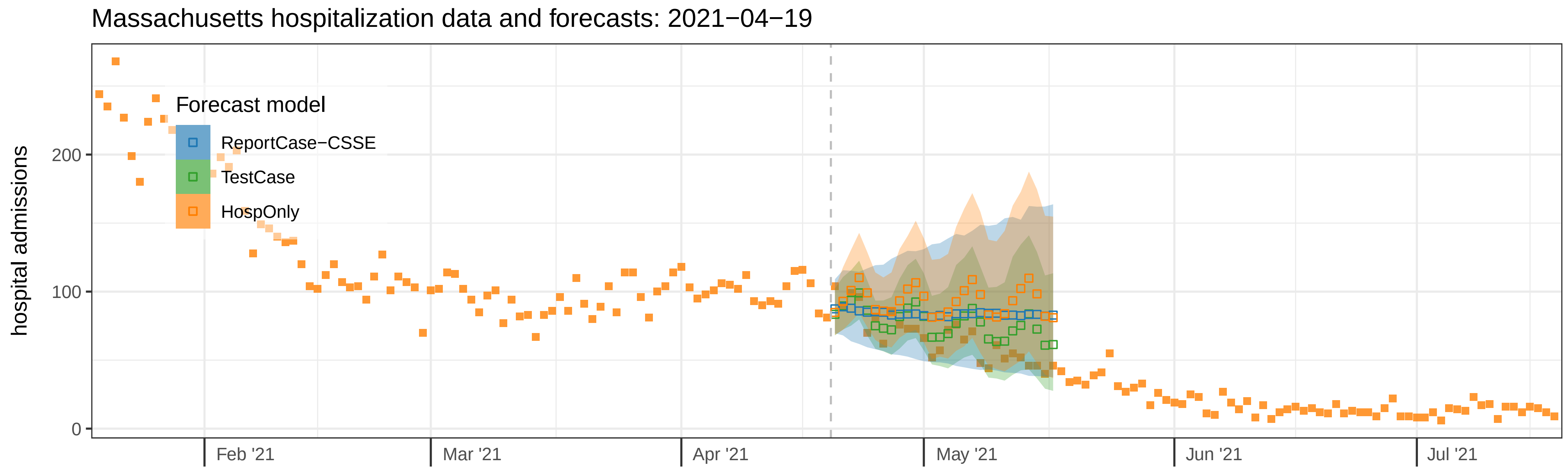

Massachusetts hospitalization data and forecasts: 2021−04−19
hospital admissions
200
100
0
Forecast model
ReportCase−CSSE
TestCase
HospOnly
Feb '21
Mar '21
Apr '21
May '21
Jun '21
Jul '21

Massachusetts case data and forecasts: 2021−04−26

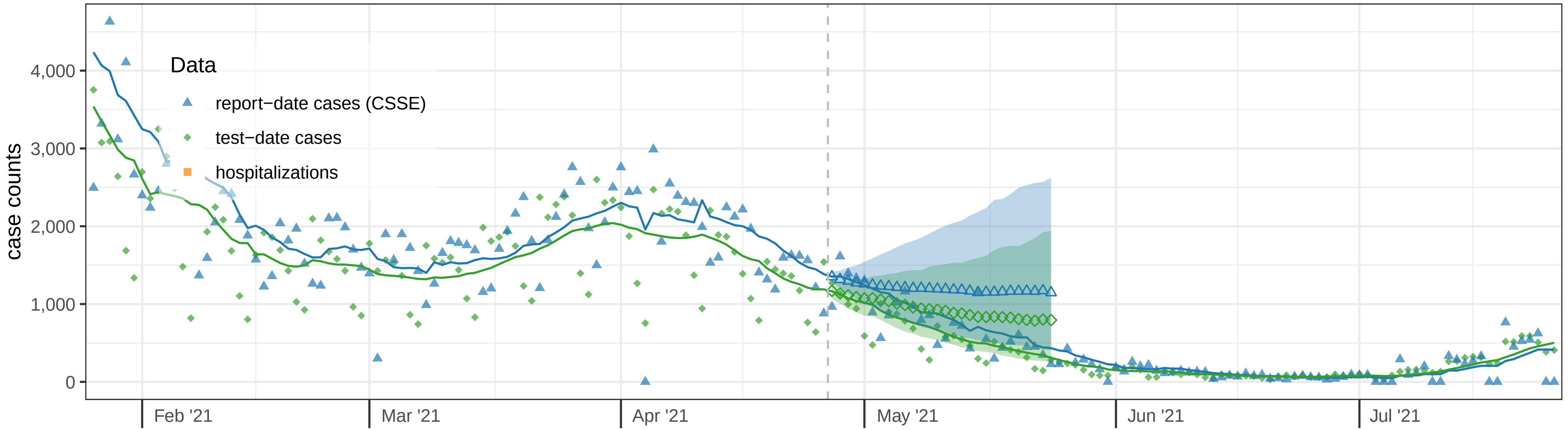


Massachusetts hospitalization data and forecasts: 2021−04−26

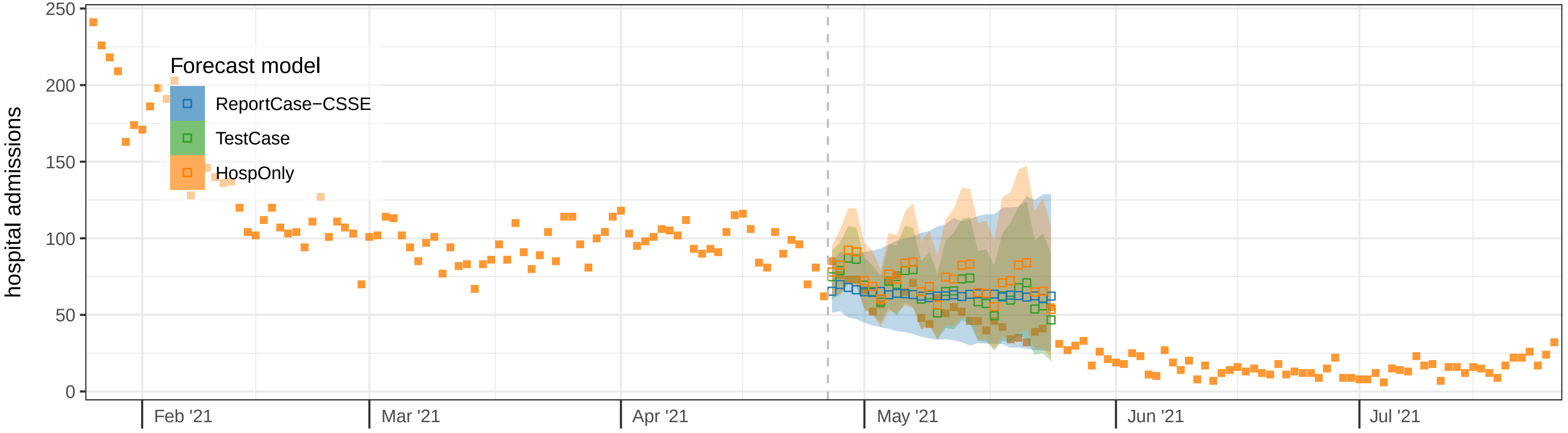

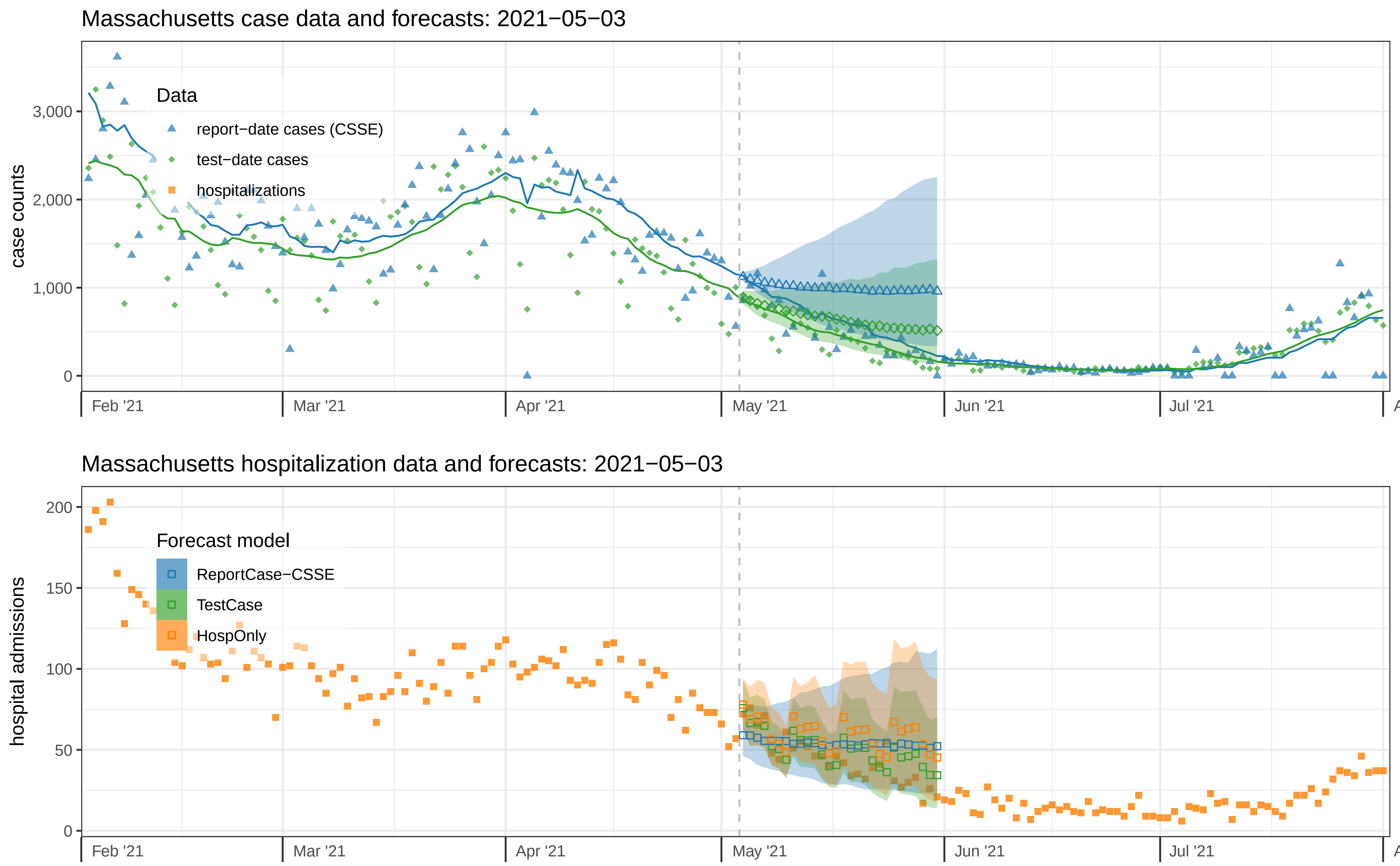

Massachusetts case data and forecasts: 2021−05−03
case counts
3,000
2,000
1,000
0
Data
report−date cases (CSSE)
test−date cases
hospitalizations
Feb '21
Mar '21
Apr '21
May '21
Jun '21
Jul '21
Massachusetts hospitalization data and forecasts: 2021−05−03
hospital admissions
200
150
100
50
0
Forecast model
ReportCase−CSSE
TestCase
HospOnly
Feb '21
Mar '21
Apr '21
May '21
Jun '21
Jul '21

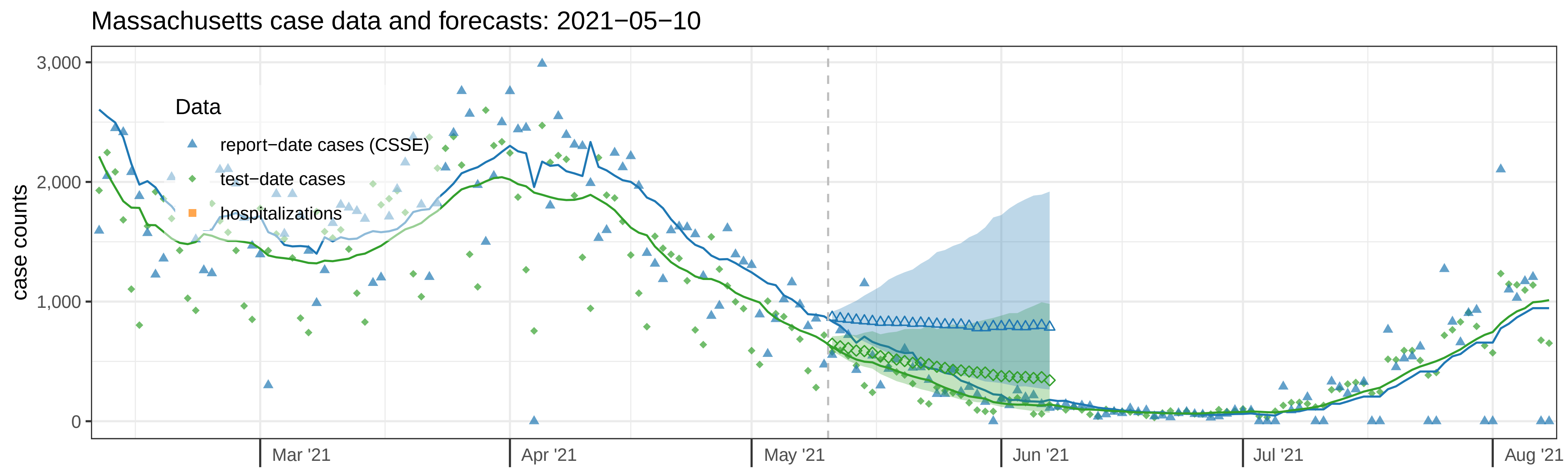

Massachusetts case data and forecasts: 2021−05−10
case counts
3,000
2,000
1,000
0
Data
report−date cases (CSSE)
test−date cases
hospitalizations
Mar '21
Apr '21
May '21
Jun '21
Jul '21
Aug '21


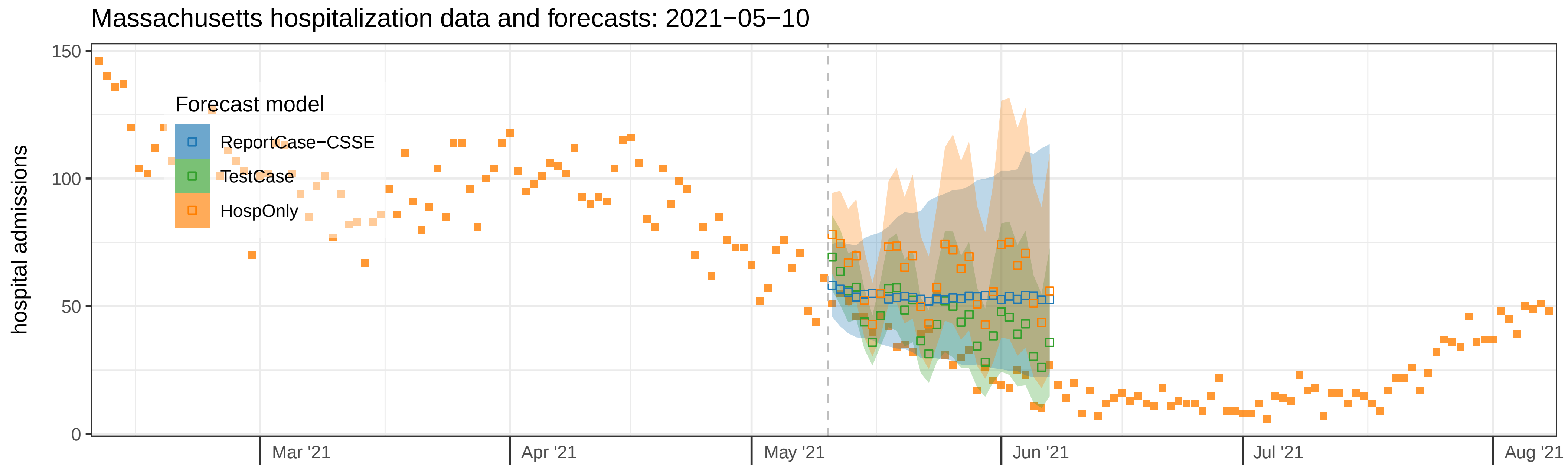

Massachusetts hospitalization data and forecasts: 2021−05−10
hospital admissions
150
100
50
0
Forecast model
ReportCase−CSSE
TestCase
HospOnly
Mar '21
Apr '21
May '21
Jun '21
Jul '21
Aug '21

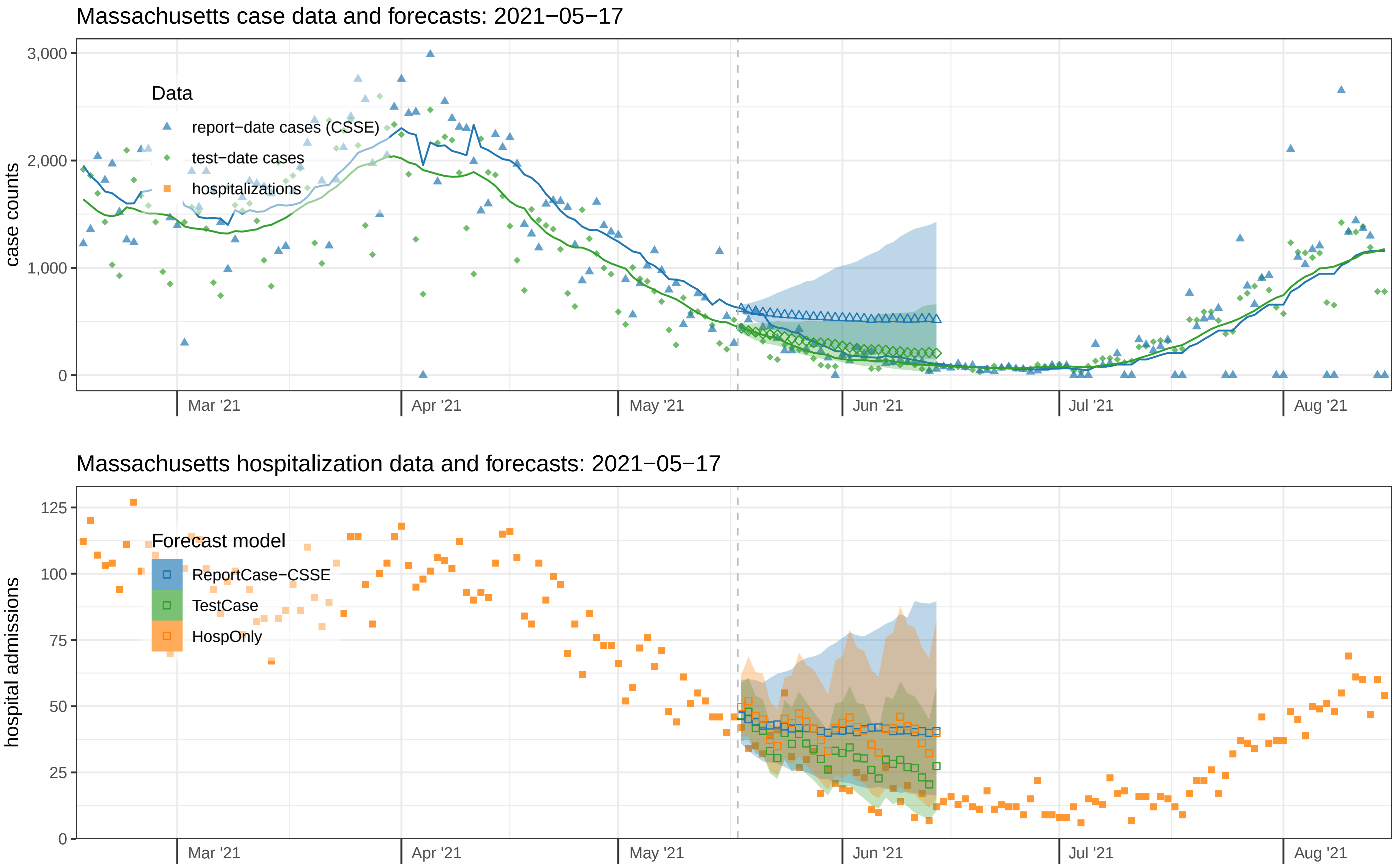

Massachusetts case data and forecasts: 2021−05−17
case counts
3,000
2,000
1,000
0
Data
report−date cases (CSSE)
test−date cases
hospitalizations
Mar '21
Apr '21
May '21
Jun '21
Jul '21
Aug '21
Massachusetts hospitalization data and forecasts: 2021−05−17
hospital admissions
125
100
75
50
25
0
Forecast model
ReportCase−CSSE
TestCase
HospOnly
Mar '21
Apr '21
May '21
Jun '21
Jul '21
Aug '21

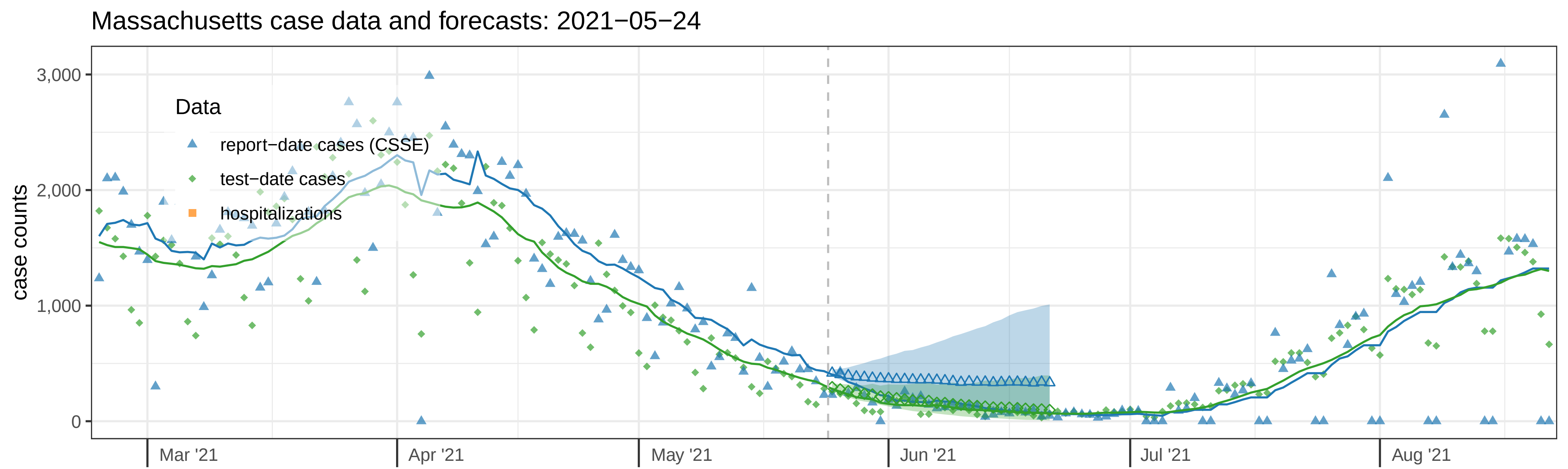
Massachusetts case data and forecasts: 2021−05−24
case counts
3,000
2,000
1,000
0
Data
report−date cases (CSSE)
test−date cases
hospitalizations
Mar '21
Apr '21
May '21
Jun '21
Jul '21
Aug '21

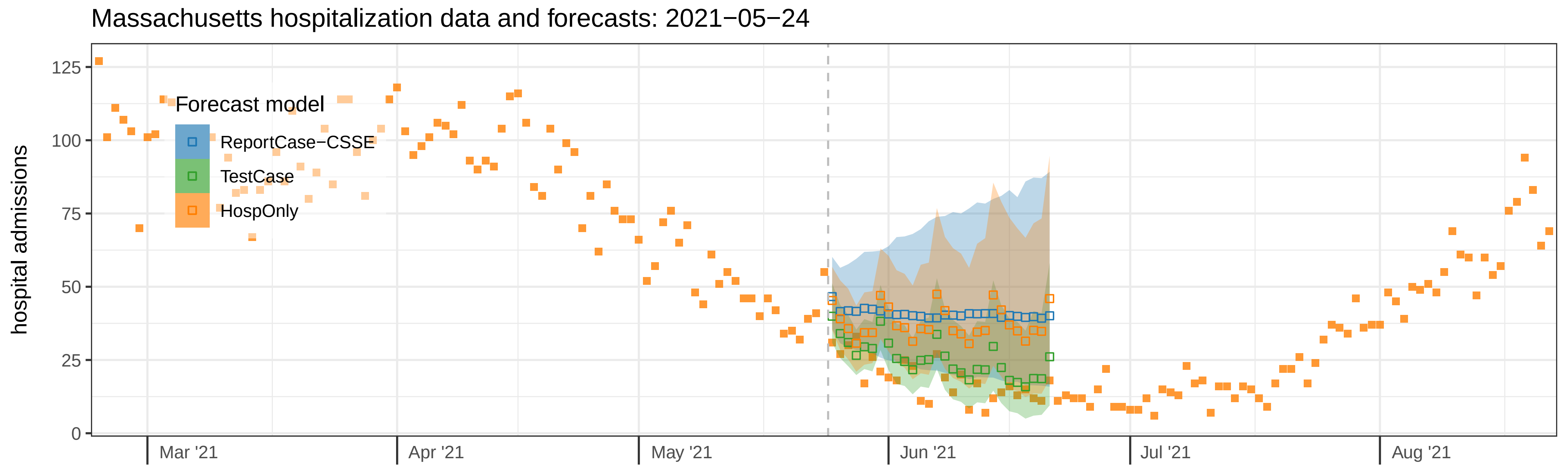
Massachusetts hospitalization data and forecasts: 2021−05−24
hospital admissions
125
100
75
50
25
0
Forecast model
ReportCase−CSSE
TestCase
HospOnly
Mar '21
Apr '21
May '21
Jun '21
Jul '21
Aug '21

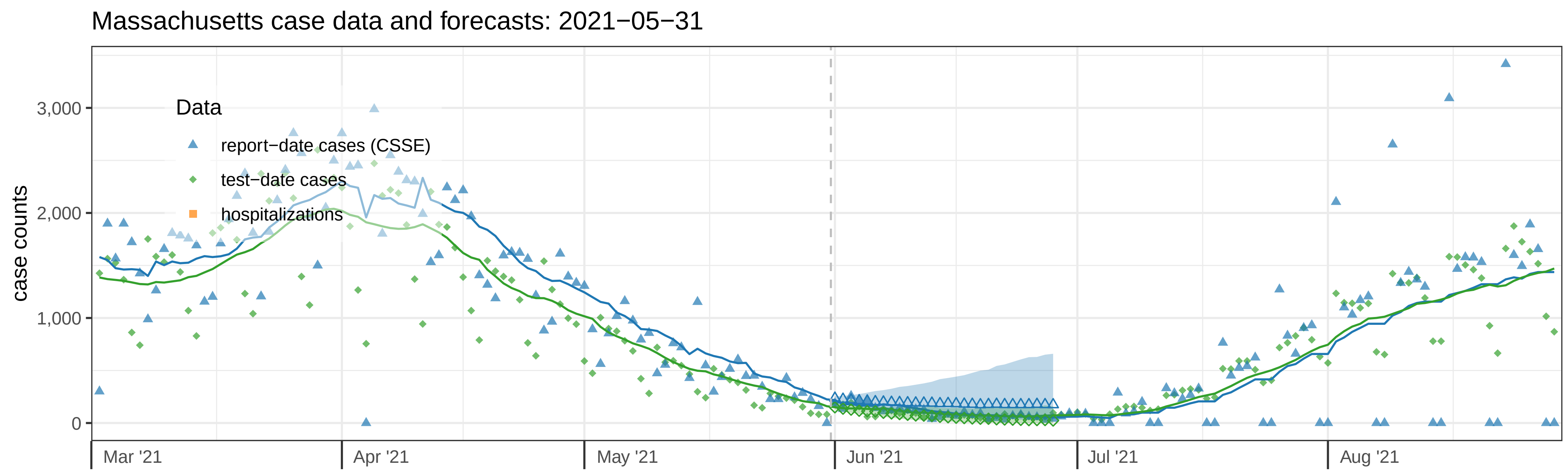

Massachusetts case data and forecasts: 2021−05−31
case counts
3,000
2,000
1,000
0
Data
report−date cases (CSSE)
test−date cases
hospitalizations
Mar '21
Apr '21
May '21
Jun '21
Jul '21
Aug '21


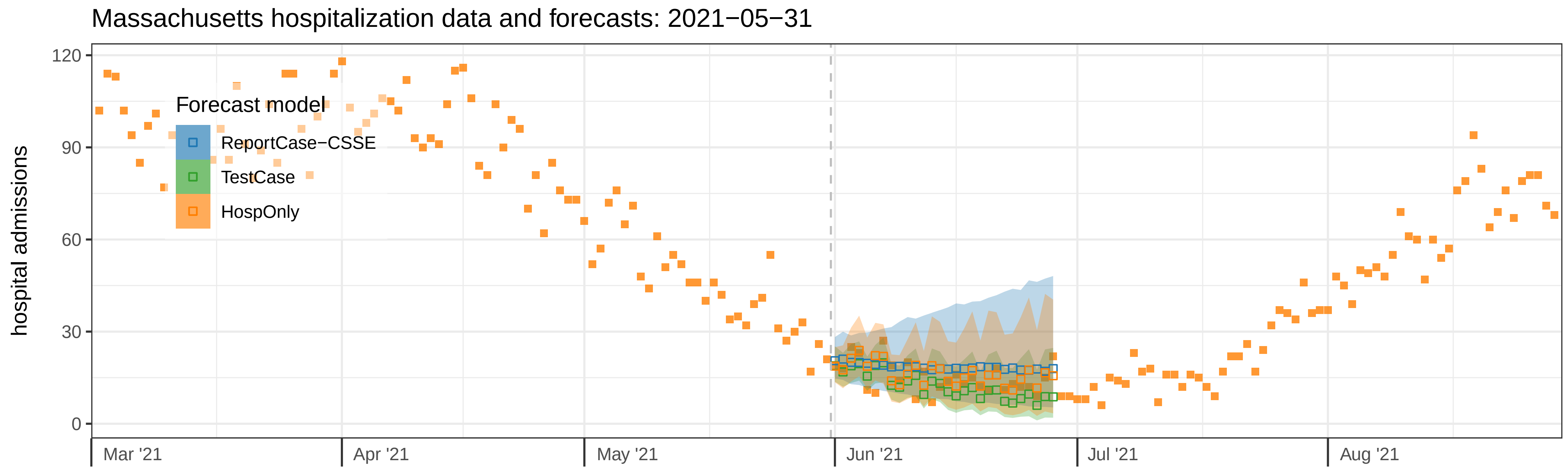

Massachusetts hospitalization data and forecasts: 2021−05−31
hospital admissions
120
90
60
30
0
Forecast model
ReportCase−CSSE
TestCase
HospOnly
Mar '21
Apr '21
May '21
Jun '21
Jul '21
Aug '21

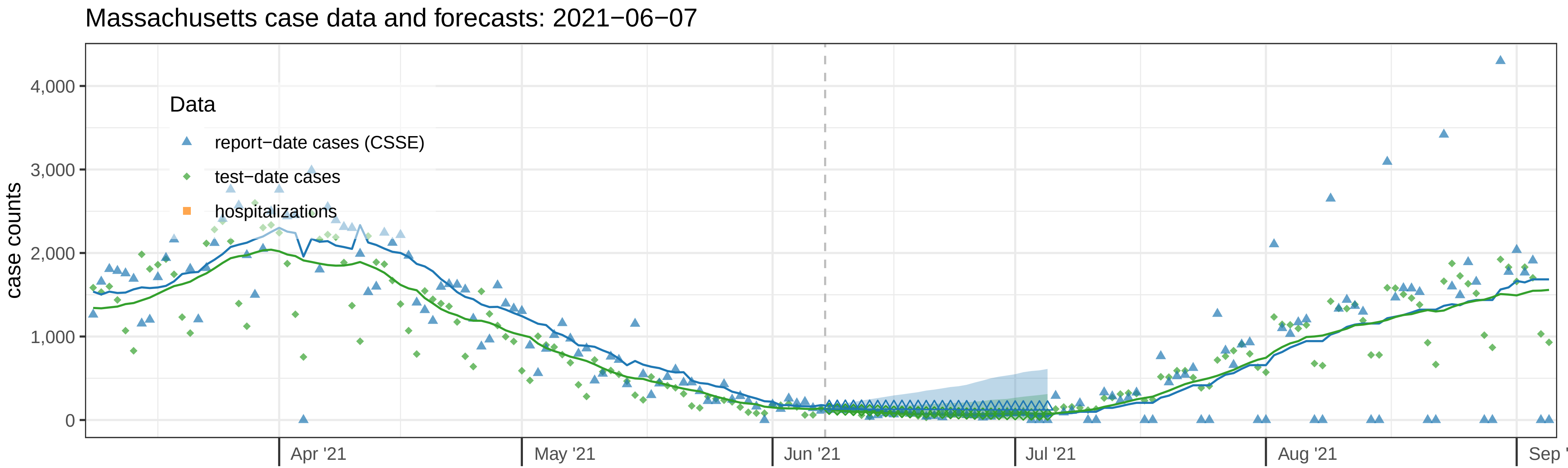
Massachusetts case data and forecasts: 2021−06−07
Data
report−date cases (CSSE)
test−date cases
hospitalizations
case counts
4,000
3,000
2,000
1,000
0
Apr '21
May '21
Jun '21
Jul '21
Aug '21
Sep '

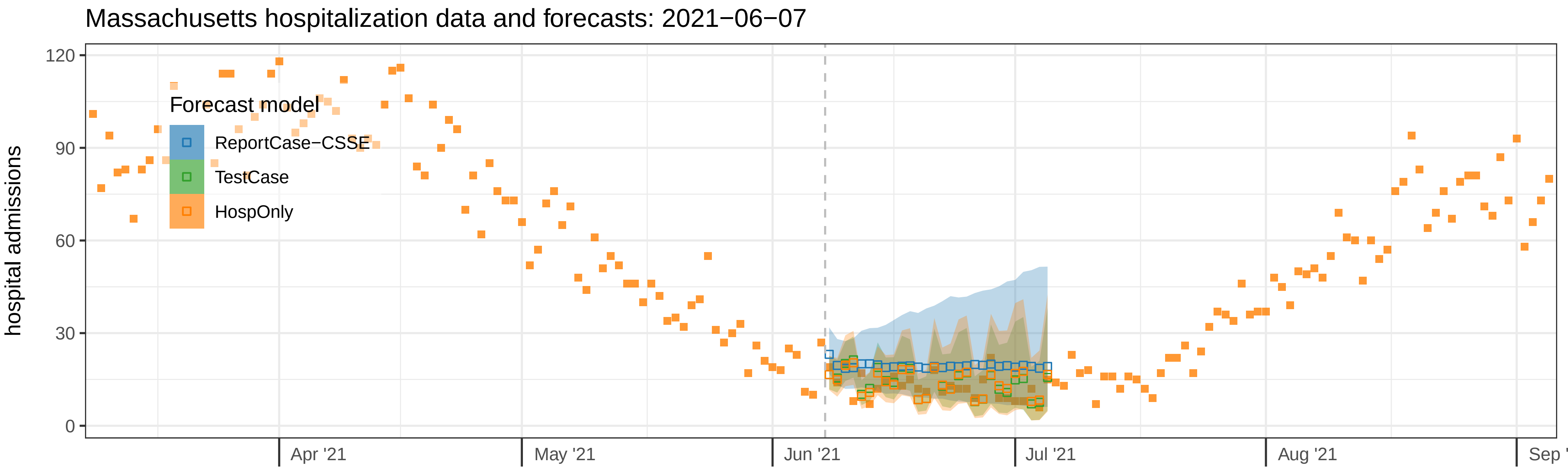
Massachusetts hospitalization data and forecasts: 2021−06−07
Forecast model
ReportCase−CSSE
TestCase
HospOnly
hospital admissions
120
90
60
30
0
Apr '21
May '21
Jun '21
Jul '21
Aug '21
Sep '

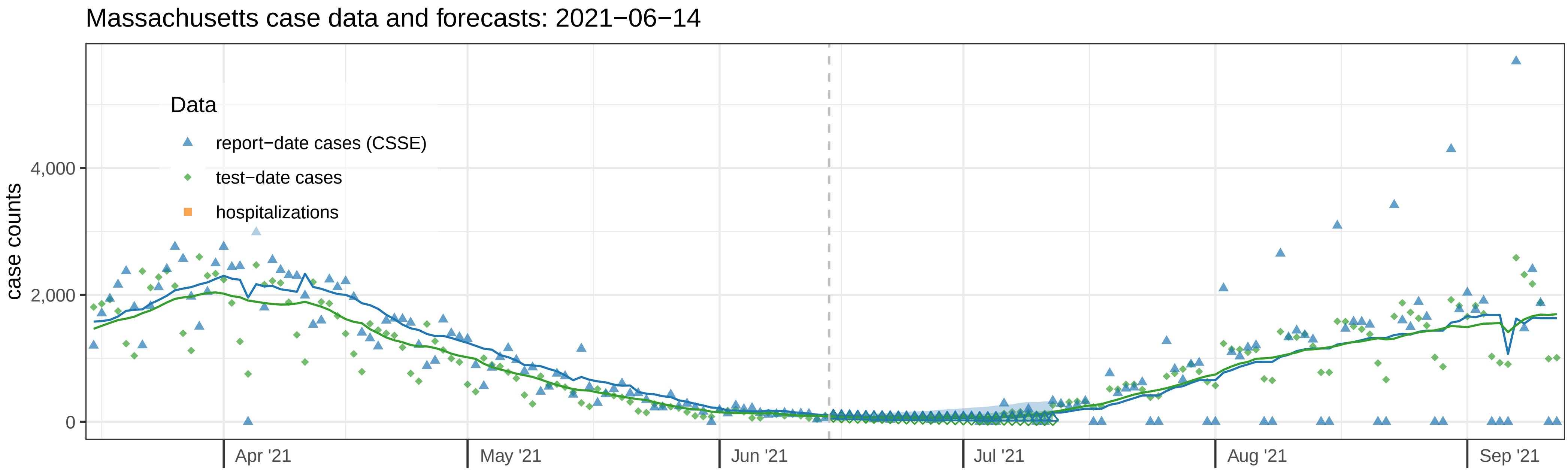

Massachusetts case data and forecasts: 2021−06−14
Data
report−date cases (CSSE)
test−date cases
hospitalizations
case counts
4,000
2,000
0
Apr '21
May '21
Jun '21
Jul '21
Aug '21
Sep '21


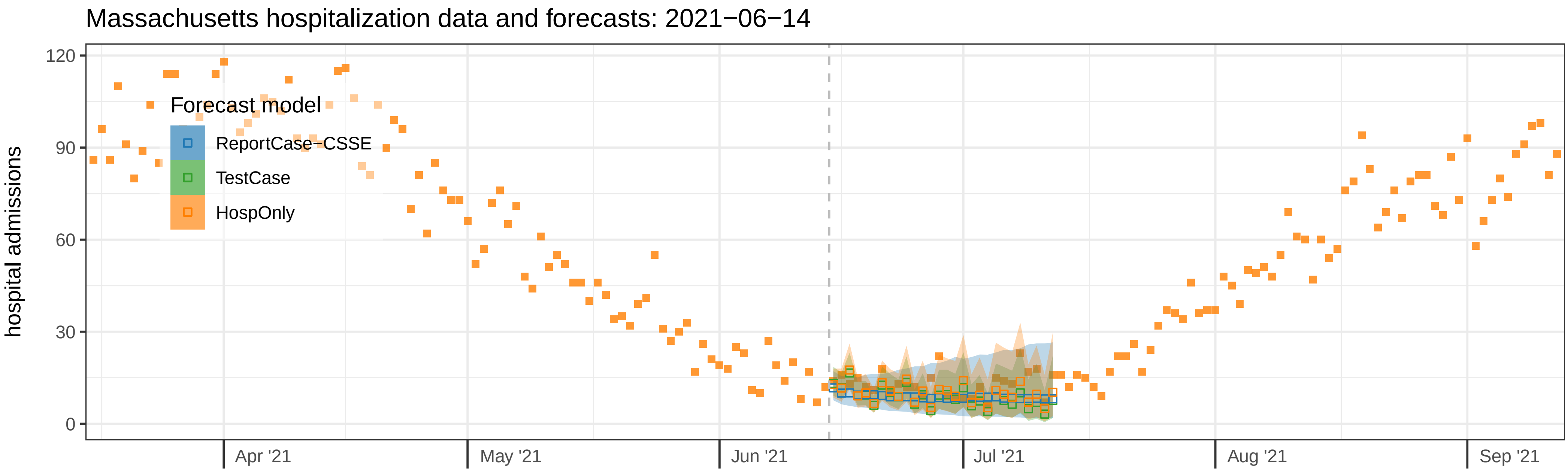

Massachusetts hospitalization data and forecasts: 2021−06−14
Forecast model
ReportCase−CSSE
TestCase
HospOnly
hospital admissions
120
90
60
30
0
Apr '21
May '21
Jun '21
Jul '21
Aug '21
Sep '21

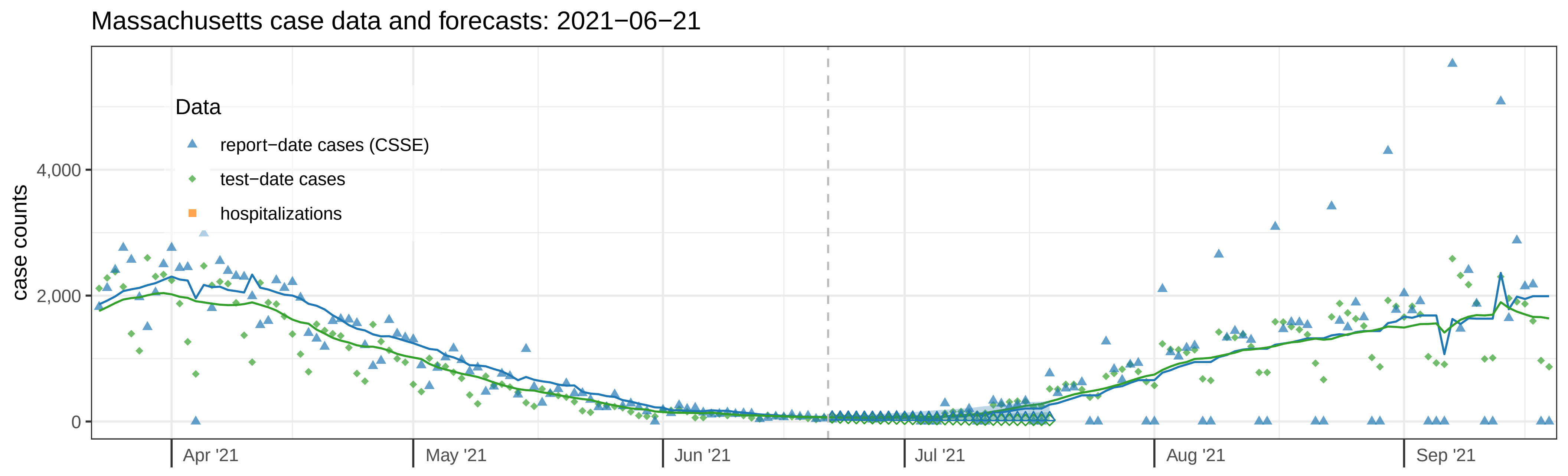
Massachusetts case data and forecasts: 2021−06−21
Data
report−date cases (CSSE)
test−date cases
hospitalizations
case counts
4,000
2,000
0
Apr '21
May '21
Jun '21
Jul '21
Aug '21
Sep '21

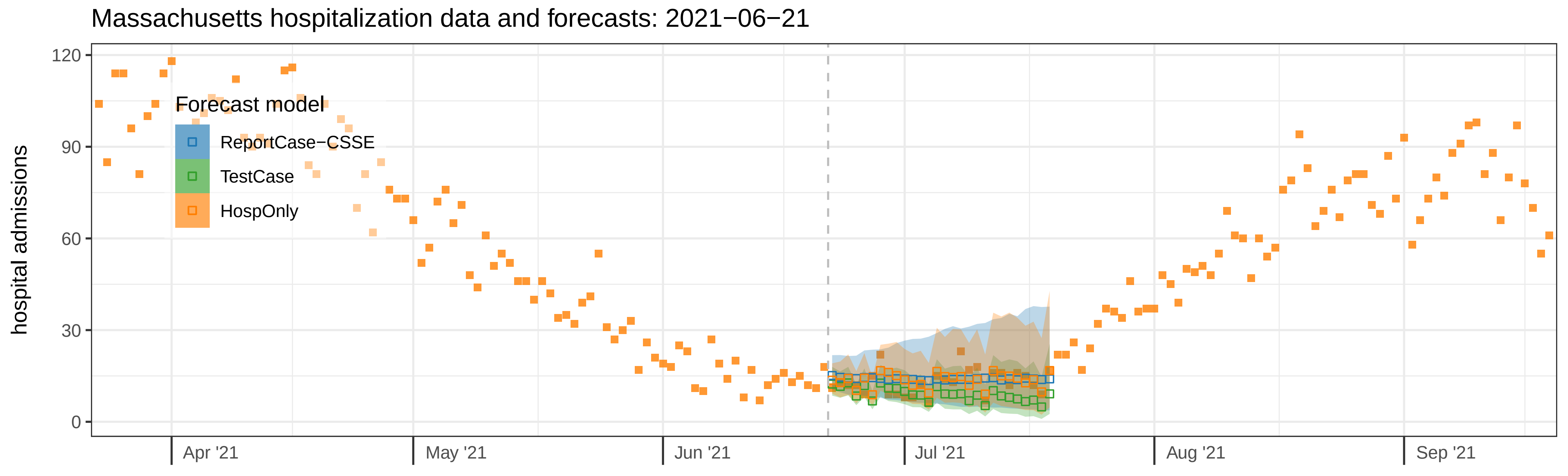
Massachusetts hospitalization data and forecasts: 2021−06−21
Forecast model
ReportCase−CSSE
TestCase
HospOnly
hospital admissions
120
90
60
30
0
Apr '21
May '21
Jun '21
Jul '21
Aug '21
Sep '21

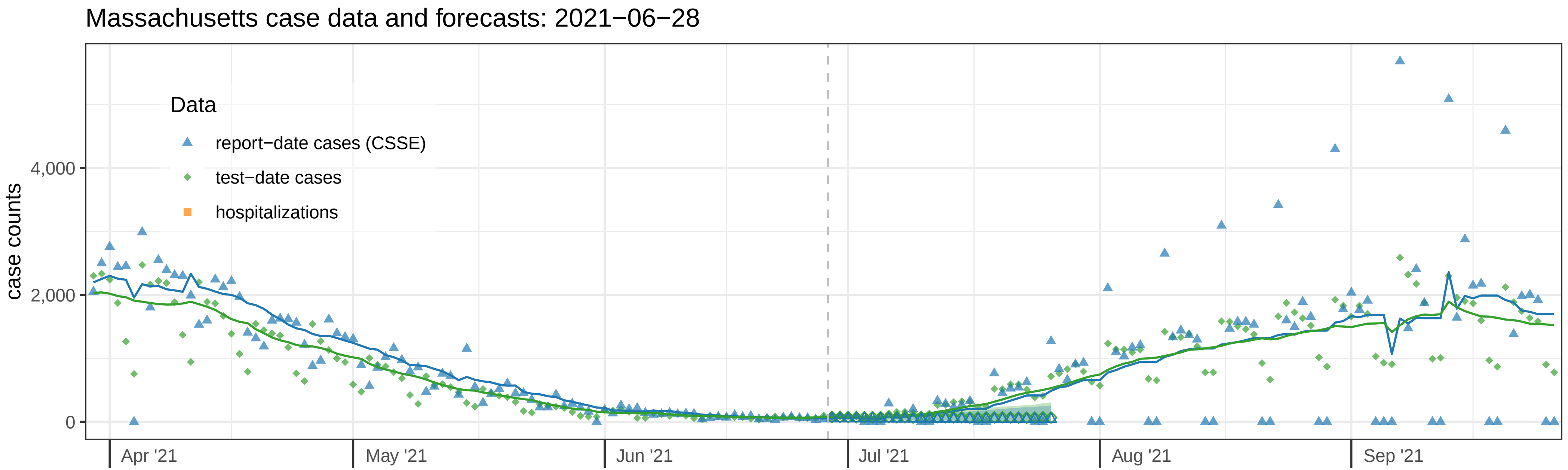

Massachusetts case data and forecasts: 2021−06−28
Data
report−date cases (CSSE)
test−date cases
hospitalizations
case counts
4,000
2,000
0
Apr '21
May '21
Jun '21
Jul '21
Aug '21
Sep '21


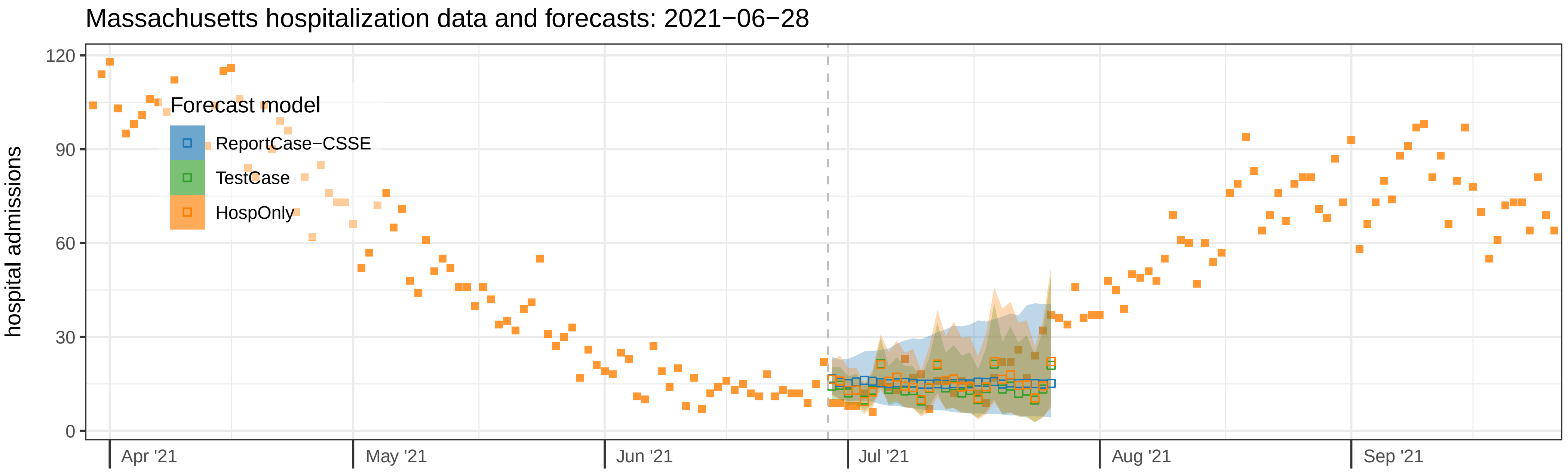

Massachusetts hospitalization data and forecasts: 2021−06−28
Forecast model
ReportCase−CSSE
TestCase
HospOnly
hospital admissions
120
90
60
30
0
Apr '21
May '21
Jun '21
Jul '21
Aug '21
Sep '21

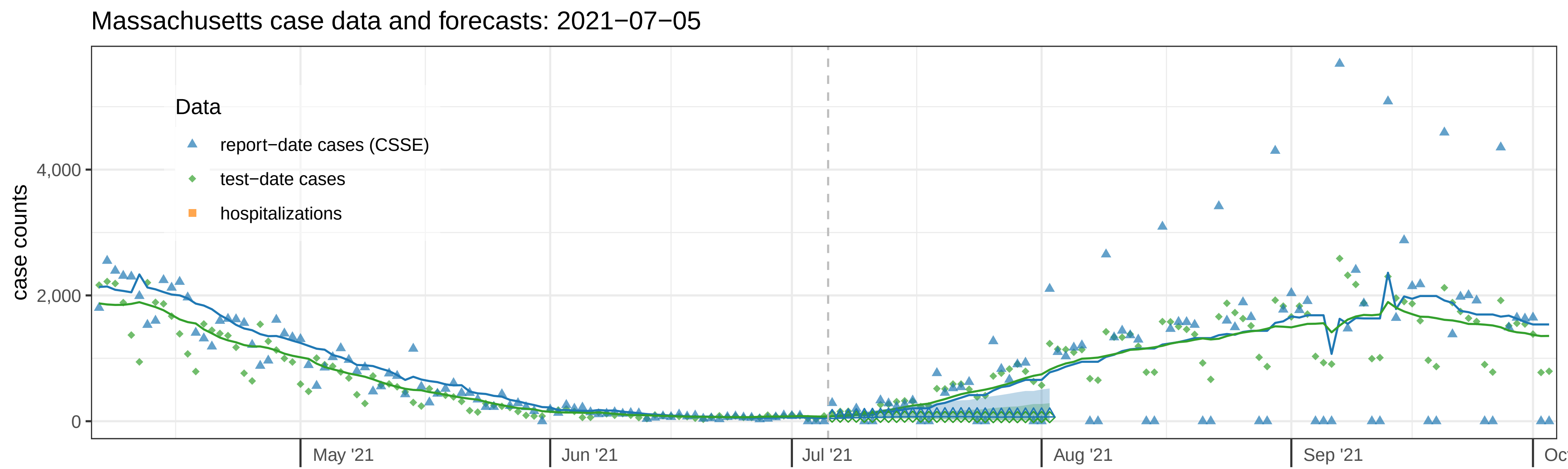

Massachusetts case data and forecasts: 2021−07−05
case counts
4,000
2,000
0
Data
report−date cases (CSSE)
test−date cases
hospitalizations
May '21
Jun '21
Jul '21
Aug '21
Sep '21


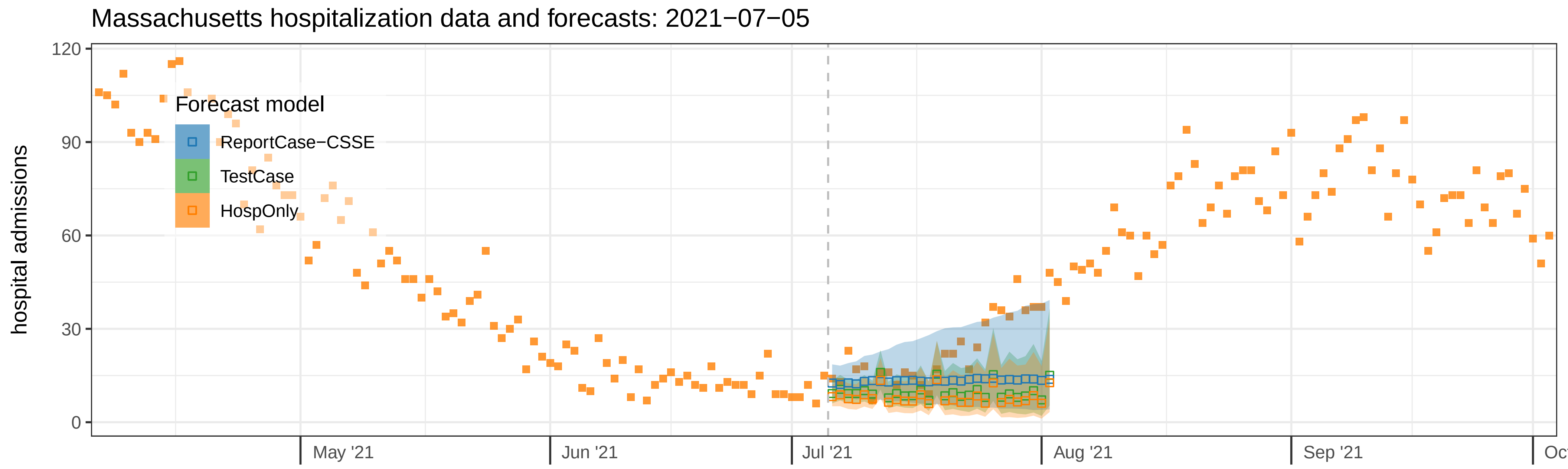

Massachusetts hospitalization data and forecasts: 2021−07−05
hospital admissions
120
90
60
30
0
Forecast model
ReportCase−CSSE
TestCase
HospOnly
May '21
Jun '21
Jul '21
Aug '21
Sep '21

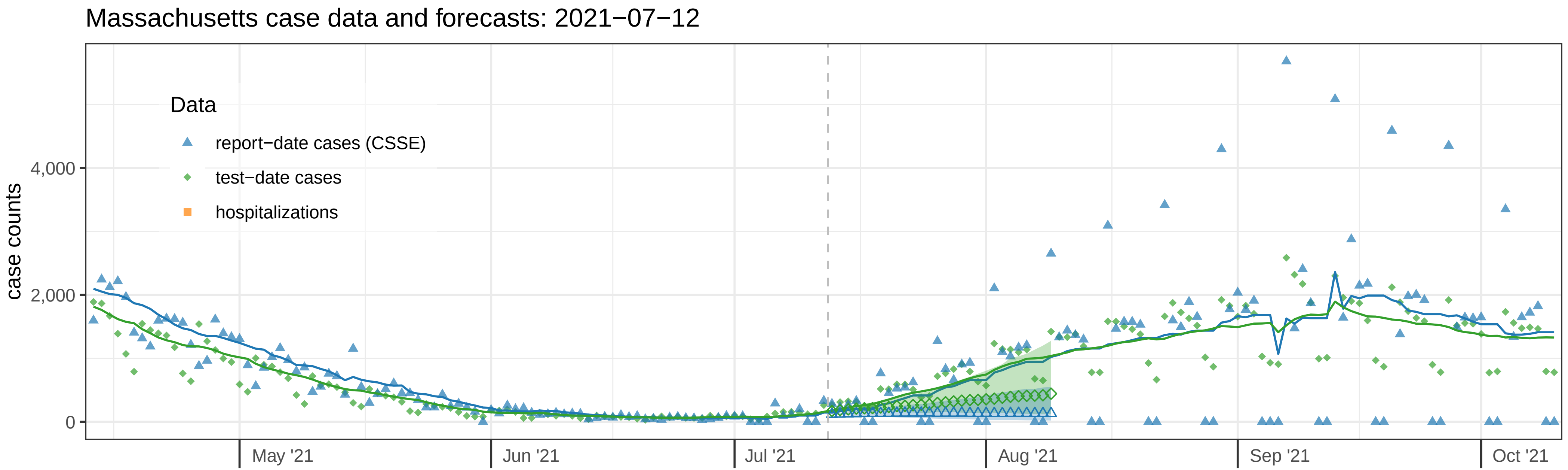
Massachusetts case data and forecasts: 2021−07−12
case counts
0
2,000
4,000
Data
report−date cases (CSSE)
test−date cases
hospitalizations
May '21
Jun '21
Jul '21
Aug '21
Sep '21
Oct '21

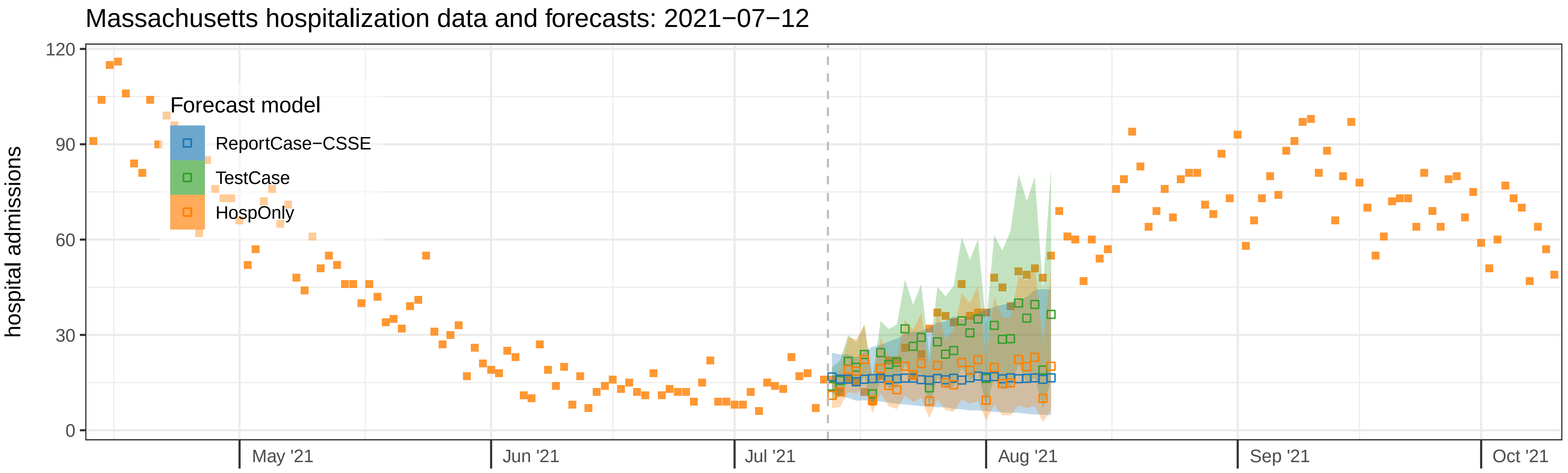
Massachusetts hospitalization data and forecasts: 2021−07−12
hospital admissions
0
30
60
90
120
Forecast model
ReportCase−CSSE
TestCase
HospOnly
May '21
Jun '21
Jul '21
Aug '21
Sep '21
Oct '21

### Massachusetts case data and forecasts: 2021−07−19

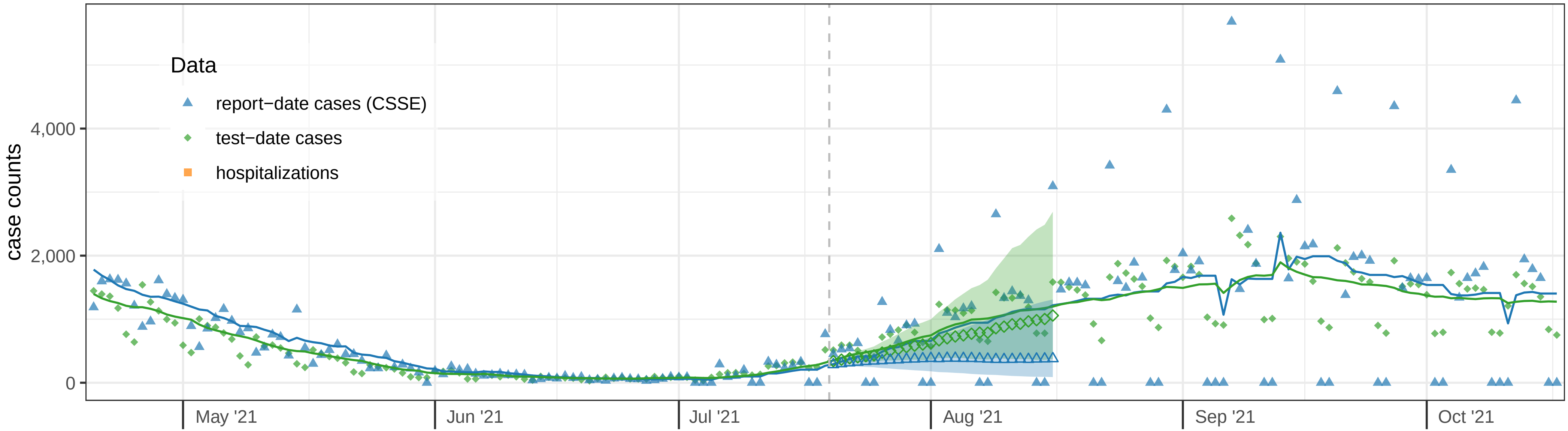


### Massachusetts hospitalization data and forecasts: 2021−07−19

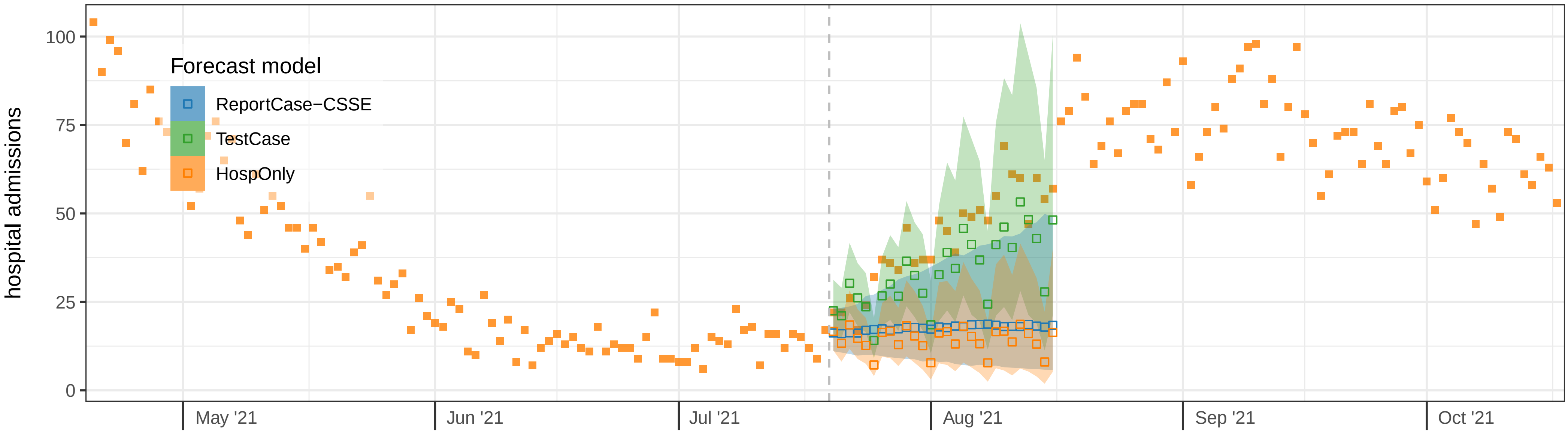

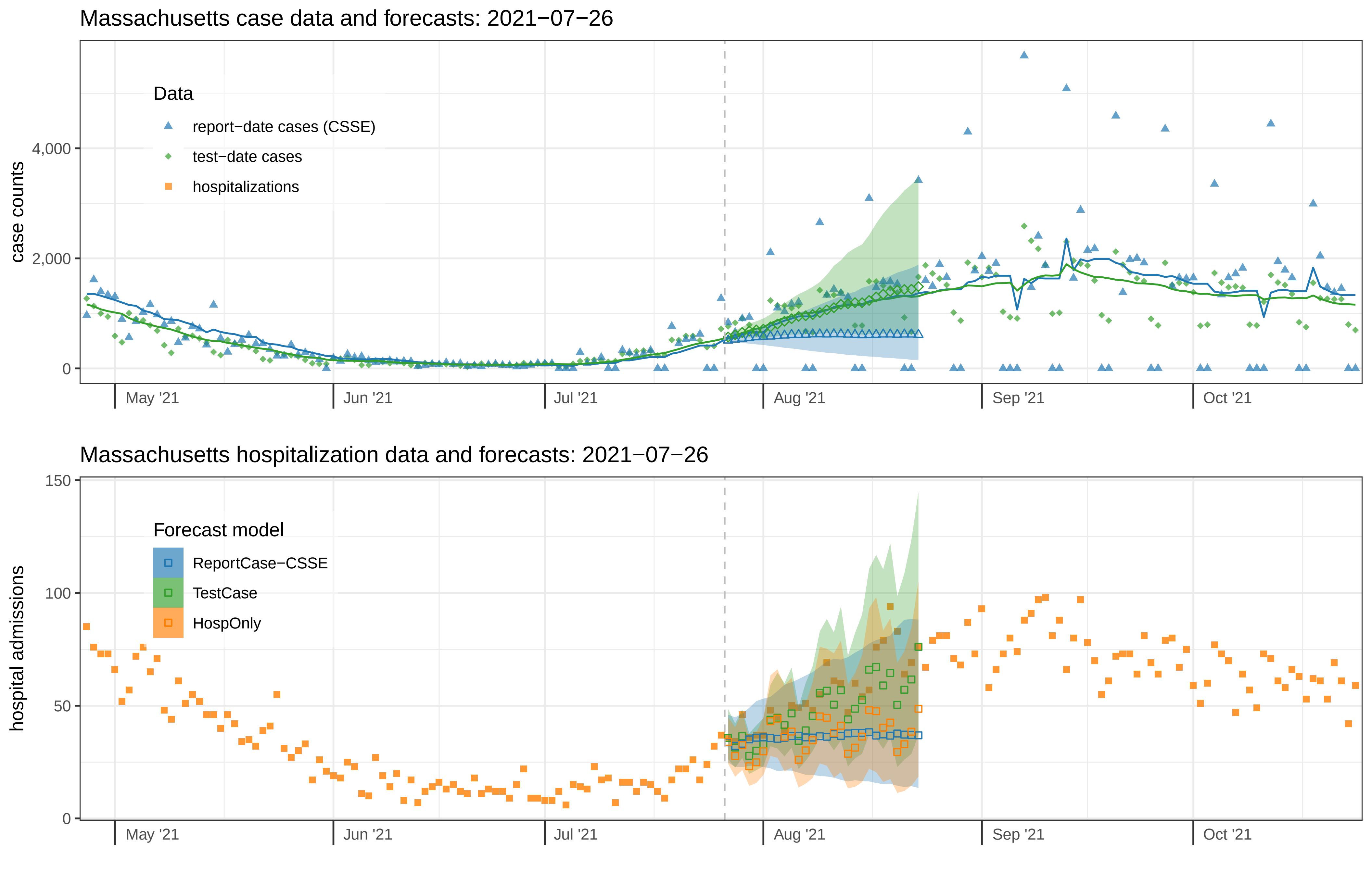

Massachusetts case data and forecasts: 2021−07−26
case counts
0
2,000
4,000
Data
report−date cases (CSSE)
test−date cases
hospitalizations
May '21
Jun '21
Jul '21
Aug '21
Sep '21
Oct '21
Massachusetts hospitalization data and forecasts: 2021−07−26
hospital admissions
0
50
100
150
Forecast model
ReportCase−CSSE
TestCase
HospOnly
May '21
Jun '21
Jul '21
Aug '21
Sep '21
Oct '21

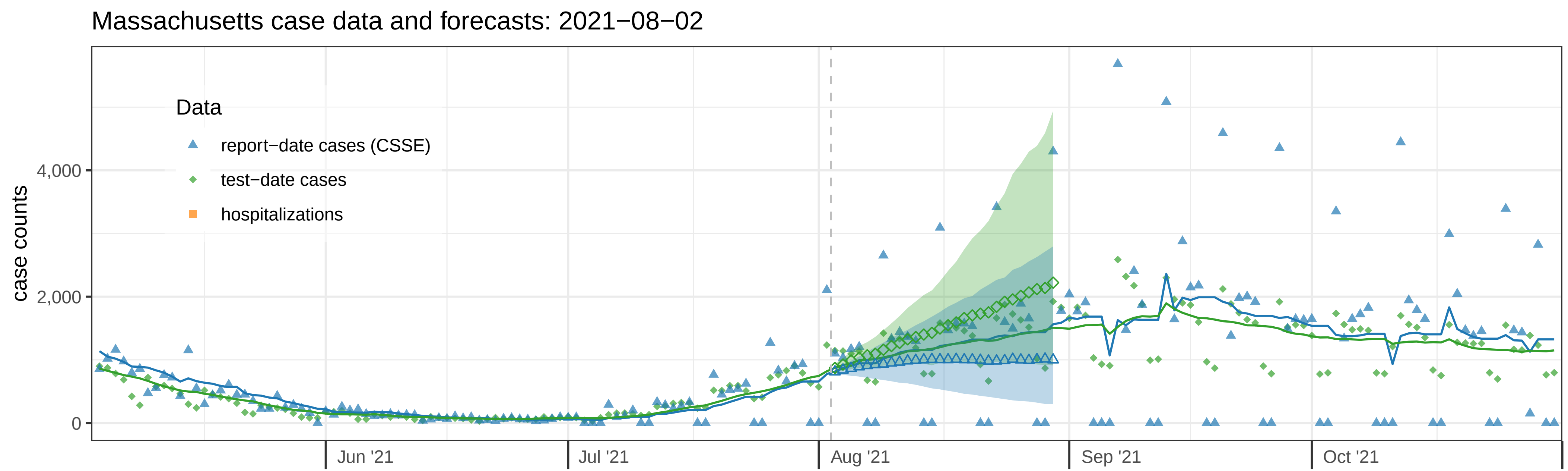
Massachusetts case data and forecasts: 2021−08−02
case counts
4,000
2,000
0
Data
report−date cases (CSSE)
test−date cases
hospitalizations
Jun '21
Jul '21
Aug '21
Sep '21
Oct '21

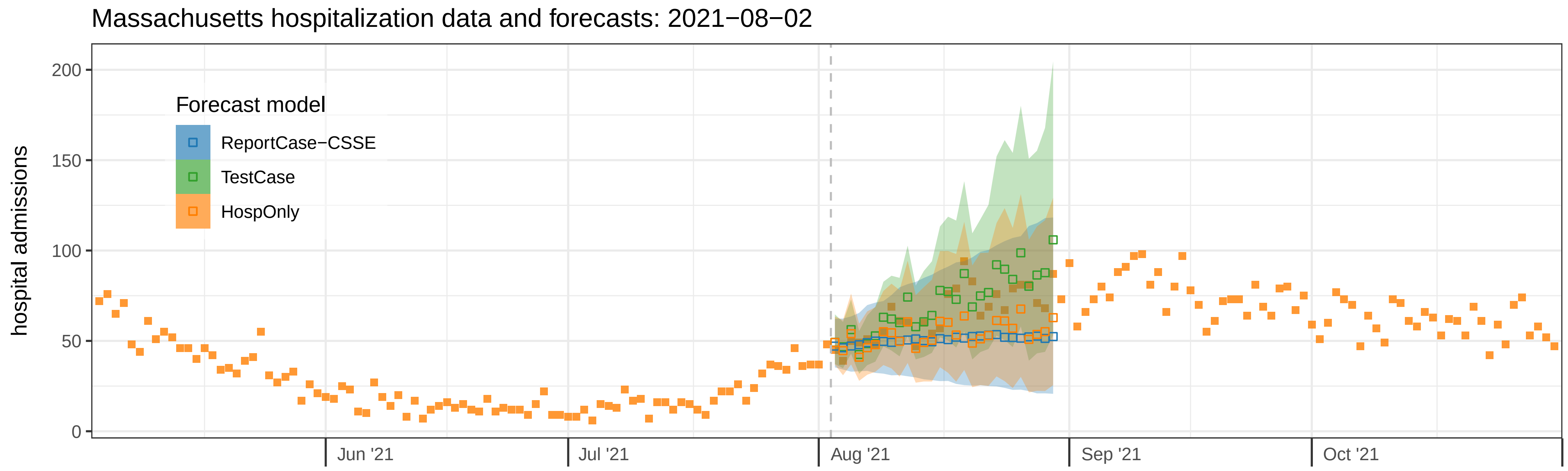
Massachusetts hospitalization data and forecasts: 2021−08−02
hospital admissions
200
150
100
50
0
Forecast model
ReportCase−CSSE
TestCase
HospOnly
Jun '21
Jul '21
Aug '21
Sep '21
Oct '21

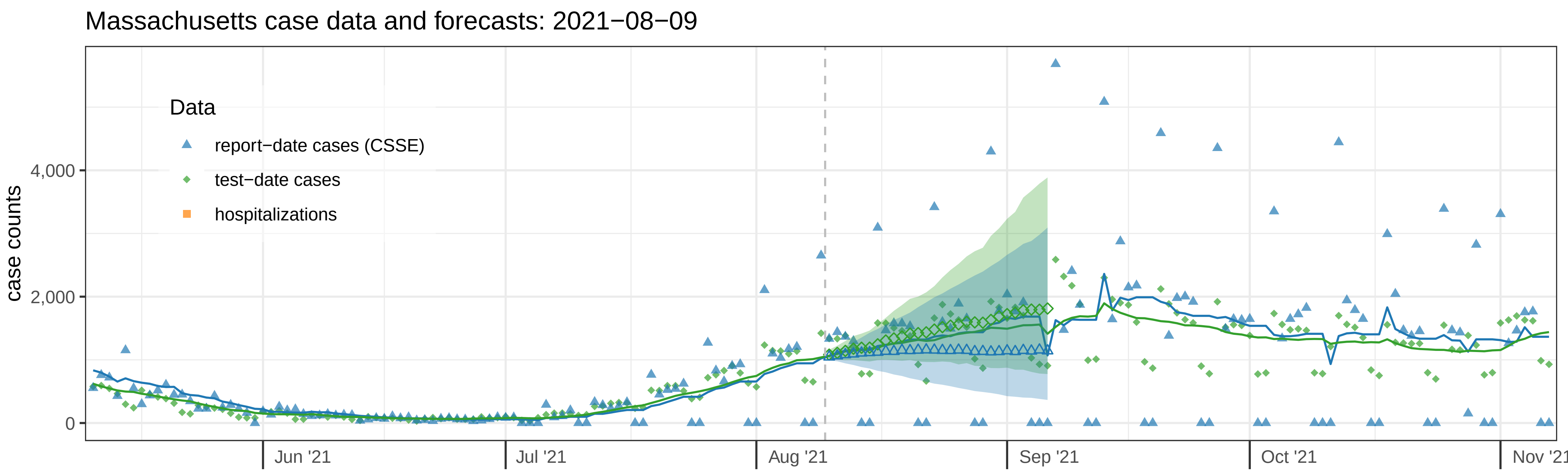

Massachusetts case data and forecasts: 2021−08−09
case counts
0
2,000
4,000
Data
report−date cases (CSSE)
test−date cases
hospitalizations
Jun '21
Jul '21
Aug '21
Sep '21
Oct '21
Nov '21


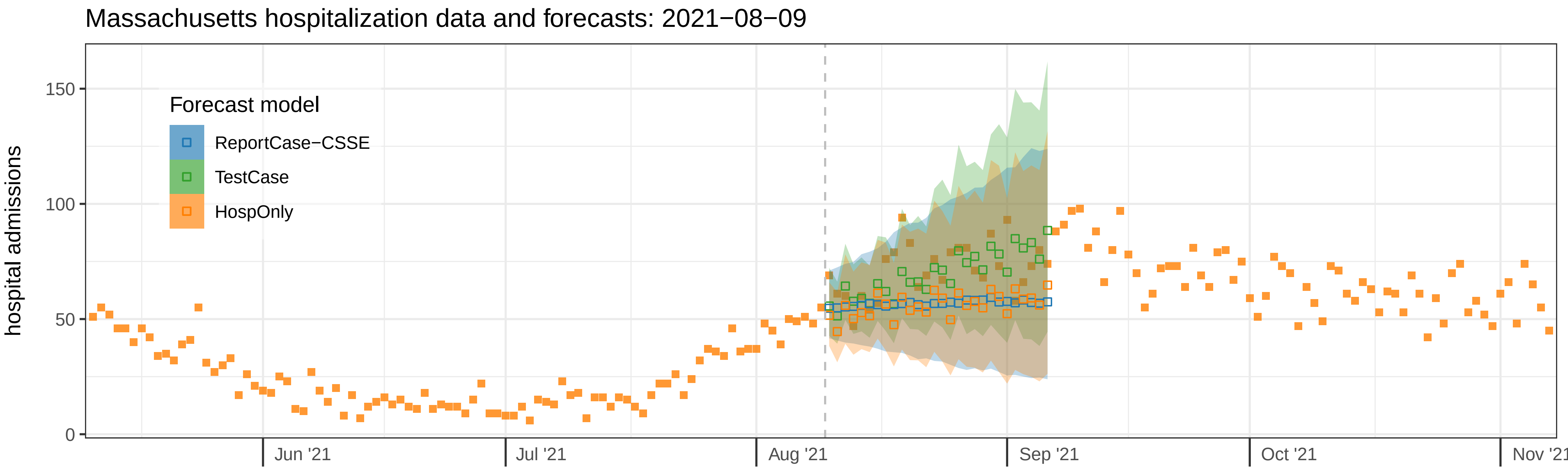

Massachusetts hospitalization data and forecasts: 2021−08−09
hospital admissions
0
50
100
150
Forecast model
ReportCase−CSSE
TestCase
HospOnly
Jun '21
Jul '21
Aug '21
Sep '21
Oct '21
Nov '21

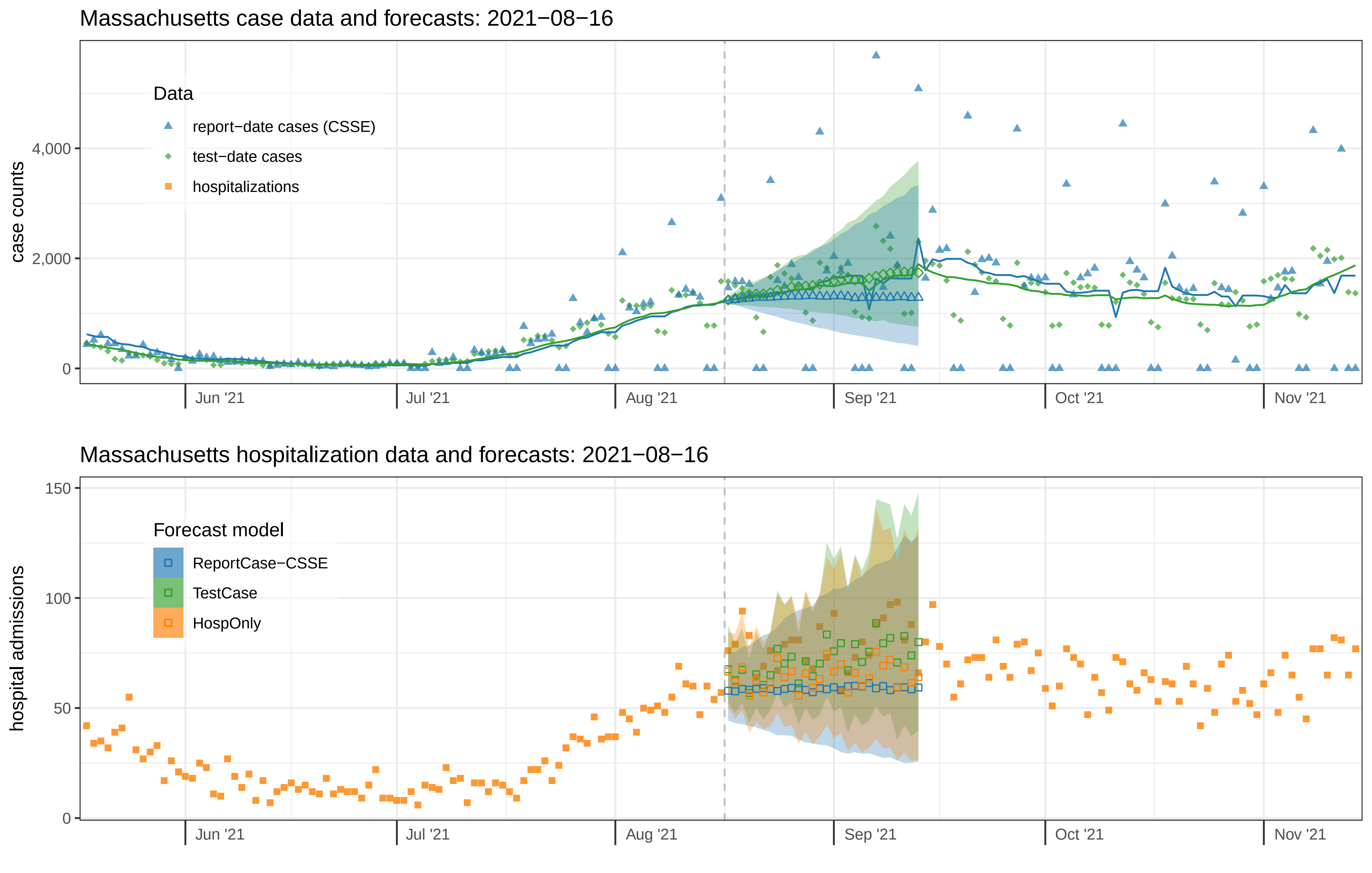
Massachusetts case data and forecasts: 2021−08−16
case counts
4,000
2,000
0
Data
report−date cases (CSSE)
test−date cases
hospitalizations
Jun '21
Jul '21
Aug '21
Sep '21
Oct '21
Nov '21
Massachusetts hospitalization data and forecasts: 2021−08−16
hospital admissions
150
100
50
0
Forecast model
ReportCase−CSSE
TestCase
HospOnly
Jun '21
Jul '21
Aug '21
Sep '21
Oct '21
Nov '21

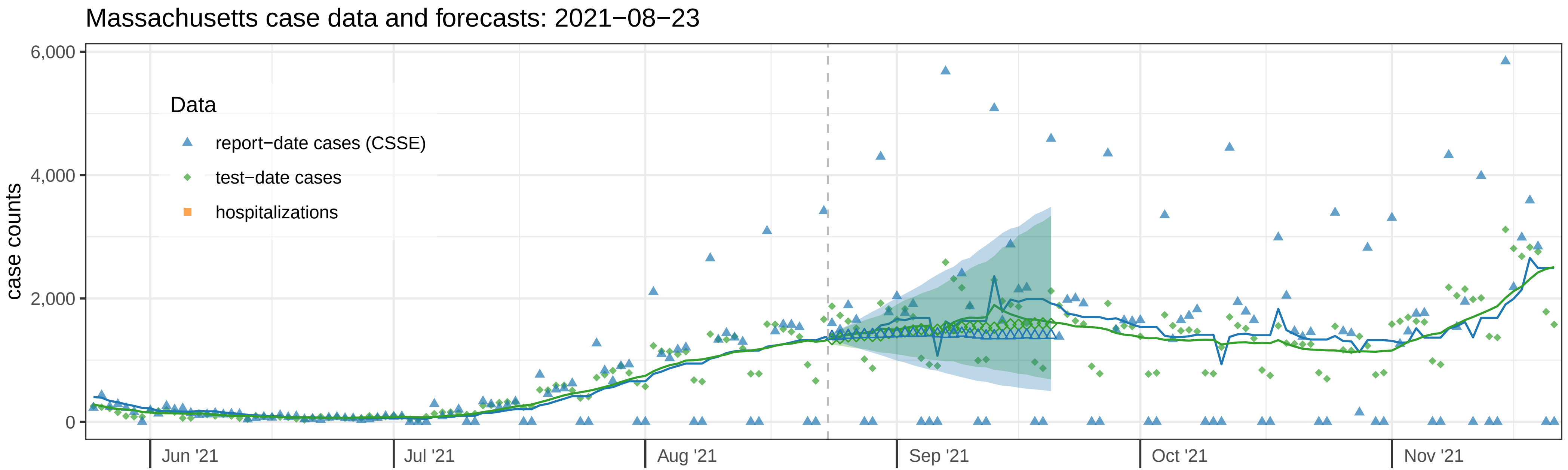

Massachusetts case data and forecasts: 2021−08−23
case counts
6,000
4,000
2,000
0
Data
report−date cases (CSSE)
test−date cases
hospitalizations
Jun '21
Jul '21
Aug '21
Sep '21
Oct '21
Nov '21


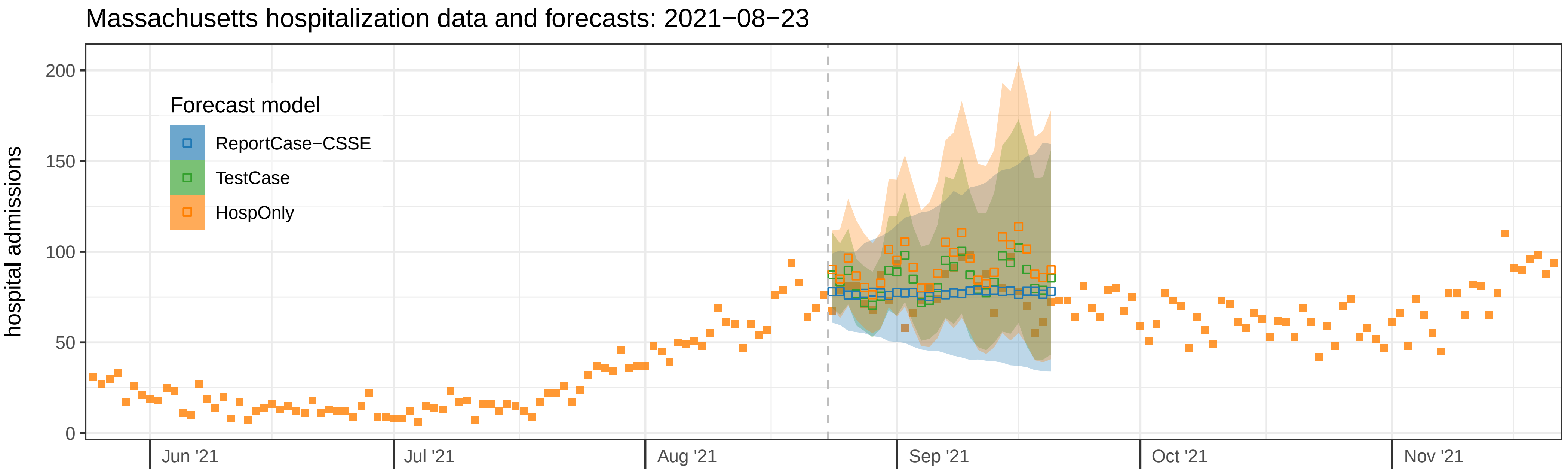

Massachusetts hospitalization data and forecasts: 2021−08−23
hospital admissions
200
150
100
50
0
Forecast model
ReportCase−CSSE
TestCase
HospOnly
Jun '21
Jul '21
Aug '21
Sep '21
Oct '21
Nov '21

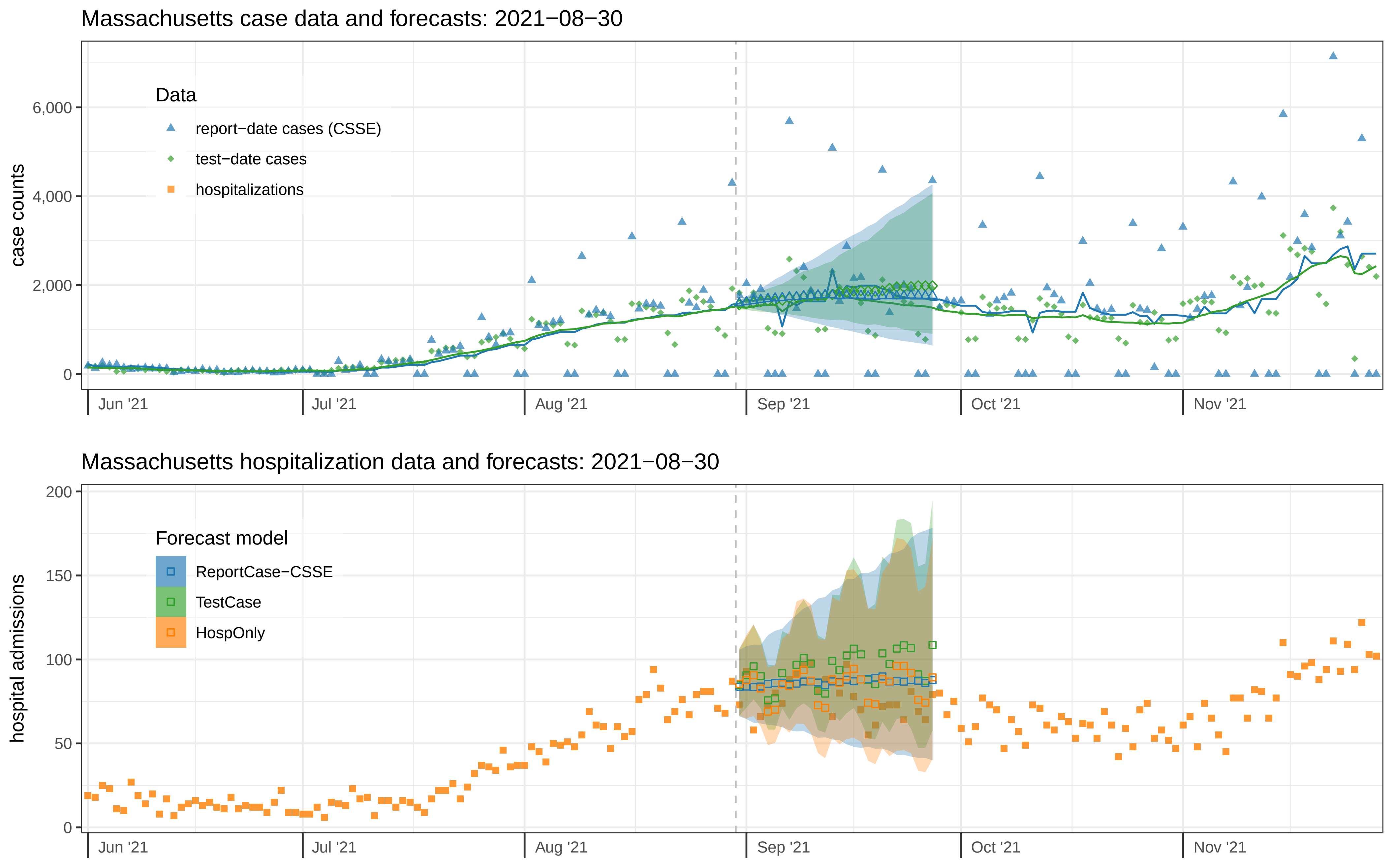

Massachusetts case data and forecasts: 2021−08−30
case counts
6,000
4,000
2,000
0
Data
report−date cases (CSSE)
test−date cases
hospitalizations
Jun '21
Jul '21
Aug '21
Sep '21
Oct '21
Nov '21
Massachusetts hospitalization data and forecasts: 2021−08−30
hospital admissions
200
150
100
50
0
Forecast model
ReportCase−CSSE
TestCase
HospOnly
Jun '21
Jul '21
Aug '21
Sep '21
Oct '21
Nov '21

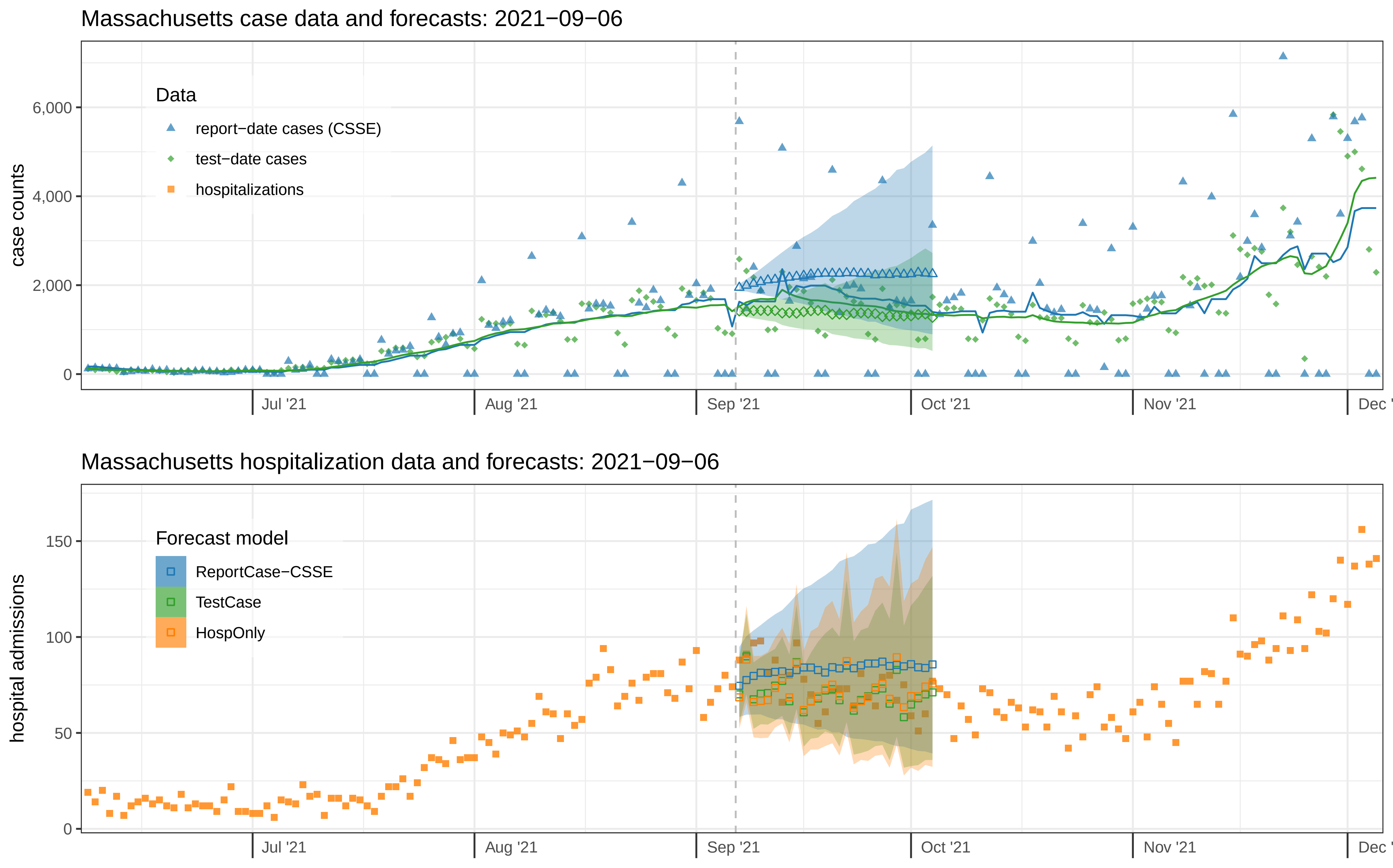

Massachusetts case data and forecasts: 2021−09−06
case counts
0
2,000
4,000
6,000
Data
report−date cases (CSSE)
test−date cases
hospitalizations
Jul '21
Aug '21
Sep '21
Oct '21
Nov '21
Dec '
Massachusetts hospitalization data and forecasts: 2021−09−06
hospital admissions
0
50
100
150
Forecast model
ReportCase−CSSE
TestCase
HospOnly
Jul '21
Aug '21
Sep '21
Oct '21
Nov '21
Dec '

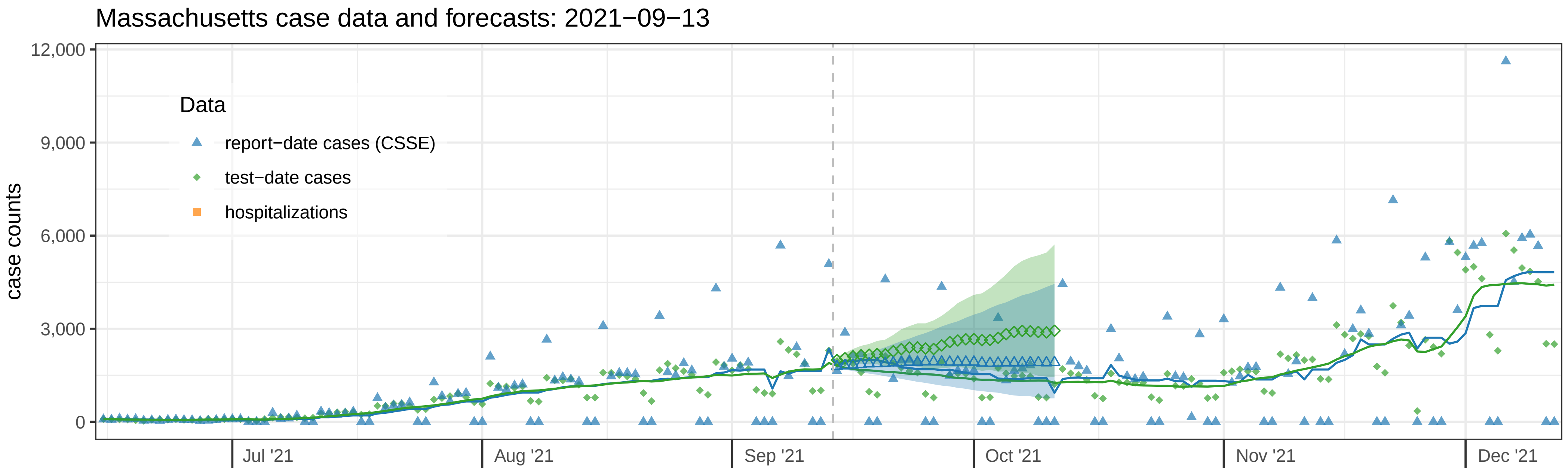

Massachusetts case data and forecasts: 2021−09−13
case counts
12,000
9,000
6,000
3,000
0
Data
report−date cases (CSSE)
test−date cases
hospitalizations
Jul '21
Aug '21
Sep '21
Oct '21
Nov '21
Dec '21


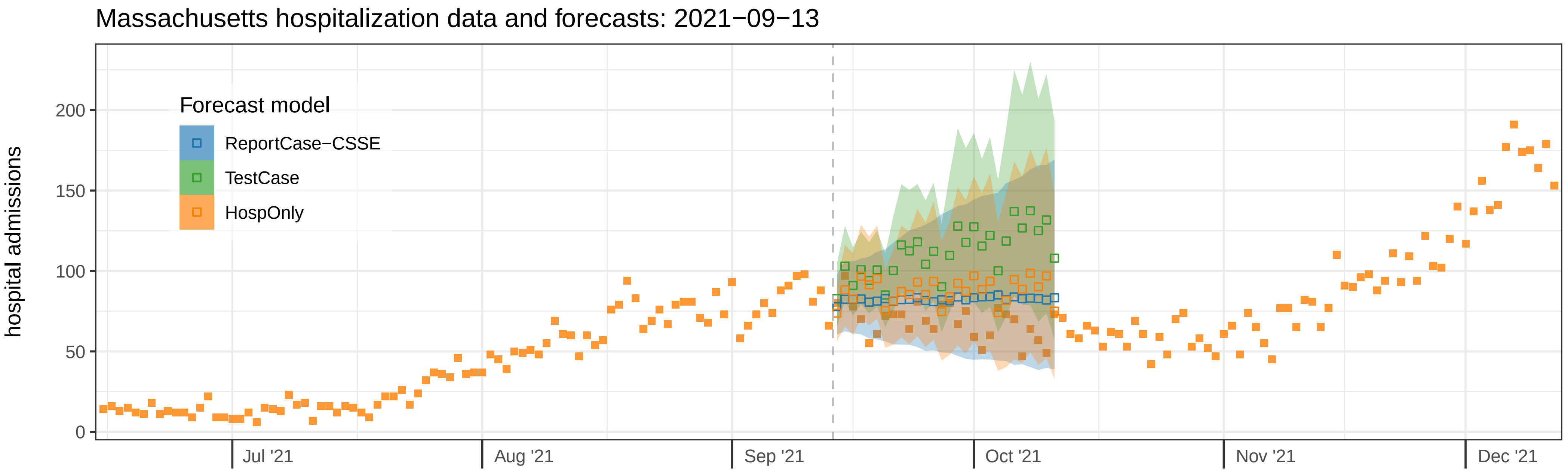

Massachusetts hospitalization data and forecasts: 2021−09−13
hospital admissions
200
150
100
50
0
Forecast model
ReportCase−CSSE
TestCase
HospOnly
Jul '21
Aug '21
Sep '21
Oct '21
Nov '21
Dec '21

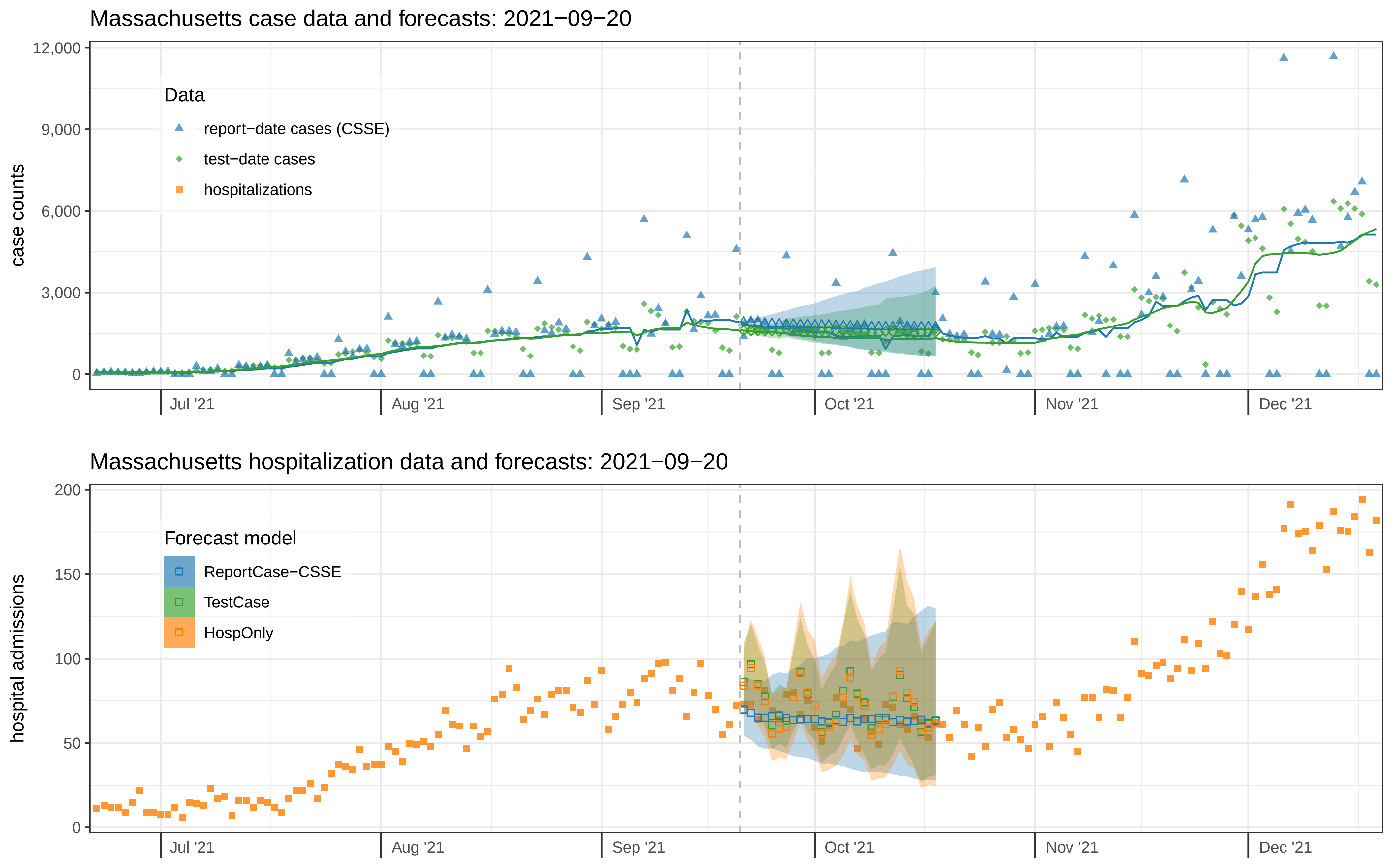

Massachusetts case data and forecasts: 2021−09−20
case counts
12,000
9,000
6,000
3,000
0
Data
report−date cases (CSSE)
test−date cases
hospitalizations
Jul '21
Aug '21
Sep '21
Oct '21
Nov '21
Dec '21
Massachusetts hospitalization data and forecasts: 2021−09−20
hospital admissions
200
150
100
50
0
Forecast model
ReportCase−CSSE
TestCase
HospOnly
Jul '21
Aug '21
Sep '21
Oct '21
Nov '21
Dec '21

Massachusetts case data and forecasts: 2021−09−27

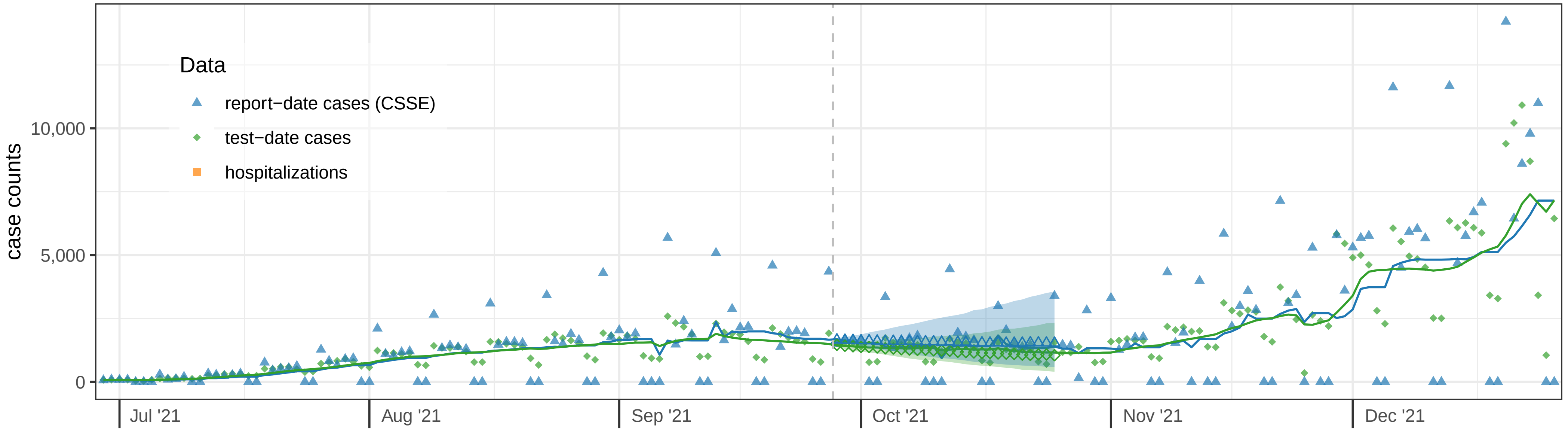


Massachusetts hospitalization data and forecasts: 2021−09−27

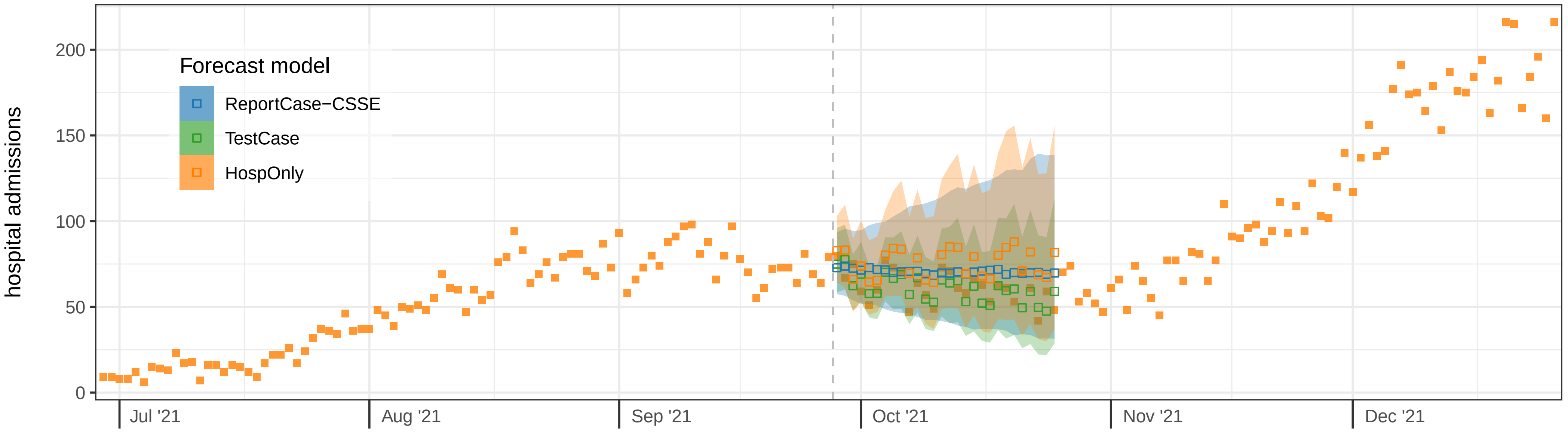

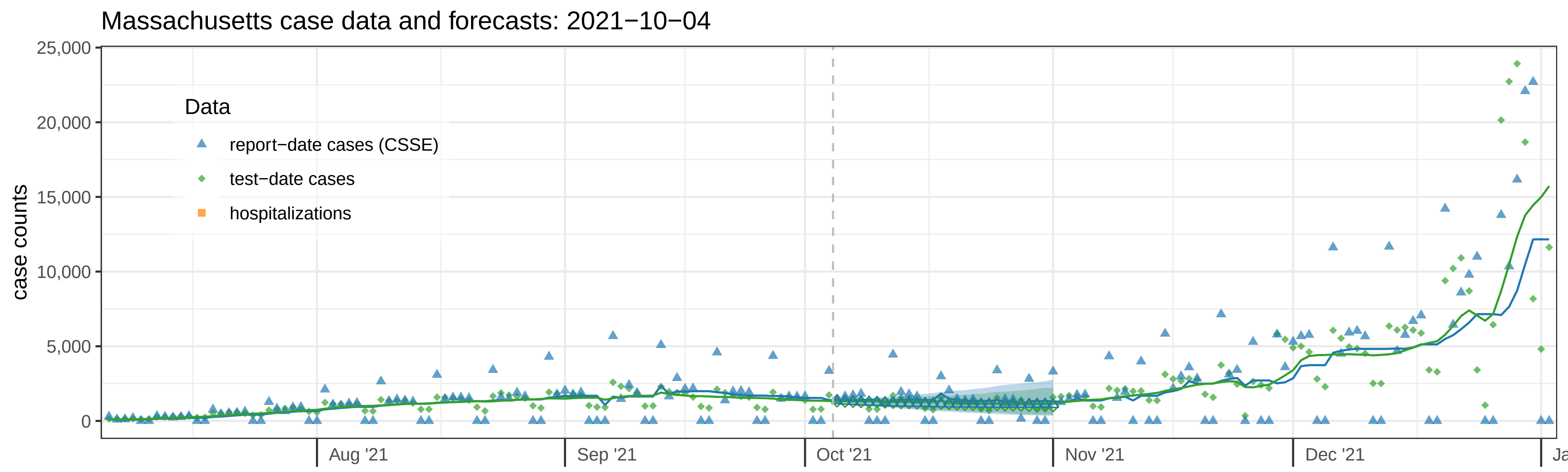

Massachusetts case data and forecasts: 2021−10−04
case counts
25,000
20,000
15,000
10,000
5,000
0
Data
report−date cases (CSSE)
test−date cases
hospitalizations
Aug '21
Sep '21
Oct '21
Nov '21
Dec '21


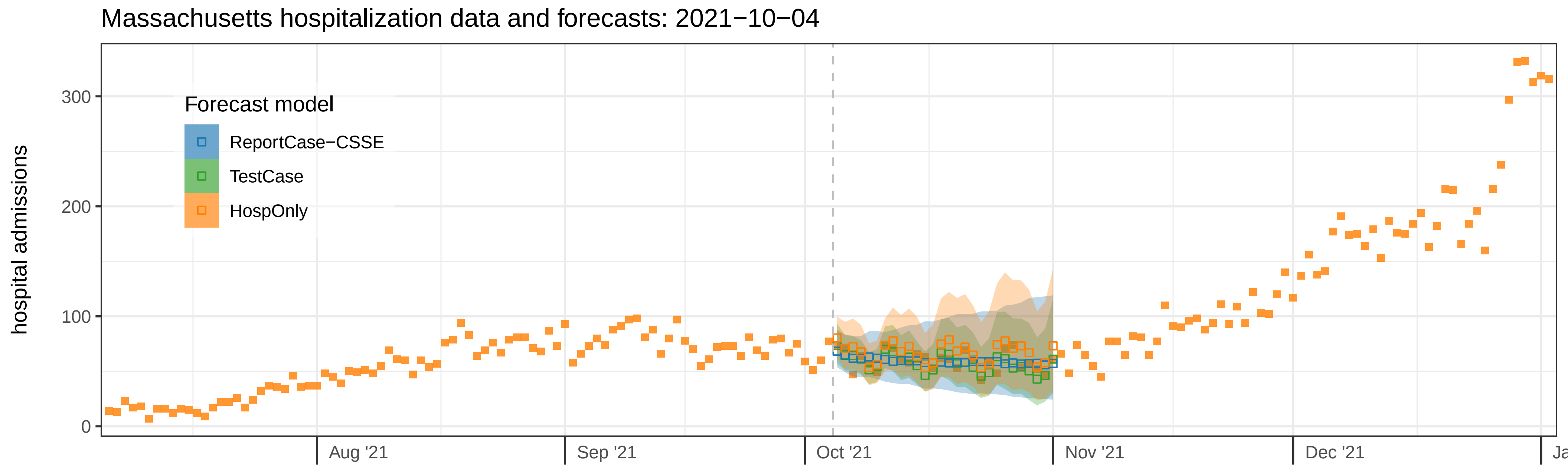

Massachusetts hospitalization data and forecasts: 2021−10−04
hospital admissions
300
200
100
0
Forecast model
ReportCase−CSSE
TestCase
HospOnly
Aug '21
Sep '21
Oct '21
Nov '21
Dec '21

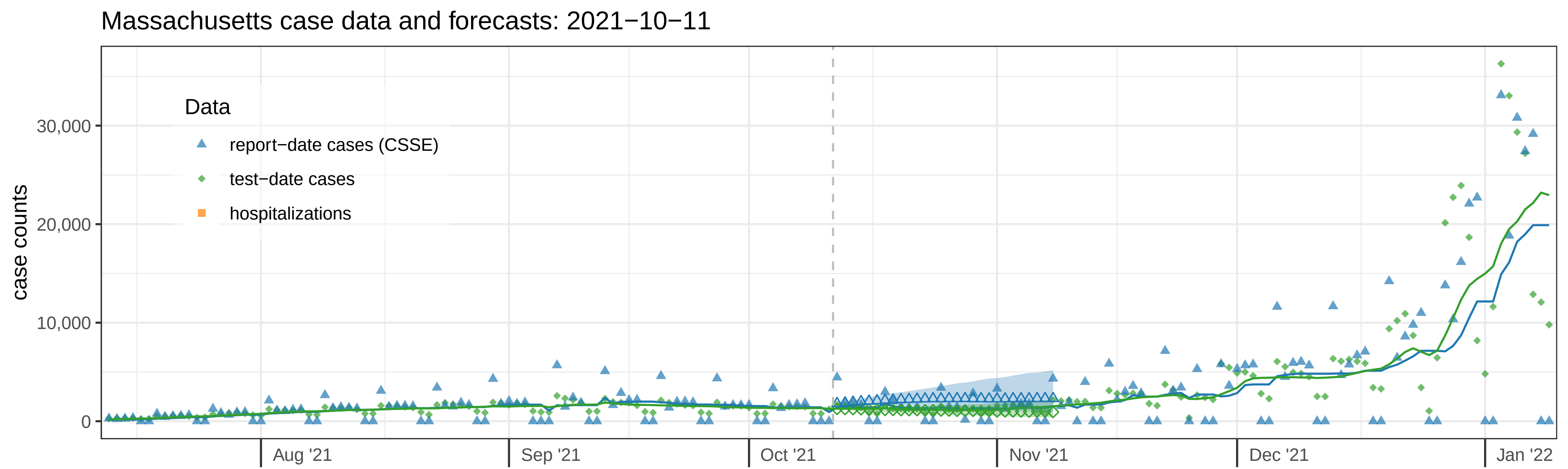
Massachusetts case data and forecasts: 2021−10−11
case counts
30,000
20,000
10,000
0
Data
report−date cases (CSSE)
test−date cases
hospitalizations
Aug '21
Sep '21
Oct '21
Nov '21
Dec '21
Jan '22

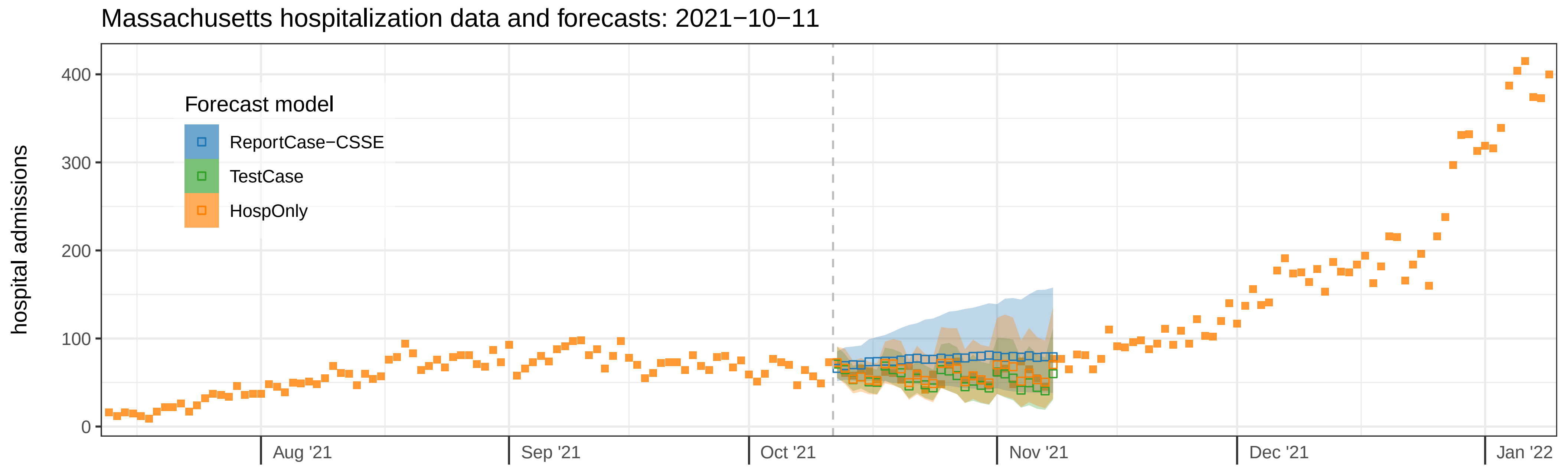
Massachusetts hospitalization data and forecasts: 2021−10−11
hospital admissions
400
300
200
100
0
Forecast model
ReportCase−CSSE
TestCase
HospOnly
Aug '21
Sep '21
Oct '21
Nov '21
Dec '21
Jan '22

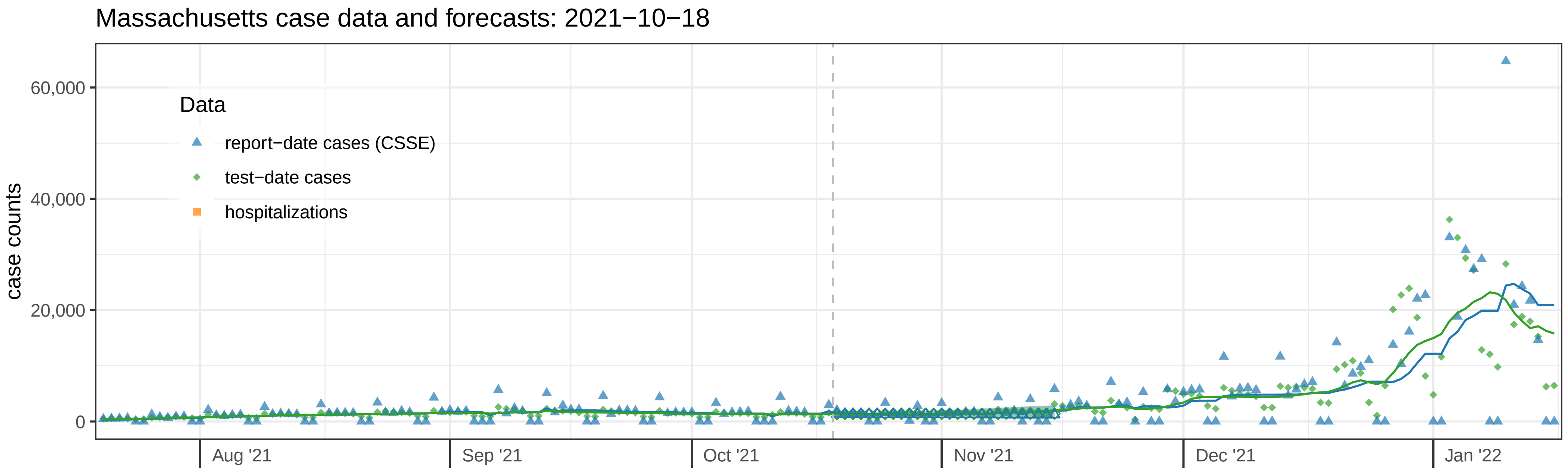
Massachusetts case data and forecasts: 2021−10−18
case counts
60,000
40,000
20,000
0
Data
report−date cases (CSSE)
test−date cases
hospitalizations
Aug '21
Sep '21
Oct '21
Nov '21
Dec '21
Jan '22

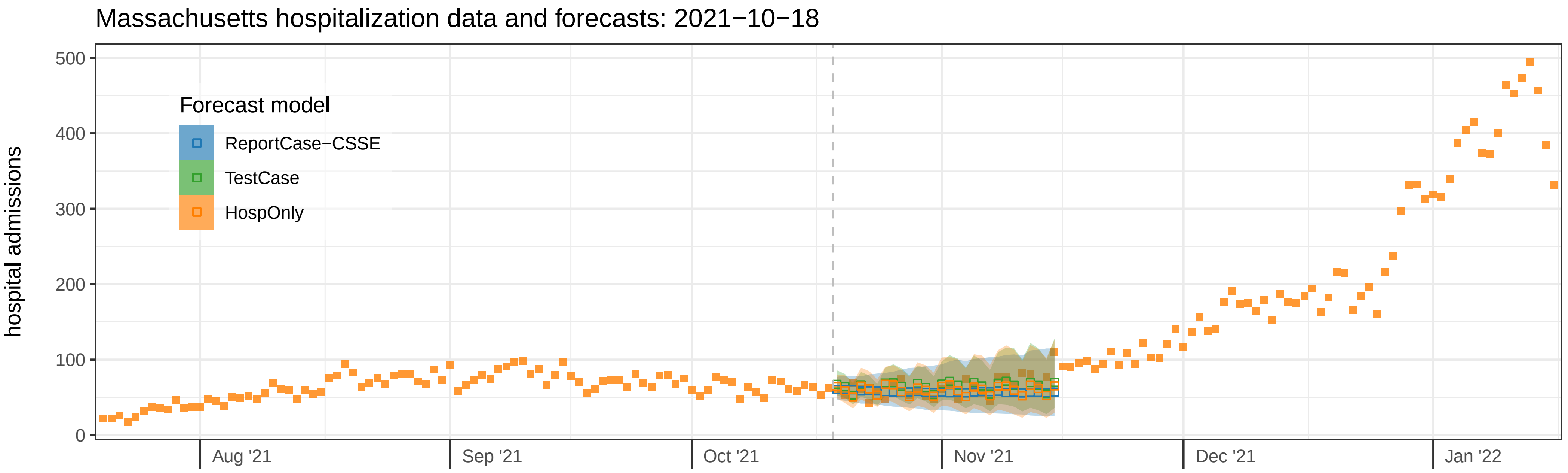
Massachusetts hospitalization data and forecasts: 2021−10−18
hospital admissions
500
400
300
200
100
0
Forecast model
ReportCase−CSSE
TestCase
HospOnly
Aug '21
Sep '21
Oct '21
Nov '21
Dec '21
Jan '22

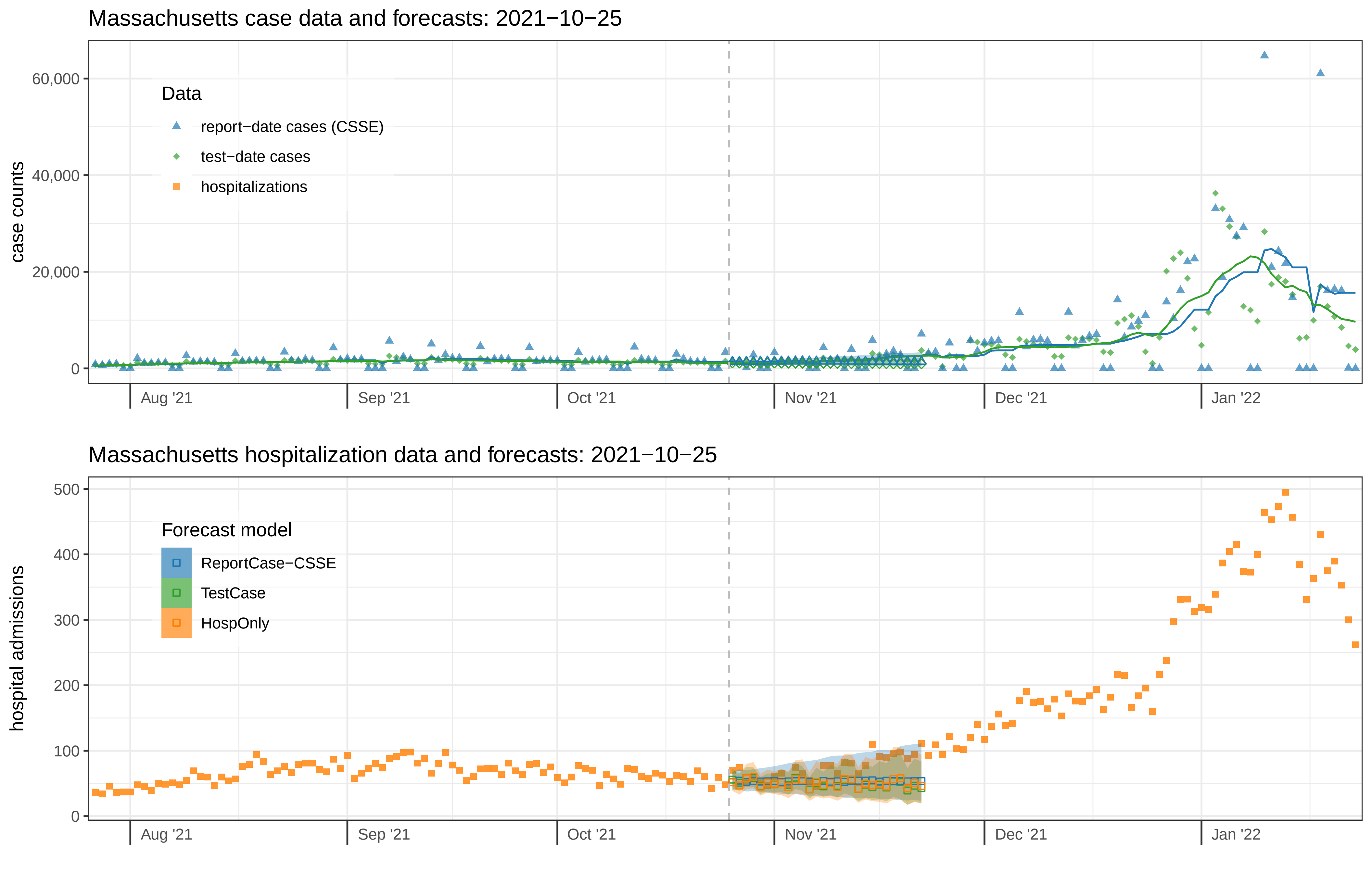

Massachusetts case data and forecasts: 2021−10−25
case counts
60,000
40,000
20,000
0
Data
report−date cases (CSSE)
test−date cases
hospitalizations
Aug '21
Sep '21
Oct '21
Nov '21
Dec '21
Jan '22
Massachusetts hospitalization data and forecasts: 2021−10−25
hospital admissions
500
400
300
200
100
0
Forecast model
ReportCase−CSSE
TestCase
HospOnly
Aug '21
Sep '21
Oct '21
Nov '21
Dec '21
Jan '22

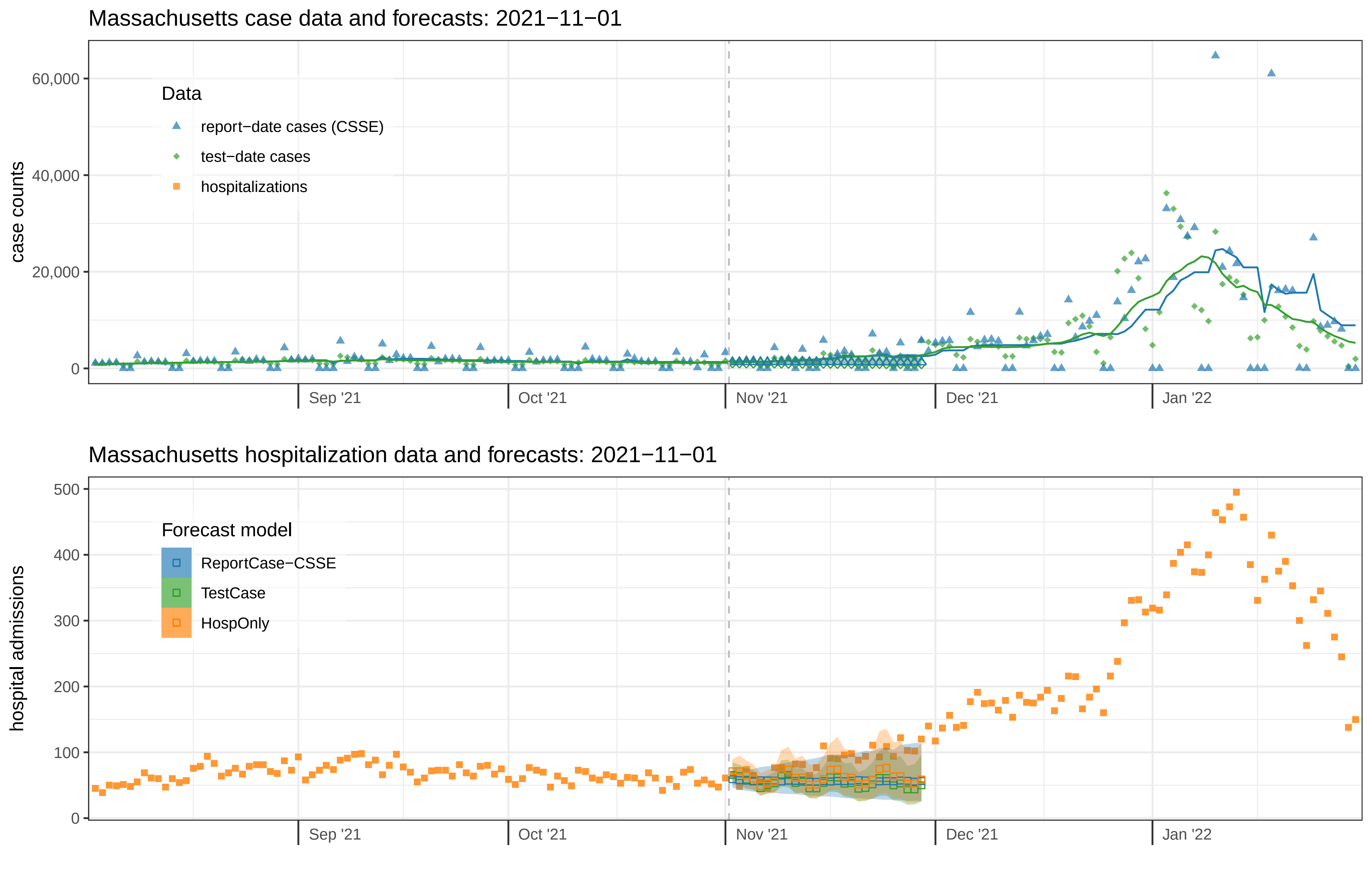
Massachusetts case data and forecasts: 2021−11−01
case counts
60,000
40,000
20,000
0
Data
report−date cases (CSSE)
test−date cases
hospitalizations
Sep '21
Oct '21
Nov '21
Dec '21
Jan '22
Massachusetts hospitalization data and forecasts: 2021−11−01
hospital admissions
500
400
300
200
100
0
Forecast model
ReportCase−CSSE
TestCase
HospOnly
Sep '21
Oct '21
Nov '21
Dec '21
Jan '22

Massachusetts case data and forecasts: 2021−11−08

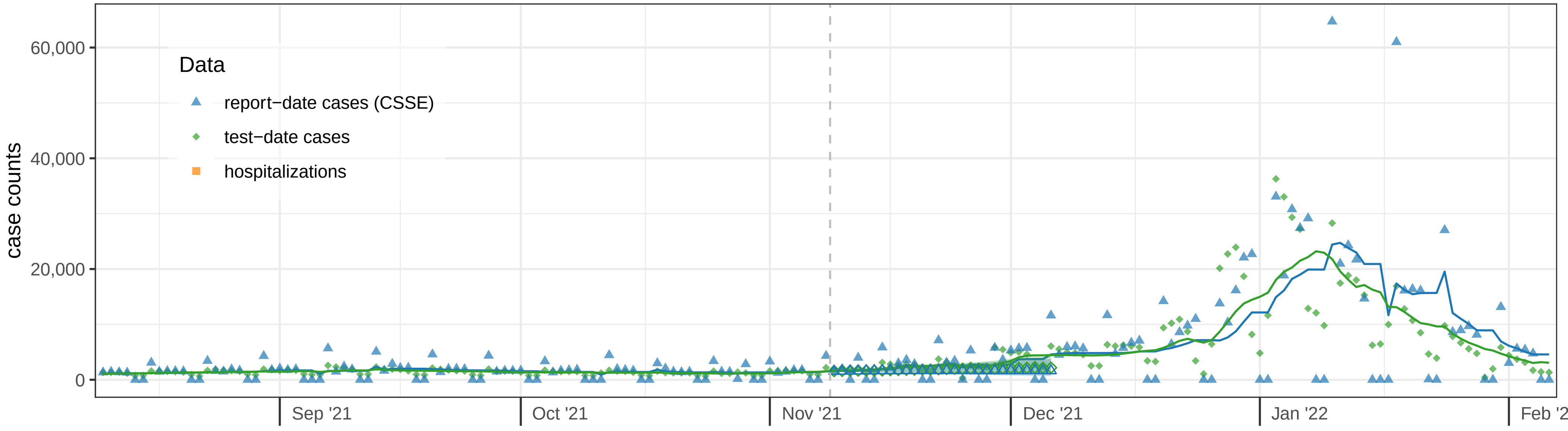


Massachusetts hospitalization data and forecasts: 2021−11−08

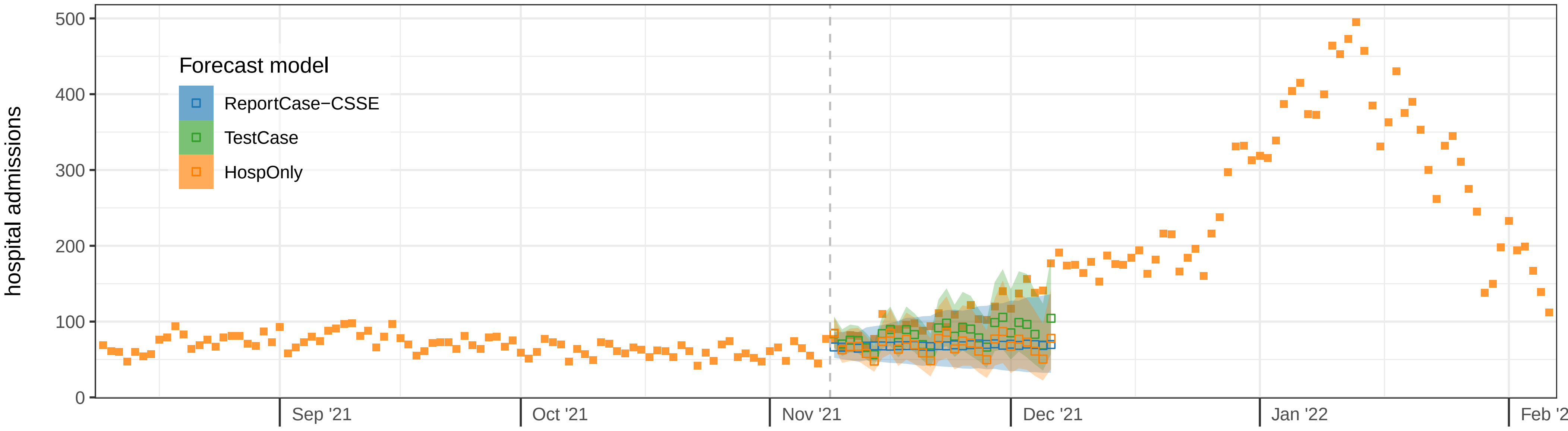

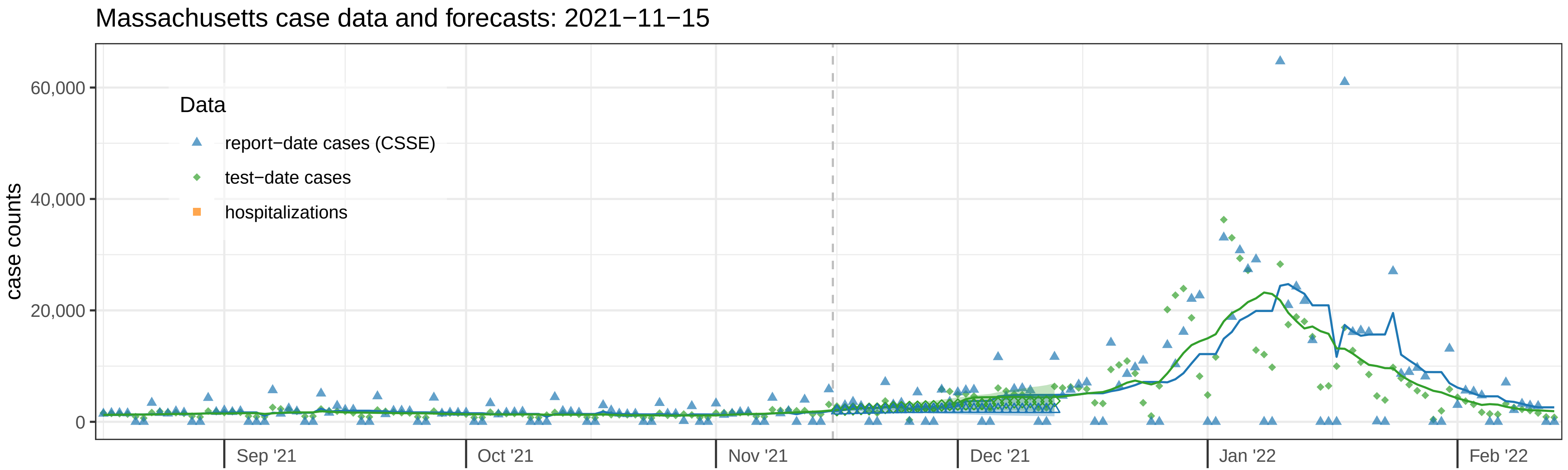

Massachusetts case data and forecasts: 2021−11−15
case counts
60,000
40,000
20,000
0
Data
report−date cases (CSSE)
test−date cases
hospitalizations
Sep '21
Oct '21
Nov '21
Dec '21
Jan '22
Feb '22


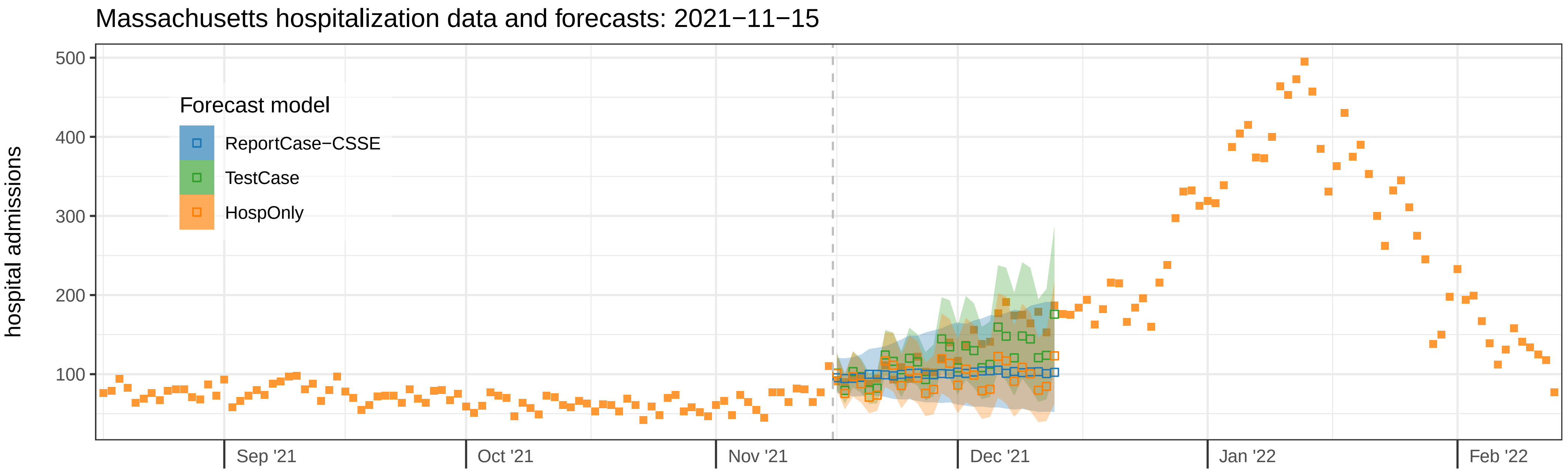

Massachusetts hospitalization data and forecasts: 2021−11−15
hospital admissions
500
400
300
200
100
Forecast model
ReportCase−CSSE
TestCase
HospOnly
Sep '21
Oct '21
Nov '21
Dec '21
Jan '22
Feb '22

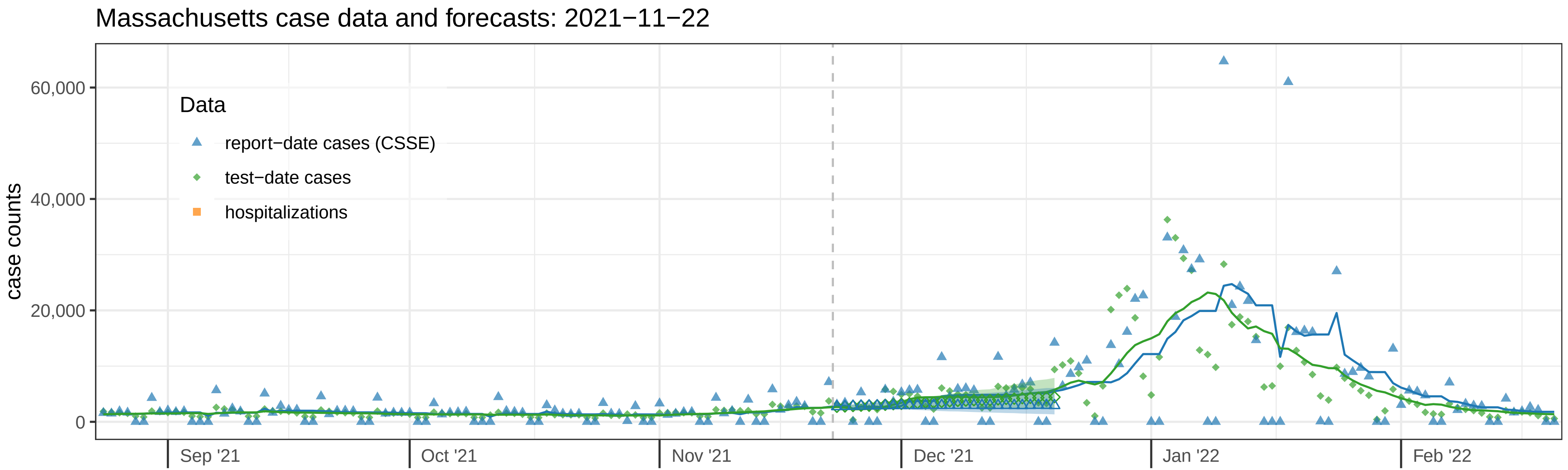
Massachusetts case data and forecasts: 2021−11−22
case counts
60,000
40,000
20,000
0
Data
report−date cases (CSSE)
test−date cases
hospitalizations
Sep '21
Oct '21
Nov '21
Dec '21
Jan '22
Feb '22

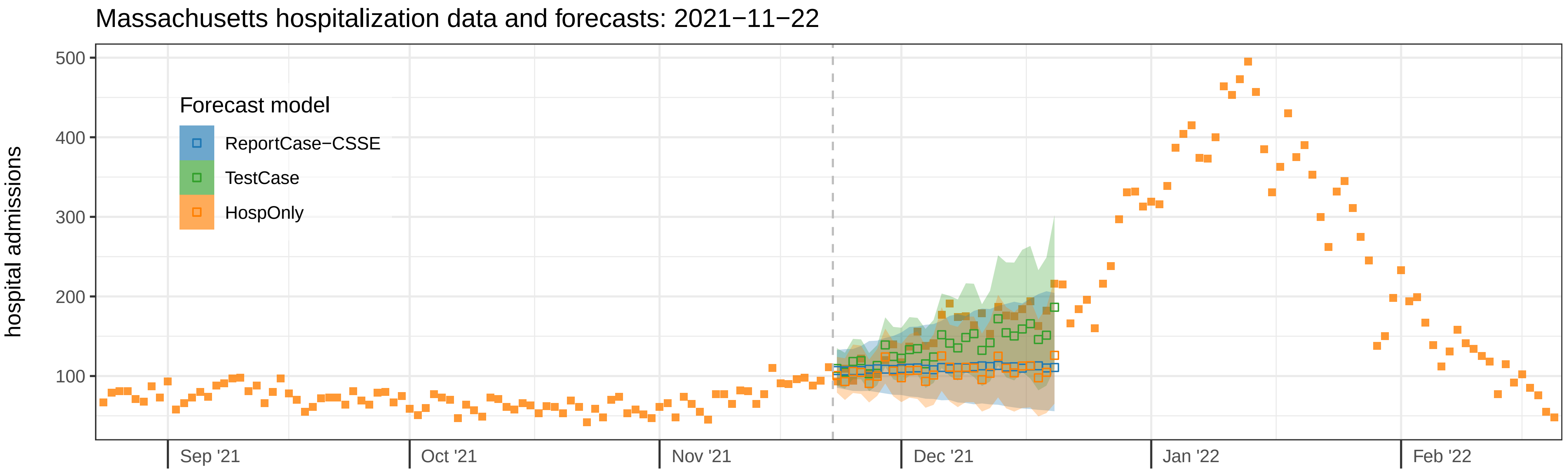
Massachusetts hospitalization data and forecasts: 2021−11−22
hospital admissions
500
400
300
200
100
Forecast model
ReportCase−CSSE
TestCase
HospOnly
Sep '21
Oct '21
Nov '21
Dec '21
Jan '22
Feb '22

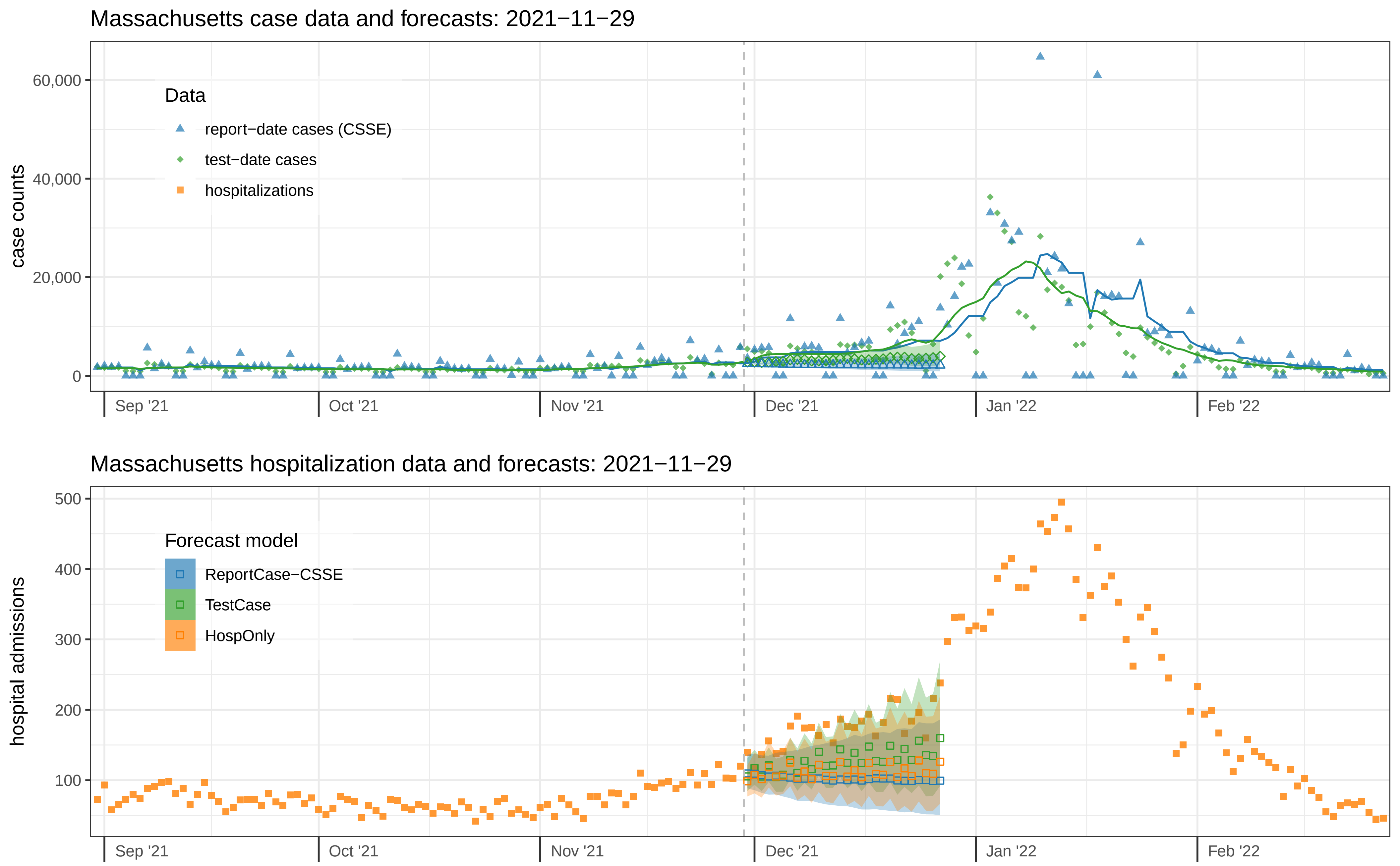

Massachusetts case data and forecasts: 2021−11−29
case counts
60,000
40,000
20,000
0
Data
report−date cases (CSSE)
test−date cases
hospitalizations
Sep '21
Oct '21
Nov '21
Dec '21
Jan '22
Feb '22
Massachusetts hospitalization data and forecasts: 2021−11−29
hospital admissions
500
400
300
200
100
Forecast model
ReportCase−CSSE
TestCase
HospOnly
Sep '21
Oct '21
Nov '21
Dec '21
Jan '22
Feb '22

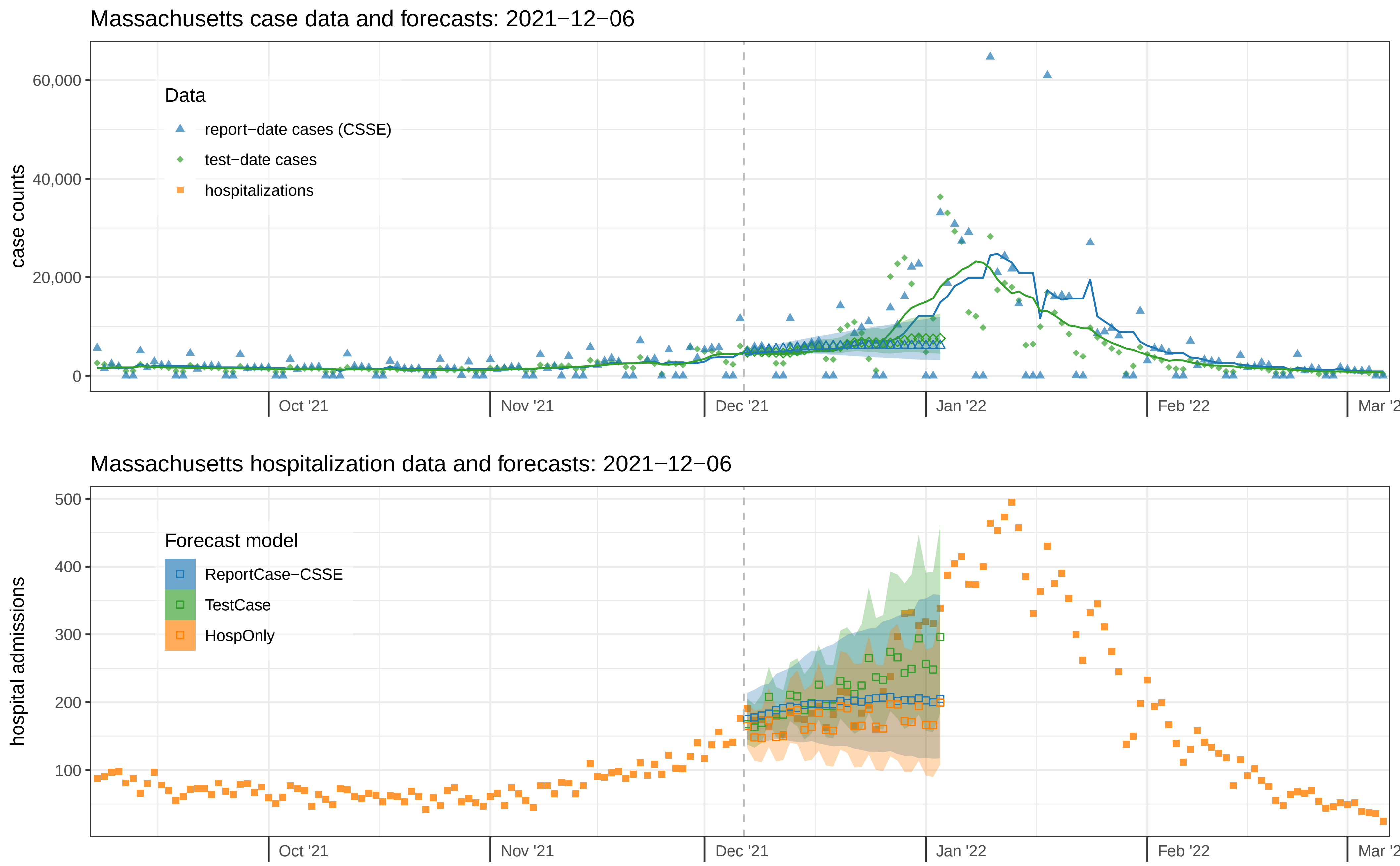

Massachusetts case data and forecasts: 2021−12−06
Data
report−date cases (CSSE)
test−date cases
hospitalizations
case counts
60,000
40,000
20,000
0
Oct '21
Nov '21
Dec '21
Jan '22
Feb '22
Massachusetts hospitalization data and forecasts: 2021−12−06
Forecast model
ReportCase−CSSE
TestCase
HospOnly
hospital admissions
500
400
300
200
100
Oct '21
Nov '21
Dec '21
Jan '22
Feb '22

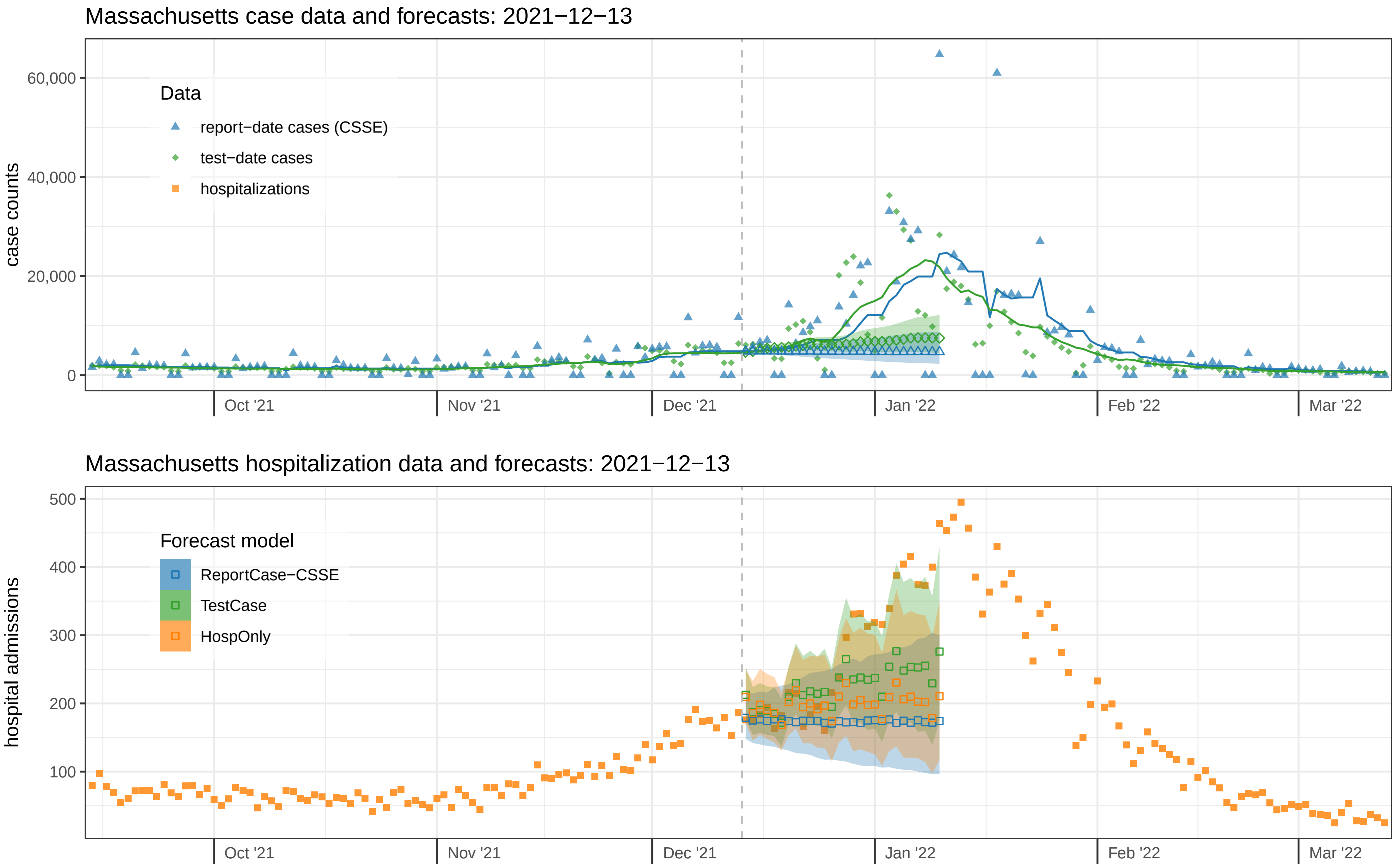

Massachusetts case data and forecasts: 2021−12−13
case counts
60,000
40,000
20,000
0
Data
report−date cases (CSSE)
test−date cases
hospitalizations
Oct '21
Nov '21
Dec '21
Jan '22
Feb '22
Mar '22
Massachusetts hospitalization data and forecasts: 2021−12−13
hospital admissions
500
400
300
200
100
Forecast model
ReportCase−CSSE
TestCase
HospOnly
Oct '21
Nov '21
Dec '21
Jan '22
Feb '22
Mar '22

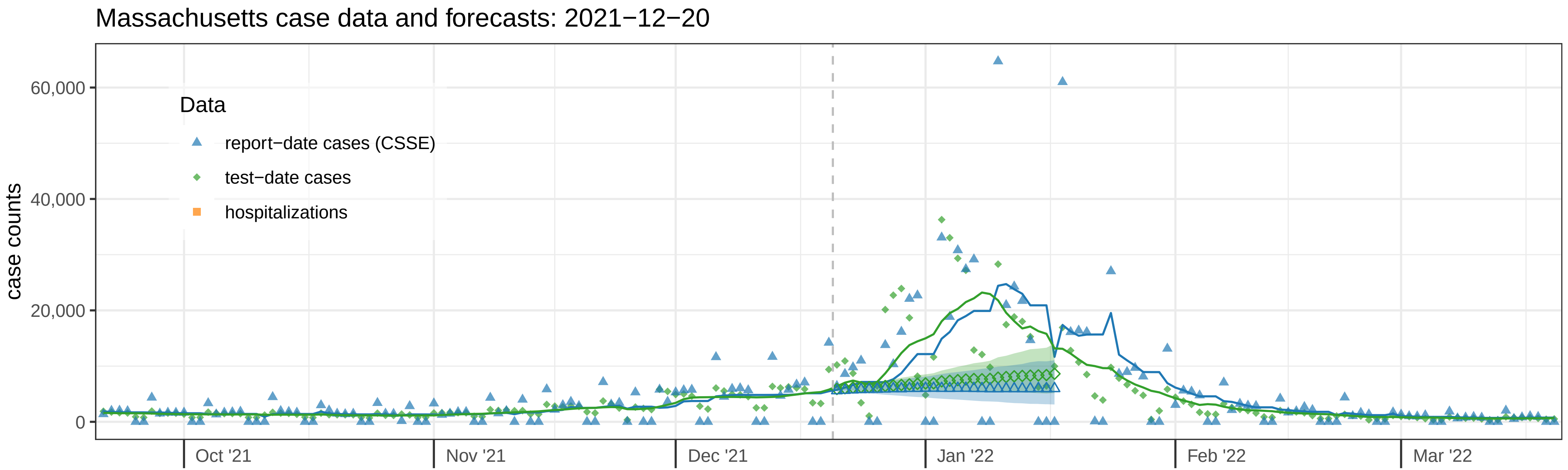

Massachusetts case data and forecasts: 2021−12−20
case counts
60,000
40,000
20,000
0
Data
report−date cases (CSSE)
test−date cases
hospitalizations
Oct '21
Nov '21
Dec '21
Jan '22
Feb '22
Mar '22


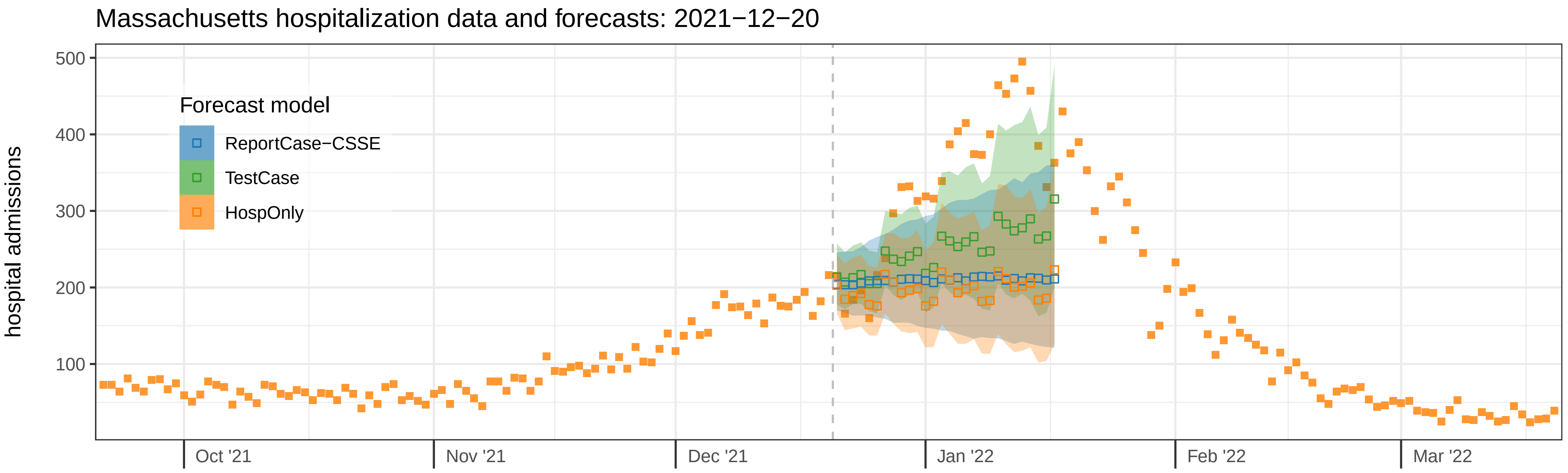

Massachusetts hospitalization data and forecasts: 2021−12−20
hospital admissions
500
400
300
200
100
Forecast model
ReportCase−CSSE
TestCase
HospOnly
Oct '21
Nov '21
Dec '21
Jan '22
Feb '22
Mar '22

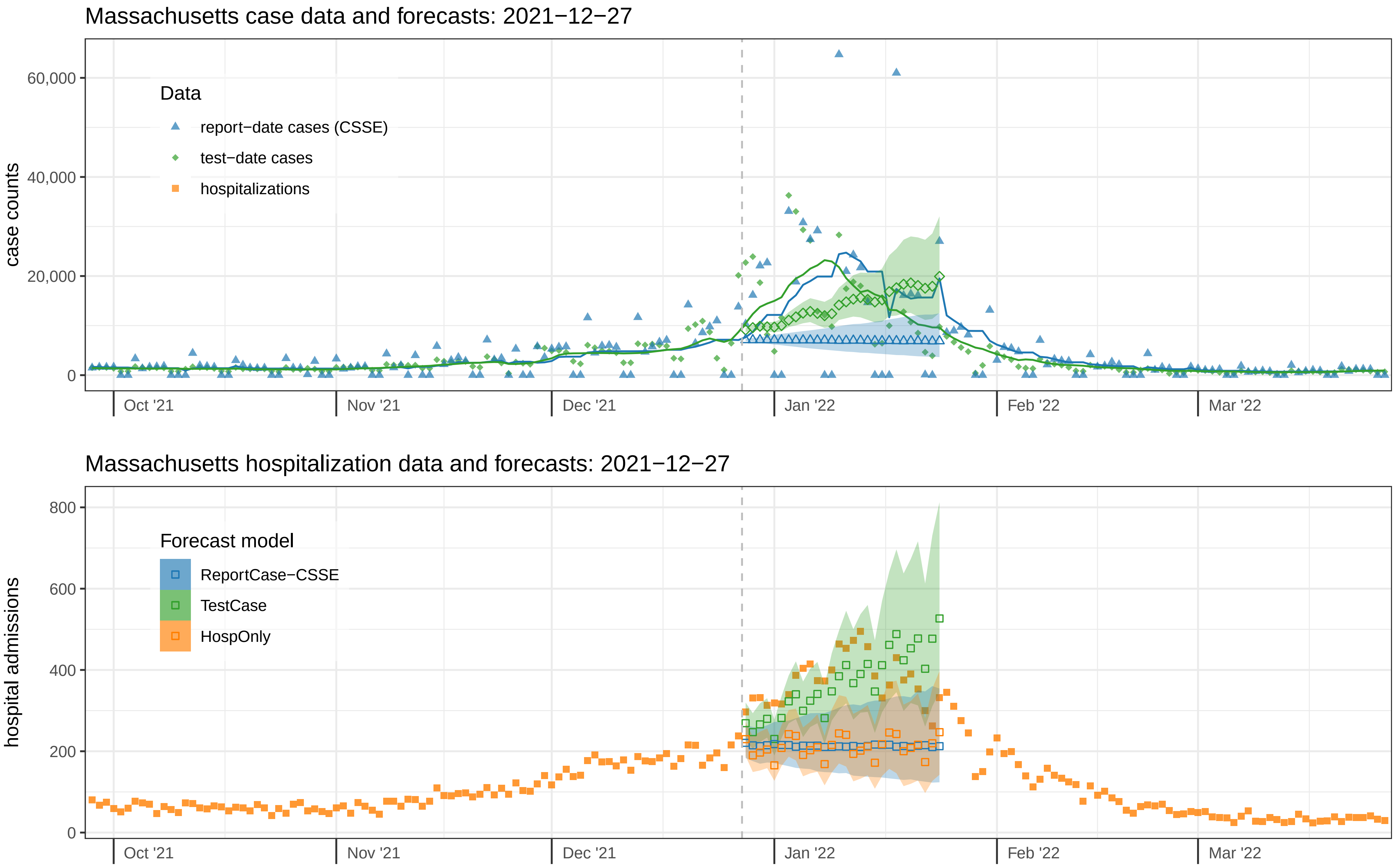

Massachusetts case data and forecasts: 2021−12−27
case counts
60,000
40,000
20,000
0
Data
report−date cases (CSSE)
test−date cases
hospitalizations
Oct '21
Nov '21
Dec '21
Jan '22
Feb '22
Mar '22
Massachusetts hospitalization data and forecasts: 2021−12−27
hospital admissions
800
600
400
200
0
Forecast model
ReportCase−CSSE
TestCase
HospOnly
Oct '21
Nov '21
Dec '21
Jan '22
Feb '22
Mar '22

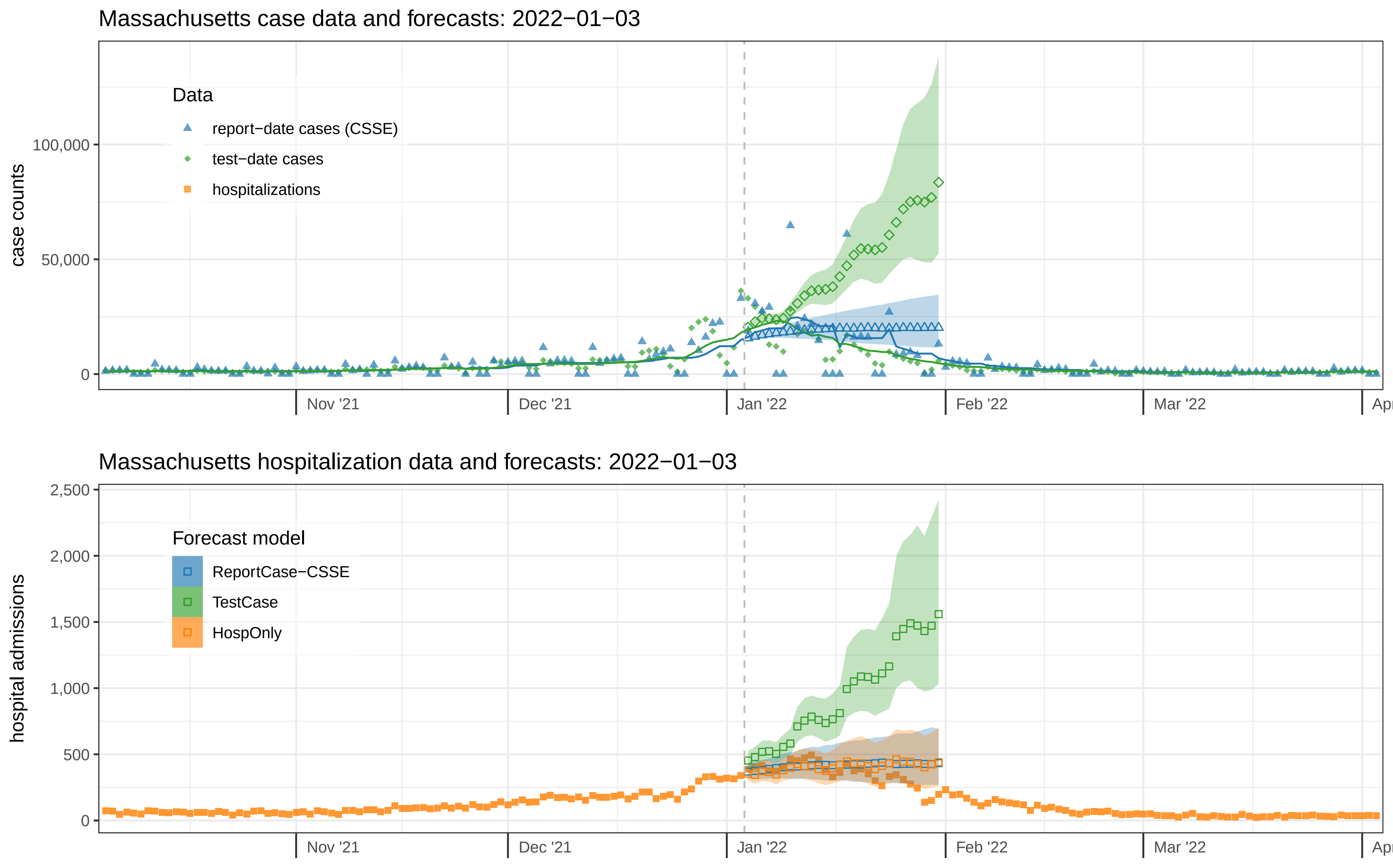
Massachusetts case data and forecasts: 2022−01−03
case counts
100,000
50,000
0
Data
report−date cases (CSSE)
test−date cases
hospitalizations
Nov '21
Dec '21
Jan '22
Feb '22
Mar '22
Apr
Massachusetts hospitalization data and forecasts: 2022−01−03
hospital admissions
2,500
2,000
1,500
1,000
500
0
Forecast model
ReportCase−CSSE
TestCase
HospOnly
Nov '21
Dec '21
Jan '22
Feb '22
Mar '22
Apr

### Massachusetts case data and forecasts: 2022−01−10

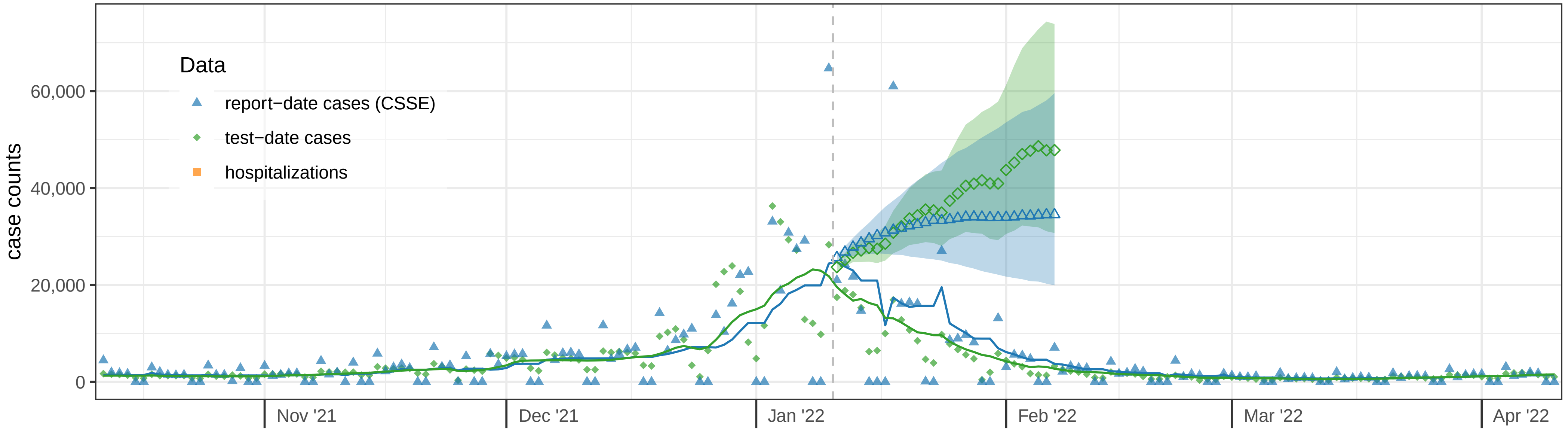


### Massachusetts hospitalization data and forecasts: 2022−01−10

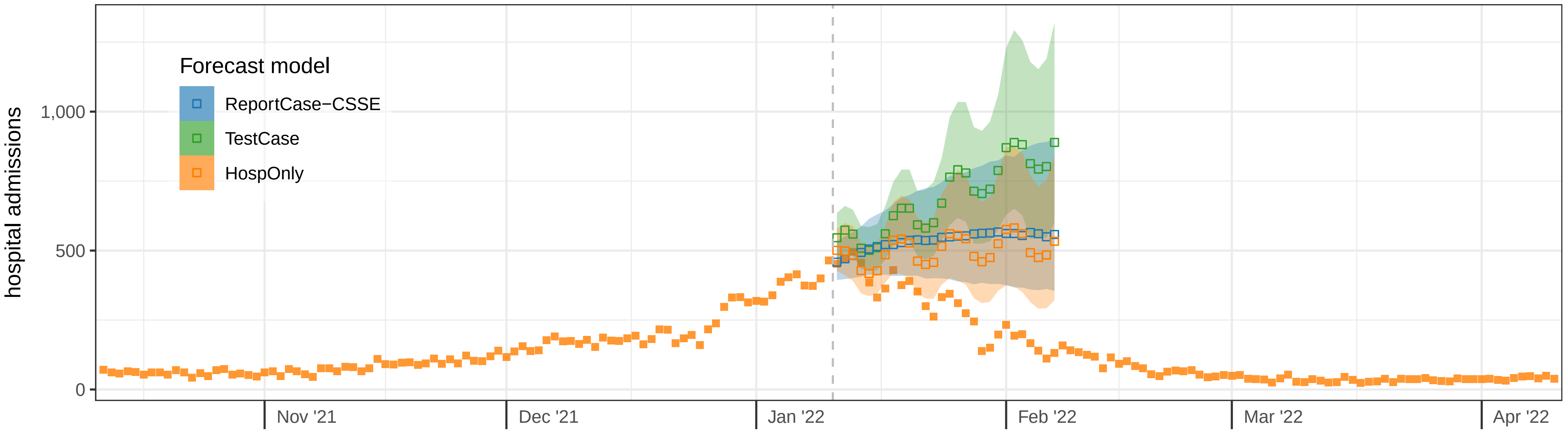

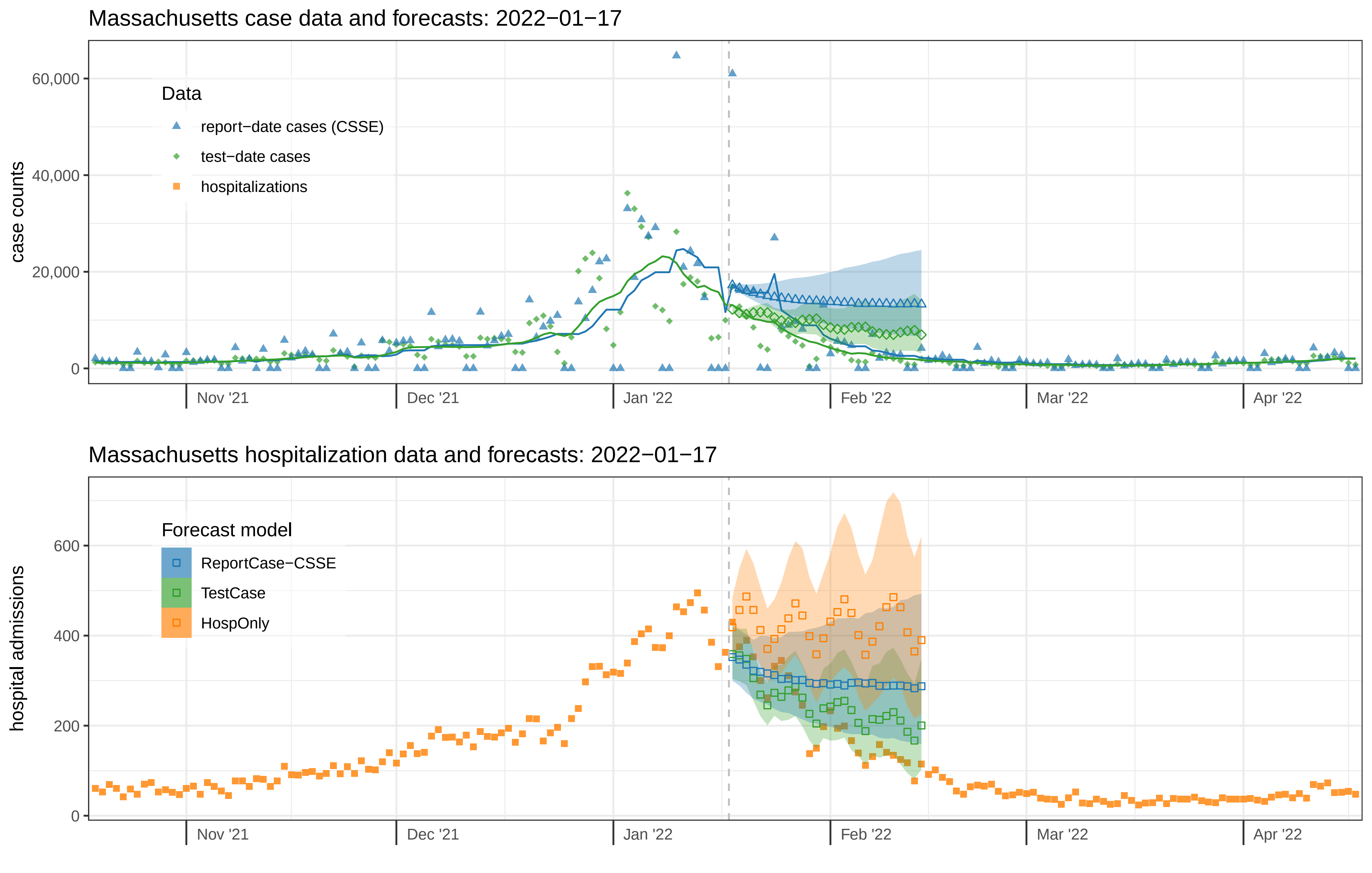
Massachusetts case data and forecasts: 2022−01−17
Data
report−date cases (CSSE)
test−date cases
hospitalizations
case counts
60,000
40,000
20,000
0
Nov '21
Dec '21
Jan '22
Feb '22
Mar '22
Apr '22
Massachusetts hospitalization data and forecasts: 2022−01−17
Forecast model
ReportCase−CSSE
TestCase
HospOnly
hospital admissions
600
400
200
0
Nov '21
Dec '21
Jan '22
Feb '22
Mar '22
Apr '22

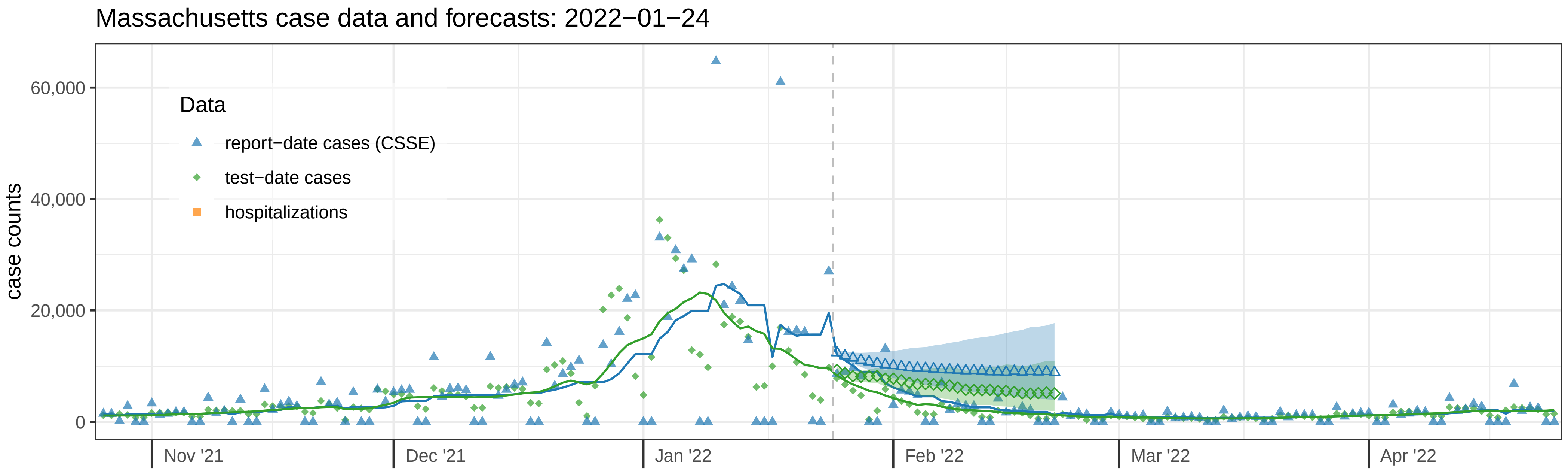

Massachusetts case data and forecasts: 2022−01−24
case counts
60,000
40,000
20,000
0
Data
report−date cases (CSSE)
test−date cases
hospitalizations
Nov '21
Dec '21
Jan '22
Feb '22
Mar '22
Apr '22


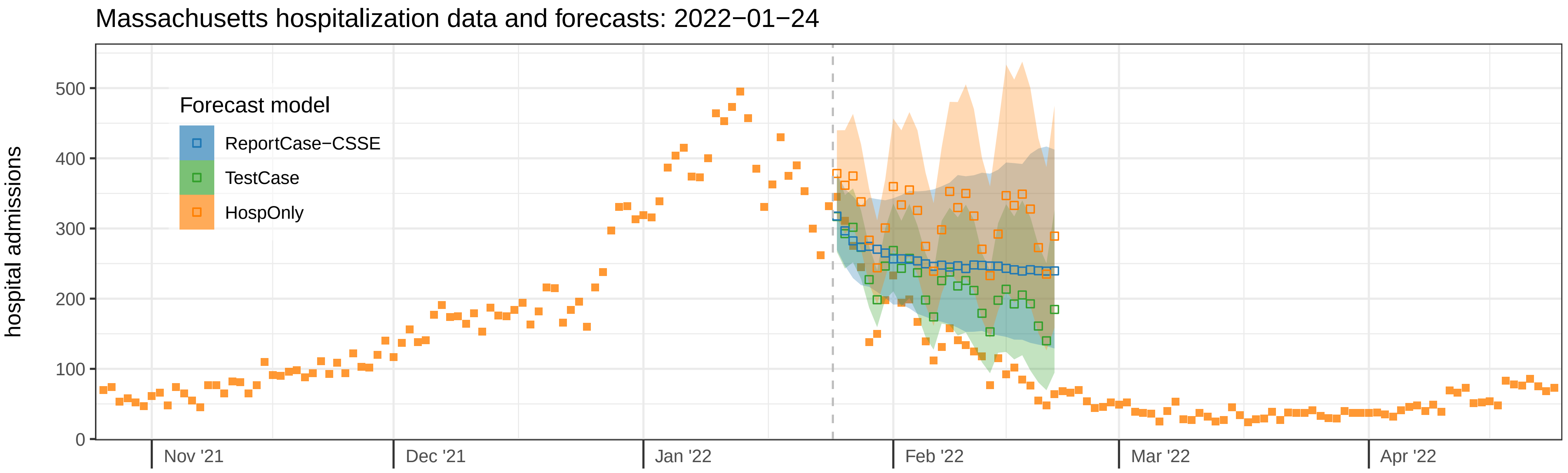

Massachusetts hospitalization data and forecasts: 2022−01−24
hospital admissions
500
400
300
200
100
0
Forecast model
ReportCase−CSSE
TestCase
HospOnly
Nov '21
Dec '21
Jan '22
Feb '22
Mar '22
Apr '22

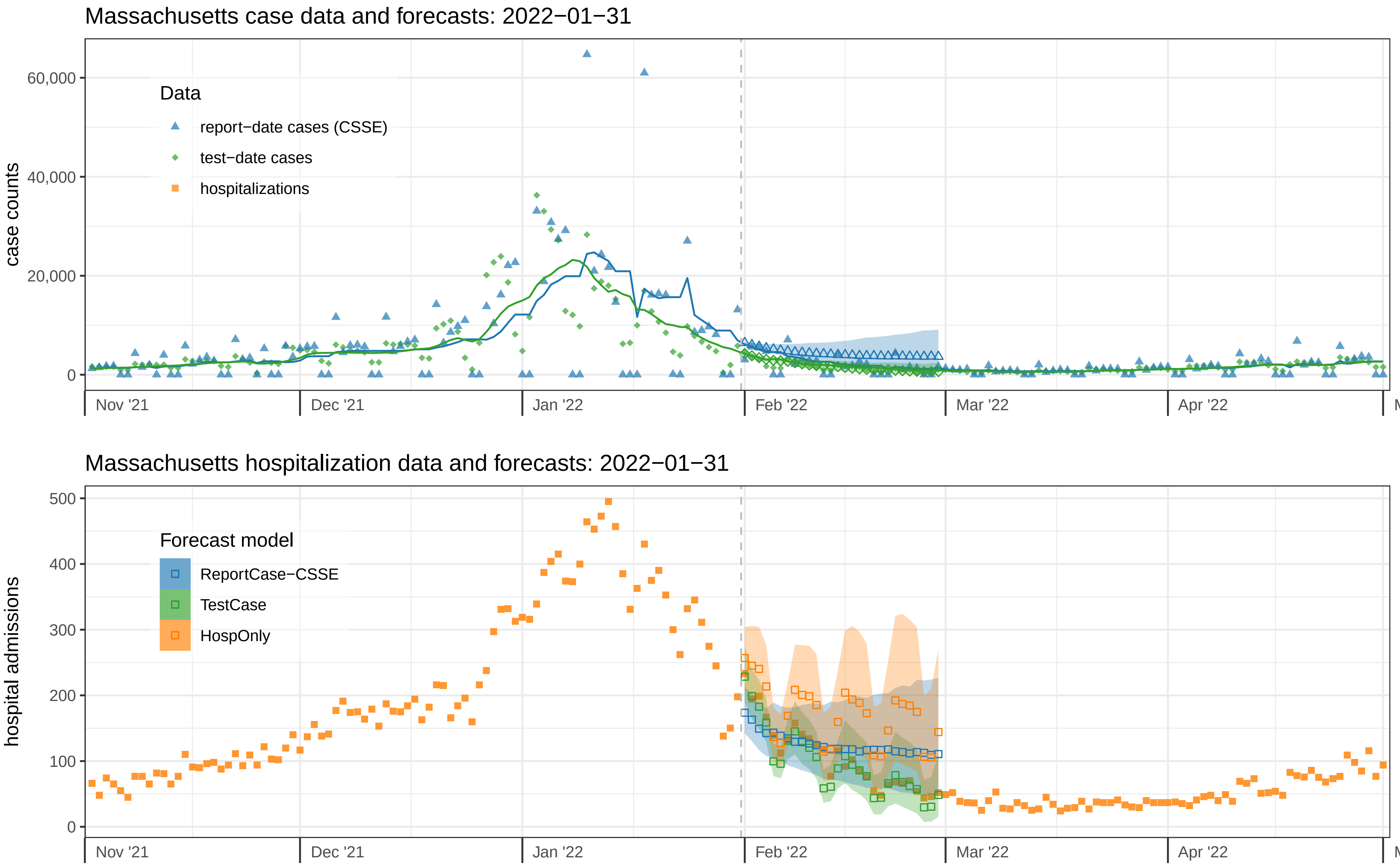
Massachusetts case data and forecasts: 2022−01−31
case counts
60,000
40,000
20,000
0
Data
report−date cases (CSSE)
test−date cases
hospitalizations
Nov '21
Dec '21
Jan '22
Feb '22
Mar '22
Apr '22
Massachusetts hospitalization data and forecasts: 2022−01−31
hospital admissions
500
400
300
200
100
0
Forecast model
ReportCase−CSSE
TestCase
HospOnly
Nov '21
Dec '21
Jan '22
Feb '22
Mar '22
Apr '22

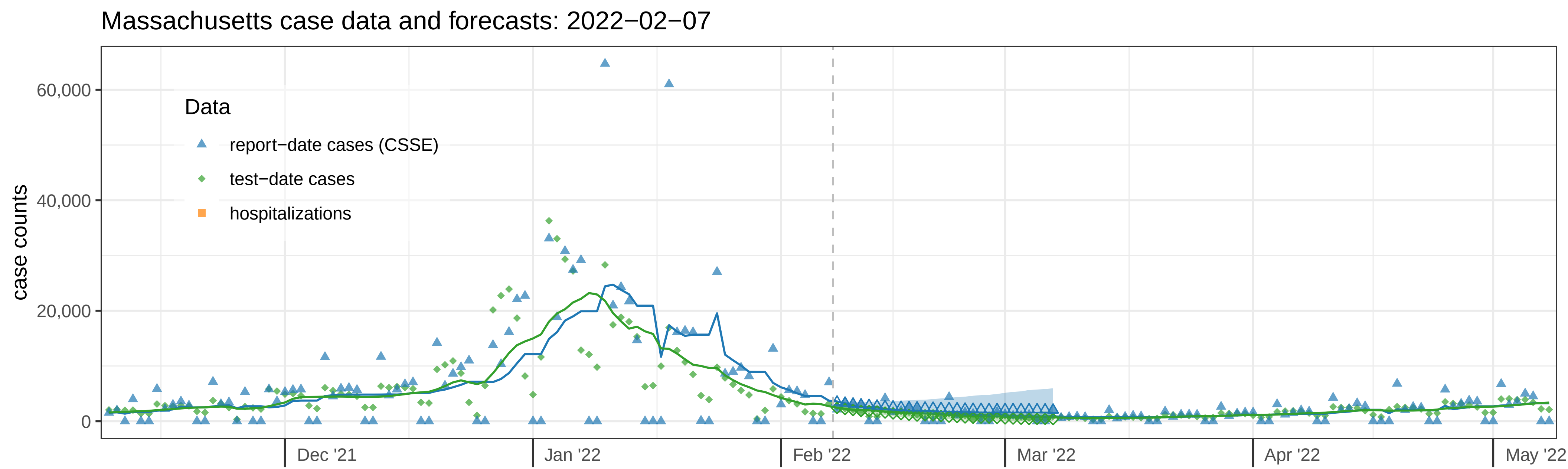
Massachusetts case data and forecasts: 2022−02−07
case counts
60,000
40,000
20,000
0
Data
report−date cases (CSSE)
test−date cases
hospitalizations
Dec '21
Jan '22
Feb '22
Mar '22
Apr '22
May '22

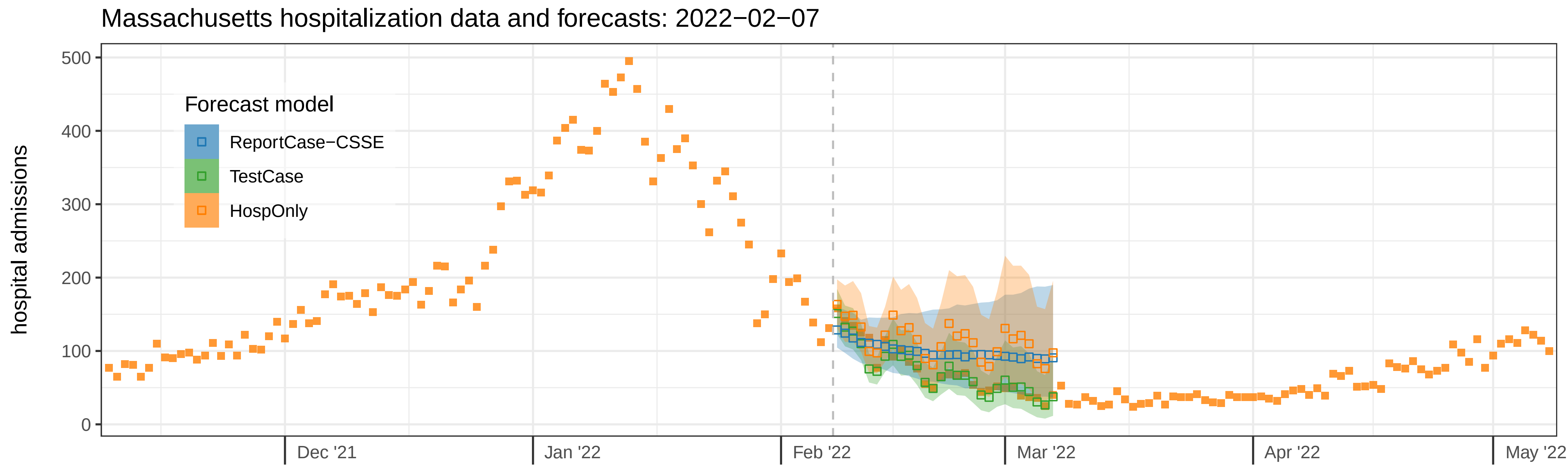
Massachusetts hospitalization data and forecasts: 2022−02−07
hospital admissions
500
400
300
200
100
0
Forecast model
ReportCase−CSSE
TestCase
HospOnly
Dec '21
Jan '22
Feb '22
Mar '22
Apr '22
May '22

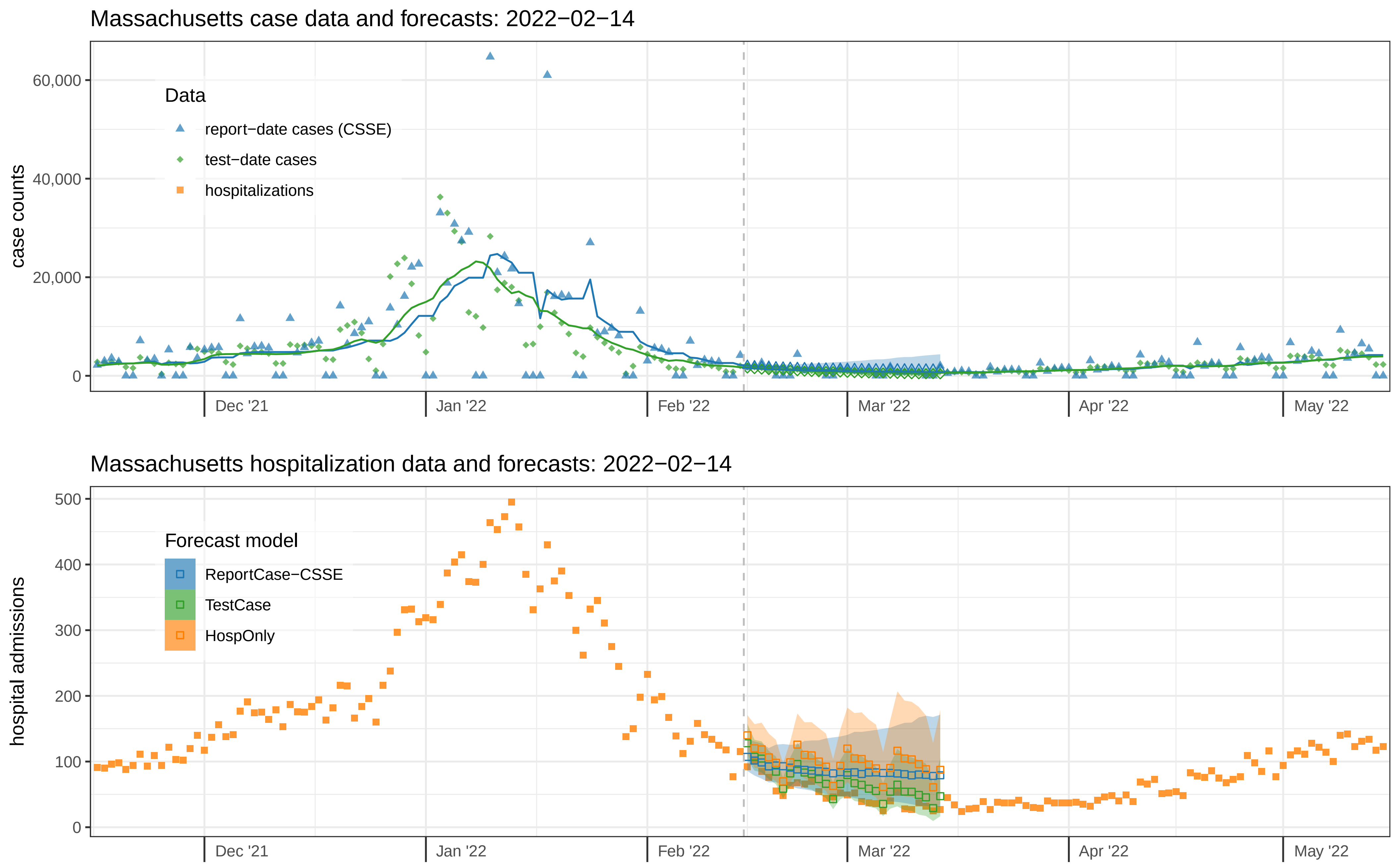

Massachusetts case data and forecasts: 2022−02−14
case counts
60,000
40,000
20,000
0
Data
report−date cases (CSSE)
test−date cases
hospitalizations
Dec '21
Jan '22
Feb '22
Mar '22
Apr '22
May '22
Massachusetts hospitalization data and forecasts: 2022−02−14
hospital admissions
500
400
300
200
100
0
Forecast model
ReportCase−CSSE
TestCase
HospOnly
Dec '21
Jan '22
Feb '22
Mar '22
Apr '22
May '22

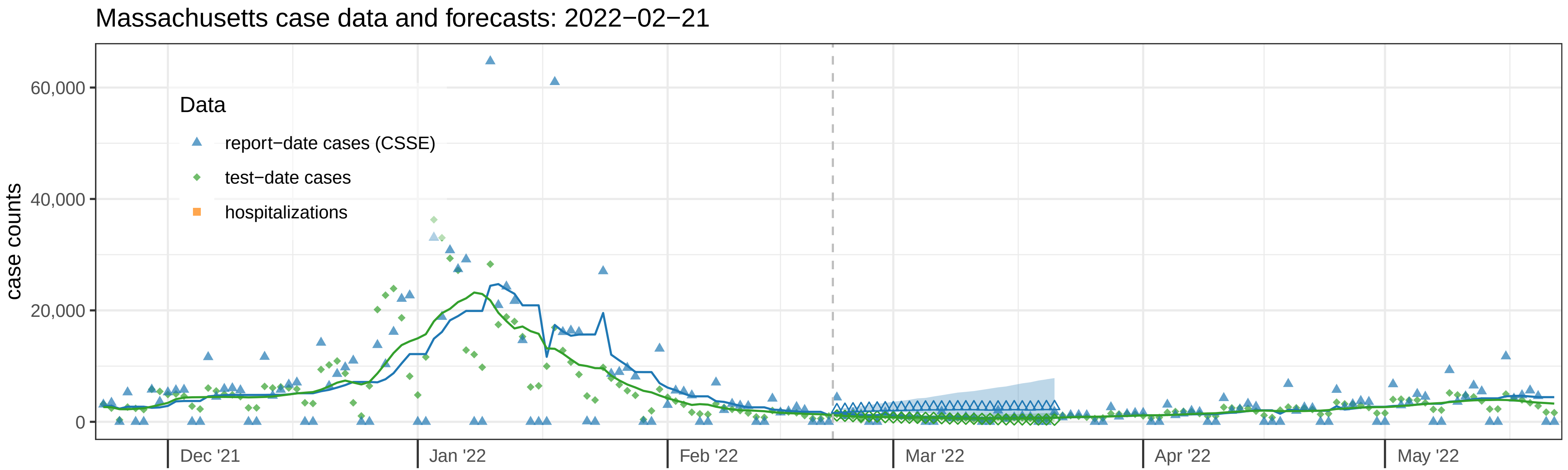
Massachusetts case data and forecasts: 2022−02−21
case counts
60,000
40,000
20,000
0
Data
report−date cases (CSSE)
test−date cases
hospitalizations
Dec '21
Jan '22
Feb '22
Mar '22
Apr '22
May '22

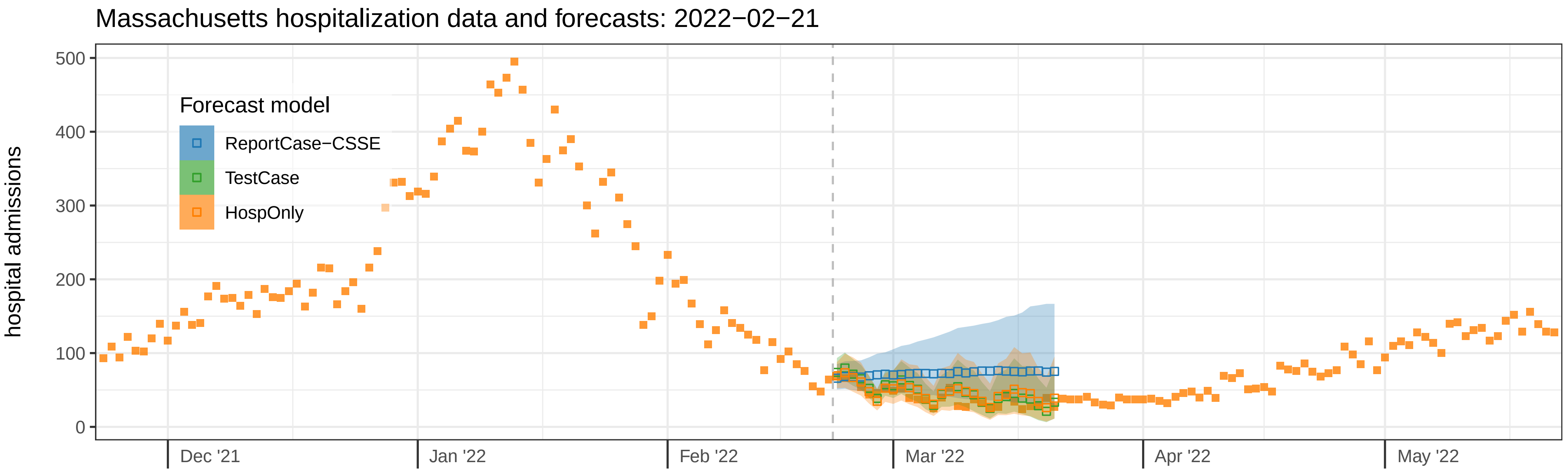
Massachusetts hospitalization data and forecasts: 2022−02−21
hospital admissions
500
400
300
200
100
0
Forecast model
ReportCase−CSSE
TestCase
HospOnly
Dec '21
Jan '22
Feb '22
Mar '22
Apr '22
May '22

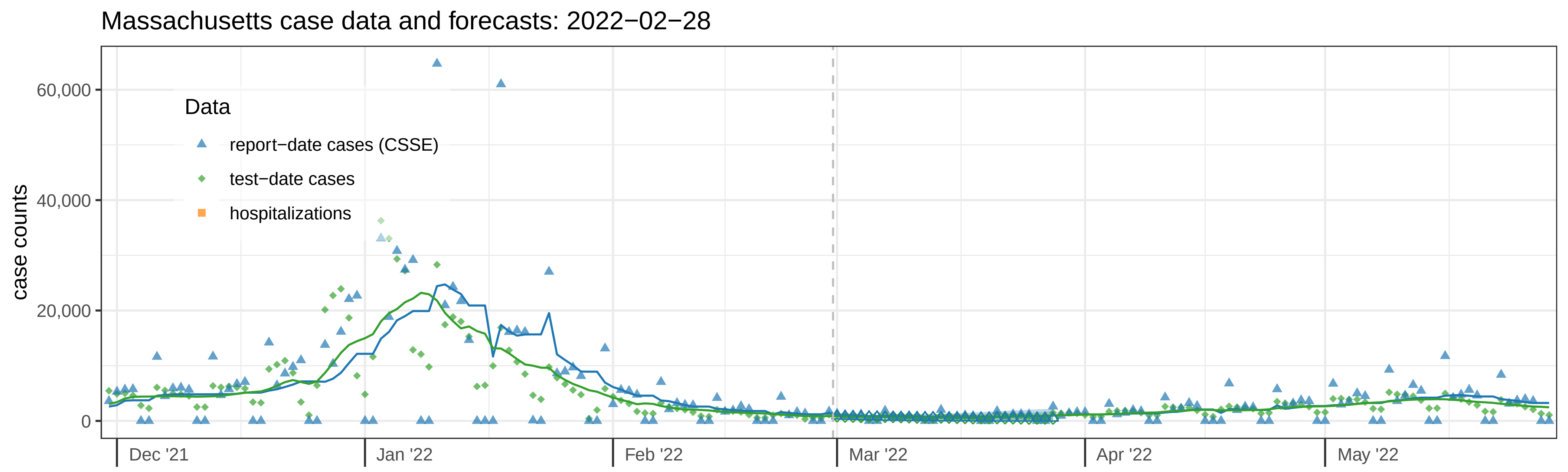
Massachusetts case data and forecasts: 2022−02−28
case counts
60,000
40,000
20,000
0
Data
report−date cases (CSSE)
test−date cases
hospitalizations
Dec '21
Jan '22
Feb '22
Mar '22
Apr '22
May '22

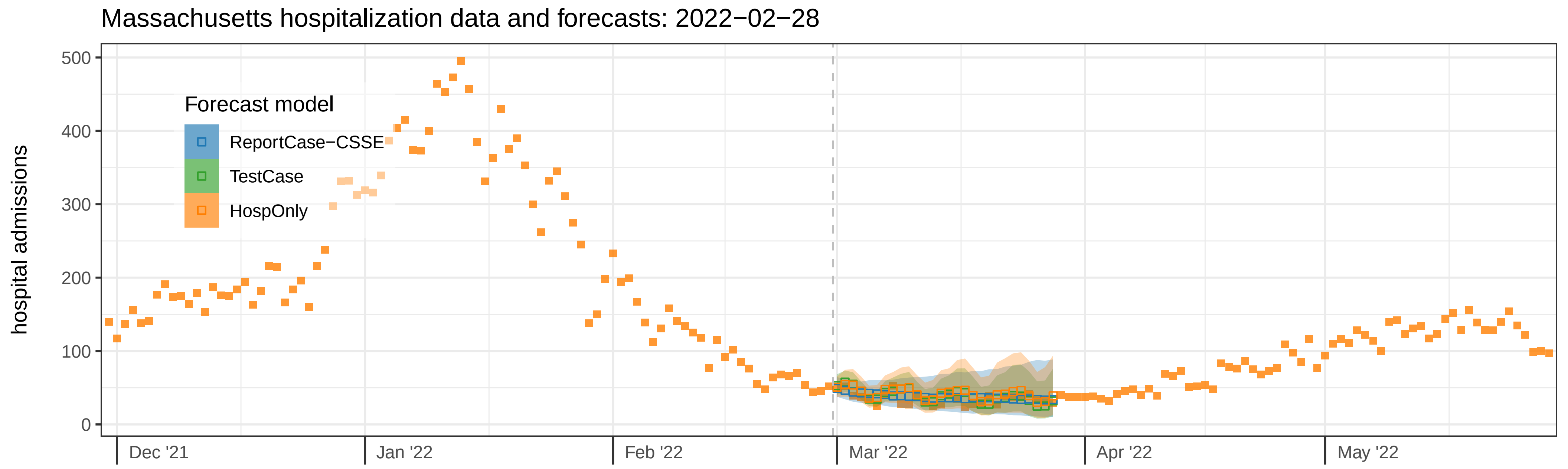
Massachusetts hospitalization data and forecasts: 2022−02−28
hospital admissions
500
400
300
200
100
0
Forecast model
ReportCase−CSSE
TestCase
HospOnly
Dec '21
Jan '22
Feb '22
Mar '22
Apr '22
May '22

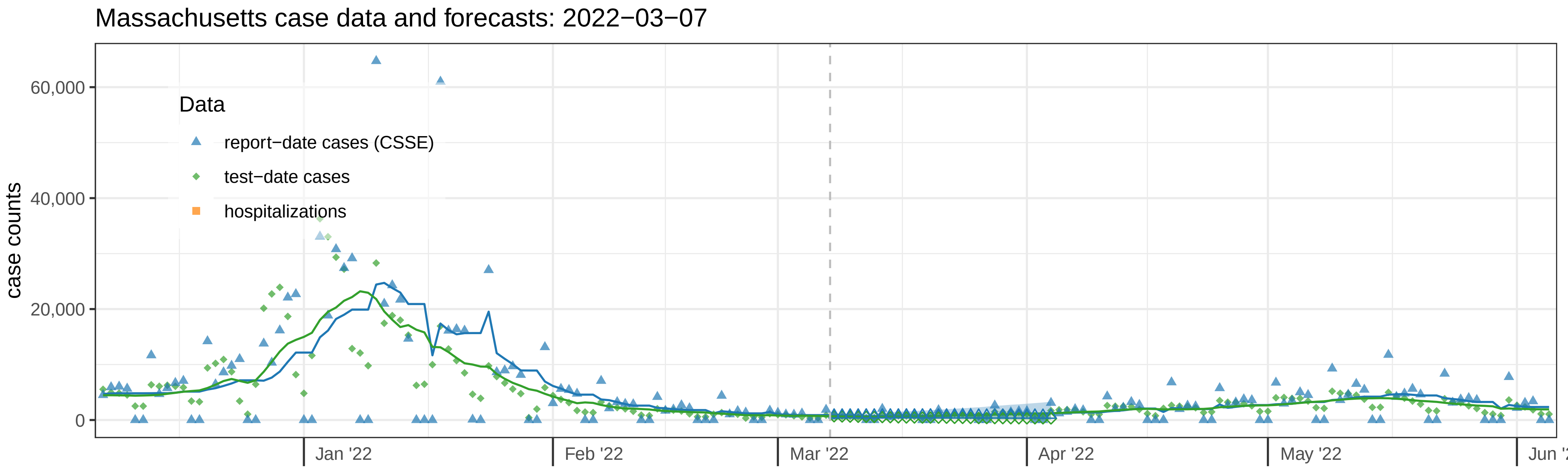

Massachusetts case data and forecasts: 2022−03−07
case counts
60,000
40,000
20,000
0
Data
report−date cases (CSSE)
test−date cases
hospitalizations
Jan '22
Feb '22
Mar '22
Apr '22
May '22


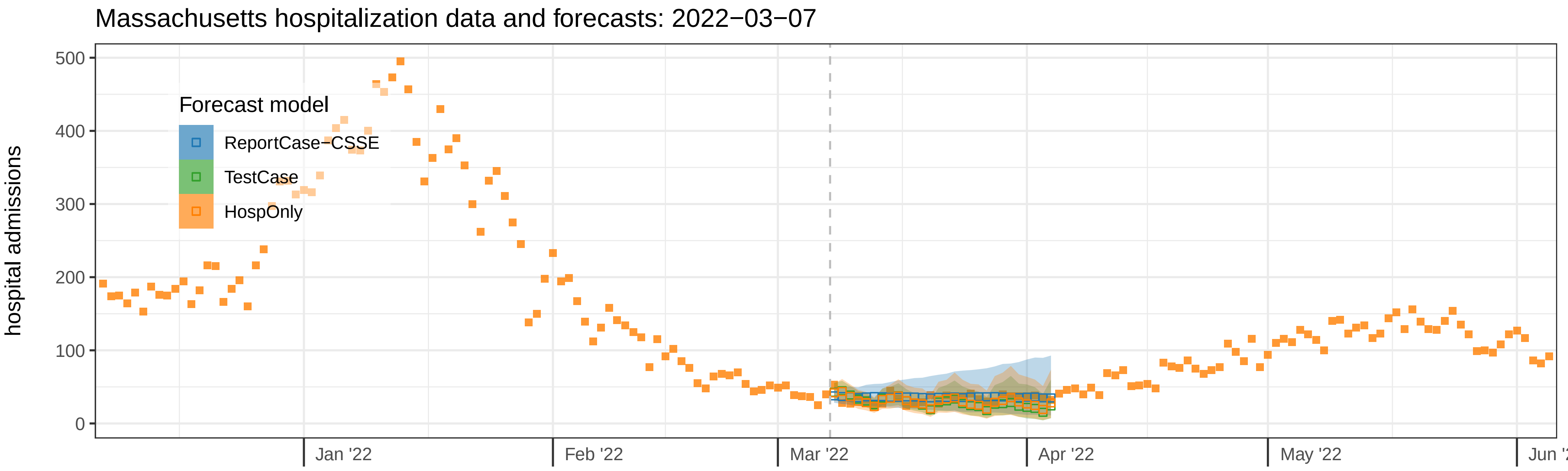

Massachusetts hospitalization data and forecasts: 2022−03−07
hospital admissions
500
400
300
200
100
0
Forecast model
ReportCase−CSSE
TestCase
HospOnly
Jan '22
Feb '22
Mar '22
Apr '22
May '22

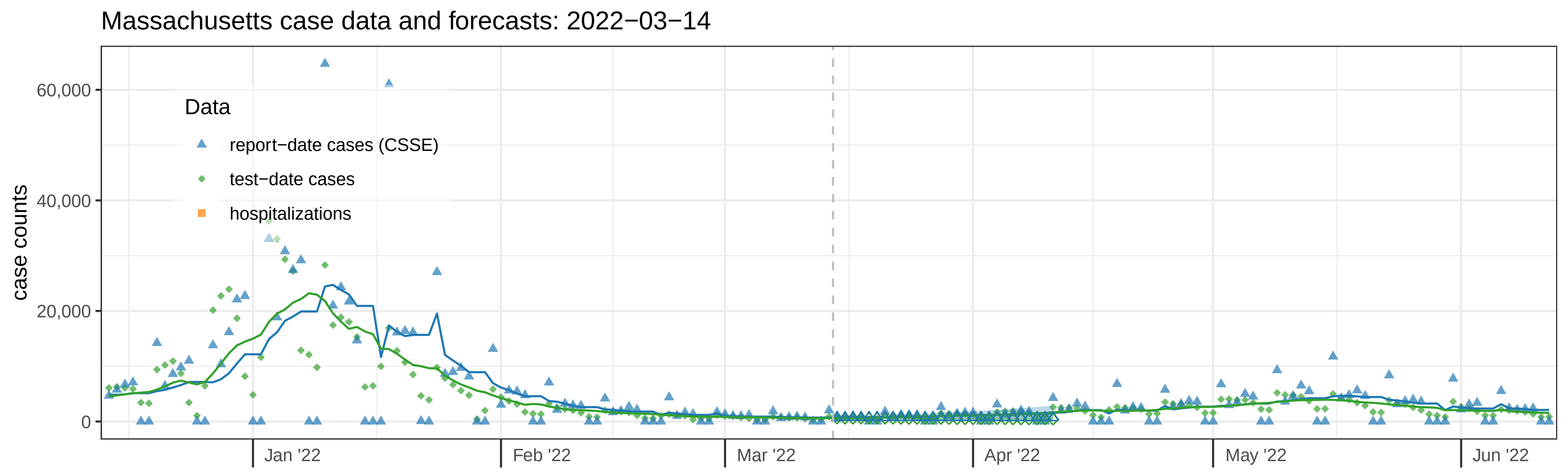

Massachusetts case data and forecasts: 2022−03−14
case counts
60,000
40,000
20,000
0
Data
report−date cases (CSSE)
test−date cases
hospitalizations
Jan '22
Feb '22
Mar '22
Apr '22
May '22
Jun '22


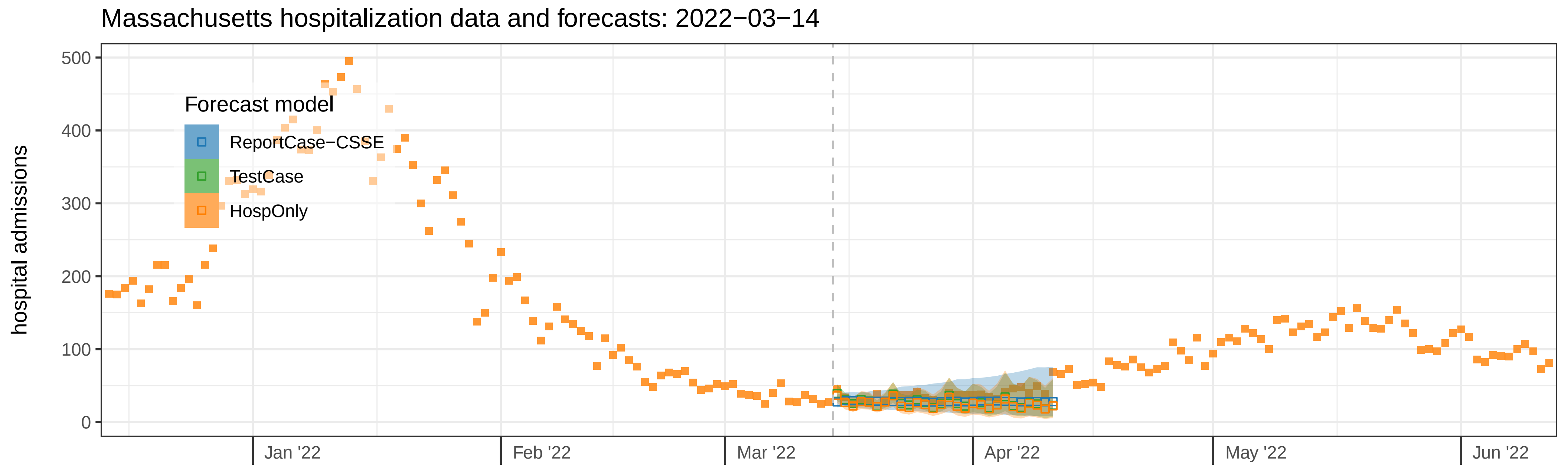

Massachusetts hospitalization data and forecasts: 2022−03−14
hospital admissions
500
400
300
200
100
0
Forecast model
ReportCase−CSSE
TestCase
HospOnly
Jan '22
Feb '22
Mar '22
Apr '22
May '22
Jun '22

Massachusetts case data and forecasts: 2022−03−21

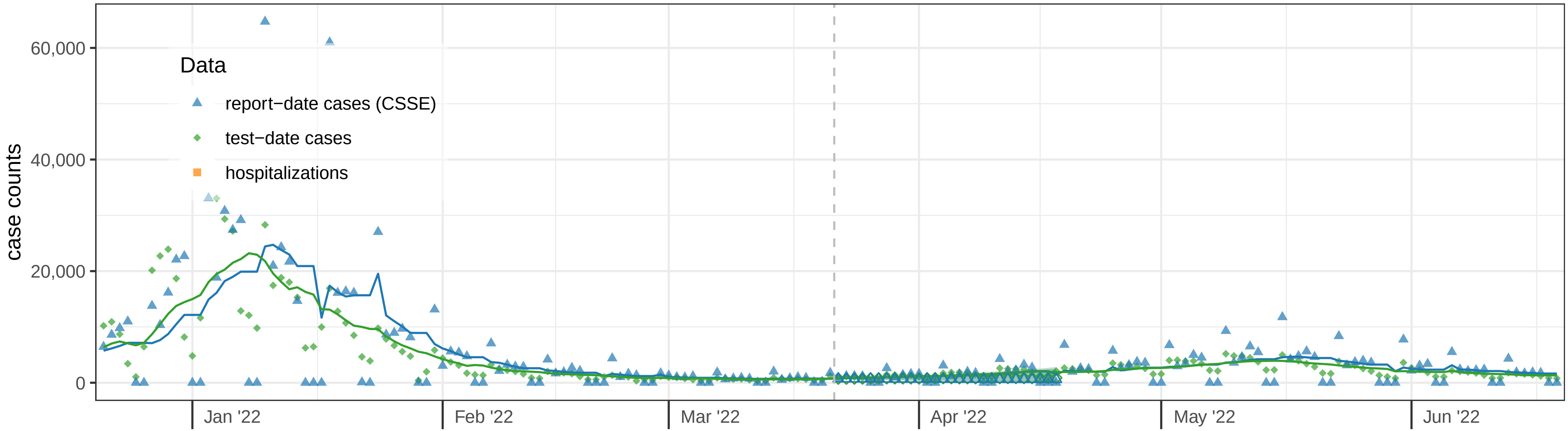


Massachusetts hospitalization data and forecasts: 2022−03−21

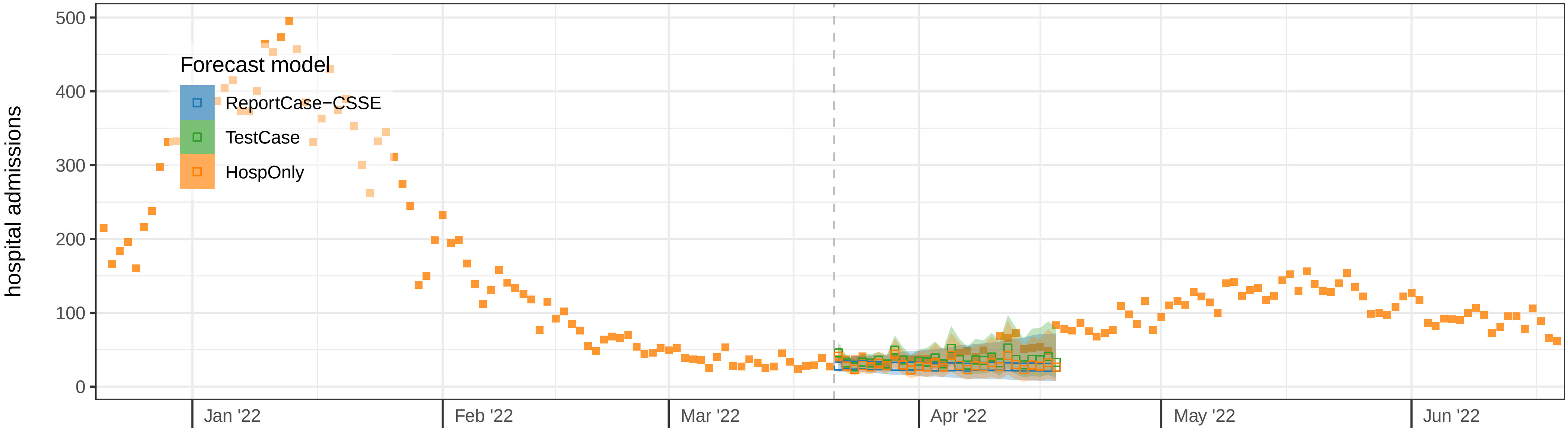

Massachusetts case data and forecasts: 2022−03−28

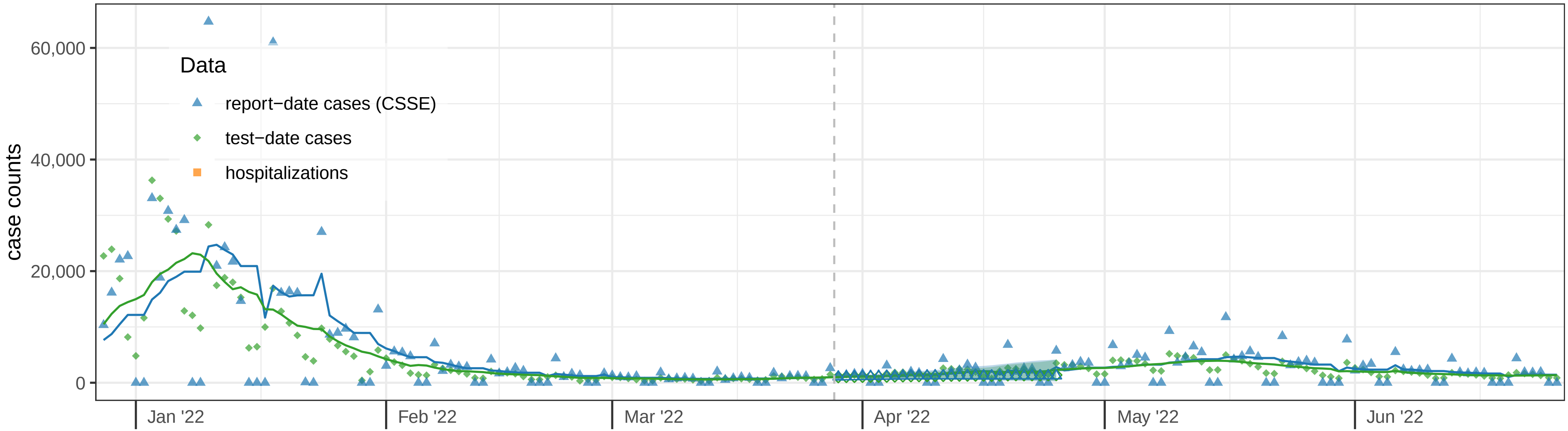


Massachusetts hospitalization data and forecasts: 2022−03−28

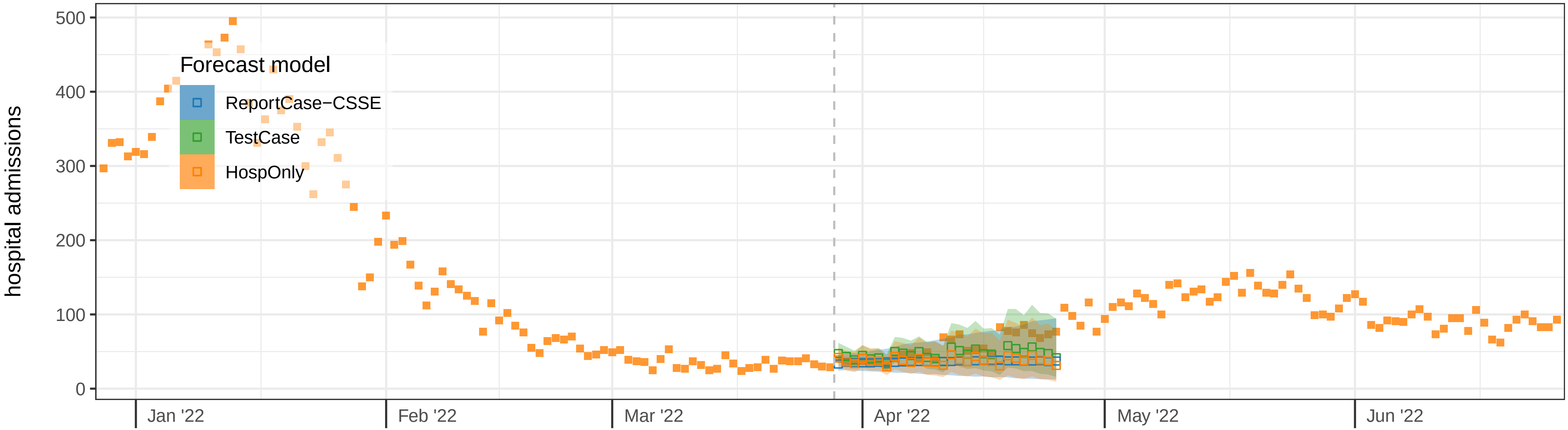

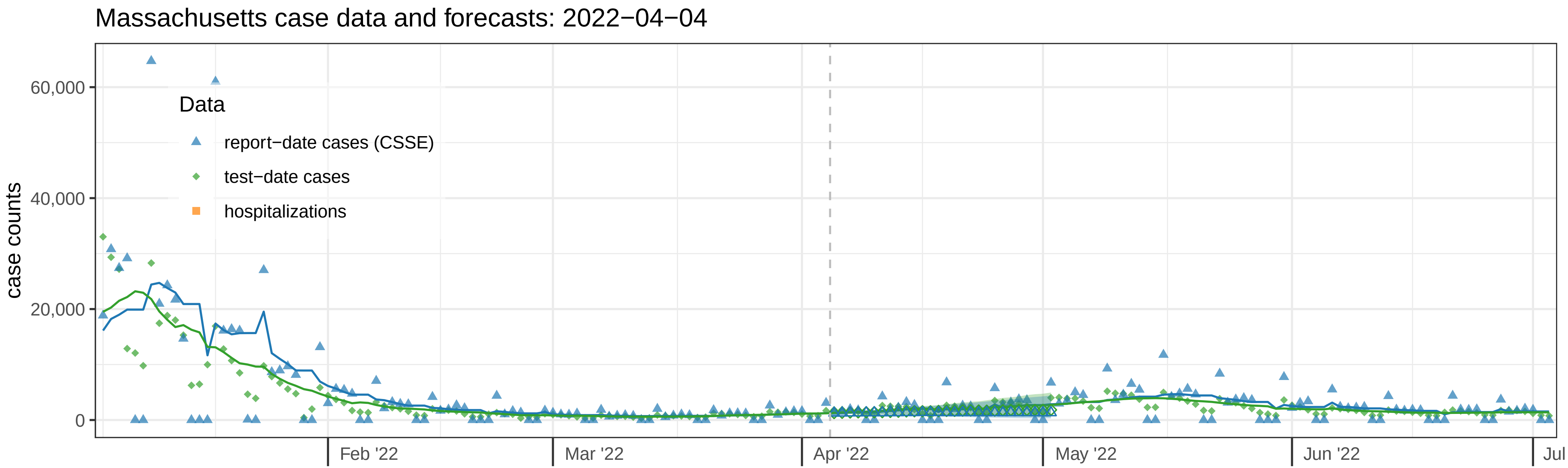
Massachusetts case data and forecasts: 2022−04−04
case counts
60,000
40,000
20,000
0
Data
report−date cases (CSSE)
test−date cases
hospitalizations
Feb '22
Mar '22
Apr '22
May '22
Jun '22
Jul

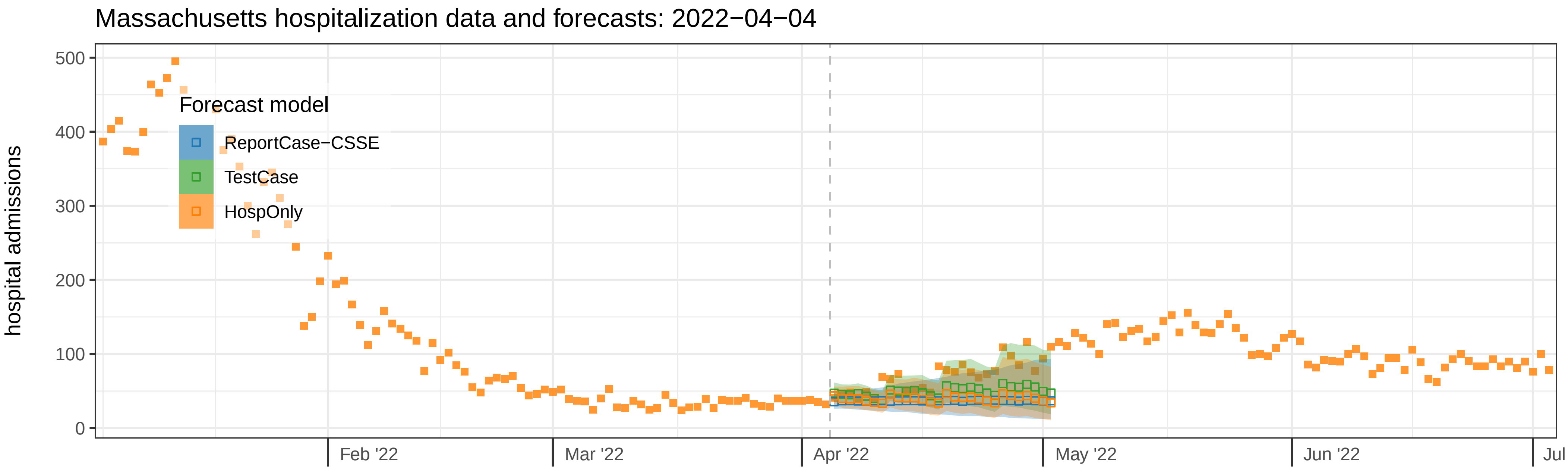
Massachusetts hospitalization data and forecasts: 2022−04−04
hospital admissions
500
400
300
200
100
0
Forecast model
ReportCase−CSSE
TestCase
HospOnly
Feb '22
Mar '22
Apr '22
May '22
Jun '22
Jul

Massachusetts case data and forecasts: 2022−04−11

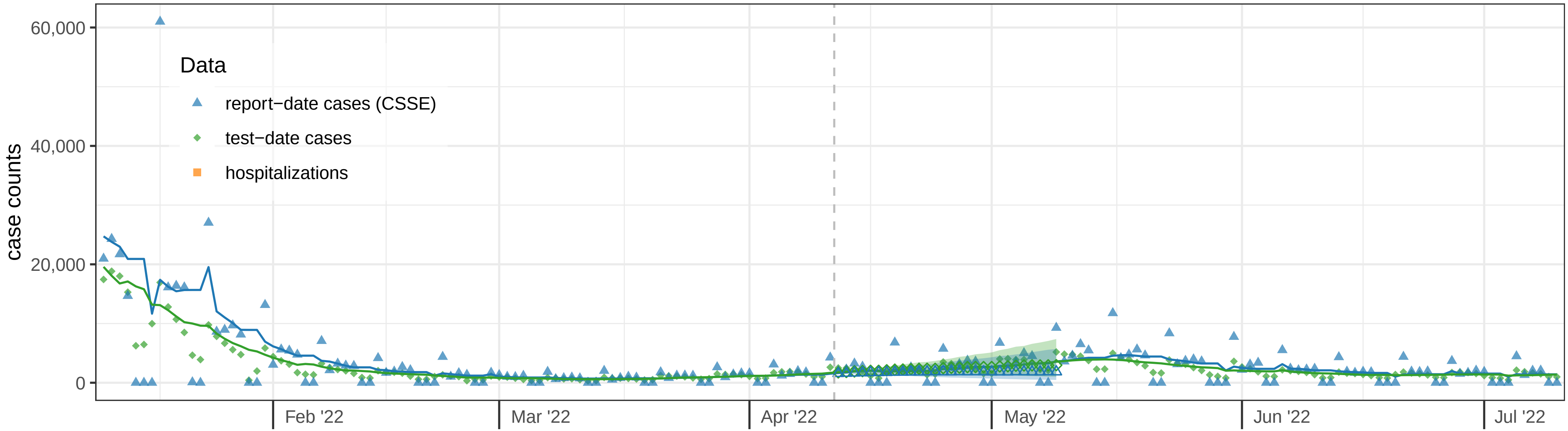


Massachusetts hospitalization data and forecasts: 2022−04−11

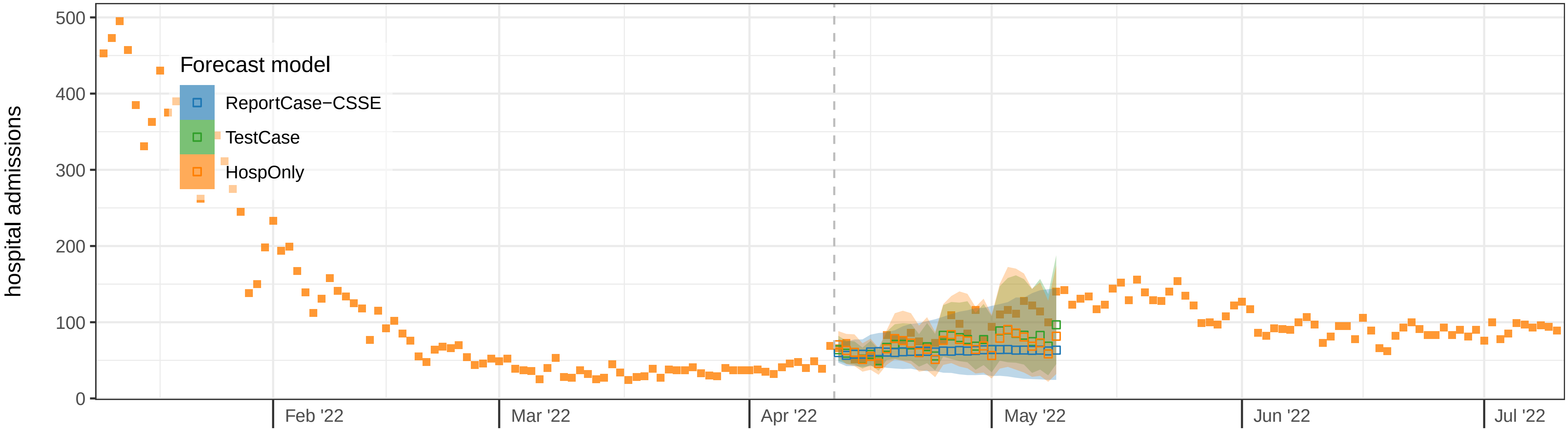

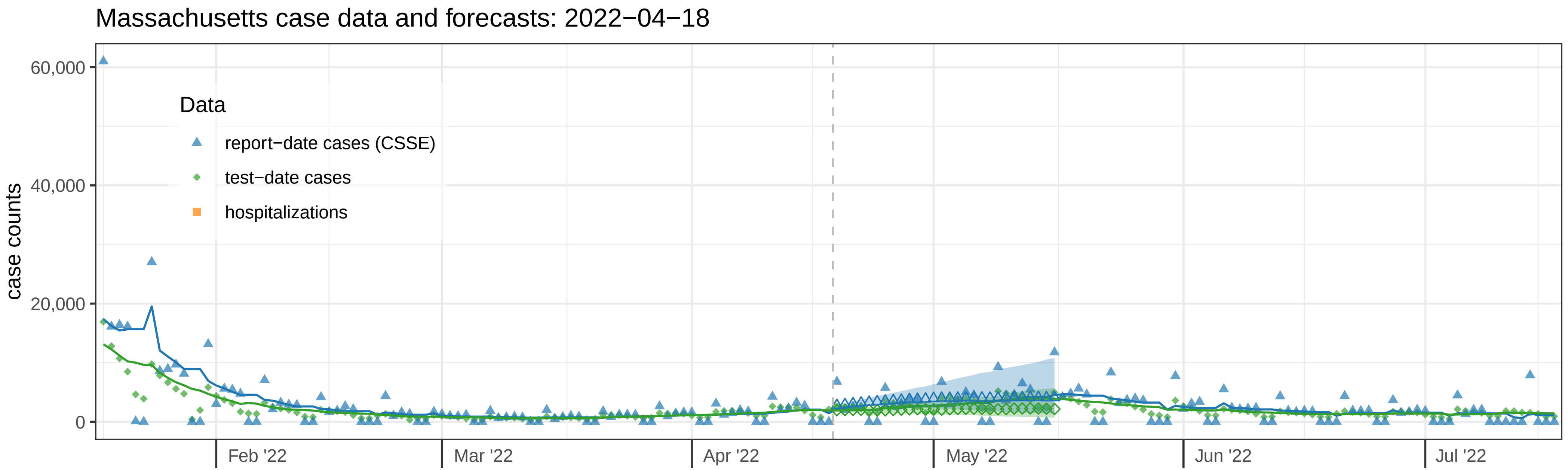
Massachusetts case data and forecasts: 2022−04−18
case counts
60,000
40,000
20,000
0
Data
report−date cases (CSSE)
test−date cases
hospitalizations
Feb '22
Mar '22
Apr '22
May '22
Jun '22
Jul '22

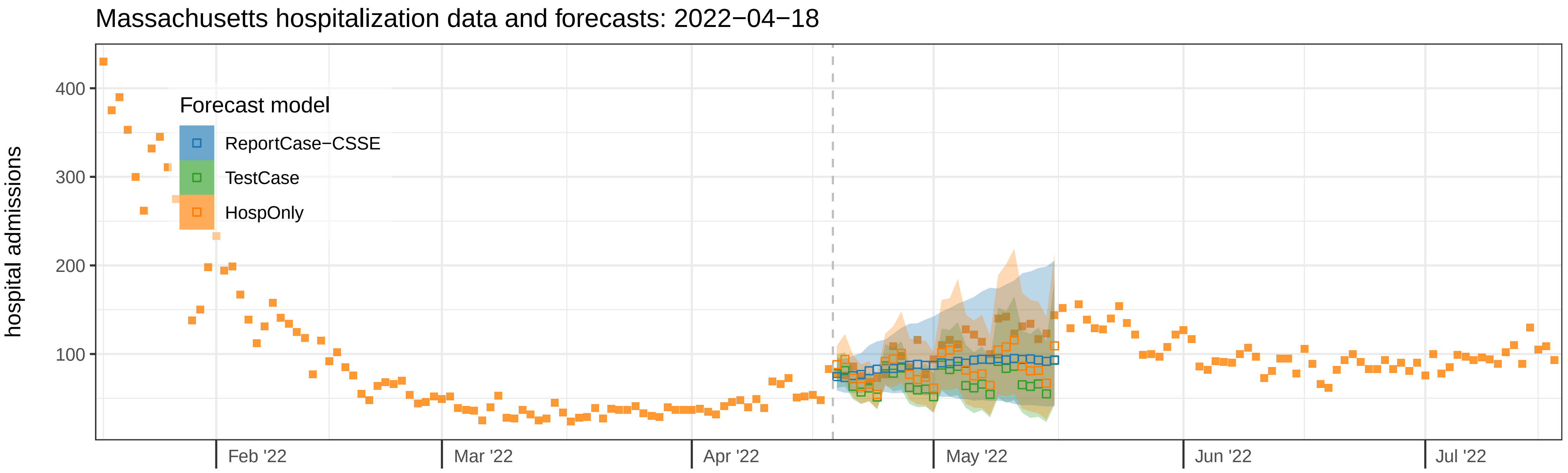
Massachusetts hospitalization data and forecasts: 2022−04−18
hospital admissions
400
300
200
100
Forecast model
ReportCase−CSSE
TestCase
HospOnly
Feb '22
Mar '22
Apr '22
May '22
Jun '22
Jul '22

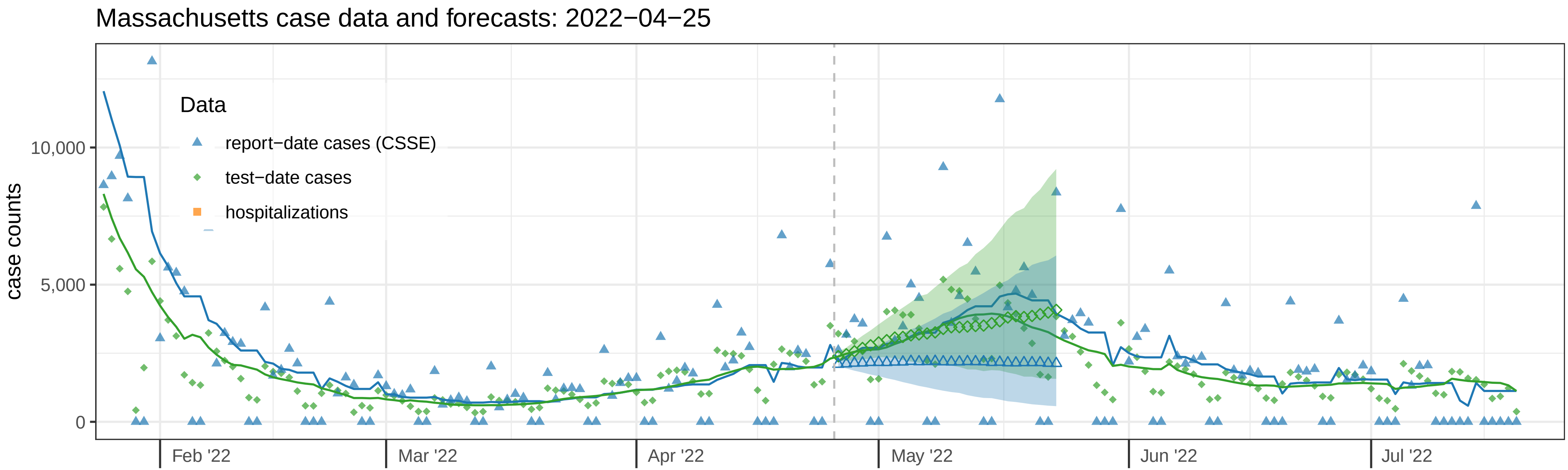
Massachusetts case data and forecasts: 2022−04−25
Data
report−date cases (CSSE)
test−date cases
hospitalizations
case counts
10,000
5,000
0
Feb '22
Mar '22
Apr '22
May '22
Jun '22
Jul '22

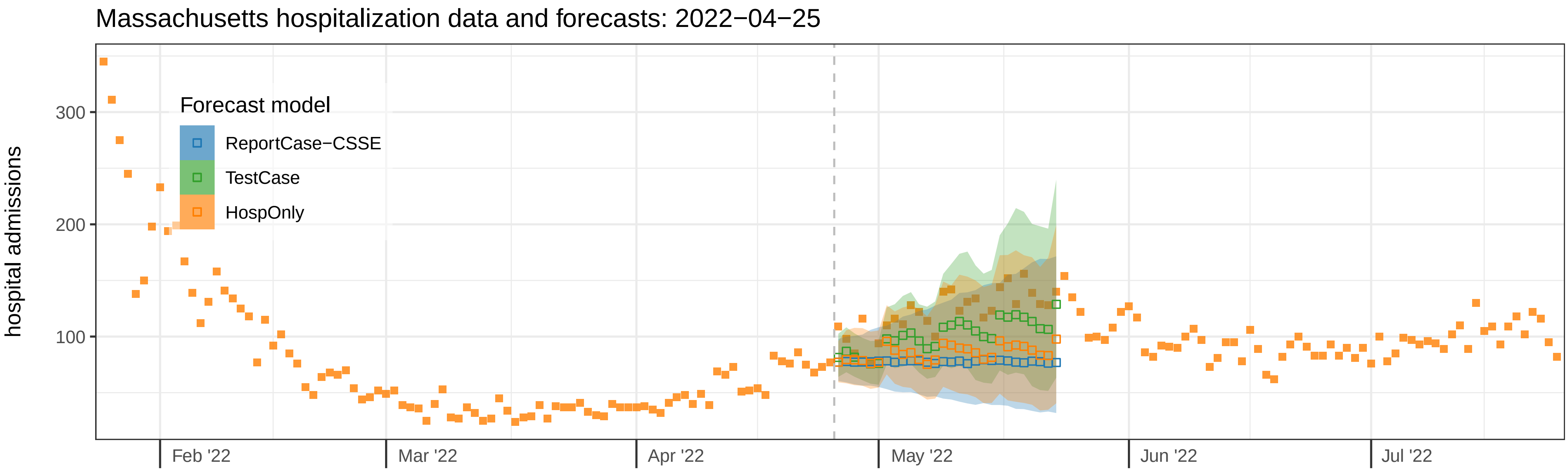
Massachusetts hospitalization data and forecasts: 2022−04−25
Forecast model
ReportCase−CSSE
TestCase
HospOnly
hospital admissions
300
200
100
Feb '22
Mar '22
Apr '22
May '22
Jun '22
Jul '22

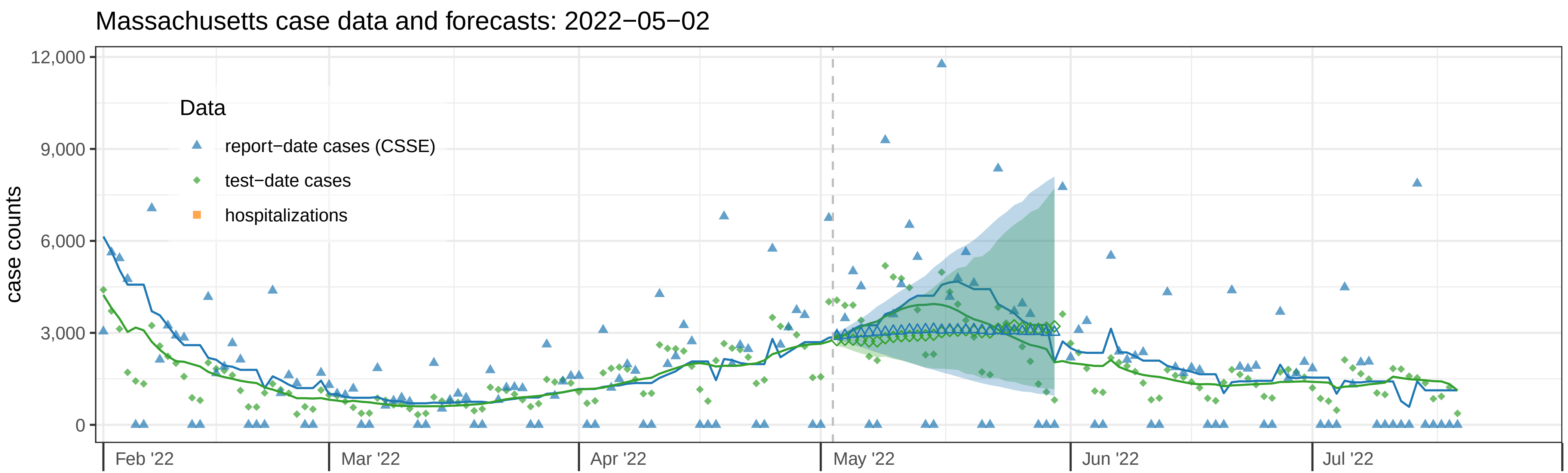
Massachusetts case data and forecasts: 2022−05−02
case counts
12,000
9,000
6,000
3,000
0
Data
report−date cases (CSSE)
test−date cases
hospitalizations
Feb '22
Mar '22
Apr '22
May '22
Jun '22
Jul '22

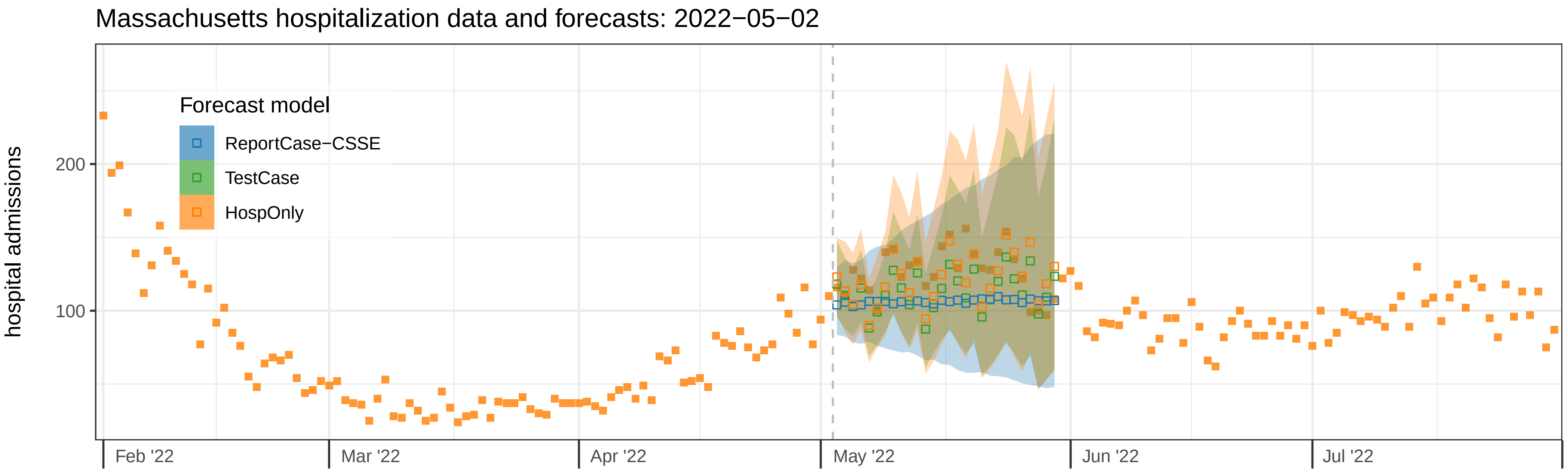
Massachusetts hospitalization data and forecasts: 2022−05−02
hospital admissions
200
100
Forecast model
ReportCase−CSSE
TestCase
HospOnly
Feb '22
Mar '22
Apr '22
May '22
Jun '22
Jul '22

Massachusetts case data and forecasts: 2022−05−09

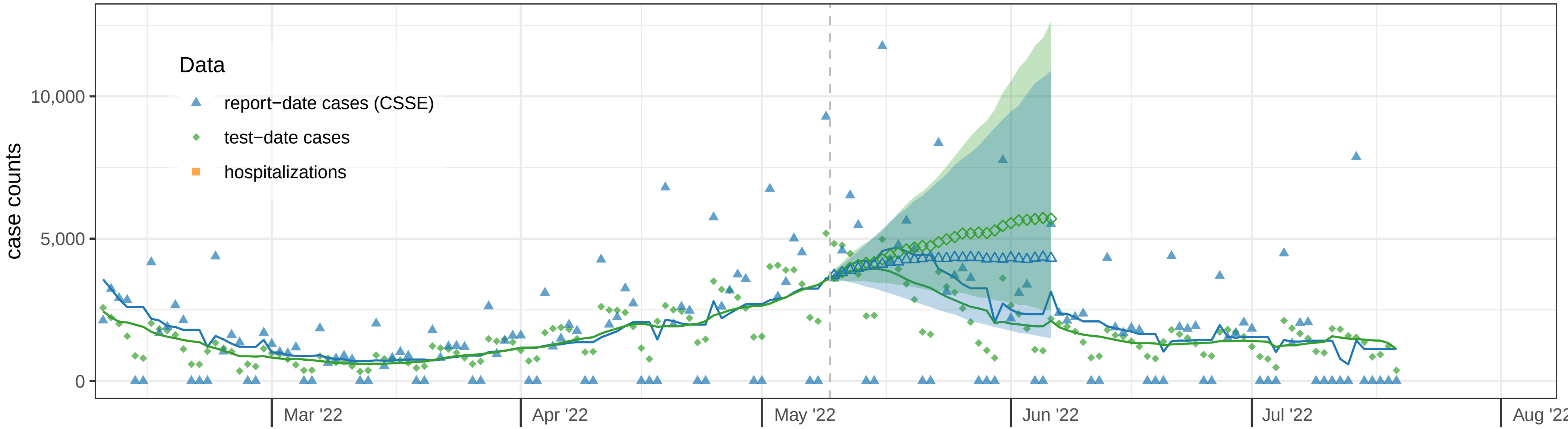


Massachusetts hospitalization data and forecasts: 2022−05−09

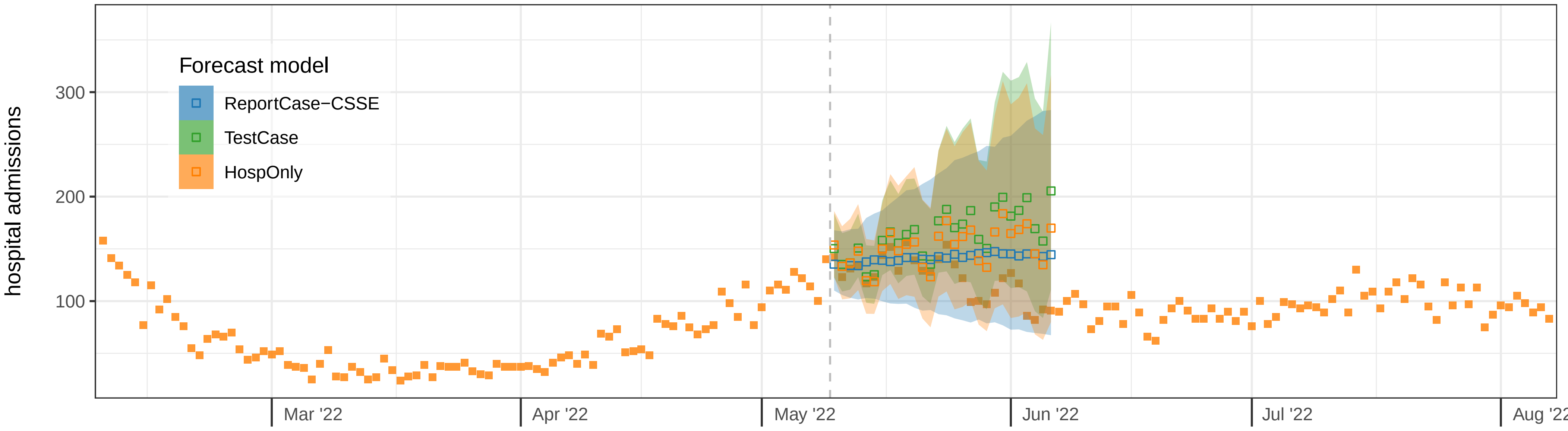

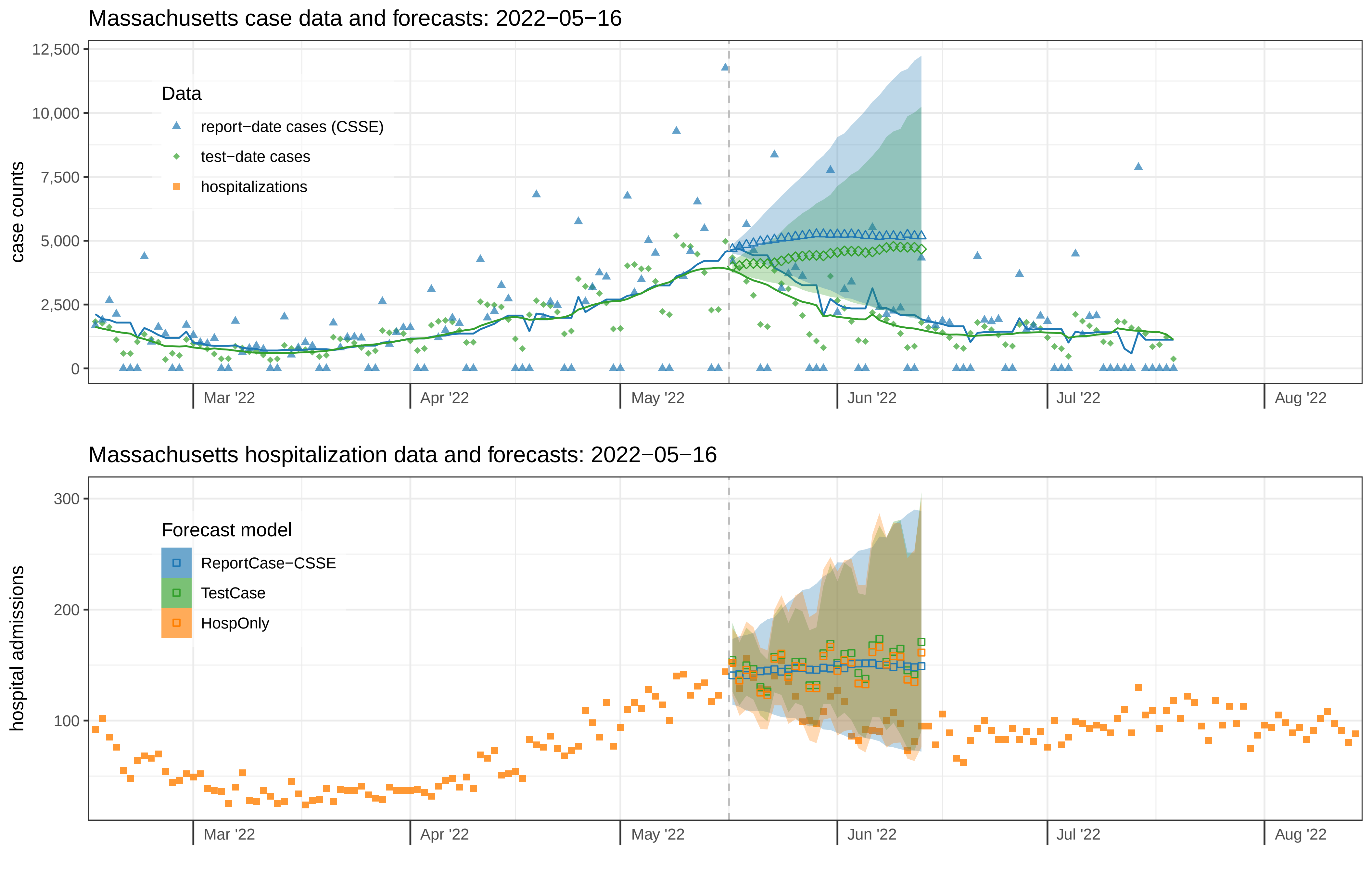

Massachusetts case data and forecasts: 2022−05−16
case counts
12,500
10,000
7,500
5,000
2,500
0
Data
report−date cases (CSSE)
test−date cases
hospitalizations
Mar '22
Apr '22
May '22
Jun '22
Jul '22
Aug '22
Massachusetts hospitalization data and forecasts: 2022−05−16
hospital admissions
300
200
100
Forecast model
ReportCase−CSSE
TestCase
HospOnly
Mar '22
Apr '22
May '22
Jun '22
Jul '22
Aug '22

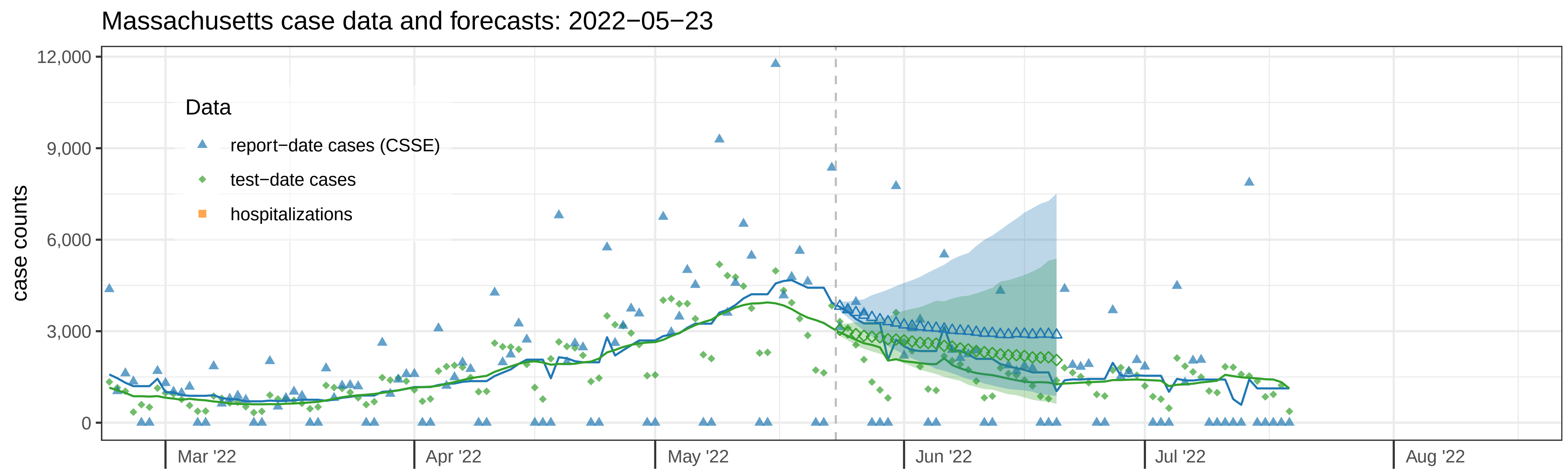

Massachusetts case data and forecasts: 2022−05−23
case counts
12,000
9,000
6,000
3,000
0
Data
report−date cases (CSSE)
test−date cases
hospitalizations
Mar '22
Apr '22
May '22
Jun '22
Jul '22
Aug '22


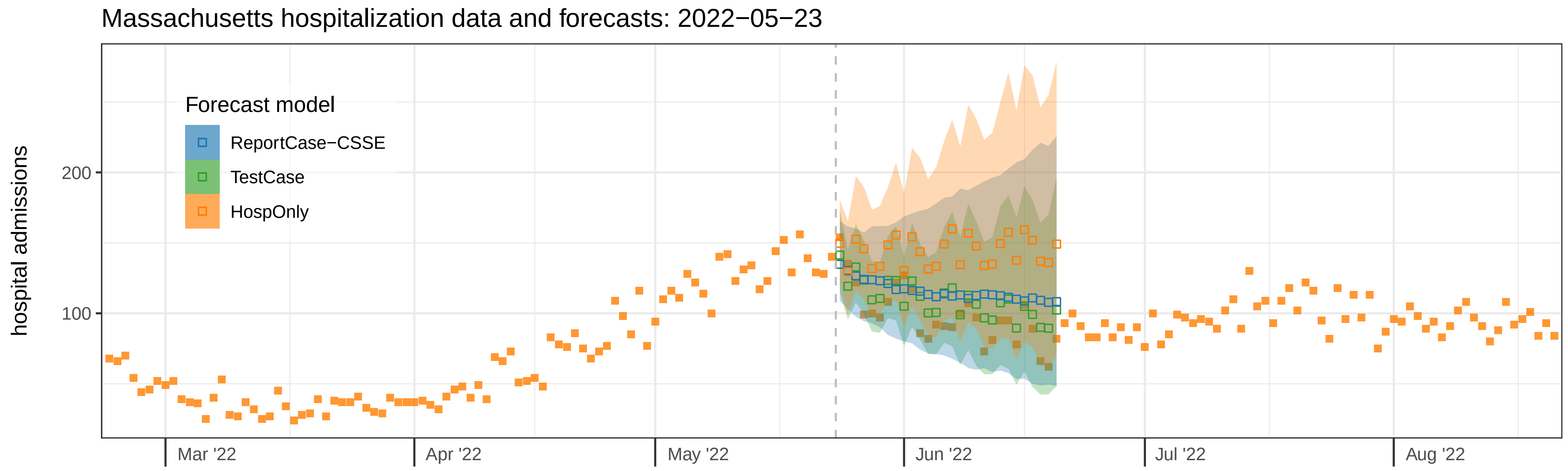

Massachusetts hospitalization data and forecasts: 2022−05−23
hospital admissions
200
100
Forecast model
ReportCase−CSSE
TestCase
HospOnly
Mar '22
Apr '22
May '22
Jun '22
Jul '22
Aug '22

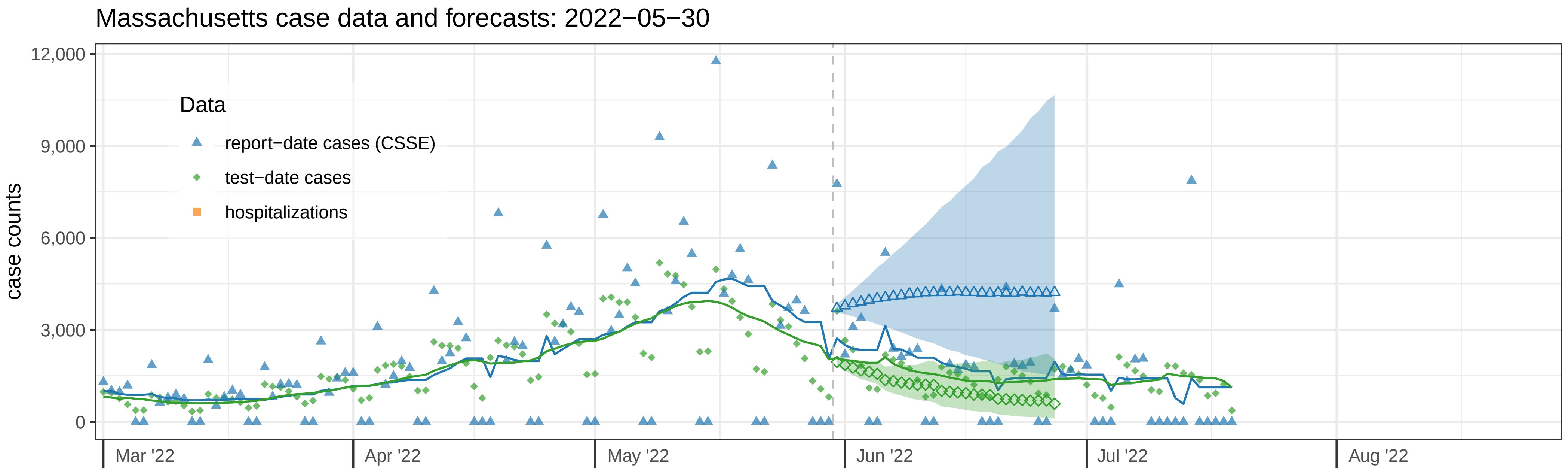

Massachusetts case data and forecasts: 2022−05−30
case counts
12,000
9,000
6,000
3,000
0
Data
report−date cases (CSSE)
test−date cases
hospitalizations
Mar '22
Apr '22
May '22
Jun '22
Jul '22
Aug '22


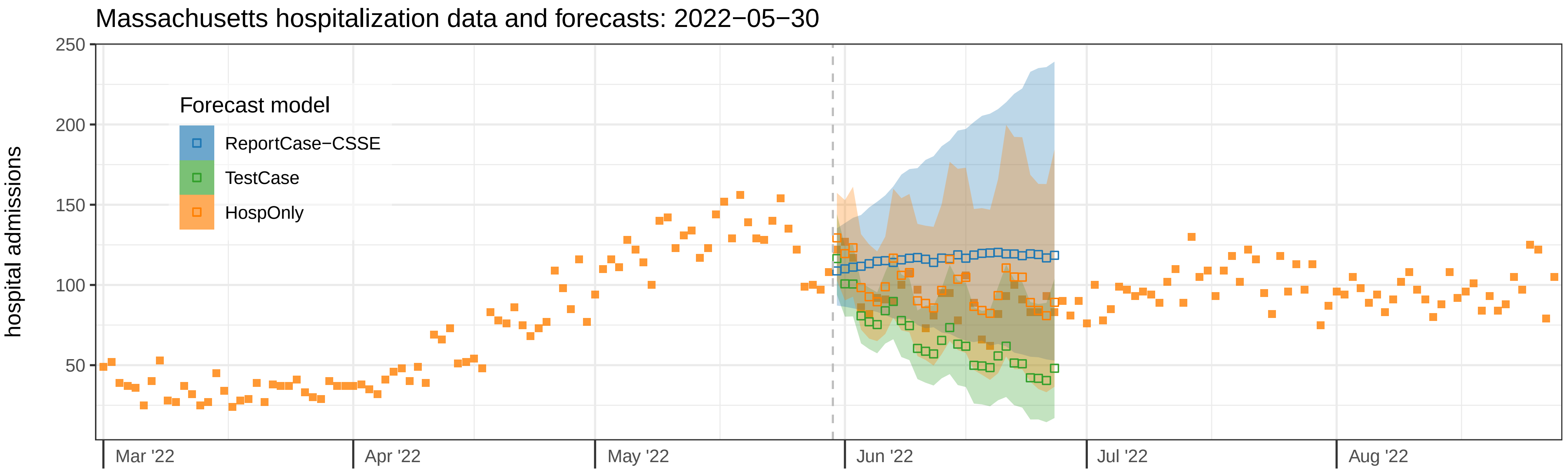

Massachusetts hospitalization data and forecasts: 2022−05−30
hospital admissions
250
200
150
100
50
Forecast model
ReportCase−CSSE
TestCase
HospOnly
Mar '22
Apr '22
May '22
Jun '22
Jul '22
Aug '22
