## Supplemental File 3 for "Assessing the utility of COVID-19 case reports as a leading indicator for hospitalization forecasting in the United States"

### Supplemental File 3 for
### "Assessing the utility of COVID-19 case reports as a leading indicator for hospitalization forecasting in the United States"

Reich et al.

March 8, 2023

#### 1 EPIFORGE 2020 reporting items

We include here in Table 1 the recommended reporting items for epidemic forecasting and prediction research.[1]

| Section of Manuscript | # | Checklist item | Reported in section |
|---|---|---|---|
| Title/Abstract | 1 | Describe the study as forecast or prediction research in at least the title or abstract | Title |
| Introduction | 2 | Define the purpose of study and forecasting targets | Introduction |
| Methods | 3 | Fully document the methods | Methods |
| Methods | 4 | Identify whether the forecast was performed prospectively, in real time, and/or retrospectively | Methods: Data vintage used in modeling |
| Methods | 5 | Explicitly describe the origin of input source data, with references | Methods: Data |
| Methods | 6 | Provide source data with publication, or document reasons as to why this was not possible | Methods: Data and code availability |
| Methods | 7 | Describe input data processing procedures in detail | Methods: Models |
| Methods | 8 | State and describe the model type, and document model assumptions, including references | Methods: Models |
| Methods | 9 | Make the model code available, or document the reasons why this was not possible | Methods: Models |
| Methods | 10 | Describe the model validation, and justify the approach | Methods: Forecast validation and testing periods |
| Methods | 11 | Describe the forecast accuracy evaluation method used, with justification | Methods: Forecast structure and evaluation |
| Methods | 12 | Where possible, compare model results to a benchmark or other comparator model, with justification of comparator choice | Methods: Models |
| Methods | 13 | Describe the forecast horizon, with justification of its length | Methods: Models |
| Results | 14 | Present and explain uncertainty of forecasting results | Results: Comparative forecast model results using different case data |
| Results | 15 | Briefly summarize the results in nontechnical terms, including a nontechnical interpretation of forecast uncertainty | Results: Comparative forecast model results using different case data |
| Results | 16 | If results are published as a data object, encourage a time-stamped version number | Methods: Data and code availability |
| Discussion | 17 | Describe the weaknesses of the forecast, including weaknesses specific to data quality and methods | Discussion |
| Discussion | 18 | If the research is applicable to a specific epidemic, comment on its potential implications and impact for public health action and decision making | Discussion |
| Discussion | 19 | If the research is applicable to a specific epidemic, comment on how generalizable it may be across populations | Discussion |

Table 1: EPIFORGE 2020 checklist reporting items.[1]

#### 2 Data revisions

Data for both hospitalizations and cases are sometimes revised by the surveillance system after initially being reported. The revisions can sometimes be substantial and occur as much as weeks or months after the data were initially reported. In the main manuscript we used “finalized” versions of the data.

To be clear about our usage of different dates, we will refer to the ‘event date’ as the date on which a particular event (e.g., a hospital admission or a case report) occurs, and the ‘issue date’ as the date on which a particular set of observations are released by a public health agency.

To provide a summary of the scale of the data revisions, we computed, for counts of new cases or hospitalizations on every event date, the ratio of the reported value on every issue date to the value as reported on the final issue date of July 26, 2022. We let $y_{t,d}$ refer to the observation of a particular data source associated with event date $t$ that was available on issue date $d$, then we compute a revision ratio $r_{t,d} = \frac{y_{t,d}}{y_t}$, where $y_t$ is the version of $y_{t,d}$ that was available on July 26, 2022. Therefore, values of $r_{t,d}$ less than one indicate that the issue of the data at date $d$ was lower than the eventual final value, and revisions would increase the observed counts. Whereas values of $r_{t,d}$ greater than one indicate that the current issue of the data as of date $d$ was higher than the eventual final value and revisions would bring the observed counts down. Boxplots of the revision ratios are shown in Figure 1, showing revisions to report-date cases from JHU CSSE for California.

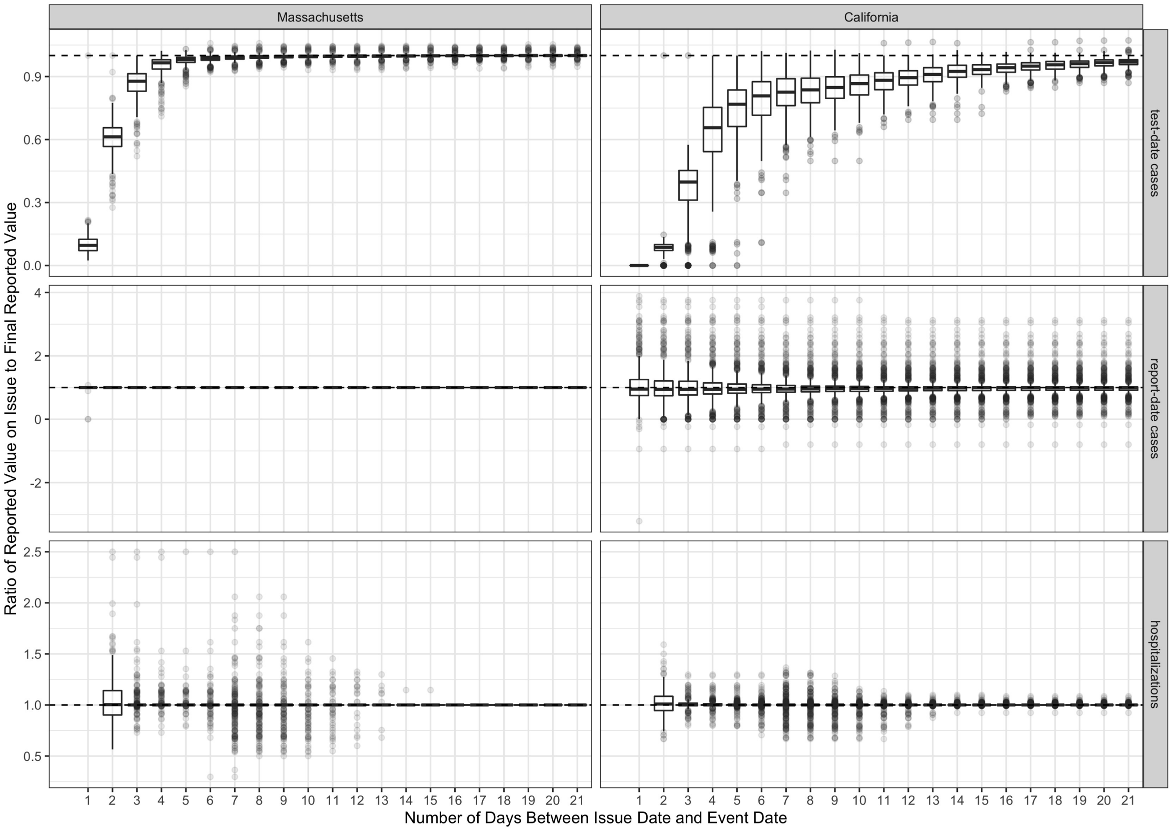


Figure 1: Boxplots of revision ratios as a function of time between the issue date for an observation and the event date. The left-hand column of plots show revision ratios for Massachusetts whereas the right-hand column show plots for California. Each row represents a different data source, from top to bottom: test-date cases, report-date cases (from JHU CSSE) and hospitalizations. The patterns of reporting vary by state and data type. For reference, each figure has a horizontal dashed line drawn at $y = 1$ to illustrate where an observation is equal to its final value. Observations below the dashed line were subsequently revised upward and observations above the line were subsequently revised downwards. The x-axis represents the difference between the issue date of the observation in the numerator of the revision ratio and the event date of the observation. For example, test-date cases in both states are typically revised upwards, with the entire inter-quartile range of revision ratios showing above 90% reporting at 4 days after the event date in Massachusetts and at 15 days in California. In Massachusetts there were rarely any revisions to report-date cases, whereas in California there were occasional substantial revisions both up and down. For hospitalizations, both states showed that after three days a majority of the observations experienced no revisions, although occasionally large revisions were made up to about two weeks past the event date.

### 3 Results with non-smoothed models

In the main manuscript, only results from models that had pre-smoothed case data as inputs were shown, as those were the only models used in the test phase analysis. Table 2 shows results from models that both did and did not use smoothed case data. The rows with "TRUE" in the column named "smoothed" are identical to the results in the main manuscript table. The results with "FALSE" are supplemental, and only shown in this supplemental table. For every model-type in each location, pre-smoothing the case data improved the model accuracy. The number of models in the rank column denominator indicates the total number of models including all variations of $(p, d, P, D)$, different case data types used, and with smoothed and unsmoothed case data (when case data were used).

| | | | validation period | | | |
|---|---|---|---|---|---|---|
| case type | smoothed | (p,d,P,D) | MWIS | MAE | $PIcov^{0.95}$ | rank |
| **California** | | | | | | |
| TestCase | TRUE | (2,0,0,1) | 104.8 | 159.4 | 0.98 | 1/392 |
| TestCase | FALSE | (4,1,1,0) | 107.5 | 172.7 | 0.99 | 3/392 |
| ReportCase-CSSE | TRUE | (4,1,0,0) | 114.1 | 172.8 | 1.00 | 14/392 |
| ReportCase-DPH | TRUE | (4,1,0,0) | 115.4 | 171.9 | 0.99 | 16/392 |
| ReportCase-CSSE | FALSE | (1,0,0,1) | 121.8 | 191.1 | 1.00 | 24/392 |
| ReportCase-DPH | FALSE | (1,1,2,0) | 122.5 | 199.0 | 0.99 | 25/392 |
| HospOnly | - | (1,1,1,0) | 124.8 | 203.7 | 0.99 | 36/392 |
| **Massachusetts** | | | | | | |
| TestCase | TRUE | (1,0,1,1) | 17.19 | 26.54 | 0.99 | 1/280 |
| TestCase | FALSE | (3,0,1,1) | 17.36 | 25.07 | 0.98 | 2/280 |
| ReportCase-CSSE | TRUE | (4,1,0,0) | 18.58 | 27.37 | 0.98 | 16/280 |
| ReportCase-CSSE | FALSE | (1,0,1,1) | 18.70 | 28.21 | 0.97 | 19/280 |
| HospOnly | - | (1,0,1,1) | 19.55 | 28.70 | 0.97 | 32/280 |

Table 2: Validation period accuracy metrics for forecasts of California and Massachusetts hospital admissions, including results from models that used smoothed case data as inputs. The models shown include the best individual autoregressive models from the validation phase that used test-date data (TestCase), report-date data (ReportCase) and no case data (HospOnly) as inputs. For the models that used case data, the best models are shown that both smoothed and did not smooth that data stream. The mean weighted interval score (MWIS), mean absolute error (MAE) and 95% prediction interval coverage ($PIcov^{0.95}$) scores are shown for each model with the best scores in the validation period highlighted. Within each state, the models are sorted by highest accuracy (lowest MWIS) scores at the top. The model parameters for the auto-regressive model are also provided in the (p,d,P,D) column.
